# Vaccine safety surveillance using routinely collected healthcare data – An empirical evaluation of epidemiological designs

**DOI:** 10.1101/2021.08.09.21261780

**Authors:** Martijn J. Schuemie, Faaizah Arshad, Nicole Pratt, Fredrik Nyberg, Thamir M Alshammari, George Hripcsak, Patrick Ryan, Daniel Prieto-Alhambra, Lana YH Lai, Xintong Li, Fortin Stephen, Evan Minty, Marc A. Suchard

**Affiliations:** Observational Health Data Sciences and Informatics, New York, NY, USA; Observational Health Data Analytics, Janssen R&D, Titusville, NJ, USA; Department of Biostatistics, University of California, Los Angeles, CA, USA; Quality Use of Medicines and Pharmacy Research Centre, Clinical and Health Sciences, University of South Australia, Australia; School of Public Health and Community Medicine, Institute of Medicine, Sahlgrenska Academy, University of Gothenburg, Gothenburg, Sweden; College of Pharmacy, Riyadh Elm University, Riyadh, Saudi Arabia; Real-World Evidence, Trial Form Support, Barcelona, Spain; Department of Biomedical Informatics, Columbia University, NY, NY, USA; Centre for Statistics in Medicine, NDORMS, University of Oxford, Oxford, UK; Department of Medical Informatics, Erasmus University Medical Center, Rotterdam, NL; O’Brien Institute for Public Health, Faculty of Medicine, University of Calgary, Canada; Division of Medical Sciences, University of Manchester, UK; Department of Human Genetics, University of California, Los Angeles, CA, USA

**Keywords:** vaccine safety, routinely collected data, adverse event, surveillance

## Abstract

**Background:** Routinely collected healthcare data such as administrative claims and electronic health records (EHR) can complement clinical trials and spontaneous reports when ensuring the safety of vaccines, but uncertainty remains about what epidemiological design to use.

**Methods:** Using 3 claims and 1 EHR database, we evaluate several variants of the case-control, comparative cohort, historical comparator, and self-controlled designs against historical vaccinations with real negative control outcomes (outcomes with no evidence to suggest that they could be caused by the vaccines) and simulated positive controls.

**Results:** Most methods show large type 1 error, often identifying false positive signals. The cohort method appears either positively or negatively biased, depending on the choice of comparator index date. Empirical calibration using effect-size estimates for negative control outcomes can restore type 1 error to close to nominal, often at the cost of increasing type 2 error. After calibration, the self-controlled case series (SCCS) design shows the shortest time to detection for small true effect sizes, while the historical comparator performs well for strong effects.

**Conclusions:** When applying any method for vaccine safety surveillance we recommend considering the potential for systematic error, especially due to confounding, which for many designs appears to be substantial. Adjusting for age and sex alone is likely not sufficient to address the differences between vaccinated and unvaccinated, and for the cohort method the choice of index date plays an important role in the comparability of the groups Inclusion of negative control outcomes allows both quantification of the systematic error and, if so desired, subsequent empirical calibration to restore type 1 error to its nominal value. In order to detect weaker signals, one may have to accept a higher type 1 error.

**Highlights:**

- Most methods used in vaccine safety surveillance show large type 1 error, which could lead to many false safety signals.
- Empirical calibration using effect-size estimates for negative control outcomes can restore type 1 error to close to nominal, often at the cost of marginal increases in type 2 error.
- After calibration, the self-controlled case series (SCCS) design shows the shortest time to detection for small true effect sizes, while the historical comparator appears best for large true effect sizes.
- Implementing negative control outcomes in a safety surveillance system is recommended to identify vulnerability to systematic error.

## 1. Introduction

Vaccines are a critical part of the public health response to communicable disease. Given the extensive rate of vaccination among the general population, including otherwise healthy individuals, considerable emphasis is placed on ensuring vaccine safety. Post-marketing safety surveillance, therefore, is critical to ensure rare events not detectable in pre-market clinical trials due to limited sample size are detected early to maintain public confidence. Regulators frequently rely on spontaneous reports of safety concerns with vaccines; however, these data are often underreported[1] and lack accurate denominator information for calculation of population rates of disease.[2] It is, therefore, of critical importance that alternate data are used to implement rapid and rigorous identification of safety signals associated with vaccines.

Routinely collected healthcare data such as administrative claims and electronic health records (EHRs) may offer information that is complementary to clinical trials and spontaneous reports. Although these data were not collected for the purpose of safety surveillance, several epidemiological designs for analyzing longitudinal healthcare data to this end have been proposed. Some of these methods were proposed as ‘signal generation’ methods, while others have been considered for ‘signal confirmation’,[3] where the main distinction seems to be the complexity of the method and its ability to scale from a single vaccine-outcome pair to many. However, recent developments in large-scale analytics such as large-scale propensity scores (PS),[4] and open-source analytics software like that developed by the Observational Health Data Sciences and Informatics (OHDSI),[5] mean the distinction between signal generation and confirmation has become blurred. It is, for example, perfectly feasible to use propensity score adjustment on a large scale, and, therefore, to use it efficiently for signal generation.

Given the current coronavirus disease 2019 (COVID-19) pandemic and mass vaccination, there is great interest in how best to monitor the safety of these vaccines. Prior research has shown that the four most commonly used epidemiological designs in vaccine safety surveillance were cohort studies, case-control studies, self-controlled case series (SCCS), and self-controlled risk-intervals (SCRI).[3, 6] One simulation study comparing the four designs indicated cohort study designs had the best performance in the sequential analysis of vaccine safety surveillance, with the lowest false positive rate, highest empirical power, and smallest risk estimate bias.[7] The SCCS and SCRI study designs were also efficient alternatives. This study, however, did not account for the potential for misclassification or confounders, such as age or seasonal effects. Another simulation study that used the cohort design as a benchmark concluded that the estimates of the case-control, SCCS and SCRI were within 5% of the true risk parameters.[8] Of the four study designs, the case-control estimates were biased by fixed confounding and less precise. While the estimates of the SCCS may be biased by unadjusted seasonal confounding, it was found to be an efficient alternative to the cohort study design, with the additional ability to avoid unmeasured between-person confounding by its self-controlled nature.

There is, however, a research gap on whether the findings from these simulation studies would apply to real-world data. To address this, we used data from three insurance claims databases and one EHR database from the United States to evaluate and compare a selection of safety surveillance methods. We studied the association between retrospective vaccinations and real outcomes assumed not to be causally related to vaccines (negative control outcomes), as well as imputed positive controls (outcomes simulated to be caused by the vaccines) to evaluate method performance.

## 2. Material and methods

### 2.1. Vaccines of interest

Our evaluation focuses on six existing (groups of) vaccines, representing different scenarios, such as a response to a pandemic, seasonal vaccinations, and continuous vaccination programs. We follow each vaccine for specific time periods (start date to end date), as shown in Table 1. For seasonal flu, we included analyses for all vaccines used during that season combined, as well as separate analyses for specific flu. For some methods, a period prior to the vaccine study period (historic start to historic end date) is used to estimate the historic incidence rate. Codes to define each vaccine group can be found in the Supplementary Materials.

**Table 1.**
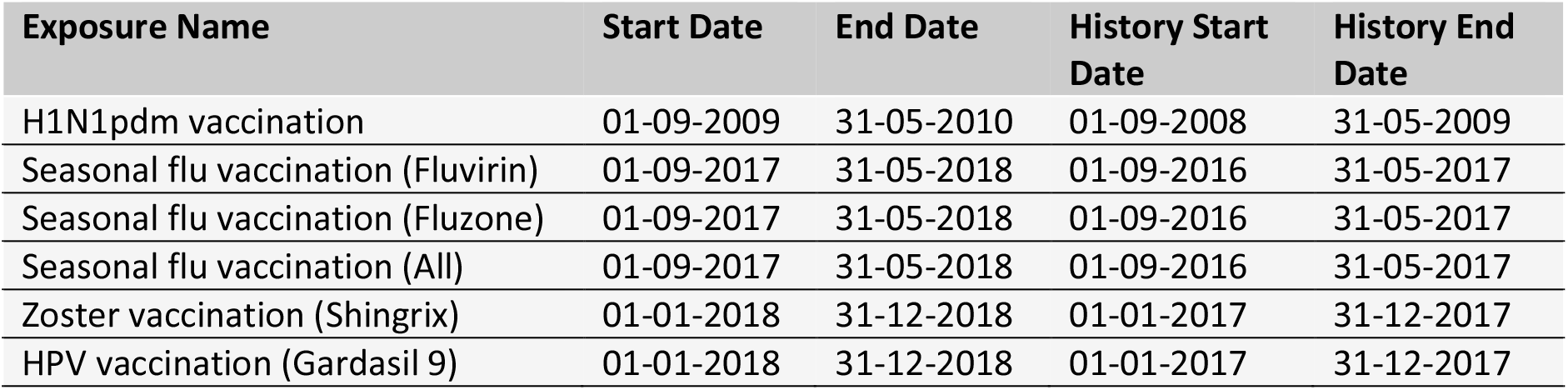
Exposures of interest.

The varicella-zoster and Human papillomavirus (HPV) vaccines require two doses. In this paper, we do not stratify by dose; anyone receiving a second dose is included in the analysis twice, with separate index dates for the two vaccinations.

### 2.2. Negative control outcomes

Negative controls are outcomes for which there is no evidence to suggest that they could be causally related to any of the vaccines, and, therefore, ideally would not be flagged as a signal by a safety surveillance system. A single set of negative control outcomes was defined for all six vaccine groups. To identify negative control outcomes that match the severity and prevalence of suspected vaccine adverse effects, a candidate list of negative controls was generated based on similarity of prevalence and percent of diagnoses that were recorded in an inpatient setting (as a proxy for severity). Manual review of this list by clinical experts created the final list of 93 negative control outcomes (see Supplementary Materials). Negative control outcomes are defined as the first occurrence of the negative control concept or any of its descendants.

### 2.3. Imputed positive control outcomes

Positive controls are outcomes known to be caused by vaccines, and ideally would be detected as signals by a safety surveillance system as early as possible. However, real positive controls are problematic for various reasons.[9] First, vaccine adverse effects that are well established are rare. Second, even when an effect is established, the magnitude is never known with precision. Third, for well-established adverse effects, actions are often taken to mitigate the risk, such as careful monitoring or even restricting use of the vaccine, masking these effects in real-world data. In our study, we therefore do not use real positive controls. To still assess the type 2 error one could expect for a given method, we instead use a simple simulation approach: for every negative control effect-size estimate produced by a method in a database, we impute three positive controls by multiplying the estimated effect size by 1.5, 2, and 4 respectively. For example, if for a negative control outcome (having true effect size = 1) a case-control design produces an odds ratio of 1.1, we can impute a positive control estimate (having true effect size = 1.5) for that design as 1.1 * 1.5 = 1.65. This simulation approach makes strong assumptions about the nature of the systematic error, most importantly that systematic error does not change as a function of true effect size.

### 2.4. Data sources

This study uses data from each of the following four observational healthcare databases:

The IBM MarketScan Commercial Claims and Encounters (CCAE) database contains adjudicated health insurance claims (e.g., inpatient, outpatient, and outpatient pharmacy) from large employers and health plans who provide private healthcare coverage to employees, their spouses and dependents.

The IBM MarketScan Medicare Supplemental Database (MDCR) database contains adjudicated health insurance claims of retirees with primary or Medicare supplemental coverage through privately insured fee-for-service, point-of-service or capitated health plans.

The IBM MarketScan Multi-State Medicaid Database (MDCD) database contains adjudicated health insurance claims for Medicaid enrollees from multiple states and includes hospital discharge diagnoses, outpatient diagnoses and procedures, and outpatient pharmacy claims.

The Optum® de-identified Electronic Health Record dataset (Optum EHR) contains clinical information, prescriptions, lab results, vital signs, body measurements, diagnoses and procedures derived from clinical notes from both inpatient and outpatient environments using natural language processing.

All data were converted to the Observational Medical Outcomes Partnership (OMOP) Common Data Model v5.3.1.

### 2.5. Evaluated method variations

The following method variations were evaluated, each using a time-at-risk (TaR) window of 1-28 days relative to the date of vaccination (both first and second dose, where applicable). For more details, see the Supplementary Materials.

- **Case-control:** The case-control design compares cases (those with the outcome) to controls (those that do not have the outcome on or before the index date), and looks back in time for exposures to a vaccine. We select up to four controls per case. We evaluate two variations:
  ◯ Age & sex matched controls, with the index date of controls set to the date of the outcome of the case to which they are matched.
  ◯ Age & sex adjusted, using random controls, with the index dates of the controls sampled from the distribution of outcome dates of the cases.
- **(Concurrent) Cohort method:** A comparative cohort study most closely emulates a randomized clinical trial, comparing the target cohort (those vaccinated) to some comparator (non-vaccinated) cohort. We define two types of comparator cohorts: one having an outpatient visit on the index date and another having a random date as the index date. For unadjusted analyses we sample a comparator cohort of equal size to the vaccinated cohort, for adjusted comparisons we sample cohorts of four times the size of the vaccinated cohort (two times for H1N1pdm) to account for loss of power due to adjustment. We exclude subjects from the comparator cohort who had a vaccination for the same disease as the target vaccine within the vaccine study period, on or before the index date. Propensity models use a large generic set of covariates, including demographics and covariates per drug, condition, procedure, measurement, etc., and are fitted using large-scale regularized regression as described previously. [10] Per-month PS matching uses only the vaccinated during that month and their comparators to create a PS and perform matching, and this matching is preserved in subsequent months. We evaluate 10 variations:
  ◯ Unadjusted, outpatient visits as comparator index
  ◯ PS matching, outpatient visits as comparator index
  ◯ Unadjusted, random days as comparator index
  ◯ PS matching, random days as comparator index
  ◯ PS stratification, outpatient visits as comparator index
  ◯ PS stratification, random days as comparator index
  ◯ PS weighting, outpatient visits as comparator index
  ◯ PS weighting, random days as comparator index
  ◯ Per-month PS matching, outpatient visits as comparator index
  ◯ Per-month PS matching, random days as comparator index The per-month PS matching variations were only executed for the H1N1 vaccines for computational reasons.
- **Historical comparator cohort design:** Traditionally, vaccine surveillance methods compute an expected count based on the incidence rate estimated during some historic time period, for example in the years prior to the initiation of the surveillance study. We use the historic period indicated in *Table 1*. Because this method proved sensitive to changes in coding practice over time, we defined variants marked as ‘filtered’ below which we filtered outcomes where the change in overall incidence rate was greater than 50% when comparing the historic period to the surveillance period so far. In total, we evaluate eight variations:
  ◯ Unadjusted, using the entire historic period
  ◯ Age & sex adjusted, using the entire historic period
  ◯ Unadjusted, using the TaR after a random outpatient visit during the historic period
  ◯ Age & sex adjusted, using the TaR after a random outpatient visit during the historic period
  ◯ Unadjusted, using the entire historic period, filtered
  ◯ Age & sex adjusted, using the entire historic period, filtered
  ◯ Unadjusted, using the TaR after a random outpatient visit during the historic period, filtered
  ◯ Age & sex adjusted, using the TaR after a random outpatient visit during the historic period, filtered
- **Self-Controlled Case Series (SCCS) / Self-Controlled Risk Interval (SCRI):** The SCCS and SCRI designs are self-controlled, comparing the TaR (the time shortly following the vaccination) to some other time in the same patient’s record. The SCCS design uses all available patient time in the vaccine study period when not at risk as the control time. [10] To account for the fact that people are either more or less likely to be vaccinated directly after a serious health outcome, the 30 days prior to vaccination are removed from the analysis. Adjustment for age and season uses 5-knot bicubic splines. The SCRI design uses a pre-specified control interval relative to the vaccination date as the control time. [9] This unexposed time can be either before or after the TaR. We evaluate five variations:
  ◯ Unadjusted SCCS excluding a 30-day pre-vaccination window
  ◯ Age & season adjusted SCCS excluding a pre-vaccination window
  ◯ SCRI with a control interval of 43 to 15 days prior to vaccination
  ◯ SCRI with a control interval of 43 to 71 days after to vaccination
  ◯ Unadjusted SCCS excluding all pre-vaccination time, so including all time after day 28 (the end of the TaR) as control time.

To adjust for multiple testing when testing the same hypothesis sequentially we apply maximum sequential probability ratio testing (MaxSPRT) to all methods by computing the log likelihood ratio (LLR) as well as a critical value for the observed power and alpha of 0.05.[11] Critical values were computed using the ‘Sequential’ R package version 3.3.1, using the Poisson model for the historical comparator design, and the binomial model for all other designs.[12]

To adjust for systematic error, we apply an empirical calibration procedure described elsewhere [13, 14] that attempts to restore p-values and LLRs to nominal (e.g. ensuring that after calibration approximately 5% of negative controls have p < 0.05). In short, this procedure first estimates the distribution of systematic error, assumed to be Gaussian, using the observed estimates for negative controls. Using the estimated distribution, we then generate calibrated p-values and LLRs considering both random and systematic error. Typically, but not necessarily, the calibrated p is higher than the nominal p, reflecting the problems unaccounted for in the standard procedure (such as unmeasured confounding, selection bias, and measurement error) but accounted for in the calibration. For the purpose of this evaluation we apply a leave-one-out approach, calibrating the estimate for a control outcome using the systematic error distribution fitted on all control outcomes except the one being calibrated.

### 2.6. Performance metrics

To evaluate timeliness, time was divided into one-month periods, where for each month the data collected in that month and those preceding it were used to compute estimates. For each database – vaccine group – outcome – method – period combination we compute the uncalibrated and calibrated effect-size estimate (odds ratio, hazard ratio or incidence ratio) with 95% confidence interval (CI) and one-sided p-value, as well the LLR and MaxSPRT critical value.

Based on these statistics, for each database – vaccine group – method, we derive the systematic error distribution for negative controls, [14] type 1 error (how often the null is rejected when the null is true), type 2 error (how often the null is not rejected when the null is not true), and time (months) to 50% sensitivity (i.e., 50% of positive controls flagged as statistically significant) stratified by true effect size. Note that we changed this metric from the initially intended 80% sensitivity, which corresponds to a more common target, to 50% sensitivity, because almost no method and database achieved 80% sensitivity in the study period. Plots showing time to 80% sensitivity are included in the Supplementary Materials.

### 2.7. Open science

The protocol as well as the analytic source code used to execute this study are available at https://github.com/ohdsi-studies/Eumaeus. The protocol has also been registered at ENCEPP with registration number EUPAS40259.

## 3. Results

Where feasible, we executed all 25 method variations on all six vaccine groups, negative and positive controls, and time periods against the four databases, briefly characterized in Table 2, thus producing a total of 1,380,672 effect size estimates. From these we derive a large set of performance metrics, which vary depending on choices of which control outcomes and data to include in the evaluation. Below we present several examples of our results, starting with a single control, single database and two analysis variants, and gradually increasing the complexity. However, it is infeasible to cover the full set of results, and instead we refer the reader to the Supplementary Materials.

**Table 2.**
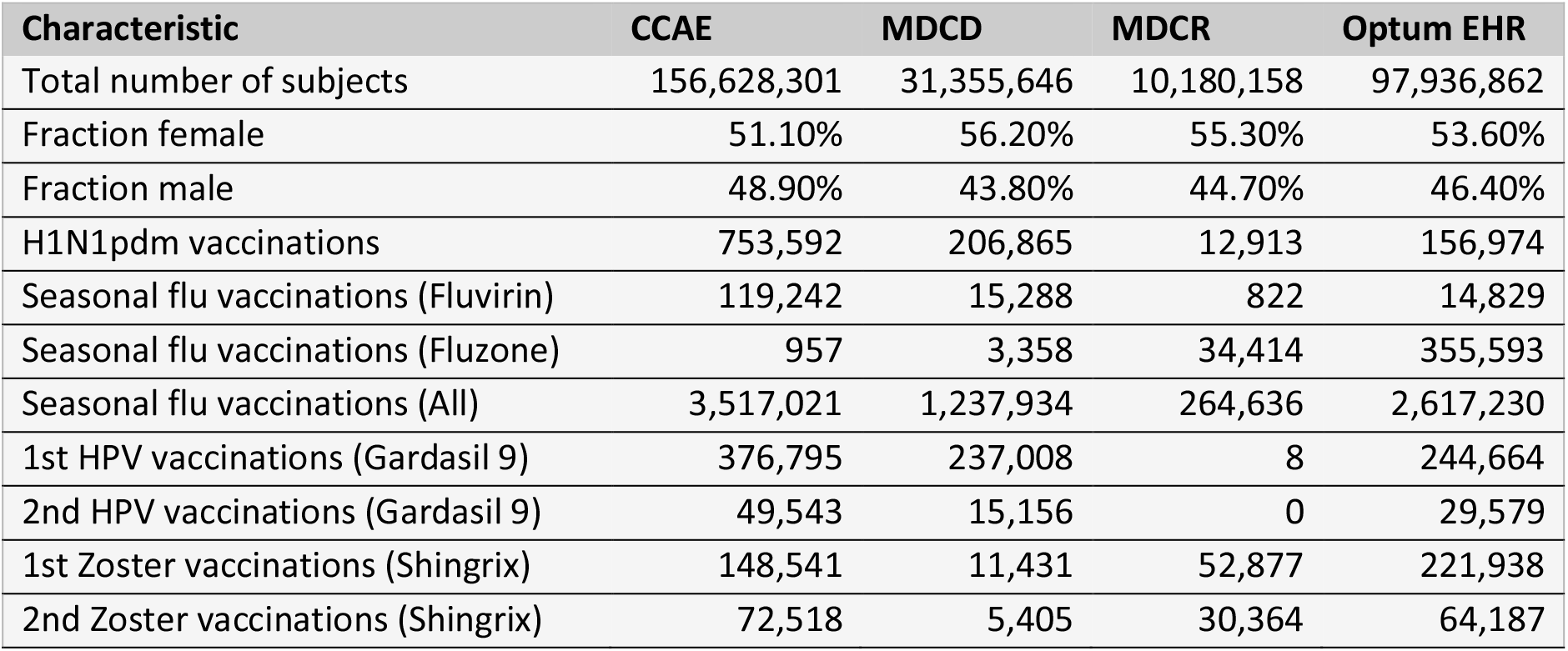
Database characteristics and vaccination counts during the vaccination study period.

### 3.1. An example control outcome for one vaccine

We illustrate our experiment with a single negative control outcome: H1N1pdm vaccines and contusion of toe. We study this relationship in the Optum EHR database, using all 9 months of data in our study period (September 2009 to May 2010), during which we observe 156,467 vaccinations. Our TaR is 1-28 days relative to the date of vaccination.

We can estimate the effect size using an unadjusted historical comparator design, using the entire historic period (September 2008 to May 2009) to estimate the background rate. During this historic period, the observed incidence rate (IR) in the Optum EHR database is 0.29 per 1,000 patient years. Based on the number of H1N1pdm vaccinations and TaR, this generates an expected count of 3.4. We observe 14 cases during TaR, leading to an incidence rate ratio (IRR) of 4.08 (95% CI: 2.30-6.62) and a LLR of 9.12, which exceeds the critical value of 1.73 computed for this analysis.

Alternatively, we could estimate the effect size using the SCCS design, adjusting for age and season, and excluding a pre-vaccination window from analysis. The total number of cases observed during the study period is 2,770. For cases that were vaccinated, 116 experienced the outcome outside the TaR, and 14 experienced the outcome during the TaR, leading to an IRR of 1.07 (95% CI: 0.59-1.81) and LLR of 0.03, which does not exceed the critical value of 1.73.

These results demonstrate that using different methods to answer the same question with the same data can lead to heterogeneous estimates, and potentially to different regulatory decisions. This is further illustrated in Fig. 1, showing the effect size estimates of all method variations for our example negative control outcome (i.e. true effect size is assumed to be 1). Note that some methods were unable to produce an estimate, for various reasons. For example, the unadjusted cohort method design using a random-day comparator found no occurrences of the outcome during the TaR in the comparator.

**Fig. 1.**
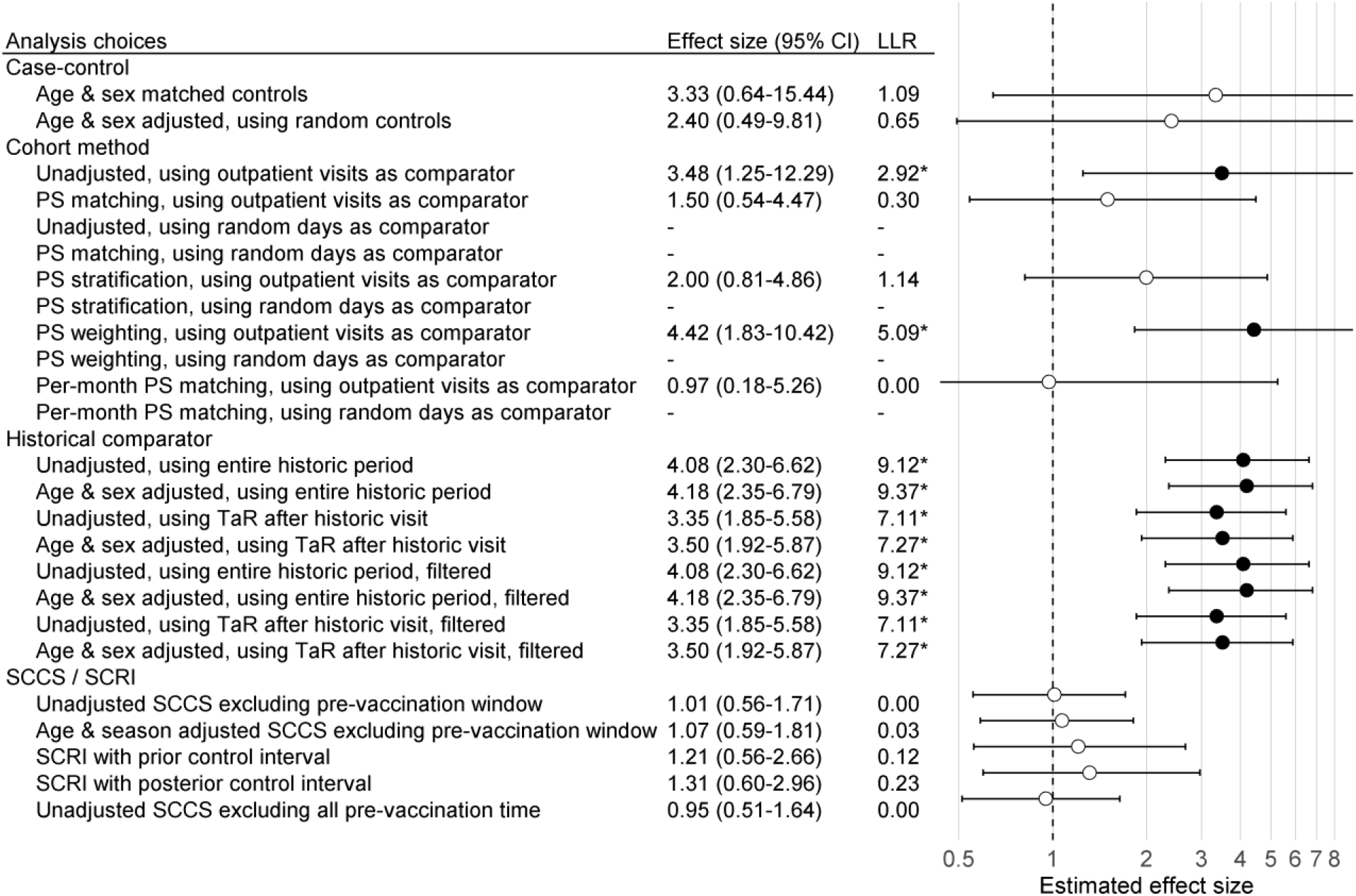
Effect size estimates, 95% CI, and LLRs for one example control. We use each analysis variation to estimate the causal effect size of H1N1pdm vaccination on the risk of ‘contusion of toe’ in the Optum EHR database, using the data across all 9 months. The true effect size is 1, as indicated by the dashed line. ‘*’ and filled dots indicates the LLR exceeds the critical value. CI = Confidence Interval, LLR = Log Likelihood Ratio, TaR = Time-at-Risk.

### 3.2. Extending the example to all negative control outcomes for one vaccine

This process was repeated for all negative control outcomes. The top row in Fig. 2 shows a compact representation of the effect size estimates of four example method variations for the negative controls, where the true effect size is assumed to be 1.

**Fig. 2.**
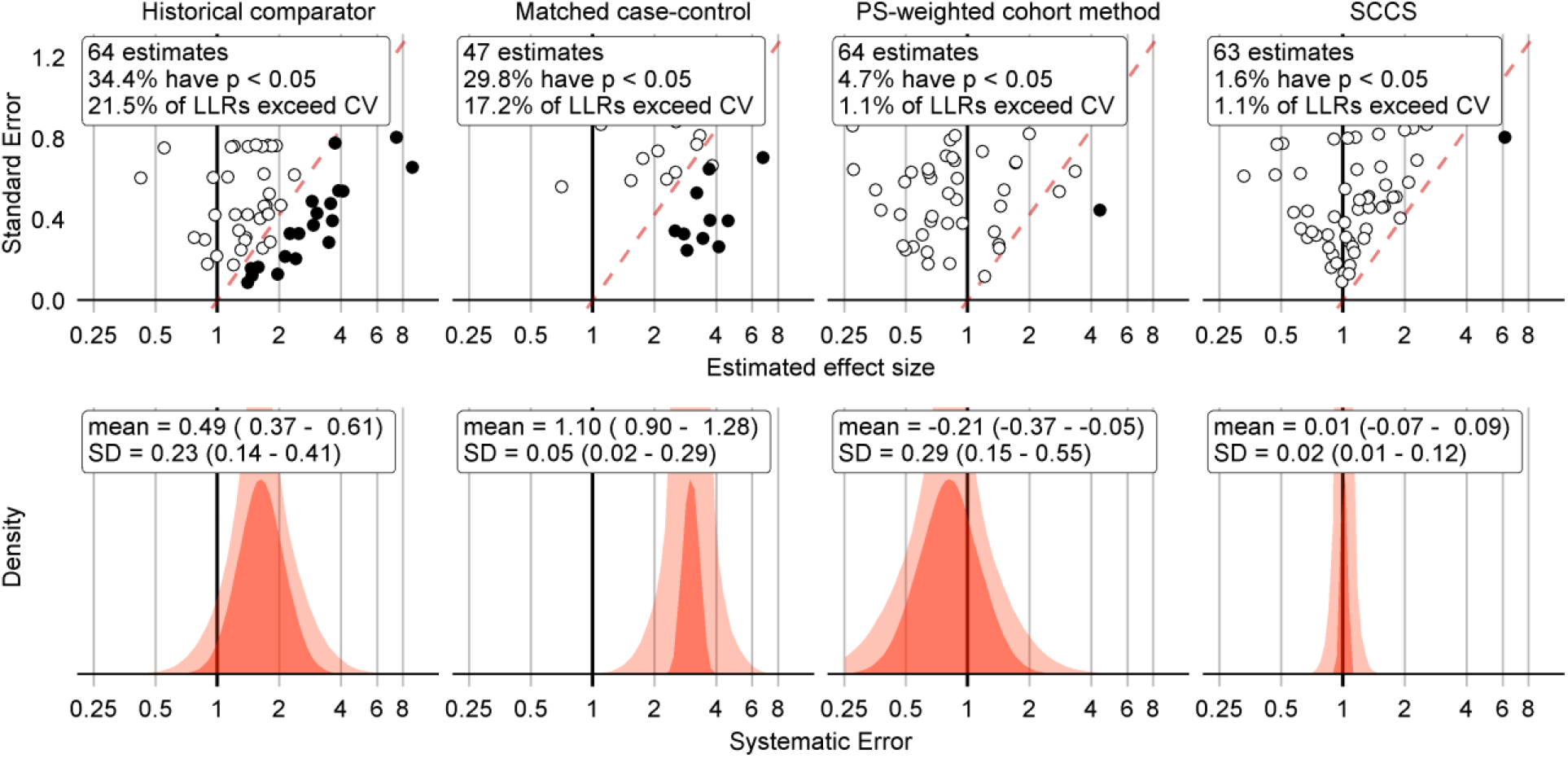
Negative control outcome effect size estimates and fitted systematic error distributions for four example method variations. In the top row, dots indicate the estimated effect size (x-axis) and corresponding standard error (y-axis), which is linearly related to the width of the confidence interval. Estimates below the red dashed line have a one-sided p-value < 0.05, and filled dots indicate the LLR exceeds the CV. The bottom row shows the systematic error distributions fitted using the negative control estimates above, for the maximum likelihood estimates of the parameters (red area), and the 95% credible interval (pink area). The historical comparator variant adjusts for age and sex, and uses the TaR after a historic outpatient visits to estimate the background rate. The case-control design matches up to 4 controls per case on age and sex. The cohort method design uses PS weighting and outpatient visits as comparator index date. The SCCS design adjusts for age and season and excludes a pre-vaccination window of 30 days from analysis. CV = Critical Value, LLR = Log Likelihood Ratio, SCCS = Self-Controlled Case Series, SD = Standard Deviation, PS = Propensity Score.

Fig. 2 shows that some methods tend to overestimate the effect size, rejecting the null when the null is assumed true more often than expected by chance alone (5% at an alpha of 0.05). Because we have a large collection of negative controls, we can use these to fit a systematic error distribution as shown in the bottom row of Fig. 2. One way to think of this distribution is that it is the distribution needed to explain the difference between the observed spread of the negative control estimates and expected spread based on random error alone. If the spread can be completely explained by random error as quantified by each method’s standard error, the systematic error distribution will have a mean and standard deviation of 0. Another way to think of this distribution is that for a future study using the same method and data, but a new outcome, the systematic error in that study will draw from this systematic error distribution.

### 3.3. Systematic error based on control outcomes across vaccines and methods

Applying this procedure to all method variations and vaccines in the Optum EHR database produces Fig. 3. For the other databases please see the Supplementary Materials. As illustrated by the plot, case-control methods and historical comparator analyses tend to be positively biased, with many negative control outcomes identified as potential safety signals in most scenarios before calibration. The cohort method appears either positively or negatively biased, depending on the choice of comparator index date. SCCS/SCRI seem less biased, with systematic error more evenly and more narrowly distributed around the null.

**Fig. 3.**
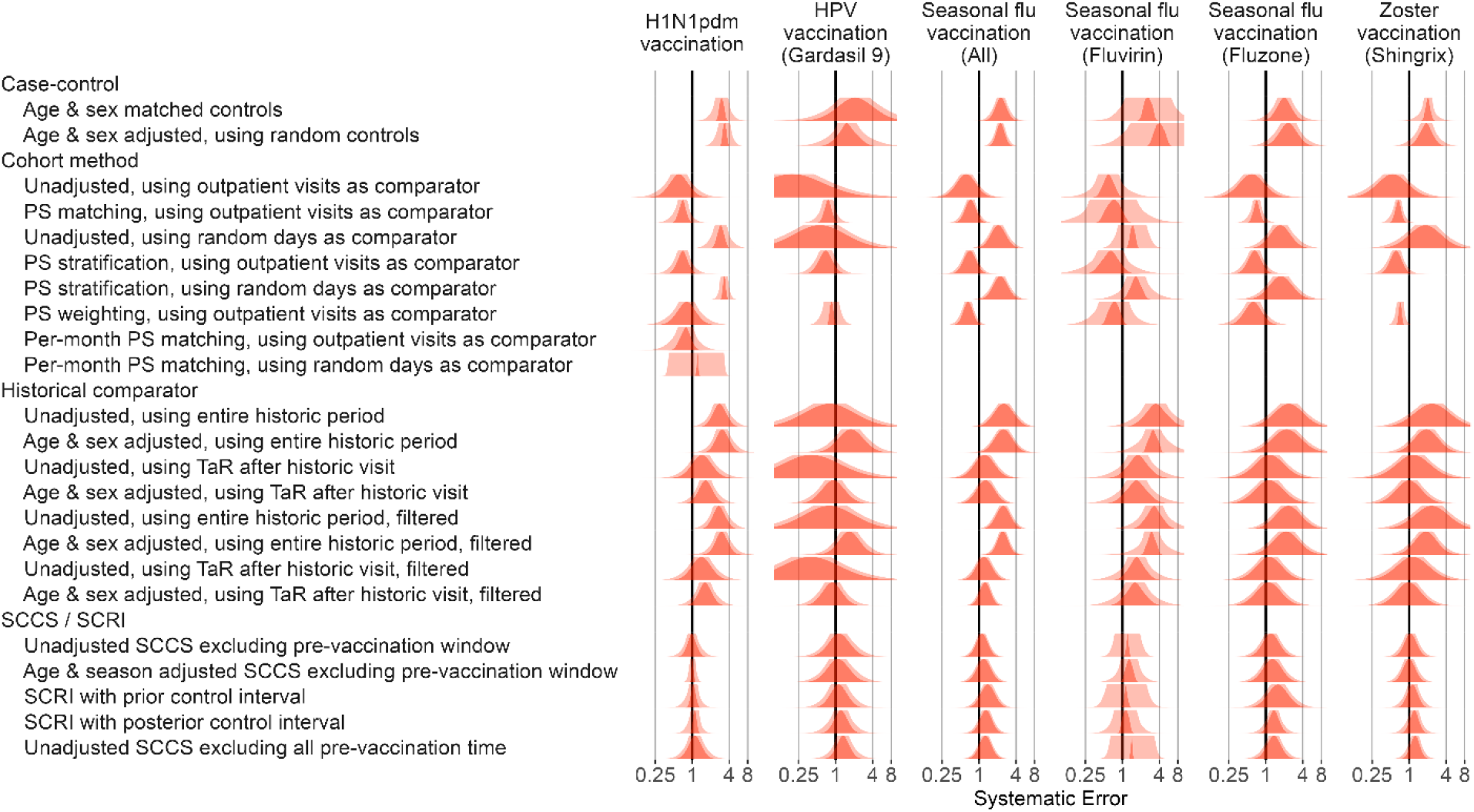
Fitted systematic error distributions. For each method variation and vaccine group, the systematic error distribution fitted on the negative control estimates in the Optum EHR database are shown. The red area indicates the maximum likelihood estimates of the distribution parameters. The pink area indicates the 95% credible interval. HPV = Human papillomavirus, PS = Propensity Score, SCCS = Self-Controlled Case Series, SCRI = Self-Controlled Risk Interval, TaR = Time-at-Risk.

### 3.4. Type 1 and 2 error tradeoffs

In addition to the systematic error inherent to a method applied to a database for a particular exposure, we consider the random error, or in other words the statistical power of a method. The estimates computed for our imputed positive control outcomes allow us to evaluate how often the null is rejected when the null is false, and thus compute type 2 error. Both type 1 and 2 error are shown in Fig. 4.

**Fig. 4.**
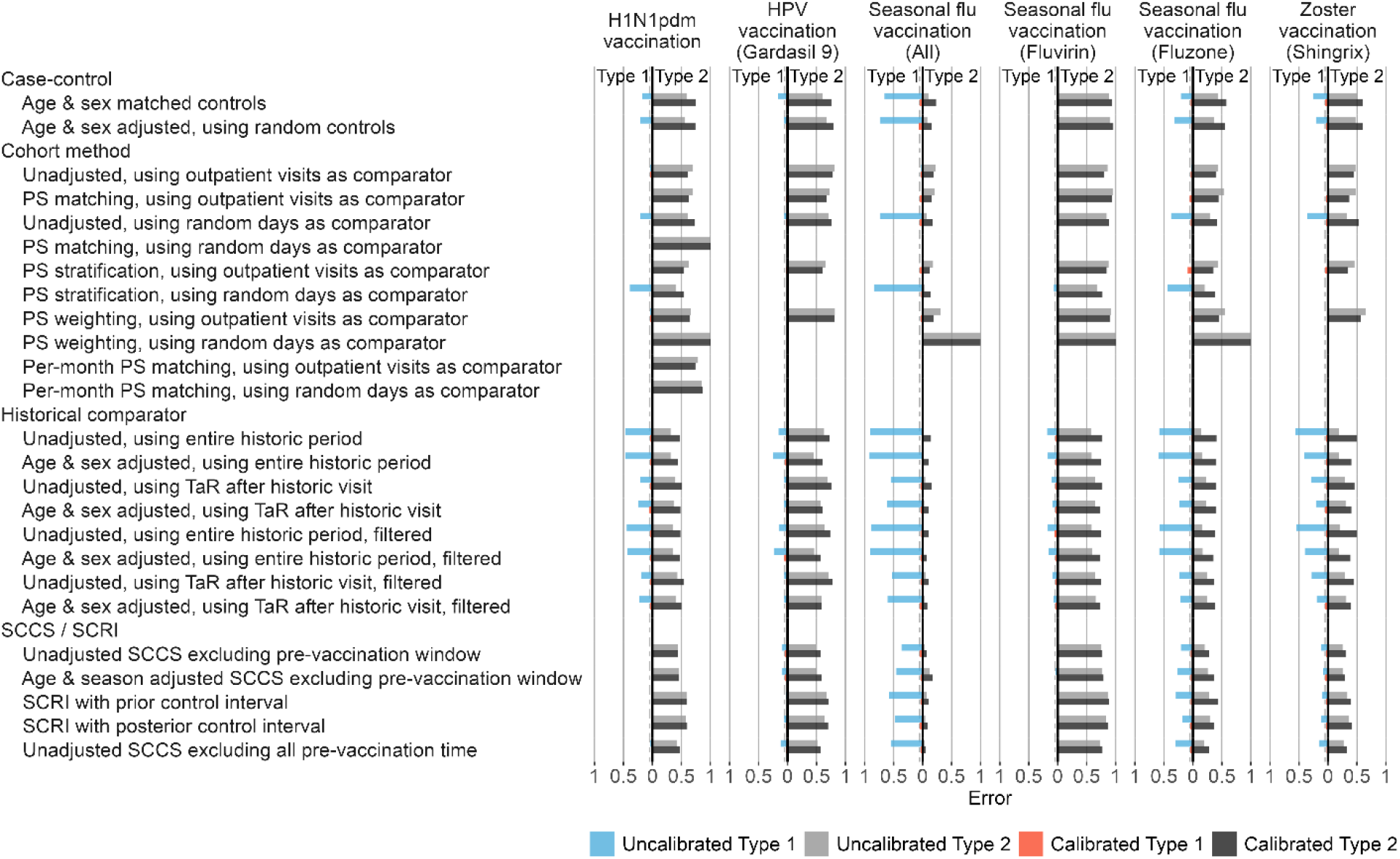
Type 1 and 2 error before and after empirical calibration. For each method variation and vaccine group, the type 1 and 2 error before and after empirical calibration in the Optum EHR database are shown. The x-axis indicates the type 1 error (higher values to the left) and type 2 error (higher values to the right), based on the (calibrated) one-sided p-value. The dashed line indicates nominal type 1 error of 5%. HPV = Human papillomavirus, PS = Propensity Score, SCCS = Self-Controlled Case Series, SCRI = Self-Controlled Risk Interval, TaR = Time-at-Risk.

Comparing type 1 and 2 error of different method variations can be complicated when both vary. Moreover, type 1 and 2 error are interchangeable, for example by changing the alpha threshold, presenting a moving target. To facilitate the comparison of methods, we apply empirical calibration, a process that uses the fitted systematic error distribution to restore the type 1 error to its nominal value (of 5% at an alpha of 0.05). [14] This typically increases type 2 error, depending on how much systematic error needs to be adjusted for in the calibration. Fig. 4 shows the type 1 and 2 error both before and after empirical calibration of the p-value. Overall, SCCS/SCRI and cohort methods provide the best combination of type 1 + type 2 error, whilst historical comparison and case-control lead to much higher type 1 error in most scenarios.

### 3.5. Timeliness

To evaluate timeliness, we divided our study periods into one-month intervals. For each month we executed the method variations on the data up to and including that month. We used MaxSPRT to account for the multiple testing of the same hypotheses each the month.[11] To facilitate comparison between methods we applied empirical calibration to restore type 1 error to nominal, and evaluate how many months must pass before type 2 error drops below 50%, in other words until 50% of the positive controls exceed our alpha threshold of 0.05. Fig. 5 depicts the results of our timeliness analyses. Overall, the SCCS analyses were the most timely methods in most scenarios. Historical comparison methods were most timely in some cases, e.g. seasonal flu vaccination, and for larger true effect sizes.

**Fig. 5.**
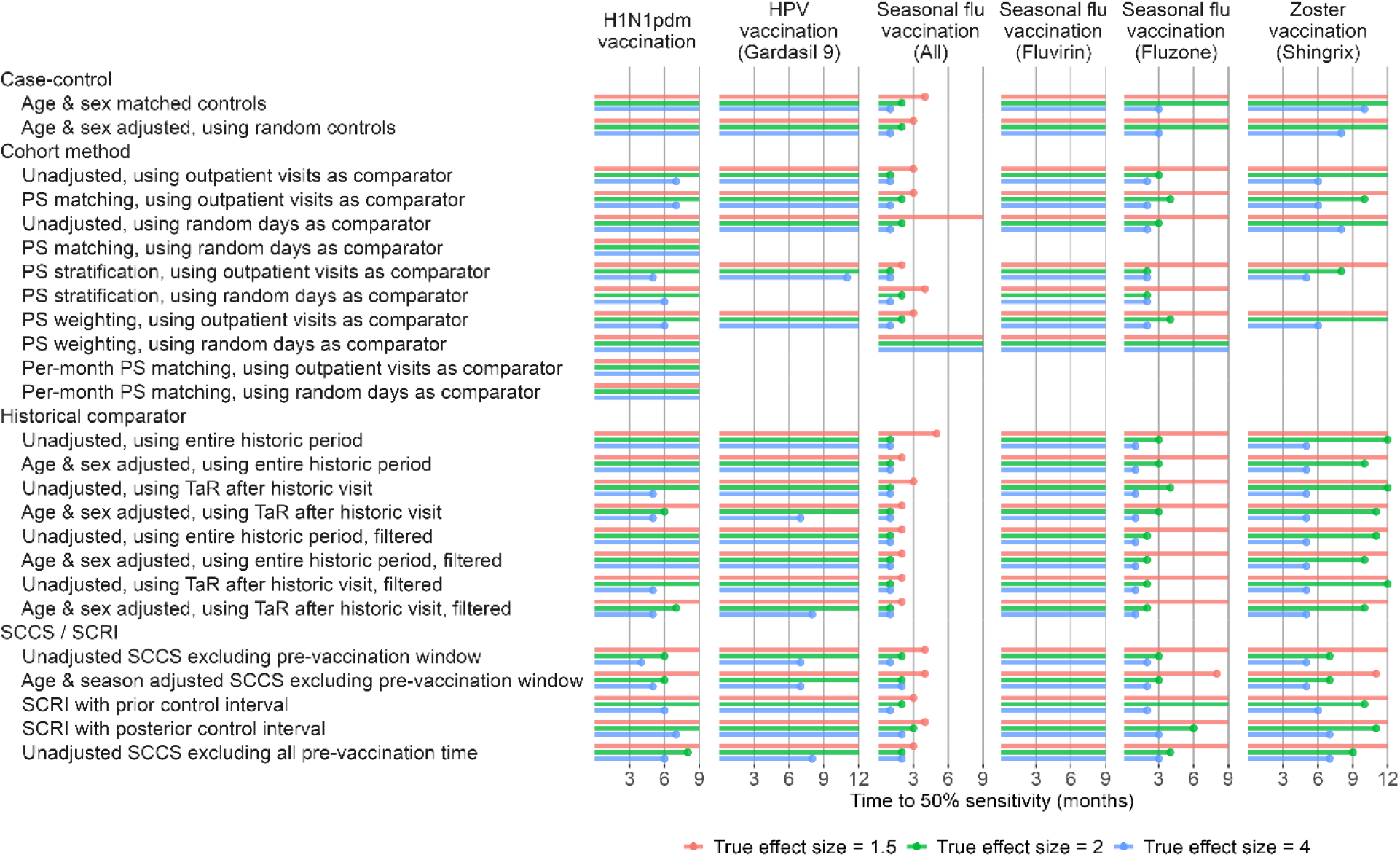
Time to 50% sensitivity after calibration. For each method variation and vaccine group, the number of months of data needed to achieve 50% sensitivity based on the calibrated MaxSPRT in the Optum EHR database are shown, stratified by true effect size of the positive controls. HPV = Human papillomavirus, PS = Propensity Score, SCCS = Self-Controlled Case Series, SCRI = Self-Controlled Risk Interval, TaR = Time-at-Risk.

## 4. Discussion

### 4.1. Key results

Most methods show large type 1 error, often rejecting the null when the null is true. This is likely due to the systematic error inherent to observational research; probably not all confounding has been adequately adjusted for, and there may be measurement error in both exposures and outcomes as well as selection biases. When using negative controls to quantify this systematic error, we found it to be substantial in the case-control, historical comparator, and unadjusted cohort method designs, but much less in the other method variations. We hypothesize that the main reason for the systematic error may be due to uncontrolled confounding, caused by important differences in the comparator group versus the vaccinated group and therefore the underlying risk of the outcomes. Many of the positively biased analyses were unadjusted or only adjusted for age and sex. It appears these analyses missed important differences in which the vaccinated group is more vulnerable to disease outcomes of any kind, including outcomes potentially not caused by the vaccine (i.e. negative control outcomes). This would explain why self-controlled designs, such as the SCCS and SCRI, are less biased, because these are less vulnerable to between-person confounding.

The cohort method appears either positively or negatively biased, depending on the choice of comparator index date. If the index date is required to have an outpatient visit, the bias is negative, probably because the comparator group at index date is sicker or at least actively participating in the health care system and being captured in the database. In contrast, when using a random day as comparator index date the bias tends to be positive, possibly because the comparator group is healthier or may not seek health care in a way captured in the data. Also interesting is the fact that the cohort method appears to remain biased even when using PS that includes not only age and sex but all other variables available in the data, suggesting important confounders are completely missing from the data.

The varying levels of type 1 error hinder the ability to compare methods. A method showing low type 2 error, flagging most positive controls as signals, may be of limited interest if it also has high type 1 error, flagging most negative controls as signals. One way to facilitate comparison is by applying empirical calibration. Empirical calibration uses the systematic error distribution inferred from the negative control estimates to restore type 1 error to its nominal value. Depending on the amount of observed systematic error, this generally leads to increased type 2 error. After applying empirical calibration, results are mixed. For large effect sizes (e.g., incidence rate ratio = 4), the historical comparator method is quickest to detect all positive controls, where this method’s efficient design appears to compensate for and overcome its inherent systematic error. Smaller effect sizes can only be detected using methods that were already fairly unbiased to begin with, such as the SCCS design. Even in large databases such as Optum EHR and CCAE, no method was clearly capable of detecting positive controls with very small outcomes at an alpha of 0.05. To allow detection of signals with such weak causal association in these databases will therefore require accepting a larger type 1 error. Larger databases or combinations of databases may lead to better detection performance, achieving shorter time to detection at lower type 1 error rates. Similarly, data with more complete information on important confounders, or data of a population where less confounding exists to begin with would improve performance, although it is unclear what type of data would meet these criteria. The four databases evaluated here showed comparable systematic error (see Supplementary Materials).

The results of our evaluation using real-world data agree to some extent to those in prior simulation studies, where SCCS was also observed to be both efficient and mostly unbiased.[7, 8] Although these prior studies indicated their unadjusted SCCS analysis was vulnerable to bias due to seasonal effects, in our study, SCCS analyses that did and did not adjust for seasonality both showed similar good performance. In contrast to the prior studies, we found the cohort method to be biased, possibly due to real data containing more unmeasured confounding than was simulated in those prior studies.

### 4.2. Strengths and limitations of this study

One of the main strengths of this study is the use of real-world data in addition to simulation of limited aspects (of positive controls), allowing the evaluation of methods in realistic scenarios. By using several different healthcare databases, we further improve the generalizability of our results.

Another strength is the inclusion of many different analysis variants, many of which have been used or proposed to be used for real vaccine safety surveillance studies. This includes more advanced designs, including SCCS adjusting for age and season using splines, and comparative cohort analyses using large-scale propensity scores.

Whereas our negative control outcomes reflect real confounding, both measured and unmeasured, as well as measurement error, our positive controls simply assume that the same systematic error also applies when the true effect size is greater than 1. In reality some forms of bias may change as a function of the true effect size, and this is not reflected in our imputed positive controls and therefore our type 2 error estimates. For example, if the TaR used in a method does not match the time when the risk of the outcome is increased by the vaccine, this can lead to bias towards the null, which is not reflected in our positive controls.

One final limitation of our study is that it included only vaccines for certain diseases in the past, and our results may not generalize to future vaccines for different diseases. For example, there are key features that are different between the situation surrounding the vaccines used as examples in this study and COVID-19 vaccines, such as the effects of nation-wide lockdowns, constraints in supply leading to highly targeted vaccinations, and decrease in regular patient care. For many vaccines, but especially for COVID-19 vaccines, vaccinations may take place outside of regular healthcare and may therefore not be completely captured in the data.

## 5. Conclusions

When applying any method for vaccine safety surveillance we recommend considering the potential for systematic error, especially due to confounding, which for many designs appears to be substantial. Adjusting for age and sex alone is likely not sufficient to address the differences between vaccinated and unvaccinated, and the choice of index date plays an important role in the comparability of the groups. Inclusion of negative control outcomes allows both quantification of the systematic error and, if so desired, subsequent empirical calibration to restore type 1 error to its nominal value. To detect weaker signals, one may have to accept a higher type 1 error, either by not calibrating (in which case type 1 error will be higher than nominal but unknown), or by calibrating and raising the alpha threshold

What levels of type 1 and 2 error are acceptable will depend on many factors. A large number of false positives may erode societal confidence in a vaccine’s safety without due cause, and exceed the downstream capacity of scientific and regulatory bodies to distinguish true positives from false positives. But false negatives, missing important safety signals, could have significant human cost.

## Data Availability

This study is part of the Evaluating Use of Methods for Adverse Event Under Surveillance (EUMAEUS) project. The protocol of this project is available at https://ohdsi-studies.github.io/Eumaeus/Protocol.html, and we publicly host the source code at (https://github.com/ohdsi-studies/Eumaeus), allowing public contribution and review, and free re-use for anyone's future research.

https://ohdsi-studies.github.io/Eumaeus/

## Abbreviations

CCAE: IBM MarketScan Commercial Claims and Encounters
CI: Confidence Interval
COVID-19: COronaVIrus Disease 2019
CV: Critical Value
H1N1pdm: Hemagglutinin Type 1 and Neuraminidase Type 1 (2009 pandemic influenza)
HPV: Human papillomavirus
LLR: Log Likelihood Ratio
MaxSPRT: Maximum Sequential Probability Ratio Testing
MDCR: IBM MarketScan Medicare Supplemental Database
MDCD: IBM MarketScan Multi-State Medicaid Database
OHDSI: Observational Health Data Sciences and Informatics
PS: Propensity Score
SCCS: Self-Controlled Case Series
SCRI: Self-Controlled Risk Interval
TaR: Time-at-Risk

## Acknowledgements

We thank Paula Casajust and Carlos Areia for helping with the literature review and reviewing the manuscript for language.

## Ethical approval

The use of Optum and IBM Marketscan databases was reviewed by the New England Institution Review Board (IRB) and was determined to be exempt from broad IRB approval, as this research project did not involve human subjects research.

## Supplementary materials

One document containing

- List of abbreviations
- Vaccine exposure definitions
- Negative control outcomes
- Data source descriptions
- Description of evaluated methods
- Fitted systematic error distributions
- Type 1 and 2 error before and after calibration
- Time to 50% sensitivity after calibration
- Time to 80% sensitivity after calibration
- Type 1 and 2 error before and after calibration, stratified by true effect size
- Type 1 and 2 error before and after calibration and when adjusting for sequential testing, stratified by true effect size
- Area Under the Receiver-Operator-Curve (AUC)
- Performance metrics based on the effect-size estimates and confidence intervals.
- Effect-size estimates for negative control outcomes.

