## Supplemental Materials for "Vaccine safety surveillance using routinely collected healthcare data – An empirical evaluation of epidemiological designs"

7 **Type 1 and 2 error before and after calibration** **22**
7.1 Type 1 and 2 error before and after calibration in the Optum EHR database 22
7.2 Type 1 and 2 error before and after calibration in the MDCD database . . 23
7.3 Type 1 and 2 error before and after calibration in the MDCR database . . 24
7.4 Type 1 and 2 error before and after calibration in the CCAE database . . 25

8 **Time to 50% sensitivity** **26**
8.1 Time to 50% sensitivity in the Optum EHR database . . . . . . . . . . . 26
8.2 Time to 50% sensitivity in the MDCD database . . . . . . . . . . . . . . . 27
8.3 Time to 50% sensitivity in the MDCR database . . . . . . . . . . . . . . . 28
8.4 Time to 50% sensitivity in the CCAE database . . . . . . . . . . . . . . . 29

9 **Time to 80% sensitivity** **30**
9.1 Time to 80% sensitivity in the Optum EHR database . . . . . . . . . . . 30
9.2 Time to 80% sensitivity in the MDCD database . . . . . . . . . . . . . . . 31
9.3 Time to 80% sensitivity in the MDCR database . . . . . . . . . . . . . . . 32
9.4 Time to 80% sensitivity in the CCAE database . . . . . . . . . . . . . . . 33

10 **Type 1 and 2 error by true effect size based on p-value** **34**
10.1 Type 1 and 2 error for H1N1 vaccination . . . . . . . . . . . . . . . . . . 34
10.2 Type 1 and 2 error for Seasonal flu vaccination (Fluvirin) . . . . . . . . . 38
10.3 Type 1 and 2 error for Seasonal flu vaccination (Fluzone) . . . . . . . . . 42
10.4 Type 1 and 2 error for Seasonal flu vaccination (All) . . . . . . . . . . . 46
10.5 Type 1 and 2 error for Zoster vaccination (Shingrix) . . . . . . . . . . . . 50
10.6 Type 1 and 2 error for HPV vaccination (Gardasil 9) . . . . . . . . . . . 54

11 **Type 1 and 2 error by true effect size based on MaxSPRT** **58**
11.1 Type 1 and 2 error for H1N1 vaccination . . . . . . . . . . . . . . . . . . 58
11.2 Type 1 and 2 error for Seasonal flu vaccination (Fluvirin) . . . . . . . . . 62
11.3 Type 1 and 2 error for Seasonal flu vaccination (Fluzone) . . . . . . . . . 66
11.4 Type 1 and 2 error for Seasonal flu vaccination (All) . . . . . . . . . . . 70
11.5 Type 1 and 2 error for Zoster vaccination (Shingrix) . . . . . . . . . . . . 74
11.6 Type 1 and 2 error for HPV vaccination (Gardasil 9) . . . . . . . . . . . 78

12 **AUC based on effect-size estimate** **82**
12.1 AUC for H1N1 vaccination . . . . . . . . . . . . . . . . . . . . . . . . . . 82
12.2 AUC for Seasonal flu vaccination (Fluvirin) . . . . . . . . . . . . . . . . 86
12.3 AUC for Seasonal flu vaccination (Fluzone) . . . . . . . . . . . . . . . . 90
12.4 AUC for Seasonal flu vaccination (All) . . . . . . . . . . . . . . . . . . . 94
12.5 AUC for Zoster vaccination (Shingrix) . . . . . . . . . . . . . . . . . . . 98
12.6 AUC for HPV vaccination (Gardasil 9) . . . . . . . . . . . . . . . . . . . 102

13 **Confidence-interval-based metrics** **106**

| | |
|---|---|
| AUC | Area Under the receiver-operator Curve |
| CCAE | IBM MarketScan Commercial Claims and Encounters |
| H1N1pdm | Hemagglutinin Type 1 and Neuraminidase Type 1 (2009 pandemic influenza) |
| HPV | Human PapillomaVirus |
| LLR | Log Likelihood Ratio |
| MDCR | IBM MarketScan Medicare Supplemental Database |
| MDCD | IBM MarketScan Multi-State Medicaid Database |
| MSE | Mean Squared Error |
| OHDSI | Observational Health Data Science and Informatics |
| OMOP | Observational Medical Outcomes Partnership |
| MaxSPRT | MAXimized Sequential Probability Ratio Test |
| PS | Propensity score |
| SCCS | Self-Controlled Case Series |
| SCRI | Self-Controlled Risk Interval |

13.1 Confidence-interval-based metrics for H1N1 vaccination . . . . . . . . . . 106
13.2 Confidence-interval-based metrics for Seasonal flu vaccination (Fluvirin) . 110
13.3 Confidence-interval-based metrics for Seasonal flu vaccination (Fluzone) . 114
13.4 Confidence-interval-based metrics for Seasonal flu vaccination (All) . . . 118
13.5 Confidence-interval-based metrics for Zoster vaccination (Shingrix) . . . . 122
13.6 Confidence-interval-based metrics for HPV vaccination (Gardasil 9) . . . 126

**14 Negative controls effect size estimates 130**
14.1 Negative controls effect size estimates in Optum EHR . . . . . . . . . . 130
14.2 Negative controls effect size estimates in MDCD . . . . . . . . . . . . . 139
14.3 Negative controls effect size estimates in MDCR . . . . . . . . . . . . . 148
14.4 Negative controls effect size estimates in CCAE . . . . . . . . . . . . . 157

**15 Negative controls effect log likelihood ratios 166**
15.1 Negative controls log likelihood ratios in Optum EHR . . . . . . . . . . . 166
15.2 Negative controls log likelihood ratios in MDCD . . . . . . . . . . . . . . 179
15.3 Negative controls log likelihood ratios in MDCR . . . . . . . . . . . . . . 192
15.4 Negative controls log likelihood ratios in CCAE . . . . . . . . . . . . . . 205

**References 218**

### 1 List of Abbreviations

Table 1: Exposures of interest.

| Exposure Name | Start Date | End Date | History Start Date | History End Date |
|---|---|---|---|---|
| H1N1pdm vaccination | 01-09-2009 | 31-05-2010 | 01-09-2008 | 31-05-2009 |
| Seasonal flu vaccination (Fluvirin) | 01-09-2017 | 31-05-2018 | 01-09-2016 | 31-05-2017 |
| Seasonal flu vaccination (Fluzone) | 01-09-2017 | 31-05-2018 | 01-09-2016 | 31-05-2017 |
| Seasonal flu vaccination (All) | 01-09-2017 | 31-05-2018 | 01-09-2016 | 31-05-2017 |
| Zoster vaccination (Shingrix) | 01-01-2018 | 31-12-2018 | 01-01-2017 | 31-12-2017 |
| HPV vaccination (Gardasil 9) | 01-01-2018 | 31-12-2018 | 01-01-2017 | 31-12-2017 |

### 2 Exposures

The evaluation centers on six existing (groups of) vaccines, for specific time periods (start date to end date), as shown in Table 1.

For some methods the period between historic start and historic end date is used to estimate the historic incidence rate. The formal cohort definitions of each exposure can be found in the next sections.

#### 2.1 H1N1pdm Vaccines

##### 2.1.1 Cohort Entry Events

People enter the cohort when observing any of the following:

1. drug exposures of 'H1N1 vaccine,' starting between September 1, 2009 and May 31, 2010.

Limit cohort entry events to the earliest event per person.

##### 2.1.2 Cohort Exit

The cohort end date will be offset from index event's start date plus 0 days.

##### 2.1.3 Cohort Eras

Entry events will be combined into cohort eras if they are within 0 days of each other.

##### 2.1.4 Concept set: H1N1 vaccine

| Concept ID | Concept Name | Code | Vocabulary | Excluded | Descendants | Mapped |
|---|---|---|---|---|---|---|
| 40213187 | Novel influenza-H1N1-09, all formulations | 128 | CVX | NO | YES | NO |
| 40166607 | influenza A-California-7-2009-(H1N1)v-like virus vaccine 0.03 MG/ML Injectable Suspension | 864704 | RxNorm | NO | YES | NO |
| 40166130 | 0.25 ML influenza A-California-7-2009-(H1N1)v-like virus vaccine 0.03 MG/ML Prefilled Syringe | 864781 | RxNorm | NO | YES | NO |

| | | | | | | |
|---|---|---|---|---|---|---|
| 40166144 | 0.5 ML influenza A-California-7-2009-(H1N1)v-like virus vaccine 0.03 MG/ML Prefilled Syringe | 864797 | RxNorm | NO | YES | NO |
| 42902936 | influenza A-California-7-2009-(H1N1)v-like virus vaccine 0.03 MG/ML Prefilled Syringe | 1360049 | RxNorm | NO | YES | NO |
| 40240135 | influenza A-California-7-2009-(H1N1)v-like virus vaccine 0.09 MG/ML | 1111367 | RxNorm | NO | YES | NO |
| 40225009 | influenza A-California-7-2009-(H1N1)v-like virus vaccine 0.12 MG/ML | 1005949 | RxNorm | NO | YES | NO |
| 40166608 | influenza A-California-7-2009-(H1N1)v-like virus vaccine 158000000 UNT/ML | 864812 | RxNorm | NO | YES | NO |
| 45776785 | influenza A-California-7-2009-(H1N1)v-like virus vaccine 50000000 MG/ML | 1543758 | RxNorm | NO | YES | NO |
| 40166609 | influenza A-California-7-2009-(H1N1)v-like virus vaccine Injectable Suspension | 864703 | RxNorm | NO | YES | NO |
| 40166611 | influenza A-California-7-2009-(H1N1)v-like virus vaccine Prefilled Syringe | 864780 | RxNorm | NO | YES | NO |

---

#### 2.2 Seasonal Flu Vaccines (Fluvirin)

##### 2.2.1 Cohort Entry Events

People enter the cohort when observing any of the following:

1. drug exposures of ‘Fluvirin,’ starting between September 1, 2017 and May 31, 2018.

Limit cohort entry events to the earliest event per person.

##### 2.2.2 Cohort Exit

The cohort end date will be offset from index event’s start date plus 0 days.

##### 2.2.3 Cohort Eras

Entry events will be combined into cohort eras if they are within 0 days of each other.

##### 2.2.4 Concept set: Fluvirin

| Concept ID | Concept Name | Code | Vocabulary | Excluded | Descendants | Mapped |
|---|---|---|---|---|---|---|
| 1593906 | influenza A virus A/Hong Kong/4801/2014 (H3N2) antigen 0.03 MG/ML / influenza A virus A/Singapore/GP1908/2015 (H1N1) antigen 0.03 MG/ML / influenza B virus B/Brisbane/60/2008 antigen 0.03 MG/ML [Fluvirin 2017-2018] | 1928971 | RxNorm | NO | YES | NO |

---

#### 2.3 Seasonal Flu Vaccines (Fluzone)

##### 2.3.1 Cohort Entry Events

People enter the cohort when observing any of the following:

1. drug exposures of ‘Fluzone,’ starting between September 1, 2017 and May 31, 2018.

Limit cohort entry events to the earliest event per person.

##### 2.3.2 Cohort Exit

The cohort end date will be offset from index event’s start date plus 0 days.

##### 2.3.3 Cohort Eras

Entry events will be combined into cohort eras if they are within 0 days of each other.

##### 2.3.4 Concept set: Fluzone

| Concept ID | Concept Name | Code | Vocabulary | Excluded | Descendants | Mapped |
|---|---|---|---|---|---|---|
| 1593354 | influenza A virus A/Hong Kong/4801/2014 (H3N2) antigen 0.12 MG/ML / influenza A virus A/Michigan/45/2015 (H1N1) antigen 0.12 MG/ML / influenza B virus B/Brisbane/60/2008 antigen 0.12 MG/ML [Fluzone 2017-2018] | 1928341 | RxNorm | NO | YES | NO |

---

#### 2.4 Seasonal Flu Vaccines (All)

##### 2.4.1 Cohort Entry Events

People enter the cohort when observing any of the following:

1. drug exposures of ‘Seasonal flu vaccine,’ starting between September 1, 2017 and May 31, 2018.

Limit cohort entry events to the earliest event per person.

##### 2.4.2 Cohort Exit

The cohort end date will be offset from index event’s start date plus 0 days.

##### 2.4.3 Cohort Eras

Entry events will be combined into cohort eras if they are within 0 days of each other.

##### 2.4.4 Concept set: Seasonal flu vaccine

| Concept ID | Concept Name | Code | Vocabulary | Excluded | Descendants | Mapped |
|---|---|---|---|---|---|---|
| 40213145 | influenza, injectable, quadrivalent, contains preservative | 158 | CVX | NO | YES | NO |
| 42903442 | influenza B virus | 1312376 | RxNorm | NO | YES | NO |
| 40213150 | influenza, live, intranasal, quadrivalent | 149 | CVX | NO | YES | NO |
| 40213159 | influenza virus vaccine, whole virus | 16 | CVX | NO | YES | NO |
| 40225028 | influenza virus vaccine, inactivated A-Victoria-210-2009 X-187 (H3N2) (A-Perth-16-2009) strain | 1005931 | RxNorm | NO | YES | NO |
| 40213156 | influenza virus vaccine, split virus (incl. purified surface antigen)-retired CODE | 15 | CVX | NO | YES | NO |
| 40213151 | Seasonal, trivalent, recombinant, injectable influenza vaccine, preservative free | 155 | CVX | NO | YES | NO |
| 40213327 | influenza nasal, unspecified formulation | 151 | CVX | NO | YES | NO |
| 40213148 | influenza, intradermal, quadrivalent, preservative free, injectable | 166 | CVX | NO | YES | NO |
| 40213158 | influenza virus vaccine, unspecified formulation | 88 | CVX | NO | YES | NO |
| 36878713 | Influenza Virus Fragmented, Inactivated, Strain B / Phuket / 3073/2013 | OMOP98957 | RxNorm Extension | NO | YES | NO |
| 42873961 | influenza B virus vaccine, B-Wisconsin-1-2010-like virus | 1303855 | RxNorm | NO | YES | NO |
| 40225038 | influenza virus vaccine, live attenuated, A-Perth-16-2009 (H3N2) strain | 1005911 | RxNorm | NO | YES | NO |
| 40213146 | Influenza, injectable, quadrivalent, preservative free | 150 | CVX | NO | YES | NO |
| 40213143 | Influenza, injectable, Madin Darby Canine Kidney, preservative free, quadrivalent | 171 | CVX | NO | YES | NO |
| 36879025 | Influenza Virus Surface Antigens, strain A / Switzerland / 9715293/2013 H3N2 - Analogue Strain Nib-88 | OMOP991645 | RxNorm Extension | NO | YES | NO |
| 40213157 | Seasonal trivalent influenza vaccine, adjuvanted, preservative free | 168 | CVX | NO | YES | NO |
| 45776076 | influenza A virus vaccine, A-Texas-50-2012 (H3N2)-like virus | 1541617 | RxNorm | NO | YES | NO |
| 40213149 | influenza virus vaccine, live, attenuated, for intranasal use | 111 | CVX | NO | YES | NO |
| 40213147 | Influenza, injectable,quadrivalent, preservative free, pediatric | 161 | CVX | NO | YES | NO |
| 40213152 | Seasonal, quadrivalent, recombinant, injectable influenza vaccine, preservative free | 185 | CVX | NO | YES | NO |
| 42903441 | influenza A virus | 1312375 | RxNorm | NO | YES | NO |
| 40213141 | influenza, high dose seasonal, preservative-free | 135 | CVX | NO | YES | NO |
| 40213153 | Influenza, seasonal, injectable | 141 | CVX | NO | YES | NO |
| 40213144 | Influenza, injectable, Madin Darby Canine Kidney, quadrivalent with preservative | 186 | CVX | NO | YES | NO |
| 40213142 | Influenza, injectable, Madin Darby Canine Kidney, preservative free | 153 | CVX | NO | YES | NO |
| 40213155 | seasonal influenza, intradermal, preservative free | 144 | CVX | NO | YES | NO |
| 40164828 | influenza B virus vaccine B/Brisbane/60/2008 antigen | 857921 | RxNorm | NO | YES | NO |

#### 2.5 HPV Vaccines

##### 2.5.1 Cohort Entry Events

People enter the cohort when observing any of the following:

1. drug exposures of ‘Gardasil 9,’ starting between January 1, 2018 and December 31, 2018.

##### 2.5.2 Cohort Exit

The cohort end date will be offset from index event’s start date plus 0 days.

##### 2.5.3 Cohort Eras

Entry events will be combined into cohort eras if they are within 0 days of each other.

##### 2.5.4 Concept set: Gardasil 9

| Concept ID | Concept Name | Code | Vocabulary | Excluded | Descendants | Mapped |
|---|---|---|---|---|---|---|
| 36248866 | Gardasil 9 Injectable Product | 1597098 | RxNorm | NO | YES | NO |
| 45892513 | L1 protein, human papillomavirus type 11 vaccine / L1 protein, human papillomavirus type 16 vaccine / L1 protein, human papillomavirus type 18 vaccine / L1 protein, human papillomavirus type 31 vaccine / L1 protein, human papillomavirus type 33 vaccine / | 1597102 | RxNorm | NO | YES | NO |
| 45892514 | 0.5 ML L1 protein, human papillomavirus type 11 vaccine 0.08 MG/ML / L1 protein, human papillomavirus type 16 vaccine 0.12 MG/ML / L1 protein, human papillomavirus type 18 vaccine 0.08 MG/ML / L1 protein, human papillomavirus type 31 vaccine 0.04 MG/ML / | 1597103 | RxNorm | NO | YES | NO |
| 45892510 | 0.5 ML L1 protein, human papillomavirus type 11 vaccine 0.08 MG/ML / L1 protein, human papillomavirus type 16 vaccine 0.12 MG/ML / L1 protein, human papillomavirus type 18 vaccine 0.08 MG/ML / L1 protein, human papillomavirus type 31 vaccine 0.04 MG/ML / | 1597099 | RxNorm | NO | YES | NO |
| 40213322 | Human Papillomavirus 9-valent vaccine | 165 | CVX | NO | YES | NO |

#### 2.6 Zoster Vaccines

##### 2.6.1 Cohort Entry Events

People enter the cohort when observing any of the following:

1. drug exposures of ‘Shingrix,’ starting between January 1, 2018 and December 31, 2018.

##### 2.6.2 Cohort Exit

The cohort end date will be offset from index event’s start date plus 0 days.

##### 2.6.3 Cohort Eras

Entry events will be combined into cohort eras if they are within 0 days of each other.

##### 2.6.4 Concept set: Shingrix

| Concept ID | Concept Name | Code | Vocabulary | Excluded | Descendants | Mapped |
|---|---|---|---|---|---|---|
| 792784 | varicella zoster virus glycoprotein E Injection [Shingrix] | 1986828 | RxNorm | NO | YES | NO |
| 792783 | varicella zoster virus glycoprotein E, recombinant 0.1 MG/ML [Shingrix] | 1986827 | RxNorm | NO | YES | NO |
| 792788 | varicella zoster virus glycoprotein E, recombinant 0.1 MG/ML Injection [Shingrix] | 1986832 | RxNorm | NO | YES | NO |
| 36421491 | Varicella-Zoster Virus Vaccine Live (Oka-Merck) strain Injectable Solution [Shingrix] | OMOP4763774 | RxNorm Extension | NO | YES | NO |
| 792785 | Shingrix Injectable Product | 1986829 | RxNorm | NO | YES | NO |
| 706103 | zoster vaccine recombinant | 187 | CVX | NO | YES | NO |

### 3 Negative control outcomes

Negative controls are outcomes believed not to be caused by any of the vaccines, and therefore ideally would not be flagged as a signal by a safety surveillance system. Any effect size estimates for negative control ideally should be close to the null.

A single set of negative control outcomes is defined for all four vaccine groups. To identify negative control outcomes that match the severity and prevalence of suspected vaccine adverse effects, a candidate list of negative controls was generated based on similarity of prevalence and percent of diagnoses that were recorded in an inpatient setting (as a proxy for severity). Manual review of this list by clinical experts created the final list of 93 negative control outcomes. The full list of negative control outcomes can be found in Table 8.

Table 8: Negative control outcomes.

| Outcome Id | Outcome Name |
|---:|---|
| 438945 | Accidental poisoning by benzodiazepine-based tranquilizer |
| 434455 | Acquired claw toes |
| 316211 | Acquired spondylolisthesis |
| 201612 | Alcoholic liver damage |
| 438730 | Alkalosis |
| 441258 | Anemia in neoplastic disease |
| 432513 | Animal bite wound |
| 4171556 | Ankle ulcer |
| 4098292 | Antiphospholipid syndrome |
| 77650 | Aseptic necrosis of bone |
| 4239873 | Benign neoplasm of ciliary body |
| 23731 | Benign neoplasm of larynx |
| 199764 | Benign neoplasm of ovary |
| 195500 | Benign neoplasm of uterus |
| 4145627 | Biliary calculus |
| 4108471 | Burn of digit of hand |
| 75121 | Burn of lower leg |
| 4284982 | Calculus of bile duct without obstruction |
| 434327 | Cannabis abuse |
| 78497 | Cellulitis and abscess of toe |
| 4001454 | Cervical spine ankylosis |
| 4068241 | Chronic instability of knee |
| 195596 | Chronic pancreatitis |
| 4206338 | Chronic salpingitis |
| 4058397 | Claustrophobia |
| 74816 | Contusion of toe |
| 73302 | Curvature of spine |
| 4151134 | Cyst of pancreas |
| 77638 | Displacement of intervertebral disc without myelopathy |
| 195864 | Diverticulum of bladder |
| 201346 | Edema of penis |
| 200461 | Endometriosis of uterus |
| 377877 | Esotropia |
| 193530 | Follicular cyst of ovary |
| 4094822 | Foreign body in respiratory tract |
| 443421 | Gallbladder and bile duct calculi |
| 4299408 | Gouty tophus |
| 135215 | Hashimoto thyroiditis |
| 442190 | Hemorrhage of colon |

| | |
|---|---|
| 43020475 | High risk heterosexual behavior |
| 194149 | Hirschsprung's disease |
| 443204 | Human ehrlichiosis |
| 4226238 | Hyperosmolar coma due to diabetes mellitus |
| 4032787 | Hyperosmolarity |
| 197032 | Hyperplasia of prostate |
| 140362 | Hypoparathyroidism |
| 435371 | Hypothermia |
| 138690 | Infestation by Pediculus |
| 4152376 | Intentional self poisoning |
| 192953 | Intestinal adhesions with obstruction |
| 196347 | Intestinal parasitism |
| 137977 | Jaundice |
| 317510 | Leukemia |
| 765053 | Lump in right breast |
| 378165 | Nystagmus |
| 434085 | Obstruction of duodenum |
| 4147016 | Open wound of buttock |
| 4129404 | Open wound of upper arm |
| 438120 | Opioid dependence |
| 75924 | Osteodystrophy |
| 432594 | Osteomalacia |
| 30365 | Panhypopituitarism |
| 4108371 | Peripheral gangrene |
| 440367 | Plasmacytosis |
| 439233 | Poisoning by antidiabetic agent |
| 442149 | Poisoning by bee sting |
| 4314086 | Poisoning due to sting of ant |
| 4147660 | Postural kyphosis |
| 434319 | Premature ejaculation |
| 199754 | Primary malignant neoplasm of pancreas |
| 4311499 | Primary malignant neoplasm of respiratory tract |
| 436635 | Primary malignant neoplasm of sigmoid colon |
| 196044 | Primary malignant neoplasm of stomach |
| 433716 | Primary malignant neoplasm of testis |
| 133424 | Primary malignant neoplasm of thyroid gland |
| 194997 | Prostatitis |
| 80286 | Prosthetic joint loosening |
| 443274 | Psychostimulant dependence |
| 314962 | Raynaud's disease |
| 37018294 | Residual osteitis |
| 4288241 | Salmonella enterica subspecies arizonae infection |
| 45757269 | Sclerosing mesenteritis |
| 74722 | Secondary localized osteoarthrosis of pelvic region |
| 200348 | Secondary malignant neoplasm of large intestine |
| 43020446 | Sedative withdrawal |
| 74194 | Sprain of spinal ligament |
| 4194207 | Tailor's bunion |
| 193521 | Tropical sprue |
| 40482801 | Type II diabetes mellitus uncontrolled |
| 74719 | Ulcer of foot |
| 196625 | Viral hepatitis A without hepatic coma |
| 197494 | Viral hepatitis C |
| 4284533 | Vitamin D-dependent rickets |

Negative control outcomes are defined as the first occurrence of the negative control concept or any of its descendants.

Table 9: Committed EUMAEUS data sources and the populations they cover.

| Data source | Population | Data capture process and short description |
|---|---|---|
| **Administrative claims** | | |
| IBM MarketScan Commercial Claims and Encounters (CCAE) | Commercially insured, younger than 65 years | Adjudicated health insurance claims (e.g. inpatient, outpatient, and outpatient pharmacy) from large employers and health plans who provide private healthcare coverage to employees, their spouses and dependents. |
| IBM MarketScan Medicare Supplemental Database (MDCR) | Commercially insured, 65 years and older | Adjudicated health insurance claims of retirees with primary or Medicare supplemental coverage through privately insured fee-for-service, point-of-service or capitated health plans. |
| IBM MarketScan Multi-State Medicaid Database (MDCD) | Medicaid enrollees, racially diverse | Adjudicated health insurance claims for Medicaid enrollees from multiple states and includes hospital discharge diagnoses, outpatient diagnoses and procedures, and outpatient pharmacy claims. |
| **Electronic health records (EHRs)** | | |
| Optum Electronic Health Records (OptumEHR) | US, general | Clinical information, prescriptions, lab results, vital signs, body measurements, diagnoses and procedures derived from clinical notes using natural language processing. |

### 4 Data sources

Table 9 lists the five data sources included in EUMAEUS; these sources encompass a large variety of practice types and populations. For each data source, we report a brief description and size of the population it represents. All data sources have received institutional review board approval or exemption for their participation before executing EUMAEUS.

#### 4.1 Database characteristics

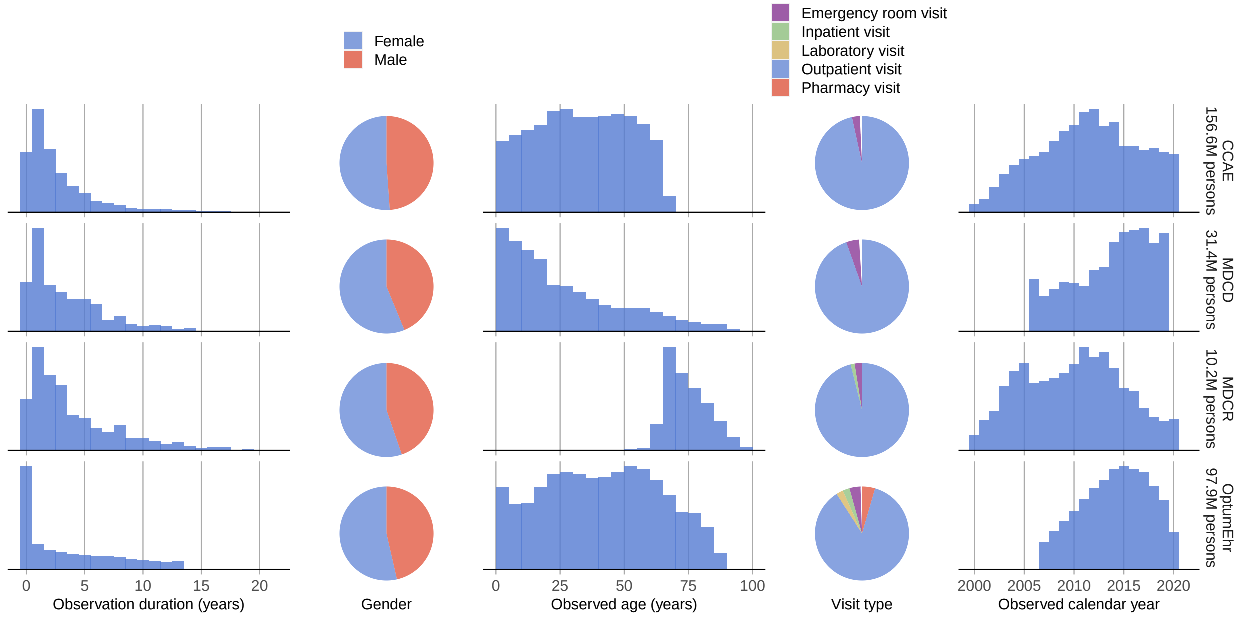

Figure 1: Overall distributions of key characteristics in each database.

### 5 Evaluated Methods

Vaccine safety surveillance methods can be broken down in to four components: construction of a *counterfactual* (often referred to as the 'expected count'), a *time-at-risk*, the *statistic* to estimate, and potentially a *decision rule* on the estimate to classify signals from non-signals.

#### 5.1 Counterfactual construction

##### Historic rates

Traditionally, vaccine surveillance methods compute an expected count based an incidence rate estimated during some historic time period, for example in the years prior to the initiation of the surveillance study. We use the historic period indicated in Table 1. We evaluate four variations:

- Unadjusted, entire year. Using a single rate computed across the entire historic year for the entire population.
- Age and sex adjusted, entire year. Using a rate stratifying by age (in 5 year increments) and sex, computed across the entire historic year. This allows the expected rate to be adjusted for the demographics of the vaccinated.
- Unadjusted, time-at-risk relative to outpatient visit. Using a single rate computed during the time-at-risk relative to a random outpatient visit in the historic year.
- Age and sex adjusted, time-at-risk relative to outpatient visit. Using a rate stratifying by age and sex, computed during the time-at-risk relative to a random outpatient visit in the historic year.

Initial results showed that this counterfactual approach is sensitive to changes in coding practices. We therefore introduce a study diagnostic: the percent change in overall incidence rate (across the entire population) between the historic and current time period. For each of the four variations listed above, we add a new variation where effect-size estimates are removed if the change in incidence rate is greater than 50%.

##### Cohort method using a contemporary non-user comparator

A comparator cohort study most closely emulates a randomized clinical trial, comparing the target cohort (those vaccinated) to some comparator cohort. We define two types of non-user comparator cohort, one having an outpatient visit on the index date, and another having a random date as the index date. For both comparator variants we exclude subjects having a vaccinations for the same disease as the target vaccine on or before the index date. When doing unadjusted comparisons, the comparator cohort is a random sample of equal size as the target cohort. When doing propensity score (PS) adjusted comparisons, the comparator cohort is a stratified (by age and sex) random sample of four times the size of the target cohort (two times for the Seasonal Flu Vaccination (all) target cohort for computational reasons). Propensity models use a large generic set of covariates, including demographics and covariates per drug, condition, procedure, measurement, etc., and are fitted using large-scale regularized regression as described previously. [2] We evaluate 10 method variations:

Anchoring the comparator on a random outpatient visit:

- Unadjusted comparison.
- 1-on-1 PS matching.
- PS stratification. Five equally-sized strata are defined in the target (vaccinated) population.
- Inverse Probability of Treatment Weighting (IPTW). We use stabilized weights to compute the average treatment effect in the treated (ATT). Weights are truncated to a maximum value of 10, similar to Izurieta et al. (2020). [7]
- 1-on-1 PS matching within each period, using only the 'new' data in that period to fit the propensity model. Once a population is matched in a period, that matching is carried forward to subsequent periods. This method is only evaluated using the H1N1pdm vaccinations for computational reasons.

Anchoring the comparator on a random date:

- Unadjusted comparison.
- 1-on-1 PS matching.
- PS stratification.
- IPTW with trimming.
- 1-on-1 PS matching within each period. This method is only be evaluated using the H1N1pdm vaccinations for computational reasons.

**Self-Controlled Case Series (SCCS) / Self-Controlled Risk Interval (SCRI)**

The SCCS and SCRI designs are self-controlled, comparing the time-at-risk (the time shortly following the vaccination) to some other time in the same patient's record. The SCCS design uses all patient time when not at risk as the control time. [6] The SCRI design uses a pre-specified control interval relative to the vaccination date as the control time. [5] This unexposed time can be both before or after the time at risk. We evaluate five variations:

- A simple SCCS, using all patient time when not at risk as the control time, with the exception of the 30 days prior to vaccination which is excluded from the analysis to avoid bias due to contra-indications.
- An SCCS adjusting for age and season. Age and season are modeled to be constant within each calendar month, and vary across months as bicubic splines.
- A simple SCCS discarding all time prior to vaccination.
- An SCRI, using a control interval of 43 to 15 days prior to vaccination.
- An SCRI, using a control interval of 43 to 71 days after to vaccination.

**Case-control**

The case-control design compares cases (those with the outcome) to controls (those that do not have the outcome), and looks back in time for exposures to a vaccine. We evaluate two variants:

- Using up to four age and sex matched controls per case. For age we use a two-year caliper.
- By sampling controls from the general non-case population, and adjusting for age and sex in the outcome model. The control sample is be four times the number of controls. Age is modeled as one variable per 5-year age category.

#### 5.2 Time-at-risk

The time-at-risk is the time window, relative to the vaccination date, when outcomes will potentially be attributed to the vaccine. We define the time-at-risk window top be 1-28 days after vaccination. Time-at-risk windows are constructed both for the first and second dose. The time-at-risk for one dose is censored at the time of the next dose.

#### 5.3 Statistic

- Effect-size estimate. Each method can be used to produce an effect-size estimates such as a hazard ratio, incidence rate ratio, or odds ratio. For example, when using a historic rate we can compute the observed to expected ratio, which can be interpreted as the incidence rate ratio.

- Log likelihood ratio (LLR). A common practice in vaccine safety surveillance is to computer the LLR, which is the log of the ratio between the likelihood of the alternative hypothesis (that there is an effect) and the likelihood of the null hypothesis (of no effect). The LLR is a convenient statistic when performing sequential testing, where the LLR can be compared to a pre-computed critical value, as is done in the MaxSPRT method. [4] Although typically MaxSPRT uses a historic rate as counterfactual, any counterfactual can be used to compute the LLR and can be used in MaxSPRT.

Effect-size estimates are computed both with and without empirical calibration. [1,3] Empirical calibration is done using leave-one-out: when calibrating the estimate for a control, the systematic error distribution is fitted uses all controls except the one being calibrated.

#### 5.4 Decision rule

To identify ‘signals’ we need a decision rule, for example in the shape of a threshold value on one of the estimates statistics. In our experiment we consider one decision rule, which is the critical value computed for the LLR at an alpha of 0.05. For the historical rates method we use a Poisson model assuming the counterfactual is known without uncertainty.

For all other methods we use a binomial model. All critical values are computed using the `Sequential` package in CRAN.

### 6 Fitted systematic error distribution

For each method variation and vaccine group, the systematic error distribution fitted on the negative control estimates are shown. The red area indicates the maximum likelihood estimates of the distribution parameters, the pink area indicates the 95% credible interval.

#### 6.1 Systematic error distributions in the Optum EHR database

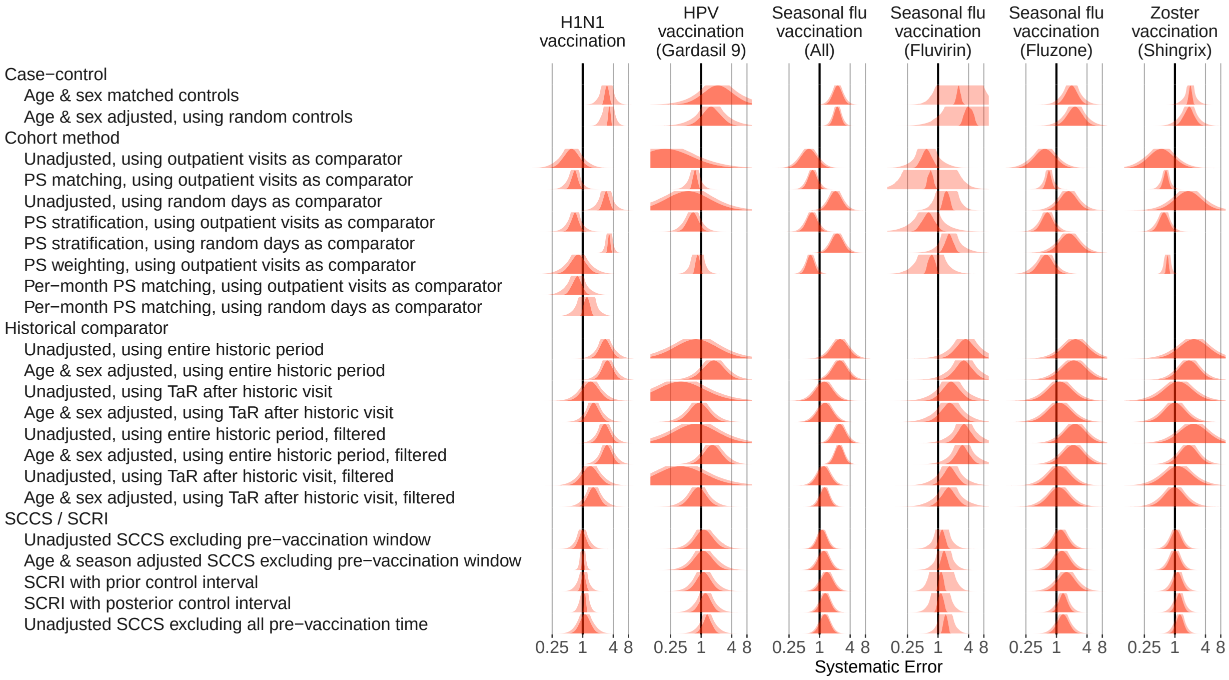

Figure 2: Fitted systematic error distributions for the Optum EHR database.

#### 6.2 Systematic error distributions in the MDCD database

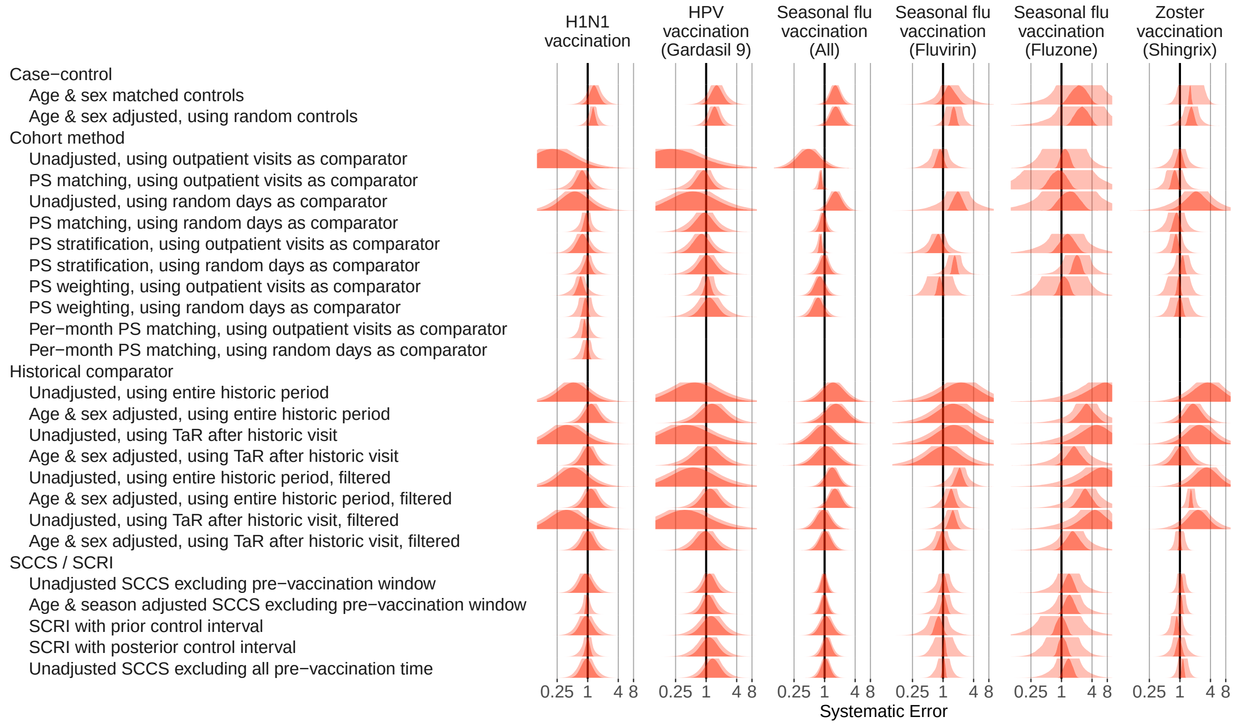

Figure 3: Fitted systematic error distributions for the MDCD database.

#### 6.3 Systematic error distributions in the MDCR database

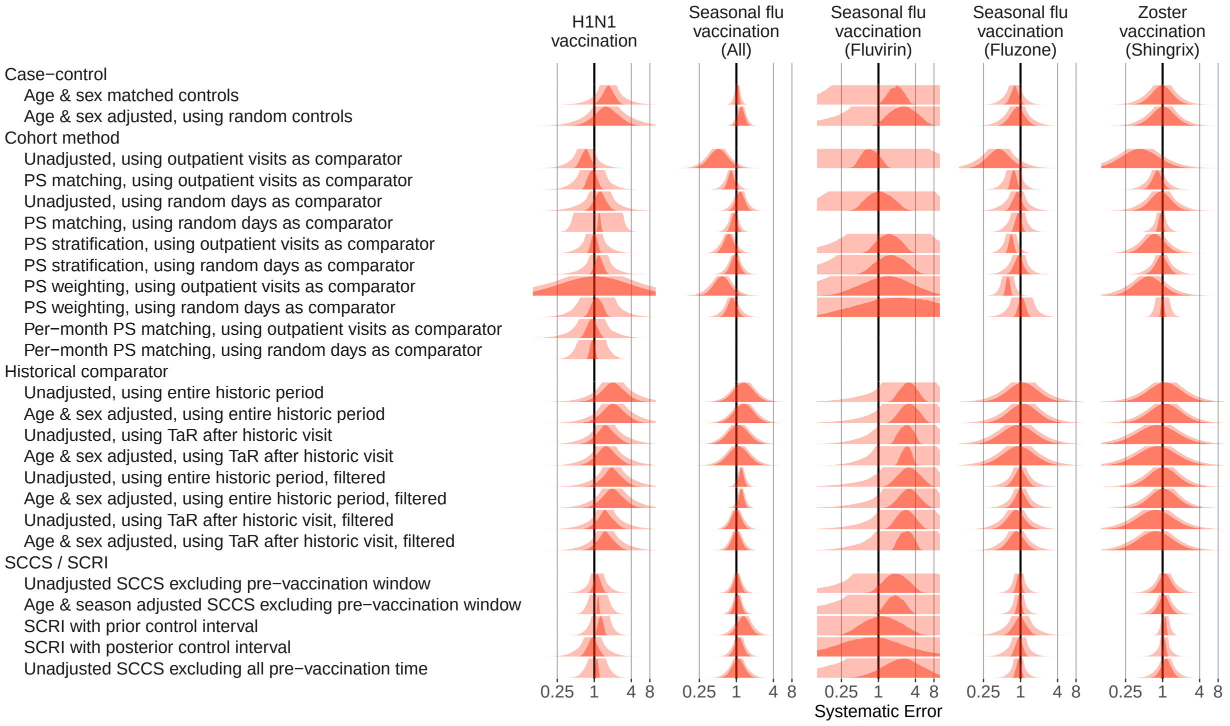

Figure 4: Fitted systematic error distributions for the MDCR database.

#### 6.4 Systematic error distributions in the CCAE database

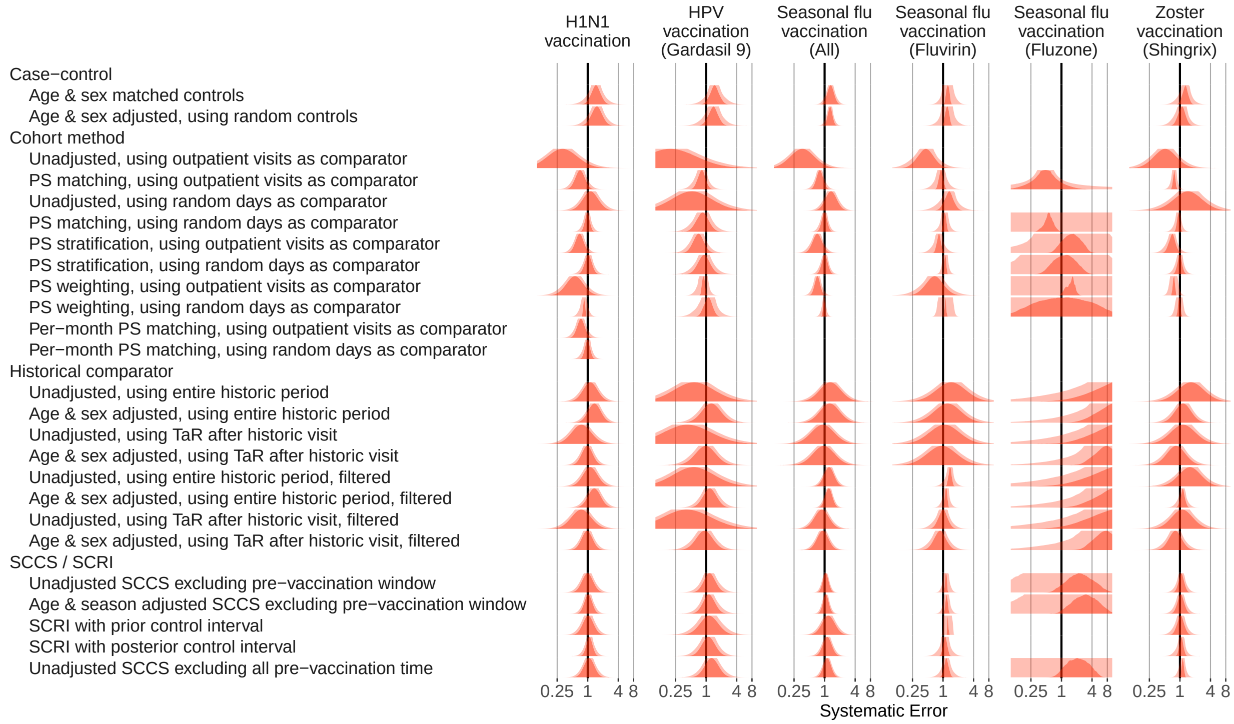

Figure 5: Fitted systematic error distributions for the CCAE database.

#### 7 Type 1 and 2 error before and after calibration

For each method variation and vaccine group, the type 1 and 2 error before and after empirical calibration are shown. The x-axis indicates the type 1 error (higher values to the left) and type 2 error (higher values to the right), based on the (calibrated) one-sided p-value. The dashed line indicates nominal type 1 error of 5%.

##### 7.1 Type 1 and 2 error before and after calibration in the Optum EHR database

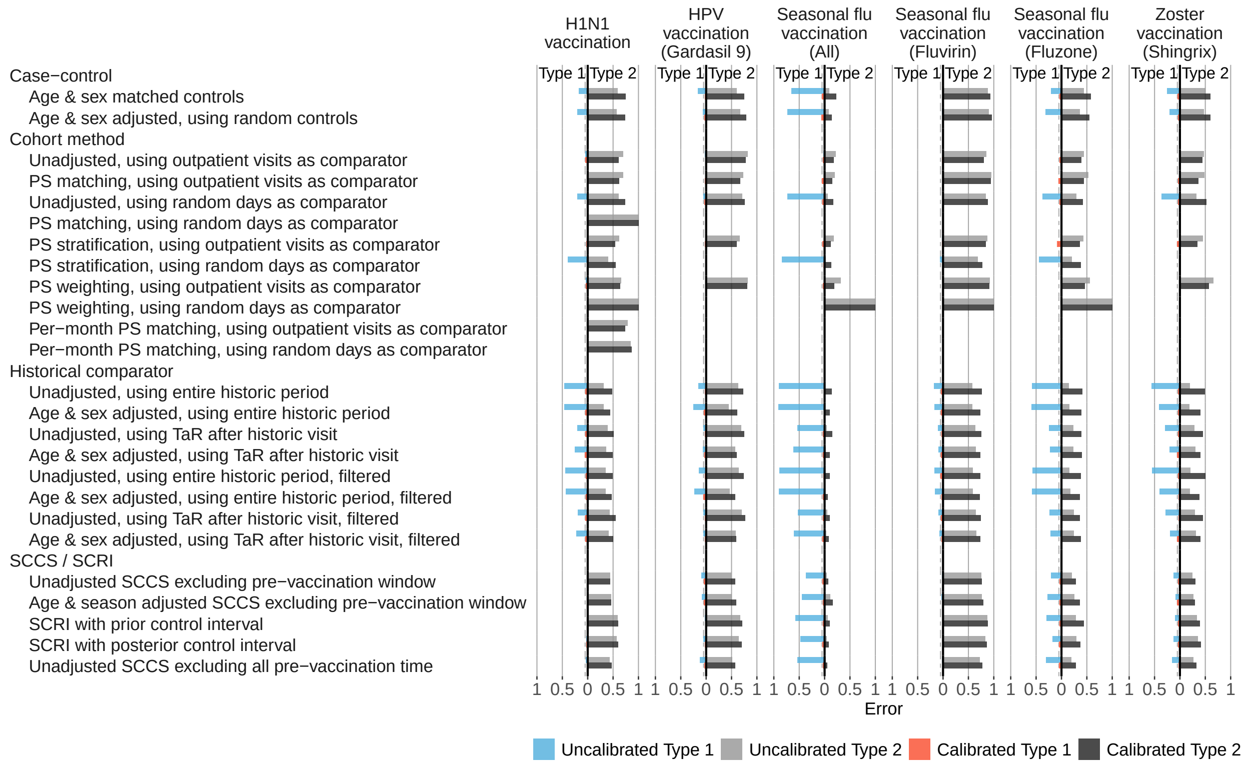

Figure 6: Type 1 and 2 error before and after empirical calibration in the Optum EHR database.

#### 7.2 Type 1 and 2 error before and after calibration in the MDCD database

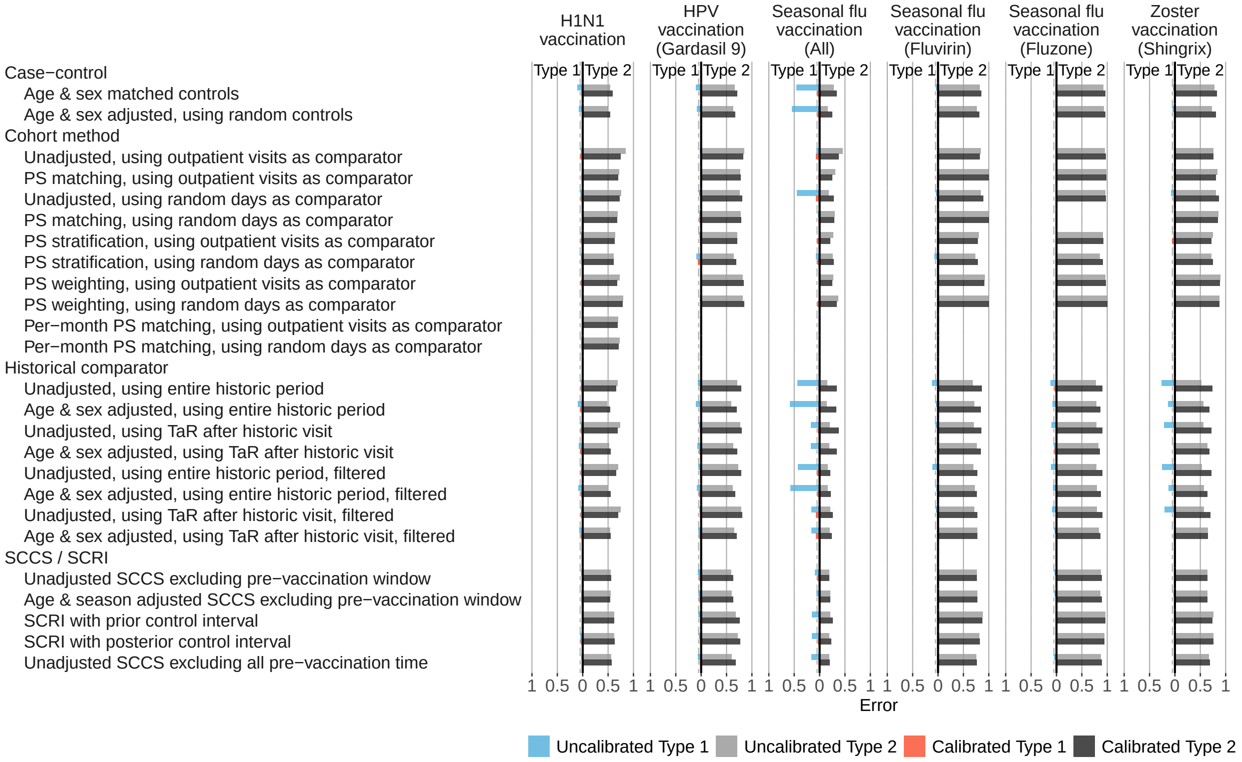

Figure 7: Type 1 and 2 error before and after empirical calibration in the MDCD database.

#### 7.3 Type 1 and 2 error before and after calibration in the MDCR database

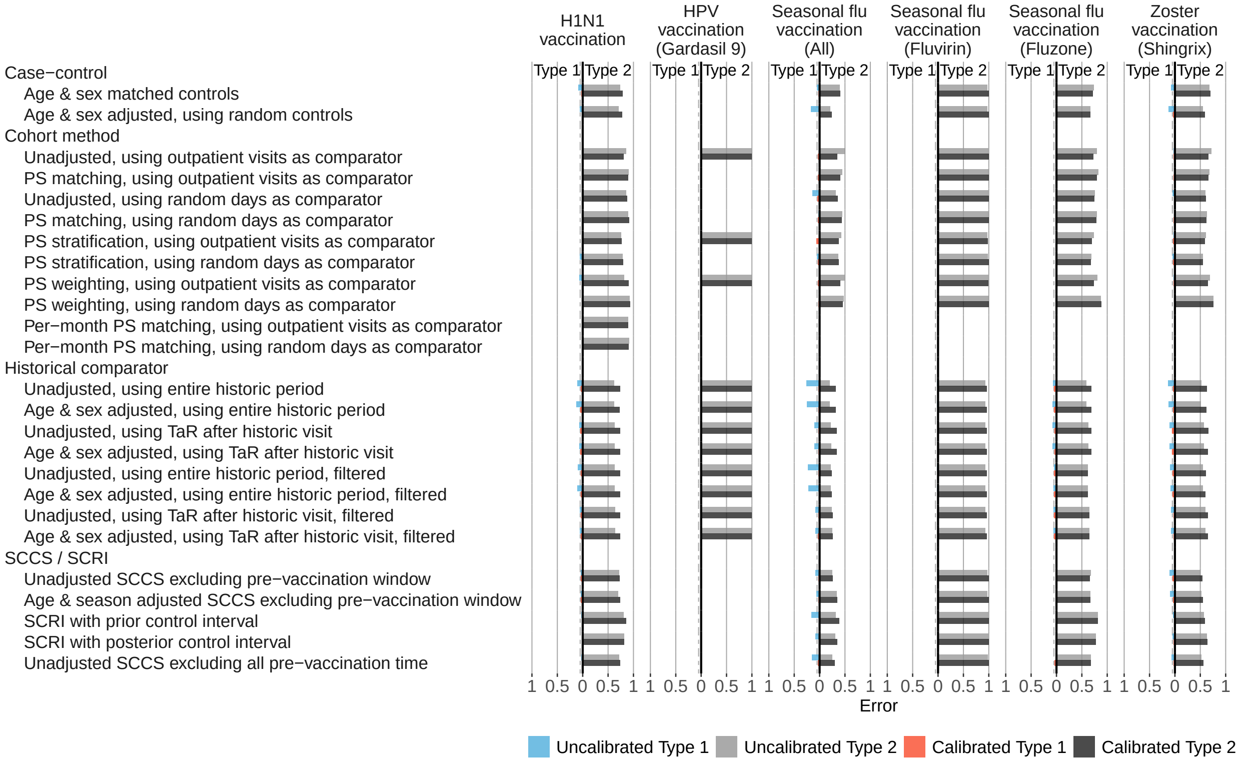

Figure 8: Type 1 and 2 error before and after empirical calibration in the MDCR database.

#### 7.4 Type 1 and 2 error before and after calibration in the CCAE database

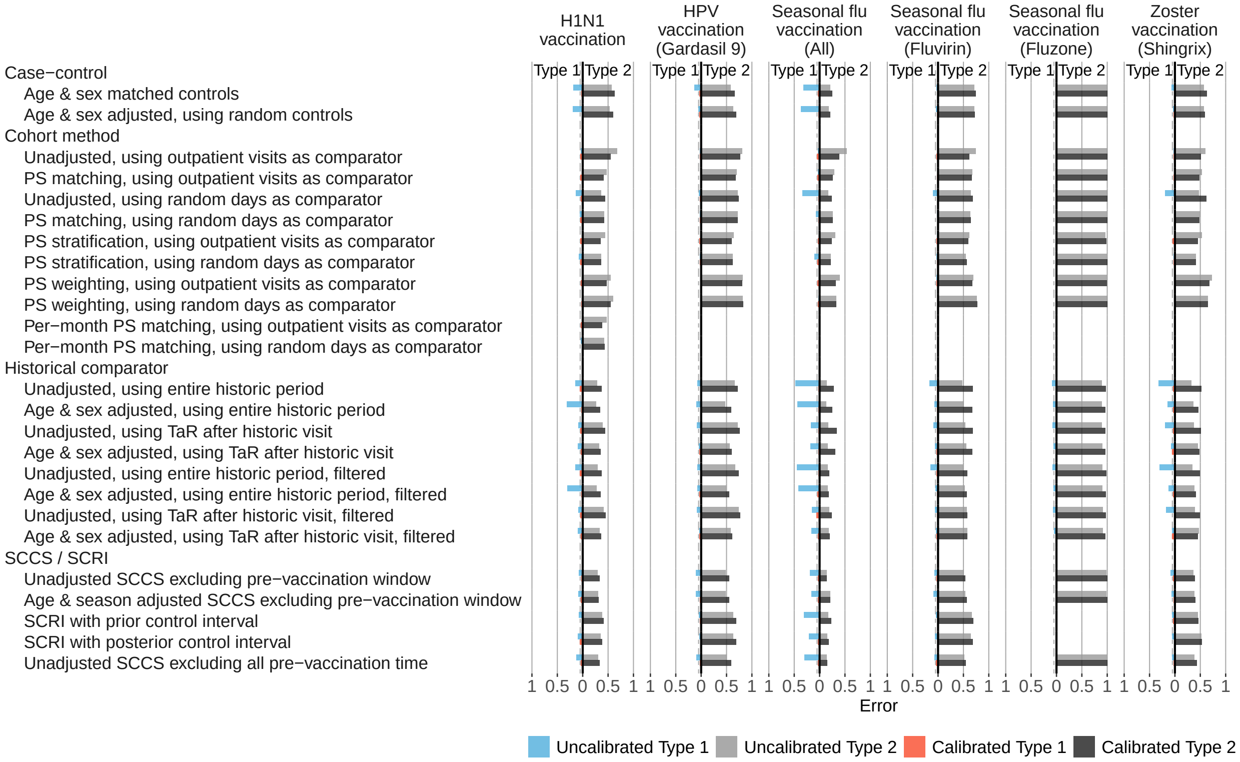

Figure 9: Type 1 and 2 error before and after empirical calibration in the CCAE database.

### 8 Time to 50% sensitivity

For each method variation and vaccine group, the number of months of data needed to achieve 50% sensitivity based on the calibrated MaxSPRT are shown, stratified by true effect size of the positive controls.

#### 8.1 Time to 50% sensitivity in the Optum EHR database

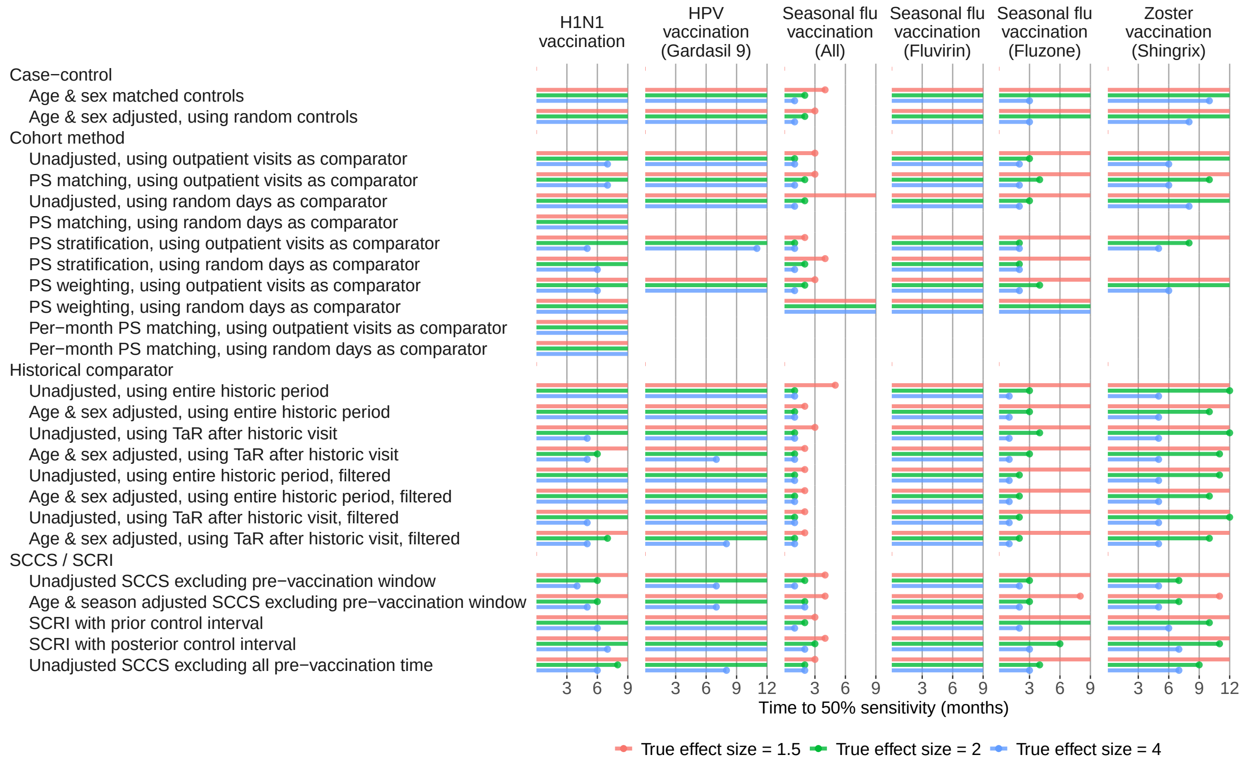

Figure 10: Time to 50% sensitivity in the Optum EHR database.

#### 8.2 Time to 50% sensitivity in the MDCD database

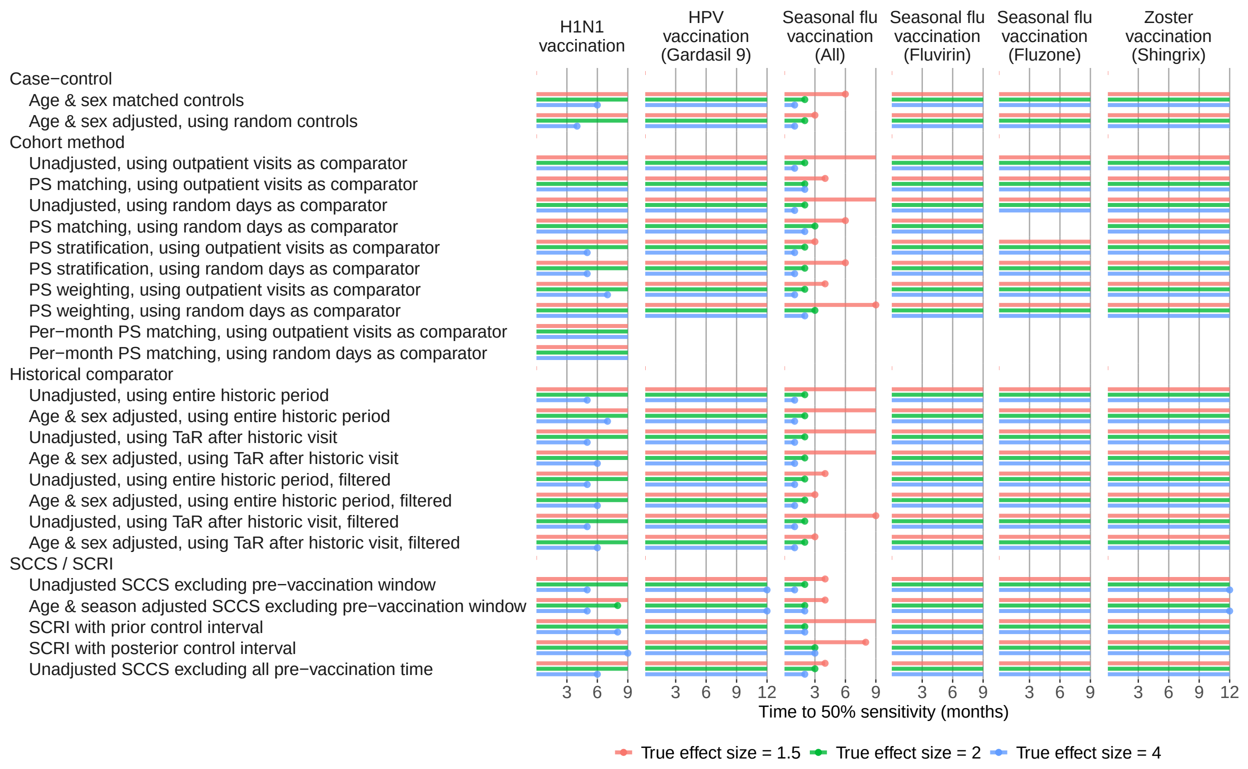

Figure 11: Time to 50% sensitivity in the MDCD database.

#### 8.3 Time to 50% sensitivity in the MDCR database

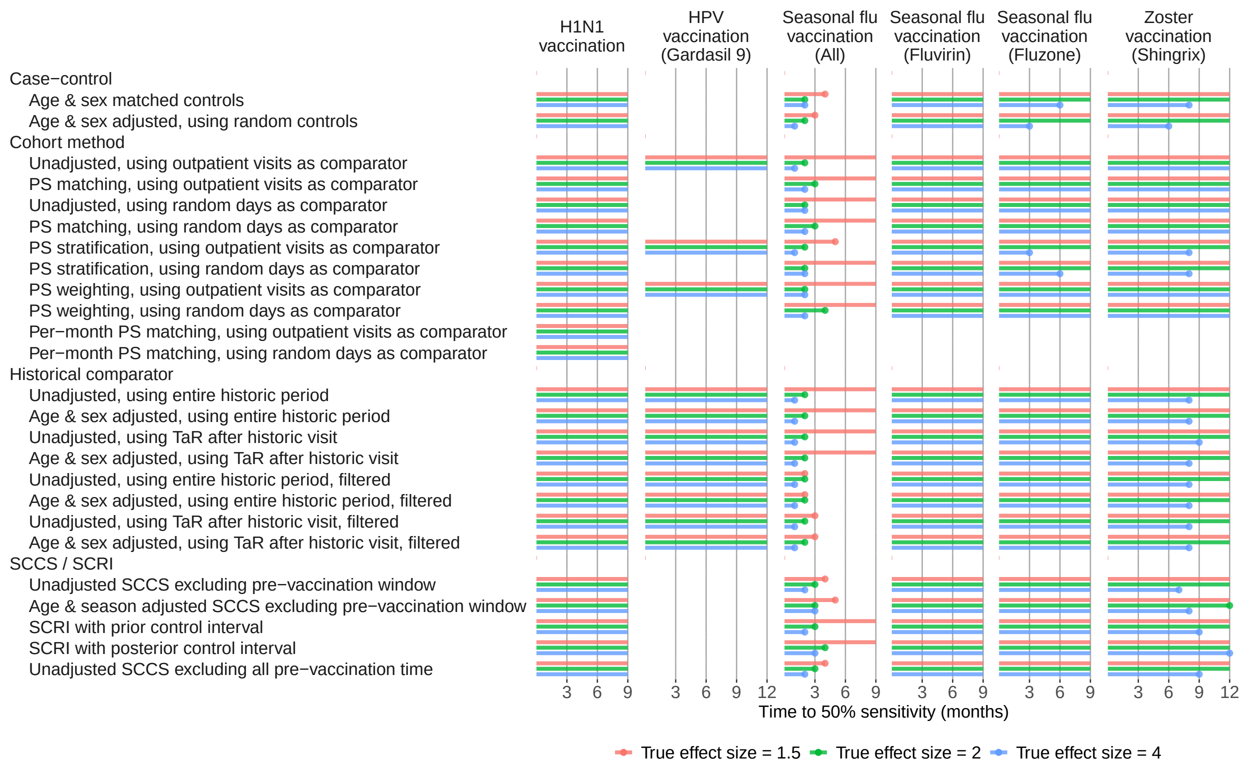

Figure 12: Time to 50% sensitivity in the MDCR database.

#### 8.4 Time to 50% sensitivity in the CCAE database

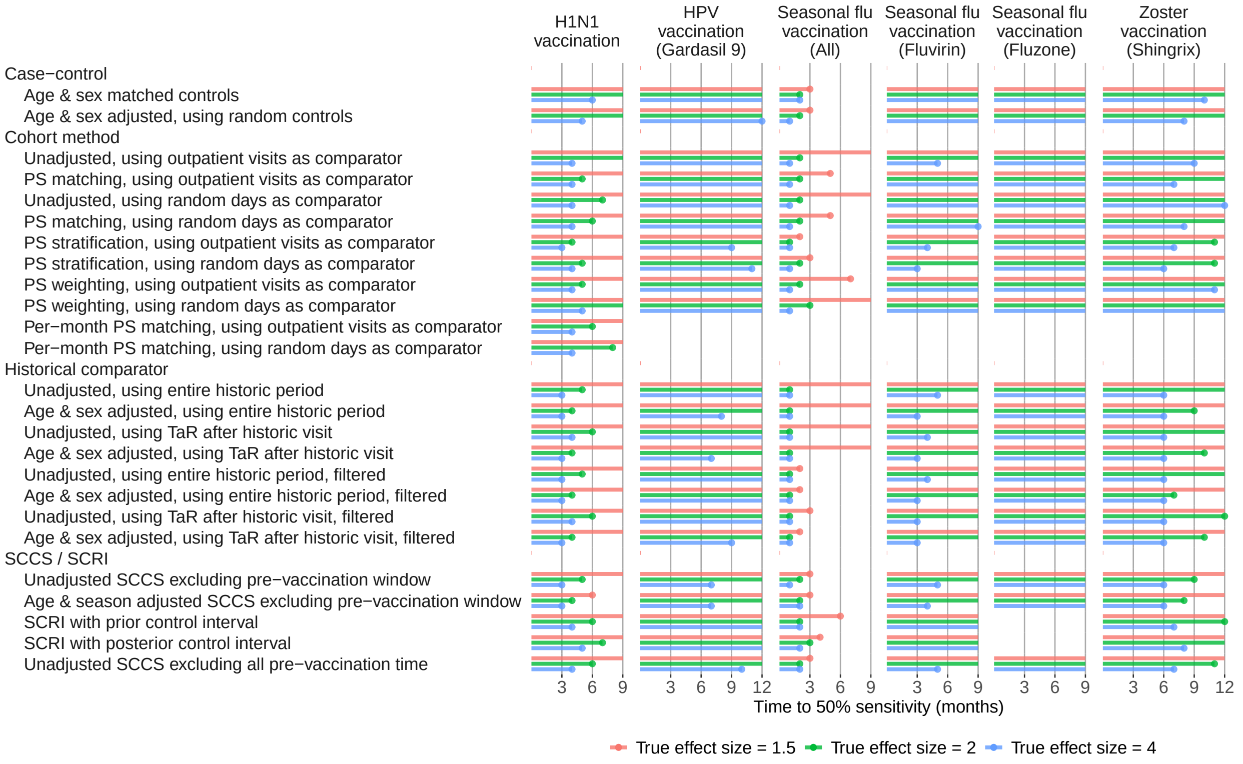

Figure 13: Time to 50% sensitivity in the CCAE database.

### 9 Time to 80% sensitivity

For each method variation and vaccine group, the number of months of data needed to achieve 80% sensitivity based on the calibrated MaxSPRT are shown, stratified by true effect size of the positive controls.

#### 9.1 Time to 80% sensitivity in the Optum EHR database

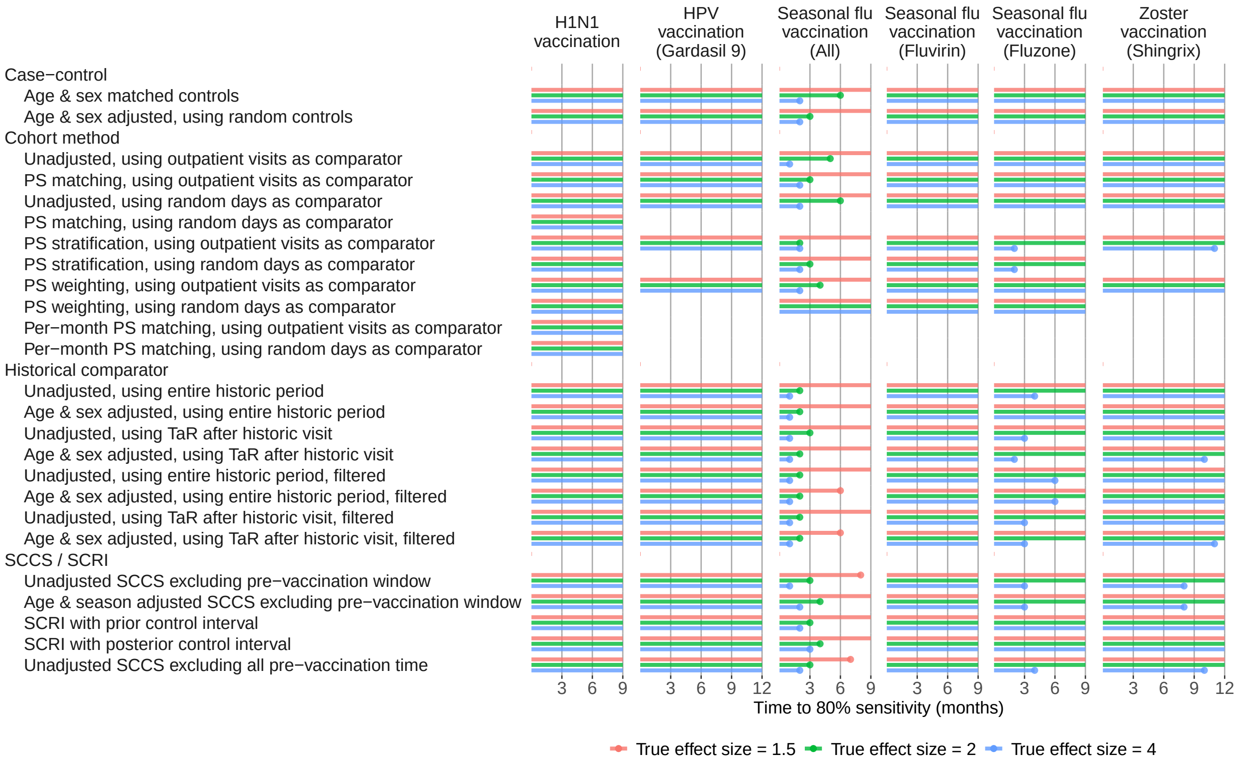

Figure 14: Time to 80% sensitivity in the Optum EHR database.

#### 9.2 Time to 80% sensitivity in the MDCD database

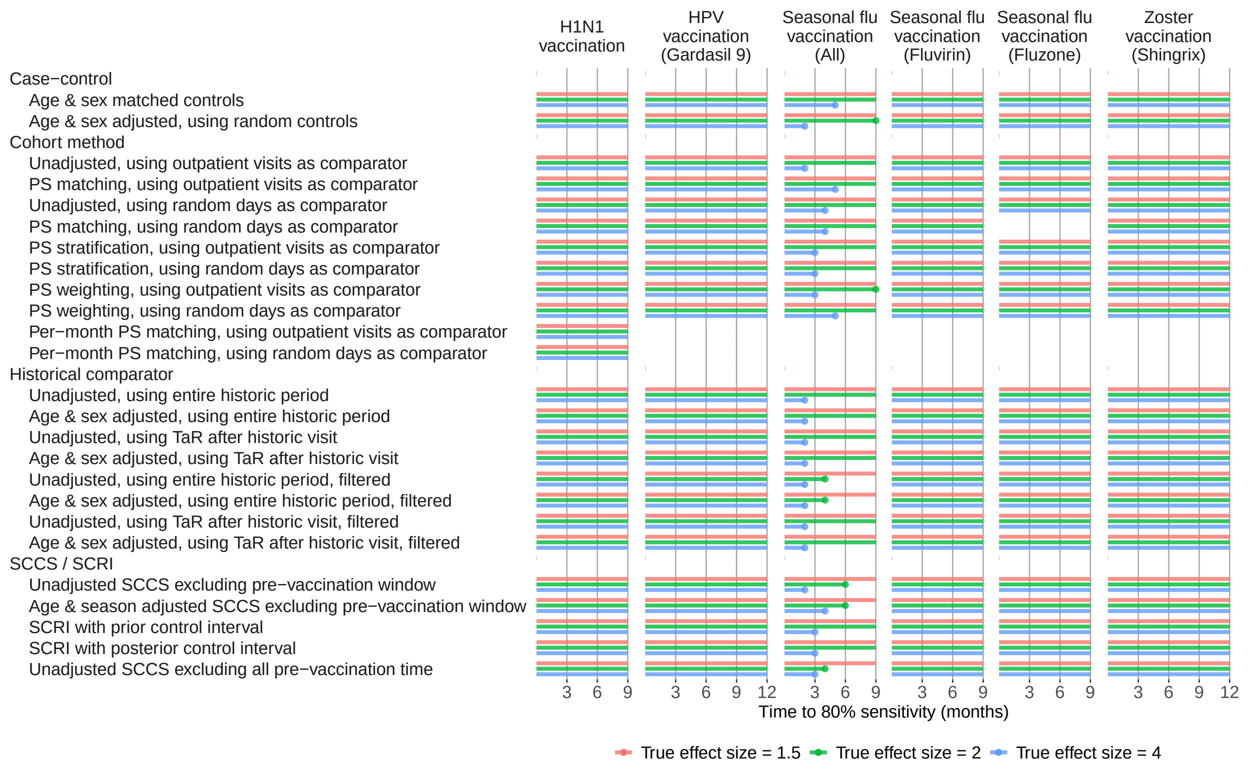

Figure 15: Time to 80% sensitivity in the MDCD database.

#### 9.3 Time to 80% sensitivity in the MDCR database

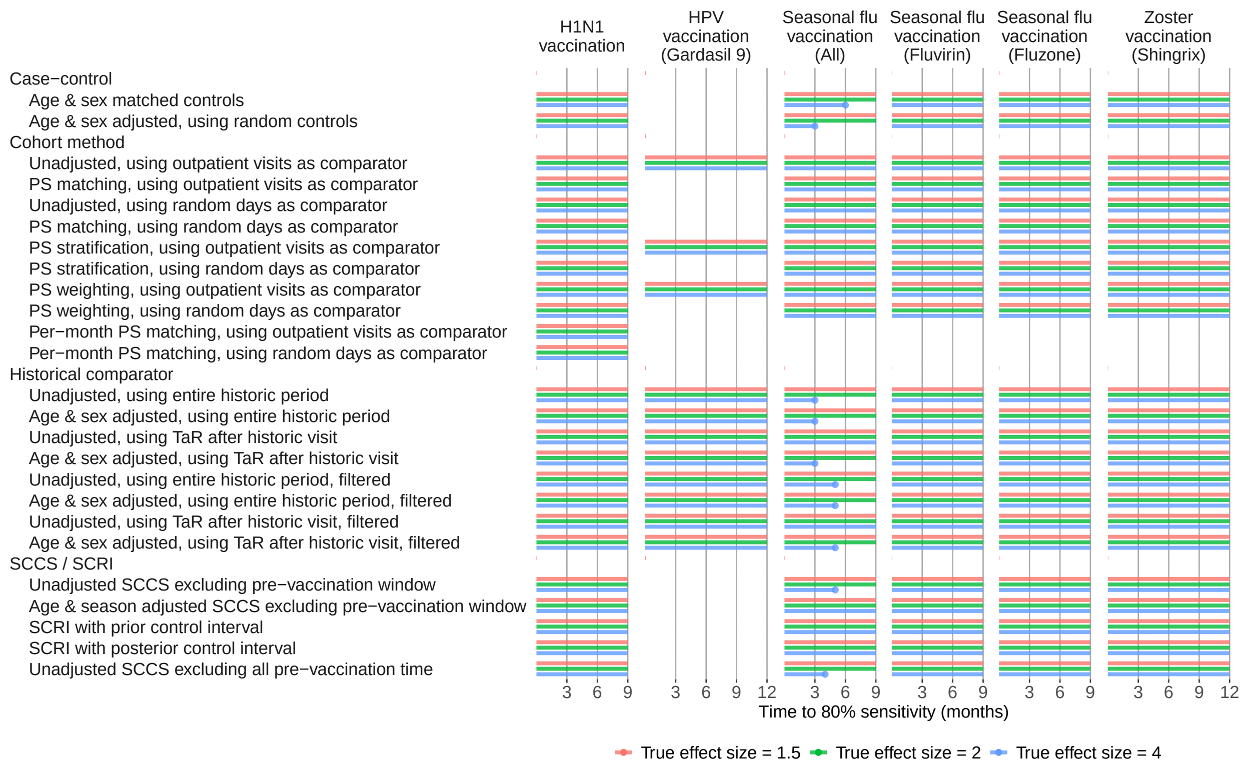

Figure 16: Time to 80% sensitivity in the MDCR database.

#### 9.4 Time to 80% sensitivity in the CCAE database

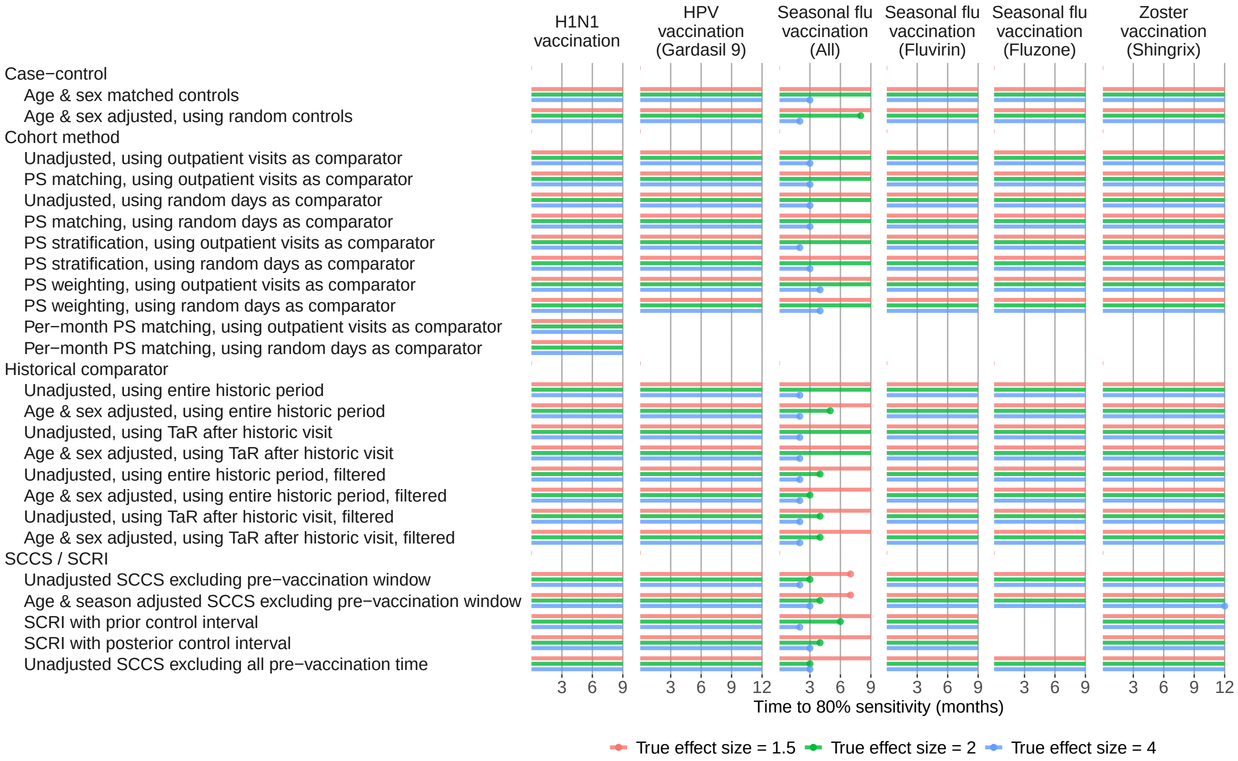

Figure 17: Time to 80% sensitivity in the CCAE database.

### 10 Type 1 and 2 error by true effect size based on p-value

For each method variation and vaccine group, at the end of the study period, the type 1 error and the type 2 error stratified by true effect size, based on whether $p < 0.05$.

#### 10.1 Type 1 and 2 error for H1N1 vaccination

| | Uncalibrated | | | | Calibrated | | | |
|---|---|---|---|---|---|---|---|---|
| | | Type 2 when true effect is | | | | Type 2 when true effect is | | |
| | Type 1 | 1.5 | 2 | 4 | Type 1 | 1.5 | 2 | 4 |
| **Case−control** | | | | | | | | |
| Age & sex matched controls | 17.2 | 66.7 | 60.2 | 50.5 | 1.1 | 88.2 | 78.5 | 55.9 |
| Age & sex adjusted, using random controls | 20.4 | 63.4 | 58.1 | 47.3 | 1.1 | 87.1 | 76.3 | 58.1 |
| **Cohort method** | | | | | | | | |
| Unadjusted, using outpatient visits as comparator | 4.3 | 89.2 | 74.2 | 44.1 | 4.3 | 82.8 | 58.1 | 40.9 |
| PS matching, using outpatient visits as comparator | 0.0 | 88.2 | 69.9 | 49.5 | 1.1 | 79.6 | 61.3 | 45.2 |
| Unadjusted, using random days as comparator | 20.4 | 67.7 | 63.4 | 51.6 | 0.0 | 87.1 | 74.2 | 58.1 |
| PS matching, using random days as comparator | 0.0 | 100.0 | 100.0 | 100.0 | 0.0 | 100.0 | 100.0 | 100.0 |
| PS stratification, using outpatient visits as comparator | 0.0 | 83.9 | 64.5 | 37.6 | 2.2 | 73.1 | 51.6 | 36.6 |
| PS stratification, using random days as comparator | 38.7 | 45.2 | 39.8 | 34.4 | 0.0 | 73.1 | 53.8 | 37.6 |
| PS weighting, using outpatient visits as comparator | 3.2 | 83.9 | 66.7 | 46.2 | 3.2 | 83.9 | 62.4 | 45.2 |
| PS weighting, using random days as comparator | 0.0 | 100.0 | 100.0 | 100.0 | 0.0 | 100.0 | 100.0 | 100.0 |
| Per−month PS matching, using outpatient visits as comparator | 1.1 | 94.6 | 86.0 | 55.9 | 1.1 | 91.4 | 78.5 | 51.6 |
| Per−month PS matching, using random days as comparator | 1.1 | 92.5 | 88.2 | 73.1 | 0.0 | 93.5 | 89.2 | 76.3 |
| **Historical comparator** | | | | | | | | |
| Unadjusted, using entire historic period | 46.2 | 35.5 | 31.2 | 26.9 | 4.3 | 67.7 | 45.2 | 30.1 |
| Age & sex adjusted, using entire historic period | 46.2 | 36.6 | 30.1 | 26.9 | 4.3 | 61.3 | 41.9 | 30.1 |
| Unadjusted, using TaR after historic visit | 20.4 | 52.7 | 35.5 | 30.1 | 4.3 | 74.2 | 49.5 | 30.1 |
| Age & sex adjusted, using TaR after historic visit | 24.7 | 45.2 | 35.5 | 29.0 | 5.4 | 68.8 | 46.2 | 30.1 |
| Unadjusted, using entire historic period, filtered | 44.1 | 39.8 | 35.5 | 31.2 | 3.2 | 62.4 | 48.4 | 34.4 |
| Age & sex adjusted, using entire historic period, filtered | 43.0 | 40.9 | 34.4 | 31.2 | 3.2 | 62.4 | 44.1 | 34.4 |
| Unadjusted, using TaR after historic visit, filtered | 19.4 | 54.8 | 39.8 | 34.4 | 4.3 | 76.3 | 52.7 | 34.4 |
| Age & sex adjusted, using TaR after historic visit, filtered | 22.6 | 48.4 | 39.8 | 33.3 | 4.3 | 66.7 | 48.4 | 34.4 |
| **SCCS / SCRI** | | | | | | | | |
| Unadjusted SCCS excluding pre−vaccination window | 1.1 | 55.9 | 44.1 | 31.2 | 1.1 | 58.1 | 44.1 | 31.2 |
| Age & season adjusted SCCS excluding pre−vaccination window | 1.1 | 59.1 | 45.2 | 32.3 | 1.1 | 59.1 | 45.2 | 32.3 |
| SCRI with prior control interval | 1.1 | 76.3 | 60.2 | 39.8 | 1.1 | 78.5 | 60.2 | 39.8 |
| SCRI with posterior control interval | 2.2 | 72.0 | 54.8 | 44.1 | 2.2 | 77.4 | 58.1 | 44.1 |
| Unadjusted SCCS excluding all pre−vaccination time | 3.2 | 53.8 | 41.9 | 32.3 | 1.1 | 61.3 | 47.3 | 33.3 |

Figure 18: Type 1 and 2 error based on $p < 0.05$ at the end of the study period in the Optum EHR database for H1N1 vaccination.

| | Uncalibrated | | | | Calibrated | | | |
|---|---|---|---|---|---|---|---|---|
| | | Type 2 when true effect is | | | | Type 2 when true effect is | | |
| | Type 1 | 1.5 | 2 | 4 | Type 1 | 1.5 | 2 | 4 |
| Case−control | | | | | | | | |
| Age & sex matched controls | 10.8 | 63.4 | 54.8 | 44.1 | 2.2 | 74.2 | 59.1 | 44.1 |
| Age & sex adjusted, using random controls | 7.5 | 61.3 | 49.5 | 38.7 | 1.1 | 67.7 | 55.9 | 38.7 |
| Cohort method | | | | | | | | |
| Unadjusted, using outpatient visits as comparator | 3.2 | 94.6 | 92.5 | 66.7 | 4.3 | 90.3 | 83.9 | 49.5 |
| PS matching, using outpatient visits as comparator | 0.0 | 86.0 | 76.3 | 51.6 | 2.2 | 83.9 | 73.1 | 50.5 |
| Unadjusted, using random days as comparator | 4.3 | 90.3 | 79.6 | 55.9 | 3.2 | 89.2 | 77.4 | 50.5 |
| PS matching, using random days as comparator | 1.1 | 82.8 | 69.9 | 53.8 | 1.1 | 81.7 | 67.7 | 53.8 |
| PS stratification, using outpatient visits as comparator | 3.2 | 81.7 | 66.7 | 43.0 | 3.2 | 79.6 | 64.5 | 43.0 |
| PS stratification, using random days as comparator | 3.2 | 76.3 | 60.2 | 45.2 | 2.2 | 76.3 | 61.3 | 45.2 |
| PS weighting, using outpatient visits as comparator | 1.1 | 84.9 | 75.3 | 57.0 | 3.2 | 80.6 | 66.7 | 54.8 |
| PS weighting, using random days as comparator | 0.0 | 91.4 | 83.9 | 64.5 | 0.0 | 90.3 | 81.7 | 64.5 |
| Per−month PS matching, using outpatient visits as comparator | 0.0 | 83.9 | 74.2 | 51.6 | 1.1 | 81.7 | 72.0 | 51.6 |
| Per−month PS matching, using random days as comparator | 2.2 | 84.9 | 74.2 | 58.1 | 2.2 | 83.9 | 73.1 | 54.8 |
| Historical comparator | | | | | | | | |
| Unadjusted, using entire historic period | 3.2 | 89.2 | 72.0 | 44.1 | 3.2 | 86.0 | 69.9 | 39.8 |
| Age & sex adjusted, using entire historic period | 9.7 | 58.1 | 49.5 | 35.5 | 4.3 | 69.9 | 54.8 | 36.6 |
| Unadjusted, using TaR after historic visit | 3.2 | 91.4 | 79.6 | 49.5 | 3.2 | 88.2 | 76.3 | 41.9 |
| Age & sex adjusted, using TaR after historic visit | 7.5 | 66.7 | 52.7 | 37.6 | 4.3 | 71.0 | 54.8 | 37.6 |
| Unadjusted, using entire historic period, filtered | 3.2 | 90.3 | 73.1 | 45.2 | 3.2 | 87.1 | 68.8 | 40.9 |
| Age & sex adjusted, using entire historic period, filtered | 8.6 | 59.1 | 50.5 | 36.6 | 3.2 | 71.0 | 54.8 | 37.6 |
| Unadjusted, using TaR after historic visit, filtered | 3.2 | 92.5 | 80.6 | 50.5 | 3.2 | 89.2 | 76.3 | 43.0 |
| Age & sex adjusted, using TaR after historic visit, filtered | 6.5 | 67.7 | 53.8 | 38.7 | 3.2 | 71.0 | 55.9 | 38.7 |
| SCCS / SCRI | | | | | | | | |
| Unadjusted SCCS excluding pre−vaccination window | 2.2 | 68.8 | 57.0 | 39.8 | 1.1 | 69.9 | 57.0 | 39.8 |
| Age & season adjusted SCCS excluding pre−vaccination window | 2.2 | 66.7 | 55.9 | 40.9 | 2.2 | 66.7 | 53.8 | 40.9 |
| SCRI with prior control interval | 0.0 | 78.5 | 61.3 | 46.2 | 0.0 | 78.5 | 61.3 | 46.2 |
| SCRI with posterior control interval | 4.3 | 75.3 | 62.4 | 48.4 | 3.2 | 77.4 | 63.4 | 48.4 |
| Unadjusted SCCS excluding all pre−vaccination time | 2.2 | 71.0 | 58.1 | 38.7 | 1.1 | 72.0 | 59.1 | 38.7 |

Figure 19: Type 1 and 2 error based on $p < 0.05$ at the end of the study period in the MDCD database for H1N1 vaccination.

| | Uncalibrated | | | | Calibrated | | | |
|---|---|---|---|---|---|---|---|---|
| | | Type 2 when true effect is | | | | Type 2 when true effect is | | |
| | Type 1 | 1.5 | 2 | 4 | Type 1 | 1.5 | 2 | 4 |
| Case−control | | | | | | | | |
| Age & sex matched controls | 8.6 | 81.7 | 75.3 | 63.4 | 2.2 | 89.2 | 81.7 | 65.6 |
| Age & sex adjusted, using random controls | 5.4 | 78.5 | 75.3 | 58.1 | 1.1 | 89.2 | 80.6 | 63.4 |
| Cohort method | | | | | | | | |
| Unadjusted, using outpatient visits as comparator | 0.0 | 96.8 | 87.1 | 72.0 | 1.1 | 93.5 | 81.7 | 66.7 |
| PS matching, using outpatient visits as comparator | 0.0 | 96.8 | 93.5 | 80.6 | 0.0 | 96.8 | 90.3 | 80.6 |
| Unadjusted, using random days as comparator | 1.1 | 89.2 | 87.1 | 78.5 | 0.0 | 93.5 | 88.2 | 79.6 |
| PS matching, using random days as comparator | 0.0 | 94.6 | 91.4 | 80.6 | 0.0 | 96.8 | 92.5 | 83.9 |
| PS stratification, using outpatient visits as comparator | 0.0 | 84.9 | 79.6 | 62.4 | 0.0 | 87.1 | 79.6 | 62.4 |
| PS stratification, using random days as comparator | 5.4 | 86.0 | 78.5 | 71.0 | 0.0 | 88.2 | 80.6 | 71.0 |
| PS weighting, using outpatient visits as comparator | 6.5 | 88.2 | 83.9 | 71.0 | 1.1 | 94.6 | 93.5 | 83.9 |
| PS weighting, using random days as comparator | 0.0 | 97.8 | 93.5 | 86.0 | 0.0 | 97.8 | 95.7 | 87.1 |
| Per−month PS matching, using outpatient visits as comparator | 0.0 | 97.8 | 90.3 | 79.6 | 0.0 | 97.8 | 90.3 | 79.6 |
| Per−month PS matching, using random days as comparator | 0.0 | 96.8 | 95.7 | 80.6 | 0.0 | 96.8 | 94.6 | 80.6 |
| Historical comparator | | | | | | | | |
| Unadjusted, using entire historic period | 10.8 | 67.7 | 63.4 | 53.8 | 3.2 | 87.1 | 73.1 | 59.1 |
| Age & sex adjusted, using entire historic period | 11.8 | 67.7 | 63.4 | 54.8 | 4.3 | 87.1 | 72.0 | 59.1 |
| Unadjusted, using TaR after historic visit | 6.5 | 69.9 | 64.5 | 54.8 | 4.3 | 89.2 | 71.0 | 59.1 |
| Age & sex adjusted, using TaR after historic visit | 6.5 | 69.9 | 64.5 | 54.8 | 4.3 | 90.3 | 69.9 | 59.1 |
| Unadjusted, using entire historic period, filtered | 9.7 | 68.8 | 64.5 | 54.8 | 3.2 | 88.2 | 73.1 | 60.2 |
| Age & sex adjusted, using entire historic period, filtered | 10.8 | 68.8 | 64.5 | 55.9 | 3.2 | 88.2 | 73.1 | 60.2 |
| Unadjusted, using TaR after historic visit, filtered | 5.4 | 71.0 | 65.6 | 55.9 | 3.2 | 88.2 | 71.0 | 60.2 |
| Age & sex adjusted, using TaR after historic visit, filtered | 5.4 | 71.0 | 65.6 | 55.9 | 3.2 | 88.2 | 71.0 | 60.2 |
| SCCS / SCRI | | | | | | | | |
| Unadjusted SCCS excluding pre−vaccination window | 3.2 | 83.9 | 71.0 | 59.1 | 3.2 | 87.1 | 72.0 | 59.1 |
| Age & season adjusted SCCS excluding pre−vaccination window | 3.2 | 80.6 | 71.0 | 58.1 | 3.2 | 87.1 | 73.1 | 59.1 |
| SCRI with prior control interval | 2.2 | 91.4 | 82.8 | 66.7 | 0.0 | 94.6 | 88.2 | 73.1 |
| SCRI with posterior control interval | 0.0 | 93.5 | 82.8 | 68.8 | 0.0 | 93.5 | 82.8 | 68.8 |
| Unadjusted SCCS excluding all pre−vaccination time | 2.2 | 81.7 | 72.0 | 61.3 | 1.1 | 84.9 | 73.1 | 61.3 |

Figure 20: Type 1 and 2 error based on $p < 0.05$ at the end of the study period in the MDCR database for H1N1 vaccination.

| | Uncalibrated | | | | Calibrated | | | |
|---|---|---|---|---|---|---|---|---|
| | | Type 2 when true effect is | | | | Type 2 when true effect is | | |
| | Type 1 | 1.5 | 2 | 4 | Type 1 | 1.5 | 2 | 4 |
| Case−control | | | | | | | | |
| Age & sex matched controls | 18.3 | 67.7 | 55.9 | 46.2 | 2.2 | 73.1 | 66.7 | 49.5 |
| Age & sex adjusted, using random controls | 19.4 | 64.5 | 54.8 | 38.7 | 1.1 | 74.2 | 59.1 | 46.2 |
| Cohort method | | | | | | | | |
| Unadjusted, using outpatient visits as comparator | 3.2 | 89.2 | 79.6 | 33.3 | 4.3 | 81.7 | 61.3 | 22.6 |
| PS matching, using outpatient visits as comparator | 0.0 | 71.0 | 41.9 | 29.0 | 4.3 | 59.1 | 35.5 | 28.0 |
| Unadjusted, using random days as comparator | 12.9 | 52.7 | 33.3 | 22.6 | 3.2 | 69.9 | 36.6 | 25.8 |
| PS matching, using random days as comparator | 5.4 | 57.0 | 40.9 | 28.0 | 4.3 | 58.1 | 40.9 | 28.0 |
| PS stratification, using outpatient visits as comparator | 0.0 | 66.7 | 38.7 | 25.8 | 4.3 | 53.8 | 29.0 | 23.7 |
| PS stratification, using random days as comparator | 7.5 | 49.5 | 34.4 | 23.7 | 4.3 | 51.6 | 34.4 | 23.7 |
| PS weighting, using outpatient visits as comparator | 0.0 | 77.4 | 57.0 | 31.2 | 3.2 | 67.7 | 47.3 | 25.8 |
| PS weighting, using random days as comparator | 1.1 | 80.6 | 65.6 | 32.3 | 2.2 | 77.4 | 57.0 | 30.1 |
| Per−month PS matching, using outpatient visits as comparator | 1.1 | 68.8 | 41.9 | 30.1 | 3.2 | 51.6 | 34.4 | 26.9 |
| Per−month PS matching, using random days as comparator | 3.2 | 57.0 | 40.9 | 29.0 | 1.1 | 58.1 | 40.9 | 29.0 |
| Historical comparator | | | | | | | | |
| Unadjusted, using entire historic period | 14.0 | 38.7 | 25.8 | 20.4 | 5.4 | 60.2 | 29.0 | 21.5 |
| Age & sex adjusted, using entire historic period | 31.2 | 33.3 | 24.7 | 20.4 | 2.2 | 49.5 | 32.3 | 20.4 |
| Unadjusted, using TaR after historic visit | 8.6 | 62.4 | 33.3 | 22.6 | 4.3 | 73.1 | 36.6 | 22.6 |
| Age & sex adjusted, using TaR after historic visit | 9.7 | 45.2 | 30.1 | 20.4 | 3.2 | 54.8 | 31.2 | 20.4 |
| Unadjusted, using entire historic period, filtered | 14.0 | 39.8 | 26.9 | 21.5 | 5.4 | 59.1 | 30.1 | 22.6 |
| Age & sex adjusted, using entire historic period, filtered | 30.1 | 34.4 | 25.8 | 21.5 | 2.2 | 49.5 | 33.3 | 21.5 |
| Unadjusted, using TaR after historic visit, filtered | 8.6 | 63.4 | 34.4 | 23.7 | 4.3 | 74.2 | 36.6 | 23.7 |
| Age & sex adjusted, using TaR after historic visit, filtered | 9.7 | 46.2 | 31.2 | 21.5 | 3.2 | 55.9 | 32.3 | 21.5 |
| SCCS / SCRI | | | | | | | | |
| Unadjusted SCCS excluding pre−vaccination window | 7.5 | 38.7 | 26.9 | 21.5 | 2.2 | 49.5 | 28.0 | 21.5 |
| Age & season adjusted SCCS excluding pre−vaccination window | 8.6 | 38.7 | 29.0 | 23.7 | 3.2 | 41.9 | 29.0 | 23.7 |
| SCRI with prior control interval | 7.5 | 57.0 | 34.4 | 22.6 | 0.0 | 63.4 | 36.6 | 22.6 |
| SCRI with posterior control interval | 9.7 | 51.6 | 30.1 | 24.7 | 5.4 | 54.8 | 33.3 | 24.7 |
| Unadjusted SCCS excluding all pre−vaccination time | 11.8 | 41.9 | 28.0 | 21.5 | 3.2 | 49.5 | 29.0 | 21.5 |

Figure 21: Type 1 and 2 error based on $p < 0.05$ at the end of the study period in the CCAE database for H1N1 vaccination.

#### 10.2 Type 1 and 2 error for Seasonal flu vaccination (Fluvirin)

| | Uncalibrated | | | | Calibrated | | | |
|---|---|---|---|---|---|---|---|---|
| | | Type 2 when true effect is | | | | Type 2 when true effect is | | |
| | Type 1 | 1.5 | 2 | 4 | Type 1 | 1.5 | 2 | 4 |
| Case−control | | | | | | | | |
| Age & sex matched controls | 1.1 | 94.6 | 87.1 | 81.7 | 0.0 | 98.9 | 96.8 | 83.9 |
| Age & sex adjusted, using random controls | 2.2 | 94.6 | 89.2 | 87.1 | 0.0 | 100.0 | 97.8 | 89.2 |
| Cohort method | | | | | | | | |
| Unadjusted, using outpatient visits as comparator | 0.0 | 95.7 | 89.2 | 72.0 | 2.2 | 90.3 | 79.6 | 69.9 |
| PS matching, using outpatient visits as comparator | 0.0 | 98.9 | 97.8 | 89.2 | 0.0 | 97.8 | 96.8 | 88.2 |
| Unadjusted, using random days as comparator | 0.0 | 92.5 | 83.9 | 75.3 | 0.0 | 95.7 | 90.3 | 79.6 |
| PS matching, using random days as comparator | | | | | | | | |
| PS stratification, using outpatient visits as comparator | 2.2 | 95.7 | 92.5 | 74.2 | 2.2 | 93.5 | 89.2 | 69.9 |
| PS stratification, using random days as comparator | 6.5 | 79.6 | 67.7 | 58.1 | 1.1 | 89.2 | 79.6 | 62.4 |
| PS weighting, using outpatient visits as comparator | 0.0 | 96.8 | 94.6 | 84.9 | 0.0 | 95.7 | 93.5 | 82.8 |
| PS weighting, using random days as comparator | 0.0 | 100.0 | 100.0 | 100.0 | 0.0 | 100.0 | 100.0 | 100.0 |
| Per−month PS matching, using outpatient visits as comparator | | | | | | | | |
| Per−month PS matching, using random days as comparator | | | | | | | | |
| Historical comparator | | | | | | | | |
| Unadjusted, using entire historic period | 18.3 | 62.4 | 55.9 | 53.8 | 4.3 | 92.5 | 78.5 | 57.0 |
| Age & sex adjusted, using entire historic period | 17.2 | 64.5 | 57.0 | 52.7 | 4.3 | 89.2 | 76.3 | 55.9 |
| Unadjusted, using TaR after historic visit | 9.7 | 73.1 | 64.5 | 53.8 | 3.2 | 89.2 | 79.6 | 58.1 |
| Age & sex adjusted, using TaR after historic visit | 8.6 | 74.2 | 64.5 | 53.8 | 4.3 | 86.0 | 76.3 | 57.0 |
| Unadjusted, using entire historic period, filtered | 17.2 | 63.4 | 57.0 | 54.8 | 5.4 | 90.3 | 75.3 | 55.9 |
| Age & sex adjusted, using entire historic period, filtered | 16.1 | 65.6 | 58.1 | 53.8 | 3.2 | 89.2 | 73.1 | 55.9 |
| Unadjusted, using TaR after historic visit, filtered | 8.6 | 74.2 | 65.6 | 54.8 | 4.3 | 89.2 | 76.3 | 57.0 |
| Age & sex adjusted, using TaR after historic visit, filtered | 7.5 | 75.3 | 65.6 | 54.8 | 3.2 | 87.1 | 75.3 | 58.1 |
| SCCS / SCRI | | | | | | | | |
| Unadjusted SCCS excluding pre−vaccination window | 2.2 | 86.0 | 76.3 | 63.4 | 1.1 | 87.1 | 78.5 | 64.5 |
| Age & season adjusted SCCS excluding pre−vaccination window | 3.2 | 87.1 | 76.3 | 65.6 | 1.1 | 89.2 | 80.6 | 66.7 |
| SCRI with prior control interval | 0.0 | 95.7 | 88.2 | 76.3 | 0.0 | 96.8 | 89.2 | 77.4 |
| SCRI with posterior control interval | 0.0 | 94.6 | 82.8 | 71.0 | 0.0 | 96.8 | 83.9 | 78.5 |
| Unadjusted SCCS excluding all pre−vaccination time | 2.2 | 80.6 | 73.1 | 63.4 | 0.0 | 91.4 | 76.3 | 63.4 |

Figure 22: Type 1 and 2 error based on $p < 0.05$ at the end of the study period in the Optum EHR database for Seasonal flu vaccination (Fluvirin).

| | Uncalibrated | | | | Calibrated | | | |
|---|---|---|---|---|---|---|---|---|
| | | Type 2 when true effect is | | | | Type 2 when true effect is | | |
| | Type 1 | 1.5 | 2 | 4 | Type 1 | 1.5 | 2 | 4 |
| Case−control | | | | | | | | |
| Age & sex matched controls | 4.3 | 87.1 | 83.9 | 75.3 | 1.1 | 92.5 | 87.1 | 77.4 |
| Age & sex adjusted, using random controls | 1.1 | 86.0 | 76.3 | 67.7 | 0.0 | 91.4 | 83.9 | 69.9 |
| Cohort method | | | | | | | | |
| Unadjusted, using outpatient visits as comparator | 1.1 | 92.5 | 88.2 | 71.0 | 2.2 | 89.2 | 86.0 | 71.0 |
| PS matching, using outpatient visits as comparator | 0.0 | 100.0 | 100.0 | 100.0 | 0.0 | 100.0 | 100.0 | 100.0 |
| Unadjusted, using random days as comparator | 4.3 | 88.2 | 86.0 | 77.4 | 2.2 | 94.6 | 90.3 | 81.7 |
| PS matching, using random days as comparator | 0.0 | 100.0 | 100.0 | 100.0 | 0.0 | 100.0 | 100.0 | 100.0 |
| PS stratification, using outpatient visits as comparator | 0.0 | 91.4 | 81.7 | 67.7 | 2.2 | 87.1 | 79.6 | 67.7 |
| PS stratification, using random days as comparator | 7.5 | 80.6 | 73.1 | 65.6 | 0.0 | 87.1 | 80.6 | 67.7 |
| PS weighting, using outpatient visits as comparator | 0.0 | 98.9 | 94.6 | 81.7 | 0.0 | 97.8 | 93.5 | 81.7 |
| PS weighting, using random days as comparator | 0.0 | 100.0 | 100.0 | 100.0 | 0.0 | 100.0 | 100.0 | 100.0 |
| Per−month PS matching, using outpatient visits as comparator | | | | | | | | |
| Per−month PS matching, using random days as comparator | | | | | | | | |
| Historical comparator | | | | | | | | |
| Unadjusted, using entire historic period | 11.8 | 75.3 | 67.7 | 61.3 | 1.1 | 98.9 | 91.4 | 67.7 |
| Age & sex adjusted, using entire historic period | 4.3 | 81.7 | 72.0 | 61.3 | 1.1 | 97.8 | 88.2 | 67.7 |
| Unadjusted, using TaR after historic visit | 5.4 | 78.5 | 72.0 | 61.3 | 1.1 | 97.8 | 90.3 | 67.7 |
| Age & sex adjusted, using TaR after historic visit | 2.2 | 86.0 | 78.5 | 64.5 | 1.1 | 96.8 | 89.2 | 66.7 |
| Unadjusted, using entire historic period, filtered | 10.8 | 76.3 | 68.8 | 62.4 | 0.0 | 88.2 | 79.6 | 64.5 |
| Age & sex adjusted, using entire historic period, filtered | 3.2 | 82.8 | 73.1 | 62.4 | 1.1 | 86.0 | 78.5 | 64.5 |
| Unadjusted, using TaR after historic visit, filtered | 4.3 | 79.6 | 73.1 | 62.4 | 2.2 | 87.1 | 79.6 | 65.6 |
| Age & sex adjusted, using TaR after historic visit, filtered | 1.1 | 87.1 | 79.6 | 65.6 | 1.1 | 88.2 | 79.6 | 65.6 |
| SCCS / SCRI | | | | | | | | |
| Unadjusted SCCS excluding pre−vaccination window | 1.1 | 86.0 | 77.4 | 64.5 | 1.1 | 87.1 | 77.4 | 64.5 |
| Age & season adjusted SCCS excluding pre−vaccination window | 1.1 | 87.1 | 78.5 | 65.6 | 1.1 | 87.1 | 78.5 | 65.6 |
| SCRI with prior control interval | 0.0 | 94.6 | 88.2 | 80.6 | 1.1 | 93.5 | 87.1 | 79.6 |
| SCRI with posterior control interval | 0.0 | 91.4 | 83.9 | 68.8 | 0.0 | 91.4 | 86.0 | 68.8 |
| Unadjusted SCCS excluding all pre−vaccination time | 2.2 | 84.9 | 77.4 | 64.5 | 1.1 | 86.0 | 78.5 | 64.5 |

Figure 23: Type 1 and 2 error based on $p < 0.05$ at the end of the study period in the MDCD database for Seasonal flu vaccination (Fluvirin).

| | Uncalibrated | | | | Calibrated | | | |
|---|---|---|---|---|---|---|---|---|
| | | Type 2 when true effect is | | | | Type 2 when true effect is | | |
| | Type 1 | 1.5 | 2 | 4 | Type 1 | 1.5 | 2 | 4 |
| Case−control | | | | | | | | |
| Age & sex matched controls | 0.0 | 98.9 | 97.8 | 95.7 | 0.0 | 100.0 | 100.0 | 100.0 |
| Age & sex adjusted, using random controls | 0.0 | 100.0 | 97.8 | 94.6 | 0.0 | 100.0 | 100.0 | 100.0 |
| Cohort method | | | | | | | | |
| Unadjusted, using outpatient visits as comparator | 0.0 | 100.0 | 100.0 | 97.8 | 0.0 | 100.0 | 100.0 | 100.0 |
| PS matching, using outpatient visits as comparator | 0.0 | 100.0 | 100.0 | 98.9 | 0.0 | 100.0 | 100.0 | 100.0 |
| Unadjusted, using random days as comparator | 0.0 | 100.0 | 98.9 | 96.8 | 0.0 | 100.0 | 100.0 | 100.0 |
| PS matching, using random days as comparator | 0.0 | 100.0 | 100.0 | 98.9 | 0.0 | 100.0 | 100.0 | 100.0 |
| PS stratification, using outpatient visits as comparator | 1.1 | 98.9 | 97.8 | 95.7 | 0.0 | 98.9 | 98.9 | 95.7 |
| PS stratification, using random days as comparator | 0.0 | 98.9 | 97.8 | 96.8 | 0.0 | 100.0 | 100.0 | 100.0 |
| PS weighting, using outpatient visits as comparator | 1.1 | 98.9 | 98.9 | 97.8 | 1.1 | 98.9 | 98.9 | 98.9 |
| PS weighting, using random days as comparator | 0.0 | 100.0 | 100.0 | 98.9 | 0.0 | 100.0 | 100.0 | 100.0 |
| Per−month PS matching, using outpatient visits as comparator | | | | | | | | |
| Per−month PS matching, using random days as comparator | | | | | | | | |
| Historical comparator | | | | | | | | |
| Unadjusted, using entire historic period | 2.2 | 94.6 | 93.5 | 91.4 | 0.0 | 97.8 | 96.8 | 93.5 |
| Age & sex adjusted, using entire historic period | 2.2 | 94.6 | 93.5 | 91.4 | 0.0 | 97.8 | 96.8 | 93.5 |
| Unadjusted, using TaR after historic visit | 1.1 | 94.6 | 93.5 | 91.4 | 0.0 | 98.9 | 96.8 | 93.5 |
| Age & sex adjusted, using TaR after historic visit | 1.1 | 94.6 | 93.5 | 91.4 | 0.0 | 97.8 | 96.8 | 93.5 |
| Unadjusted, using entire historic period, filtered | 2.2 | 94.6 | 93.5 | 91.4 | 0.0 | 97.8 | 96.8 | 93.5 |
| Age & sex adjusted, using entire historic period, filtered | 2.2 | 94.6 | 93.5 | 91.4 | 0.0 | 97.8 | 96.8 | 93.5 |
| Unadjusted, using TaR after historic visit, filtered | 1.1 | 94.6 | 93.5 | 91.4 | 0.0 | 98.9 | 96.8 | 93.5 |
| Age & sex adjusted, using TaR after historic visit, filtered | 1.1 | 94.6 | 93.5 | 91.4 | 0.0 | 97.8 | 96.8 | 93.5 |
| SCCS / SCRI | | | | | | | | |
| Unadjusted SCCS excluding pre−vaccination window | 0.0 | 97.8 | 96.8 | 95.7 | 0.0 | 100.0 | 100.0 | 100.0 |
| Age & season adjusted SCCS excluding pre−vaccination window | 0.0 | 97.8 | 96.8 | 95.7 | 0.0 | 100.0 | 100.0 | 100.0 |
| SCRI with prior control interval | 0.0 | 100.0 | 98.9 | 97.8 | 0.0 | 100.0 | 100.0 | 100.0 |
| SCRI with posterior control interval | 0.0 | 100.0 | 98.9 | 98.9 | 0.0 | 100.0 | 100.0 | 100.0 |
| Unadjusted SCCS excluding all pre−vaccination time | 0.0 | 98.9 | 97.8 | 96.8 | 0.0 | 100.0 | 100.0 | 100.0 |

Figure 24: Type 1 and 2 error based on $p < 0.05$ at the end of the study period in the MDCR database for Seasonal flu vaccination (Fluvirin).

| | Uncalibrated | | | | Calibrated | | | |
|---|---|---|---|---|---|---|---|---|
| | | Type 2 when true effect is | | | | Type 2 when true effect is | | |
| | Type 1 | 1.5 | 2 | 4 | Type 1 | 1.5 | 2 | 4 |
| Case−control | | | | | | | | |
| Age & sex matched controls | 3.2 | 82.8 | 73.1 | 57.0 | 0.0 | 84.9 | 78.5 | 60.2 |
| Age & sex adjusted, using random controls | 4.3 | 83.9 | 73.1 | 55.9 | 2.2 | 86.0 | 76.3 | 55.9 |
| Cohort method | | | | | | | | |
| Unadjusted, using outpatient visits as comparator | 0.0 | 91.4 | 78.5 | 52.7 | 3.2 | 77.4 | 64.5 | 43.0 |
| PS matching, using outpatient visits as comparator | 1.1 | 82.8 | 69.9 | 50.5 | 2.2 | 79.6 | 68.8 | 50.5 |
| Unadjusted, using random days as comparator | 9.7 | 75.3 | 71.0 | 48.4 | 1.1 | 82.8 | 72.0 | 51.6 |
| PS matching, using random days as comparator | 2.2 | 77.4 | 66.7 | 45.2 | 1.1 | 78.5 | 66.7 | 47.3 |
| PS stratification, using outpatient visits as comparator | 2.2 | 76.3 | 64.5 | 44.1 | 3.2 | 75.3 | 60.2 | 43.0 |
| PS stratification, using random days as comparator | 3.2 | 69.9 | 58.1 | 36.6 | 2.2 | 72.0 | 59.1 | 38.7 |
| PS weighting, using outpatient visits as comparator | 3.2 | 81.7 | 71.0 | 54.8 | 3.2 | 81.7 | 66.7 | 53.8 |
| PS weighting, using random days as comparator | 0.0 | 87.1 | 81.7 | 60.2 | 0.0 | 87.1 | 83.9 | 61.3 |
| Per−month PS matching, using outpatient visits as comparator | | | | | | | | |
| Per−month PS matching, using random days as comparator | | | | | | | | |
| Historical comparator | | | | | | | | |
| Unadjusted, using entire historic period | 17.2 | 60.2 | 49.5 | 34.4 | 2.2 | 92.5 | 74.2 | 39.8 |
| Age & sex adjusted, using entire historic period | 7.5 | 63.4 | 53.8 | 35.5 | 2.2 | 92.5 | 69.9 | 39.8 |
| Unadjusted, using TaR after historic visit | 8.6 | 71.0 | 53.8 | 37.6 | 2.2 | 92.5 | 74.2 | 39.8 |
| Age & sex adjusted, using TaR after historic visit | 6.5 | 72.0 | 55.9 | 38.7 | 3.2 | 91.4 | 71.0 | 39.8 |
| Unadjusted, using entire historic period, filtered | 15.1 | 62.4 | 51.6 | 36.6 | 3.2 | 74.2 | 59.1 | 38.7 |
| Age & sex adjusted, using entire historic period, filtered | 5.4 | 65.6 | 55.9 | 37.6 | 2.2 | 72.0 | 55.9 | 40.9 |
| Unadjusted, using TaR after historic visit, filtered | 6.5 | 73.1 | 55.9 | 39.8 | 2.2 | 75.3 | 58.1 | 39.8 |
| Age & sex adjusted, using TaR after historic visit, filtered | 4.3 | 74.2 | 58.1 | 40.9 | 3.2 | 73.1 | 58.1 | 40.9 |
| SCCS / SCRI | | | | | | | | |
| Unadjusted SCCS excluding pre−vaccination window | 7.5 | 62.4 | 53.8 | 36.6 | 3.2 | 67.7 | 54.8 | 37.6 |
| Age & season adjusted SCCS excluding pre−vaccination window | 8.6 | 66.7 | 55.9 | 39.8 | 2.2 | 71.0 | 58.1 | 39.8 |
| SCRI with prior control interval | 4.3 | 77.4 | 67.7 | 53.8 | 1.1 | 81.7 | 71.0 | 54.8 |
| SCRI with posterior control interval | 6.5 | 75.3 | 66.7 | 52.7 | 1.1 | 78.5 | 71.0 | 54.8 |
| Unadjusted SCCS excluding all pre−vaccination time | 7.5 | 62.4 | 54.8 | 38.7 | 4.3 | 66.7 | 57.0 | 39.8 |

Figure 25: Type 1 and 2 error based on $p < 0.05$ at the end of the study period in the CCAE database for Seasonal flu vaccination (Fluvirin).

#### 10.3 Type 1 and 2 error for Seasonal flu vaccination (Fluzone)

| | Uncalibrated | | | | Calibrated | | | |
|---|---|---|---|---|---|---|---|---|
| | | Type 2 when true effect is | | | | Type 2 when true effect is | | |
| | Type 1 | 1.5 | 2 | 4 | Type 1 | 1.5 | 2 | 4 |
| Case−control | | | | | | | | |
| Age & sex matched controls | 20.4 | 52.7 | 43.0 | 35.5 | 3.2 | 79.6 | 57.0 | 36.6 |
| Age & sex adjusted, using random controls | 31.2 | 45.2 | 36.6 | 26.9 | 2.2 | 75.3 | 57.0 | 32.3 |
| Cohort method | | | | | | | | |
| Unadjusted, using outpatient visits as comparator | 2.2 | 69.9 | 43.0 | 17.2 | 3.2 | 63.4 | 41.9 | 12.9 |
| PS matching, using outpatient visits as comparator | 1.1 | 76.3 | 49.5 | 33.3 | 5.4 | 58.1 | 44.1 | 30.1 |
| Unadjusted, using random days as comparator | 37.6 | 38.7 | 25.8 | 21.5 | 3.2 | 63.4 | 40.9 | 21.5 |
| PS matching, using random days as comparator | | | | | | | | |
| PS stratification, using outpatient visits as comparator | 1.1 | 66.7 | 45.2 | 16.1 | 8.6 | 57.0 | 37.6 | 11.8 |
| PS stratification, using random days as comparator | 44.1 | 29.0 | 20.4 | 10.8 | 2.2 | 62.4 | 38.7 | 12.9 |
| PS weighting, using outpatient visits as comparator | 1.1 | 78.5 | 53.8 | 34.4 | 2.2 | 66.7 | 44.1 | 25.8 |
| PS weighting, using random days as comparator | 0.0 | 100.0 | 100.0 | 100.0 | 0.0 | 100.0 | 100.0 | 100.0 |
| Per−month PS matching, using outpatient visits as comparator | | | | | | | | |
| Per−month PS matching, using random days as comparator | | | | | | | | |
| Historical comparator | | | | | | | | |
| Unadjusted, using entire historic period | 58.1 | 22.6 | 12.9 | 8.6 | 2.2 | 69.9 | 39.8 | 11.8 |
| Age & sex adjusted, using entire historic period | 59.1 | 23.7 | 15.1 | 8.6 | 2.2 | 68.8 | 37.6 | 11.8 |
| Unadjusted, using TaR after historic visit | 24.7 | 37.6 | 22.6 | 8.6 | 2.2 | 66.7 | 39.8 | 11.8 |
| Age & sex adjusted, using TaR after historic visit | 22.6 | 37.6 | 23.7 | 8.6 | 2.2 | 73.1 | 37.6 | 9.7 |
| Unadjusted, using entire historic period, filtered | 57.0 | 23.7 | 14.0 | 9.7 | 1.1 | 65.6 | 35.5 | 12.9 |
| Age & sex adjusted, using entire historic period, filtered | 58.1 | 24.7 | 16.1 | 9.7 | 2.2 | 61.3 | 34.4 | 10.8 |
| Unadjusted, using TaR after historic visit, filtered | 23.7 | 38.7 | 23.7 | 9.7 | 1.1 | 61.3 | 36.6 | 10.8 |
| Age & sex adjusted, using TaR after historic visit, filtered | 21.5 | 38.7 | 24.7 | 9.7 | 1.1 | 69.9 | 34.4 | 10.8 |
| SCCS / SCRI | | | | | | | | |
| Unadjusted SCCS excluding pre−vaccination window | 20.4 | 31.2 | 19.4 | 9.7 | 3.2 | 47.3 | 26.9 | 9.7 |
| Age & season adjusted SCCS excluding pre−vaccination window | 26.9 | 37.6 | 23.7 | 15.1 | 3.2 | 58.1 | 35.5 | 15.1 |
| SCRI with prior control interval | 29.0 | 36.6 | 28.0 | 20.4 | 3.2 | 68.8 | 40.9 | 20.4 |
| SCRI with posterior control interval | 17.2 | 44.1 | 28.0 | 15.1 | 4.3 | 53.8 | 36.6 | 19.4 |
| Unadjusted SCCS excluding all pre−vaccination time | 30.1 | 29.0 | 20.4 | 8.6 | 4.3 | 47.3 | 25.8 | 10.8 |

Figure 26: Type 1 and 2 error based on $p < 0.05$ at the end of the study period in the Optum EHR database for Seasonal flu vaccination (Fluzone).

| | Uncalibrated | | | | Calibrated | | | |
|---|---|---|---|---|---|---|---|---|
| | | Type 2 when true effect is | | | | Type 2 when true effect is | | |
| | Type 1 | 1.5 | 2 | 4 | Type 1 | 1.5 | 2 | 4 |
| Case−control | | | | | | | | |
| Age & sex matched controls | 0.0 | 95.7 | 93.5 | 87.1 | 0.0 | 100.0 | 96.8 | 91.4 |
| Age & sex adjusted, using random controls | 1.1 | 95.7 | 93.5 | 91.4 | 0.0 | 98.9 | 96.8 | 93.5 |
| Cohort method | | | | | | | | |
| Unadjusted, using outpatient visits as comparator | 0.0 | 98.9 | 97.8 | 89.2 | 0.0 | 98.9 | 98.9 | 94.6 |
| PS matching, using outpatient visits as comparator | 0.0 | 100.0 | 98.9 | 95.7 | 0.0 | 100.0 | 98.9 | 95.7 |
| Unadjusted, using random days as comparator | 1.1 | 97.8 | 97.8 | 93.5 | 0.0 | 98.9 | 97.8 | 95.7 |
| PS matching, using random days as comparator | | | | | | | | |
| PS stratification, using outpatient visits as comparator | 1.1 | 93.5 | 92.5 | 86.0 | 1.1 | 94.6 | 93.5 | 87.1 |
| PS stratification, using random days as comparator | 2.2 | 90.3 | 86.0 | 80.6 | 0.0 | 97.8 | 90.3 | 83.9 |
| PS weighting, using outpatient visits as comparator | 0.0 | 98.9 | 97.8 | 92.5 | 0.0 | 98.9 | 98.9 | 92.5 |
| PS weighting, using random days as comparator | 0.0 | 100.0 | 100.0 | 100.0 | 0.0 | 100.0 | 100.0 | 100.0 |
| Per−month PS matching, using outpatient visits as comparator | | | | | | | | |
| Per−month PS matching, using random days as comparator | | | | | | | | |
| Historical comparator | | | | | | | | |
| Unadjusted, using entire historic period | 11.8 | 80.6 | 76.3 | 75.3 | 3.2 | 95.7 | 92.5 | 81.7 |
| Age & sex adjusted, using entire historic period | 7.5 | 81.7 | 78.5 | 75.3 | 1.1 | 92.5 | 88.2 | 77.4 |
| Unadjusted, using TaR after historic visit | 9.7 | 82.8 | 77.4 | 75.3 | 2.2 | 94.6 | 92.5 | 82.8 |
| Age & sex adjusted, using TaR after historic visit | 4.3 | 87.1 | 83.9 | 76.3 | 3.2 | 92.5 | 87.1 | 77.4 |
| Unadjusted, using entire historic period, filtered | 10.8 | 81.7 | 77.4 | 76.3 | 2.2 | 95.7 | 92.5 | 81.7 |
| Age & sex adjusted, using entire historic period, filtered | 6.5 | 82.8 | 79.6 | 76.3 | 2.2 | 93.5 | 88.2 | 78.5 |
| Unadjusted, using TaR after historic visit, filtered | 8.6 | 83.9 | 78.5 | 76.3 | 2.2 | 94.6 | 92.5 | 82.8 |
| Age & sex adjusted, using TaR after historic visit, filtered | 3.2 | 88.2 | 84.9 | 77.4 | 2.2 | 92.5 | 88.2 | 78.5 |
| SCCS / SCRI | | | | | | | | |
| Unadjusted SCCS excluding pre−vaccination window | 3.2 | 92.5 | 87.1 | 80.6 | 1.1 | 94.6 | 91.4 | 81.7 |
| Age & season adjusted SCCS excluding pre−vaccination window | 3.2 | 91.4 | 87.1 | 80.6 | 1.1 | 96.8 | 90.3 | 81.7 |
| SCRI with prior control interval | 1.1 | 98.9 | 96.8 | 91.4 | 0.0 | 98.9 | 96.8 | 91.4 |
| SCRI with posterior control interval | 0.0 | 98.9 | 96.8 | 87.1 | 0.0 | 98.9 | 96.8 | 87.1 |
| Unadjusted SCCS excluding all pre−vaccination time | 4.3 | 92.5 | 88.2 | 80.6 | 1.1 | 95.7 | 91.4 | 81.7 |

Figure 27: Type 1 and 2 error based on $p < 0.05$ at the end of the study period in the MDCD database for Seasonal flu vaccination (Fluzone).

| | Uncalibrated | | | | Calibrated | | | |
|---|---|---|---|---|---|---|---|---|
| | | Type 2 when true effect is | | | | Type 2 when true effect is | | |
| | Type 1 | 1.5 | 2 | 4 | Type 1 | 1.5 | 2 | 4 |
| Case−control | | | | | | | | |
| Age & sex matched controls | 0.0 | 88.2 | 77.4 | 55.9 | 1.1 | 87.1 | 73.1 | 54.8 |
| Age & sex adjusted, using random controls | 1.1 | 83.9 | 68.8 | 46.2 | 1.1 | 83.9 | 68.8 | 46.2 |
| Cohort method | | | | | | | | |
| Unadjusted, using outpatient visits as comparator | 0.0 | 92.5 | 86.0 | 59.1 | 2.2 | 86.0 | 80.6 | 50.5 |
| PS matching, using outpatient visits as comparator | 0.0 | 93.5 | 84.9 | 67.7 | 0.0 | 88.2 | 83.9 | 65.6 |
| Unadjusted, using random days as comparator | 1.1 | 88.2 | 81.7 | 57.0 | 1.1 | 88.2 | 79.6 | 57.0 |
| PS matching, using random days as comparator | 1.1 | 93.5 | 84.9 | 59.1 | 1.1 | 91.4 | 83.9 | 59.1 |
| PS stratification, using outpatient visits as comparator | 0.0 | 89.2 | 79.6 | 51.6 | 0.0 | 83.9 | 76.3 | 48.4 |
| PS stratification, using random days as comparator | 1.1 | 87.1 | 72.0 | 47.3 | 1.1 | 86.0 | 69.9 | 47.3 |
| PS weighting, using outpatient visits as comparator | 0.0 | 92.5 | 82.8 | 66.7 | 1.1 | 82.8 | 76.3 | 62.4 |
| PS weighting, using random days as comparator | 2.2 | 94.6 | 90.3 | 77.4 | 2.2 | 94.6 | 91.4 | 77.4 |
| Per−month PS matching, using outpatient visits as comparator | | | | | | | | |
| Per−month PS matching, using random days as comparator | | | | | | | | |
| Historical comparator | | | | | | | | |
| Unadjusted, using entire historic period | 6.5 | 76.3 | 61.3 | 39.8 | 4.3 | 87.1 | 75.3 | 41.9 |
| Age & sex adjusted, using entire historic period | 7.5 | 76.3 | 60.2 | 40.9 | 4.3 | 87.1 | 75.3 | 41.9 |
| Unadjusted, using TaR after historic visit | 7.5 | 78.5 | 67.7 | 40.9 | 3.2 | 87.1 | 76.3 | 43.0 |
| Age & sex adjusted, using TaR after historic visit | 7.5 | 78.5 | 68.8 | 40.9 | 3.2 | 87.1 | 75.3 | 41.9 |
| Unadjusted, using entire historic period, filtered | 4.3 | 78.5 | 63.4 | 41.9 | 4.3 | 78.5 | 64.5 | 43.0 |
| Age & sex adjusted, using entire historic period, filtered | 5.4 | 78.5 | 62.4 | 43.0 | 4.3 | 79.6 | 63.4 | 43.0 |
| Unadjusted, using TaR after historic visit, filtered | 5.4 | 80.6 | 69.9 | 43.0 | 4.3 | 82.8 | 68.8 | 43.0 |
| Age & sex adjusted, using TaR after historic visit, filtered | 5.4 | 80.6 | 71.0 | 43.0 | 4.3 | 81.7 | 67.7 | 43.0 |
| SCCS / SCRI | | | | | | | | |
| Unadjusted SCCS excluding pre−vaccination window | 2.2 | 82.8 | 71.0 | 49.5 | 2.2 | 79.6 | 68.8 | 49.5 |
| Age & season adjusted SCCS excluding pre−vaccination window | 1.1 | 79.6 | 68.8 | 50.5 | 1.1 | 79.6 | 68.8 | 50.5 |
| SCRI with prior control interval | 1.1 | 90.3 | 86.0 | 68.8 | 1.1 | 90.3 | 86.0 | 68.8 |
| SCRI with posterior control interval | 2.2 | 87.1 | 81.7 | 62.4 | 2.2 | 87.1 | 81.7 | 62.4 |
| Unadjusted SCCS excluding all pre−vaccination time | 1.1 | 82.8 | 71.0 | 49.5 | 3.2 | 81.7 | 71.0 | 49.5 |

Figure 28: Type 1 and 2 error based on $p < 0.05$ at the end of the study period in the MDCR database for Seasonal flu vaccination (Fluzone).

| | Uncalibrated | | | | Calibrated | | | |
|---|---|---|---|---|---|---|---|---|
| | | Type 2 when true effect is | | | | Type 2 when true effect is | | |
| | Type 1 | 1.5 | 2 | 4 | Type 1 | 1.5 | 2 | 4 |
| Case−control | | | | | | | | |
| Age & sex matched controls | 0.0 | 100.0 | 100.0 | 100.0 | 0.0 | 100.0 | 100.0 | 100.0 |
| Age & sex adjusted, using random controls | 0.0 | 100.0 | 100.0 | 100.0 | 0.0 | 100.0 | 100.0 | 100.0 |
| Cohort method | | | | | | | | |
| Unadjusted, using outpatient visits as comparator | 0.0 | 100.0 | 100.0 | 98.9 | 0.0 | 100.0 | 100.0 | 100.0 |
| PS matching, using outpatient visits as comparator | 0.0 | 100.0 | 100.0 | 98.9 | 0.0 | 100.0 | 100.0 | 100.0 |
| Unadjusted, using random days as comparator | 0.0 | 100.0 | 100.0 | 100.0 | 0.0 | 100.0 | 100.0 | 100.0 |
| PS matching, using random days as comparator | 0.0 | 100.0 | 100.0 | 97.8 | 0.0 | 100.0 | 100.0 | 100.0 |
| PS stratification, using outpatient visits as comparator | 1.1 | 98.9 | 95.7 | 94.6 | 0.0 | 98.9 | 98.9 | 95.7 |
| PS stratification, using random days as comparator | 0.0 | 100.0 | 100.0 | 95.7 | 0.0 | 100.0 | 100.0 | 100.0 |
| PS weighting, using outpatient visits as comparator | 0.0 | 100.0 | 98.9 | 96.8 | 0.0 | 100.0 | 100.0 | 96.8 |
| PS weighting, using random days as comparator | 0.0 | 100.0 | 100.0 | 100.0 | 0.0 | 100.0 | 100.0 | 100.0 |
| Per−month PS matching, using outpatient visits as comparator | | | | | | | | |
| Per−month PS matching, using random days as comparator | | | | | | | | |
| Historical comparator | | | | | | | | |
| Unadjusted, using entire historic period | 8.6 | 91.4 | 88.2 | 88.2 | 1.1 | 98.9 | 97.8 | 94.6 |
| Age & sex adjusted, using entire historic period | 6.5 | 90.3 | 89.2 | 88.2 | 1.1 | 98.9 | 97.8 | 91.4 |
| Unadjusted, using TaR after historic visit | 7.5 | 91.4 | 89.2 | 88.2 | 1.1 | 98.9 | 97.8 | 92.5 |
| Age & sex adjusted, using TaR after historic visit | 5.4 | 91.4 | 90.3 | 88.2 | 1.1 | 98.9 | 97.8 | 91.4 |
| Unadjusted, using entire historic period, filtered | 7.5 | 92.5 | 89.2 | 89.2 | 1.1 | 98.9 | 98.9 | 95.7 |
| Age & sex adjusted, using entire historic period, filtered | 5.4 | 91.4 | 90.3 | 89.2 | 1.1 | 98.9 | 98.9 | 93.5 |
| Unadjusted, using TaR after historic visit, filtered | 6.5 | 92.5 | 90.3 | 89.2 | 1.1 | 98.9 | 97.8 | 94.6 |
| Age & sex adjusted, using TaR after historic visit, filtered | 4.3 | 92.5 | 91.4 | 89.2 | 1.1 | 98.9 | 97.8 | 92.5 |
| SCCS / SCRI | | | | | | | | |
| Unadjusted SCCS excluding pre−vaccination window | 0.0 | 100.0 | 97.8 | 96.8 | 0.0 | 100.0 | 100.0 | 100.0 |
| Age & season adjusted SCCS excluding pre−vaccination window | 0.0 | 98.9 | 97.8 | 96.8 | 0.0 | 100.0 | 100.0 | 100.0 |
| SCRI with prior control interval | | | | | | | | |
| SCRI with posterior control interval | | | | | | | | |
| Unadjusted SCCS excluding all pre−vaccination time | 0.0 | 100.0 | 98.9 | 96.8 | 0.0 | 100.0 | 100.0 | 100.0 |

Figure 29: Type 1 and 2 error based on $p < 0.05$ at the end of the study period in the CCAE database for Seasonal flu vaccination (Fluzone).

#### 10.4 Type 1 and 2 error for Seasonal flu vaccination (All)

| | Uncalibrated | | | | Calibrated | | | |
|---|---|---|---|---|---|---|---|---|
| | | Type 2 when true effect is | | | | Type 2 when true effect is | | |
| | Type 1 | 1.5 | 2 | 4 | Type 1 | 1.5 | 2 | 4 |
| Case−control | | | | | | | | |
| Age & sex matched controls | 65.6 | 12.9 | 9.7 | 5.4 | 5.4 | 40.9 | 19.4 | 8.6 |
| Age & sex adjusted, using random controls | 73.1 | 9.7 | 9.7 | 4.3 | 6.5 | 26.9 | 11.8 | 5.4 |
| Cohort method | | | | | | | | |
| Unadjusted, using outpatient visits as comparator | 2.2 | 48.4 | 18.3 | 1.1 | 3.2 | 41.9 | 11.8 | 1.1 |
| PS matching, using outpatient visits as comparator | 0.0 | 37.6 | 17.2 | 5.4 | 5.4 | 29.0 | 10.8 | 5.4 |
| Unadjusted, using random days as comparator | 73.1 | 11.8 | 4.3 | 3.2 | 4.3 | 34.4 | 12.9 | 3.2 |
| PS matching, using random days as comparator | | | | | | | | |
| PS stratification, using outpatient visits as comparator | 1.1 | 38.7 | 11.8 | 3.2 | 4.3 | 25.8 | 8.6 | 3.2 |
| PS stratification, using random days as comparator | 83.9 | 4.3 | 3.2 | 1.1 | 2.2 | 32.3 | 5.4 | 2.2 |
| PS weighting, using outpatient visits as comparator | 0.0 | 63.4 | 23.7 | 7.5 | 3.2 | 35.5 | 14.0 | 7.5 |
| PS weighting, using random days as comparator | 0.0 | 100.0 | 100.0 | 100.0 | 0.0 | 100.0 | 100.0 | 100.0 |
| Per−month PS matching, using outpatient visits as comparator | | | | | | | | |
| Per−month PS matching, using random days as comparator | | | | | | | | |
| Historical comparator | | | | | | | | |
| Unadjusted, using entire historic period | 90.3 | 1.1 | 1.1 | 0.0 | 1.1 | 34.4 | 7.5 | 1.1 |
| Age & sex adjusted, using entire historic period | 91.4 | 1.1 | 1.1 | 0.0 | 1.1 | 24.7 | 4.3 | 1.1 |
| Unadjusted, using TaR after historic visit | 53.8 | 9.7 | 2.2 | 1.1 | 3.2 | 37.6 | 7.5 | 1.1 |
| Age & sex adjusted, using TaR after historic visit | 61.3 | 5.4 | 2.2 | 0.0 | 3.2 | 25.8 | 4.3 | 1.1 |
| Unadjusted, using entire historic period, filtered | 89.2 | 2.2 | 2.2 | 1.1 | 2.2 | 24.7 | 5.4 | 2.2 |
| Age & sex adjusted, using entire historic period, filtered | 90.3 | 2.2 | 2.2 | 1.1 | 3.2 | 12.9 | 5.4 | 2.2 |
| Unadjusted, using TaR after historic visit, filtered | 52.7 | 10.8 | 3.2 | 2.2 | 3.2 | 23.7 | 6.5 | 2.2 |
| Age & sex adjusted, using TaR after historic visit, filtered | 60.2 | 6.5 | 3.2 | 1.1 | 4.3 | 17.2 | 4.3 | 2.2 |
| SCCS / SCRI | | | | | | | | |
| Unadjusted SCCS excluding pre−vaccination window | 36.6 | 8.6 | 2.2 | 1.1 | 4.3 | 16.1 | 4.3 | 1.1 |
| Age & season adjusted SCCS excluding pre−vaccination window | 45.2 | 15.1 | 9.7 | 8.6 | 3.2 | 28.0 | 12.9 | 8.6 |
| SCRI with prior control interval | 58.1 | 11.8 | 5.4 | 2.2 | 3.2 | 19.4 | 9.7 | 2.2 |
| SCRI with posterior control interval | 47.3 | 6.5 | 4.3 | 2.2 | 4.3 | 16.1 | 6.5 | 3.2 |
| Unadjusted SCCS excluding all pre−vaccination time | 53.8 | 4.3 | 3.2 | 1.1 | 3.2 | 11.8 | 4.3 | 1.1 |

Figure 30: Type 1 and 2 error based on $p < 0.05$ at the end of the study period in the Optum EHR database for Seasonal flu vaccination (All).

| | Uncalibrated | | | | Calibrated | | | |
|---|---|---|---|---|---|---|---|---|
| | | Type 2 when true effect is | | | | Type 2 when true effect is | | |
| | Type 1 | 1.5 | 2 | 4 | Type 1 | 1.5 | 2 | 4 |
| Case−control | | | | | | | | |
| Age & sex matched controls | 46.2 | 32.3 | 26.9 | 23.7 | 5.4 | 46.2 | 32.3 | 23.7 |
| Age & sex adjusted, using random controls | 54.8 | 21.5 | 15.1 | 12.9 | 4.3 | 37.6 | 22.6 | 14.0 |
| Cohort method | | | | | | | | |
| Unadjusted, using outpatient visits as comparator | 6.5 | 78.5 | 45.2 | 15.1 | 7.5 | 69.9 | 31.2 | 14.0 |
| PS matching, using outpatient visits as comparator | 0.0 | 51.6 | 25.8 | 15.1 | 2.2 | 37.6 | 23.7 | 15.1 |
| Unadjusted, using random days as comparator | 45.2 | 22.6 | 19.4 | 14.0 | 7.5 | 48.4 | 22.6 | 14.0 |
| PS matching, using random days as comparator | 3.2 | 44.1 | 32.3 | 14.0 | 2.2 | 41.9 | 30.1 | 14.0 |
| PS stratification, using outpatient visits as comparator | 1.1 | 46.2 | 22.6 | 12.9 | 5.4 | 31.2 | 19.4 | 12.9 |
| PS stratification, using random days as comparator | 7.5 | 40.9 | 22.6 | 15.1 | 4.3 | 46.2 | 23.7 | 15.1 |
| PS weighting, using outpatient visits as comparator | 1.1 | 46.2 | 21.5 | 12.9 | 1.1 | 40.9 | 20.4 | 12.9 |
| PS weighting, using random days as comparator | 1.1 | 59.1 | 34.4 | 17.2 | 4.3 | 53.8 | 31.2 | 16.1 |
| Per−month PS matching, using outpatient visits as comparator | | | | | | | | |
| Per−month PS matching, using random days as comparator | | | | | | | | |
| Historical comparator | | | | | | | | |
| Unadjusted, using entire historic period | 44.1 | 18.3 | 15.1 | 12.9 | 2.2 | 69.9 | 19.4 | 12.9 |
| Age & sex adjusted, using entire historic period | 59.1 | 18.3 | 12.9 | 12.9 | 2.2 | 67.7 | 19.4 | 12.9 |
| Unadjusted, using TaR after historic visit | 17.2 | 30.1 | 18.3 | 12.9 | 3.2 | 73.1 | 28.0 | 12.9 |
| Age & sex adjusted, using TaR after historic visit | 17.2 | 25.8 | 18.3 | 12.9 | 2.2 | 66.7 | 22.6 | 12.9 |
| Unadjusted, using entire historic period, filtered | 43.0 | 19.4 | 16.1 | 14.0 | 5.4 | 32.3 | 18.3 | 14.0 |
| Age & sex adjusted, using entire historic period, filtered | 58.1 | 19.4 | 14.0 | 14.0 | 4.3 | 32.3 | 19.4 | 14.0 |
| Unadjusted, using TaR after historic visit, filtered | 16.1 | 31.2 | 19.4 | 14.0 | 7.5 | 43.0 | 20.4 | 14.0 |
| Age & sex adjusted, using TaR after historic visit, filtered | 16.1 | 26.9 | 19.4 | 14.0 | 7.5 | 39.8 | 19.4 | 14.0 |
| SCCS / SCRI | | | | | | | | |
| Unadjusted SCCS excluding pre−vaccination window | 9.7 | 24.7 | 19.4 | 12.9 | 4.3 | 24.7 | 19.4 | 12.9 |
| Age & season adjusted SCCS excluding pre−vaccination window | 6.5 | 24.7 | 22.6 | 16.1 | 2.2 | 24.7 | 22.6 | 16.1 |
| SCRI with prior control interval | 15.1 | 31.2 | 20.4 | 12.9 | 1.1 | 41.9 | 22.6 | 15.1 |
| SCRI with posterior control interval | 15.1 | 24.7 | 19.4 | 12.9 | 3.2 | 36.6 | 19.4 | 12.9 |
| Unadjusted SCCS excluding all pre−vaccination time | 16.1 | 24.7 | 19.4 | 12.9 | 2.2 | 26.9 | 19.4 | 12.9 |

Figure 31: Type 1 and 2 error based on $p < 0.05$ at the end of the study period in the MDCD database for Seasonal flu vaccination (All).

| | Uncalibrated | | | | Calibrated | | | |
|---|---|---|---|---|---|---|---|---|
| | | Type 2 when true effect is | | | | Type 2 when true effect is | | |
| | Type 1 | 1.5 | 2 | 4 | Type 1 | 1.5 | 2 | 4 |
| Case−control | | | | | | | | |
| Age & sex matched controls | 5.4 | 48.4 | 37.6 | 32.3 | 2.2 | 49.5 | 39.8 | 32.3 |
| Age & sex adjusted, using random controls | 17.2 | 26.9 | 22.6 | 15.1 | 1.1 | 33.3 | 23.7 | 15.1 |
| Cohort method | | | | | | | | |
| Unadjusted, using outpatient visits as comparator | 0.0 | 80.6 | 50.5 | 21.5 | 3.2 | 54.8 | 30.1 | 19.4 |
| PS matching, using outpatient visits as comparator | 1.1 | 63.4 | 44.1 | 25.8 | 3.2 | 60.2 | 36.6 | 25.8 |
| Unadjusted, using random days as comparator | 14.0 | 45.2 | 29.0 | 22.6 | 4.3 | 51.6 | 32.3 | 23.7 |
| PS matching, using random days as comparator | 2.2 | 63.4 | 44.1 | 25.8 | 3.2 | 62.4 | 43.0 | 25.8 |
| PS stratification, using outpatient visits as comparator | 1.1 | 64.5 | 39.8 | 24.7 | 6.5 | 53.8 | 35.5 | 23.7 |
| PS stratification, using random days as comparator | 5.4 | 51.6 | 37.6 | 22.6 | 3.2 | 52.7 | 37.6 | 22.6 |
| PS weighting, using outpatient visits as comparator | 0.0 | 74.2 | 48.4 | 25.8 | 3.2 | 58.1 | 39.8 | 24.7 |
| PS weighting, using random days as comparator | 1.1 | 69.9 | 46.2 | 28.0 | 2.2 | 65.6 | 44.1 | 28.0 |
| Per−month PS matching, using outpatient visits as comparator | | | | | | | | |
| Per−month PS matching, using random days as comparator | | | | | | | | |
| Historical comparator | | | | | | | | |
| Unadjusted, using entire historic period | 25.8 | 26.9 | 19.4 | 14.0 | 2.2 | 55.9 | 24.7 | 15.1 |
| Age & sex adjusted, using entire historic period | 24.7 | 26.9 | 19.4 | 14.0 | 2.2 | 57.0 | 24.7 | 15.1 |
| Unadjusted, using TaR after historic visit | 10.8 | 31.2 | 21.5 | 14.0 | 2.2 | 59.1 | 26.9 | 15.1 |
| Age & sex adjusted, using TaR after historic visit | 10.8 | 31.2 | 22.6 | 15.1 | 2.2 | 58.1 | 28.0 | 15.1 |
| Unadjusted, using entire historic period, filtered | 23.7 | 29.0 | 21.5 | 16.1 | 2.2 | 31.2 | 24.7 | 17.2 |
| Age & sex adjusted, using entire historic period, filtered | 22.6 | 29.0 | 21.5 | 16.1 | 1.1 | 31.2 | 24.7 | 17.2 |
| Unadjusted, using TaR after historic visit, filtered | 8.6 | 33.3 | 23.7 | 16.1 | 2.2 | 37.6 | 24.7 | 17.2 |
| Age & sex adjusted, using TaR after historic visit, filtered | 8.6 | 33.3 | 24.7 | 17.2 | 3.2 | 35.5 | 24.7 | 17.2 |
| SCCS / SCRI | | | | | | | | |
| Unadjusted SCCS excluding pre−vaccination window | 8.6 | 35.5 | 23.7 | 17.2 | 2.2 | 36.6 | 23.7 | 17.2 |
| Age & season adjusted SCCS excluding pre−vaccination window | 6.5 | 38.7 | 32.3 | 30.1 | 2.2 | 41.9 | 32.3 | 30.1 |
| SCRI with prior control interval | 16.1 | 43.0 | 31.2 | 22.6 | 1.1 | 55.9 | 38.7 | 22.6 |
| SCRI with posterior control interval | 8.6 | 43.0 | 29.0 | 21.5 | 1.1 | 50.5 | 30.1 | 23.7 |
| Unadjusted SCCS excluding all pre−vaccination time | 15.1 | 34.4 | 21.5 | 18.3 | 3.2 | 45.2 | 25.8 | 19.4 |

Figure 32: Type 1 and 2 error based on $p < 0.05$ at the end of the study period in the MDCR database for Seasonal flu vaccination (All).

| | Uncalibrated | | | | Calibrated | | | |
|---|---|---|---|---|---|---|---|---|
| | | Type 2 when true effect is | | | | Type 2 when true effect is | | |
| | Type 1 | 1.5 | 2 | 4 | Type 1 | 1.5 | 2 | 4 |
| Case−control | | | | | | | | |
| Age & sex matched controls | 32.3 | 29.0 | 19.4 | 16.1 | 3.2 | 35.5 | 22.6 | 17.2 |
| Age & sex adjusted, using random controls | 36.6 | 25.8 | 17.2 | 12.9 | 2.2 | 31.2 | 18.3 | 12.9 |
| Cohort method | | | | | | | | |
| Unadjusted, using outpatient visits as comparator | 3.2 | 83.9 | 58.1 | 19.4 | 5.4 | 67.7 | 34.4 | 15.1 |
| PS matching, using outpatient visits as comparator | 3.2 | 47.3 | 25.8 | 14.0 | 4.3 | 38.7 | 24.7 | 14.0 |
| Unadjusted, using random days as comparator | 34.4 | 20.4 | 18.3 | 12.9 | 2.2 | 40.9 | 18.3 | 12.9 |
| PS matching, using random days as comparator | 7.5 | 39.8 | 23.7 | 15.1 | 2.2 | 39.8 | 24.7 | 15.1 |
| PS stratification, using outpatient visits as comparator | 1.1 | 51.6 | 28.0 | 12.9 | 3.2 | 37.6 | 20.4 | 12.9 |
| PS stratification, using random days as comparator | 10.8 | 34.4 | 20.4 | 12.9 | 4.3 | 34.4 | 20.4 | 12.9 |
| PS weighting, using outpatient visits as comparator | 0.0 | 64.5 | 37.6 | 18.3 | 4.3 | 51.6 | 28.0 | 17.2 |
| PS weighting, using random days as comparator | 2.2 | 49.5 | 32.3 | 16.1 | 3.2 | 49.5 | 32.3 | 16.1 |
| Per−month PS matching, using outpatient visits as comparator | | | | | | | | |
| Per−month PS matching, using random days as comparator | | | | | | | | |
| Historical comparator | | | | | | | | |
| Unadjusted, using entire historic period | 47.3 | 18.3 | 12.9 | 10.8 | 2.2 | 54.8 | 18.3 | 10.8 |
| Age & sex adjusted, using entire historic period | 44.1 | 17.2 | 12.9 | 10.8 | 2.2 | 48.4 | 17.2 | 10.8 |
| Unadjusted, using TaR after historic visit | 17.2 | 23.7 | 17.2 | 10.8 | 2.2 | 69.9 | 21.5 | 10.8 |
| Age & sex adjusted, using TaR after historic visit | 18.3 | 21.5 | 16.1 | 10.8 | 2.2 | 63.4 | 19.4 | 10.8 |
| Unadjusted, using entire historic period, filtered | 45.2 | 20.4 | 15.1 | 12.9 | 2.2 | 25.8 | 18.3 | 12.9 |
| Age & sex adjusted, using entire historic period, filtered | 41.9 | 19.4 | 15.1 | 12.9 | 4.3 | 23.7 | 17.2 | 12.9 |
| Unadjusted, using TaR after historic visit, filtered | 15.1 | 25.8 | 19.4 | 12.9 | 6.5 | 40.9 | 19.4 | 12.9 |
| Age & sex adjusted, using TaR after historic visit, filtered | 16.1 | 23.7 | 18.3 | 12.9 | 3.2 | 29.0 | 19.4 | 12.9 |
| SCCS / SCRI | | | | | | | | |
| Unadjusted SCCS excluding pre−vaccination window | 19.4 | 18.3 | 14.0 | 10.8 | 3.2 | 18.3 | 14.0 | 10.8 |
| Age & season adjusted SCCS excluding pre−vaccination window | 16.1 | 24.7 | 21.5 | 18.3 | 3.2 | 24.7 | 21.5 | 18.3 |
| SCRI with prior control interval | 31.2 | 22.6 | 16.1 | 11.8 | 1.1 | 36.6 | 20.4 | 11.8 |
| SCRI with posterior control interval | 21.5 | 19.4 | 15.1 | 11.8 | 3.2 | 25.8 | 15.1 | 12.9 |
| Unadjusted SCCS excluding all pre−vaccination time | 30.1 | 16.1 | 14.0 | 11.8 | 3.2 | 20.4 | 14.0 | 11.8 |

Figure 33: Type 1 and 2 error based on $p < 0.05$ at the end of the study period in the CCAE database for Seasonal flu vaccination (All).

#### 10.5 Type 1 and 2 error for Zoster vaccination (Shingrix)

| | Uncalibrated | | | | Calibrated | | | |
|---|---|---|---|---|---|---|---|---|
| | | Type 2 when true effect is | | | | Type 2 when true effect is | | |
| | Type 1 | 1.5 | 2 | 4 | Type 1 | 1.5 | 2 | 4 |
| Case−control | | | | | | | | |
| Age & sex matched controls | 24.7 | 61.3 | 48.4 | 39.8 | 4.3 | 73.1 | 63.4 | 41.9 |
| Age & sex adjusted, using random controls | 20.4 | 60.2 | 46.2 | 34.4 | 3.2 | 77.4 | 62.4 | 38.7 |
| Cohort method | | | | | | | | |
| Unadjusted, using outpatient visits as comparator | 1.1 | 68.8 | 51.6 | 20.4 | 1.1 | 67.7 | 45.2 | 19.4 |
| PS matching, using outpatient visits as comparator | 0.0 | 74.2 | 48.4 | 20.4 | 3.2 | 59.1 | 33.3 | 17.2 |
| Unadjusted, using random days as comparator | 35.5 | 43.0 | 32.3 | 21.5 | 3.2 | 77.4 | 52.7 | 26.9 |
| PS matching, using random days as comparator | | | | | | | | |
| PS stratification, using outpatient visits as comparator | 0.0 | 75.3 | 43.0 | 17.2 | 5.4 | 54.8 | 32.3 | 15.1 |
| PS stratification, using random days as comparator | | | | | | | | |
| PS weighting, using outpatient visits as comparator | 0.0 | 87.1 | 68.8 | 40.9 | 1.1 | 76.3 | 59.1 | 34.4 |
| PS weighting, using random days as comparator | | | | | | | | |
| Per−month PS matching, using outpatient visits as comparator | | | | | | | | |
| Per−month PS matching, using random days as comparator | | | | | | | | |
| Historical comparator | | | | | | | | |
| Unadjusted, using entire historic period | 55.9 | 24.7 | 20.4 | 11.8 | 3.2 | 78.5 | 48.4 | 20.4 |
| Age & sex adjusted, using entire historic period | 40.9 | 25.8 | 17.2 | 11.8 | 3.2 | 68.8 | 36.6 | 15.1 |
| Unadjusted, using TaR after historic visit | 29.0 | 40.9 | 29.0 | 15.1 | 3.2 | 73.1 | 43.0 | 20.4 |
| Age & sex adjusted, using TaR after historic visit | 20.4 | 48.4 | 28.0 | 15.1 | 5.4 | 65.6 | 39.8 | 15.1 |
| Unadjusted, using entire historic period, filtered | 54.8 | 25.8 | 21.5 | 12.9 | 3.2 | 79.6 | 48.4 | 21.5 |
| Age & sex adjusted, using entire historic period, filtered | 39.8 | 26.9 | 18.3 | 12.9 | 2.2 | 64.5 | 34.4 | 16.1 |
| Unadjusted, using TaR after historic visit, filtered | 28.0 | 41.9 | 30.1 | 16.1 | 2.2 | 69.9 | 43.0 | 21.5 |
| Age & sex adjusted, using TaR after historic visit, filtered | 19.4 | 49.5 | 29.0 | 16.1 | 5.4 | 65.6 | 37.6 | 16.1 |
| SCCS / SCRI | | | | | | | | |
| Unadjusted SCCS excluding pre−vaccination window | 11.8 | 38.7 | 22.6 | 12.9 | 3.2 | 52.7 | 24.7 | 14.0 |
| Age & season adjusted SCCS excluding pre−vaccination window | 8.6 | 41.9 | 22.6 | 14.0 | 5.4 | 50.5 | 23.7 | 14.0 |
| SCRI with prior control interval | 9.7 | 48.4 | 33.3 | 18.3 | 3.2 | 59.1 | 35.5 | 21.5 |
| SCRI with posterior control interval | 11.8 | 51.6 | 36.6 | 18.3 | 3.2 | 60.2 | 40.9 | 21.5 |
| Unadjusted SCCS excluding all pre−vaccination time | 15.1 | 39.8 | 26.9 | 12.9 | 2.2 | 49.5 | 30.1 | 16.1 |

Figure 34: Type 1 and 2 error based on $p < 0.05$ at the end of the study period in the Optum EHR database for Zoster vaccination (Shingrix).

| | Uncalibrated | | | | Calibrated | | | |
|---|---|---|---|---|---|---|---|---|
| | | Type 2 when true effect is | | | | Type 2 when true effect is | | |
| | Type 1 | 1.5 | 2 | 4 | Type 1 | 1.5 | 2 | 4 |
| Case−control | | | | | | | | |
| Age & sex matched controls | 3.2 | 86.0 | 77.4 | 68.8 | 1.1 | 91.4 | 86.0 | 71.0 |
| Age & sex adjusted, using random controls | 4.3 | 86.0 | 72.0 | 61.3 | 1.1 | 92.5 | 86.0 | 63.4 |
| Cohort method | | | | | | | | |
| Unadjusted, using outpatient visits as comparator | 1.1 | 87.1 | 79.6 | 61.3 | 0.0 | 87.1 | 79.6 | 61.3 |
| PS matching, using outpatient visits as comparator | 0.0 | 97.8 | 84.9 | 68.8 | 1.1 | 91.4 | 83.9 | 67.7 |
| Unadjusted, using random days as comparator | 7.5 | 86.0 | 82.8 | 72.0 | 2.2 | 93.5 | 89.2 | 75.3 |
| PS matching, using random days as comparator | 0.0 | 94.6 | 90.3 | 72.0 | 0.0 | 93.5 | 87.1 | 72.0 |
| PS stratification, using outpatient visits as comparator | 2.2 | 90.3 | 79.6 | 54.8 | 5.4 | 86.0 | 76.3 | 53.8 |
| PS stratification, using random days as comparator | 1.1 | 86.0 | 74.2 | 54.8 | 0.0 | 90.3 | 77.4 | 55.9 |
| PS weighting, using outpatient visits as comparator | 0.0 | 98.9 | 90.3 | 78.5 | 0.0 | 97.8 | 90.3 | 78.5 |
| PS weighting, using random days as comparator | 0.0 | 95.7 | 89.2 | 77.4 | 0.0 | 95.7 | 89.2 | 77.4 |
| Per−month PS matching, using outpatient visits as comparator | | | | | | | | |
| Per−month PS matching, using random days as comparator | | | | | | | | |
| Historical comparator | | | | | | | | |
| Unadjusted, using entire historic period | 25.8 | 55.9 | 50.5 | 48.4 | 1.1 | 89.2 | 79.6 | 51.6 |
| Age & sex adjusted, using entire historic period | 12.9 | 63.4 | 57.0 | 48.4 | 2.2 | 84.9 | 67.7 | 49.5 |
| Unadjusted, using TaR after historic visit | 21.5 | 66.7 | 51.6 | 48.4 | 1.1 | 88.2 | 75.3 | 50.5 |
| Age & sex adjusted, using TaR after historic visit | 2.2 | 76.3 | 65.6 | 50.5 | 1.1 | 86.0 | 66.7 | 50.5 |
| Unadjusted, using entire historic period, filtered | 24.7 | 57.0 | 51.6 | 49.5 | 1.1 | 88.2 | 74.2 | 52.7 |
| Age & sex adjusted, using entire historic period, filtered | 11.8 | 64.5 | 58.1 | 49.5 | 1.1 | 78.5 | 64.5 | 49.5 |
| Unadjusted, using TaR after historic visit, filtered | 20.4 | 67.7 | 52.7 | 49.5 | 0.0 | 86.0 | 73.1 | 49.5 |
| Age & sex adjusted, using TaR after historic visit, filtered | 1.1 | 77.4 | 66.7 | 51.6 | 2.2 | 77.4 | 66.7 | 51.6 |
| SCCS / SCRI | | | | | | | | |
| Unadjusted SCCS excluding pre−vaccination window | 1.1 | 74.2 | 66.7 | 49.5 | 1.1 | 75.3 | 66.7 | 49.5 |
| Age & season adjusted SCCS excluding pre−vaccination window | 2.2 | 76.3 | 66.7 | 49.5 | 1.1 | 76.3 | 66.7 | 49.5 |
| SCRI with prior control interval | 0.0 | 90.3 | 78.5 | 59.1 | 0.0 | 86.0 | 77.4 | 58.1 |
| SCRI with posterior control interval | 1.1 | 89.2 | 77.4 | 60.2 | 1.1 | 89.2 | 77.4 | 60.2 |
| Unadjusted SCCS excluding all pre−vaccination time | 1.1 | 80.6 | 65.6 | 52.7 | 0.0 | 84.9 | 68.8 | 52.7 |

Figure 35: Type 1 and 2 error based on $p < 0.05$ at the end of the study period in the MDCD database for Zoster vaccination (Shingrix).

| | Uncalibrated | | | | Calibrated | | | |
|---|---|---|---|---|---|---|---|---|
| | | Type 2 when true effect is | | | | Type 2 when true effect is | | |
| | Type 1 | 1.5 | 2 | 4 | Type 1 | 1.5 | 2 | 4 |
| Case−control | | | | | | | | |
| Age & sex matched controls | 7.5 | 78.5 | 67.7 | 57.0 | 1.1 | 80.6 | 71.0 | 57.0 |
| Age & sex adjusted, using random controls | 11.8 | 69.9 | 58.1 | 35.5 | 3.2 | 75.3 | 63.4 | 37.6 |
| Cohort method | | | | | | | | |
| Unadjusted, using outpatient visits as comparator | 2.2 | 88.2 | 77.4 | 50.5 | 2.2 | 83.9 | 72.0 | 41.9 |
| PS matching, using outpatient visits as comparator | 1.1 | 83.9 | 69.9 | 50.5 | 2.2 | 80.6 | 65.6 | 50.5 |
| Unadjusted, using random days as comparator | 4.3 | 75.3 | 63.4 | 39.8 | 1.1 | 77.4 | 64.5 | 39.8 |
| PS matching, using random days as comparator | 1.1 | 80.6 | 63.4 | 45.2 | 2.2 | 76.3 | 63.4 | 45.2 |
| PS stratification, using outpatient visits as comparator | 2.2 | 77.4 | 64.5 | 41.9 | 3.2 | 75.3 | 61.3 | 38.7 |
| PS stratification, using random days as comparator | 4.3 | 74.2 | 54.8 | 35.5 | 3.2 | 75.3 | 54.8 | 35.5 |
| PS weighting, using outpatient visits as comparator | 2.2 | 80.6 | 71.0 | 54.8 | 2.2 | 78.5 | 67.7 | 49.5 |
| PS weighting, using random days as comparator | 1.1 | 86.0 | 79.6 | 60.2 | 1.1 | 86.0 | 79.6 | 60.2 |
| Per−month PS matching, using outpatient visits as comparator | | | | | | | | |
| Per−month PS matching, using random days as comparator | | | | | | | | |
| Historical comparator | | | | | | | | |
| Unadjusted, using entire historic period | 12.9 | 65.6 | 55.9 | 34.4 | 3.2 | 83.9 | 67.7 | 37.6 |
| Age & sex adjusted, using entire historic period | 11.8 | 64.5 | 54.8 | 34.4 | 3.2 | 82.8 | 66.7 | 36.6 |
| Unadjusted, using TaR after historic visit | 10.8 | 71.0 | 61.3 | 37.6 | 5.4 | 87.1 | 71.0 | 39.8 |
| Age & sex adjusted, using TaR after historic visit | 10.8 | 71.0 | 61.3 | 37.6 | 5.4 | 87.1 | 69.9 | 38.7 |
| Unadjusted, using entire historic period, filtered | 9.7 | 68.8 | 59.1 | 37.6 | 4.3 | 78.5 | 65.6 | 38.7 |
| Age & sex adjusted, using entire historic period, filtered | 8.6 | 67.7 | 58.1 | 37.6 | 4.3 | 77.4 | 63.4 | 38.7 |
| Unadjusted, using TaR after historic visit, filtered | 7.5 | 74.2 | 64.5 | 40.9 | 2.2 | 83.9 | 71.0 | 40.9 |
| Age & sex adjusted, using TaR after historic visit, filtered | 7.5 | 74.2 | 64.5 | 40.9 | 2.2 | 84.9 | 68.8 | 40.9 |
| SCCS / SCRI | | | | | | | | |
| Unadjusted SCCS excluding pre−vaccination window | 10.8 | 64.5 | 50.5 | 35.5 | 4.3 | 69.9 | 54.8 | 36.6 |
| Age & season adjusted SCCS excluding pre−vaccination window | 9.7 | 65.6 | 51.6 | 39.8 | 3.2 | 69.9 | 53.8 | 40.9 |
| SCRI with prior control interval | 3.2 | 66.7 | 58.1 | 47.3 | 1.1 | 66.7 | 60.2 | 48.4 |
| SCRI with posterior control interval | 4.3 | 76.3 | 63.4 | 47.3 | 2.2 | 79.6 | 64.5 | 47.3 |
| Unadjusted SCCS excluding all pre−vaccination time | 6.5 | 67.7 | 51.6 | 37.6 | 2.2 | 72.0 | 57.0 | 37.6 |

Figure 36: Type 1 and 2 error based on $p < 0.05$ at the end of the study period in the MDCR database for Zoster vaccination (Shingrix).

| | Uncalibrated | | | | Calibrated | | | |
|---|---|---|---|---|---|---|---|---|
| | | Type 2 when true effect is | | | | Type 2 when true effect is | | |
| | Type 1 | 1.5 | 2 | 4 | Type 1 | 1.5 | 2 | 4 |
| Case−control | | | | | | | | |
| Age & sex matched controls | 6.5 | 73.1 | 61.3 | 37.6 | 1.1 | 77.4 | 66.7 | 43.0 |
| Age & sex adjusted, using random controls | 3.2 | 78.5 | 58.1 | 33.3 | 2.2 | 81.7 | 61.3 | 34.4 |
| Cohort method | | | | | | | | |
| Unadjusted, using outpatient visits as comparator | 2.2 | 82.8 | 62.4 | 33.3 | 2.2 | 73.1 | 52.7 | 28.0 |
| PS matching, using outpatient visits as comparator | 0.0 | 74.2 | 53.8 | 31.2 | 2.2 | 67.7 | 45.2 | 30.1 |
| Unadjusted, using random days as comparator | 19.4 | 59.1 | 48.4 | 33.3 | 1.1 | 82.8 | 61.3 | 41.9 |
| PS matching, using random days as comparator | 1.1 | 68.8 | 48.4 | 30.1 | 1.1 | 66.7 | 46.2 | 30.1 |
| PS stratification, using outpatient visits as comparator | 1.1 | 77.4 | 53.8 | 29.0 | 4.3 | 62.4 | 45.2 | 26.9 |
| PS stratification, using random days as comparator | 2.2 | 64.5 | 37.6 | 22.6 | 2.2 | 64.5 | 37.6 | 22.6 |
| PS weighting, using outpatient visits as comparator | 0.0 | 91.4 | 80.6 | 46.2 | 0.0 | 88.2 | 73.1 | 41.9 |
| PS weighting, using random days as comparator | 0.0 | 83.9 | 74.2 | 37.6 | 0.0 | 83.9 | 74.2 | 37.6 |
| Per−month PS matching, using outpatient visits as comparator | | | | | | | | |
| Per−month PS matching, using random days as comparator | | | | | | | | |
| Historical comparator | | | | | | | | |
| Unadjusted, using entire historic period | 32.3 | 40.9 | 34.4 | 20.4 | 3.2 | 78.5 | 50.5 | 25.8 |
| Age & sex adjusted, using entire historic period | 14.0 | 53.8 | 33.3 | 20.4 | 3.2 | 68.8 | 44.1 | 24.7 |
| Unadjusted, using TaR after historic visit | 19.4 | 52.7 | 37.6 | 20.4 | 3.2 | 75.3 | 51.6 | 24.7 |
| Age & sex adjusted, using TaR after historic visit | 7.5 | 63.4 | 47.3 | 23.7 | 5.4 | 69.9 | 50.5 | 23.7 |
| Unadjusted, using entire historic period, filtered | 30.1 | 43.0 | 36.6 | 22.6 | 1.1 | 72.0 | 50.5 | 24.7 |
| Age & sex adjusted, using entire historic period, filtered | 11.8 | 55.9 | 35.5 | 22.6 | 3.2 | 60.2 | 38.7 | 24.7 |
| Unadjusted, using TaR after historic visit, filtered | 17.2 | 54.8 | 39.8 | 22.6 | 1.1 | 71.0 | 51.6 | 24.7 |
| Age & sex adjusted, using TaR after historic visit, filtered | 5.4 | 65.6 | 49.5 | 25.8 | 5.4 | 65.6 | 46.2 | 24.7 |
| SCCS / SCRI | | | | | | | | |
| Unadjusted SCCS excluding pre−vaccination window | 8.6 | 54.8 | 33.3 | 20.4 | 3.2 | 59.1 | 35.5 | 21.5 |
| Age & season adjusted SCCS excluding pre−vaccination window | 6.5 | 57.0 | 35.5 | 22.6 | 2.2 | 62.4 | 35.5 | 23.7 |
| SCRI with prior control interval | 5.4 | 64.5 | 44.1 | 25.8 | 3.2 | 66.7 | 46.2 | 25.8 |
| SCRI with posterior control interval | 5.4 | 67.7 | 51.6 | 37.6 | 1.1 | 69.9 | 51.6 | 37.6 |
| Unadjusted SCCS excluding all pre−vaccination time | 5.4 | 53.8 | 38.7 | 22.6 | 2.2 | 61.3 | 44.1 | 23.7 |

Figure 37: Type 1 and 2 error based on $p < 0.05$ at the end of the study period in the CCAE database for Zoster vaccination (Shingrix).

#### 10.6 Type 1 and 2 error for HPV vaccination (Gardasil 9)

| | Uncalibrated | | | | Calibrated | | | |
|---|---|---|---|---|---|---|---|---|
| | | Type 2 when true effect is | | | | Type 2 when true effect is | | |
| | Type 1 | 1.5 | 2 | 4 | Type 1 | 1.5 | 2 | 4 |
| **Case−control** | | | | | | | | |
| Age & sex matched controls | 16.1 | 66.7 | 60.2 | 52.7 | 2.2 | 89.2 | 79.6 | 57.0 |
| Age & sex adjusted, using random controls | 6.5 | 78.5 | 66.7 | 57.0 | 3.2 | 93.5 | 80.6 | 61.3 |
| **Cohort method** | | | | | | | | |
| Unadjusted, using outpatient visits as comparator | 2.2 | 93.5 | 81.7 | 68.8 | 1.1 | 89.2 | 78.5 | 66.7 |
| PS matching, using outpatient visits as comparator | 1.1 | 91.4 | 72.0 | 55.9 | 2.2 | 80.6 | 66.7 | 54.8 |
| Unadjusted, using random days as comparator | 5.4 | 81.7 | 75.3 | 57.0 | 3.2 | 90.3 | 79.6 | 57.0 |
| PS matching, using random days as comparator | | | | | | | | |
| PS stratification, using outpatient visits as comparator | 1.1 | 89.2 | 66.7 | 41.9 | 2.2 | 80.6 | 58.1 | 41.9 |
| PS stratification, using random days as comparator | | | | | | | | |
| PS weighting, using outpatient visits as comparator | 1.1 | 94.6 | 86.0 | 64.5 | 1.1 | 93.5 | 83.9 | 64.5 |
| PS weighting, using random days as comparator | | | | | | | | |
| Per−month PS matching, using outpatient visits as comparator | | | | | | | | |
| Per−month PS matching, using random days as comparator | | | | | | | | |
| **Historical comparator** | | | | | | | | |
| Unadjusted, using entire historic period | 15.1 | 71.0 | 68.8 | 48.4 | 2.2 | 87.1 | 76.3 | 54.8 |
| Age & sex adjusted, using entire historic period | 24.7 | 51.6 | 45.2 | 37.6 | 4.3 | 82.8 | 61.3 | 38.7 |
| Unadjusted, using TaR after historic visit | 5.4 | 79.6 | 73.1 | 54.8 | 2.2 | 88.2 | 77.4 | 60.2 |
| Age & sex adjusted, using TaR after historic visit | 6.5 | 71.0 | 60.2 | 40.9 | 3.2 | 77.4 | 62.4 | 41.9 |
| Unadjusted, using entire historic period, filtered | 14.0 | 72.0 | 69.9 | 49.5 | 2.2 | 87.1 | 77.4 | 55.9 |
| Age & sex adjusted, using entire historic period, filtered | 23.7 | 52.7 | 46.2 | 38.7 | 5.4 | 76.3 | 55.9 | 39.8 |
| Unadjusted, using TaR after historic visit, filtered | 5.4 | 80.6 | 74.2 | 55.9 | 2.2 | 89.2 | 80.6 | 61.3 |
| Age & sex adjusted, using TaR after historic visit, filtered | 5.4 | 72.0 | 61.3 | 41.9 | 2.2 | 73.1 | 61.3 | 41.9 |
| **SCCS / SCRI** | | | | | | | | |
| Unadjusted SCCS excluding pre−vaccination window | 9.7 | 61.3 | 48.4 | 37.6 | 4.3 | 76.3 | 58.1 | 37.6 |
| Age & season adjusted SCCS excluding pre−vaccination window | 8.6 | 63.4 | 49.5 | 37.6 | 4.3 | 77.4 | 60.2 | 38.7 |
| SCRI with prior control interval | 2.2 | 74.2 | 67.7 | 58.1 | 1.1 | 80.6 | 71.0 | 60.2 |
| SCRI with posterior control interval | 5.4 | 74.2 | 65.6 | 52.7 | 2.2 | 81.7 | 68.8 | 59.1 |
| Unadjusted SCCS excluding all pre−vaccination time | 11.8 | 61.3 | 53.8 | 38.7 | 3.2 | 74.2 | 57.0 | 39.8 |

Figure 38: Type 1 and 2 error based on $p < 0.05$ at the end of the study period in the Optum EHR database for HPV vaccination (Gardasil 9).

| | Uncalibrated | | | | Calibrated | | | |
|---|---|---|---|---|---|---|---|---|
| | | Type 2 when true effect is | | | | Type 2 when true effect is | | |
| | Type 1 | 1.5 | 2 | 4 | Type 1 | 1.5 | 2 | 4 |
| Case−control | | | | | | | | |
| Age & sex matched controls | 10.8 | 67.7 | 66.7 | 62.4 | 2.2 | 82.8 | 67.7 | 63.4 |
| Age & sex adjusted, using random controls | 8.6 | 71.0 | 65.6 | 53.8 | 1.1 | 78.5 | 67.7 | 54.8 |
| Cohort method | | | | | | | | |
| Unadjusted, using outpatient visits as comparator | 2.2 | 91.4 | 84.9 | 77.4 | 0.0 | 91.4 | 86.0 | 71.0 |
| PS matching, using outpatient visits as comparator | 1.1 | 83.9 | 79.6 | 66.7 | 1.1 | 87.1 | 78.5 | 66.7 |
| Unadjusted, using random days as comparator | 4.3 | 82.8 | 79.6 | 64.5 | 1.1 | 92.5 | 80.6 | 68.8 |
| PS matching, using random days as comparator | 3.2 | 86.0 | 80.6 | 66.7 | 3.2 | 89.2 | 80.6 | 67.7 |
| PS stratification, using outpatient visits as comparator | 3.2 | 83.9 | 76.3 | 51.6 | 2.2 | 88.2 | 72.0 | 51.6 |
| PS stratification, using random days as comparator | 9.7 | 72.0 | 69.9 | 51.6 | 6.5 | 80.6 | 71.0 | 54.8 |
| PS weighting, using outpatient visits as comparator | 1.1 | 89.2 | 83.9 | 73.1 | 0.0 | 92.5 | 84.9 | 73.1 |
| PS weighting, using random days as comparator | 2.2 | 89.2 | 82.8 | 74.2 | 1.1 | 95.7 | 84.9 | 74.2 |
| Per−month PS matching, using outpatient visits as comparator | | | | | | | | |
| Per−month PS matching, using random days as comparator | | | | | | | | |
| Historical comparator | | | | | | | | |
| Unadjusted, using entire historic period | 6.5 | 80.6 | 74.2 | 58.1 | 2.2 | 93.5 | 80.6 | 63.4 |
| Age & sex adjusted, using entire historic period | 10.8 | 68.8 | 62.4 | 47.3 | 2.2 | 86.0 | 74.2 | 49.5 |
| Unadjusted, using TaR after historic visit | 4.3 | 84.9 | 79.6 | 66.7 | 2.2 | 91.4 | 82.8 | 65.6 |
| Age & sex adjusted, using TaR after historic visit | 7.5 | 73.1 | 64.5 | 50.5 | 3.2 | 89.2 | 72.0 | 52.7 |
| Unadjusted, using entire historic period, filtered | 5.4 | 82.8 | 76.3 | 60.2 | 2.2 | 92.5 | 81.7 | 63.4 |
| Age & sex adjusted, using entire historic period, filtered | 8.6 | 71.0 | 64.5 | 49.5 | 4.3 | 82.8 | 68.8 | 49.5 |
| Unadjusted, using TaR after historic visit, filtered | 3.2 | 86.0 | 80.6 | 68.8 | 2.2 | 92.5 | 82.8 | 66.7 |
| Age & sex adjusted, using TaR after historic visit, filtered | 5.4 | 75.3 | 66.7 | 52.7 | 3.2 | 84.9 | 71.0 | 53.8 |
| SCCS / SCRI | | | | | | | | |
| Unadjusted SCCS excluding pre−vaccination window | 6.5 | 69.9 | 61.3 | 47.3 | 3.2 | 77.4 | 63.4 | 47.3 |
| Age & season adjusted SCCS excluding pre−vaccination window | 5.4 | 71.0 | 62.4 | 48.4 | 3.2 | 77.4 | 63.4 | 48.4 |
| SCRI with prior control interval | 5.4 | 77.4 | 68.8 | 59.1 | 2.2 | 91.4 | 74.2 | 63.4 |
| SCRI with posterior control interval | 4.3 | 79.6 | 72.0 | 63.4 | 2.2 | 89.2 | 74.2 | 66.7 |
| Unadjusted SCCS excluding all pre−vaccination time | 6.5 | 68.8 | 60.2 | 52.7 | 2.2 | 83.9 | 67.7 | 53.8 |

Figure 39: Type 1 and 2 error based on $p < 0.05$ at the end of the study period in the MDCD database for HPV vaccination (Gardasil 9).

| | Uncalibrated | | | | Calibrated | | | |
|---|---|---|---|---|---|---|---|---|
| | | Type 2 when true effect is | | | | Type 2 when true effect is | | |
| | Type 1 | 1.5 | 2 | 4 | Type 1 | 1.5 | 2 | 4 |
| Case−control | | | | | | | | |
| Age & sex matched controls | | | | | | | | |
| Age & sex adjusted, using random controls | | | | | | | | |
| Cohort method | | | | | | | | |
| Unadjusted, using outpatient visits as comparator | 0.0 | 100.0 | 100.0 | 100.0 | 0.0 | 100.0 | 100.0 | 100.0 |
| PS matching, using outpatient visits as comparator | | | | | | | | |
| Unadjusted, using random days as comparator | | | | | | | | |
| PS matching, using random days as comparator | | | | | | | | |
| PS stratification, using outpatient visits as comparator | 0.0 | 100.0 | 100.0 | 100.0 | 0.0 | 100.0 | 100.0 | 100.0 |
| PS stratification, using random days as comparator | | | | | | | | |
| PS weighting, using outpatient visits as comparator | 0.0 | 100.0 | 100.0 | 100.0 | 0.0 | 100.0 | 100.0 | 100.0 |
| PS weighting, using random days as comparator | | | | | | | | |
| Per−month PS matching, using outpatient visits as comparator | | | | | | | | |
| Per−month PS matching, using random days as comparator | | | | | | | | |
| Historical comparator | | | | | | | | |
| Unadjusted, using entire historic period | 0.0 | 100.0 | 100.0 | 100.0 | 0.0 | 100.0 | 100.0 | 100.0 |
| Age & sex adjusted, using entire historic period | 0.0 | 100.0 | 100.0 | 100.0 | 0.0 | 100.0 | 100.0 | 100.0 |
| Unadjusted, using TaR after historic visit | 0.0 | 100.0 | 100.0 | 100.0 | 0.0 | 100.0 | 100.0 | 100.0 |
| Age & sex adjusted, using TaR after historic visit | 0.0 | 100.0 | 100.0 | 100.0 | 0.0 | 100.0 | 100.0 | 100.0 |
| Unadjusted, using entire historic period, filtered | 0.0 | 100.0 | 100.0 | 100.0 | 0.0 | 100.0 | 100.0 | 100.0 |
| Age & sex adjusted, using entire historic period, filtered | 0.0 | 100.0 | 100.0 | 100.0 | 0.0 | 100.0 | 100.0 | 100.0 |
| Unadjusted, using TaR after historic visit, filtered | 0.0 | 100.0 | 100.0 | 100.0 | 0.0 | 100.0 | 100.0 | 100.0 |
| Age & sex adjusted, using TaR after historic visit, filtered | 0.0 | 100.0 | 100.0 | 100.0 | 0.0 | 100.0 | 100.0 | 100.0 |

Figure 40: Type 1 and 2 error based on $p < 0.05$ at the end of the study period in the MDCR database for HPV vaccination (Gardasil 9).

| | Uncalibrated | | | | Calibrated | | | |
|---|---|---|---|---|---|---|---|---|
| | | Type 2 when true effect is | | | | Type 2 when true effect is | | |
| | Type 1 | 1.5 | 2 | 4 | Type 1 | 1.5 | 2 | 4 |
| Case−control | | | | | | | | |
| Age & sex matched controls | 12.9 | 66.7 | 57.0 | 50.5 | 3.2 | 79.6 | 65.6 | 51.6 |
| Age & sex adjusted, using random controls | 5.4 | 73.1 | 64.5 | 52.7 | 3.2 | 81.7 | 71.0 | 54.8 |
| Cohort method | | | | | | | | |
| Unadjusted, using outpatient visits as comparator | 2.2 | 89.2 | 84.9 | 68.8 | 2.2 | 89.2 | 78.5 | 63.4 |
| PS matching, using outpatient visits as comparator | 0.0 | 83.9 | 68.8 | 55.9 | 1.1 | 80.6 | 67.7 | 55.9 |
| Unadjusted, using random days as comparator | 4.3 | 83.9 | 77.4 | 55.9 | 2.2 | 89.2 | 78.5 | 54.8 |
| PS matching, using random days as comparator | 2.2 | 86.0 | 72.0 | 57.0 | 2.2 | 87.1 | 72.0 | 57.0 |
| PS stratification, using outpatient visits as comparator | 1.1 | 83.9 | 64.5 | 45.2 | 3.2 | 76.3 | 59.1 | 44.1 |
| PS stratification, using random days as comparator | 2.2 | 78.5 | 61.3 | 47.3 | 2.2 | 79.6 | 60.2 | 47.3 |
| PS weighting, using outpatient visits as comparator | 0.0 | 93.5 | 83.9 | 66.7 | 0.0 | 93.5 | 82.8 | 65.6 |
| PS weighting, using random days as comparator | 1.1 | 93.5 | 81.7 | 71.0 | 1.1 | 93.5 | 83.9 | 71.0 |
| Per−month PS matching, using outpatient visits as comparator | | | | | | | | |
| Per−month PS matching, using random days as comparator | | | | | | | | |
| Historical comparator | | | | | | | | |
| Unadjusted, using entire historic period | 7.5 | 78.5 | 67.7 | 50.5 | 1.1 | 89.2 | 75.3 | 51.6 |
| Age & sex adjusted, using entire historic period | 9.7 | 58.1 | 45.2 | 37.6 | 1.1 | 80.6 | 57.0 | 39.8 |
| Unadjusted, using TaR after historic visit | 8.6 | 81.7 | 76.3 | 58.1 | 1.1 | 87.1 | 78.5 | 61.3 |
| Age & sex adjusted, using TaR after historic visit | 4.3 | 74.2 | 52.7 | 40.9 | 3.2 | 81.7 | 59.1 | 40.9 |
| Unadjusted, using entire historic period, filtered | 7.5 | 79.6 | 69.9 | 52.7 | 1.1 | 89.2 | 77.4 | 53.8 |
| Age & sex adjusted, using entire historic period, filtered | 7.5 | 60.2 | 47.3 | 39.8 | 3.2 | 73.1 | 52.7 | 40.9 |
| Unadjusted, using TaR after historic visit, filtered | 8.6 | 82.8 | 78.5 | 60.2 | 1.1 | 87.1 | 79.6 | 63.4 |
| Age & sex adjusted, using TaR after historic visit, filtered | 3.2 | 76.3 | 54.8 | 43.0 | 3.2 | 81.7 | 59.1 | 43.0 |
| SCCS / SCRI | | | | | | | | |
| Unadjusted SCCS excluding pre−vaccination window | 10.8 | 59.1 | 48.4 | 38.7 | 1.1 | 71.0 | 53.8 | 39.8 |
| Age & season adjusted SCCS excluding pre−vaccination window | 10.8 | 59.1 | 48.4 | 38.7 | 2.2 | 71.0 | 53.8 | 39.8 |
| SCRI with prior control interval | 4.3 | 73.1 | 61.3 | 53.8 | 2.2 | 86.0 | 63.4 | 57.0 |
| SCRI with posterior control interval | 3.2 | 73.1 | 63.4 | 53.8 | 1.1 | 86.0 | 65.6 | 54.8 |
| Unadjusted SCCS excluding all pre−vaccination time | 9.7 | 60.2 | 49.5 | 40.9 | 2.2 | 77.4 | 58.1 | 40.9 |

Figure 41: Type 1 and 2 error based on $p < 0.05$ at the end of the study period in the CCAE database for HPV vaccination (Gardasil 9).

### 11 Type 1 and 2 error by true effect size based on MaxSPRT

For each method variation and vaccine group, the type 1 error and the type 2 error stratified by true effect size, based on whether the log likelihood ratio exceeded the critical value any time during the study period.

#### 11.1 Type 1 and 2 error for H1N1 vaccination

| | Uncalibrated | | | | Calibrated | | | |
|---|---|---|---|---|---|---|---|---|
| | | Type 2 when true effect is | | | | Type 2 when true effect is | | |
| | Type 1 | 1.5 | 2 | 4 | Type 1 | 1.5 | 2 | 4 |
| Case−control | | | | | | | | |
| Age & sex matched controls | 29.0 | 64.5 | 52.7 | 38.7 | 4.3 | 76.3 | 69.9 | 51.6 |
| Age & sex adjusted, using random controls | 29.0 | 59.1 | 53.8 | 37.6 | 4.3 | 82.8 | 73.1 | 49.5 |
| Cohort method | | | | | | | | |
| Unadjusted, using outpatient visits as comparator | 4.3 | 89.2 | 76.3 | 53.8 | 7.5 | 83.9 | 69.9 | 48.4 |
| PS matching, using outpatient visits as comparator | 0.0 | 90.3 | 73.1 | 50.5 | 3.2 | 77.4 | 65.6 | 48.4 |
| Unadjusted, using random days as comparator | 20.4 | 73.1 | 66.7 | 61.3 | 3.2 | 90.3 | 81.7 | 63.4 |
| PS matching, using random days as comparator | 1.1 | 96.8 | 95.7 | 93.5 | 0.0 | 98.9 | 95.7 | 93.5 |
| PS stratification, using outpatient visits as comparator | 0.0 | 81.7 | 63.4 | 39.8 | 4.3 | 73.1 | 50.5 | 36.6 |
| PS stratification, using random days as comparator | 35.5 | 49.5 | 41.9 | 36.6 | 0.0 | 78.5 | 58.1 | 39.8 |
| PS weighting, using outpatient visits as comparator | 5.4 | 78.5 | 61.3 | 46.2 | 5.4 | 77.4 | 62.4 | 44.1 |
| PS weighting, using random days as comparator | 0.0 | 98.9 | 97.8 | 93.5 | 0.0 | 100.0 | 97.8 | 94.6 |
| Per−month PS matching, using outpatient visits as comparator | 1.1 | 95.7 | 90.3 | 71.0 | 3.2 | 92.5 | 89.2 | 68.8 |
| Per−month PS matching, using random days as comparator | 1.1 | 94.6 | 89.2 | 84.9 | 0.0 | 96.8 | 90.3 | 84.9 |
| Historical comparator | | | | | | | | |
| Unadjusted, using entire historic period | 36.6 | 58.1 | 55.9 | 53.8 | 4.3 | 73.1 | 63.4 | 55.9 |
| Age & sex adjusted, using entire historic period | 33.3 | 61.3 | 58.1 | 55.9 | 4.3 | 67.7 | 63.4 | 58.1 |
| Unadjusted, using TaR after historic visit | 16.1 | 51.6 | 46.2 | 41.9 | 5.4 | 80.6 | 55.9 | 41.9 |
| Age & sex adjusted, using TaR after historic visit | 18.3 | 47.3 | 46.2 | 39.8 | 5.4 | 69.9 | 47.3 | 41.9 |
| Unadjusted, using entire historic period, filtered | 36.6 | 58.1 | 55.9 | 53.8 | 4.3 | 73.1 | 63.4 | 55.9 |
| Age & sex adjusted, using entire historic period, filtered | 33.3 | 61.3 | 58.1 | 55.9 | 4.3 | 68.8 | 62.4 | 58.1 |
| Unadjusted, using TaR after historic visit, filtered | 16.1 | 53.8 | 48.4 | 44.1 | 5.4 | 79.6 | 55.9 | 44.1 |
| Age & sex adjusted, using TaR after historic visit, filtered | 18.3 | 48.4 | 47.3 | 40.9 | 6.5 | 65.6 | 48.4 | 43.0 |
| SCCS / SCRI | | | | | | | | |
| Unadjusted SCCS excluding pre−vaccination window | 4.3 | 64.5 | 47.3 | 30.1 | 4.3 | 63.4 | 47.3 | 30.1 |
| Age & season adjusted SCCS excluding pre−vaccination window | 2.2 | 57.0 | 43.0 | 30.1 | 4.3 | 54.8 | 41.9 | 30.1 |
| SCRI with prior control interval | 3.2 | 78.5 | 64.5 | 40.9 | 0.0 | 79.6 | 65.6 | 40.9 |
| SCRI with posterior control interval | 4.3 | 78.5 | 61.3 | 44.1 | 3.2 | 82.8 | 61.3 | 44.1 |
| Unadjusted SCCS excluding all pre−vaccination time | 6.5 | 59.1 | 45.2 | 32.3 | 3.2 | 64.5 | 46.2 | 32.3 |

Figure 42: Type 1 and 2 error based on whether the log likelihood ratio exceeded the critical value any time during the study period in the Optum EHR database for H1N1 vaccination.

| | Uncalibrated | | | | Calibrated | | | |
|---|---|---|---|---|---|---|---|---|
| | | Type 2 when true effect is | | | | Type 2 when true effect is | | |
| | Type 1 | 1.5 | 2 | 4 | Type 1 | 1.5 | 2 | 4 |
| Case−control | | | | | | | | |
| Age & sex matched controls | 15.1 | 62.4 | 52.7 | 39.8 | 4.3 | 72.0 | 59.1 | 40.9 |
| Age & sex adjusted, using random controls | 9.7 | 63.4 | 54.8 | 36.6 | 2.2 | 73.1 | 57.0 | 37.6 |
| Cohort method | | | | | | | | |
| Unadjusted, using outpatient visits as comparator | 3.2 | 67.7 | 81.7 | 68.8 | 4.3 | 94.6 | 90.3 | 60.2 |
| PS matching, using outpatient visits as comparator | 0.0 | 89.2 | 76.3 | 60.2 | 4.3 | 86.0 | 74.2 | 58.1 |
| Unadjusted, using random days as comparator | 3.2 | 91.4 | 82.8 | 58.1 | 2.2 | 92.5 | 83.9 | 58.1 |
| PS matching, using random days as comparator | 3.2 | 84.9 | 72.0 | 57.0 | 4.3 | 84.9 | 72.0 | 57.0 |
| PS stratification, using outpatient visits as comparator | 4.3 | 76.3 | 68.8 | 43.0 | 6.5 | 75.3 | 61.3 | 41.9 |
| PS stratification, using random days as comparator | 4.3 | 77.4 | 64.5 | 48.4 | 3.2 | 77.4 | 65.6 | 48.4 |
| PS weighting, using outpatient visits as comparator | 6.5 | 51.6 | 57.0 | 52.7 | 7.5 | 77.4 | 63.4 | 48.4 |
| PS weighting, using random days as comparator | 0.0 | 92.5 | 86.0 | 69.9 | 0.0 | 91.4 | 86.0 | 68.8 |
| Per−month PS matching, using outpatient visits as comparator | 1.1 | 87.1 | 79.6 | 63.4 | 2.2 | 84.9 | 76.3 | 61.3 |
| Per−month PS matching, using random days as comparator | 1.1 | 88.2 | 78.5 | 65.6 | 1.1 | 87.1 | 78.5 | 65.6 |
| Historical comparator | | | | | | | | |
| Unadjusted, using entire historic period | 4.3 | 81.7 | 68.8 | 44.1 | 2.2 | 88.2 | 75.3 | 43.0 |
| Age & sex adjusted, using entire historic period | 7.5 | 65.6 | 57.0 | 48.4 | 0.0 | 76.3 | 62.4 | 49.5 |
| Unadjusted, using TaR after historic visit | 4.3 | 81.7 | 78.5 | 51.6 | 3.2 | 90.3 | 79.6 | 46.2 |
| Age & sex adjusted, using TaR after historic visit | 5.4 | 66.7 | 59.1 | 48.4 | 1.1 | 74.2 | 63.4 | 49.5 |
| Unadjusted, using entire historic period, filtered | 3.2 | 81.7 | 68.8 | 44.1 | 2.2 | 89.2 | 73.1 | 43.0 |
| Age & sex adjusted, using entire historic period, filtered | 7.5 | 65.6 | 57.0 | 48.4 | 0.0 | 76.3 | 63.4 | 49.5 |
| Unadjusted, using TaR after historic visit, filtered | 3.2 | 81.7 | 78.5 | 51.6 | 3.2 | 90.3 | 76.3 | 45.2 |
| Age & sex adjusted, using TaR after historic visit, filtered | 5.4 | 66.7 | 59.1 | 48.4 | 1.1 | 74.2 | 62.4 | 49.5 |
| SCCS / SCRI | | | | | | | | |
| Unadjusted SCCS excluding pre−vaccination window | 4.3 | 71.0 | 57.0 | 40.9 | 4.3 | 75.3 | 57.0 | 39.8 |
| Age & season adjusted SCCS excluding pre−vaccination window | 4.3 | 65.6 | 51.6 | 39.8 | 6.5 | 62.4 | 48.4 | 39.8 |
| SCRI with prior control interval | 4.3 | 82.8 | 67.7 | 49.5 | 3.2 | 86.0 | 67.7 | 49.5 |
| SCRI with posterior control interval | 6.5 | 87.1 | 66.7 | 49.5 | 4.3 | 87.1 | 66.7 | 49.5 |
| Unadjusted SCCS excluding all pre−vaccination time | 6.5 | 72.0 | 55.9 | 41.9 | 3.2 | 74.2 | 55.9 | 43.0 |

Figure 43: Type 1 and 2 error based on whether the log likelihood ratio exceeded the critical value any time during the study period in the MDCD database for H1N1 vaccination.

| | Uncalibrated | | | | Calibrated | | | |
|---|---|---|---|---|---|---|---|---|
| | | Type 2 when true effect is | | | | Type 2 when true effect is | | |
| | Type 1 | 1.5 | 2 | 4 | Type 1 | 1.5 | 2 | 4 |
| Case−control | | | | | | | | |
| Age & sex matched controls | 15.1 | 76.3 | 71.0 | 55.9 | 3.2 | 84.9 | 76.3 | 59.1 |
| Age & sex adjusted, using random controls | 11.8 | 78.5 | 74.2 | 55.9 | 5.4 | 87.1 | 76.3 | 57.0 |
| Cohort method | | | | | | | | |
| Unadjusted, using outpatient visits as comparator | 0.0 | 94.6 | 91.4 | 79.6 | 1.1 | 92.5 | 89.2 | 77.4 |
| PS matching, using outpatient visits as comparator | 1.1 | 95.7 | 95.7 | 86.0 | 1.1 | 95.7 | 93.5 | 86.0 |
| Unadjusted, using random days as comparator | 2.2 | 92.5 | 89.2 | 83.9 | 1.1 | 92.5 | 89.2 | 83.9 |
| PS matching, using random days as comparator | 0.0 | 95.7 | 93.5 | 87.1 | 0.0 | 95.7 | 92.5 | 87.1 |
| PS stratification, using outpatient visits as comparator | 0.0 | 82.8 | 75.3 | 65.6 | 4.3 | 82.8 | 74.2 | 64.5 |
| PS stratification, using random days as comparator | 5.4 | 80.6 | 78.5 | 68.8 | 2.2 | 83.9 | 79.6 | 72.0 |
| PS weighting, using outpatient visits as comparator | 7.5 | 87.1 | 82.8 | 71.0 | 2.2 | 95.7 | 87.1 | 78.5 |
| PS weighting, using random days as comparator | 0.0 | 98.9 | 97.8 | 91.4 | 0.0 | 100.0 | 97.8 | 91.4 |
| Per−month PS matching, using outpatient visits as comparator | 0.0 | 97.8 | 93.5 | 89.2 | 0.0 | 96.8 | 93.5 | 89.2 |
| Per−month PS matching, using random days as comparator | 0.0 | 96.8 | 95.7 | 90.3 | 0.0 | 96.8 | 95.7 | 90.3 |
| Historical comparator | | | | | | | | |
| Unadjusted, using entire historic period | 7.5 | 82.8 | 81.7 | 75.3 | 1.1 | 94.6 | 83.9 | 79.6 |
| Age & sex adjusted, using entire historic period | 8.6 | 82.8 | 79.6 | 75.3 | 1.1 | 93.5 | 82.8 | 78.5 |
| Unadjusted, using TaR after historic visit | 2.2 | 82.8 | 80.6 | 74.2 | 1.1 | 93.5 | 82.8 | 77.4 |
| Age & sex adjusted, using TaR after historic visit | 4.3 | 81.7 | 79.6 | 72.0 | 1.1 | 91.4 | 81.7 | 75.3 |
| Unadjusted, using entire historic period, filtered | 7.5 | 82.8 | 81.7 | 75.3 | 1.1 | 93.5 | 82.8 | 79.6 |
| Age & sex adjusted, using entire historic period, filtered | 8.6 | 82.8 | 79.6 | 75.3 | 1.1 | 93.5 | 82.8 | 78.5 |
| Unadjusted, using TaR after historic visit, filtered | 2.2 | 82.8 | 80.6 | 74.2 | 1.1 | 92.5 | 82.8 | 77.4 |
| Age & sex adjusted, using TaR after historic visit, filtered | 4.3 | 81.7 | 79.6 | 72.0 | 1.1 | 91.4 | 81.7 | 75.3 |
| SCCS / SCRI | | | | | | | | |
| Unadjusted SCCS excluding pre−vaccination window | 5.4 | 80.6 | 71.0 | 59.1 | 4.3 | 83.9 | 73.1 | 61.3 |
| Age & season adjusted SCCS excluding pre−vaccination window | 7.5 | 73.1 | 68.8 | 57.0 | 7.5 | 75.3 | 69.9 | 57.0 |
| SCRI with prior control interval | 2.2 | 92.5 | 84.9 | 71.0 | 0.0 | 95.7 | 88.2 | 72.0 |
| SCRI with posterior control interval | 0.0 | 95.7 | 88.2 | 74.2 | 1.1 | 95.7 | 86.0 | 74.2 |
| Unadjusted SCCS excluding all pre−vaccination time | 3.2 | 83.9 | 74.2 | 61.3 | 3.2 | 84.9 | 75.3 | 62.4 |

Figure 44: Type 1 and 2 error based on whether the log likelihood ratio exceeded the critical value any time during the study period in the MDCR database for H1N1 vaccination.

| | Uncalibrated | | | | Calibrated | | | |
|---|---|---|---|---|---|---|---|---|
| | | Type 2 when true effect is | | | | Type 2 when true effect is | | |
| | Type 1 | 1.5 | 2 | 4 | Type 1 | 1.5 | 2 | 4 |
| Case−control | | | | | | | | |
| Age & sex matched controls | 21.5 | 63.4 | 53.8 | 43.0 | 3.2 | 74.2 | 61.3 | 44.1 |
| Age & sex adjusted, using random controls | 19.4 | 59.1 | 53.8 | 38.7 | 2.2 | 72.0 | 58.1 | 40.9 |
| Cohort method | | | | | | | | |
| Unadjusted, using outpatient visits as comparator | 4.3 | 59.1 | 71.0 | 35.5 | 5.4 | 88.2 | 76.3 | 32.3 |
| PS matching, using outpatient visits as comparator | 0.0 | 73.1 | 45.2 | 29.0 | 5.4 | 60.2 | 33.3 | 29.0 |
| Unadjusted, using random days as comparator | 10.8 | 58.1 | 37.6 | 29.0 | 3.2 | 79.6 | 48.4 | 29.0 |
| PS matching, using random days as comparator | 5.4 | 59.1 | 40.9 | 25.8 | 5.4 | 60.2 | 40.9 | 25.8 |
| PS stratification, using outpatient visits as comparator | 1.1 | 69.9 | 41.9 | 25.8 | 4.3 | 57.0 | 33.3 | 24.7 |
| PS stratification, using random days as comparator | 6.5 | 51.6 | 37.6 | 23.7 | 5.4 | 57.0 | 40.9 | 23.7 |
| PS weighting, using outpatient visits as comparator | 6.5 | 66.7 | 47.3 | 26.9 | 2.2 | 68.8 | 43.0 | 26.9 |
| PS weighting, using random days as comparator | 1.1 | 83.9 | 68.8 | 37.6 | 2.2 | 78.5 | 66.7 | 36.6 |
| Per−month PS matching, using outpatient visits as comparator | 1.1 | 74.2 | 50.5 | 33.3 | 5.4 | 60.2 | 43.0 | 32.3 |
| Per−month PS matching, using random days as comparator | 4.3 | 64.5 | 49.5 | 32.3 | 4.3 | 64.5 | 49.5 | 32.3 |
| Historical comparator | | | | | | | | |
| Unadjusted, using entire historic period | 16.1 | 37.6 | 29.0 | 24.7 | 6.5 | 71.0 | 29.0 | 24.7 |
| Age & sex adjusted, using entire historic period | 29.0 | 32.3 | 29.0 | 25.8 | 5.4 | 52.7 | 33.3 | 25.8 |
| Unadjusted, using TaR after historic visit | 10.8 | 62.4 | 33.3 | 24.7 | 6.5 | 79.6 | 43.0 | 22.6 |
| Age & sex adjusted, using TaR after historic visit | 11.8 | 47.3 | 29.0 | 23.7 | 5.4 | 57.0 | 29.0 | 23.7 |
| Unadjusted, using entire historic period, filtered | 16.1 | 38.7 | 30.1 | 25.8 | 6.5 | 72.0 | 30.1 | 25.8 |
| Age & sex adjusted, using entire historic period, filtered | 28.0 | 33.3 | 30.1 | 26.9 | 4.3 | 52.7 | 34.4 | 26.9 |
| Unadjusted, using TaR after historic visit, filtered | 10.8 | 63.4 | 34.4 | 25.8 | 5.4 | 79.6 | 44.1 | 23.7 |
| Age & sex adjusted, using TaR after historic visit, filtered | 11.8 | 48.4 | 30.1 | 24.7 | 5.4 | 58.1 | 30.1 | 24.7 |
| SCCS / SCRI | | | | | | | | |
| Unadjusted SCCS excluding pre−vaccination window | 5.4 | 44.1 | 28.0 | 21.5 | 2.2 | 52.7 | 28.0 | 21.5 |
| Age & season adjusted SCCS excluding pre−vaccination window | 6.5 | 38.7 | 26.9 | 21.5 | 5.4 | 39.8 | 28.0 | 20.4 |
| SCRI with prior control interval | 6.5 | 62.4 | 40.9 | 26.9 | 2.2 | 67.7 | 46.2 | 26.9 |
| SCRI with posterior control interval | 10.8 | 57.0 | 35.5 | 26.9 | 5.4 | 60.2 | 37.6 | 26.9 |
| Unadjusted SCCS excluding all pre−vaccination time | 12.9 | 46.2 | 29.0 | 21.5 | 4.3 | 51.6 | 31.2 | 21.5 |

Figure 45: Type 1 and 2 error based on whether the log likelihood ratio exceeded the critical value any time during the study period in the CCAE database for H1N1 vaccination.

#### 11.2 Type 1 and 2 error for Seasonal flu vaccination (Fluvirin)

| | Uncalibrated | | | | Calibrated | | | |
|---|---|---|---|---|---|---|---|---|
| | | Type 2 when true effect is | | | | Type 2 when true effect is | | |
| | Type 1 | 1.5 | 2 | 4 | Type 1 | 1.5 | 2 | 4 |
| Case−control | | | | | | | | |
| Age & sex matched controls | 5.4 | 93.5 | 87.1 | 76.3 | 1.1 | 100.0 | 95.7 | 80.6 |
| Age & sex adjusted, using random controls | 4.3 | 93.5 | 91.4 | 75.3 | 0.0 | 98.9 | 97.8 | 87.1 |
| Cohort method | | | | | | | | |
| Unadjusted, using outpatient visits as comparator | 1.1 | 96.8 | 95.7 | 79.6 | 3.2 | 95.7 | 87.1 | 79.6 |
| PS matching, using outpatient visits as comparator | 0.0 | 100.0 | 98.9 | 92.5 | 1.1 | 98.9 | 95.7 | 92.5 |
| Unadjusted, using random days as comparator | 1.1 | 95.7 | 88.2 | 86.0 | 0.0 | 97.8 | 95.7 | 86.0 |
| PS matching, using random days as comparator | | | | | | | | |
| PS stratification, using outpatient visits as comparator | 2.2 | 95.7 | 92.5 | 79.6 | 2.2 | 93.5 | 89.2 | 73.1 |
| PS stratification, using random days as comparator | 9.7 | 80.6 | 73.1 | 60.2 | 2.2 | 88.2 | 79.6 | 68.8 |
| PS weighting, using outpatient visits as comparator | 0.0 | 98.9 | 94.6 | 83.9 | 0.0 | 97.8 | 92.5 | 81.7 |
| PS weighting, using random days as comparator | 0.0 | 100.0 | 100.0 | 100.0 | 0.0 | 100.0 | 100.0 | 100.0 |
| Per−month PS matching, using outpatient visits as comparator | | | | | | | | |
| Per−month PS matching, using random days as comparator | | | | | | | | |
| Historical comparator | | | | | | | | |
| Unadjusted, using entire historic period | 5.4 | 87.1 | 87.1 | 87.1 | 0.0 | 96.8 | 90.3 | 88.2 |
| Age & sex adjusted, using entire historic period | 8.6 | 84.9 | 84.9 | 84.9 | 0.0 | 96.8 | 89.2 | 86.0 |
| Unadjusted, using TaR after historic visit | 6.5 | 82.8 | 76.3 | 73.1 | 1.1 | 93.5 | 83.9 | 75.3 |
| Age & sex adjusted, using TaR after historic visit | 6.5 | 82.8 | 75.3 | 72.0 | 1.1 | 90.3 | 83.9 | 73.1 |
| Unadjusted, using entire historic period, filtered | 5.4 | 87.1 | 87.1 | 87.1 | 0.0 | 96.8 | 90.3 | 88.2 |
| Age & sex adjusted, using entire historic period, filtered | 8.6 | 84.9 | 84.9 | 84.9 | 0.0 | 94.6 | 89.2 | 86.0 |
| Unadjusted, using TaR after historic visit, filtered | 6.5 | 82.8 | 76.3 | 73.1 | 1.1 | 93.5 | 83.9 | 75.3 |
| Age & sex adjusted, using TaR after historic visit, filtered | 6.5 | 82.8 | 75.3 | 72.0 | 1.1 | 91.4 | 82.8 | 73.1 |
| SCCS / SCRI | | | | | | | | |
| Unadjusted SCCS excluding pre−vaccination window | 3.2 | 84.9 | 82.8 | 64.5 | 3.2 | 91.4 | 83.9 | 64.5 |
| Age & season adjusted SCCS excluding pre−vaccination window | 2.2 | 84.9 | 78.5 | 65.6 | 3.2 | 89.2 | 81.7 | 64.5 |
| SCRI with prior control interval | 3.2 | 97.8 | 90.3 | 76.3 | 1.1 | 97.8 | 93.5 | 76.3 |
| SCRI with posterior control interval | 2.2 | 97.8 | 88.2 | 78.5 | 1.1 | 97.8 | 92.5 | 78.5 |
| Unadjusted SCCS excluding all pre−vaccination time | 2.2 | 87.1 | 75.3 | 63.4 | 1.1 | 92.5 | 81.7 | 64.5 |

Figure 46: Type 1 and 2 error based on whether the log likelihood ratio exceeded the critical value any time during the study period in the Optum EHR database for Seasonal flu vaccination (Fluvirin).

| | Uncalibrated | | | | Calibrated | | | |
|---|---|---|---|---|---|---|---|---|
| | | Type 2 when true effect is | | | | Type 2 when true effect is | | |
| | Type 1 | 1.5 | 2 | 4 | Type 1 | 1.5 | 2 | 4 |
| Case−control | | | | | | | | |
| Age & sex matched controls | 8.6 | 83.9 | 80.6 | 66.7 | 6.5 | 90.3 | 83.9 | 73.1 |
| Age & sex adjusted, using random controls | 1.1 | 86.0 | 82.8 | 65.6 | 1.1 | 91.4 | 83.9 | 69.9 |
| Cohort method | | | | | | | | |
| Unadjusted, using outpatient visits as comparator | 0.0 | 94.6 | 88.2 | 78.5 | 1.1 | 93.5 | 87.1 | 77.4 |
| PS matching, using outpatient visits as comparator | 0.0 | 98.9 | 97.8 | 93.5 | 1.1 | 97.8 | 93.5 | 91.4 |
| Unadjusted, using random days as comparator | 6.5 | 91.4 | 88.2 | 83.9 | 2.2 | 96.8 | 91.4 | 84.9 |
| PS matching, using random days as comparator | 0.0 | 95.7 | 91.4 | 86.0 | 1.1 | 92.5 | 89.2 | 84.9 |
| PS stratification, using outpatient visits as comparator | 0.0 | 86.0 | 79.6 | 67.7 | 3.2 | 86.0 | 77.4 | 67.7 |
| PS stratification, using random days as comparator | 5.4 | 81.7 | 76.3 | 68.8 | 1.1 | 87.1 | 77.4 | 69.9 |
| PS weighting, using outpatient visits as comparator | 0.0 | 97.8 | 95.7 | 80.6 | 0.0 | 97.8 | 94.6 | 80.6 |
| PS weighting, using random days as comparator | 0.0 | 100.0 | 96.8 | 88.2 | 0.0 | 100.0 | 96.8 | 88.2 |
| Per−month PS matching, using outpatient visits as comparator | | | | | | | | |
| Per−month PS matching, using random days as comparator | | | | | | | | |
| Historical comparator | | | | | | | | |
| Unadjusted, using entire historic period | 7.5 | 86.0 | 82.8 | 79.6 | 0.0 | 96.8 | 91.4 | 81.7 |
| Age & sex adjusted, using entire historic period | 4.3 | 83.9 | 78.5 | 74.2 | 1.1 | 90.3 | 84.9 | 76.3 |
| Unadjusted, using TaR after historic visit | 4.3 | 81.7 | 79.6 | 75.3 | 0.0 | 94.6 | 84.9 | 78.5 |
| Age & sex adjusted, using TaR after historic visit | 3.2 | 91.4 | 80.6 | 73.1 | 2.2 | 93.5 | 90.3 | 74.2 |
| Unadjusted, using entire historic period, filtered | 7.5 | 86.0 | 82.8 | 79.6 | 2.2 | 92.5 | 87.1 | 80.6 |
| Age & sex adjusted, using entire historic period, filtered | 4.3 | 83.9 | 78.5 | 74.2 | 2.2 | 87.1 | 80.6 | 74.2 |
| Unadjusted, using TaR after historic visit, filtered | 4.3 | 81.7 | 79.6 | 75.3 | 2.2 | 90.3 | 80.6 | 76.3 |
| Age & sex adjusted, using TaR after historic visit, filtered | 3.2 | 91.4 | 80.6 | 73.1 | 3.2 | 91.4 | 82.8 | 73.1 |
| SCCS / SCRI | | | | | | | | |
| Unadjusted SCCS excluding pre−vaccination window | 1.1 | 88.2 | 79.6 | 65.6 | 1.1 | 88.2 | 80.6 | 65.6 |
| Age & season adjusted SCCS excluding pre−vaccination window | 1.1 | 88.2 | 80.6 | 64.5 | 1.1 | 88.2 | 80.6 | 64.5 |
| SCRI with prior control interval | 2.2 | 96.8 | 91.4 | 81.7 | 0.0 | 93.5 | 90.3 | 80.6 |
| SCRI with posterior control interval | 1.1 | 93.5 | 87.1 | 75.3 | 0.0 | 93.5 | 88.2 | 75.3 |
| Unadjusted SCCS excluding all pre−vaccination time | 0.0 | 89.2 | 79.6 | 66.7 | 0.0 | 90.3 | 79.6 | 66.7 |

Figure 47: Type 1 and 2 error based on whether the log likelihood ratio exceeded the critical value any time during the study period in the MDCD database for Seasonal flu vaccination (Fluvirin).

| | Uncalibrated | | | | Calibrated | | | |
|---|---|---|---|---|---|---|---|---|
| | | Type 2 when true effect is | | | | Type 2 when true effect is | | |
| | Type 1 | 1.5 | 2 | 4 | Type 1 | 1.5 | 2 | 4 |
| Case−control | | | | | | | | |
| Age & sex matched controls | 0.0 | 98.9 | 98.9 | 94.6 | 0.0 | 100.0 | 100.0 | 100.0 |
| Age & sex adjusted, using random controls | 0.0 | 100.0 | 100.0 | 92.5 | 0.0 | 100.0 | 100.0 | 97.8 |
| Cohort method | | | | | | | | |
| Unadjusted, using outpatient visits as comparator | 0.0 | 100.0 | 100.0 | 98.9 | 0.0 | 100.0 | 100.0 | 100.0 |
| PS matching, using outpatient visits as comparator | 0.0 | 100.0 | 100.0 | 100.0 | 0.0 | 100.0 | 100.0 | 100.0 |
| Unadjusted, using random days as comparator | 0.0 | 100.0 | 100.0 | 100.0 | 0.0 | 100.0 | 100.0 | 100.0 |
| PS matching, using random days as comparator | 0.0 | 100.0 | 100.0 | 100.0 | 0.0 | 100.0 | 100.0 | 100.0 |
| PS stratification, using outpatient visits as comparator | 0.0 | 98.9 | 96.8 | 94.6 | 0.0 | 100.0 | 98.9 | 96.8 |
| PS stratification, using random days as comparator | 0.0 | 98.9 | 98.9 | 96.8 | 0.0 | 100.0 | 100.0 | 100.0 |
| PS weighting, using outpatient visits as comparator | 1.1 | 98.9 | 98.9 | 96.8 | 1.1 | 98.9 | 98.9 | 96.8 |
| PS weighting, using random days as comparator | 0.0 | 100.0 | 100.0 | 100.0 | 0.0 | 100.0 | 100.0 | 100.0 |
| Per−month PS matching, using outpatient visits as comparator | | | | | | | | |
| Per−month PS matching, using random days as comparator | | | | | | | | |
| Historical comparator | | | | | | | | |
| Unadjusted, using entire historic period | 0.0 | 98.9 | 98.9 | 97.8 | 0.0 | 100.0 | 98.9 | 98.9 |
| Age & sex adjusted, using entire historic period | 0.0 | 98.9 | 98.9 | 97.8 | 0.0 | 100.0 | 98.9 | 98.9 |
| Unadjusted, using TaR after historic visit | 0.0 | 98.9 | 98.9 | 97.8 | 0.0 | 100.0 | 98.9 | 97.8 |
| Age & sex adjusted, using TaR after historic visit | 0.0 | 98.9 | 98.9 | 97.8 | 0.0 | 100.0 | 98.9 | 98.9 |
| Unadjusted, using entire historic period, filtered | 0.0 | 98.9 | 98.9 | 97.8 | 0.0 | 100.0 | 98.9 | 98.9 |
| Age & sex adjusted, using entire historic period, filtered | 0.0 | 98.9 | 98.9 | 97.8 | 0.0 | 100.0 | 98.9 | 98.9 |
| Unadjusted, using TaR after historic visit, filtered | 0.0 | 98.9 | 98.9 | 97.8 | 0.0 | 100.0 | 98.9 | 97.8 |
| Age & sex adjusted, using TaR after historic visit, filtered | 0.0 | 98.9 | 98.9 | 97.8 | 0.0 | 100.0 | 98.9 | 98.9 |
| SCCS / SCRI | | | | | | | | |
| Unadjusted SCCS excluding pre−vaccination window | 0.0 | 97.8 | 97.8 | 95.7 | 0.0 | 100.0 | 100.0 | 100.0 |
| Age & season adjusted SCCS excluding pre−vaccination window | 0.0 | 100.0 | 98.9 | 96.8 | 0.0 | 100.0 | 100.0 | 100.0 |
| SCRI with prior control interval | 0.0 | 100.0 | 100.0 | 98.9 | 0.0 | 100.0 | 100.0 | 100.0 |
| SCRI with posterior control interval | 0.0 | 100.0 | 100.0 | 98.9 | 0.0 | 100.0 | 100.0 | 100.0 |
| Unadjusted SCCS excluding all pre−vaccination time | 0.0 | 98.9 | 97.8 | 96.8 | 0.0 | 100.0 | 100.0 | 100.0 |

Figure 48: Type 1 and 2 error based on whether the log likelihood ratio exceeded the critical value any time during the study period in the MDCR database for Seasonal flu vaccination (Fluvirin).

| | Uncalibrated | | | | Calibrated | | | |
|---|---|---|---|---|---|---|---|---|
| | | Type 2 when true effect is | | | | Type 2 when true effect is | | |
| | Type 1 | 1.5 | 2 | 4 | Type 1 | 1.5 | 2 | 4 |
| Case−control | | | | | | | | |
| Age & sex matched controls | 6.5 | 82.8 | 72.0 | 55.9 | 2.2 | 83.9 | 75.3 | 55.9 |
| Age & sex adjusted, using random controls | 4.3 | 78.5 | 71.0 | 52.7 | 3.2 | 82.8 | 75.3 | 52.7 |
| Cohort method | | | | | | | | |
| Unadjusted, using outpatient visits as comparator | 0.0 | 87.1 | 79.6 | 58.1 | 4.3 | 80.6 | 69.9 | 48.4 |
| PS matching, using outpatient visits as comparator | 3.2 | 83.9 | 74.2 | 53.8 | 5.4 | 81.7 | 73.1 | 53.8 |
| Unadjusted, using random days as comparator | 11.8 | 77.4 | 73.1 | 60.2 | 3.2 | 83.9 | 76.3 | 61.3 |
| PS matching, using random days as comparator | 2.2 | 77.4 | 67.7 | 49.5 | 2.2 | 79.6 | 68.8 | 49.5 |
| PS stratification, using outpatient visits as comparator | 2.2 | 74.2 | 63.4 | 47.3 | 4.3 | 73.1 | 58.1 | 45.2 |
| PS stratification, using random days as comparator | 5.4 | 68.8 | 59.1 | 35.5 | 3.2 | 71.0 | 60.2 | 36.6 |
| PS weighting, using outpatient visits as comparator | 3.2 | 78.5 | 66.7 | 53.8 | 3.2 | 76.3 | 63.4 | 52.7 |
| PS weighting, using random days as comparator | 0.0 | 89.2 | 83.9 | 67.7 | 0.0 | 90.3 | 84.9 | 68.8 |
| Per−month PS matching, using outpatient visits as comparator | | | | | | | | |
| Per−month PS matching, using random days as comparator | | | | | | | | |
| Historical comparator | | | | | | | | |
| Unadjusted, using entire historic period | 16.1 | 62.4 | 54.8 | 46.2 | 2.2 | 91.4 | 74.2 | 48.4 |
| Age & sex adjusted, using entire historic period | 12.9 | 61.3 | 50.5 | 40.9 | 2.2 | 87.1 | 68.8 | 41.9 |
| Unadjusted, using TaR after historic visit | 8.6 | 67.7 | 53.8 | 40.9 | 1.1 | 89.2 | 75.3 | 45.2 |
| Age & sex adjusted, using TaR after historic visit | 8.6 | 66.7 | 53.8 | 37.6 | 4.3 | 84.9 | 71.0 | 43.0 |
| Unadjusted, using entire historic period, filtered | 15.1 | 63.4 | 55.9 | 47.3 | 3.2 | 73.1 | 61.3 | 47.3 |
| Age & sex adjusted, using entire historic period, filtered | 10.8 | 63.4 | 52.7 | 43.0 | 1.1 | 68.8 | 57.0 | 43.0 |
| Unadjusted, using TaR after historic visit, filtered | 7.5 | 68.8 | 54.8 | 41.9 | 3.2 | 72.0 | 54.8 | 41.9 |
| Age & sex adjusted, using TaR after historic visit, filtered | 6.5 | 68.8 | 55.9 | 39.8 | 5.4 | 69.9 | 54.8 | 40.9 |
| SCCS / SCRI | | | | | | | | |
| Unadjusted SCCS excluding pre−vaccination window | 6.5 | 64.5 | 55.9 | 38.7 | 4.3 | 67.7 | 58.1 | 39.8 |
| Age & season adjusted SCCS excluding pre−vaccination window | 9.7 | 63.4 | 51.6 | 35.5 | 4.3 | 65.6 | 57.0 | 35.5 |
| SCRI with prior control interval | 4.3 | 83.9 | 68.8 | 54.8 | 2.2 | 87.1 | 74.2 | 54.8 |
| SCRI with posterior control interval | 7.5 | 78.5 | 72.0 | 55.9 | 2.2 | 82.8 | 72.0 | 57.0 |
| Unadjusted SCCS excluding all pre−vaccination time | 8.6 | 65.6 | 58.1 | 39.8 | 2.2 | 72.0 | 61.3 | 39.8 |

Figure 49: Type 1 and 2 error based on whether the log likelihood ratio exceeded the critical value any time during the study period in the CCAE database for Seasonal flu vaccination (Fluvirin).

#### 11.3 Type 1 and 2 error for Seasonal flu vaccination (Fluzone)

| | Uncalibrated | | | | Calibrated | | | |
|---|---|---|---|---|---|---|---|---|
| | | Type 2 when true effect is | | | | Type 2 when true effect is | | |
| | Type 1 | 1.5 | 2 | 4 | Type 1 | 1.5 | 2 | 4 |
| Case−control | | | | | | | | |
| Age & sex matched controls | 30.1 | 49.5 | 37.6 | 26.9 | 8.6 | 72.0 | 53.8 | 32.3 |
| Age & sex adjusted, using random controls | 34.4 | 43.0 | 38.7 | 22.6 | 5.4 | 68.8 | 48.4 | 25.8 |
| Cohort method | | | | | | | | |
| Unadjusted, using outpatient visits as comparator | 3.2 | 66.7 | 49.5 | 23.7 | 5.4 | 68.8 | 41.9 | 21.5 |
| PS matching, using outpatient visits as comparator | 2.2 | 73.1 | 47.3 | 20.4 | 10.8 | 58.1 | 37.6 | 20.4 |
| Unadjusted, using random days as comparator | 34.4 | 43.0 | 32.3 | 23.7 | 4.3 | 73.1 | 47.3 | 25.8 |
| PS matching, using random days as comparator | | | | | | | | |
| PS stratification, using outpatient visits as comparator | 1.1 | 69.9 | 39.8 | 14.0 | 8.6 | 53.8 | 34.4 | 10.8 |
| PS stratification, using random days as comparator | 36.6 | 26.9 | 20.4 | 11.8 | 4.3 | 62.4 | 34.4 | 12.9 |
| PS weighting, using outpatient visits as comparator | 1.1 | 71.0 | 54.8 | 25.8 | 3.2 | 60.2 | 41.9 | 21.5 |
| PS weighting, using random days as comparator | 0.0 | 100.0 | 100.0 | 100.0 | 0.0 | 100.0 | 100.0 | 100.0 |
| Per−month PS matching, using outpatient visits as comparator | | | | | | | | |
| Per−month PS matching, using random days as comparator | | | | | | | | |
| Historical comparator | | | | | | | | |
| Unadjusted, using entire historic period | 58.1 | 24.7 | 19.4 | 18.3 | 3.2 | 68.8 | 41.9 | 18.3 |
| Age & sex adjusted, using entire historic period | 54.8 | 28.0 | 21.5 | 18.3 | 1.1 | 68.8 | 43.0 | 20.4 |
| Unadjusted, using TaR after historic visit | 25.8 | 34.4 | 24.7 | 15.1 | 2.2 | 72.0 | 43.0 | 15.1 |
| Age & sex adjusted, using TaR after historic visit | 23.7 | 35.5 | 24.7 | 14.0 | 3.2 | 76.3 | 35.5 | 15.1 |
| Unadjusted, using entire historic period, filtered | 58.1 | 24.7 | 20.4 | 18.3 | 4.3 | 54.8 | 31.2 | 18.3 |
| Age & sex adjusted, using entire historic period, filtered | 53.8 | 28.0 | 22.6 | 18.3 | 6.5 | 58.1 | 31.2 | 19.4 |
| Unadjusted, using TaR after historic visit, filtered | 23.7 | 34.4 | 25.8 | 15.1 | 2.2 | 55.9 | 31.2 | 15.1 |
| Age & sex adjusted, using TaR after historic visit, filtered | 22.6 | 35.5 | 25.8 | 14.0 | 6.5 | 54.8 | 26.9 | 14.0 |
| SCCS / SCRI | | | | | | | | |
| Unadjusted SCCS excluding pre−vaccination window | 17.2 | 32.3 | 23.7 | 10.8 | 4.3 | 49.5 | 26.9 | 9.7 |
| Age & season adjusted SCCS excluding pre−vaccination window | 23.7 | 32.3 | 21.5 | 9.7 | 6.5 | 47.3 | 28.0 | 9.7 |
| SCRI with prior control interval | 28.0 | 45.2 | 30.1 | 20.4 | 3.2 | 78.5 | 51.6 | 22.6 |
| SCRI with posterior control interval | 18.3 | 48.4 | 33.3 | 19.4 | 7.5 | 62.4 | 44.1 | 20.4 |
| Unadjusted SCCS excluding all pre−vaccination time | 22.6 | 34.4 | 22.6 | 9.7 | 8.6 | 53.8 | 28.0 | 12.9 |

Figure 50: Type 1 and 2 error based on whether the log likelihood ratio exceeded the critical value any time during the study period in the Optum EHR database for Seasonal flu vaccination (Fluzone).

| | Uncalibrated | | | | Calibrated | | | |
|---|---|---|---|---|---|---|---|---|
| | | Type 2 when true effect is | | | | Type 2 when true effect is | | |
| | Type 1 | 1.5 | 2 | 4 | Type 1 | 1.5 | 2 | 4 |
| Case−control | | | | | | | | |
| Age & sex matched controls | 3.2 | 96.8 | 91.4 | 82.8 | 0.0 | 98.9 | 96.8 | 86.0 |
| Age & sex adjusted, using random controls | 3.2 | 96.8 | 92.5 | 82.8 | 0.0 | 98.9 | 96.8 | 90.3 |
| Cohort method | | | | | | | | |
| Unadjusted, using outpatient visits as comparator | 0.0 | 98.9 | 98.9 | 97.8 | 0.0 | 98.9 | 98.9 | 97.8 |
| PS matching, using outpatient visits as comparator | 0.0 | 100.0 | 98.9 | 98.9 | 0.0 | 100.0 | 98.9 | 98.9 |
| Unadjusted, using random days as comparator | 1.1 | 97.8 | 97.8 | 96.8 | 1.1 | 98.9 | 97.8 | 97.8 |
| PS matching, using random days as comparator | | | | | | | | |
| PS stratification, using outpatient visits as comparator | 1.1 | 92.5 | 90.3 | 87.1 | 1.1 | 94.6 | 90.3 | 88.2 |
| PS stratification, using random days as comparator | 4.3 | 90.3 | 87.1 | 80.6 | 0.0 | 98.9 | 91.4 | 82.8 |
| PS weighting, using outpatient visits as comparator | 0.0 | 97.8 | 95.7 | 92.5 | 0.0 | 97.8 | 96.8 | 92.5 |
| PS weighting, using random days as comparator | 0.0 | 100.0 | 100.0 | 100.0 | 0.0 | 100.0 | 100.0 | 100.0 |
| Per−month PS matching, using outpatient visits as comparator | | | | | | | | |
| Per−month PS matching, using random days as comparator | | | | | | | | |
| Historical comparator | | | | | | | | |
| Unadjusted, using entire historic period | 2.2 | 96.8 | 94.6 | 94.6 | 0.0 | 100.0 | 100.0 | 96.8 |
| Age & sex adjusted, using entire historic period | 1.1 | 94.6 | 93.5 | 92.5 | 0.0 | 98.9 | 96.8 | 93.5 |
| Unadjusted, using TaR after historic visit | 1.1 | 96.8 | 94.6 | 94.6 | 0.0 | 100.0 | 100.0 | 96.8 |
| Age & sex adjusted, using TaR after historic visit | 1.1 | 95.7 | 91.4 | 89.2 | 0.0 | 97.8 | 96.8 | 90.3 |
| Unadjusted, using entire historic period, filtered | 2.2 | 96.8 | 94.6 | 94.6 | 0.0 | 100.0 | 98.9 | 96.8 |
| Age & sex adjusted, using entire historic period, filtered | 1.1 | 94.6 | 93.5 | 92.5 | 0.0 | 98.9 | 96.8 | 93.5 |
| Unadjusted, using TaR after historic visit, filtered | 1.1 | 96.8 | 94.6 | 94.6 | 0.0 | 100.0 | 98.9 | 95.7 |
| Age & sex adjusted, using TaR after historic visit, filtered | 1.1 | 95.7 | 91.4 | 89.2 | 0.0 | 97.8 | 96.8 | 90.3 |
| SCCS / SCRI | | | | | | | | |
| Unadjusted SCCS excluding pre−vaccination window | 2.2 | 93.5 | 89.2 | 81.7 | 2.2 | 94.6 | 92.5 | 82.8 |
| Age & season adjusted SCCS excluding pre−vaccination window | 2.2 | 95.7 | 89.2 | 81.7 | 2.2 | 96.8 | 92.5 | 83.9 |
| SCRI with prior control interval | 1.1 | 98.9 | 98.9 | 91.4 | 1.1 | 98.9 | 97.8 | 91.4 |
| SCRI with posterior control interval | 0.0 | 98.9 | 97.8 | 90.3 | 0.0 | 98.9 | 97.8 | 90.3 |
| Unadjusted SCCS excluding all pre−vaccination time | 2.2 | 94.6 | 90.3 | 80.6 | 1.1 | 95.7 | 90.3 | 81.7 |

Figure 51: Type 1 and 2 error based on whether the log likelihood ratio exceeded the critical value any time during the study period in the MDCD database for Seasonal flu vaccination (Fluzone).

| | Uncalibrated | | | | Calibrated | | | |
|---|---|---|---|---|---|---|---|---|
| | | Type 2 when true effect is | | | | Type 2 when true effect is | | |
| | Type 1 | 1.5 | 2 | 4 | Type 1 | 1.5 | 2 | 4 |
| Case−control | | | | | | | | |
| Age & sex matched controls | 2.2 | 81.7 | 72.0 | 45.2 | 3.2 | 80.6 | 69.9 | 45.2 |
| Age & sex adjusted, using random controls | 4.3 | 82.8 | 66.7 | 39.8 | 4.3 | 80.6 | 66.7 | 39.8 |
| Cohort method | | | | | | | | |
| Unadjusted, using outpatient visits as comparator | 0.0 | 92.5 | 89.2 | 72.0 | 2.2 | 88.2 | 83.9 | 62.4 |
| PS matching, using outpatient visits as comparator | 0.0 | 92.5 | 84.9 | 71.0 | 1.1 | 89.2 | 82.8 | 69.9 |
| Unadjusted, using random days as comparator | 1.1 | 91.4 | 84.9 | 69.9 | 1.1 | 89.2 | 83.9 | 68.8 |
| PS matching, using random days as comparator | 1.1 | 91.4 | 86.0 | 64.5 | 1.1 | 89.2 | 82.8 | 64.5 |
| PS stratification, using outpatient visits as comparator | 0.0 | 87.1 | 80.6 | 53.8 | 2.2 | 81.7 | 68.8 | 46.2 |
| PS stratification, using random days as comparator | 1.1 | 89.2 | 77.4 | 50.5 | 1.1 | 88.2 | 73.1 | 49.5 |
| PS weighting, using outpatient visits as comparator | 1.1 | 90.3 | 81.7 | 64.5 | 2.2 | 81.7 | 76.3 | 61.3 |
| PS weighting, using random days as comparator | 0.0 | 95.7 | 91.4 | 82.8 | 1.1 | 96.8 | 91.4 | 82.8 |
| Per−month PS matching, using outpatient visits as comparator | | | | | | | | |
| Per−month PS matching, using random days as comparator | | | | | | | | |
| Historical comparator | | | | | | | | |
| Unadjusted, using entire historic period | 4.3 | 81.7 | 65.6 | 53.8 | 3.2 | 92.5 | 81.7 | 54.8 |
| Age & sex adjusted, using entire historic period | 4.3 | 81.7 | 66.7 | 54.8 | 3.2 | 92.5 | 80.6 | 54.8 |
| Unadjusted, using TaR after historic visit | 3.2 | 81.7 | 73.1 | 51.6 | 3.2 | 89.2 | 81.7 | 51.6 |
| Age & sex adjusted, using TaR after historic visit | 3.2 | 81.7 | 71.0 | 52.7 | 3.2 | 90.3 | 77.4 | 53.8 |
| Unadjusted, using entire historic period, filtered | 3.2 | 82.8 | 66.7 | 54.8 | 3.2 | 82.8 | 68.8 | 54.8 |
| Age & sex adjusted, using entire historic period, filtered | 3.2 | 82.8 | 67.7 | 55.9 | 3.2 | 82.8 | 68.8 | 54.8 |
| Unadjusted, using TaR after historic visit, filtered | 2.2 | 83.9 | 74.2 | 52.7 | 5.4 | 82.8 | 74.2 | 52.7 |
| Age & sex adjusted, using TaR after historic visit, filtered | 2.2 | 83.9 | 72.0 | 53.8 | 5.4 | 82.8 | 71.0 | 53.8 |
| SCCS / SCRI | | | | | | | | |
| Unadjusted SCCS excluding pre−vaccination window | 2.2 | 82.8 | 72.0 | 52.7 | 2.2 | 82.8 | 71.0 | 51.6 |
| Age & season adjusted SCCS excluding pre−vaccination window | 2.2 | 78.5 | 69.9 | 50.5 | 2.2 | 78.5 | 69.9 | 50.5 |
| SCRI with prior control interval | 4.3 | 91.4 | 87.1 | 71.0 | 1.1 | 91.4 | 87.1 | 71.0 |
| SCRI with posterior control interval | 3.2 | 89.2 | 86.0 | 69.9 | 1.1 | 89.2 | 86.0 | 68.8 |
| Unadjusted SCCS excluding all pre−vaccination time | 3.2 | 83.9 | 77.4 | 54.8 | 2.2 | 83.9 | 76.3 | 54.8 |

Figure 52: Type 1 and 2 error based on whether the log likelihood ratio exceeded the critical value any time during the study period in the MDCR database for Seasonal flu vaccination (Fluzone).

| | Uncalibrated | | | | Calibrated | | | |
|---|---|---|---|---|---|---|---|---|
| | | Type 2 when true effect is | | | | Type 2 when true effect is | | |
| | Type 1 | 1.5 | 2 | 4 | Type 1 | 1.5 | 2 | 4 |
| Case−control | | | | | | | | |
| Age & sex matched controls | 0.0 | 100.0 | 100.0 | 98.9 | 0.0 | 100.0 | 100.0 | 100.0 |
| Age & sex adjusted, using random controls | 0.0 | 100.0 | 100.0 | 98.9 | 0.0 | 100.0 | 100.0 | 100.0 |
| Cohort method | | | | | | | | |
| Unadjusted, using outpatient visits as comparator | 0.0 | 100.0 | 100.0 | 100.0 | 0.0 | 100.0 | 100.0 | 100.0 |
| PS matching, using outpatient visits as comparator | 0.0 | 100.0 | 100.0 | 100.0 | 0.0 | 100.0 | 100.0 | 100.0 |
| Unadjusted, using random days as comparator | 0.0 | 100.0 | 100.0 | 100.0 | 0.0 | 100.0 | 100.0 | 100.0 |
| PS matching, using random days as comparator | 0.0 | 100.0 | 100.0 | 100.0 | 0.0 | 100.0 | 100.0 | 100.0 |
| PS stratification, using outpatient visits as comparator | 0.0 | 100.0 | 98.9 | 95.7 | 0.0 | 100.0 | 100.0 | 96.8 |
| PS stratification, using random days as comparator | 0.0 | 98.9 | 98.9 | 97.8 | 0.0 | 100.0 | 100.0 | 100.0 |
| PS weighting, using outpatient visits as comparator | 0.0 | 100.0 | 100.0 | 96.8 | 0.0 | 100.0 | 100.0 | 100.0 |
| PS weighting, using random days as comparator | 0.0 | 100.0 | 100.0 | 100.0 | 0.0 | 100.0 | 100.0 | 100.0 |
| Per−month PS matching, using outpatient visits as comparator | | | | | | | | |
| Per−month PS matching, using random days as comparator | | | | | | | | |
| Historical comparator | | | | | | | | |
| Unadjusted, using entire historic period | 0.0 | 100.0 | 100.0 | 100.0 | 0.0 | 100.0 | 100.0 | 100.0 |
| Age & sex adjusted, using entire historic period | 0.0 | 100.0 | 100.0 | 98.9 | 0.0 | 100.0 | 100.0 | 100.0 |
| Unadjusted, using TaR after historic visit | 0.0 | 100.0 | 100.0 | 100.0 | 0.0 | 100.0 | 100.0 | 100.0 |
| Age & sex adjusted, using TaR after historic visit | 0.0 | 100.0 | 100.0 | 98.9 | 0.0 | 100.0 | 100.0 | 100.0 |
| Unadjusted, using entire historic period, filtered | 0.0 | 100.0 | 100.0 | 100.0 | 0.0 | 100.0 | 100.0 | 100.0 |
| Age & sex adjusted, using entire historic period, filtered | 0.0 | 100.0 | 100.0 | 98.9 | 0.0 | 100.0 | 100.0 | 100.0 |
| Unadjusted, using TaR after historic visit, filtered | 0.0 | 100.0 | 100.0 | 100.0 | 0.0 | 100.0 | 100.0 | 100.0 |
| Age & sex adjusted, using TaR after historic visit, filtered | 0.0 | 100.0 | 100.0 | 98.9 | 0.0 | 100.0 | 100.0 | 100.0 |
| SCCS / SCRI | | | | | | | | |
| Unadjusted SCCS excluding pre−vaccination window | 0.0 | 100.0 | 100.0 | 96.8 | 0.0 | 100.0 | 100.0 | 100.0 |
| Age & season adjusted SCCS excluding pre−vaccination window | 0.0 | 100.0 | 100.0 | 97.8 | 0.0 | 100.0 | 100.0 | 100.0 |
| SCRI with prior control interval | | | | | | | | |
| SCRI with posterior control interval | | | | | | | | |
| Unadjusted SCCS excluding all pre−vaccination time | 0.0 | 100.0 | 100.0 | 96.8 | 0.0 | 100.0 | 100.0 | 100.0 |

Figure 53: Type 1 and 2 error based on whether the log likelihood ratio exceeded the critical value any time during the study period in the CCAE database for Seasonal flu vaccination (Fluzone).

#### 11.4 Type 1 and 2 error for Seasonal flu vaccination (All)

| | Uncalibrated | | | | Calibrated | | | |
|---|---|---|---|---|---|---|---|---|
| | | Type 2 when true effect is | | | | Type 2 when true effect is | | |
| | Type 1 | 1.5 | 2 | 4 | Type 1 | 1.5 | 2 | 4 |
| Case−control | | | | | | | | |
| Age & sex matched controls | 78.5 | 10.8 | 7.5 | 4.3 | 5.4 | 24.7 | 15.1 | 7.5 |
| Age & sex adjusted, using random controls | 81.7 | 7.5 | 5.4 | 2.2 | 7.5 | 20.4 | 11.8 | 4.3 |
| Cohort method | | | | | | | | |
| Unadjusted, using outpatient visits as comparator | 2.2 | 44.1 | 22.6 | 3.2 | 4.3 | 44.1 | 16.1 | 2.2 |
| PS matching, using outpatient visits as comparator | 1.1 | 40.9 | 12.9 | 5.4 | 4.3 | 24.7 | 10.8 | 5.4 |
| Unadjusted, using random days as comparator | 67.7 | 11.8 | 7.5 | 6.5 | 5.4 | 51.6 | 15.1 | 6.5 |
| PS matching, using random days as comparator | | | | | | | | |
| PS stratification, using outpatient visits as comparator | 1.1 | 43.0 | 11.8 | 4.3 | 4.3 | 23.7 | 8.6 | 4.3 |
| PS stratification, using random days as comparator | 79.6 | 4.3 | 4.3 | 2.2 | 1.1 | 38.7 | 9.7 | 4.3 |
| PS weighting, using outpatient visits as comparator | 3.2 | 39.8 | 15.1 | 5.4 | 6.5 | 24.7 | 12.9 | 4.3 |
| PS weighting, using random days as comparator | 0.0 | 100.0 | 100.0 | 100.0 | 0.0 | 100.0 | 100.0 | 100.0 |
| Per−month PS matching, using outpatient visits as comparator | | | | | | | | |
| Per−month PS matching, using random days as comparator | | | | | | | | |
| Historical comparator | | | | | | | | |
| Unadjusted, using entire historic period | 91.4 | 2.2 | 1.1 | 1.1 | 1.1 | 43.0 | 8.6 | 1.1 |
| Age & sex adjusted, using entire historic period | 92.5 | 2.2 | 1.1 | 1.1 | 3.2 | 25.8 | 5.4 | 1.1 |
| Unadjusted, using TaR after historic visit | 59.1 | 9.7 | 3.2 | 1.1 | 3.2 | 44.1 | 11.8 | 1.1 |
| Age & sex adjusted, using TaR after historic visit | 60.2 | 4.3 | 3.2 | 1.1 | 4.3 | 30.1 | 5.4 | 1.1 |
| Unadjusted, using entire historic period, filtered | 90.3 | 2.2 | 1.1 | 1.1 | 2.2 | 31.2 | 3.2 | 1.1 |
| Age & sex adjusted, using entire historic period, filtered | 91.4 | 2.2 | 1.1 | 1.1 | 3.2 | 18.3 | 3.2 | 1.1 |
| Unadjusted, using TaR after historic visit, filtered | 58.1 | 10.8 | 3.2 | 1.1 | 3.2 | 34.4 | 5.4 | 1.1 |
| Age & sex adjusted, using TaR after historic visit, filtered | 59.1 | 5.4 | 3.2 | 1.1 | 5.4 | 19.4 | 3.2 | 1.1 |
| SCCS / SCRI | | | | | | | | |
| Unadjusted SCCS excluding pre−vaccination window | 34.4 | 7.5 | 4.3 | 1.1 | 4.3 | 16.1 | 5.4 | 1.1 |
| Age & season adjusted SCCS excluding pre−vaccination window | 45.2 | 7.5 | 2.2 | 1.1 | 3.2 | 24.7 | 5.4 | 1.1 |
| SCRI with prior control interval | 53.8 | 10.8 | 7.5 | 2.2 | 6.5 | 30.1 | 10.8 | 3.2 |
| SCRI with posterior control interval | 43.0 | 10.8 | 4.3 | 2.2 | 6.5 | 29.0 | 6.5 | 2.2 |
| Unadjusted SCCS excluding all pre−vaccination time | 49.5 | 5.4 | 3.2 | 0.0 | 6.5 | 12.9 | 4.3 | 1.1 |

Figure 54: Type 1 and 2 error based on whether the log likelihood ratio exceeded the critical value any time during the study period in the Optum EHR database for Seasonal flu vaccination (All).

| | Uncalibrated | | | | Calibrated | | | |
|---|---|---|---|---|---|---|---|---|
| | | Type 2 when true effect is | | | | Type 2 when true effect is | | |
| | Type 1 | 1.5 | 2 | 4 | Type 1 | 1.5 | 2 | 4 |
| Case−control | | | | | | | | |
| Age & sex matched controls | 54.8 | 23.7 | 21.5 | 17.2 | 5.4 | 40.9 | 23.7 | 17.2 |
| Age & sex adjusted, using random controls | 63.4 | 17.2 | 15.1 | 12.9 | 6.5 | 34.4 | 18.3 | 12.9 |
| Cohort method | | | | | | | | |
| Unadjusted, using outpatient visits as comparator | 7.5 | 64.5 | 35.5 | 17.2 | 8.6 | 69.9 | 30.1 | 15.1 |
| PS matching, using outpatient visits as comparator | 3.2 | 47.3 | 23.7 | 17.2 | 8.6 | 37.6 | 23.7 | 17.2 |
| Unadjusted, using random days as comparator | 47.3 | 23.7 | 19.4 | 17.2 | 8.6 | 51.6 | 24.7 | 17.2 |
| PS matching, using random days as comparator | 3.2 | 47.3 | 29.0 | 16.1 | 4.3 | 45.2 | 24.7 | 16.1 |
| PS stratification, using outpatient visits as comparator | 3.2 | 39.8 | 22.6 | 12.9 | 7.5 | 34.4 | 21.5 | 12.9 |
| PS stratification, using random days as comparator | 8.6 | 43.0 | 23.7 | 12.9 | 3.2 | 46.2 | 24.7 | 12.9 |
| PS weighting, using outpatient visits as comparator | 4.3 | 37.6 | 20.4 | 12.9 | 3.2 | 40.9 | 18.3 | 12.9 |
| PS weighting, using random days as comparator | 8.6 | 59.1 | 34.4 | 18.3 | 8.6 | 49.5 | 31.2 | 17.2 |
| Per−month PS matching, using outpatient visits as comparator | | | | | | | | |
| Per−month PS matching, using random days as comparator | | | | | | | | |
| Historical comparator | | | | | | | | |
| Unadjusted, using entire historic period | 53.8 | 17.2 | 12.9 | 12.9 | 4.3 | 78.5 | 23.7 | 12.9 |
| Age & sex adjusted, using entire historic period | 60.2 | 17.2 | 12.9 | 12.9 | 3.2 | 72.0 | 22.6 | 12.9 |
| Unadjusted, using TaR after historic visit | 28.0 | 25.8 | 17.2 | 12.9 | 4.3 | 78.5 | 30.1 | 12.9 |
| Age & sex adjusted, using TaR after historic visit | 22.6 | 24.7 | 17.2 | 12.9 | 4.3 | 76.3 | 29.0 | 12.9 |
| Unadjusted, using entire historic period, filtered | 52.7 | 18.3 | 14.0 | 12.9 | 4.3 | 40.9 | 18.3 | 14.0 |
| Age & sex adjusted, using entire historic period, filtered | 59.1 | 18.3 | 14.0 | 12.9 | 3.2 | 34.4 | 18.3 | 14.0 |
| Unadjusted, using TaR after historic visit, filtered | 26.9 | 26.9 | 18.3 | 12.9 | 6.5 | 49.5 | 21.5 | 14.0 |
| Age & sex adjusted, using TaR after historic visit, filtered | 21.5 | 25.8 | 18.3 | 14.0 | 7.5 | 39.8 | 20.4 | 14.0 |
| SCCS / SCRI | | | | | | | | |
| Unadjusted SCCS excluding pre−vaccination window | 7.5 | 23.7 | 19.4 | 12.9 | 6.5 | 24.7 | 19.4 | 12.9 |
| Age & season adjusted SCCS excluding pre−vaccination window | 6.5 | 23.7 | 18.3 | 12.9 | 3.2 | 23.7 | 18.3 | 12.9 |
| SCRI with prior control interval | 12.9 | 36.6 | 22.6 | 15.1 | 4.3 | 54.8 | 26.9 | 15.1 |
| SCRI with posterior control interval | 20.4 | 31.2 | 19.4 | 12.9 | 3.2 | 47.3 | 21.5 | 12.9 |
| Unadjusted SCCS excluding all pre−vaccination time | 19.4 | 22.6 | 18.3 | 12.9 | 3.2 | 32.3 | 19.4 | 12.9 |

Figure 55: Type 1 and 2 error based on whether the log likelihood ratio exceeded the critical value any time during the study period in the MDCD database for Seasonal flu vaccination (All).

| | Uncalibrated | | | | Calibrated | | | |
|---|---|---|---|---|---|---|---|---|
| | | Type 2 when true effect is | | | | Type 2 when true effect is | | |
| | Type 1 | 1.5 | 2 | 4 | Type 1 | 1.5 | 2 | 4 |
| Case−control | | | | | | | | |
| Age & sex matched controls | 22.6 | 29.0 | 24.7 | 16.1 | 6.5 | 30.1 | 24.7 | 16.1 |
| Age & sex adjusted, using random controls | 24.7 | 25.8 | 19.4 | 14.0 | 11.8 | 28.0 | 21.5 | 14.0 |
| Cohort method | | | | | | | | |
| Unadjusted, using outpatient visits as comparator | 1.1 | 76.3 | 53.8 | 23.7 | 3.2 | 64.5 | 39.8 | 23.7 |
| PS matching, using outpatient visits as comparator | 2.2 | 58.1 | 44.1 | 25.8 | 7.5 | 53.8 | 37.6 | 25.8 |
| Unadjusted, using random days as comparator | 12.9 | 48.4 | 32.3 | 25.8 | 5.4 | 54.8 | 38.7 | 26.9 |
| PS matching, using random days as comparator | 2.2 | 57.0 | 39.8 | 28.0 | 6.5 | 55.9 | 38.7 | 28.0 |
| PS stratification, using outpatient visits as comparator | 5.4 | 58.1 | 39.8 | 23.7 | 7.5 | 49.5 | 35.5 | 21.5 |
| PS stratification, using random days as comparator | 7.5 | 52.7 | 34.4 | 22.6 | 6.5 | 52.7 | 34.4 | 22.6 |
| PS weighting, using outpatient visits as comparator | 1.1 | 65.6 | 48.4 | 23.7 | 6.5 | 58.1 | 40.9 | 22.6 |
| PS weighting, using random days as comparator | 3.2 | 73.1 | 48.4 | 30.1 | 3.2 | 67.7 | 44.1 | 28.0 |
| Per−month PS matching, using outpatient visits as comparator | | | | | | | | |
| Per−month PS matching, using random days as comparator | | | | | | | | |
| Historical comparator | | | | | | | | |
| Unadjusted, using entire historic period | 30.1 | 25.8 | 19.4 | 17.2 | 7.5 | 57.0 | 23.7 | 17.2 |
| Age & sex adjusted, using entire historic period | 29.0 | 25.8 | 20.4 | 18.3 | 6.5 | 58.1 | 24.7 | 18.3 |
| Unadjusted, using TaR after historic visit | 14.0 | 33.3 | 22.6 | 20.4 | 8.6 | 64.5 | 29.0 | 20.4 |
| Age & sex adjusted, using TaR after historic visit | 15.1 | 31.2 | 20.4 | 18.3 | 8.6 | 63.4 | 28.0 | 18.3 |
| Unadjusted, using entire historic period, filtered | 26.9 | 28.0 | 20.4 | 18.3 | 8.6 | 31.2 | 22.6 | 18.3 |
| Age & sex adjusted, using entire historic period, filtered | 25.8 | 28.0 | 21.5 | 19.4 | 7.5 | 32.3 | 23.7 | 19.4 |
| Unadjusted, using TaR after historic visit, filtered | 11.8 | 35.5 | 24.7 | 21.5 | 7.5 | 38.7 | 25.8 | 21.5 |
| Age & sex adjusted, using TaR after historic visit, filtered | 12.9 | 33.3 | 22.6 | 19.4 | 8.6 | 36.6 | 23.7 | 19.4 |
| SCCS / SCRI | | | | | | | | |
| Unadjusted SCCS excluding pre−vaccination window | 7.5 | 34.4 | 24.7 | 18.3 | 5.4 | 41.9 | 23.7 | 18.3 |
| Age & season adjusted SCCS excluding pre−vaccination window | 6.5 | 39.8 | 32.3 | 30.1 | 3.2 | 41.9 | 32.3 | 30.1 |
| SCRI with prior control interval | 17.2 | 47.3 | 36.6 | 22.6 | 1.1 | 63.4 | 41.9 | 23.7 |
| SCRI with posterior control interval | 11.8 | 51.6 | 32.3 | 23.7 | 4.3 | 58.1 | 33.3 | 23.7 |
| Unadjusted SCCS excluding all pre−vaccination time | 17.2 | 39.8 | 24.7 | 17.2 | 5.4 | 46.2 | 28.0 | 18.3 |

Figure 56: Type 1 and 2 error based on whether the log likelihood ratio exceeded the critical value any time during the study period in the MDCR database for Seasonal flu vaccination (All).

| | Uncalibrated | | | | Calibrated | | | |
|---|---|---|---|---|---|---|---|---|
| | | Type 2 when true effect is | | | | Type 2 when true effect is | | |
| | Type 1 | 1.5 | 2 | 4 | Type 1 | 1.5 | 2 | 4 |
| Case−control | | | | | | | | |
| Age & sex matched controls | 41.9 | 22.6 | 18.3 | 14.0 | 5.4 | 31.2 | 21.5 | 14.0 |
| Age & sex adjusted, using random controls | 45.2 | 21.5 | 16.1 | 11.8 | 9.7 | 28.0 | 19.4 | 12.9 |
| Cohort method | | | | | | | | |
| Unadjusted, using outpatient visits as comparator | 1.1 | 50.5 | 49.5 | 20.4 | 4.3 | 73.1 | 41.9 | 18.3 |
| PS matching, using outpatient visits as comparator | 2.2 | 47.3 | 25.8 | 12.9 | 9.7 | 38.7 | 23.7 | 12.9 |
| Unadjusted, using random days as comparator | 33.3 | 28.0 | 17.2 | 14.0 | 3.2 | 53.8 | 20.4 | 15.1 |
| PS matching, using random days as comparator | 10.8 | 37.6 | 26.9 | 14.0 | 7.5 | 38.7 | 26.9 | 14.0 |
| PS stratification, using outpatient visits as comparator | 2.2 | 48.4 | 25.8 | 15.1 | 7.5 | 37.6 | 21.5 | 14.0 |
| PS stratification, using random days as comparator | 12.9 | 36.6 | 23.7 | 12.9 | 5.4 | 37.6 | 23.7 | 14.0 |
| PS weighting, using outpatient visits as comparator | 2.2 | 57.0 | 32.3 | 16.1 | 6.5 | 48.4 | 26.9 | 15.1 |
| PS weighting, using random days as comparator | 1.1 | 55.9 | 37.6 | 17.2 | 2.2 | 53.8 | 36.6 | 17.2 |
| Per−month PS matching, using outpatient visits as comparator | | | | | | | | |
| Per−month PS matching, using random days as comparator | | | | | | | | |
| Historical comparator | | | | | | | | |
| Unadjusted, using entire historic period | 52.7 | 18.3 | 12.9 | 10.8 | 5.4 | 65.6 | 21.5 | 10.8 |
| Age & sex adjusted, using entire historic period | 48.4 | 17.2 | 11.8 | 9.7 | 5.4 | 55.9 | 19.4 | 9.7 |
| Unadjusted, using TaR after historic visit | 21.5 | 23.7 | 16.1 | 9.7 | 5.4 | 73.1 | 33.3 | 9.7 |
| Age & sex adjusted, using TaR after historic visit | 18.3 | 22.6 | 16.1 | 10.8 | 5.4 | 66.7 | 24.7 | 10.8 |
| Unadjusted, using entire historic period, filtered | 50.5 | 18.3 | 12.9 | 10.8 | 3.2 | 26.9 | 17.2 | 10.8 |
| Age & sex adjusted, using entire historic period, filtered | 46.2 | 17.2 | 11.8 | 9.7 | 4.3 | 21.5 | 14.0 | 9.7 |
| Unadjusted, using TaR after historic visit, filtered | 20.4 | 23.7 | 16.1 | 9.7 | 8.6 | 43.0 | 16.1 | 9.7 |
| Age & sex adjusted, using TaR after historic visit, filtered | 17.2 | 22.6 | 16.1 | 10.8 | 7.5 | 35.5 | 16.1 | 10.8 |
| SCCS / SCRI | | | | | | | | |
| Unadjusted SCCS excluding pre−vaccination window | 20.4 | 16.1 | 15.1 | 10.8 | 6.5 | 18.3 | 15.1 | 10.8 |
| Age & season adjusted SCCS excluding pre−vaccination window | 19.4 | 17.2 | 15.1 | 11.8 | 2.2 | 18.3 | 15.1 | 11.8 |
| SCRI with prior control interval | 30.1 | 24.7 | 16.1 | 11.8 | 2.2 | 44.1 | 19.4 | 11.8 |
| SCRI with posterior control interval | 25.8 | 19.4 | 15.1 | 12.9 | 5.4 | 29.0 | 15.1 | 12.9 |
| Unadjusted SCCS excluding all pre−vaccination time | 30.1 | 19.4 | 14.0 | 11.8 | 5.4 | 23.7 | 15.1 | 11.8 |

Figure 57: Type 1 and 2 error based on whether the log likelihood ratio exceeded the critical value any time during the study period in the CCAE database for Seasonal flu vaccination (All).

#### 11.5 Type 1 and 2 error for Zoster vaccination (Shingrix)

| | Uncalibrated | | | | Calibrated | | | |
|---|---|---|---|---|---|---|---|---|
| | | Type 2 when true effect is | | | | Type 2 when true effect is | | |
| | Type 1 | 1.5 | 2 | 4 | Type 1 | 1.5 | 2 | 4 |
| Case−control | | | | | | | | |
| Age & sex matched controls | 30.1 | 59.1 | 48.4 | 32.3 | 8.6 | 72.0 | 63.4 | 40.9 |
| Age & sex adjusted, using random controls | 24.7 | 54.8 | 46.2 | 31.2 | 10.8 | 74.2 | 55.9 | 34.4 |
| Cohort method | | | | | | | | |
| Unadjusted, using outpatient visits as comparator | 3.2 | 63.4 | 55.9 | 26.9 | 5.4 | 73.1 | 55.9 | 24.7 |
| PS matching, using outpatient visits as comparator | 1.1 | 75.3 | 52.7 | 23.7 | 6.5 | 61.3 | 43.0 | 21.5 |
| Unadjusted, using random days as comparator | 34.4 | 46.2 | 39.8 | 26.9 | 3.2 | 83.9 | 61.3 | 32.3 |
| PS matching, using random days as comparator | | | | | | | | |
| PS stratification, using outpatient visits as comparator | 1.1 | 76.3 | 51.6 | 18.3 | 6.5 | 63.4 | 34.4 | 17.2 |
| PS stratification, using random days as comparator | | | | | | | | |
| PS weighting, using outpatient visits as comparator | 1.1 | 76.3 | 62.4 | 38.7 | 3.2 | 69.9 | 53.8 | 32.3 |
| PS weighting, using random days as comparator | | | | | | | | |
| Per−month PS matching, using outpatient visits as comparator | | | | | | | | |
| Per−month PS matching, using random days as comparator | | | | | | | | |
| Historical comparator | | | | | | | | |
| Unadjusted, using entire historic period | 57.0 | 31.2 | 29.0 | 22.6 | 3.2 | 80.6 | 49.5 | 28.0 |
| Age & sex adjusted, using entire historic period | 46.2 | 28.0 | 24.7 | 21.5 | 2.2 | 73.1 | 46.2 | 22.6 |
| Unadjusted, using TaR after historic visit | 36.6 | 39.8 | 29.0 | 18.3 | 2.2 | 75.3 | 49.5 | 23.7 |
| Age & sex adjusted, using TaR after historic visit | 24.7 | 43.0 | 28.0 | 14.0 | 6.5 | 73.1 | 47.3 | 16.1 |
| Unadjusted, using entire historic period, filtered | 55.9 | 32.3 | 30.1 | 23.7 | 3.2 | 77.4 | 47.3 | 29.0 |
| Age & sex adjusted, using entire historic period, filtered | 45.2 | 29.0 | 25.8 | 22.6 | 2.2 | 67.7 | 46.2 | 23.7 |
| Unadjusted, using TaR after historic visit, filtered | 35.5 | 40.9 | 30.1 | 19.4 | 2.2 | 76.3 | 49.5 | 24.7 |
| Age & sex adjusted, using TaR after historic visit, filtered | 23.7 | 44.1 | 29.0 | 15.1 | 6.5 | 71.0 | 43.0 | 17.2 |
| SCCS / SCRI | | | | | | | | |
| Unadjusted SCCS excluding pre−vaccination window | 12.9 | 40.9 | 19.4 | 11.8 | 4.3 | 55.9 | 23.7 | 11.8 |
| Age & season adjusted SCCS excluding pre−vaccination window | 11.8 | 39.8 | 19.4 | 11.8 | 8.6 | 45.2 | 20.4 | 11.8 |
| SCRI with prior control interval | 21.5 | 51.6 | 32.3 | 20.4 | 10.8 | 60.2 | 39.8 | 21.5 |
| SCRI with posterior control interval | 16.1 | 57.0 | 39.8 | 20.4 | 2.2 | 66.7 | 44.1 | 23.7 |
| Unadjusted SCCS excluding all pre−vaccination time | 21.5 | 40.9 | 26.9 | 14.0 | 6.5 | 55.9 | 31.2 | 15.1 |

Figure 58: Type 1 and 2 error based on whether the log likelihood ratio exceeded the critical value any time during the study period in the Optum EHR database for Zoster vaccination (Shingrix).

| | Uncalibrated | | | | Calibrated | | | |
|---|---|---|---|---|---|---|---|---|
| | | Type 2 when true effect is | | | | Type 2 when true effect is | | |
| | Type 1 | 1.5 | 2 | 4 | Type 1 | 1.5 | 2 | 4 |
| Case−control | | | | | | | | |
| Age & sex matched controls | 9.7 | 81.7 | 69.9 | 58.1 | 2.2 | 92.5 | 80.6 | 59.1 |
| Age & sex adjusted, using random controls | 6.5 | 76.3 | 69.9 | 55.9 | 2.2 | 89.2 | 76.3 | 55.9 |
| Cohort method | | | | | | | | |
| Unadjusted, using outpatient visits as comparator | 0.0 | 91.4 | 82.8 | 72.0 | 0.0 | 91.4 | 84.9 | 72.0 |
| PS matching, using outpatient visits as comparator | 0.0 | 97.8 | 91.4 | 79.6 | 0.0 | 95.7 | 86.0 | 77.4 |
| Unadjusted, using random days as comparator | 8.6 | 90.3 | 84.9 | 79.6 | 2.2 | 95.7 | 90.3 | 82.8 |
| PS matching, using random days as comparator | 0.0 | 95.7 | 89.2 | 78.5 | 0.0 | 94.6 | 88.2 | 76.3 |
| PS stratification, using outpatient visits as comparator | 2.2 | 91.4 | 77.4 | 59.1 | 2.2 | 88.2 | 74.2 | 59.1 |
| PS stratification, using random days as comparator | 3.2 | 86.0 | 73.1 | 57.0 | 0.0 | 92.5 | 74.2 | 58.1 |
| PS weighting, using outpatient visits as comparator | 0.0 | 100.0 | 96.8 | 82.8 | 0.0 | 100.0 | 95.7 | 82.8 |
| PS weighting, using random days as comparator | 0.0 | 97.8 | 93.5 | 82.8 | 0.0 | 97.8 | 93.5 | 81.7 |
| Per−month PS matching, using outpatient visits as comparator | | | | | | | | |
| Per−month PS matching, using random days as comparator | | | | | | | | |
| Historical comparator | | | | | | | | |
| Unadjusted, using entire historic period | 15.1 | 78.5 | 77.4 | 76.3 | 0.0 | 95.7 | 90.3 | 78.5 |
| Age & sex adjusted, using entire historic period | 12.9 | 68.8 | 66.7 | 64.5 | 1.1 | 90.3 | 74.2 | 64.5 |
| Unadjusted, using TaR after historic visit | 16.1 | 74.2 | 71.0 | 69.9 | 1.1 | 93.5 | 86.0 | 71.0 |
| Age & sex adjusted, using TaR after historic visit | 5.4 | 73.1 | 67.7 | 59.1 | 1.1 | 84.9 | 72.0 | 59.1 |
| Unadjusted, using entire historic period, filtered | 15.1 | 78.5 | 77.4 | 76.3 | 1.1 | 92.5 | 87.1 | 78.5 |
| Age & sex adjusted, using entire historic period, filtered | 10.8 | 69.9 | 67.7 | 65.6 | 1.1 | 80.6 | 71.0 | 65.6 |
| Unadjusted, using TaR after historic visit, filtered | 15.1 | 75.3 | 72.0 | 71.0 | 3.2 | 91.4 | 84.9 | 72.0 |
| Age & sex adjusted, using TaR after historic visit, filtered | 3.2 | 75.3 | 68.8 | 60.2 | 2.2 | 79.6 | 72.0 | 60.2 |
| SCCS / SCRI | | | | | | | | |
| Unadjusted SCCS excluding pre−vaccination window | 2.2 | 77.4 | 62.4 | 49.5 | 2.2 | 81.7 | 62.4 | 49.5 |
| Age & season adjusted SCCS excluding pre−vaccination window | 4.3 | 73.1 | 62.4 | 49.5 | 3.2 | 74.2 | 64.5 | 49.5 |
| SCRI with prior control interval | 5.4 | 90.3 | 80.6 | 60.2 | 1.1 | 90.3 | 79.6 | 59.1 |
| SCRI with posterior control interval | 0.0 | 91.4 | 84.9 | 61.3 | 1.1 | 91.4 | 84.9 | 61.3 |
| Unadjusted SCCS excluding all pre−vaccination time | 6.5 | 86.0 | 67.7 | 52.7 | 3.2 | 87.1 | 72.0 | 52.7 |

Figure 59: Type 1 and 2 error based on whether the log likelihood ratio exceeded the critical value any time during the study period in the MDCD database for Zoster vaccination (Shingrix).

| | Uncalibrated | | | | Calibrated | | | |
|---|---|---|---|---|---|---|---|---|
| | | Type 2 when true effect is | | | | Type 2 when true effect is | | |
| | Type 1 | 1.5 | 2 | 4 | Type 1 | 1.5 | 2 | 4 |
| Case−control | | | | | | | | |
| Age & sex matched controls | 15.1 | 66.7 | 57.0 | 35.5 | 5.4 | 72.0 | 58.1 | 35.5 |
| Age & sex adjusted, using random controls | 14.0 | 65.6 | 50.5 | 31.2 | 8.6 | 73.1 | 54.8 | 31.2 |
| Cohort method | | | | | | | | |
| Unadjusted, using outpatient visits as comparator | 1.1 | 83.9 | 77.4 | 58.1 | 2.2 | 87.1 | 76.3 | 54.8 |
| PS matching, using outpatient visits as comparator | 2.2 | 82.8 | 72.0 | 54.8 | 4.3 | 80.6 | 69.9 | 54.8 |
| Unadjusted, using random days as comparator | 2.2 | 78.5 | 66.7 | 50.5 | 1.1 | 81.7 | 71.0 | 50.5 |
| PS matching, using random days as comparator | 2.2 | 84.9 | 67.7 | 52.7 | 2.2 | 81.7 | 67.7 | 52.7 |
| PS stratification, using outpatient visits as comparator | 3.2 | 78.5 | 65.6 | 43.0 | 3.2 | 75.3 | 63.4 | 40.9 |
| PS stratification, using random days as comparator | 4.3 | 77.4 | 55.9 | 37.6 | 4.3 | 77.4 | 58.1 | 38.7 |
| PS weighting, using outpatient visits as comparator | 1.1 | 79.6 | 67.7 | 50.5 | 2.2 | 79.6 | 65.6 | 50.5 |
| PS weighting, using random days as comparator | 0.0 | 93.5 | 86.0 | 66.7 | 0.0 | 93.5 | 86.0 | 66.7 |
| Per−month PS matching, using outpatient visits as comparator | | | | | | | | |
| Per−month PS matching, using random days as comparator | | | | | | | | |
| Historical comparator | | | | | | | | |
| Unadjusted, using entire historic period | 15.1 | 66.7 | 52.7 | 40.9 | 4.3 | 82.8 | 69.9 | 43.0 |
| Age & sex adjusted, using entire historic period | 12.9 | 63.4 | 51.6 | 39.8 | 5.4 | 82.8 | 67.7 | 40.9 |
| Unadjusted, using TaR after historic visit | 12.9 | 66.7 | 57.0 | 39.8 | 5.4 | 84.9 | 72.0 | 45.2 |
| Age & sex adjusted, using TaR after historic visit | 9.7 | 68.8 | 58.1 | 39.8 | 4.3 | 86.0 | 71.0 | 43.0 |
| Unadjusted, using entire historic period, filtered | 14.0 | 67.7 | 53.8 | 41.9 | 5.4 | 78.5 | 66.7 | 43.0 |
| Age & sex adjusted, using entire historic period, filtered | 10.8 | 65.6 | 53.8 | 41.9 | 4.3 | 75.3 | 64.5 | 43.0 |
| Unadjusted, using TaR after historic visit, filtered | 9.7 | 68.8 | 59.1 | 41.9 | 3.2 | 82.8 | 68.8 | 41.9 |
| Age & sex adjusted, using TaR after historic visit, filtered | 8.6 | 69.9 | 59.1 | 40.9 | 3.2 | 80.6 | 67.7 | 40.9 |
| SCCS / SCRI | | | | | | | | |
| Unadjusted SCCS excluding pre−vaccination window | 14.0 | 62.4 | 48.4 | 33.3 | 6.5 | 68.8 | 51.6 | 34.4 |
| Age & season adjusted SCCS excluding pre−vaccination window | 10.8 | 61.3 | 48.4 | 37.6 | 9.7 | 62.4 | 48.4 | 38.7 |
| SCRI with prior control interval | 7.5 | 72.0 | 59.1 | 47.3 | 2.2 | 75.3 | 62.4 | 48.4 |
| SCRI with posterior control interval | 8.6 | 81.7 | 65.6 | 49.5 | 4.3 | 81.7 | 66.7 | 49.5 |
| Unadjusted SCCS excluding all pre−vaccination time | 8.6 | 72.0 | 52.7 | 35.5 | 3.2 | 73.1 | 55.9 | 36.6 |

Figure 60: Type 1 and 2 error based on whether the log likelihood ratio exceeded the critical value any time during the study period in the MDCR database for Zoster vaccination (Shingrix).

| | Uncalibrated | | | | Calibrated | | | |
|---|---|---|---|---|---|---|---|---|
| | | Type 2 when true effect is | | | | Type 2 when true effect is | | |
| | Type 1 | 1.5 | 2 | 4 | Type 1 | 1.5 | 2 | 4 |
| Case−control | | | | | | | | |
| Age & sex matched controls | 8.6 | 73.1 | 62.4 | 32.3 | 3.2 | 76.3 | 65.6 | 34.4 |
| Age & sex adjusted, using random controls | 9.7 | 67.7 | 58.1 | 30.1 | 4.3 | 77.4 | 59.1 | 30.1 |
| Cohort method | | | | | | | | |
| Unadjusted, using outpatient visits as comparator | 2.2 | 80.6 | 69.9 | 38.7 | 2.2 | 83.9 | 61.3 | 37.6 |
| PS matching, using outpatient visits as comparator | 0.0 | 76.3 | 64.5 | 35.5 | 2.2 | 69.9 | 54.8 | 34.4 |
| Unadjusted, using random days as comparator | 18.3 | 67.7 | 55.9 | 41.9 | 2.2 | 89.2 | 75.3 | 48.4 |
| PS matching, using random days as comparator | 1.1 | 78.5 | 54.8 | 37.6 | 2.2 | 77.4 | 54.8 | 37.6 |
| PS stratification, using outpatient visits as comparator | 0.0 | 80.6 | 55.9 | 31.2 | 4.3 | 66.7 | 48.4 | 29.0 |
| PS stratification, using random days as comparator | 4.3 | 67.7 | 44.1 | 23.7 | 3.2 | 67.7 | 45.2 | 23.7 |
| PS weighting, using outpatient visits as comparator | 0.0 | 84.9 | 75.3 | 51.6 | 3.2 | 80.6 | 73.1 | 46.2 |
| PS weighting, using random days as comparator | 0.0 | 91.4 | 82.8 | 52.7 | 0.0 | 89.2 | 82.8 | 52.7 |
| Per−month PS matching, using outpatient visits as comparator | | | | | | | | |
| Per−month PS matching, using random days as comparator | | | | | | | | |
| Historical comparator | | | | | | | | |
| Unadjusted, using entire historic period | 35.5 | 41.9 | 38.7 | 30.1 | 5.4 | 76.3 | 57.0 | 32.3 |
| Age & sex adjusted, using entire historic period | 18.3 | 41.9 | 28.0 | 21.5 | 5.4 | 66.7 | 40.9 | 22.6 |
| Unadjusted, using TaR after historic visit | 24.7 | 44.1 | 36.6 | 23.7 | 3.2 | 74.2 | 51.6 | 28.0 |
| Age & sex adjusted, using TaR after historic visit | 9.7 | 55.9 | 44.1 | 24.7 | 5.4 | 73.1 | 47.3 | 24.7 |
| Unadjusted, using entire historic period, filtered | 33.3 | 44.1 | 40.9 | 32.3 | 4.3 | 69.9 | 52.7 | 34.4 |
| Age & sex adjusted, using entire historic period, filtered | 15.1 | 44.1 | 30.1 | 23.7 | 5.4 | 51.6 | 33.3 | 24.7 |
| Unadjusted, using TaR after historic visit, filtered | 22.6 | 46.2 | 38.7 | 25.8 | 3.2 | 67.7 | 49.5 | 29.0 |
| Age & sex adjusted, using TaR after historic visit, filtered | 6.5 | 58.1 | 46.2 | 26.9 | 5.4 | 58.1 | 45.2 | 25.8 |
| SCCS / SCRI | | | | | | | | |
| Unadjusted SCCS excluding pre−vaccination window | 8.6 | 54.8 | 32.3 | 20.4 | 5.4 | 58.1 | 36.6 | 20.4 |
| Age & season adjusted SCCS excluding pre−vaccination window | 12.9 | 49.5 | 24.7 | 19.4 | 6.5 | 53.8 | 26.9 | 19.4 |
| SCRI with prior control interval | 7.5 | 73.1 | 47.3 | 26.9 | 3.2 | 73.1 | 49.5 | 26.9 |
| SCRI with posterior control interval | 6.5 | 73.1 | 53.8 | 37.6 | 3.2 | 73.1 | 57.0 | 37.6 |
| Unadjusted SCCS excluding all pre−vaccination time | 10.8 | 57.0 | 43.0 | 22.6 | 3.2 | 62.4 | 44.1 | 22.6 |

Figure 61: Type 1 and 2 error based on whether the log likelihood ratio exceeded the critical value any time during the study period in the CCAE database for Zoster vaccination (Shingrix).

#### 11.6 Type 1 and 2 error for HPV vaccination (Gardasil 9)

| | Uncalibrated | | | | Calibrated | | | |
|---|---|---|---|---|---|---|---|---|
| | | Type 2 when true effect is | | | | Type 2 when true effect is | | |
| | Type 1 | 1.5 | 2 | 4 | Type 1 | 1.5 | 2 | 4 |
| Case−control | | | | | | | | |
| Age & sex matched controls | 19.4 | 66.7 | 60.2 | 51.6 | 2.2 | 87.1 | 75.3 | 55.9 |
| Age & sex adjusted, using random controls | 8.6 | 77.4 | 73.1 | 52.7 | 5.4 | 86.0 | 77.4 | 52.7 |
| Cohort method | | | | | | | | |
| Unadjusted, using outpatient visits as comparator | 1.1 | 72.0 | 76.3 | 71.0 | 3.2 | 93.5 | 86.0 | 72.0 |
| PS matching, using outpatient visits as comparator | 1.1 | 89.2 | 77.4 | 61.3 | 4.3 | 87.1 | 69.9 | 59.1 |
| Unadjusted, using random days as comparator | 4.3 | 79.6 | 74.2 | 61.3 | 3.2 | 94.6 | 86.0 | 63.4 |
| PS matching, using random days as comparator | | | | | | | | |
| PS stratification, using outpatient visits as comparator | 1.1 | 87.1 | 72.0 | 47.3 | 3.2 | 82.8 | 61.3 | 45.2 |
| PS stratification, using random days as comparator | | | | | | | | |
| PS weighting, using outpatient visits as comparator | 0.0 | 95.7 | 91.4 | 73.1 | 0.0 | 94.6 | 88.2 | 73.1 |
| PS weighting, using random days as comparator | | | | | | | | |
| Per−month PS matching, using outpatient visits as comparator | | | | | | | | |
| Per−month PS matching, using random days as comparator | | | | | | | | |
| Historical comparator | | | | | | | | |
| Unadjusted, using entire historic period | 12.9 | 68.8 | 63.4 | 47.3 | 1.1 | 88.2 | 83.9 | 54.8 |
| Age & sex adjusted, using entire historic period | 18.3 | 61.3 | 59.1 | 57.0 | 3.2 | 89.2 | 73.1 | 57.0 |
| Unadjusted, using TaR after historic visit | 7.5 | 71.0 | 67.7 | 53.8 | 2.2 | 91.4 | 82.8 | 63.4 |
| Age & sex adjusted, using TaR after historic visit | 7.5 | 68.8 | 54.8 | 46.2 | 2.2 | 81.7 | 61.3 | 47.3 |
| Unadjusted, using entire historic period, filtered | 11.8 | 69.9 | 64.5 | 48.4 | 1.1 | 89.2 | 84.9 | 55.9 |
| Age & sex adjusted, using entire historic period, filtered | 17.2 | 62.4 | 60.2 | 58.1 | 2.2 | 79.6 | 63.4 | 58.1 |
| Unadjusted, using TaR after historic visit, filtered | 7.5 | 72.0 | 68.8 | 54.8 | 1.1 | 92.5 | 84.9 | 63.4 |
| Age & sex adjusted, using TaR after historic visit, filtered | 6.5 | 69.9 | 55.9 | 47.3 | 2.2 | 75.3 | 58.1 | 47.3 |
| SCCS / SCRI | | | | | | | | |
| Unadjusted SCCS excluding pre−vaccination window | 8.6 | 59.1 | 51.6 | 37.6 | 4.3 | 79.6 | 57.0 | 37.6 |
| Age & season adjusted SCCS excluding pre−vaccination window | 7.5 | 59.1 | 52.7 | 38.7 | 3.2 | 77.4 | 63.4 | 39.8 |
| SCRI with prior control interval | 11.8 | 78.5 | 68.8 | 60.2 | 2.2 | 90.3 | 71.0 | 60.2 |
| SCRI with posterior control interval | 11.8 | 78.5 | 68.8 | 59.1 | 2.2 | 82.8 | 72.0 | 59.1 |
| Unadjusted SCCS excluding all pre−vaccination time | 10.8 | 63.4 | 55.9 | 39.8 | 5.4 | 75.3 | 60.2 | 41.9 |

Figure 62: Type 1 and 2 error based on whether the log likelihood ratio exceeded the critical value any time during the study period in the Optum EHR database for HPV vaccination (Gardasil 9).

| | Uncalibrated | | | | Calibrated | | | |
|---|---|---|---|---|---|---|---|---|
| | | Type 2 when true effect is | | | | Type 2 when true effect is | | |
| | Type 1 | 1.5 | 2 | 4 | Type 1 | 1.5 | 2 | 4 |
| Case−control | | | | | | | | |
| Age & sex matched controls | 15.1 | 65.6 | 59.1 | 55.9 | 2.2 | 78.5 | 65.6 | 55.9 |
| Age & sex adjusted, using random controls | 14.0 | 69.9 | 64.5 | 51.6 | 3.2 | 77.4 | 66.7 | 51.6 |
| Cohort method | | | | | | | | |
| Unadjusted, using outpatient visits as comparator | 2.2 | 78.5 | 76.3 | 76.3 | 1.1 | 97.8 | 90.3 | 77.4 |
| PS matching, using outpatient visits as comparator | 2.2 | 84.9 | 79.6 | 69.9 | 1.1 | 87.1 | 80.6 | 69.9 |
| Unadjusted, using random days as comparator | 5.4 | 77.4 | 78.5 | 67.7 | 2.2 | 97.8 | 89.2 | 69.9 |
| PS matching, using random days as comparator | 3.2 | 87.1 | 80.6 | 68.8 | 2.2 | 92.5 | 80.6 | 68.8 |
| PS stratification, using outpatient visits as comparator | 3.2 | 80.6 | 76.3 | 54.8 | 3.2 | 84.9 | 77.4 | 54.8 |
| PS stratification, using random days as comparator | 7.5 | 74.2 | 66.7 | 59.1 | 5.4 | 80.6 | 69.9 | 60.2 |
| PS weighting, using outpatient visits as comparator | 1.1 | 93.5 | 86.0 | 80.6 | 1.1 | 93.5 | 86.0 | 81.7 |
| PS weighting, using random days as comparator | 2.2 | 93.5 | 86.0 | 78.5 | 1.1 | 95.7 | 90.3 | 79.6 |
| Per−month PS matching, using outpatient visits as comparator | | | | | | | | |
| Per−month PS matching, using random days as comparator | | | | | | | | |
| Historical comparator | | | | | | | | |
| Unadjusted, using entire historic period | 8.6 | 72.0 | 67.7 | 57.0 | 2.2 | 93.5 | 83.9 | 63.4 |
| Age & sex adjusted, using entire historic period | 10.8 | 67.7 | 65.6 | 60.2 | 4.3 | 91.4 | 78.5 | 60.2 |
| Unadjusted, using TaR after historic visit | 5.4 | 68.8 | 68.8 | 60.2 | 2.2 | 96.8 | 80.6 | 64.5 |
| Age & sex adjusted, using TaR after historic visit | 5.4 | 74.2 | 65.6 | 53.8 | 4.3 | 91.4 | 79.6 | 57.0 |
| Unadjusted, using entire historic period, filtered | 6.5 | 75.3 | 69.9 | 59.1 | 2.2 | 96.8 | 86.0 | 65.6 |
| Age & sex adjusted, using entire historic period, filtered | 9.7 | 68.8 | 66.7 | 61.3 | 3.2 | 87.1 | 67.7 | 61.3 |
| Unadjusted, using TaR after historic visit, filtered | 4.3 | 73.1 | 71.0 | 62.4 | 2.2 | 97.8 | 84.9 | 66.7 |
| Age & sex adjusted, using TaR after historic visit, filtered | 4.3 | 75.3 | 66.7 | 54.8 | 2.2 | 83.9 | 72.0 | 55.9 |
| SCCS / SCRI | | | | | | | | |
| Unadjusted SCCS excluding pre−vaccination window | 4.3 | 69.9 | 62.4 | 47.3 | 2.2 | 75.3 | 63.4 | 47.3 |
| Age & season adjusted SCCS excluding pre−vaccination window | 4.3 | 68.8 | 59.1 | 47.3 | 3.2 | 72.0 | 60.2 | 48.4 |
| SCRI with prior control interval | 12.9 | 79.6 | 73.1 | 61.3 | 2.2 | 89.2 | 77.4 | 67.7 |
| SCRI with posterior control interval | 5.4 | 82.8 | 74.2 | 66.7 | 1.1 | 91.4 | 81.7 | 66.7 |
| Unadjusted SCCS excluding all pre−vaccination time | 7.5 | 71.0 | 63.4 | 52.7 | 1.1 | 89.2 | 68.8 | 54.8 |

Figure 63: Type 1 and 2 error based on whether the log likelihood ratio exceeded the critical value any time during the study period in the MDCD database for HPV vaccination (Gardasil 9).

| | Uncalibrated | | | | Calibrated | | | |
|---|---|---|---|---|---|---|---|---|
| | | Type 2 when true effect is | | | | Type 2 when true effect is | | |
| | Type 1 | 1.5 | 2 | 4 | Type 1 | 1.5 | 2 | 4 |
| Case−control | | | | | | | | |
| Age & sex matched controls | | | | | | | | |
| Age & sex adjusted, using random controls | | | | | | | | |
| Cohort method | | | | | | | | |
| Unadjusted, using outpatient visits as comparator | 0.0 | 100.0 | 100.0 | 100.0 | 0.0 | 100.0 | 100.0 | 100.0 |
| PS matching, using outpatient visits as comparator | | | | | | | | |
| Unadjusted, using random days as comparator | | | | | | | | |
| PS matching, using random days as comparator | | | | | | | | |
| PS stratification, using outpatient visits as comparator | 0.0 | 100.0 | 100.0 | 100.0 | 0.0 | 100.0 | 100.0 | 100.0 |
| PS stratification, using random days as comparator | | | | | | | | |
| PS weighting, using outpatient visits as comparator | 0.0 | 100.0 | 100.0 | 100.0 | 0.0 | 100.0 | 100.0 | 100.0 |
| PS weighting, using random days as comparator | | | | | | | | |
| Per−month PS matching, using outpatient visits as comparator | | | | | | | | |
| Per−month PS matching, using random days as comparator | | | | | | | | |
| Historical comparator | | | | | | | | |
| Unadjusted, using entire historic period | 0.0 | 100.0 | 100.0 | 100.0 | 0.0 | 100.0 | 100.0 | 100.0 |
| Age & sex adjusted, using entire historic period | 0.0 | 100.0 | 100.0 | 100.0 | 0.0 | 100.0 | 100.0 | 100.0 |
| Unadjusted, using TaR after historic visit | 0.0 | 100.0 | 100.0 | 100.0 | 0.0 | 100.0 | 100.0 | 100.0 |
| Age & sex adjusted, using TaR after historic visit | 0.0 | 100.0 | 100.0 | 100.0 | 0.0 | 100.0 | 100.0 | 100.0 |
| Unadjusted, using entire historic period, filtered | 0.0 | 100.0 | 100.0 | 100.0 | 0.0 | 100.0 | 100.0 | 100.0 |
| Age & sex adjusted, using entire historic period, filtered | 0.0 | 100.0 | 100.0 | 100.0 | 0.0 | 100.0 | 100.0 | 100.0 |
| Unadjusted, using TaR after historic visit, filtered | 0.0 | 100.0 | 100.0 | 100.0 | 0.0 | 100.0 | 100.0 | 100.0 |
| Age & sex adjusted, using TaR after historic visit, filtered | 0.0 | 100.0 | 100.0 | 100.0 | 0.0 | 100.0 | 100.0 | 100.0 |

Figure 64: Type 1 and 2 error based on whether the log likelihood ratio exceeded the critical value any time during the study period in the MDCR database for HPV vaccination (Gardasil 9).

| | Uncalibrated | | | | Calibrated | | | |
|---|---|---|---|---|---|---|---|---|
| | | Type 2 when true effect is | | | | Type 2 when true effect is | | |
| | Type 1 | 1.5 | 2 | 4 | Type 1 | 1.5 | 2 | 4 |
| Case−control | | | | | | | | |
| Age & sex matched controls | 12.9 | 63.4 | 54.8 | 49.5 | 4.3 | 75.3 | 62.4 | 50.5 |
| Age & sex adjusted, using random controls | 11.8 | 69.9 | 61.3 | 49.5 | 4.3 | 79.6 | 64.5 | 49.5 |
| Cohort method | | | | | | | | |
| Unadjusted, using outpatient visits as comparator | 2.2 | 64.5 | 74.2 | 68.8 | 2.2 | 96.8 | 91.4 | 74.2 |
| PS matching, using outpatient visits as comparator | 1.1 | 87.1 | 71.0 | 60.2 | 1.1 | 82.8 | 69.9 | 58.1 |
| Unadjusted, using random days as comparator | 5.4 | 81.7 | 75.3 | 62.4 | 2.2 | 93.5 | 90.3 | 65.6 |
| PS matching, using random days as comparator | 2.2 | 88.2 | 73.1 | 62.4 | 2.2 | 90.3 | 73.1 | 62.4 |
| PS stratification, using outpatient visits as comparator | 1.1 | 83.9 | 69.9 | 47.3 | 4.3 | 78.5 | 62.4 | 46.2 |
| PS stratification, using random days as comparator | 2.2 | 78.5 | 61.3 | 48.4 | 2.2 | 84.9 | 63.4 | 48.4 |
| PS weighting, using outpatient visits as comparator | 0.0 | 95.7 | 88.2 | 73.1 | 0.0 | 95.7 | 87.1 | 71.0 |
| PS weighting, using random days as comparator | 1.1 | 91.4 | 88.2 | 73.1 | 1.1 | 94.6 | 88.2 | 74.2 |
| Per−month PS matching, using outpatient visits as comparator | | | | | | | | |
| Per−month PS matching, using random days as comparator | | | | | | | | |
| Historical comparator | | | | | | | | |
| Unadjusted, using entire historic period | 9.7 | 68.8 | 63.4 | 47.3 | 2.2 | 93.5 | 79.6 | 51.6 |
| Age & sex adjusted, using entire historic period | 12.9 | 55.9 | 52.7 | 48.4 | 3.2 | 82.8 | 61.3 | 48.4 |
| Unadjusted, using TaR after historic visit | 8.6 | 71.0 | 65.6 | 50.5 | 2.2 | 94.6 | 84.9 | 59.1 |
| Age & sex adjusted, using TaR after historic visit | 9.7 | 69.9 | 52.7 | 45.2 | 4.3 | 83.9 | 65.6 | 45.2 |
| Unadjusted, using entire historic period, filtered | 9.7 | 71.0 | 66.7 | 49.5 | 2.2 | 94.6 | 82.8 | 53.8 |
| Age & sex adjusted, using entire historic period, filtered | 10.8 | 59.1 | 54.8 | 50.5 | 4.3 | 74.2 | 55.9 | 50.5 |
| Unadjusted, using TaR after historic visit, filtered | 8.6 | 73.1 | 67.7 | 52.7 | 1.1 | 94.6 | 88.2 | 61.3 |
| Age & sex adjusted, using TaR after historic visit, filtered | 7.5 | 72.0 | 54.8 | 47.3 | 4.3 | 81.7 | 60.2 | 47.3 |
| SCCS / SCRI | | | | | | | | |
| Unadjusted SCCS excluding pre−vaccination window | 11.8 | 59.1 | 47.3 | 38.7 | 5.4 | 71.0 | 54.8 | 39.8 |
| Age & season adjusted SCCS excluding pre−vaccination window | 14.0 | 55.9 | 48.4 | 38.7 | 7.5 | 67.7 | 51.6 | 38.7 |
| SCRI with prior control interval | 12.9 | 77.4 | 62.4 | 55.9 | 2.2 | 90.3 | 76.3 | 55.9 |
| SCRI with posterior control interval | 4.3 | 80.6 | 65.6 | 54.8 | 1.1 | 90.3 | 69.9 | 54.8 |
| Unadjusted SCCS excluding all pre−vaccination time | 9.7 | 64.5 | 53.8 | 40.9 | 3.2 | 78.5 | 58.1 | 40.9 |

Figure 65: Type 1 and 2 error based on whether the log likelihood ratio exceeded the critical value any time during the study period in the CCAE database for HPV vaccination (Gardasil 9).

### 12 AUC based on effect-size estimate

For each method variation and vaccine group, the area under the receiver operator curve (AUC) stratified by true effect size, based on the effect-size estimate at the end of the study period.

#### 12.1 AUC for H1N1 vaccination

| | Uncalibrated AUC when true effect is | | | Calibrated AUC when true effect is | | |
|---|---|---|---|---|---|---|
| | 1.5 | 2 | 4 | 1.5 | 2 | 4 |
| Case−control | | | | | | |
| Age & sex matched controls | 55.1 | 57.2 | 61.1 | 64.5 | 69.1 | 72.1 |
| Age & sex adjusted, using random controls | 57.4 | 59.9 | 64.2 | 67.6 | 75.4 | 78.5 |
| Cohort method | | | | | | |
| Unadjusted, using outpatient visits as comparator | 62.3 | 68.2 | 83.1 | 71.6 | 77.8 | 82.8 |
| PS matching, using outpatient visits as comparator | 62.8 | 69.6 | 82.1 | 73.5 | 80.6 | 84.4 |
| Unadjusted, using random days as comparator | 55.9 | 58.0 | 61.8 | 65.1 | 73.0 | 75.0 |
| PS matching, using random days as comparator | 50.0 | 50.0 | 50.0 | 50.0 | 50.0 | 50.0 |
| PS stratification, using outpatient visits as comparator | 62.9 | 71.0 | 86.5 | 72.4 | 79.1 | 84.8 |
| PS stratification, using random days as comparator | 59.3 | 64.1 | 72.2 | 72.1 | 78.0 | 81.0 |
| PS weighting, using outpatient visits as comparator | 62.3 | 70.3 | 86.2 | 70.3 | 78.2 | 84.4 |
| PS weighting, using random days as comparator | 50.0 | 50.1 | 50.1 | 50.0 | 50.0 | 50.0 |
| Per−month PS matching, using outpatient visits as comparator | 61.2 | 66.6 | 76.9 | 68.3 | 74.8 | 76.3 |
| Per−month PS matching, using random days as comparator | 57.9 | 59.6 | 65.6 | 60.6 | 66.1 | 67.6 |
| Historical comparator | | | | | | |
| Unadjusted, using entire historic period | 60.4 | 66.8 | 74.4 | 74.9 | 81.6 | 85.9 |
| Age & sex adjusted, using entire historic period | 61.3 | 67.1 | 74.2 | 75.2 | 81.7 | 85.9 |
| Unadjusted, using TaR after historic visit | 63.0 | 68.6 | 77.9 | 75.1 | 82.2 | 87.0 |
| Age & sex adjusted, using TaR after historic visit | 62.9 | 68.5 | 76.9 | 75.4 | 81.3 | 85.9 |
| Unadjusted, using entire historic period, filtered | 59.5 | 65.5 | 72.0 | 74.0 | 79.7 | 83.7 |
| Age & sex adjusted, using entire historic period, filtered | 60.5 | 65.8 | 71.9 | 73.4 | 79.3 | 83.0 |
| Unadjusted, using TaR after historic visit, filtered | 62.5 | 67.6 | 76.2 | 74.5 | 81.2 | 85.6 |
| Age & sex adjusted, using TaR after historic visit, filtered | 62.3 | 67.3 | 75.1 | 74.4 | 79.7 | 84.0 |
| SCCS / SCRI | | | | | | |
| Unadjusted SCCS excluding pre−vaccination window | 65.3 | 72.4 | 80.7 | 75.0 | 80.7 | 84.0 |
| Age & season adjusted SCCS excluding pre−vaccination window | 64.5 | 71.6 | 80.3 | 74.4 | 79.7 | 82.9 |
| SCRI with prior control interval | 64.4 | 70.6 | 79.3 | 71.9 | 79.3 | 82.3 |
| SCRI with posterior control interval | 63.7 | 69.0 | 75.2 | 72.1 | 76.9 | 80.5 |
| Unadjusted SCCS excluding all pre−vaccination time | 64.4 | 69.8 | 76.2 | 73.4 | 78.6 | 83.0 |

Figure 66: The area under the receiver operator curve (AUC) based on the effect-size estimate at the end of the study period in the Optum EHR database for H1N1 vaccination.

| | Uncalibrated AUC when true effect is | | | Calibrated AUC when true effect is | | |
|---|---|---|---|---|---|---|
| | 1.5 | 2 | 4 | 1.5 | 2 | 4 |
| Case−control | | | | | | |
| Age & sex matched controls | 58.8 | 62.9 | 68.4 | 68.2 | 73.4 | 76.9 |
| Age & sex adjusted, using random controls | 62.5 | 67.2 | 73.8 | 72.6 | 77.7 | 80.3 |
| Cohort method | | | | | | |
| Unadjusted, using outpatient visits as comparator | 60.4 | 65.7 | 75.4 | 67.9 | 75.4 | 83.0 |
| PS matching, using outpatient visits as comparator | 60.4 | 67.3 | 75.0 | 70.5 | 74.3 | 76.8 |
| Unadjusted, using random days as comparator | 58.5 | 63.9 | 78.3 | 68.9 | 76.4 | 81.8 |
| PS matching, using random days as comparator | 61.6 | 66.6 | 74.8 | 67.2 | 73.9 | 75.5 |
| PS stratification, using outpatient visits as comparator | 61.6 | 69.0 | 82.4 | 71.9 | 77.8 | 81.4 |
| PS stratification, using random days as comparator | 62.4 | 69.8 | 81.4 | 72.2 | 77.8 | 82.2 |
| PS weighting, using outpatient visits as comparator | 62.7 | 68.0 | 85.4 | 68.8 | 76.4 | 81.2 |
| PS weighting, using random days as comparator | 59.3 | 65.1 | 77.8 | 66.3 | 74.4 | 80.4 |
| Per−month PS matching, using outpatient visits as comparator | 64.9 | 70.3 | 76.5 | 69.3 | 73.6 | 75.3 |
| Per−month PS matching, using random days as comparator | 61.1 | 66.9 | 74.3 | 70.9 | 75.5 | 78.2 |
| Historical comparator | | | | | | |
| Unadjusted, using entire historic period | 58.7 | 64.9 | 79.7 | 70.0 | 78.0 | 84.1 |
| Age & sex adjusted, using entire historic period | 63.1 | 70.2 | 77.1 | 73.1 | 79.2 | 82.5 |
| Unadjusted, using TaR after historic visit | 57.4 | 62.9 | 76.1 | 67.9 | 76.8 | 84.2 |
| Age & sex adjusted, using TaR after historic visit | 63.6 | 70.3 | 79.6 | 73.6 | 80.4 | 83.4 |
| Unadjusted, using entire historic period, filtered | 58.7 | 65.0 | 79.9 | 70.5 | 78.1 | 83.9 |
| Age & sex adjusted, using entire historic period, filtered | 63.2 | 70.5 | 77.3 | 73.0 | 78.8 | 81.6 |
| Unadjusted, using TaR after historic visit, filtered | 57.4 | 62.9 | 76.2 | 67.7 | 76.6 | 83.6 |
| Age & sex adjusted, using TaR after historic visit, filtered | 63.8 | 70.5 | 80.0 | 73.1 | 80.0 | 82.7 |
| SCCS / SCRI | | | | | | |
| Unadjusted SCCS excluding pre−vaccination window | 66.2 | 74.2 | 80.7 | 74.7 | 79.2 | 81.4 |
| Age & season adjusted SCCS excluding pre−vaccination window | 66.7 | 73.9 | 79.2 | 75.1 | 78.4 | 80.7 |
| SCRI with prior control interval | 62.8 | 68.4 | 75.1 | 69.9 | 74.9 | 77.2 |
| SCRI with posterior control interval | 61.3 | 70.0 | 77.9 | 73.4 | 78.3 | 80.4 |
| Unadjusted SCCS excluding all pre−vaccination time | 65.9 | 73.3 | 81.3 | 75.5 | 80.5 | 82.3 |

Figure 67: The area under the receiver operator curve (AUC) based on the effect-size estimate at the end of the study period in the MDCD database for H1N1 vaccination.

| | Uncalibrated AUC when true effect is 1.5 | 2 | 4 | Calibrated AUC when true effect is 1.5 | 2 | 4 |
|---|---|---|---|---|---|---|
| Case−control | | | | | | |
| Age & sex matched controls | 54.6 | 57.1 | 60.5 | 61.4 | 65.6 | 67.2 |
| Age & sex adjusted, using random controls | 54.7 | 58.9 | 63.0 | 59.1 | 65.5 | 68.7 |
| Cohort method | | | | | | |
| Unadjusted, using outpatient visits as comparator | 57.6 | 61.5 | 72.6 | 66.3 | 69.1 | 71.5 |
| PS matching, using outpatient visits as comparator | 57.2 | 59.9 | 64.1 | 56.6 | 59.8 | 61.0 |
| Unadjusted, using random days as comparator | 54.0 | 55.5 | 60.5 | 56.6 | 60.5 | 62.9 |
| PS matching, using random days as comparator | 56.4 | 57.9 | 63.7 | 58.5 | 64.0 | 65.3 |
| PS stratification, using outpatient visits as comparator | 59.2 | 62.4 | 76.5 | 61.9 | 65.3 | 69.1 |
| PS stratification, using random days as comparator | 56.4 | 57.5 | 69.3 | 58.8 | 63.0 | 66.7 |
| PS weighting, using outpatient visits as comparator | 59.8 | 64.5 | 81.2 | 63.8 | 67.7 | 74.2 |
| PS weighting, using random days as comparator | 55.9 | 58.0 | 70.3 | 58.4 | 61.6 | 65.7 |
| Per−month PS matching, using outpatient visits as comparator | 55.7 | 57.6 | 60.2 | 55.5 | 57.9 | 60.2 |
| Per−month PS matching, using random days as comparator | 58.8 | 61.7 | 67.5 | 60.6 | 61.9 | 62.1 |
| Historical comparator | | | | | | |
| Unadjusted, using entire historic period | 53.9 | 56.8 | 61.6 | 63.2 | 67.9 | 72.3 |
| Age & sex adjusted, using entire historic period | 54.0 | 57.4 | 61.6 | 63.2 | 68.6 | 72.3 |
| Unadjusted, using TaR after historic visit | 55.8 | 58.0 | 63.3 | 65.5 | 70.2 | 73.8 |
| Age & sex adjusted, using TaR after historic visit | 54.7 | 56.9 | 62.2 | 64.9 | 69.7 | 73.3 |
| Unadjusted, using entire historic period, filtered | 53.9 | 56.8 | 61.6 | 63.3 | 68.1 | 72.3 |
| Age & sex adjusted, using entire historic period, filtered | 54.0 | 57.4 | 61.7 | 63.4 | 68.8 | 72.3 |
| Unadjusted, using TaR after historic visit, filtered | 55.8 | 58.0 | 63.4 | 65.1 | 69.9 | 73.4 |
| Age & sex adjusted, using TaR after historic visit, filtered | 54.7 | 56.9 | 62.3 | 63.9 | 69.3 | 72.2 |
| SCCS / SCRI | | | | | | |
| Unadjusted SCCS excluding pre−vaccination window | 59.9 | 62.2 | 67.5 | 64.6 | 69.3 | 71.4 |
| Age & season adjusted SCCS excluding pre−vaccination window | 58.9 | 61.2 | 67.3 | 64.7 | 70.1 | 71.5 |
| SCRI with prior control interval | 54.0 | 55.2 | 59.6 | 65.1 | 69.3 | 70.2 |
| SCRI with posterior control interval | 55.3 | 59.3 | 65.7 | 60.5 | 65.7 | 66.7 |
| Unadjusted SCCS excluding all pre−vaccination time | 58.9 | 61.7 | 66.9 | 64.2 | 68.7 | 71.1 |

Figure 68: The area under the receiver operator curve (AUC) based on the effect-size estimate at the end of the study period in the MDCR database for H1N1 vaccination.

| | Uncalibrated AUC when true effect is 1.5 | 2 | 4 | Calibrated AUC when true effect is 1.5 | 2 | 4 |
|---|---|---|---|---|---|---|
| Case−control | | | | | | |
| Age & sex matched controls | 58.2 | 63.7 | 70.6 | 69.2 | 76.8 | 79.4 |
| Age & sex adjusted, using random controls | 61.8 | 67.0 | 74.4 | 72.2 | 79.4 | 81.6 |
| Cohort method | | | | | | |
| Unadjusted, using outpatient visits as comparator | 61.3 | 68.4 | 82.6 | 74.2 | 83.2 | 90.1 |
| PS matching, using outpatient visits as comparator | 70.4 | 79.6 | 90.2 | 78.6 | 83.7 | 88.3 |
| Unadjusted, using random days as comparator | 69.2 | 76.2 | 85.9 | 79.0 | 87.0 | 90.2 |
| PS matching, using random days as comparator | 70.5 | 78.5 | 87.0 | 81.0 | 87.0 | 90.9 |
| PS stratification, using outpatient visits as comparator | 71.4 | 83.1 | 92.8 | 83.4 | 88.4 | 91.5 |
| PS stratification, using random days as comparator | 69.3 | 77.8 | 87.3 | 81.2 | 87.9 | 91.0 |
| PS weighting, using outpatient visits as comparator | 63.8 | 73.5 | 89.0 | 75.4 | 85.7 | 91.0 |
| PS weighting, using random days as comparator | 66.8 | 77.2 | 88.0 | 78.3 | 85.2 | 89.8 |
| Per−month PS matching, using outpatient visits as comparator | 68.6 | 77.8 | 87.4 | 79.7 | 85.3 | 89.2 |
| Per−month PS matching, using random days as comparator | 69.8 | 78.2 | 84.8 | 80.8 | 85.4 | 88.6 |
| Historical comparator | | | | | | |
| Unadjusted, using entire historic period | 71.8 | 79.5 | 87.7 | 84.1 | 90.0 | 91.7 |
| Age & sex adjusted, using entire historic period | 70.4 | 77.4 | 84.2 | 83.7 | 88.6 | 90.0 |
| Unadjusted, using TaR after historic visit | 68.9 | 80.5 | 91.9 | 82.1 | 89.0 | 91.6 |
| Age & sex adjusted, using TaR after historic visit | 74.8 | 83.9 | 90.7 | 85.9 | 89.8 | 91.1 |
| Unadjusted, using entire historic period, filtered | 71.9 | 79.4 | 87.2 | 84.0 | 89.6 | 91.3 |
| Age & sex adjusted, using entire historic period, filtered | 70.4 | 77.2 | 83.6 | 83.6 | 88.2 | 89.5 |
| Unadjusted, using TaR after historic visit, filtered | 68.8 | 80.5 | 91.7 | 82.0 | 88.6 | 91.3 |
| Age & sex adjusted, using TaR after historic visit, filtered | 75.0 | 84.0 | 90.4 | 85.8 | 89.5 | 90.7 |
| SCCS / SCRI | | | | | | |
| Unadjusted SCCS excluding pre−vaccination window | 73.5 | 81.0 | 89.0 | 83.0 | 88.8 | 89.8 |
| Age & season adjusted SCCS excluding pre−vaccination window | 73.0 | 80.5 | 87.5 | 82.9 | 87.7 | 88.7 |
| SCRI with prior control interval | 73.0 | 81.1 | 87.3 | 82.7 | 87.3 | 90.1 |
| SCRI with posterior control interval | 68.5 | 76.2 | 84.5 | 80.3 | 86.6 | 89.3 |
| Unadjusted SCCS excluding all pre−vaccination time | 72.9 | 80.4 | 88.0 | 84.5 | 89.9 | 90.8 |

Figure 69: The area under the receiver operator curve (AUC) based on the effect-size estimate at the end of the study period in the CCAE database for H1N1 vaccination.

#### 12.2 AUC for Seasonal flu vaccination (Fluvirin)

| | Uncalibrated AUC when true effect is 1.5 | Uncalibrated 2 | Uncalibrated 4 | Calibrated AUC when true effect is 1.5 | Calibrated 2 | Calibrated 4 |
|---|---|---|---|---|---|---|
| Case−control | | | | | | |
| Age & sex matched controls | 50.4 | 51.6 | 52.1 | 55.9 | 56.3 | 57.6 |
| Age & sex adjusted, using random controls | 50.3 | 50.4 | 50.7 | 56.1 | 58.2 | 58.3 |
| Cohort method | | | | | | |
| Unadjusted, using outpatient visits as comparator | 57.9 | 60.8 | 70.1 | 61.2 | 66.8 | 69.8 |
| PS matching, using outpatient visits as comparator | 55.3 | 56.3 | 59.3 | 54.3 | 55.4 | 56.6 |
| Unadjusted, using random days as comparator | 59.4 | 60.3 | 62.4 | 64.4 | 66.1 | 66.6 |
| PS matching, using random days as comparator | | | | | | |
| PS stratification, using outpatient visits as comparator | 66.5 | 71.6 | 88.8 | 67.5 | 69.6 | 75.0 |
| PS stratification, using random days as comparator | 57.8 | 60.6 | 74.1 | 62.3 | 67.0 | 70.1 |
| PS weighting, using outpatient visits as comparator | 66.8 | 72.9 | 89.5 | 63.9 | 70.9 | 74.3 |
| PS weighting, using random days as comparator | 50.0 | 50.0 | 50.0 | 50.0 | 50.0 | 50.0 |
| Per−month PS matching, using outpatient visits as comparator | | | | | | |
| Per−month PS matching, using random days as comparator | | | | | | |
| Historical comparator | | | | | | |
| Unadjusted, using entire historic period | 53.4 | 55.5 | 59.3 | 66.3 | 71.7 | 74.4 |
| Age & sex adjusted, using entire historic period | 53.3 | 55.2 | 59.3 | 65.8 | 71.8 | 74.5 |
| Unadjusted, using TaR after historic visit | 57.3 | 60.4 | 64.4 | 65.0 | 71.7 | 74.3 |
| Age & sex adjusted, using TaR after historic visit | 56.5 | 60.0 | 64.1 | 63.7 | 69.8 | 72.5 |
| Unadjusted, using entire historic period, filtered | 53.4 | 55.4 | 59.0 | 65.8 | 70.0 | 72.5 |
| Age & sex adjusted, using entire historic period, filtered | 53.3 | 55.1 | 59.1 | 65.8 | 71.2 | 73.7 |
| Unadjusted, using TaR after historic visit, filtered | 57.3 | 60.4 | 64.2 | 66.2 | 71.7 | 74.1 |
| Age & sex adjusted, using TaR after historic visit, filtered | 56.5 | 60.0 | 63.9 | 64.3 | 70.3 | 72.9 |
| SCCS / SCRI | | | | | | |
| Unadjusted SCCS excluding pre−vaccination window | 55.8 | 58.7 | 63.5 | 61.8 | 65.5 | 67.1 |
| Age & season adjusted SCCS excluding pre−vaccination window | 56.0 | 60.1 | 63.0 | 60.9 | 64.6 | 66.0 |
| SCRI with prior control interval | 55.7 | 57.1 | 58.8 | 58.3 | 59.9 | 61.0 |
| SCRI with posterior control interval | 54.2 | 55.2 | 58.6 | 62.7 | 65.4 | 66.7 |
| Unadjusted SCCS excluding all pre−vaccination time | 55.9 | 58.6 | 61.5 | 60.4 | 63.5 | 65.1 |

Figure 70: The area under the receiver operator curve (AUC) based on the effect-size estimate at the end of the study period in the Optum EHR database for Seasonal flu vaccination (Fluvirin).

| | Uncalibrated AUC when true effect is | | | Calibrated AUC when true effect is | | |
|---|---|---|---|---|---|---|
| | 1.5 | 2 | 4 | 1.5 | 2 | 4 |
| Case−control | | | | | | |
| Age & sex matched controls | 55.8 | 57.3 | 59.2 | 59.8 | 61.5 | 63.0 |
| Age & sex adjusted, using random controls | 54.0 | 55.9 | 58.2 | 62.1 | 64.7 | 66.3 |
| Cohort method | | | | | | |
| Unadjusted, using outpatient visits as comparator | 58.0 | 60.9 | 67.7 | 61.2 | 64.0 | 65.4 |
| PS matching, using outpatient visits as comparator | 50.0 | 50.0 | 50.0 | 50.0 | 50.0 | 50.0 |
| Unadjusted, using random days as comparator | 55.4 | 55.7 | 59.1 | 57.8 | 59.3 | 61.8 |
| PS matching, using random days as comparator | 50.0 | 50.0 | 50.0 | 50.0 | 50.0 | 50.0 |
| PS stratification, using outpatient visits as comparator | 64.1 | 70.1 | 87.6 | 63.3 | 66.9 | 70.3 |
| PS stratification, using random days as comparator | 59.0 | 60.4 | 75.2 | 63.1 | 65.4 | 68.9 |
| PS weighting, using outpatient visits as comparator | 65.8 | 71.0 | 90.2 | 64.0 | 67.6 | 71.0 |
| PS weighting, using random days as comparator | 50.0 | 50.0 | 50.0 | 50.0 | 50.0 | 50.0 |
| Per−month PS matching, using outpatient visits as comparator | | | | | | |
| Per−month PS matching, using random days as comparator | | | | | | |
| Historical comparator | | | | | | |
| Unadjusted, using entire historic period | 54.5 | 56.6 | 59.6 | 63.0 | 66.8 | 69.0 |
| Age & sex adjusted, using entire historic period | 56.1 | 59.0 | 62.5 | 66.9 | 71.7 | 72.3 |
| Unadjusted, using TaR after historic visit | 56.0 | 58.8 | 63.0 | 62.6 | 68.3 | 69.6 |
| Age & sex adjusted, using TaR after historic visit | 60.0 | 64.7 | 69.6 | 66.5 | 70.9 | 72.4 |
| Unadjusted, using entire historic period, filtered | 54.5 | 56.6 | 59.6 | 62.5 | 66.3 | 68.5 |
| Age & sex adjusted, using entire historic period, filtered | 56.1 | 59.1 | 62.6 | 64.4 | 68.0 | 68.5 |
| Unadjusted, using TaR after historic visit, filtered | 56.1 | 58.9 | 63.1 | 63.4 | 68.5 | 69.2 |
| Age & sex adjusted, using TaR after historic visit, filtered | 60.2 | 64.9 | 69.8 | 64.7 | 69.8 | 70.5 |
| SCCS / SCRI | | | | | | |
| Unadjusted SCCS excluding pre−vaccination window | 57.1 | 59.9 | 65.3 | 62.4 | 66.6 | 68.0 |
| Age & season adjusted SCCS excluding pre−vaccination window | 57.2 | 59.9 | 65.3 | 61.5 | 65.7 | 67.1 |
| SCRI with prior control interval | 54.7 | 57.5 | 62.2 | 58.1 | 60.5 | 62.7 |
| SCRI with posterior control interval | 53.8 | 57.8 | 59.1 | 66.4 | 67.4 | 70.4 |
| Unadjusted SCCS excluding all pre−vaccination time | 57.1 | 60.6 | 66.1 | 63.1 | 67.4 | 68.9 |

Figure 71: The area under the receiver operator curve (AUC) based on the effect-size estimate at the end of the study period in the MDCD database for Seasonal flu vaccination (Fluvirin).

| | Uncalibrated AUC when true effect is | | | Calibrated AUC when true effect is | | |
|---|---|---|---|---|---|---|
| | 1.5 | 2 | 4 | 1.5 | 2 | 4 |
| Case−control | | | | | | |
| Age & sex matched controls | 51.1 | 51.1 | 51.1 | 50.0 | 50.0 | 50.0 |
| Age & sex adjusted, using random controls | 50.1 | 50.1 | 50.1 | 50.0 | 50.0 | 50.0 |
| Cohort method | | | | | | |
| Unadjusted, using outpatient visits as comparator | 52.9 | 52.9 | 55.0 | 50.0 | 50.0 | 50.0 |
| PS matching, using outpatient visits as comparator | 50.3 | 50.3 | 50.3 | 50.0 | 50.0 | 50.0 |
| Unadjusted, using random days as comparator | 53.1 | 53.1 | 54.2 | 50.0 | 50.0 | 50.0 |
| PS matching, using random days as comparator | 50.1 | 50.1 | 50.1 | 50.0 | 50.0 | 50.0 |
| PS stratification, using outpatient visits as comparator | 55.6 | 57.3 | 76.8 | 52.1 | 54.2 | 54.2 |
| PS stratification, using random days as comparator | 51.1 | 51.2 | 61.9 | 50.0 | 50.0 | 50.0 |
| PS weighting, using outpatient visits as comparator | 57.0 | 57.4 | 79.1 | 51.1 | 53.1 | 55.2 |
| PS weighting, using random days as comparator | 51.3 | 51.5 | 65.2 | 50.0 | 50.0 | 50.0 |
| Per−month PS matching, using outpatient visits as comparator | | | | | | |
| Per−month PS matching, using random days as comparator | | | | | | |
| Historical comparator | | | | | | |
| Unadjusted, using entire historic period | 51.1 | 51.1 | 51.2 | 52.1 | 54.2 | 54.3 |
| Age & sex adjusted, using entire historic period | 51.1 | 51.1 | 51.2 | 53.1 | 55.1 | 55.3 |
| Unadjusted, using TaR after historic visit | 51.1 | 51.1 | 51.2 | 52.2 | 54.2 | 54.3 |
| Age & sex adjusted, using TaR after historic visit | 51.1 | 51.1 | 51.2 | 52.1 | 54.2 | 54.3 |
| Unadjusted, using entire historic period, filtered | 51.1 | 51.1 | 51.2 | 52.1 | 54.2 | 54.3 |
| Age & sex adjusted, using entire historic period, filtered | 51.1 | 51.1 | 51.2 | 53.1 | 55.1 | 55.3 |
| Unadjusted, using TaR after historic visit, filtered | 51.1 | 51.1 | 51.2 | 52.2 | 54.2 | 54.3 |
| Age & sex adjusted, using TaR after historic visit, filtered | 51.1 | 51.1 | 51.2 | 52.1 | 54.2 | 54.3 |
| SCCS / SCRI | | | | | | |
| Unadjusted SCCS excluding pre−vaccination window | 51.1 | 51.1 | 51.1 | 50.0 | 50.0 | 50.0 |
| Age & season adjusted SCCS excluding pre−vaccination window | 51.1 | 51.1 | 51.1 | 50.0 | 50.0 | 50.0 |
| SCRI with prior control interval | 50.0 | 50.0 | 51.1 | 50.0 | 50.0 | 50.0 |
| SCRI with posterior control interval | 50.0 | 51.1 | 51.1 | 50.0 | 50.0 | 50.0 |
| Unadjusted SCCS excluding all pre−vaccination time | 51.1 | 51.1 | 51.1 | 50.0 | 50.0 | 50.0 |

Figure 72: The area under the receiver operator curve (AUC) based on the effect-size estimate at the end of the study period in the MDCR database for Seasonal flu vaccination (Fluvirin).

| | Uncalibrated AUC when true effect is | | | Calibrated AUC when true effect is | | |
|---|---|---|---|---|---|---|
| | 1.5 | 2 | 4 | 1.5 | 2 | 4 |
| Case−control | | | | | | |
| Age & sex matched controls | 55.2 | 59.5 | 64.4 | 62.7 | 68.1 | 69.7 |
| Age & sex adjusted, using random controls | 59.4 | 64.1 | 69.7 | 67.5 | 72.0 | 73.8 |
| Cohort method | | | | | | |
| Unadjusted, using outpatient visits as comparator | 61.3 | 68.4 | 82.5 | 68.7 | 78.1 | 83.5 |
| PS matching, using outpatient visits as comparator | 60.8 | 66.8 | 74.6 | 70.1 | 74.7 | 77.1 |
| Unadjusted, using random days as comparator | 60.8 | 65.0 | 74.3 | 69.2 | 76.6 | 80.3 |
| PS matching, using random days as comparator | 62.6 | 67.9 | 78.4 | 76.3 | 81.3 | 83.6 |
| PS stratification, using outpatient visits as comparator | 64.0 | 72.2 | 89.3 | 71.6 | 79.1 | 84.2 |
| PS stratification, using random days as comparator | 63.5 | 71.4 | 86.7 | 76.1 | 82.3 | 83.4 |
| PS weighting, using outpatient visits as comparator | 64.4 | 71.6 | 87.7 | 66.8 | 75.2 | 83.3 |
| PS weighting, using random days as comparator | 63.9 | 70.0 | 86.5 | 76.1 | 82.1 | 83.8 |
| Per−month PS matching, using outpatient visits as comparator | | | | | | |
| Per−month PS matching, using random days as comparator | | | | | | |
| Historical comparator | | | | | | |
| Unadjusted, using entire historic period | 62.6 | 68.8 | 76.2 | 76.2 | 83.3 | 85.3 |
| Age & sex adjusted, using entire historic period | 64.9 | 71.7 | 79.7 | 75.8 | 83.4 | 85.6 |
| Unadjusted, using TaR after historic visit | 65.2 | 72.1 | 81.9 | 74.4 | 82.7 | 84.9 |
| Age & sex adjusted, using TaR after historic visit | 63.6 | 72.8 | 82.6 | 74.2 | 82.2 | 85.0 |
| Unadjusted, using entire historic period, filtered | 62.8 | 69.2 | 76.6 | 75.4 | 82.1 | 83.5 |
| Age & sex adjusted, using entire historic period, filtered | 65.2 | 72.2 | 80.3 | 76.0 | 82.6 | 83.9 |
| Unadjusted, using TaR after historic visit, filtered | 65.6 | 72.6 | 82.7 | 75.4 | 82.7 | 84.3 |
| Age & sex adjusted, using TaR after historic visit, filtered | 63.9 | 73.5 | 83.4 | 74.5 | 81.8 | 83.8 |
| SCCS / SCRI | | | | | | |
| Unadjusted SCCS excluding pre−vaccination window | 64.5 | 70.5 | 78.0 | 74.9 | 80.9 | 84.0 |
| Age & season adjusted SCCS excluding pre−vaccination window | 64.3 | 71.1 | 77.9 | 74.9 | 80.4 | 83.1 |
| SCRI with prior control interval | 59.1 | 63.3 | 69.8 | 68.4 | 74.3 | 77.1 |
| SCRI with posterior control interval | 58.2 | 64.9 | 71.3 | 69.5 | 75.0 | 79.2 |
| Unadjusted SCCS excluding all pre−vaccination time | 61.9 | 67.8 | 76.7 | 71.9 | 79.4 | 82.2 |

Figure 73: The area under the receiver operator curve (AUC) based on the effect-size estimate at the end of the study period in the CCAE database for Seasonal flu vaccination (Fluvirin).

#### 12.3 AUC for Seasonal flu vaccination (Fluzone)

| | Uncalibrated AUC when true effect is | | | Calibrated AUC when true effect is | | |
|---|---|---|---|---|---|---|
| | 1.5 | 2 | 4 | 1.5 | 2 | 4 |
| Case−control | | | | | | |
| Age & sex matched controls | 58.9 | 64.4 | 72.3 | 72.3 | 78.6 | 82.9 |
| Age & sex adjusted, using random controls | 58.9 | 65.4 | 75.1 | 73.4 | 82.3 | 87.6 |
| Cohort method | | | | | | |
| Unadjusted, using outpatient visits as comparator | 66.0 | 75.5 | 89.1 | 78.4 | 88.2 | 95.2 |
| PS matching, using outpatient visits as comparator | 66.5 | 74.6 | 87.5 | 76.7 | 86.0 | 90.6 |
| Unadjusted, using random days as comparator | 61.9 | 69.5 | 80.2 | 75.1 | 84.5 | 89.5 |
| PS matching, using random days as comparator | | | | | | |
| PS stratification, using outpatient visits as comparator | 69.5 | 80.6 | 94.7 | 83.6 | 92.2 | 96.1 |
| PS stratification, using random days as comparator | 65.6 | 74.2 | 88.4 | 79.3 | 89.6 | 94.6 |
| PS weighting, using outpatient visits as comparator | 67.1 | 76.8 | 93.0 | 77.1 | 87.6 | 94.1 |
| PS weighting, using random days as comparator | 50.3 | 50.4 | 50.6 | 50.0 | 50.0 | 50.0 |
| Per−month PS matching, using outpatient visits as comparator | | | | | | |
| Per−month PS matching, using random days as comparator | | | | | | |
| Historical comparator | | | | | | |
| Unadjusted, using entire historic period | 66.7 | 75.8 | 87.6 | 80.7 | 90.3 | 95.9 |
| Age & sex adjusted, using entire historic period | 66.6 | 76.0 | 87.7 | 80.7 | 90.2 | 96.0 |
| Unadjusted, using TaR after historic visit | 67.7 | 77.6 | 90.0 | 81.0 | 90.2 | 95.4 |
| Age & sex adjusted, using TaR after historic visit | 68.5 | 77.7 | 89.9 | 81.7 | 91.0 | 96.2 |
| Unadjusted, using entire historic period, filtered | 66.8 | 75.9 | 87.5 | 80.9 | 90.5 | 95.3 |
| Age & sex adjusted, using entire historic period, filtered | 66.7 | 76.1 | 87.7 | 81.0 | 90.5 | 95.6 |
| Unadjusted, using TaR after historic visit, filtered | 67.9 | 78.0 | 90.2 | 81.4 | 90.5 | 94.9 |
| Age & sex adjusted, using TaR after historic visit, filtered | 68.7 | 78.0 | 90.1 | 82.0 | 91.3 | 95.8 |
| SCCS / SCRI | | | | | | |
| Unadjusted SCCS excluding pre−vaccination window | 71.1 | 80.9 | 91.4 | 84.7 | 92.2 | 95.8 |
| Age & season adjusted SCCS excluding pre−vaccination window | 69.5 | 78.9 | 90.4 | 81.0 | 89.1 | 93.0 |
| SCRI with prior control interval | 64.2 | 72.7 | 85.2 | 78.0 | 88.4 | 93.3 |
| SCRI with posterior control interval | 67.7 | 75.9 | 85.9 | 81.5 | 89.6 | 92.5 |
| Unadjusted SCCS excluding all pre−vaccination time | 70.2 | 79.6 | 90.6 | 84.5 | 92.5 | 96.4 |

Figure 74: The area under the receiver operator curve (AUC) based on the effect-size estimate at the end of the study period in the Optum EHR database for Seasonal flu vaccination (Fluzone).

| | Uncalibrated AUC when true effect is | | | Calibrated AUC when true effect is | | |
|---|---|---|---|---|---|---|
| | 1.5 | 2 | 4 | 1.5 | 2 | 4 |
| Case−control | | | | | | |
| Age & sex matched controls | 51.2 | 51.3 | 52.5 | 52.3 | 53.4 | 54.7 |
| Age & sex adjusted, using random controls | 51.1 | 51.1 | 51.2 | 53.1 | 54.2 | 54.3 |
| Cohort method | | | | | | |
| Unadjusted, using outpatient visits as comparator | 56.2 | 57.1 | 58.3 | 58.1 | 59.2 | 59.2 |
| PS matching, using outpatient visits as comparator | 52.2 | 52.4 | 54.5 | 52.2 | 53.2 | 53.3 |
| Unadjusted, using random days as comparator | 52.1 | 52.1 | 54.2 | 53.2 | 55.2 | 55.3 |
| PS matching, using random days as comparator | | | | | | |
| PS stratification, using outpatient visits as comparator | 57.4 | 59.7 | 77.5 | 52.5 | 54.6 | 58.8 |
| PS stratification, using random days as comparator | 54.6 | 55.8 | 70.7 | 58.9 | 61.1 | 61.4 |
| PS weighting, using outpatient visits as comparator | 59.7 | 60.3 | 80.4 | 53.4 | 57.4 | 60.6 |
| PS weighting, using random days as comparator | 50.0 | 50.0 | 50.0 | 50.0 | 50.0 | 50.0 |
| Per−month PS matching, using outpatient visits as comparator | | | | | | |
| Per−month PS matching, using random days as comparator | | | | | | |
| Historical comparator | | | | | | |
| Unadjusted, using entire historic period | 51.4 | 51.8 | 52.6 | 56.8 | 59.9 | 63.3 |
| Age & sex adjusted, using entire historic period | 51.7 | 52.0 | 53.6 | 58.6 | 60.8 | 63.5 |
| Unadjusted, using TaR after historic visit | 52.2 | 52.6 | 53.5 | 56.0 | 59.1 | 62.5 |
| Age & sex adjusted, using TaR after historic visit | 51.6 | 53.6 | 55.1 | 57.8 | 61.6 | 63.5 |
| Unadjusted, using entire historic period, filtered | 51.4 | 51.8 | 52.5 | 57.7 | 59.9 | 63.2 |
| Age & sex adjusted, using entire historic period, filtered | 51.7 | 52.0 | 53.5 | 58.6 | 61.7 | 63.4 |
| Unadjusted, using TaR after historic visit, filtered | 52.2 | 52.6 | 53.4 | 56.8 | 60.0 | 62.5 |
| Age & sex adjusted, using TaR after historic visit, filtered | 51.5 | 53.6 | 55.1 | 57.8 | 60.8 | 62.6 |
| SCCS / SCRI | | | | | | |
| Unadjusted SCCS excluding pre−vaccination window | 54.0 | 55.1 | 57.6 | 58.7 | 61.0 | 62.3 |
| Age & season adjusted SCCS excluding pre−vaccination window | 53.2 | 54.3 | 56.7 | 57.9 | 60.1 | 61.5 |
| SCRI with prior control interval | 55.7 | 55.9 | 56.0 | 56.1 | 56.2 | 59.3 |
| SCRI with posterior control interval | 51.4 | 54.4 | 55.6 | 59.0 | 59.3 | 60.3 |
| Unadjusted SCCS excluding all pre−vaccination time | 53.2 | 56.1 | 57.5 | 57.9 | 60.1 | 61.5 |

Figure 75: The area under the receiver operator curve (AUC) based on the effect-size estimate at the end of the study period in the MDCD database for Seasonal flu vaccination (Fluzone).

| | Uncalibrated AUC when true effect is 1.5 | 2 | 4 | Calibrated AUC when true effect is 1.5 | 2 | 4 |
|---|---|---|---|---|---|---|
| Case−control | | | | | | |
| Age & sex matched controls | 58.8 | 66.8 | 75.1 | 68.0 | 71.1 | 73.5 |
| Age & sex adjusted, using random controls | 60.4 | 66.2 | 76.5 | 70.1 | 75.2 | 79.8 |
| Cohort method | | | | | | |
| Unadjusted, using outpatient visits as comparator | 59.5 | 64.8 | 77.8 | 67.7 | 75.5 | 80.0 |
| PS matching, using outpatient visits as comparator | 59.8 | 64.2 | 73.8 | 64.2 | 69.3 | 70.9 |
| Unadjusted, using random days as comparator | 59.7 | 64.5 | 75.7 | 64.3 | 72.3 | 74.4 |
| PS matching, using random days as comparator | 62.6 | 67.3 | 75.0 | 65.4 | 70.1 | 73.3 |
| PS stratification, using outpatient visits as comparator | 63.8 | 72.1 | 88.3 | 71.5 | 77.6 | 82.3 |
| PS stratification, using random days as comparator | 59.9 | 66.8 | 81.2 | 68.0 | 74.3 | 79.1 |
| PS weighting, using outpatient visits as comparator | 63.9 | 72.1 | 89.4 | 63.9 | 72.1 | 77.5 |
| PS weighting, using random days as comparator | 60.1 | 65.2 | 80.0 | 66.0 | 71.6 | 77.8 |
| Per−month PS matching, using outpatient visits as comparator | | | | | | |
| Per−month PS matching, using random days as comparator | | | | | | |
| Historical comparator | | | | | | |
| Unadjusted, using entire historic period | 62.5 | 68.3 | 76.3 | 70.6 | 77.5 | 82.5 |
| Age & sex adjusted, using entire historic period | 61.4 | 67.9 | 75.9 | 71.6 | 77.9 | 83.0 |
| Unadjusted, using TaR after historic visit | 60.8 | 68.1 | 78.1 | 70.4 | 77.3 | 82.5 |
| Age & sex adjusted, using TaR after historic visit | 61.0 | 68.3 | 77.9 | 71.0 | 77.5 | 82.5 |
| Unadjusted, using entire historic period, filtered | 62.8 | 68.7 | 76.7 | 71.5 | 76.7 | 80.9 |
| Age & sex adjusted, using entire historic period, filtered | 61.6 | 68.3 | 76.3 | 70.4 | 75.9 | 80.1 |
| Unadjusted, using TaR after historic visit, filtered | 61.1 | 68.5 | 78.7 | 70.9 | 76.5 | 81.1 |
| Age & sex adjusted, using TaR after historic visit, filtered | 61.3 | 68.8 | 78.4 | 71.5 | 77.2 | 81.6 |
| SCCS / SCRI | | | | | | |
| Unadjusted SCCS excluding pre−vaccination window | 64.3 | 70.6 | 76.7 | 71.9 | 76.2 | 79.5 |
| Age & season adjusted SCCS excluding pre−vaccination window | 63.0 | 69.3 | 75.8 | 70.2 | 74.8 | 77.8 |
| SCRI with prior control interval | 55.3 | 59.9 | 66.6 | 62.1 | 66.7 | 69.5 |
| SCRI with posterior control interval | 57.4 | 62.9 | 68.7 | 63.7 | 68.3 | 72.7 |
| Unadjusted SCCS excluding all pre−vaccination time | 64.7 | 70.3 | 78.0 | 71.6 | 77.1 | 80.0 |

Figure 76: The area under the receiver operator curve (AUC) based on the effect-size estimate at the end of the study period in the MDCR database for Seasonal flu vaccination (Fluzone).

| | Uncalibrated AUC when true effect is | | | Calibrated AUC when true effect is | | |
|---|---|---|---|---|---|---|
| | 1.5 | 2 | 4 | 1.5 | 2 | 4 |
| Case−control | | | | | | |
| Age & sex matched controls | 50.0 | 50.0 | 50.0 | 50.0 | 50.0 | 50.0 |
| Age & sex adjusted, using random controls | 50.0 | 50.0 | 50.0 | 50.0 | 50.0 | 50.0 |
| Cohort method | | | | | | |
| Unadjusted, using outpatient visits as comparator | 50.3 | 50.3 | 50.3 | 50.0 | 50.0 | 50.0 |
| PS matching, using outpatient visits as comparator | 51.1 | 52.1 | 52.2 | 50.0 | 50.0 | 50.0 |
| Unadjusted, using random days as comparator | 50.1 | 50.1 | 50.1 | 50.0 | 50.0 | 50.0 |
| PS matching, using random days as comparator | 52.2 | 52.2 | 54.3 | 50.0 | 50.0 | 50.0 |
| PS stratification, using outpatient visits as comparator | 53.1 | 54.3 | 69.1 | 50.1 | 52.2 | 53.3 |
| PS stratification, using random days as comparator | 51.6 | 52.8 | 59.2 | 50.0 | 50.0 | 50.0 |
| PS weighting, using outpatient visits as comparator | 53.7 | 53.7 | 72.2 | 52.1 | 53.2 | 54.2 |
| PS weighting, using random days as comparator | 51.2 | 51.2 | 63.4 | 50.0 | 50.0 | 50.0 |
| Per−month PS matching, using outpatient visits as comparator | | | | | | |
| Per−month PS matching, using random days as comparator | | | | | | |
| Historical comparator | | | | | | |
| Unadjusted, using entire historic period | 50.1 | 50.2 | 50.3 | 53.1 | 53.1 | 56.2 |
| Age & sex adjusted, using entire historic period | 51.1 | 51.2 | 51.3 | 52.1 | 53.2 | 55.3 |
| Unadjusted, using TaR after historic visit | 50.1 | 50.2 | 50.3 | 51.2 | 52.2 | 55.3 |
| Age & sex adjusted, using TaR after historic visit | 51.1 | 51.2 | 51.3 | 52.2 | 53.2 | 55.3 |
| Unadjusted, using entire historic period, filtered | 50.1 | 50.2 | 50.2 | 52.1 | 52.1 | 55.2 |
| Age & sex adjusted, using entire historic period, filtered | 51.1 | 51.1 | 51.3 | 52.1 | 54.1 | 55.3 |
| Unadjusted, using TaR after historic visit, filtered | 50.1 | 50.2 | 50.3 | 51.1 | 52.2 | 55.2 |
| Age & sex adjusted, using TaR after historic visit, filtered | 51.1 | 51.1 | 51.3 | 52.1 | 53.2 | 55.3 |
| SCCS / SCRI | | | | | | |
| Unadjusted SCCS excluding pre−vaccination window | 50.0 | 50.0 | 50.1 | 50.0 | 50.0 | 50.0 |
| Age & season adjusted SCCS excluding pre−vaccination window | 50.0 | 50.0 | 50.1 | 50.0 | 50.0 | 50.0 |
| SCRI with prior control interval | | | | | | |
| SCRI with posterior control interval | | | | | | |
| Unadjusted SCCS excluding all pre−vaccination time | 50.0 | 50.0 | 50.1 | 50.0 | 50.0 | 50.0 |

Figure 77: The area under the receiver operator curve (AUC) based on the effect-size estimate at the end of the study period in the CCAE database for Seasonal flu vaccination (Fluzone).

#### 12.4 AUC for Seasonal flu vaccination (All)

| | Uncalibrated AUC when true effect is | | | Calibrated AUC when true effect is | | |
|---|---|---|---|---|---|---|
| | 1.5 | 2 | 4 | 1.5 | 2 | 4 |
| Case−control | | | | | | |
| Age & sex matched controls | 76.1 | 86.9 | 94.1 | 91.9 | 96.9 | 97.6 |
| Age & sex adjusted, using random controls | 80.8 | 89.9 | 95.1 | 93.7 | 97.4 | 98.2 |
| Cohort method | | | | | | |
| Unadjusted, using outpatient visits as comparator | 75.0 | 86.8 | 98.1 | 90.8 | 98.3 | 99.5 |
| PS matching, using outpatient visits as comparator | 77.3 | 88.1 | 96.3 | 91.2 | 96.4 | 98.4 |
| Unadjusted, using random days as comparator | 73.7 | 85.3 | 95.5 | 89.7 | 96.8 | 98.2 |
| PS matching, using random days as comparator | | | | | | |
| PS stratification, using outpatient visits as comparator | 79.5 | 90.3 | 97.0 | 93.2 | 97.3 | 99.5 |
| PS stratification, using random days as comparator | 78.1 | 90.6 | 98.6 | 94.2 | 99.0 | 99.4 |
| PS weighting, using outpatient visits as comparator | 76.1 | 87.0 | 97.0 | 89.9 | 96.7 | 99.2 |
| PS weighting, using random days as comparator | 50.7 | 51.1 | 51.8 | 50.0 | 50.0 | 50.0 |
| Per−month PS matching, using outpatient visits as comparator | | | | | | |
| Per−month PS matching, using random days as comparator | | | | | | |
| Historical comparator | | | | | | |
| Unadjusted, using entire historic period | 79.3 | 91.2 | 98.6 | 94.4 | 98.6 | 100.0 |
| Age & sex adjusted, using entire historic period | 84.5 | 94.1 | 98.6 | 95.9 | 98.6 | 99.9 |
| Unadjusted, using TaR after historic visit | 79.5 | 90.7 | 98.4 | 93.2 | 98.4 | 100.0 |
| Age & sex adjusted, using TaR after historic visit | 83.5 | 94.0 | 98.8 | 95.7 | 98.8 | 100.0 |
| Unadjusted, using entire historic period, filtered | 79.3 | 91.2 | 98.6 | 94.9 | 99.1 | 99.4 |
| Age & sex adjusted, using entire historic period, filtered | 84.5 | 94.1 | 98.6 | 96.4 | 99.1 | 99.4 |
| Unadjusted, using TaR after historic visit, filtered | 79.8 | 91.0 | 98.7 | 93.7 | 98.9 | 99.5 |
| Age & sex adjusted, using TaR after historic visit, filtered | 83.7 | 94.2 | 99.0 | 96.2 | 99.3 | 99.5 |
| SCCS / SCRI | | | | | | |
| Unadjusted SCCS excluding pre−vaccination window | 84.5 | 95.4 | 99.4 | 97.2 | 99.4 | 100.0 |
| Age & season adjusted SCCS excluding pre−vaccination window | 80.7 | 92.7 | 98.5 | 92.3 | 95.5 | 96.2 |
| SCRI with prior control interval | 79.7 | 87.4 | 95.4 | 89.6 | 95.8 | 98.3 |
| SCRI with posterior control interval | 81.0 | 90.7 | 97.7 | 93.2 | 98.0 | 99.5 |
| Unadjusted SCCS excluding all pre−vaccination time | 85.7 | 94.2 | 99.6 | 95.9 | 99.6 | 100.0 |

Figure 78: The area under the receiver operator curve (AUC) based on the effect-size estimate at the end of the study period in the Optum EHR database for Seasonal flu vaccination (All).

| | Uncalibrated AUC when true effect is | | | Calibrated AUC when true effect is | | |
|---|---|---|---|---|---|---|
| | 1.5 | 2 | 4 | 1.5 | 2 | 4 |
| Case−control | | | | | | |
| Age & sex matched controls | 68.1 | 75.2 | 80.0 | 84.3 | 87.8 | 89.3 |
| Age & sex adjusted, using random controls | 73.8 | 82.4 | 88.8 | 89.6 | 93.7 | 94.0 |
| Cohort method | | | | | | |
| Unadjusted, using outpatient visits as comparator | 67.8 | 78.1 | 93.7 | 82.3 | 91.1 | 94.0 |
| PS matching, using outpatient visits as comparator | 83.5 | 92.0 | 97.1 | 91.0 | 93.7 | 93.8 |
| Unadjusted, using random days as comparator | 69.9 | 78.7 | 86.1 | 85.5 | 90.6 | 92.7 |
| PS matching, using random days as comparator | 75.3 | 85.5 | 94.2 | 87.5 | 92.9 | 93.6 |
| PS stratification, using outpatient visits as comparator | 78.8 | 88.2 | 95.9 | 87.8 | 92.4 | 93.9 |
| PS stratification, using random days as comparator | 71.1 | 81.5 | 91.7 | 84.9 | 91.4 | 93.3 |
| PS weighting, using outpatient visits as comparator | 77.7 | 88.0 | 96.0 | 88.5 | 92.5 | 93.2 |
| PS weighting, using random days as comparator | 69.2 | 78.8 | 91.4 | 80.2 | 88.6 | 93.4 |
| Per−month PS matching, using outpatient visits as comparator | | | | | | |
| Per−month PS matching, using random days as comparator | | | | | | |
| Historical comparator | | | | | | |
| Unadjusted, using entire historic period | 74.5 | 83.1 | 88.1 | 90.2 | 93.6 | 94.1 |
| Age & sex adjusted, using entire historic period | 75.2 | 83.3 | 87.6 | 90.9 | 93.7 | 94.2 |
| Unadjusted, using TaR after historic visit | 75.3 | 85.7 | 93.5 | 88.3 | 93.4 | 93.9 |
| Age & sex adjusted, using TaR after historic visit | 77.0 | 86.5 | 92.8 | 89.3 | 93.6 | 93.9 |
| Unadjusted, using entire historic period, filtered | 74.7 | 83.2 | 88.3 | 90.1 | 93.4 | 93.8 |
| Age & sex adjusted, using entire historic period, filtered | 75.3 | 83.4 | 87.7 | 90.3 | 93.1 | 93.5 |
| Unadjusted, using TaR after historic visit, filtered | 75.7 | 86.2 | 94.1 | 88.4 | 93.5 | 93.9 |
| Age & sex adjusted, using TaR after historic visit, filtered | 77.4 | 87.0 | 93.3 | 89.4 | 93.7 | 93.9 |
| SCCS / SCRI | | | | | | |
| Unadjusted SCCS excluding pre−vaccination window | 82.7 | 89.8 | 93.9 | 91.0 | 93.7 | 93.8 |
| Age & season adjusted SCCS excluding pre−vaccination window | 81.3 | 88.3 | 92.6 | 88.9 | 91.6 | 91.8 |
| SCRI with prior control interval | 77.0 | 85.7 | 92.3 | 88.6 | 93.3 | 93.8 |
| SCRI with posterior control interval | 78.6 | 85.9 | 91.5 | 89.6 | 93.7 | 94.6 |
| Unadjusted SCCS excluding all pre−vaccination time | 79.2 | 87.3 | 93.0 | 89.6 | 93.6 | 93.9 |

Figure 79: The area under the receiver operator curve (AUC) based on the effect-size estimate at the end of the study period in the MDCD database for Seasonal flu vaccination (All).

| | Uncalibrated AUC when true effect is | | | Calibrated AUC when true effect is | | |
|---|---|---|---|---|---|---|
| | 1.5 | 2 | 4 | 1.5 | 2 | 4 |
| Case−control | | | | | | |
| Age & sex matched controls | 70.3 | 77.3 | 81.5 | 79.6 | 82.2 | 83.4 |
| Age & sex adjusted, using random controls | 75.0 | 84.2 | 88.7 | 89.1 | 91.6 | 92.3 |
| Cohort method | | | | | | |
| Unadjusted, using outpatient visits as comparator | 66.3 | 75.9 | 91.2 | 79.7 | 87.4 | 91.0 |
| PS matching, using outpatient visits as comparator | 67.3 | 76.4 | 87.0 | 78.6 | 85.9 | 88.2 |
| Unadjusted, using random days as comparator | 68.8 | 76.9 | 85.7 | 81.0 | 87.5 | 91.0 |
| PS matching, using random days as comparator | 70.0 | 77.7 | 88.2 | 78.7 | 87.2 | 89.6 |
| PS stratification, using outpatient visits as comparator | 69.0 | 79.2 | 92.6 | 80.8 | 87.7 | 91.5 |
| PS stratification, using random days as comparator | 69.0 | 77.9 | 87.8 | 79.9 | 87.1 | 89.3 |
| PS weighting, using outpatient visits as comparator | 66.7 | 76.4 | 92.2 | 78.0 | 86.2 | 90.9 |
| PS weighting, using random days as comparator | 65.1 | 72.6 | 85.4 | 75.4 | 84.5 | 89.8 |
| Per−month PS matching, using outpatient visits as comparator | | | | | | |
| Per−month PS matching, using random days as comparator | | | | | | |
| Historical comparator | | | | | | |
| Unadjusted, using entire historic period | 75.2 | 83.1 | 87.6 | 89.3 | 92.3 | 94.2 |
| Age & sex adjusted, using entire historic period | 75.0 | 82.9 | 87.4 | 89.3 | 92.3 | 94.2 |
| Unadjusted, using TaR after historic visit | 72.5 | 83.1 | 90.2 | 86.0 | 91.0 | 92.8 |
| Age & sex adjusted, using TaR after historic visit | 73.0 | 83.3 | 90.1 | 86.7 | 91.6 | 93.5 |
| Unadjusted, using entire historic period, filtered | 75.6 | 83.5 | 87.9 | 88.6 | 91.5 | 91.8 |
| Age & sex adjusted, using entire historic period, filtered | 75.2 | 83.2 | 87.6 | 88.8 | 91.7 | 91.9 |
| Unadjusted, using TaR after historic visit, filtered | 73.1 | 83.8 | 90.8 | 86.0 | 90.8 | 91.1 |
| Age & sex adjusted, using TaR after historic visit, filtered | 73.6 | 84.0 | 90.6 | 86.0 | 90.6 | 90.9 |
| SCCS / SCRI | | | | | | |
| Unadjusted SCCS excluding pre−vaccination window | 75.2 | 83.1 | 89.6 | 85.4 | 90.3 | 91.8 |
| Age & season adjusted SCCS excluding pre−vaccination window | 74.1 | 80.1 | 82.0 | 83.8 | 85.0 | 85.8 |
| SCRI with prior control interval | 66.5 | 75.1 | 83.5 | 81.2 | 87.9 | 89.6 |
| SCRI with posterior control interval | 71.7 | 79.8 | 86.6 | 83.8 | 88.6 | 90.6 |
| Unadjusted SCCS excluding all pre−vaccination time | 73.2 | 81.4 | 88.3 | 84.1 | 89.4 | 92.0 |

Figure 80: The area under the receiver operator curve (AUC) based on the effect-size estimate at the end of the study period in the MDCR database for Seasonal flu vaccination (All).

| | Uncalibrated AUC when true effect is | | | Calibrated AUC when true effect is | | |
|---|---|---|---|---|---|---|
| | 1.5 | 2 | 4 | 1.5 | 2 | 4 |
| **Case−control** | | | | | | |
| Age & sex matched controls | 72.3 | 81.4 | 88.3 | 86.1 | 91.1 | 92.5 |
| Age & sex adjusted, using random controls | 76.5 | 84.8 | 90.2 | 89.1 | 93.1 | 94.2 |
| **Cohort method** | | | | | | |
| Unadjusted, using outpatient visits as comparator | 65.6 | 75.1 | 91.2 | 80.1 | 88.9 | 94.3 |
| PS matching, using outpatient visits as comparator | 76.7 | 86.7 | 95.1 | 88.1 | 93.7 | 94.3 |
| Unadjusted, using random days as comparator | 73.3 | 83.1 | 89.8 | 88.2 | 92.3 | 93.7 |
| PS matching, using random days as comparator | 76.7 | 84.5 | 90.9 | 87.8 | 92.2 | 94.7 |
| PS stratification, using outpatient visits as comparator | 74.7 | 86.3 | 96.1 | 87.9 | 93.8 | 94.7 |
| PS stratification, using random days as comparator | 77.0 | 85.0 | 92.6 | 88.2 | 93.7 | 94.3 |
| PS weighting, using outpatient visits as comparator | 76.0 | 87.9 | 97.0 | 89.9 | 94.8 | 95.4 |
| PS weighting, using random days as comparator | 71.4 | 81.3 | 92.5 | 85.0 | 92.3 | 94.6 |
| Per−month PS matching, using outpatient visits as comparator | | | | | | |
| Per−month PS matching, using random days as comparator | | | | | | |
| **Historical comparator** | | | | | | |
| Unadjusted, using entire historic period | 76.9 | 84.4 | 89.9 | 88.5 | 92.4 | 94.6 |
| Age & sex adjusted, using entire historic period | 79.7 | 86.1 | 90.1 | 89.7 | 92.6 | 94.9 |
| Unadjusted, using TaR after historic visit | 75.4 | 86.4 | 93.8 | 88.4 | 93.0 | 95.5 |
| Age & sex adjusted, using TaR after historic visit | 78.0 | 88.1 | 93.5 | 89.8 | 92.6 | 95.0 |
| Unadjusted, using entire historic period, filtered | 77.1 | 84.8 | 90.2 | 88.9 | 92.7 | 93.8 |
| Age & sex adjusted, using entire historic period, filtered | 80.0 | 86.5 | 90.4 | 90.2 | 93.0 | 94.0 |
| Unadjusted, using TaR after historic visit, filtered | 76.2 | 87.6 | 95.0 | 88.7 | 93.3 | 94.6 |
| Age & sex adjusted, using TaR after historic visit, filtered | 78.9 | 89.3 | 94.7 | 90.1 | 93.0 | 94.1 |
| **SCCS / SCRI** | | | | | | |
| Unadjusted SCCS excluding pre−vaccination window | 84.7 | 89.6 | 92.6 | 91.7 | 93.6 | 94.6 |
| Age & season adjusted SCCS excluding pre−vaccination window | 83.8 | 88.0 | 91.2 | 87.4 | 89.6 | 90.6 |
| SCRI with prior control interval | 75.1 | 83.2 | 90.6 | 86.8 | 92.1 | 93.6 |
| SCRI with posterior control interval | 83.0 | 88.5 | 91.3 | 91.1 | 93.2 | 95.5 |
| Unadjusted SCCS excluding all pre−vaccination time | 84.6 | 88.6 | 91.8 | 91.0 | 93.2 | 95.0 |

Figure 81: The area under the receiver operator curve (AUC) based on the effect-size estimate at the end of the study period in the CCAE database for Seasonal flu vaccination (All).

#### 12.5 AUC for Zoster vaccination (Shingrix)

| | Uncalibrated AUC when true effect is | | | Calibrated AUC when true effect is | | |
|---|---|---|---|---|---|---|
| | 1.5 | 2 | 4 | 1.5 | 2 | 4 |
| Case−control | | | | | | |
| Age & sex matched controls | 58.6 | 63.1 | 71.0 | 69.7 | 76.0 | 81.7 |
| Age & sex adjusted, using random controls | 60.4 | 66.3 | 74.2 | 71.8 | 80.6 | 83.7 |
| Cohort method | | | | | | |
| Unadjusted, using outpatient visits as comparator | 63.3 | 71.0 | 85.6 | 73.7 | 84.0 | 93.0 |
| PS matching, using outpatient visits as comparator | 69.0 | 78.0 | 90.3 | 82.6 | 89.6 | 93.6 |
| Unadjusted, using random days as comparator | 61.2 | 67.8 | 77.5 | 72.3 | 80.5 | 88.8 |
| PS matching, using random days as comparator | | | | | | |
| PS stratification, using outpatient visits as comparator | 70.1 | 81.4 | 95.7 | 83.6 | 91.6 | 95.0 |
| PS stratification, using random days as comparator | | | | | | |
| PS weighting, using outpatient visits as comparator | 70.8 | 81.2 | 95.7 | 84.0 | 92.3 | 95.5 |
| PS weighting, using random days as comparator | | | | | | |
| Per−month PS matching, using outpatient visits as comparator | | | | | | |
| Per−month PS matching, using random days as comparator | | | | | | |
| Historical comparator | | | | | | |
| Unadjusted, using entire historic period | 63.6 | 70.6 | 82.6 | 76.3 | 85.6 | 93.7 |
| Age & sex adjusted, using entire historic period | 65.3 | 74.1 | 86.1 | 81.2 | 90.6 | 95.2 |
| Unadjusted, using TaR after historic visit | 64.7 | 73.0 | 83.8 | 75.7 | 84.4 | 93.4 |
| Age & sex adjusted, using TaR after historic visit | 65.8 | 75.5 | 89.1 | 78.4 | 89.1 | 94.6 |
| Unadjusted, using entire historic period, filtered | 63.5 | 70.6 | 82.2 | 76.3 | 85.5 | 93.2 |
| Age & sex adjusted, using entire historic period, filtered | 65.4 | 74.0 | 85.7 | 81.4 | 90.5 | 94.7 |
| Unadjusted, using TaR after historic visit, filtered | 64.8 | 73.0 | 83.4 | 75.7 | 84.1 | 93.0 |
| Age & sex adjusted, using TaR after historic visit, filtered | 66.0 | 75.6 | 89.0 | 78.5 | 88.9 | 94.0 |
| SCCS / SCRI | | | | | | |
| Unadjusted SCCS excluding pre−vaccination window | 73.2 | 83.1 | 91.9 | 87.1 | 92.8 | 95.1 |
| Age & season adjusted SCCS excluding pre−vaccination window | 73.2 | 83.7 | 92.4 | 86.6 | 92.4 | 94.9 |
| SCRI with prior control interval | 67.3 | 76.6 | 87.1 | 81.8 | 90.0 | 93.3 |
| SCRI with posterior control interval | 66.9 | 76.4 | 87.7 | 81.3 | 90.4 | 94.5 |
| Unadjusted SCCS excluding all pre−vaccination time | 71.5 | 81.2 | 89.7 | 86.1 | 92.1 | 95.0 |

Figure 82: The area under the receiver operator curve (AUC) based on the effect-size estimate at the end of the study period in the Optum EHR database for Zoster vaccination (Shingrix).

| | Uncalibrated AUC when true effect is | | | Calibrated AUC when true effect is | | |
|---|---|---|---|---|---|---|
| | 1.5 | 2 | 4 | 1.5 | 2 | 4 |
| Case−control | | | | | | |
| Age & sex matched controls | 54.6 | 56.3 | 58.6 | 60.3 | 62.3 | 63.9 |
| Age & sex adjusted, using random controls | 56.8 | 58.4 | 61.2 | 69.5 | 72.4 | 72.7 |
| Cohort method | | | | | | |
| Unadjusted, using outpatient visits as comparator | 55.7 | 60.0 | 69.8 | 61.9 | 69.2 | 72.6 |
| PS matching, using outpatient visits as comparator | 57.8 | 60.2 | 64.4 | 58.1 | 60.9 | 62.2 |
| Unadjusted, using random days as comparator | 55.6 | 57.0 | 59.8 | 63.7 | 66.2 | 68.6 |
| PS matching, using random days as comparator | 58.1 | 62.7 | 70.5 | 64.0 | 66.4 | 67.2 |
| PS stratification, using outpatient visits as comparator | 62.0 | 68.8 | 83.9 | 70.2 | 72.9 | 76.3 |
| PS stratification, using random days as comparator | 59.8 | 64.9 | 78.3 | 68.5 | 73.6 | 75.4 |
| PS weighting, using outpatient visits as comparator | 59.5 | 63.3 | 77.3 | 63.6 | 66.8 | 71.4 |
| PS weighting, using random days as comparator | 58.6 | 62.9 | 77.5 | 63.8 | 68.3 | 71.2 |
| Per−month PS matching, using outpatient visits as comparator | | | | | | |
| Per−month PS matching, using random days as comparator | | | | | | |
| Historical comparator | | | | | | |
| Unadjusted, using entire historic period | 54.4 | 57.7 | 61.9 | 64.9 | 70.2 | 74.6 |
| Age & sex adjusted, using entire historic period | 58.6 | 62.1 | 65.6 | 74.8 | 78.6 | 80.0 |
| Unadjusted, using TaR after historic visit | 56.2 | 59.9 | 64.5 | 68.0 | 73.9 | 77.3 |
| Age & sex adjusted, using TaR after historic visit | 61.8 | 67.2 | 74.4 | 69.7 | 76.0 | 77.4 |
| Unadjusted, using entire historic period, filtered | 54.4 | 57.7 | 61.8 | 64.4 | 69.8 | 73.3 |
| Age & sex adjusted, using entire historic period, filtered | 58.7 | 62.1 | 65.5 | 71.3 | 75.1 | 76.1 |
| Unadjusted, using TaR after historic visit, filtered | 56.2 | 59.9 | 64.4 | 66.6 | 72.4 | 75.5 |
| Age & sex adjusted, using TaR after historic visit, filtered | 61.9 | 67.3 | 74.4 | 68.8 | 74.4 | 75.5 |
| SCCS / SCRI | | | | | | |
| Unadjusted SCCS excluding pre−vaccination window | 64.2 | 68.4 | 72.3 | 71.5 | 73.9 | 75.0 |
| Age & season adjusted SCCS excluding pre−vaccination window | 63.0 | 67.8 | 71.7 | 71.3 | 73.8 | 75.0 |
| SCRI with prior control interval | 60.0 | 64.3 | 69.4 | 61.9 | 65.9 | 66.6 |
| SCRI with posterior control interval | 56.6 | 62.7 | 68.7 | 64.8 | 69.1 | 70.3 |
| Unadjusted SCCS excluding all pre−vaccination time | 60.9 | 65.4 | 69.5 | 69.0 | 72.4 | 73.4 |

Figure 83: The area under the receiver operator curve (AUC) based on the effect-size estimate at the end of the study period in the MDCD database for Zoster vaccination (Shingrix).

| | Uncalibrated AUC when true effect is | | | Calibrated AUC when true effect is | | |
|---|---|---|---|---|---|---|
| | 1.5 | 2 | 4 | 1.5 | 2 | 4 |
| Case−control | | | | | | |
| Age & sex matched controls | 58.3 | 64.1 | 71.8 | 64.9 | 71.8 | 74.5 |
| Age & sex adjusted, using random controls | 61.4 | 67.5 | 80.2 | 69.7 | 80.2 | 85.3 |
| Cohort method | | | | | | |
| Unadjusted, using outpatient visits as comparator | 57.6 | 64.8 | 78.4 | 66.4 | 73.4 | 81.6 |
| PS matching, using outpatient visits as comparator | 61.1 | 68.1 | 79.2 | 70.8 | 78.5 | 80.6 |
| Unadjusted, using random days as comparator | 59.2 | 65.2 | 74.9 | 66.9 | 75.3 | 80.7 |
| PS matching, using random days as comparator | 63.6 | 69.7 | 78.4 | 70.6 | 76.0 | 78.8 |
| PS stratification, using outpatient visits as comparator | 62.6 | 70.0 | 86.0 | 68.3 | 77.5 | 82.8 |
| PS stratification, using random days as comparator | 63.6 | 69.3 | 84.2 | 72.0 | 79.9 | 84.5 |
| PS weighting, using outpatient visits as comparator | 63.3 | 68.7 | 85.9 | 63.9 | 71.9 | 80.4 |
| PS weighting, using random days as comparator | 62.6 | 69.0 | 84.0 | 72.2 | 79.7 | 84.1 |
| Per−month PS matching, using outpatient visits as comparator | | | | | | |
| Per−month PS matching, using random days as comparator | | | | | | |
| Historical comparator | | | | | | |
| Unadjusted, using entire historic period | 61.3 | 66.6 | 78.2 | 68.6 | 78.5 | 85.1 |
| Age & sex adjusted, using entire historic period | 61.4 | 66.9 | 78.7 | 69.9 | 79.3 | 85.5 |
| Unadjusted, using TaR after historic visit | 59.8 | 65.9 | 77.2 | 67.1 | 75.9 | 84.3 |
| Age & sex adjusted, using TaR after historic visit | 60.2 | 66.1 | 78.1 | 67.6 | 76.8 | 85.1 |
| Unadjusted, using entire historic period, filtered | 61.6 | 66.9 | 78.3 | 70.0 | 78.3 | 83.6 |
| Age & sex adjusted, using entire historic period, filtered | 61.7 | 67.2 | 78.7 | 70.0 | 78.7 | 83.6 |
| Unadjusted, using TaR after historic visit, filtered | 60.1 | 66.4 | 77.7 | 67.2 | 75.9 | 83.0 |
| Age & sex adjusted, using TaR after historic visit, filtered | 60.6 | 66.6 | 78.6 | 67.7 | 76.8 | 83.4 |
| SCCS / SCRI | | | | | | |
| Unadjusted SCCS excluding pre−vaccination window | 61.9 | 70.0 | 79.3 | 74.1 | 80.9 | 84.4 |
| Age & season adjusted SCCS excluding pre−vaccination window | 61.9 | 69.6 | 78.1 | 73.6 | 79.7 | 82.2 |
| SCRI with prior control interval | 59.0 | 64.0 | 74.2 | 69.2 | 77.9 | 81.6 |
| SCRI with posterior control interval | 60.5 | 68.7 | 74.5 | 73.9 | 77.6 | 80.8 |
| Unadjusted SCCS excluding all pre−vaccination time | 65.0 | 71.0 | 79.1 | 74.1 | 79.8 | 83.9 |

Figure 84: The area under the receiver operator curve (AUC) based on the effect-size estimate at the end of the study period in the MDCR database for Zoster vaccination (Shingrix).

| | Uncalibrated AUC when true effect is | | | Calibrated AUC when true effect is | | |
|---|---|---|---|---|---|---|
| | 1.5 | 2 | 4 | 1.5 | 2 | 4 |
| Case−control | | | | | | |
| Age & sex matched controls | 60.5 | 67.2 | 76.8 | 72.1 | 80.9 | 84.2 |
| Age & sex adjusted, using random controls | 64.8 | 73.9 | 84.2 | 77.6 | 85.4 | 88.6 |
| Cohort method | | | | | | |
| Unadjusted, using outpatient visits as comparator | 62.5 | 70.3 | 84.7 | 72.8 | 82.5 | 90.1 |
| PS matching, using outpatient visits as comparator | 66.9 | 74.8 | 86.2 | 76.4 | 84.3 | 88.2 |
| Unadjusted, using random days as comparator | 59.4 | 63.3 | 74.0 | 67.7 | 75.3 | 84.2 |
| PS matching, using random days as comparator | 70.1 | 78.6 | 87.5 | 79.8 | 84.5 | 87.3 |
| PS stratification, using outpatient visits as comparator | 67.7 | 78.4 | 93.1 | 79.8 | 88.2 | 92.1 |
| PS stratification, using random days as comparator | 68.7 | 77.1 | 88.9 | 81.3 | 88.3 | 91.1 |
| PS weighting, using outpatient visits as comparator | 68.7 | 78.6 | 93.4 | 80.8 | 88.8 | 91.2 |
| PS weighting, using random days as comparator | 67.6 | 76.4 | 89.0 | 80.3 | 87.6 | 91.6 |
| Per−month PS matching, using outpatient visits as comparator | | | | | | |
| Per−month PS matching, using random days as comparator | | | | | | |
| Historical comparator | | | | | | |
| Unadjusted, using entire historic period | 62.7 | 69.7 | 80.9 | 75.0 | 83.7 | 90.1 |
| Age & sex adjusted, using entire historic period | 69.5 | 77.9 | 87.8 | 81.7 | 89.2 | 91.8 |
| Unadjusted, using TaR after historic visit | 64.9 | 73.0 | 83.3 | 76.7 | 84.5 | 90.6 |
| Age & sex adjusted, using TaR after historic visit | 68.2 | 77.3 | 89.4 | 79.1 | 87.9 | 92.2 |
| Unadjusted, using entire historic period, filtered | 62.8 | 69.8 | 80.5 | 75.1 | 83.4 | 89.1 |
| Age & sex adjusted, using entire historic period, filtered | 69.9 | 78.4 | 87.5 | 81.7 | 88.6 | 90.6 |
| Unadjusted, using TaR after historic visit, filtered | 65.3 | 73.4 | 83.0 | 76.6 | 83.7 | 89.2 |
| Age & sex adjusted, using TaR after historic visit, filtered | 68.7 | 78.0 | 89.6 | 79.0 | 87.2 | 90.7 |
| SCCS / SCRI | | | | | | |
| Unadjusted SCCS excluding pre−vaccination window | 70.1 | 79.8 | 89.5 | 82.8 | 90.1 | 91.3 |
| Age & season adjusted SCCS excluding pre−vaccination window | 69.7 | 79.6 | 88.8 | 82.3 | 89.0 | 90.0 |
| SCRI with prior control interval | 67.3 | 74.5 | 83.0 | 78.2 | 84.6 | 88.2 |
| SCRI with posterior control interval | 63.6 | 69.4 | 80.8 | 74.3 | 83.8 | 87.0 |
| Unadjusted SCCS excluding all pre−vaccination time | 66.8 | 75.4 | 84.6 | 80.2 | 87.0 | 91.0 |

Figure 85: The area under the receiver operator curve (AUC) based on the effect-size estimate at the end of the study period in the CCAE database for Zoster vaccination (Shingrix).

#### 12.6 AUC for HPV vaccination (Gardasil 9)

| | Uncalibrated AUC when true effect is | | | Calibrated AUC when true effect is | | |
|---|---|---|---|---|---|---|
| | 1.5 | 2 | 4 | 1.5 | 2 | 4 |
| Case−control | | | | | | |
| Age & sex matched controls | 56.9 | 59.5 | 63.3 | 59.5 | 67.7 | 71.8 |
| Age & sex adjusted, using random controls | 55.0 | 58.1 | 63.7 | 60.7 | 66.7 | 69.8 |
| Cohort method | | | | | | |
| Unadjusted, using outpatient visits as comparator | 63.4 | 70.2 | 81.8 | 60.6 | 67.9 | 77.8 |
| PS matching, using outpatient visits as comparator | 64.5 | 69.8 | 77.9 | 68.6 | 72.9 | 74.5 |
| Unadjusted, using random days as comparator | 60.3 | 65.1 | 75.4 | 63.7 | 72.4 | 80.0 |
| PS matching, using random days as comparator | | | | | | |
| PS stratification, using outpatient visits as comparator | 67.6 | 77.4 | 93.7 | 72.2 | 78.3 | 80.8 |
| PS stratification, using random days as comparator | | | | | | |
| PS weighting, using outpatient visits as comparator | 63.9 | 70.9 | 86.3 | 68.7 | 74.4 | 79.3 |
| PS weighting, using random days as comparator | | | | | | |
| Per−month PS matching, using outpatient visits as comparator | | | | | | |
| Per−month PS matching, using random days as comparator | | | | | | |
| Historical comparator | | | | | | |
| Unadjusted, using entire historic period | 57.3 | 61.4 | 74.3 | 61.9 | 71.9 | 78.3 |
| Age & sex adjusted, using entire historic period | 60.4 | 64.5 | 70.6 | 73.4 | 79.5 | 81.6 |
| Unadjusted, using TaR after historic visit | 56.4 | 61.2 | 69.4 | 61.6 | 70.7 | 78.7 |
| Age & sex adjusted, using TaR after historic visit | 62.9 | 70.7 | 79.9 | 72.8 | 79.1 | 82.3 |
| Unadjusted, using entire historic period, filtered | 57.2 | 61.3 | 74.2 | 62.1 | 71.7 | 78.0 |
| Age & sex adjusted, using entire historic period, filtered | 60.4 | 64.5 | 70.4 | 73.1 | 78.3 | 80.0 |
| Unadjusted, using TaR after historic visit, filtered | 55.9 | 60.6 | 68.9 | 61.0 | 70.1 | 77.9 |
| Age & sex adjusted, using TaR after historic visit, filtered | 63.0 | 70.9 | 80.0 | 73.0 | 78.7 | 81.6 |
| SCCS / SCRI | | | | | | |
| Unadjusted SCCS excluding pre−vaccination window | 62.1 | 68.8 | 75.4 | 72.6 | 77.8 | 80.4 |
| Age & season adjusted SCCS excluding pre−vaccination window | 62.2 | 69.0 | 76.0 | 71.6 | 77.2 | 79.9 |
| SCRI with prior control interval | 60.4 | 62.4 | 67.1 | 65.2 | 69.6 | 70.5 |
| SCRI with posterior control interval | 60.9 | 64.2 | 70.3 | 68.4 | 73.7 | 76.6 |
| Unadjusted SCCS excluding all pre−vaccination time | 62.8 | 67.8 | 73.8 | 72.0 | 77.6 | 81.2 |

Figure 86: The area under the receiver operator curve (AUC) based on the effect-size estimate at the end of the study period in the Optum EHR database for HPV vaccination (Gardasil 9).

| | Uncalibrated AUC when true effect is | | | Calibrated AUC when true effect is | | |
|---|---|---|---|---|---|---|
| | 1.5 | 2 | 4 | 1.5 | 2 | 4 |
| Case−control | | | | | | |
| Age & sex matched controls | 54.0 | 56.8 | 59.0 | 67.3 | 70.2 | 71.3 |
| Age & sex adjusted, using random controls | 57.5 | 61.3 | 63.9 | 71.2 | 73.7 | 74.6 |
| Cohort method | | | | | | |
| Unadjusted, using outpatient visits as comparator | 62.7 | 65.8 | 74.8 | 60.6 | 64.9 | 73.6 |
| PS matching, using outpatient visits as comparator | 58.5 | 62.6 | 67.7 | 62.9 | 67.3 | 70.0 |
| Unadjusted, using random days as comparator | 61.1 | 64.9 | 72.5 | 59.5 | 66.3 | 73.0 |
| PS matching, using random days as comparator | 60.9 | 64.1 | 68.7 | 60.7 | 67.5 | 68.7 |
| PS stratification, using outpatient visits as comparator | 62.5 | 70.5 | 86.9 | 66.3 | 72.4 | 75.0 |
| PS stratification, using random days as comparator | 60.6 | 65.9 | 80.2 | 66.3 | 71.6 | 75.1 |
| PS weighting, using outpatient visits as comparator | 61.0 | 65.7 | 81.2 | 64.3 | 68.3 | 71.4 |
| PS weighting, using random days as comparator | 58.6 | 60.8 | 74.9 | 60.2 | 65.5 | 71.4 |
| Per−month PS matching, using outpatient visits as comparator | | | | | | |
| Per−month PS matching, using random days as comparator | | | | | | |
| Historical comparator | | | | | | |
| Unadjusted, using entire historic period | 58.3 | 61.0 | 68.0 | 60.2 | 65.5 | 73.2 |
| Age & sex adjusted, using entire historic period | 61.9 | 66.6 | 71.7 | 72.0 | 77.6 | 79.8 |
| Unadjusted, using TaR after historic visit | 57.4 | 62.1 | 68.6 | 59.3 | 65.6 | 73.5 |
| Age & sex adjusted, using TaR after historic visit | 60.2 | 66.1 | 73.8 | 67.5 | 73.8 | 77.6 |
| Unadjusted, using entire historic period, filtered | 58.4 | 61.0 | 67.8 | 60.0 | 65.2 | 72.7 |
| Age & sex adjusted, using entire historic period, filtered | 62.1 | 66.8 | 71.7 | 72.4 | 76.9 | 78.3 |
| Unadjusted, using TaR after historic visit, filtered | 56.8 | 61.5 | 67.9 | 59.0 | 65.3 | 73.0 |
| Age & sex adjusted, using TaR after historic visit, filtered | 60.4 | 66.5 | 74.2 | 66.3 | 72.7 | 75.7 |
| SCCS / SCRI | | | | | | |
| Unadjusted SCCS excluding pre−vaccination window | 61.8 | 66.7 | 70.7 | 71.8 | 74.8 | 75.7 |
| Age & season adjusted SCCS excluding pre−vaccination window | 62.3 | 67.4 | 71.4 | 72.6 | 75.5 | 76.5 |
| SCRI with prior control interval | 58.5 | 62.0 | 67.9 | 69.4 | 75.5 | 76.7 |
| SCRI with posterior control interval | 57.7 | 61.9 | 68.1 | 68.6 | 73.0 | 75.2 |
| Unadjusted SCCS excluding all pre−vaccination time | 60.2 | 64.5 | 69.5 | 69.7 | 74.3 | 75.5 |

Figure 87: The area under the receiver operator curve (AUC) based on the effect-size estimate at the end of the study period in the MDCD database for HPV vaccination (Gardasil 9).

| | Uncalibrated AUC when true effect is | | | Calibrated AUC when true effect is | | |
|---|---|---|---|---|---|---|
| | 1.5 | 2 | 4 | 1.5 | 2 | 4 |
| Case−control | | | | | | |
| Age & sex matched controls | | | | | | |
| Age & sex adjusted, using random controls | | | | | | |
| Cohort method | | | | | | |
| Unadjusted, using outpatient visits as comparator | 50.0 | 50.0 | 50.0 | 50.0 | 50.0 | 50.0 |
| PS matching, using outpatient visits as comparator | | | | | | |
| Unadjusted, using random days as comparator | | | | | | |
| PS matching, using random days as comparator | | | | | | |
| PS stratification, using outpatient visits as comparator | 50.0 | 50.0 | 52.1 | 50.0 | 50.0 | 50.0 |
| PS stratification, using random days as comparator | | | | | | |
| PS weighting, using outpatient visits as comparator | 50.0 | 50.0 | 52.1 | 50.0 | 50.0 | 50.0 |
| PS weighting, using random days as comparator | | | | | | |
| Per−month PS matching, using outpatient visits as comparator | | | | | | |
| Per−month PS matching, using random days as comparator | | | | | | |
| Historical comparator | | | | | | |
| Unadjusted, using entire historic period | 50.0 | 50.0 | 50.0 | 50.0 | 50.0 | 50.0 |
| Age & sex adjusted, using entire historic period | 50.0 | 50.0 | 50.0 | 50.0 | 50.0 | 50.0 |
| Unadjusted, using TaR after historic visit | 50.0 | 50.0 | 50.0 | 50.0 | 50.0 | 50.0 |
| Age & sex adjusted, using TaR after historic visit | 50.0 | 50.0 | 50.0 | 50.0 | 50.0 | 50.0 |
| Unadjusted, using entire historic period, filtered | 50.0 | 50.0 | 50.0 | 50.0 | 50.0 | 50.0 |
| Age & sex adjusted, using entire historic period, filtered | 50.0 | 50.0 | 50.0 | 50.0 | 50.0 | 50.0 |
| Unadjusted, using TaR after historic visit, filtered | 50.0 | 50.0 | 50.0 | 50.0 | 50.0 | 50.0 |
| Age & sex adjusted, using TaR after historic visit, filtered | 50.0 | 50.0 | 50.0 | 50.0 | 50.0 | 50.0 |

Figure 88: The area under the receiver operator curve (AUC) based on the effect-size estimate at the end of the study period in the MDCR database for HPV vaccination (Gardasil 9).

| | Uncalibrated AUC when true effect is | | | Calibrated AUC when true effect is | | |
|---|---|---|---|---|---|---|
| | 1.5 | 2 | 4 | 1.5 | 2 | 4 |
| Case−control | | | | | | |
| Age & sex matched controls | 57.5 | 61.0 | 64.7 | 67.1 | 71.2 | 73.2 |
| Age & sex adjusted, using random controls | 57.7 | 62.2 | 66.4 | 68.1 | 71.5 | 73.4 |
| Cohort method | | | | | | |
| Unadjusted, using outpatient visits as comparator | 62.6 | 68.1 | 76.4 | 61.2 | 68.3 | 78.0 |
| PS matching, using outpatient visits as comparator | 60.7 | 65.9 | 73.6 | 68.3 | 72.8 | 73.7 |
| Unadjusted, using random days as comparator | 61.3 | 66.8 | 78.0 | 65.1 | 72.6 | 79.9 |
| PS matching, using random days as comparator | 63.9 | 69.2 | 75.6 | 70.1 | 73.3 | 73.8 |
| PS stratification, using outpatient visits as comparator | 66.7 | 74.1 | 91.9 | 70.5 | 75.9 | 77.7 |
| PS stratification, using random days as comparator | 63.2 | 69.2 | 83.1 | 69.8 | 74.9 | 79.1 |
| PS weighting, using outpatient visits as comparator | 63.3 | 70.6 | 87.1 | 70.7 | 75.0 | 78.1 |
| PS weighting, using random days as comparator | 58.7 | 64.3 | 78.3 | 64.2 | 70.5 | 75.9 |
| Per−month PS matching, using outpatient visits as comparator | | | | | | |
| Per−month PS matching, using random days as comparator | | | | | | |
| Historical comparator | | | | | | |
| Unadjusted, using entire historic period | 57.9 | 62.7 | 74.4 | 65.4 | 72.8 | 80.8 |
| Age & sex adjusted, using entire historic period | 64.1 | 69.7 | 75.5 | 74.8 | 80.0 | 81.6 |
| Unadjusted, using TaR after historic visit | 57.1 | 61.1 | 69.3 | 65.1 | 72.1 | 81.1 |
| Age & sex adjusted, using TaR after historic visit | 63.3 | 70.1 | 79.5 | 73.1 | 79.5 | 82.3 |
| Unadjusted, using entire historic period, filtered | 57.2 | 62.0 | 73.7 | 65.1 | 71.9 | 79.7 |
| Age & sex adjusted, using entire historic period, filtered | 64.3 | 69.8 | 75.1 | 75.1 | 79.3 | 80.5 |
| Unadjusted, using TaR after historic visit, filtered | 57.0 | 60.0 | 67.8 | 65.1 | 71.3 | 80.2 |
| Age & sex adjusted, using TaR after historic visit, filtered | 63.4 | 70.2 | 79.4 | 72.8 | 78.5 | 80.8 |
| SCCS / SCRI | | | | | | |
| Unadjusted SCCS excluding pre−vaccination window | 63.5 | 69.3 | 74.7 | 71.8 | 76.3 | 78.3 |
| Age & season adjusted SCCS excluding pre−vaccination window | 63.7 | 69.9 | 75.4 | 72.5 | 76.6 | 78.7 |
| SCRI with prior control interval | 60.9 | 65.0 | 69.5 | 69.9 | 73.8 | 75.6 |
| SCRI with posterior control interval | 64.6 | 69.9 | 73.0 | 73.1 | 75.3 | 75.5 |
| Unadjusted SCCS excluding all pre−vaccination time | 62.5 | 67.7 | 72.5 | 72.6 | 76.0 | 77.1 |

Figure 89: The area under the receiver operator curve (AUC) based on the effect-size estimate at the end of the study period in the CCAE database for HPV vaccination (Gardasil 9).

### 13 Confidence-interval-based metrics

For each method variation and vaccine group, the coverage of the 95% confidence interval, (geometric) mean of the precision (1/SE^2), mean squared error (MSE), and fraction non-estimable, based on the negative control effect-size estimates at the end of the study period.

#### 13.1 Confidence-interval-based metrics for H1N1 vaccination

| | Uncalibrated | | | | Calibrated | | | |
|---|---|---|---|---|---|---|---|---|
| | Coverage | Mean precision | MSE | Non estimable | Coverage | Mean precision | MSE | Non estimable |
| Case−control | | | | | | | | |
| Age & sex matched controls | 0.83 | 0.60 | 81.26 | 0.49 | 0.98 | 0.60 | 76.12 | 0.49 |
| Age & sex adjusted, using random controls | 0.81 | 0.81 | 0.79 | 0.47 | 1.00 | 0.81 | 0.27 | 0.47 |
| Cohort method | | | | | | | | |
| Unadjusted, using outpatient visits as comparator | 0.80 | 1.70 | 1.11 | 0.34 | 0.96 | 1.16 | 0.41 | 0.34 |
| PS matching, using outpatient visits as comparator | 0.89 | 1.37 | 0.95 | 0.35 | 0.99 | 1.36 | 0.32 | 0.35 |
| Unadjusted, using random days as comparator | 0.78 | 0.73 | 0.82 | 0.52 | 1.00 | 0.72 | 0.23 | 0.52 |
| PS matching, using random days as comparator | 1.00 | 0.00 | 0.00 | 1.00 | 1.00 | 0.00 | 0.00 | 1.00 |
| PS stratification, using outpatient visits as comparator | 0.82 | 1.88 | 0.96 | 0.29 | 0.97 | 1.66 | 0.42 | 0.29 |
| PS stratification, using random days as comparator | 0.67 | 1.60 | 1.34 | 0.34 | 1.00 | 1.60 | 0.25 | 0.34 |
| PS weighting, using outpatient visits as comparator | 0.82 | 1.51 | 0.93 | 0.31 | 0.97 | 1.10 | 0.57 | 0.31 |
| PS weighting, using random days as comparator | 1.00 | 0.00 | 2.78 | 1.00 | 1.00 | 0.00 | 0.00 | 1.00 |
| Per−month PS matching, using outpatient visits as comparator | 0.97 | 0.69 | 0.97 | 0.48 | 0.98 | 0.64 | 0.20 | 0.48 |
| Per−month PS matching, using random days as comparator | 0.99 | 0.32 | 0.44 | 0.68 | 1.00 | 0.31 | 0.15 | 0.68 |
| Historical comparator | | | | | | | | |
| Unadjusted, using entire historic period | 0.59 | 2.87 | 1.12 | 0.27 | 0.94 | 2.20 | 0.30 | 0.27 |
| Age & sex adjusted, using entire historic period | 0.59 | 2.86 | 1.30 | 0.27 | 0.94 | 2.14 | 0.31 | 0.27 |
| Unadjusted, using TaR after historic visit | 0.82 | 2.78 | 0.43 | 0.27 | 0.97 | 1.58 | 0.34 | 0.27 |
| Age & sex adjusted, using TaR after historic visit | 0.80 | 2.77 | 0.50 | 0.27 | 0.94 | 2.07 | 0.31 | 0.27 |
| Unadjusted, using entire historic period, filtered | 0.61 | 2.69 | 0.99 | 0.31 | 0.94 | 2.06 | 0.28 | 0.31 |
| Age & sex adjusted, using entire historic period, filtered | 0.61 | 2.69 | 1.15 | 0.31 | 0.95 | 2.05 | 0.28 | 0.31 |
| Unadjusted, using TaR after historic visit, filtered | 0.83 | 2.62 | 0.39 | 0.31 | 0.98 | 1.50 | 0.32 | 0.31 |
| Age & sex adjusted, using TaR after historic visit, filtered | 0.81 | 2.60 | 0.44 | 0.31 | 0.95 | 1.98 | 0.29 | 0.31 |
| SCCS / SCRI | | | | | | | | |
| Unadjusted SCCS excluding pre−vaccination window | 0.95 | 2.55 | 0.26 | 0.28 | 0.98 | 2.28 | 0.26 | 0.28 |
| Age & season adjusted SCCS excluding pre−vaccination window | 0.96 | 2.41 | 0.26 | 0.29 | 0.99 | 2.40 | 0.25 | 0.29 |
| SCRI with prior control interval | 0.97 | 1.52 | 0.25 | 0.33 | 0.98 | 1.50 | 0.25 | 0.33 |
| SCRI with posterior control interval | 0.97 | 1.45 | 4.52 | 0.38 | 0.99 | 1.45 | 4.57 | 0.38 |
| Unadjusted SCCS excluding all pre−vaccination time | 0.95 | 2.29 | 4.50 | 0.29 | 0.99 | 1.75 | 0.35 | 0.29 |

Figure 90: The coverage, mean precision, mean squared error (MSE), and fraction non-estimable based on the negative control effect-size estimate at the end of the study period in the Optum EHR database for H1N1 vaccination.

| | Uncalibrated | | | | Calibrated | | | |
|---|---|---|---|---|---|---|---|---|
| Case−control | Coverage | Mean precision | MSE | Non estimable | Coverage | Mean precision | MSE | Non estimable |
| Age & sex matched controls | 0.88 | 1.48 | 15.22 | 0.42 | 0.99 | 1.23 | 0.37 | 0.42 |
| Age & sex adjusted, using random controls | 0.97 | 1.90 | 0.28 | 0.37 | 0.98 | 1.80 | 0.23 | 0.37 |
| Cohort method | | | | | | | | |
| Unadjusted, using outpatient visits as comparator | 0.48 | 1.90 | 3.17 | 0.35 | 0.97 | 0.45 | 0.74 | 0.35 |
| PS matching, using outpatient visits as comparator | 0.92 | 1.10 | 0.71 | 0.48 | 0.97 | 0.93 | 0.20 | 0.48 |
| Unadjusted, using random days as comparator | 0.73 | 1.54 | 1.12 | 0.42 | 0.98 | 0.65 | 0.40 | 0.42 |
| PS matching, using random days as comparator | 0.97 | 1.05 | 0.51 | 0.49 | 0.98 | 1.04 | 0.20 | 0.49 |
| PS stratification, using outpatient visits as comparator | 0.91 | 1.56 | 0.67 | 0.38 | 0.96 | 1.32 | 0.34 | 0.38 |
| PS stratification, using random days as comparator | 0.95 | 1.51 | 0.50 | 0.38 | 0.97 | 1.39 | 0.31 | 0.38 |
| PS weighting, using outpatient visits as comparator | 0.92 | 1.18 | 0.65 | 0.40 | 0.96 | 1.15 | 0.36 | 0.40 |
| PS weighting, using random days as comparator | 0.98 | 0.55 | 1.16 | 0.39 | 1.00 | 0.54 | 1.02 | 0.39 |
| Per−month PS matching, using outpatient visits as comparator | 0.97 | 0.99 | 0.52 | 0.48 | 0.98 | 0.98 | 0.15 | 0.48 |
| Per−month PS matching, using random days as comparator | 0.99 | 0.98 | 0.61 | 0.49 | 0.99 | 0.97 | 0.22 | 0.49 |
| Historical comparator | | | | | | | | |
| Unadjusted, using entire historic period | 0.62 | 2.44 | 0.86 | 0.33 | 0.98 | 0.79 | 0.51 | 0.33 |
| Age & sex adjusted, using entire historic period | 0.92 | 2.43 | 0.41 | 0.33 | 0.96 | 1.51 | 0.38 | 0.33 |
| Unadjusted, using TaR after historic visit | 0.54 | 2.35 | 1.31 | 0.33 | 0.98 | 0.66 | 0.62 | 0.33 |
| Age & sex adjusted, using TaR after historic visit | 0.91 | 2.28 | 0.40 | 0.33 | 0.94 | 1.54 | 0.40 | 0.33 |
| Unadjusted, using entire historic period, filtered | 0.62 | 2.40 | 0.85 | 0.34 | 0.98 | 0.79 | 0.48 | 0.34 |
| Age & sex adjusted, using entire historic period, filtered | 0.94 | 2.39 | 0.34 | 0.34 | 0.97 | 1.53 | 0.31 | 0.34 |
| Unadjusted, using TaR after historic visit, filtered | 0.54 | 2.32 | 1.30 | 0.34 | 0.98 | 0.65 | 0.58 | 0.34 |
| Age & sex adjusted, using TaR after historic visit, filtered | 0.92 | 2.25 | 0.31 | 0.34 | 0.96 | 1.54 | 0.31 | 0.34 |
| SCCS / SCRI | | | | | | | | |
| Unadjusted SCCS excluding pre−vaccination window | 0.94 | 2.17 | 0.23 | 0.37 | 0.98 | 1.46 | 0.23 | 0.37 |
| Age & season adjusted SCCS excluding pre−vaccination window | 0.94 | 2.07 | 0.21 | 0.38 | 0.97 | 1.95 | 0.21 | 0.38 |
| SCRI with prior control interval | 0.99 | 1.30 | 14.02 | 0.42 | 0.99 | 0.98 | 0.22 | 0.42 |
| SCRI with posterior control interval | 0.97 | 1.30 | 14.04 | 0.42 | 0.98 | 1.28 | 0.23 | 0.42 |
| Unadjusted SCCS excluding all pre−vaccination time | 0.96 | 1.97 | 0.21 | 0.37 | 0.98 | 1.45 | 0.21 | 0.37 |

Figure 91: The coverage, mean precision, mean squared error (MSE), and fraction non-estimable based on the negative control effect-size estimate at the end of the study period in the MDCD database for H1N1 vaccination.

| Case−control | Uncalibrated Coverage | Uncalibrated Mean precision | Uncalibrated MSE | Uncalibrated Non estimable | Calibrated Coverage | Calibrated Mean precision | Calibrated MSE | Calibrated Non estimable |
|---|---|---|---|---|---|---|---|---|
| Age & sex matched controls | 0.92 | 0.46 | 17.76 | 0.63 | 0.98 | 0.46 | 17.12 | 0.63 |
| Age & sex adjusted, using random controls | 0.97 | 0.46 | 0.50 | 0.58 | 1.00 | 0.39 | 0.34 | 0.58 |
| Cohort method | | | | | | | | |
| Unadjusted, using outpatient visits as comparator | 0.98 | 0.47 | 0.55 | 0.60 | 0.98 | 0.47 | 0.18 | 0.60 |
| PS matching, using outpatient visits as comparator | 1.00 | 0.22 | 0.51 | 0.76 | 1.00 | 0.22 | 0.11 | 0.76 |
| Unadjusted, using random days as comparator | 0.97 | 0.29 | 0.52 | 0.74 | 0.99 | 0.29 | 0.17 | 0.74 |
| PS matching, using random days as comparator | 1.00 | 0.22 | 0.29 | 0.74 | 1.00 | 0.22 | 0.13 | 0.74 |
| PS stratification, using outpatient visits as comparator | 0.96 | 0.47 | 0.72 | 0.58 | 0.98 | 0.47 | 0.36 | 0.58 |
| PS stratification, using random days as comparator | 0.99 | 0.37 | 0.69 | 0.66 | 0.99 | 0.37 | 0.43 | 0.66 |
| PS weighting, using outpatient visits as comparator | 0.86 | 0.58 | 1.26 | 0.58 | 0.99 | 0.14 | 0.87 | 0.58 |
| PS weighting, using random days as comparator | 1.00 | 0.13 | 0.91 | 0.66 | 1.00 | 0.13 | 0.62 | 0.66 |
| Per−month PS matching, using outpatient visits as comparator | 0.98 | 0.22 | 0.38 | 0.76 | 1.00 | 0.21 | 0.16 | 0.76 |
| Per−month PS matching, using random days as comparator | 1.00 | 0.21 | 0.42 | 0.74 | 1.00 | 0.21 | 0.07 | 0.74 |
| Historical comparator | | | | | | | | |
| Unadjusted, using entire historic period | 0.92 | 0.79 | 0.72 | 0.54 | 0.95 | 0.58 | 0.43 | 0.54 |
| Age & sex adjusted, using entire historic period | 0.92 | 0.79 | 0.72 | 0.54 | 0.95 | 0.63 | 0.43 | 0.54 |
| Unadjusted, using TaR after historic visit | 0.92 | 0.78 | 0.61 | 0.54 | 0.94 | 0.65 | 0.48 | 0.54 |
| Age & sex adjusted, using TaR after historic visit | 0.94 | 0.78 | 0.59 | 0.54 | 0.94 | 0.63 | 0.45 | 0.54 |
| Unadjusted, using entire historic period, filtered | 0.94 | 0.78 | 0.59 | 0.55 | 0.96 | 0.61 | 0.36 | 0.55 |
| Age & sex adjusted, using entire historic period, filtered | 0.94 | 0.78 | 0.59 | 0.55 | 0.96 | 0.62 | 0.35 | 0.55 |
| Unadjusted, using TaR after historic visit, filtered | 0.94 | 0.77 | 0.47 | 0.55 | 0.95 | 0.64 | 0.38 | 0.55 |
| Age & sex adjusted, using TaR after historic visit, filtered | 0.95 | 0.77 | 0.47 | 0.55 | 0.95 | 0.65 | 0.37 | 0.55 |
| SCCS / SCRI | | | | | | | | |
| Unadjusted SCCS excluding pre−vaccination window | 0.96 | 0.68 | 0.24 | 0.57 | 1.00 | 0.68 | 0.22 | 0.57 |
| Age & season adjusted SCCS excluding pre−vaccination window | 0.97 | 0.68 | 0.22 | 0.57 | 0.99 | 0.68 | 0.20 | 0.57 |
| SCRI with prior control interval | 1.00 | 0.35 | 13.96 | 0.65 | 1.00 | 0.35 | 0.14 | 0.65 |
| SCRI with posterior control interval | 0.98 | 0.41 | 0.17 | 0.61 | 0.98 | 0.41 | 0.17 | 0.61 |
| Unadjusted SCCS excluding all pre−vaccination time | 0.96 | 0.62 | 15.75 | 0.57 | 0.98 | 0.61 | 15.65 | 0.57 |

Figure 92: The coverage, mean precision, mean squared error (MSE), and fraction non-estimable based on the negative control effect-size estimate at the end of the study period in the MDCR database for H1N1 vaccination.

| | Uncalibrated | | | | Calibrated | | | |
|---|---|---|---|---|---|---|---|---|
| | Coverage | Mean precision | MSE | Non estimable | Coverage | Mean precision | MSE | Non estimable |
| **Case−control** | | | | | | | | |
| Age & sex matched controls | 0.88 | 1.61 | 29.95 | 0.42 | 0.97 | 1.31 | 29.33 | 0.42 |
| Age & sex adjusted, using random controls | 0.88 | 1.87 | 0.31 | 0.39 | 0.99 | 1.46 | 0.24 | 0.39 |
| **Cohort method** | | | | | | | | |
| Unadjusted, using outpatient visits as comparator | 0.39 | 6.32 | 1.67 | 0.20 | 0.96 | 1.14 | 0.51 | 0.20 |
| PS matching, using outpatient visits as comparator | 0.77 | 4.50 | 0.46 | 0.22 | 0.97 | 2.93 | 0.34 | 0.22 |
| Unadjusted, using random days as comparator | 0.80 | 4.38 | 0.32 | 0.20 | 0.97 | 2.06 | 0.31 | 0.20 |
| PS matching, using random days as comparator | 0.92 | 4.19 | 0.32 | 0.23 | 0.95 | 3.84 | 0.32 | 0.23 |
| PS stratification, using outpatient visits as comparator | 0.70 | 6.25 | 0.39 | 0.19 | 0.96 | 4.04 | 0.22 | 0.19 |
| PS stratification, using random days as comparator | 0.90 | 5.52 | 0.26 | 0.20 | 0.95 | 4.00 | 0.25 | 0.20 |
| PS weighting, using outpatient visits as comparator | 0.55 | 5.59 | 0.82 | 0.19 | 0.99 | 2.11 | 0.37 | 0.19 |
| PS weighting, using random days as comparator | 0.96 | 2.07 | 0.38 | 0.20 | 0.97 | 2.05 | 0.34 | 0.20 |
| Per−month PS matching, using outpatient visits as comparator | 0.81 | 4.16 | 0.47 | 0.24 | 0.99 | 3.06 | 0.30 | 0.24 |
| Per−month PS matching, using random days as comparator | 0.95 | 3.92 | 0.30 | 0.26 | 0.97 | 3.66 | 0.21 | 0.26 |
| **Historical comparator** | | | | | | | | |
| Unadjusted, using entire historic period | 0.80 | 8.60 | 0.19 | 0.19 | 0.96 | 3.05 | 0.19 | 0.19 |
| Age & sex adjusted, using entire historic period | 0.74 | 8.59 | 0.22 | 0.19 | 0.97 | 3.49 | 0.16 | 0.19 |
| Unadjusted, using TaR after historic visit | 0.61 | 8.35 | 0.33 | 0.19 | 0.96 | 2.18 | 0.24 | 0.19 |
| Age & sex adjusted, using TaR after historic visit | 0.82 | 8.27 | 0.15 | 0.19 | 0.97 | 3.97 | 0.14 | 0.19 |
| Unadjusted, using entire historic period, filtered | 0.80 | 8.44 | 0.18 | 0.20 | 0.96 | 3.02 | 0.18 | 0.20 |
| Age & sex adjusted, using entire historic period, filtered | 0.74 | 8.43 | 0.21 | 0.20 | 0.97 | 3.44 | 0.15 | 0.20 |
| Unadjusted, using TaR after historic visit, filtered | 0.61 | 8.20 | 0.33 | 0.20 | 0.96 | 2.15 | 0.23 | 0.20 |
| Age & sex adjusted, using TaR after historic visit, filtered | 0.82 | 8.12 | 0.14 | 0.20 | 0.97 | 3.89 | 0.14 | 0.20 |
| **SCCS / SCRI** | | | | | | | | |
| Unadjusted SCCS excluding pre−vaccination window | 0.89 | 7.96 | 0.15 | 0.19 | 0.98 | 3.84 | 0.15 | 0.19 |
| Age & season adjusted SCCS excluding pre−vaccination window | 0.91 | 7.13 | 0.14 | 0.22 | 0.96 | 5.37 | 0.14 | 0.22 |
| SCRI with prior control interval | 0.90 | 4.72 | 0.22 | 0.20 | 0.99 | 2.87 | 0.22 | 0.20 |
| SCRI with posterior control interval | 0.91 | 4.55 | 15.08 | 0.22 | 0.97 | 3.69 | 0.23 | 0.22 |
| Unadjusted SCCS excluding all pre−vaccination time | 0.87 | 7.06 | 0.15 | 0.19 | 0.97 | 3.74 | 0.15 | 0.19 |

Figure 93: The coverage, mean precision, mean squared error (MSE), and fraction non-estimable based on the negative control effect-size estimate at the end of the study period in the CCAE database for H1N1 vaccination.

#### 13.2 Confidence-interval-based metrics for Seasonal flu vaccination (Fluvirin)

| | Uncalibrated | | | | Calibrated | | | |
|---|---|---|---|---|---|---|---|---|
| Case−control | Coverage | Mean precision | MSE | Non estimable | Coverage | Mean precision | MSE | Non estimable |
| Age & sex matched controls | 0.97 | 0.10 | 33.76 | 0.82 | 1.00 | 0.09 | 31.89 | 0.82 |
| Age & sex adjusted, using random controls | 0.98 | 0.06 | 0.32 | 0.87 | 1.00 | 0.06 | 0.06 | 0.87 |
| Cohort method | | | | | | | | |
| Unadjusted, using outpatient visits as comparator | 0.95 | 0.38 | 1.45 | 0.60 | 0.98 | 0.37 | 0.40 | 0.60 |
| PS matching, using outpatient visits as comparator | 0.99 | 0.08 | 0.58 | 0.85 | 1.00 | 0.08 | 0.09 | 0.85 |
| Unadjusted, using random days as comparator | 0.99 | 0.23 | 0.57 | 0.70 | 1.00 | 0.23 | 0.10 | 0.70 |
| PS matching, using random days as comparator | | | | | | | | |
| PS stratification, using outpatient visits as comparator | 0.96 | 0.33 | 1.02 | 0.54 | 0.97 | 0.32 | 0.50 | 0.54 |
| PS stratification, using random days as comparator | 0.95 | 0.44 | 0.99 | 0.57 | 0.98 | 0.44 | 0.36 | 0.57 |
| PS weighting, using outpatient visits as comparator | 0.98 | 0.19 | 0.91 | 0.57 | 1.00 | 0.19 | 0.35 | 0.57 |
| PS weighting, using random days as comparator | 1.00 | 0.00 | 0.00 | 1.00 | 1.00 | 0.00 | 0.00 | 1.00 |
| Per−month PS matching, using outpatient visits as comparator | | | | | | | | |
| Per−month PS matching, using random days as comparator | | | | | | | | |
| Historical comparator | | | | | | | | |
| Unadjusted, using entire historic period | 0.86 | 0.70 | 1.09 | 0.53 | 0.96 | 0.56 | 0.34 | 0.53 |
| Age & sex adjusted, using entire historic period | 0.87 | 0.70 | 1.00 | 0.53 | 0.95 | 0.61 | 0.32 | 0.53 |
| Unadjusted, using TaR after historic visit | 0.92 | 0.70 | 0.51 | 0.53 | 0.95 | 0.61 | 0.37 | 0.53 |
| Age & sex adjusted, using TaR after historic visit | 0.94 | 0.70 | 0.52 | 0.53 | 0.92 | 0.65 | 0.39 | 0.53 |
| Unadjusted, using entire historic period, filtered | 0.87 | 0.67 | 1.02 | 0.54 | 0.94 | 0.64 | 0.32 | 0.54 |
| Age & sex adjusted, using entire historic period, filtered | 0.88 | 0.67 | 0.94 | 0.54 | 0.96 | 0.65 | 0.31 | 0.54 |
| Unadjusted, using TaR after historic visit, filtered | 0.94 | 0.67 | 0.48 | 0.54 | 0.94 | 0.63 | 0.35 | 0.54 |
| Age & sex adjusted, using TaR after historic visit, filtered | 0.95 | 0.67 | 0.49 | 0.54 | 0.95 | 0.61 | 0.38 | 0.54 |
| SCCS / SCRI | | | | | | | | |
| Unadjusted SCCS excluding pre−vaccination window | 0.99 | 0.51 | 0.17 | 0.62 | 1.00 | 0.51 | 0.15 | 0.62 |
| Age & season adjusted SCCS excluding pre−vaccination window | 0.99 | 0.47 | 0.18 | 0.65 | 1.00 | 0.46 | 0.14 | 0.65 |
| SCRI with prior control interval | 1.00 | 0.21 | 39.95 | 0.74 | 1.00 | 0.21 | 4.63 | 0.74 |
| SCRI with posterior control interval | 1.00 | 0.26 | 13.95 | 0.69 | 1.00 | 0.26 | 0.13 | 0.69 |
| Unadjusted SCCS excluding all pre−vaccination time | 0.99 | 0.47 | 0.26 | 0.62 | 1.00 | 0.47 | 0.17 | 0.62 |

Figure 94: The coverage, mean precision, mean squared error (MSE), and fraction non-estimable based on the negative control effect-size estimate at the end of the study period in the Optum EHR database for Seasonal flu vaccination (Fluvirin).

| | Uncalibrated | | | | Calibrated | | | |
|---|---|---|---|---|---|---|---|---|
| Case−control | Coverage | Mean precision | MSE | Non estimable | Coverage | Mean precision | MSE | Non estimable |
| Age & sex matched controls | 0.90 | 0.47 | 0.34 | 0.72 | 0.98 | 0.27 | 0.27 | 0.72 |
| Age & sex adjusted, using random controls | 1.00 | 0.35 | 0.26 | 0.67 | 1.00 | 0.35 | 0.16 | 0.67 |
| Cohort method | | | | | | | | |
| Unadjusted, using outpatient visits as comparator | 0.97 | 0.41 | 1.07 | 0.68 | 0.99 | 0.41 | 0.15 | 0.68 |
| PS matching, using outpatient visits as comparator | 1.00 | 0.00 | 0.00 | 1.00 | 1.00 | 0.00 | 0.00 | 1.00 |
| Unadjusted, using random days as comparator | 0.95 | 0.22 | 0.51 | 0.75 | 0.98 | 0.21 | 0.25 | 0.75 |
| PS matching, using random days as comparator | 1.00 | 0.00 | 0.00 | 1.00 | 1.00 | 0.00 | 0.00 | 1.00 |
| PS stratification, using outpatient visits as comparator | 0.96 | 0.60 | 0.92 | 0.62 | 0.98 | 0.59 | 0.26 | 0.62 |
| PS stratification, using random days as comparator | 0.96 | 0.51 | 0.81 | 0.63 | 0.99 | 0.50 | 0.22 | 0.63 |
| PS weighting, using outpatient visits as comparator | 0.99 | 0.18 | 0.75 | 0.62 | 1.00 | 0.18 | 0.26 | 0.62 |
| PS weighting, using random days as comparator | 1.00 | 0.00 | 0.72 | 1.00 | 1.00 | 0.00 | 0.00 | 1.00 |
| Per−month PS matching, using outpatient visits as comparator | | | | | | | | |
| Per−month PS matching, using random days as comparator | | | | | | | | |
| Historical comparator | | | | | | | | |
| Unadjusted, using entire historic period | 0.92 | 0.70 | 0.49 | 0.61 | 0.99 | 0.28 | 0.29 | 0.61 |
| Age & sex adjusted, using entire historic period | 0.97 | 0.70 | 0.28 | 0.61 | 0.99 | 0.32 | 0.23 | 0.61 |
| Unadjusted, using TaR after historic visit | 0.96 | 0.70 | 0.31 | 0.61 | 0.99 | 0.31 | 0.26 | 0.61 |
| Age & sex adjusted, using TaR after historic visit | 0.97 | 0.69 | 0.23 | 0.61 | 0.99 | 0.33 | 0.23 | 0.61 |
| Unadjusted, using entire historic period, filtered | 0.94 | 0.65 | 0.29 | 0.62 | 0.99 | 0.62 | 0.15 | 0.62 |
| Age & sex adjusted, using entire historic period, filtered | 0.98 | 0.65 | 0.14 | 0.62 | 0.97 | 0.64 | 0.11 | 0.62 |
| Unadjusted, using TaR after historic visit, filtered | 0.97 | 0.65 | 0.16 | 0.62 | 0.98 | 0.64 | 0.14 | 0.62 |
| Age & sex adjusted, using TaR after historic visit, filtered | 0.98 | 0.65 | 0.14 | 0.62 | 0.98 | 0.63 | 0.13 | 0.62 |
| SCCS / SCRI | | | | | | | | |
| Unadjusted SCCS excluding pre−vaccination window | 0.97 | 0.63 | 0.14 | 0.61 | 0.98 | 0.63 | 0.14 | 0.61 |
| Age & season adjusted SCCS excluding pre−vaccination window | 0.97 | 0.60 | 0.14 | 0.62 | 0.98 | 0.60 | 0.14 | 0.62 |
| SCRI with prior control interval | 0.99 | 0.30 | 51.47 | 0.75 | 1.00 | 0.29 | 0.17 | 0.75 |
| SCRI with posterior control interval | 0.98 | 0.39 | 4.09 | 0.65 | 0.99 | 0.39 | 4.11 | 0.65 |
| Unadjusted SCCS excluding all pre−vaccination time | 0.98 | 0.61 | 3.94 | 0.61 | 0.99 | 0.60 | 3.96 | 0.61 |

Figure 95: The coverage, mean precision, mean squared error (MSE), and fraction non-estimable based on the negative control effect-size estimate at the end of the study period in the MDCD database for Seasonal flu vaccination (Fluvirin).

| | Uncalibrated | | | | Calibrated | | | |
|---|---|---|---|---|---|---|---|---|
| Case−control | Coverage | Mean precision | MSE | Non estimable | Coverage | Mean precision | MSE | Non estimable |
| Age & sex matched controls | 1.00 | 0.02 | 4.46 | 0.96 | 1.00 | 0.00 | 0.00 | 1.00 |
| Age & sex adjusted, using random controls | 1.00 | 0.02 | 0.06 | 0.95 | 1.00 | 0.00 | 0.00 | 1.00 |
| Cohort method | | | | | | | | |
| Unadjusted, using outpatient visits as comparator | 1.00 | 0.02 | 0.61 | 0.96 | 1.00 | 0.00 | 0.00 | 1.00 |
| PS matching, using outpatient visits as comparator | 1.00 | 0.00 | 0.29 | 0.99 | 1.00 | 0.00 | 0.00 | 1.00 |
| Unadjusted, using random days as comparator | 1.00 | 0.02 | 0.06 | 0.95 | 1.00 | 0.00 | 0.00 | 1.00 |
| PS matching, using random days as comparator | 1.00 | 0.00 | 0.13 | 0.99 | 1.00 | 0.00 | 0.00 | 1.00 |
| PS stratification, using outpatient visits as comparator | 1.00 | 0.03 | 0.56 | 0.92 | 1.00 | 0.03 | 0.10 | 0.92 |
| PS stratification, using random days as comparator | 1.00 | 0.02 | 0.29 | 0.96 | 1.00 | 0.00 | 0.00 | 1.00 |
| PS weighting, using outpatient visits as comparator | 0.99 | 0.02 | 0.72 | 0.92 | 0.99 | 0.02 | 0.22 | 0.92 |
| PS weighting, using random days as comparator | 1.00 | 0.00 | 0.36 | 0.96 | 1.00 | 0.00 | 0.00 | 1.00 |
| Per−month PS matching, using outpatient visits as comparator | | | | | | | | |
| Per−month PS matching, using random days as comparator | | | | | | | | |
| Historical comparator | | | | | | | | |
| Unadjusted, using entire historic period | 0.99 | 0.06 | 0.22 | 0.91 | 1.00 | 0.06 | 0.09 | 0.91 |
| Age & sex adjusted, using entire historic period | 1.00 | 0.06 | 0.22 | 0.91 | 1.00 | 0.06 | 0.08 | 0.91 |
| Unadjusted, using TaR after historic visit | 1.00 | 0.06 | 0.19 | 0.91 | 1.00 | 0.06 | 0.08 | 0.91 |
| Age & sex adjusted, using TaR after historic visit | 1.00 | 0.06 | 0.19 | 0.91 | 1.00 | 0.06 | 0.08 | 0.91 |
| Unadjusted, using entire historic period, filtered | 0.99 | 0.06 | 0.22 | 0.91 | 1.00 | 0.06 | 0.09 | 0.91 |
| Age & sex adjusted, using entire historic period, filtered | 1.00 | 0.06 | 0.22 | 0.91 | 1.00 | 0.06 | 0.08 | 0.91 |
| Unadjusted, using TaR after historic visit, filtered | 1.00 | 0.06 | 0.19 | 0.91 | 1.00 | 0.06 | 0.08 | 0.91 |
| Age & sex adjusted, using TaR after historic visit, filtered | 1.00 | 0.06 | 0.19 | 0.91 | 1.00 | 0.06 | 0.08 | 0.91 |
| SCCS / SCRI | | | | | | | | |
| Unadjusted SCCS excluding pre−vaccination window | 1.00 | 0.03 | 0.06 | 0.96 | 1.00 | 0.00 | 0.00 | 1.00 |
| Age & season adjusted SCCS excluding pre−vaccination window | 1.00 | 0.03 | 0.06 | 0.96 | 1.00 | 0.00 | 0.00 | 1.00 |
| SCRI with prior control interval | 1.00 | 0.01 | 0.02 | 0.97 | 1.00 | 0.00 | 0.00 | 1.00 |
| SCRI with posterior control interval | 1.00 | 0.01 | 3.81 | 0.97 | 1.00 | 0.00 | 0.00 | 1.00 |
| Unadjusted SCCS excluding all pre−vaccination time | 1.00 | 0.02 | 3.94 | 0.97 | 1.00 | 0.00 | 0.00 | 1.00 |

Figure 96: The coverage, mean precision, mean squared error (MSE), and fraction non-estimable based on the negative control effect-size estimate at the end of the study period in the MDCR database for Seasonal flu vaccination (Fluvirin).

| Case−control | Uncalibrated Coverage | Uncalibrated Mean precision | Uncalibrated MSE | Uncalibrated Non estimable | Calibrated Coverage | Calibrated Mean precision | Calibrated MSE | Calibrated Non estimable |
|---|---|---|---|---|---|---|---|---|
| Age & sex matched controls | 1.00 | 0.61 | 0.28 | 0.57 | 1.00 | 0.60 | 0.22 | 0.57 |
| Age & sex adjusted, using random controls | 0.97 | 0.66 | 0.28 | 0.54 | 0.99 | 0.65 | 0.26 | 0.54 |
| Cohort method | | | | | | | | |
| Unadjusted, using outpatient visits as comparator | 0.70 | 1.77 | 1.69 | 0.35 | 0.97 | 1.13 | 0.42 | 0.35 |
| PS matching, using outpatient visits as comparator | 0.95 | 1.03 | 0.96 | 0.45 | 0.98 | 1.02 | 0.22 | 0.45 |
| Unadjusted, using random days as comparator | 0.90 | 1.16 | 0.52 | 0.44 | 0.99 | 1.06 | 0.26 | 0.44 |
| PS matching, using random days as comparator | 0.99 | 1.28 | 0.70 | 0.38 | 1.00 | 1.28 | 0.23 | 0.38 |
| PS stratification, using outpatient visits as comparator | 0.90 | 1.69 | 0.83 | 0.35 | 0.96 | 1.65 | 0.39 | 0.35 |
| PS stratification, using random days as comparator | 0.96 | 1.87 | 0.52 | 0.35 | 0.98 | 1.86 | 0.17 | 0.35 |
| PS weighting, using outpatient visits as comparator | 0.76 | 1.70 | 1.32 | 0.38 | 0.95 | 0.99 | 0.70 | 0.38 |
| PS weighting, using random days as comparator | 1.00 | 0.56 | 0.54 | 0.35 | 1.00 | 0.56 | 0.20 | 0.35 |
| Per−month PS matching, using outpatient visits as comparator | | | | | | | | |
| Per−month PS matching, using random days as comparator | | | | | | | | |
| Historical comparator | | | | | | | | |
| Unadjusted, using entire historic period | 0.86 | 2.40 | 0.39 | 0.34 | 0.98 | 0.77 | 0.34 | 0.34 |
| Age & sex adjusted, using entire historic period | 0.95 | 2.39 | 0.31 | 0.34 | 0.98 | 0.85 | 0.30 | 0.34 |
| Unadjusted, using TaR after historic visit | 0.88 | 2.38 | 0.36 | 0.34 | 0.98 | 0.75 | 0.36 | 0.34 |
| Age & sex adjusted, using TaR after historic visit | 0.88 | 2.37 | 0.38 | 0.34 | 0.98 | 0.77 | 0.37 | 0.34 |
| Unadjusted, using entire historic period, filtered | 0.88 | 2.15 | 0.21 | 0.37 | 0.97 | 2.15 | 0.19 | 0.37 |
| Age & sex adjusted, using entire historic period, filtered | 0.97 | 2.15 | 0.17 | 0.37 | 0.97 | 2.14 | 0.18 | 0.37 |
| Unadjusted, using TaR after historic visit, filtered | 0.90 | 2.14 | 0.20 | 0.37 | 0.97 | 1.93 | 0.20 | 0.37 |
| Age & sex adjusted, using TaR after historic visit, filtered | 0.90 | 2.14 | 0.26 | 0.37 | 0.98 | 1.52 | 0.23 | 0.37 |
| SCCS / SCRI | | | | | | | | |
| Unadjusted SCCS excluding pre−vaccination window | 0.96 | 2.08 | 0.26 | 0.34 | 0.98 | 2.06 | 0.25 | 0.34 |
| Age & season adjusted SCCS excluding pre−vaccination window | 0.97 | 1.74 | 0.25 | 0.38 | 0.98 | 1.73 | 0.24 | 0.38 |
| SCRI with prior control interval | 0.97 | 0.98 | 57.53 | 0.45 | 0.99 | 0.97 | 0.25 | 0.45 |
| SCRI with posterior control interval | 0.95 | 1.19 | 4.22 | 0.46 | 1.00 | 1.18 | 0.31 | 0.46 |
| Unadjusted SCCS excluding all pre−vaccination time | 0.95 | 1.95 | 0.30 | 0.35 | 0.96 | 1.93 | 0.28 | 0.35 |

Figure 97: The coverage, mean precision, mean squared error (MSE), and fraction non-estimable based on the negative control effect-size estimate at the end of the study period in the CCAE database for Seasonal flu vaccination (Fluvirin).

#### 13.3 Confidence-interval-based metrics for Seasonal flu vaccination (Fluzone)

| | Uncalibrated | | | | Calibrated | | | |
|---|---|---|---|---|---|---|---|---|
| Case−control | Coverage | Mean precision | MSE | Non estimable | Coverage | Mean precision | MSE | Non estimable |
| Age & sex matched controls | 0.81 | 1.64 | 48.65 | 0.30 | 0.99 | 1.34 | 46.63 | 0.30 |
| Age & sex adjusted, using random controls | 0.75 | 1.92 | 0.89 | 0.26 | 0.99 | 1.38 | 0.39 | 0.26 |
| Cohort method | | | | | | | | |
| Unadjusted, using outpatient visits as comparator | 0.60 | 6.78 | 0.95 | 0.08 | 0.96 | 2.17 | 0.51 | 0.08 |
| PS matching, using outpatient visits as comparator | 0.82 | 3.06 | 0.80 | 0.20 | 0.95 | 3.05 | 0.34 | 0.20 |
| Unadjusted, using random days as comparator | 0.61 | 4.06 | 0.75 | 0.18 | 0.95 | 2.02 | 0.46 | 0.18 |
| PS matching, using random days as comparator | | | | | | | | |
| PS stratification, using outpatient visits as comparator | 0.71 | 5.64 | 0.69 | 0.09 | 0.95 | 3.59 | 0.35 | 0.09 |
| PS stratification, using random days as comparator | 0.66 | 7.31 | 0.76 | 0.10 | 0.97 | 2.46 | 0.40 | 0.10 |
| PS weighting, using outpatient visits as comparator | 0.69 | 3.67 | 1.02 | 0.12 | 0.96 | 2.20 | 0.51 | 0.12 |
| PS weighting, using random days as comparator | 1.00 | 0.00 | 13.14 | 1.00 | 1.00 | 0.00 | 0.00 | 1.00 |
| Per−month PS matching, using outpatient visits as comparator | | | | | | | | |
| Per−month PS matching, using random days as comparator | | | | | | | | |
| Historical comparator | | | | | | | | |
| Unadjusted, using entire historic period | 0.43 | 10.56 | 1.02 | 0.08 | 0.95 | 2.04 | 0.44 | 0.08 |
| Age & sex adjusted, using entire historic period | 0.45 | 10.55 | 0.86 | 0.08 | 0.95 | 2.07 | 0.40 | 0.08 |
| Unadjusted, using TaR after historic visit | 0.70 | 10.38 | 0.43 | 0.08 | 0.96 | 2.09 | 0.43 | 0.08 |
| Age & sex adjusted, using TaR after historic visit | 0.71 | 10.32 | 0.39 | 0.08 | 0.97 | 2.15 | 0.39 | 0.08 |
| Unadjusted, using entire historic period, filtered | 0.44 | 9.98 | 0.94 | 0.09 | 0.95 | 2.37 | 0.40 | 0.09 |
| Age & sex adjusted, using entire historic period, filtered | 0.46 | 9.98 | 0.79 | 0.09 | 0.95 | 2.43 | 0.36 | 0.09 |
| Unadjusted, using TaR after historic visit, filtered | 0.71 | 9.82 | 0.40 | 0.09 | 0.95 | 2.42 | 0.40 | 0.09 |
| Age & sex adjusted, using TaR after historic visit, filtered | 0.72 | 9.77 | 0.35 | 0.09 | 0.95 | 2.52 | 0.36 | 0.09 |
| SCCS / SCRI | | | | | | | | |
| Unadjusted SCCS excluding pre−vaccination window | 0.80 | 8.63 | 0.30 | 0.08 | 0.97 | 3.82 | 0.27 | 0.08 |
| Age & season adjusted SCCS excluding pre−vaccination window | 0.77 | 7.00 | 0.33 | 0.13 | 0.96 | 2.95 | 0.29 | 0.13 |
| SCRI with prior control interval | 0.74 | 3.78 | 28.95 | 0.14 | 0.96 | 1.98 | 0.35 | 0.14 |
| SCRI with posterior control interval | 0.84 | 4.09 | 14.33 | 0.15 | 0.95 | 3.37 | 0.29 | 0.15 |
| Unadjusted SCCS excluding all pre−vaccination time | 0.76 | 7.37 | 0.38 | 0.08 | 0.95 | 4.13 | 0.30 | 0.08 |

Figure 98: The coverage, mean precision, mean squared error (MSE), and fraction non-estimable based on the negative control effect-size estimate at the end of the study period in the Optum EHR database for Seasonal flu vaccination (Fluzone).

| Case−control | Uncalibrated Coverage | Uncalibrated Mean precision | Uncalibrated MSE | Uncalibrated Non estimable | Calibrated Coverage | Calibrated Mean precision | Calibrated MSE | Calibrated Non estimable |
|---|---|---|---|---|---|---|---|---|
| Age & sex matched controls | 1.00 | 0.06 | 15.00 | 0.86 | 1.00 | 0.06 | 0.13 | 0.86 |
| Age & sex adjusted, using random controls | 0.99 | 0.05 | 0.16 | 0.91 | 1.00 | 0.05 | 0.06 | 0.91 |
| Cohort method | | | | | | | | |
| Unadjusted, using outpatient visits as comparator | 1.00 | 0.07 | 0.63 | 0.88 | 1.00 | 0.07 | 0.03 | 0.88 |
| PS matching, using outpatient visits as comparator | 1.00 | 0.03 | 0.39 | 0.92 | 1.00 | 0.03 | 0.05 | 0.92 |
| Unadjusted, using random days as comparator | 0.99 | 0.05 | 0.30 | 0.91 | 1.00 | 0.05 | 0.08 | 0.91 |
| PS matching, using random days as comparator | | | | | | | | |
| PS stratification, using outpatient visits as comparator | 0.99 | 0.12 | 0.79 | 0.80 | 1.00 | 0.12 | 0.30 | 0.80 |
| PS stratification, using random days as comparator | 0.99 | 0.16 | 0.57 | 0.81 | 1.00 | 0.16 | 0.09 | 0.81 |
| PS weighting, using outpatient visits as comparator | 1.00 | 0.07 | 0.62 | 0.80 | 1.00 | 0.07 | 0.21 | 0.80 |
| PS weighting, using random days as comparator | 1.00 | 0.00 | 0.00 | 1.00 | 1.00 | 0.00 | 0.00 | 1.00 |
| Per−month PS matching, using outpatient visits as comparator | | | | | | | | |
| Per−month PS matching, using random days as comparator | | | | | | | | |
| Historical comparator | | | | | | | | |
| Unadjusted, using entire historic period | 0.90 | 0.27 | 1.24 | 0.75 | 1.00 | 0.16 | 0.35 | 0.75 |
| Age & sex adjusted, using entire historic period | 0.96 | 0.27 | 0.58 | 0.75 | 0.98 | 0.26 | 0.23 | 0.75 |
| Unadjusted, using TaR after historic visit | 0.91 | 0.27 | 0.90 | 0.75 | 0.99 | 0.17 | 0.31 | 0.75 |
| Age & sex adjusted, using TaR after historic visit | 0.97 | 0.27 | 0.32 | 0.75 | 1.00 | 0.26 | 0.23 | 0.75 |
| Unadjusted, using entire historic period, filtered | 0.91 | 0.26 | 1.07 | 0.76 | 0.99 | 0.17 | 0.31 | 0.76 |
| Age & sex adjusted, using entire historic period, filtered | 0.97 | 0.26 | 0.51 | 0.76 | 0.99 | 0.25 | 0.21 | 0.76 |
| Unadjusted, using TaR after historic visit, filtered | 0.92 | 0.25 | 0.76 | 0.76 | 0.99 | 0.20 | 0.27 | 0.76 |
| Age & sex adjusted, using TaR after historic visit, filtered | 0.98 | 0.25 | 0.28 | 0.76 | 0.99 | 0.25 | 0.21 | 0.76 |
| SCCS / SCRI | | | | | | | | |
| Unadjusted SCCS excluding pre−vaccination window | 0.99 | 0.20 | 0.17 | 0.80 | 0.99 | 0.20 | 0.15 | 0.80 |
| Age & season adjusted SCCS excluding pre−vaccination window | 0.99 | 0.20 | 0.17 | 0.80 | 1.00 | 0.20 | 0.15 | 0.80 |
| SCRI with prior control interval | 0.99 | 0.08 | 43.71 | 0.86 | 1.00 | 0.08 | 0.12 | 0.86 |
| SCRI with posterior control interval | 1.00 | 0.11 | 0.07 | 0.84 | 1.00 | 0.10 | 0.07 | 0.84 |
| Unadjusted SCCS excluding all pre−vaccination time | 0.99 | 0.20 | 0.18 | 0.80 | 1.00 | 0.19 | 0.16 | 0.80 |

Figure 99: The coverage, mean precision, mean squared error (MSE), and fraction non-estimable based on the negative control effect-size estimate at the end of the study period in the MDCD database for Seasonal flu vaccination (Fluzone).

| | Uncalibrated | | | | Calibrated | | | |
|---|---|---|---|---|---|---|---|---|
| Case−control | Coverage | Mean precision | MSE | Non estimable | Coverage | Mean precision | MSE | Non estimable |
| Age & sex matched controls | 0.95 | 0.90 | 0.28 | 0.53 | 0.97 | 0.89 | 0.26 | 0.53 |
| Age & sex adjusted, using random controls | 0.94 | 1.03 | 0.34 | 0.42 | 0.98 | 0.88 | 0.34 | 0.42 |
| Cohort method | | | | | | | | |
| Unadjusted, using outpatient visits as comparator | 0.81 | 0.91 | 1.45 | 0.44 | 0.99 | 0.64 | 0.40 | 0.44 |
| PS matching, using outpatient visits as comparator | 0.98 | 0.52 | 0.93 | 0.60 | 1.00 | 0.51 | 0.15 | 0.60 |
| Unadjusted, using random days as comparator | 0.97 | 0.74 | 0.59 | 0.52 | 0.99 | 0.72 | 0.22 | 0.52 |
| PS matching, using random days as comparator | 0.99 | 0.61 | 0.70 | 0.52 | 0.99 | 0.60 | 0.21 | 0.52 |
| PS stratification, using outpatient visits as comparator | 0.94 | 0.92 | 0.87 | 0.39 | 0.97 | 0.91 | 0.34 | 0.39 |
| PS stratification, using random days as comparator | 0.96 | 0.93 | 0.78 | 0.41 | 0.98 | 0.91 | 0.36 | 0.41 |
| PS weighting, using outpatient visits as comparator | 0.91 | 0.73 | 1.00 | 0.43 | 0.99 | 0.73 | 0.40 | 0.43 |
| PS weighting, using random days as comparator | 1.00 | 0.28 | 1.31 | 0.41 | 0.99 | 0.28 | 0.95 | 0.42 |
| Per−month PS matching, using outpatient visits as comparator | | | | | | | | |
| Per−month PS matching, using random days as comparator | | | | | | | | |
| Historical comparator | | | | | | | | |
| Unadjusted, using entire historic period | 0.91 | 1.39 | 0.49 | 0.38 | 0.97 | 0.72 | 0.49 | 0.38 |
| Age & sex adjusted, using entire historic period | 0.91 | 1.39 | 0.48 | 0.38 | 0.97 | 0.74 | 0.47 | 0.38 |
| Unadjusted, using TaR after historic visit | 0.88 | 1.36 | 0.61 | 0.38 | 0.97 | 0.65 | 0.60 | 0.38 |
| Age & sex adjusted, using TaR after historic visit | 0.89 | 1.35 | 0.59 | 0.38 | 0.96 | 0.69 | 0.59 | 0.38 |
| Unadjusted, using entire historic period, filtered | 0.94 | 1.28 | 0.40 | 0.40 | 0.96 | 1.16 | 0.40 | 0.40 |
| Age & sex adjusted, using entire historic period, filtered | 0.94 | 1.28 | 0.39 | 0.40 | 0.96 | 1.19 | 0.40 | 0.40 |
| Unadjusted, using TaR after historic visit, filtered | 0.90 | 1.25 | 0.51 | 0.40 | 0.95 | 0.98 | 0.49 | 0.40 |
| Age & sex adjusted, using TaR after historic visit, filtered | 0.91 | 1.25 | 0.49 | 0.40 | 0.95 | 1.04 | 0.48 | 0.40 |
| SCCS / SCRI | | | | | | | | |
| Unadjusted SCCS excluding pre−vaccination window | 0.96 | 1.11 | 0.26 | 0.45 | 0.97 | 1.11 | 0.25 | 0.45 |
| Age & season adjusted SCCS excluding pre−vaccination window | 0.97 | 1.09 | 0.23 | 0.46 | 0.98 | 1.08 | 0.23 | 0.46 |
| SCRI with prior control interval | 0.97 | 0.49 | 44.75 | 0.60 | 0.98 | 0.47 | 0.23 | 0.60 |
| SCRI with posterior control interval | 0.95 | 0.66 | 15.20 | 0.54 | 1.00 | 0.65 | 0.29 | 0.54 |
| Unadjusted SCCS excluding all pre−vaccination time | 0.95 | 1.07 | 0.28 | 0.45 | 0.98 | 1.07 | 0.28 | 0.45 |

Figure 100: The coverage, mean precision, mean squared error (MSE), and fraction non-estimable based on the negative control effect-size estimate at the end of the study period in the MDCR database for Seasonal flu vaccination (Fluzone).

| Case−control | Uncalibrated Coverage | Uncalibrated Mean precision | Uncalibrated MSE | Uncalibrated Non estimable | Calibrated Coverage | Calibrated Mean precision | Calibrated MSE | Calibrated Non estimable |
|---|---|---|---|---|---|---|---|---|
| Age & sex matched controls | 1.00 | 0.00 | 0.00 | 1.00 | 1.00 | 0.00 | 0.00 | 1.00 |
| Age & sex adjusted, using random controls | 1.00 | 0.00 | 0.00 | 1.00 | 1.00 | 0.00 | 0.00 | 1.00 |
| Cohort method | | | | | | | | |
| Unadjusted, using outpatient visits as comparator | 1.00 | 0.00 | 0.29 | 0.99 | 1.00 | 0.00 | 0.00 | 1.00 |
| PS matching, using outpatient visits as comparator | 1.00 | 0.01 | 0.18 | 0.98 | 1.00 | 0.00 | 0.00 | 1.00 |
| Unadjusted, using random days as comparator | 1.00 | 0.00 | 0.20 | 1.00 | 1.00 | 0.00 | 0.00 | 1.00 |
| PS matching, using random days as comparator | 1.00 | 0.02 | 0.28 | 0.96 | 1.00 | 0.00 | 0.00 | 1.00 |
| PS stratification, using outpatient visits as comparator | 1.00 | 0.03 | 0.52 | 0.91 | 1.00 | 0.03 | 0.17 | 0.91 |
| PS stratification, using random days as comparator | 1.00 | 0.02 | 0.32 | 0.95 | 1.00 | 0.00 | 0.00 | 1.00 |
| PS weighting, using outpatient visits as comparator | 1.00 | 0.02 | 0.42 | 0.92 | 1.00 | 0.02 | 0.08 | 0.92 |
| PS weighting, using random days as comparator | 1.00 | 0.00 | 0.25 | 0.95 | 1.00 | 0.00 | 0.00 | 1.00 |
| Per−month PS matching, using outpatient visits as comparator | | | | | | | | |
| Per−month PS matching, using random days as comparator | | | | | | | | |
| Historical comparator | | | | | | | | |
| Unadjusted, using entire historic period | 0.94 | 0.07 | 1.37 | 0.88 | 0.99 | 0.05 | 0.33 | 0.88 |
| Age & sex adjusted, using entire historic period | 0.97 | 0.07 | 0.93 | 0.88 | 0.98 | 0.06 | 0.25 | 0.88 |
| Unadjusted, using TaR after historic visit | 0.95 | 0.07 | 1.09 | 0.88 | 0.99 | 0.06 | 0.30 | 0.88 |
| Age & sex adjusted, using TaR after historic visit | 0.98 | 0.07 | 0.73 | 0.88 | 0.98 | 0.07 | 0.23 | 0.88 |
| Unadjusted, using entire historic period, filtered | 0.95 | 0.07 | 1.22 | 0.89 | 0.99 | 0.04 | 0.32 | 0.89 |
| Age & sex adjusted, using entire historic period, filtered | 0.97 | 0.07 | 0.85 | 0.89 | 0.99 | 0.05 | 0.25 | 0.89 |
| Unadjusted, using TaR after historic visit, filtered | 0.96 | 0.07 | 0.98 | 0.89 | 0.99 | 0.05 | 0.29 | 0.89 |
| Age & sex adjusted, using TaR after historic visit, filtered | 0.98 | 0.07 | 0.68 | 0.89 | 0.99 | 0.06 | 0.23 | 0.89 |
| SCCS / SCRI | | | | | | | | |
| Unadjusted SCCS excluding pre−vaccination window | 1.00 | 0.01 | 0.04 | 0.97 | 1.00 | 0.00 | 0.00 | 1.00 |
| Age & season adjusted SCCS excluding pre−vaccination window | 1.00 | 0.01 | 0.05 | 0.97 | 1.00 | 0.00 | 0.00 | 1.00 |
| SCRI with prior control interval | | | | | | | | |
| SCRI with posterior control interval | | | | | | | | |
| Unadjusted SCCS excluding all pre−vaccination time | 1.00 | 0.01 | 0.03 | 0.97 | 1.00 | 0.00 | 0.00 | 1.00 |

Figure 101: The coverage, mean precision, mean squared error (MSE), and fraction non-estimable based on the negative control effect-size estimate at the end of the study period in the CCAE database for Seasonal flu vaccination (Fluzone).

#### 13.4 Confidence-interval-based metrics for Seasonal flu vaccination (All)

| | Uncalibrated | | | | Calibrated | | | |
|---|---|---|---|---|---|---|---|---|
| Case−control | Coverage | Mean precision | MSE | Non estimable | Coverage | Mean precision | MSE | Non estimable |
| Age & sex matched controls | 0.38 | 12.24 | 0.70 | 0.05 | 0.94 | 6.60 | 0.14 | 0.05 |
| Age & sex adjusted, using random controls | 0.33 | 14.93 | 0.65 | 0.04 | 0.94 | 9.54 | 0.12 | 0.04 |
| Cohort method | | | | | | | | |
| Unadjusted, using outpatient visits as comparator | 0.35 | 32.94 | 0.45 | 0.01 | 0.95 | 5.28 | 0.18 | 0.01 |
| PS matching, using outpatient visits as comparator | 0.60 | 21.16 | 0.41 | 0.03 | 0.92 | 9.35 | 0.19 | 0.03 |
| Unadjusted, using random days as comparator | 0.29 | 18.56 | 0.75 | 0.03 | 0.99 | 6.00 | 0.19 | 0.03 |
| PS matching, using random days as comparator | | | | | | | | |
| PS stratification, using outpatient visits as comparator | 0.48 | 28.66 | 0.36 | 0.01 | 0.91 | 9.08 | 0.17 | 0.01 |
| PS stratification, using random days as comparator | 0.19 | 26.91 | 0.79 | 0.01 | 0.97 | 7.38 | 0.13 | 0.01 |
| PS weighting, using outpatient visits as comparator | 0.49 | 13.43 | 0.48 | 0.01 | 0.95 | 8.16 | 0.26 | 0.01 |
| PS weighting, using random days as comparator | 1.00 | 0.00 | 23.17 | 1.00 | 1.00 | 0.00 | 0.00 | 1.00 |
| Per−month PS matching, using outpatient visits as comparator | | | | | | | | |
| Per−month PS matching, using random days as comparator | | | | | | | | |
| Historical comparator | | | | | | | | |
| Unadjusted, using entire historic period | 0.12 | 90.76 | 0.98 | 0.00 | 0.99 | 6.20 | 0.17 | 0.00 |
| Age & sex adjusted, using entire historic period | 0.12 | 90.50 | 0.92 | 0.00 | 0.99 | 7.44 | 0.14 | 0.00 |
| Unadjusted, using TaR after historic visit | 0.45 | 79.37 | 0.21 | 0.00 | 0.96 | 5.89 | 0.18 | 0.00 |
| Age & sex adjusted, using TaR after historic visit | 0.44 | 78.42 | 0.19 | 0.00 | 0.99 | 7.23 | 0.14 | 0.00 |
| Unadjusted, using entire historic period, filtered | 0.13 | 84.07 | 0.88 | 0.01 | 0.97 | 10.22 | 0.12 | 0.01 |
| Age & sex adjusted, using entire historic period, filtered | 0.13 | 83.84 | 0.81 | 0.01 | 0.96 | 15.62 | 0.09 | 0.01 |
| Unadjusted, using TaR after historic visit, filtered | 0.46 | 74.03 | 0.15 | 0.01 | 0.96 | 8.96 | 0.13 | 0.01 |
| Age & sex adjusted, using TaR after historic visit, filtered | 0.45 | 73.16 | 0.13 | 0.01 | 0.94 | 14.93 | 0.09 | 0.01 |
| SCCS / SCRI | | | | | | | | |
| Unadjusted SCCS excluding pre−vaccination window | 0.61 | 78.64 | 0.10 | 0.00 | 0.97 | 14.28 | 0.10 | 0.00 |
| Age & season adjusted SCCS excluding pre−vaccination window | 0.51 | 49.23 | 0.14 | 0.08 | 0.96 | 8.60 | 0.12 | 0.08 |
| SCRI with prior control interval | 0.43 | 31.65 | 18.65 | 0.01 | 0.95 | 7.87 | 4.62 | 0.01 |
| SCRI with posterior control interval | 0.60 | 38.69 | 0.21 | 0.01 | 0.97 | 10.94 | 0.15 | 0.01 |
| Unadjusted SCCS excluding all pre−vaccination time | 0.51 | 70.99 | 0.14 | 0.00 | 0.97 | 13.11 | 0.10 | 0.00 |

Figure 102: The coverage, mean precision, mean squared error (MSE), and fraction non-estimable based on the negative control effect-size estimate at the end of the study period in the Optum EHR database for Seasonal flu vaccination (All).

| | Uncalibrated | | | | Calibrated | | | |
|---|---|---|---|---|---|---|---|---|
| Case−control | Coverage | Mean precision | MSE | Non estimable | Coverage | Mean precision | MSE | Non estimable |
| Age & sex matched controls | 0.60 | 9.03 | 0.34 | 0.23 | 0.96 | 4.47 | 0.18 | 0.23 |
| Age & sex adjusted, using random controls | 0.53 | 14.63 | 0.31 | 0.13 | 0.95 | 5.89 | 0.11 | 0.13 |
| Cohort method | | | | | | | | |
| Unadjusted, using outpatient visits as comparator | 0.28 | 16.72 | 0.81 | 0.13 | 0.91 | 2.48 | 0.26 | 0.13 |
| PS matching, using outpatient visits as comparator | 0.83 | 9.62 | 0.13 | 0.15 | 0.96 | 9.41 | 0.08 | 0.15 |
| Unadjusted, using random days as comparator | 0.58 | 10.55 | 0.47 | 0.14 | 0.96 | 4.35 | 0.22 | 0.14 |
| PS matching, using random days as comparator | 0.89 | 7.71 | 0.20 | 0.13 | 0.97 | 5.98 | 0.14 | 0.13 |
| PS stratification, using outpatient visits as comparator | 0.73 | 12.63 | 0.21 | 0.13 | 0.92 | 10.39 | 0.15 | 0.13 |
| PS stratification, using random days as comparator | 0.86 | 9.81 | 0.24 | 0.13 | 0.99 | 5.50 | 0.19 | 0.13 |
| PS weighting, using outpatient visits as comparator | 0.77 | 12.60 | 0.19 | 0.13 | 0.94 | 5.77 | 0.12 | 0.13 |
| PS weighting, using random days as comparator | 0.73 | 7.48 | 0.48 | 0.13 | 0.94 | 4.29 | 0.36 | 0.13 |
| Per−month PS matching, using outpatient visits as comparator | | | | | | | | |
| Per−month PS matching, using random days as comparator | | | | | | | | |
| Historical comparator | | | | | | | | |
| Unadjusted, using entire historic period | 0.57 | 28.61 | 0.32 | 0.13 | 0.98 | 2.82 | 0.21 | 0.13 |
| Age & sex adjusted, using entire historic period | 0.46 | 28.57 | 0.41 | 0.13 | 0.98 | 2.89 | 0.20 | 0.13 |
| Unadjusted, using TaR after historic visit | 0.61 | 24.99 | 0.24 | 0.13 | 0.99 | 2.60 | 0.24 | 0.13 |
| Age & sex adjusted, using TaR after historic visit | 0.72 | 24.83 | 0.22 | 0.13 | 0.99 | 2.80 | 0.22 | 0.13 |
| Unadjusted, using entire historic period, filtered | 0.58 | 26.84 | 0.19 | 0.14 | 0.96 | 6.05 | 0.10 | 0.14 |
| Age & sex adjusted, using entire historic period, filtered | 0.47 | 26.81 | 0.27 | 0.14 | 0.96 | 6.33 | 0.09 | 0.14 |
| Unadjusted, using TaR after historic visit, filtered | 0.62 | 23.76 | 0.14 | 0.14 | 0.94 | 4.94 | 0.13 | 0.14 |
| Age & sex adjusted, using TaR after historic visit, filtered | 0.73 | 23.62 | 0.11 | 0.14 | 0.95 | 6.42 | 0.11 | 0.14 |
| SCCS / SCRI | | | | | | | | |
| Unadjusted SCCS excluding pre−vaccination window | 0.86 | 25.96 | 0.08 | 0.13 | 0.95 | 12.77 | 0.08 | 0.13 |
| Age & season adjusted SCCS excluding pre−vaccination window | 0.90 | 21.28 | 0.08 | 0.16 | 0.94 | 9.88 | 0.08 | 0.16 |
| SCRI with prior control interval | 0.84 | 12.60 | 0.12 | 0.13 | 0.97 | 4.76 | 0.12 | 0.13 |
| SCRI with posterior control interval | 0.86 | 15.26 | 0.12 | 0.13 | 0.96 | 7.36 | 0.12 | 0.13 |
| Unadjusted SCCS excluding all pre−vaccination time | 0.83 | 24.53 | 0.10 | 0.13 | 0.97 | 9.00 | 0.10 | 0.13 |

Figure 103: The coverage, mean precision, mean squared error (MSE), and fraction non-estimable based on the negative control effect-size estimate at the end of the study period in the MDCD database for Seasonal flu vaccination (All).

| Case−control | Uncalibrated Coverage | Uncalibrated Mean precision | Uncalibrated MSE | Uncalibrated Non estimable | Calibrated Coverage | Calibrated Mean precision | Calibrated MSE | Calibrated Non estimable |
|---|---|---|---|---|---|---|---|---|
| Age & sex matched controls | 0.98 | 5.52 | 0.13 | 0.32 | 0.98 | 5.34 | 0.13 | 0.32 |
| Age & sex adjusted, using random controls | 0.88 | 8.60 | 0.14 | 0.14 | 0.99 | 6.90 | 0.10 | 0.14 |
| Cohort method | | | | | | | | |
| Unadjusted, using outpatient visits as comparator | 0.53 | 6.47 | 0.88 | 0.17 | 0.96 | 2.75 | 0.32 | 0.17 |
| PS matching, using outpatient visits as comparator | 0.87 | 3.91 | 0.63 | 0.24 | 0.96 | 3.59 | 0.25 | 0.24 |
| Unadjusted, using random days as comparator | 0.87 | 4.97 | 0.27 | 0.19 | 0.99 | 3.87 | 0.26 | 0.19 |
| PS matching, using random days as comparator | 0.91 | 3.56 | 0.36 | 0.23 | 0.96 | 3.45 | 0.26 | 0.23 |
| PS stratification, using outpatient visits as comparator | 0.80 | 5.64 | 0.50 | 0.18 | 0.96 | 4.46 | 0.29 | 0.18 |
| PS stratification, using random days as comparator | 0.91 | 4.63 | 0.33 | 0.19 | 0.97 | 3.69 | 0.24 | 0.19 |
| PS weighting, using outpatient visits as comparator | 0.62 | 5.31 | 0.77 | 0.18 | 0.98 | 2.68 | 0.36 | 0.18 |
| PS weighting, using random days as comparator | 0.92 | 2.70 | 0.53 | 0.19 | 0.95 | 2.46 | 0.49 | 0.19 |
| Per−month PS matching, using outpatient visits as comparator | | | | | | | | |
| Per−month PS matching, using random days as comparator | | | | | | | | |
| Historical comparator | | | | | | | | |
| Unadjusted, using entire historic period | 0.81 | 11.18 | 0.22 | 0.14 | 0.98 | 3.32 | 0.16 | 0.14 |
| Age & sex adjusted, using entire historic period | 0.81 | 11.18 | 0.22 | 0.14 | 0.98 | 3.32 | 0.16 | 0.14 |
| Unadjusted, using TaR after historic visit | 0.85 | 9.70 | 0.21 | 0.14 | 0.98 | 2.95 | 0.20 | 0.14 |
| Age & sex adjusted, using TaR after historic visit | 0.85 | 9.69 | 0.20 | 0.14 | 0.98 | 3.03 | 0.20 | 0.14 |
| Unadjusted, using entire historic period, filtered | 0.83 | 10.18 | 0.13 | 0.16 | 0.97 | 8.56 | 0.09 | 0.16 |
| Age & sex adjusted, using entire historic period, filtered | 0.83 | 10.18 | 0.13 | 0.16 | 0.97 | 8.63 | 0.09 | 0.16 |
| Unadjusted, using TaR after historic visit, filtered | 0.87 | 8.96 | 0.13 | 0.16 | 0.99 | 5.36 | 0.13 | 0.16 |
| Age & sex adjusted, using TaR after historic visit, filtered | 0.87 | 8.95 | 0.12 | 0.16 | 0.99 | 5.66 | 0.12 | 0.16 |
| SCCS / SCRI | | | | | | | | |
| Unadjusted SCCS excluding pre−vaccination window | 0.90 | 9.38 | 0.15 | 0.16 | 0.98 | 7.51 | 0.15 | 0.16 |
| Age & season adjusted SCCS excluding pre−vaccination window | 0.91 | 7.84 | 0.07 | 0.30 | 0.98 | 5.86 | 0.07 | 0.30 |
| SCRI with prior control interval | 0.85 | 4.20 | 18.13 | 0.20 | 0.97 | 2.84 | 0.19 | 0.20 |
| SCRI with posterior control interval | 0.94 | 5.36 | 15.49 | 0.19 | 0.99 | 4.16 | 0.18 | 0.19 |
| Unadjusted SCCS excluding all pre−vaccination time | 0.88 | 8.69 | 0.23 | 0.16 | 0.98 | 4.70 | 0.22 | 0.16 |

Figure 104: The coverage, mean precision, mean squared error (MSE), and fraction non-estimable based on the negative control effect-size estimate at the end of the study period in the MDCR database for Seasonal flu vaccination (All).

| | Uncalibrated | | | | Calibrated | | | |
|---|---|---|---|---|---|---|---|---|
| Case−control | Coverage | Mean precision | MSE | Non estimable | Coverage | Mean precision | MSE | Non estimable |
| Age & sex matched controls | 0.72 | 11.55 | 0.22 | 0.14 | 0.96 | 7.45 | 0.16 | 0.14 |
| Age & sex adjusted, using random controls | 0.73 | 14.63 | 0.15 | 0.12 | 0.95 | 11.30 | 0.13 | 0.12 |
| Cohort method | | | | | | | | |
| Unadjusted, using outpatient visits as comparator | 0.26 | 18.02 | 1.44 | 0.12 | 0.92 | 2.20 | 0.37 | 0.12 |
| PS matching, using outpatient visits as comparator | 0.76 | 11.27 | 0.28 | 0.13 | 0.92 | 7.38 | 0.14 | 0.13 |
| Unadjusted, using random days as comparator | 0.67 | 12.28 | 0.26 | 0.12 | 0.95 | 4.98 | 0.15 | 0.12 |
| PS matching, using random days as comparator | 0.88 | 10.69 | 0.34 | 0.13 | 0.95 | 7.86 | 0.21 | 0.13 |
| PS stratification, using outpatient visits as comparator | 0.54 | 15.98 | 0.35 | 0.12 | 0.97 | 6.97 | 0.15 | 0.12 |
| PS stratification, using random days as comparator | 0.85 | 13.70 | 0.24 | 0.13 | 0.96 | 8.96 | 0.13 | 0.13 |
| PS weighting, using outpatient visits as comparator | 0.77 | 6.60 | 0.31 | 0.12 | 0.97 | 5.31 | 0.13 | 0.12 |
| PS weighting, using random days as comparator | 0.97 | 6.83 | 0.29 | 0.13 | 0.98 | 6.70 | 0.22 | 0.13 |
| Per−month PS matching, using outpatient visits as comparator | | | | | | | | |
| Per−month PS matching, using random days as comparator | | | | | | | | |
| Historical comparator | | | | | | | | |
| Unadjusted, using entire historic period | 0.56 | 50.06 | 0.29 | 0.10 | 0.97 | 3.24 | 0.25 | 0.10 |
| Age & sex adjusted, using entire historic period | 0.56 | 49.93 | 0.25 | 0.10 | 0.98 | 3.62 | 0.22 | 0.10 |
| Unadjusted, using TaR after historic visit | 0.53 | 42.23 | 0.29 | 0.10 | 0.98 | 2.74 | 0.28 | 0.10 |
| Age & sex adjusted, using TaR after historic visit | 0.58 | 41.91 | 0.26 | 0.10 | 0.98 | 3.03 | 0.26 | 0.10 |
| Unadjusted, using entire historic period, filtered | 0.58 | 43.78 | 0.15 | 0.12 | 0.95 | 9.34 | 0.13 | 0.12 |
| Age & sex adjusted, using entire historic period, filtered | 0.58 | 43.68 | 0.11 | 0.12 | 0.96 | 14.09 | 0.10 | 0.12 |
| Unadjusted, using TaR after historic visit, filtered | 0.55 | 37.68 | 0.18 | 0.12 | 0.96 | 5.41 | 0.16 | 0.12 |
| Age & sex adjusted, using TaR after historic visit, filtered | 0.60 | 37.41 | 0.14 | 0.12 | 0.97 | 7.52 | 0.13 | 0.12 |
| SCCS / SCRI | | | | | | | | |
| Unadjusted SCCS excluding pre−vaccination window | 0.73 | 44.73 | 0.09 | 0.10 | 0.92 | 18.20 | 0.09 | 0.10 |
| Age & season adjusted SCCS excluding pre−vaccination window | 0.76 | 28.80 | 0.09 | 0.17 | 0.94 | 13.65 | 0.09 | 0.17 |
| SCRI with prior control interval | 0.67 | 20.73 | 4.30 | 0.10 | 0.95 | 5.42 | 4.34 | 0.10 |
| SCRI with posterior control interval | 0.83 | 25.87 | 0.16 | 0.10 | 0.97 | 9.13 | 0.16 | 0.10 |
| Unadjusted SCCS excluding all pre−vaccination time | 0.73 | 41.73 | 0.12 | 0.10 | 0.96 | 12.99 | 0.11 | 0.10 |

Figure 105: The coverage, mean precision, mean squared error (MSE), and fraction non-estimable based on the negative control effect-size estimate at the end of the study period in the CCAE database for Seasonal flu vaccination (All).

#### 13.5 Confidence-interval-based metrics for Zoster vaccination (Shingrix)

| | Uncalibrated | | | | Calibrated | | | |
|---|---|---|---|---|---|---|---|---|
| Case−control | Coverage | Mean precision | MSE | Non estimable | Coverage | Mean precision | MSE | Non estimable |
| Age & sex matched controls | 0.78 | 1.66 | 30.98 | 0.35 | 0.94 | 1.66 | 29.68 | 0.35 |
| Age & sex adjusted, using random controls | 0.80 | 1.79 | 0.51 | 0.33 | 0.95 | 1.59 | 0.33 | 0.33 |
| Cohort method | | | | | | | | |
| Unadjusted, using outpatient visits as comparator | 0.56 | 6.08 | 1.35 | 0.12 | 0.94 | 1.61 | 0.62 | 0.12 |
| PS matching, using outpatient visits as comparator | 0.69 | 4.78 | 0.82 | 0.14 | 0.94 | 4.60 | 0.28 | 0.14 |
| Unadjusted, using random days as comparator | 0.61 | 3.41 | 1.14 | 0.18 | 0.96 | 1.15 | 0.65 | 0.18 |
| PS matching, using random days as comparator | | | | | | | | |
| PS stratification, using outpatient visits as comparator | 0.62 | 6.53 | 0.76 | 0.11 | 0.96 | 4.14 | 0.28 | 0.11 |
| PS stratification, using random days as comparator | | | | | | | | |
| PS weighting, using outpatient visits as comparator | 0.92 | 1.83 | 0.58 | 0.11 | 0.99 | 1.82 | 0.26 | 0.11 |
| PS weighting, using random days as comparator | | | | | | | | |
| Per−month PS matching, using outpatient visits as comparator | | | | | | | | |
| Per−month PS matching, using random days as comparator | | | | | | | | |
| Historical comparator | | | | | | | | |
| Unadjusted, using entire historic period | 0.46 | 9.19 | 1.17 | 0.11 | 0.94 | 1.48 | 0.63 | 0.11 |
| Age & sex adjusted, using entire historic period | 0.58 | 9.18 | 0.65 | 0.11 | 0.96 | 2.43 | 0.37 | 0.11 |
| Unadjusted, using TaR after historic visit | 0.61 | 9.06 | 0.60 | 0.11 | 0.92 | 1.58 | 0.61 | 0.11 |
| Age & sex adjusted, using TaR after historic visit | 0.60 | 8.95 | 0.44 | 0.11 | 0.97 | 2.23 | 0.44 | 0.11 |
| Unadjusted, using entire historic period, filtered | 0.47 | 8.62 | 1.12 | 0.12 | 0.95 | 1.51 | 0.61 | 0.12 |
| Age & sex adjusted, using entire historic period, filtered | 0.59 | 8.61 | 0.61 | 0.12 | 0.97 | 2.67 | 0.35 | 0.12 |
| Unadjusted, using TaR after historic visit, filtered | 0.62 | 8.50 | 0.59 | 0.12 | 0.92 | 1.59 | 0.59 | 0.12 |
| Age & sex adjusted, using TaR after historic visit, filtered | 0.61 | 8.40 | 0.43 | 0.12 | 0.97 | 2.35 | 0.42 | 0.12 |
| SCCS / SCRI | | | | | | | | |
| Unadjusted SCCS excluding pre−vaccination window | 0.88 | 7.78 | 0.24 | 0.11 | 0.97 | 4.36 | 0.24 | 0.11 |
| Age & season adjusted SCCS excluding pre−vaccination window | 0.85 | 7.75 | 0.25 | 0.11 | 0.97 | 4.35 | 0.25 | 0.11 |
| SCRI with prior control interval | 0.90 | 4.30 | 45.63 | 0.14 | 0.96 | 3.17 | 0.26 | 0.14 |
| SCRI with posterior control interval | 0.91 | 3.87 | 30.27 | 0.13 | 0.98 | 3.27 | 0.32 | 0.13 |
| Unadjusted SCCS excluding all pre−vaccination time | 0.84 | 6.13 | 0.33 | 0.11 | 0.98 | 4.50 | 0.32 | 0.11 |

Figure 106: The coverage, mean precision, mean squared error (MSE), and fraction non-estimable based on the negative control effect-size estimate at the end of the study period in the Optum EHR database for Zoster vaccination (Shingrix).

| | Uncalibrated | | | | Calibrated | | | |
|---|---|---|---|---|---|---|---|---|
| Case−control | Coverage | Mean precision | MSE | Non estimable | Coverage | Mean precision | MSE | Non estimable |
| Age & sex matched controls | 0.98 | 0.35 | 15.64 | 0.68 | 0.99 | 0.34 | 0.17 | 0.68 |
| Age & sex adjusted, using random controls | 0.97 | 0.51 | 0.14 | 0.61 | 1.00 | 0.51 | 0.09 | 0.61 |
| Cohort method | | | | | | | | |
| Unadjusted, using outpatient visits as comparator | 0.99 | 0.64 | 0.78 | 0.55 | 0.99 | 0.63 | 0.26 | 0.55 |
| PS matching, using outpatient visits as comparator | 0.98 | 0.36 | 0.75 | 0.66 | 1.00 | 0.36 | 0.15 | 0.66 |
| Unadjusted, using random days as comparator | 0.91 | 0.34 | 0.48 | 0.71 | 0.98 | 0.30 | 0.20 | 0.71 |
| PS matching, using random days as comparator | 0.99 | 0.34 | 0.83 | 0.66 | 1.00 | 0.34 | 0.14 | 0.66 |
| PS stratification, using outpatient visits as comparator | 0.97 | 0.68 | 0.94 | 0.47 | 0.98 | 0.67 | 0.43 | 0.47 |
| PS stratification, using random days as comparator | 1.00 | 0.57 | 0.72 | 0.49 | 1.00 | 0.57 | 0.25 | 0.49 |
| PS weighting, using outpatient visits as comparator | 1.00 | 0.24 | 1.24 | 0.47 | 1.00 | 0.24 | 0.84 | 0.47 |
| PS weighting, using random days as comparator | 1.00 | 0.26 | 1.47 | 0.49 | 1.00 | 0.26 | 1.07 | 0.49 |
| Per−month PS matching, using outpatient visits as comparator | | | | | | | | |
| Per−month PS matching, using random days as comparator | | | | | | | | |
| Historical comparator | | | | | | | | |
| Unadjusted, using entire historic period | 0.76 | 1.15 | 1.14 | 0.48 | 0.97 | 0.52 | 0.33 | 0.48 |
| Age & sex adjusted, using entire historic period | 0.92 | 1.15 | 0.35 | 0.48 | 0.99 | 0.83 | 0.17 | 0.48 |
| Unadjusted, using TaR after historic visit | 0.81 | 1.14 | 0.62 | 0.48 | 0.98 | 0.59 | 0.27 | 0.48 |
| Age & sex adjusted, using TaR after historic visit | 0.98 | 1.13 | 0.19 | 0.48 | 0.99 | 0.95 | 0.19 | 0.48 |
| Unadjusted, using entire historic period, filtered | 0.77 | 1.08 | 1.04 | 0.49 | 0.98 | 0.60 | 0.29 | 0.49 |
| Age & sex adjusted, using entire historic period, filtered | 0.94 | 1.08 | 0.30 | 0.49 | 1.00 | 1.07 | 0.15 | 0.49 |
| Unadjusted, using TaR after historic visit, filtered | 0.82 | 1.07 | 0.56 | 0.49 | 0.98 | 0.68 | 0.24 | 0.49 |
| Age & sex adjusted, using TaR after historic visit, filtered | 0.99 | 1.07 | 0.17 | 0.49 | 1.00 | 1.06 | 0.17 | 0.49 |
| SCCS / SCRI | | | | | | | | |
| Unadjusted SCCS excluding pre−vaccination window | 1.00 | 0.99 | 0.17 | 0.49 | 1.00 | 0.99 | 0.16 | 0.49 |
| Age & season adjusted SCCS excluding pre−vaccination window | 1.00 | 0.97 | 0.18 | 0.49 | 1.00 | 0.97 | 0.17 | 0.49 |
| SCRI with prior control interval | 0.98 | 0.61 | 58.54 | 0.57 | 1.00 | 0.61 | 0.14 | 0.57 |
| SCRI with posterior control interval | 0.99 | 0.56 | 29.74 | 0.57 | 0.99 | 0.56 | 0.18 | 0.57 |
| Unadjusted SCCS excluding all pre−vaccination time | 1.00 | 0.85 | 16.39 | 0.52 | 1.00 | 0.85 | 0.14 | 0.52 |

Figure 107: The coverage, mean precision, mean squared error (MSE), and fraction non-estimable based on the negative control effect-size estimate at the end of the study period in the MDCD database for Zoster vaccination (Shingrix).

| Case−control | Uncalibrated Coverage | Uncalibrated Mean precision | Uncalibrated MSE | Uncalibrated Non estimable | Calibrated Coverage | Calibrated Mean precision | Calibrated MSE | Calibrated Non estimable |
|---|---|---|---|---|---|---|---|---|
| Age & sex matched controls | 0.89 | 1.57 | 0.21 | 0.52 | 0.98 | 0.94 | 0.21 | 0.52 |
| Age & sex adjusted, using random controls | 0.85 | 2.37 | 0.39 | 0.30 | 0.96 | 1.23 | 0.39 | 0.30 |
| Cohort method | | | | | | | | |
| Unadjusted, using outpatient visits as comparator | 0.65 | 2.01 | 1.63 | 0.33 | 0.96 | 0.69 | 0.65 | 0.33 |
| PS matching, using outpatient visits as comparator | 0.94 | 1.37 | 0.79 | 0.41 | 0.98 | 1.31 | 0.22 | 0.41 |
| Unadjusted, using random days as comparator | 0.91 | 1.65 | 0.77 | 0.34 | 0.95 | 1.05 | 0.42 | 0.34 |
| PS matching, using random days as comparator | 0.97 | 1.34 | 0.51 | 0.40 | 0.99 | 1.33 | 0.25 | 0.40 |
| PS stratification, using outpatient visits as comparator | 0.83 | 2.01 | 0.80 | 0.33 | 0.97 | 1.24 | 0.38 | 0.33 |
| PS stratification, using random days as comparator | 0.91 | 1.96 | 0.60 | 0.31 | 0.96 | 1.65 | 0.38 | 0.31 |
| PS weighting, using outpatient visits as comparator | 0.77 | 1.72 | 1.22 | 0.38 | 0.96 | 0.83 | 0.61 | 0.38 |
| PS weighting, using random days as comparator | 1.00 | 0.59 | 0.97 | 0.31 | 0.99 | 0.59 | 0.75 | 0.31 |
| Per−month PS matching, using outpatient visits as comparator | | | | | | | | |
| Per−month PS matching, using random days as comparator | | | | | | | | |
| Historical comparator | | | | | | | | |
| Unadjusted, using entire historic period | 0.81 | 2.89 | 0.45 | 0.30 | 0.98 | 0.93 | 0.45 | 0.30 |
| Age & sex adjusted, using entire historic period | 0.81 | 2.89 | 0.42 | 0.30 | 0.98 | 0.99 | 0.43 | 0.30 |
| Unadjusted, using TaR after historic visit | 0.72 | 2.74 | 0.66 | 0.30 | 0.96 | 0.74 | 0.60 | 0.30 |
| Age & sex adjusted, using TaR after historic visit | 0.74 | 2.74 | 0.63 | 0.30 | 0.97 | 0.77 | 0.58 | 0.30 |
| Unadjusted, using entire historic period, filtered | 0.83 | 2.58 | 0.37 | 0.33 | 0.98 | 1.02 | 0.37 | 0.33 |
| Age & sex adjusted, using entire historic period, filtered | 0.83 | 2.58 | 0.35 | 0.33 | 0.97 | 1.09 | 0.36 | 0.33 |
| Unadjusted, using TaR after historic visit, filtered | 0.75 | 2.47 | 0.56 | 0.33 | 0.97 | 0.79 | 0.47 | 0.33 |
| Age & sex adjusted, using TaR after historic visit, filtered | 0.77 | 2.47 | 0.54 | 0.33 | 0.97 | 0.82 | 0.45 | 0.33 |
| SCCS / SCRI | | | | | | | | |
| Unadjusted SCCS excluding pre−vaccination window | 0.92 | 2.50 | 0.27 | 0.31 | 0.96 | 1.92 | 0.27 | 0.31 |
| Age & season adjusted SCCS excluding pre−vaccination window | 0.95 | 2.41 | 0.23 | 0.35 | 0.95 | 2.13 | 0.23 | 0.35 |
| SCRI with prior control interval | 0.97 | 1.55 | 29.60 | 0.38 | 0.99 | 1.54 | 0.31 | 0.38 |
| SCRI with posterior control interval | 0.95 | 1.50 | 45.39 | 0.42 | 0.97 | 1.48 | 0.23 | 0.42 |
| Unadjusted SCCS excluding all pre−vaccination time | 0.91 | 2.17 | 0.30 | 0.32 | 0.97 | 1.84 | 0.30 | 0.32 |

Figure 108: The coverage, mean precision, mean squared error (MSE), and fraction non-estimable based on the negative control effect-size estimate at the end of the study period in the MDCR database for Zoster vaccination (Shingrix).

| Case−control | Uncalibrated Coverage | Uncalibrated Mean precision | Uncalibrated MSE | Uncalibrated Non estimable | Calibrated Coverage | Calibrated Mean precision | Calibrated MSE | Calibrated Non estimable |
|---|---|---|---|---|---|---|---|---|
| Age & sex matched controls | 0.97 | 1.41 | 15.00 | 0.32 | 0.98 | 1.38 | 0.31 | 0.32 |
| Age & sex adjusted, using random controls | 0.97 | 1.59 | 0.26 | 0.28 | 0.98 | 1.50 | 0.28 | 0.28 |
| Cohort method | | | | | | | | |
| Unadjusted, using outpatient visits as comparator | 0.62 | 3.56 | 1.35 | 0.18 | 0.94 | 1.36 | 0.56 | 0.18 |
| PS matching, using outpatient visits as comparator | 0.87 | 2.59 | 0.59 | 0.24 | 0.99 | 2.56 | 0.31 | 0.24 |
| Unadjusted, using random days as comparator | 0.74 | 2.10 | 0.76 | 0.28 | 0.97 | 0.81 | 0.59 | 0.28 |
| PS matching, using random days as comparator | 0.97 | 2.63 | 0.36 | 0.25 | 0.98 | 2.58 | 0.23 | 0.25 |
| PS stratification, using outpatient visits as comparator | 0.76 | 3.82 | 0.62 | 0.18 | 0.96 | 3.14 | 0.33 | 0.18 |
| PS stratification, using random days as comparator | 0.94 | 3.57 | 0.35 | 0.17 | 0.97 | 3.32 | 0.24 | 0.17 |
| PS weighting, using outpatient visits as comparator | 0.98 | 1.02 | 0.44 | 0.18 | 1.00 | 1.02 | 0.24 | 0.18 |
| PS weighting, using random days as comparator | 1.00 | 1.14 | 0.39 | 0.17 | 1.00 | 1.14 | 0.31 | 0.17 |
| Per−month PS matching, using outpatient visits as comparator | | | | | | | | |
| Per−month PS matching, using random days as comparator | | | | | | | | |
| Historical comparator | | | | | | | | |
| Unadjusted, using entire historic period | 0.68 | 5.06 | 0.58 | 0.18 | 0.96 | 1.32 | 0.44 | 0.18 |
| Age & sex adjusted, using entire historic period | 0.88 | 5.04 | 0.24 | 0.18 | 0.98 | 2.26 | 0.26 | 0.18 |
| Unadjusted, using TaR after historic visit | 0.76 | 4.98 | 0.41 | 0.18 | 0.95 | 1.41 | 0.42 | 0.18 |
| Age & sex adjusted, using TaR after historic visit | 0.76 | 4.86 | 0.41 | 0.18 | 0.95 | 1.90 | 0.35 | 0.18 |
| Unadjusted, using entire historic period, filtered | 0.70 | 4.52 | 0.50 | 0.20 | 0.96 | 1.54 | 0.39 | 0.20 |
| Age & sex adjusted, using entire historic period, filtered | 0.90 | 4.51 | 0.21 | 0.20 | 0.96 | 4.10 | 0.22 | 0.20 |
| Unadjusted, using TaR after historic visit, filtered | 0.78 | 4.46 | 0.36 | 0.20 | 0.95 | 1.58 | 0.37 | 0.20 |
| Age & sex adjusted, using TaR after historic visit, filtered | 0.78 | 4.36 | 0.39 | 0.20 | 0.92 | 2.49 | 0.32 | 0.20 |
| SCCS / SCRI | | | | | | | | |
| Unadjusted SCCS excluding pre−vaccination window | 0.96 | 4.42 | 0.18 | 0.18 | 0.97 | 3.74 | 0.18 | 0.18 |
| Age & season adjusted SCCS excluding pre−vaccination window | 0.96 | 4.11 | 0.17 | 0.20 | 0.97 | 3.30 | 0.17 | 0.20 |
| SCRI with prior control interval | 0.94 | 2.74 | 18.91 | 0.23 | 0.97 | 2.33 | 0.26 | 0.23 |
| SCRI with posterior control interval | 0.96 | 2.40 | 59.36 | 0.30 | 0.98 | 2.34 | 0.29 | 0.30 |
| Unadjusted SCCS excluding all pre−vaccination time | 0.96 | 3.66 | 0.37 | 0.18 | 0.97 | 3.64 | 0.37 | 0.18 |

Figure 109: The coverage, mean precision, mean squared error (MSE), and fraction non-estimable based on the negative control effect-size estimate at the end of the study period in the CCAE database for Zoster vaccination (Shingrix).

#### 13.6 Confidence-interval-based metrics for HPV vaccination (Gardasil 9)

| | Uncalibrated | | | | Calibrated | | | |
|---|---|---|---|---|---|---|---|---|
| Case−control | Coverage | Mean precision | MSE | Non estimable | Coverage | Mean precision | MSE | Non estimable |
| Age & sex matched controls | 0.90 | 0.74 | 1.04 | 0.52 | 0.99 | 0.40 | 0.62 | 0.52 |
| Age & sex adjusted, using random controls | 0.96 | 0.63 | 0.47 | 0.56 | 0.98 | 0.50 | 0.33 | 0.56 |
| Cohort method | | | | | | | | |
| Unadjusted, using outpatient visits as comparator | 0.53 | 2.03 | 4.33 | 0.39 | 0.97 | 0.32 | 1.13 | 0.39 |
| PS matching, using outpatient visits as comparator | 0.95 | 1.09 | 0.81 | 0.52 | 0.98 | 1.07 | 0.15 | 0.52 |
| Unadjusted, using random days as comparator | 0.74 | 1.68 | 1.92 | 0.40 | 0.96 | 0.41 | 0.73 | 0.40 |
| PS matching, using random days as comparator | | | | | | | | |
| PS stratification, using outpatient visits as comparator | 0.87 | 1.65 | 0.76 | 0.39 | 0.98 | 1.29 | 0.23 | 0.39 |
| PS stratification, using random days as comparator | | | | | | | | |
| PS weighting, using outpatient visits as comparator | 1.00 | 0.54 | 0.92 | 0.39 | 0.99 | 0.53 | 0.47 | 0.39 |
| PS weighting, using random days as comparator | | | | | | | | |
| Per−month PS matching, using outpatient visits as comparator | | | | | | | | |
| Per−month PS matching, using random days as comparator | | | | | | | | |
| Historical comparator | | | | | | | | |
| Unadjusted, using entire historic period | 0.70 | 2.48 | 0.89 | 0.38 | 0.97 | 0.47 | 0.82 | 0.38 |
| Age & sex adjusted, using entire historic period | 0.83 | 2.47 | 0.45 | 0.38 | 0.98 | 1.07 | 0.23 | 0.38 |
| Unadjusted, using TaR after historic visit | 0.56 | 2.47 | 1.66 | 0.38 | 0.97 | 0.40 | 0.97 | 0.38 |
| Age & sex adjusted, using TaR after historic visit | 0.87 | 2.35 | 0.29 | 0.38 | 0.95 | 1.13 | 0.28 | 0.38 |
| Unadjusted, using entire historic period, filtered | 0.71 | 2.35 | 0.89 | 0.39 | 0.97 | 0.45 | 0.81 | 0.39 |
| Age & sex adjusted, using entire historic period, filtered | 0.84 | 2.33 | 0.41 | 0.39 | 0.97 | 1.23 | 0.21 | 0.39 |
| Unadjusted, using TaR after historic visit, filtered | 0.57 | 2.33 | 1.65 | 0.39 | 0.97 | 0.38 | 0.96 | 0.39 |
| Age & sex adjusted, using TaR after historic visit, filtered | 0.88 | 2.23 | 0.28 | 0.39 | 0.96 | 1.27 | 0.27 | 0.39 |
| SCCS / SCRI | | | | | | | | |
| Unadjusted SCCS excluding pre−vaccination window | 0.90 | 2.17 | 0.40 | 0.38 | 0.97 | 1.16 | 0.36 | 0.38 |
| Age & season adjusted SCCS excluding pre−vaccination window | 0.90 | 2.17 | 0.41 | 0.38 | 0.97 | 1.12 | 0.37 | 0.38 |
| SCRI with prior control interval | 0.96 | 1.07 | 74.44 | 0.56 | 0.98 | 0.76 | 0.15 | 0.56 |
| SCRI with posterior control interval | 0.96 | 1.10 | 30.11 | 0.47 | 0.99 | 0.92 | 0.22 | 0.47 |
| Unadjusted SCCS excluding all pre−vaccination time | 0.91 | 1.77 | 0.43 | 0.39 | 0.98 | 1.40 | 0.34 | 0.39 |

Figure 110: The coverage, mean precision, mean squared error (MSE), and fraction non-estimable based on the negative control effect-size estimate at the end of the study period in the Optum EHR database for HPV vaccination (Gardasil 9).

| | Uncalibrated | | | | Calibrated | | | |
|---|---|---|---|---|---|---|---|---|
| Case−control | Coverage | Mean precision | MSE | Non estimable | Coverage | Mean precision | MSE | Non estimable |
| Age & sex matched controls | 0.90 | 1.07 | 0.34 | 0.61 | 0.98 | 0.78 | 0.20 | 0.61 |
| Age & sex adjusted, using random controls | 0.94 | 1.22 | 0.23 | 0.54 | 0.98 | 1.00 | 0.14 | 0.54 |
| Cohort method | | | | | | | | |
| Unadjusted, using outpatient visits as comparator | 0.59 | 1.55 | 4.09 | 0.47 | 1.00 | 0.23 | 1.14 | 0.47 |
| PS matching, using outpatient visits as comparator | 0.94 | 0.82 | 0.88 | 0.61 | 0.98 | 0.61 | 0.21 | 0.61 |
| Unadjusted, using random days as comparator | 0.78 | 1.30 | 2.30 | 0.49 | 0.96 | 0.31 | 0.79 | 0.49 |
| PS matching, using random days as comparator | 0.94 | 0.77 | 0.59 | 0.61 | 0.99 | 0.50 | 0.19 | 0.61 |
| PS stratification, using outpatient visits as comparator | 0.88 | 1.25 | 0.73 | 0.49 | 0.98 | 0.79 | 0.23 | 0.49 |
| PS stratification, using random days as comparator | 0.92 | 1.12 | 1.03 | 0.47 | 0.98 | 0.73 | 0.67 | 0.47 |
| PS weighting, using outpatient visits as comparator | 0.97 | 0.47 | 1.06 | 0.49 | 0.98 | 0.46 | 0.57 | 0.49 |
| PS weighting, using random days as comparator | 0.99 | 0.44 | 1.83 | 0.48 | 0.99 | 0.35 | 1.40 | 0.49 |
| Per−month PS matching, using outpatient visits as comparator | | | | | | | | |
| Per−month PS matching, using random days as comparator | | | | | | | | |
| Historical comparator | | | | | | | | |
| Unadjusted, using entire historic period | 0.75 | 1.92 | 1.07 | 0.46 | 0.95 | 0.36 | 0.81 | 0.46 |
| Age & sex adjusted, using entire historic period | 0.89 | 1.91 | 0.30 | 0.46 | 0.98 | 0.72 | 0.26 | 0.46 |
| Unadjusted, using TaR after historic visit | 0.71 | 1.89 | 1.55 | 0.46 | 0.97 | 0.32 | 0.91 | 0.46 |
| Age & sex adjusted, using TaR after historic visit | 0.88 | 1.78 | 0.35 | 0.46 | 0.97 | 0.66 | 0.35 | 0.46 |
| Unadjusted, using entire historic period, filtered | 0.76 | 1.77 | 1.06 | 0.48 | 0.95 | 0.35 | 0.76 | 0.48 |
| Age & sex adjusted, using entire historic period, filtered | 0.90 | 1.76 | 0.22 | 0.48 | 0.97 | 0.90 | 0.20 | 0.48 |
| Unadjusted, using TaR after historic visit, filtered | 0.72 | 1.75 | 1.55 | 0.48 | 0.96 | 0.30 | 0.87 | 0.48 |
| Age & sex adjusted, using TaR after historic visit, filtered | 0.90 | 1.66 | 0.26 | 0.48 | 0.95 | 0.83 | 0.26 | 0.48 |
| SCCS / SCRI | | | | | | | | |
| Unadjusted SCCS excluding pre−vaccination window | 0.94 | 1.71 | 0.27 | 0.47 | 0.97 | 1.15 | 0.23 | 0.47 |
| Age & season adjusted SCCS excluding pre−vaccination window | 0.94 | 1.60 | 0.28 | 0.48 | 0.97 | 1.07 | 0.26 | 0.48 |
| SCRI with prior control interval | 0.96 | 1.03 | 0.21 | 0.55 | 0.99 | 0.59 | 0.19 | 0.55 |
| SCRI with posterior control interval | 0.95 | 1.03 | 0.17 | 0.57 | 0.98 | 0.62 | 0.18 | 0.57 |
| Unadjusted SCCS excluding all pre−vaccination time | 0.92 | 1.48 | 0.22 | 0.49 | 0.97 | 0.89 | 0.18 | 0.49 |

Figure 111: The coverage, mean precision, mean squared error (MSE), and fraction non-estimable based on the negative control effect-size estimate at the end of the study period in the MDCD database for HPV vaccination (Gardasil 9).

| | Uncalibrated | | | | Calibrated | | | |
|---|---|---|---|---|---|---|---|---|
| Case−control | Coverage | Mean precision | MSE | Non estimable | Coverage | Mean precision | MSE | Non estimable |
| Age & sex matched controls | | | | | | | | |
| Age & sex adjusted, using random controls | | | | | | | | |
| Cohort method | | | | | | | | |
| Unadjusted, using outpatient visits as comparator | 1.00 | 0.00 | 0.04 | 1.00 | 1.00 | 0.00 | 0.00 | 1.00 |
| PS matching, using outpatient visits as comparator | | | | | | | | |
| Unadjusted, using random days as comparator | | | | | | | | |
| PS matching, using random days as comparator | | | | | | | | |
| PS stratification, using outpatient visits as comparator | 1.00 | 0.00 | 0.03 | 1.00 | 1.00 | 0.00 | 0.00 | 1.00 |
| PS stratification, using random days as comparator | | | | | | | | |
| PS weighting, using outpatient visits as comparator | 1.00 | 0.00 | 0.02 | 1.00 | 1.00 | 0.00 | 0.00 | 1.00 |
| PS weighting, using random days as comparator | | | | | | | | |
| Per−month PS matching, using outpatient visits as comparator | | | | | | | | |
| Per−month PS matching, using random days as comparator | | | | | | | | |
| Historical comparator | | | | | | | | |
| Unadjusted, using entire historic period | 1.00 | 0.00 | 0.00 | 1.00 | 1.00 | 0.00 | 0.00 | 1.00 |
| Age & sex adjusted, using entire historic period | 1.00 | 0.00 | 0.00 | 1.00 | 1.00 | 0.00 | 0.00 | 1.00 |
| Unadjusted, using TaR after historic visit | 1.00 | 0.00 | 0.00 | 1.00 | 1.00 | 0.00 | 0.00 | 1.00 |
| Age & sex adjusted, using TaR after historic visit | 1.00 | 0.00 | 0.00 | 1.00 | 1.00 | 0.00 | 0.00 | 1.00 |
| Unadjusted, using entire historic period, filtered | 1.00 | 0.00 | 0.00 | 1.00 | 1.00 | 0.00 | 0.00 | 1.00 |
| Age & sex adjusted, using entire historic period, filtered | 1.00 | 0.00 | 0.00 | 1.00 | 1.00 | 0.00 | 0.00 | 1.00 |
| Unadjusted, using TaR after historic visit, filtered | 1.00 | 0.00 | 0.00 | 1.00 | 1.00 | 0.00 | 0.00 | 1.00 |
| Age & sex adjusted, using TaR after historic visit, filtered | 1.00 | 0.00 | 0.00 | 1.00 | 1.00 | 0.00 | 0.00 | 1.00 |

Figure 112: The coverage, mean precision, mean squared error (MSE), and fraction non-estimable based on the negative control effect-size estimate at the end of the study period in the MDCR database for HPV vaccination (Gardasil 9).

| | Uncalibrated | | | | Calibrated | | | |
|---|---|---|---|---|---|---|---|---|
| | Coverage | Mean precision | MSE | Non estimable | Coverage | Mean precision | MSE | Non estimable |
| **Case−control** | | | | | | | | |
| Age & sex matched controls | 0.92 | 1.16 | 0.50 | 0.51 | 0.99 | 1.01 | 0.35 | 0.51 |
| Age & sex adjusted, using random controls | 0.95 | 0.99 | 0.37 | 0.53 | 0.96 | 0.92 | 0.29 | 0.53 |
| **Cohort method** | | | | | | | | |
| Unadjusted, using outpatient visits as comparator | 0.54 | 2.27 | 4.01 | 0.39 | 0.98 | 0.32 | 1.02 | 0.39 |
| PS matching, using outpatient visits as comparator | 0.96 | 1.26 | 0.80 | 0.53 | 0.98 | 1.12 | 0.14 | 0.53 |
| Unadjusted, using random days as comparator | 0.75 | 1.93 | 1.88 | 0.40 | 0.97 | 0.45 | 0.57 | 0.40 |
| PS matching, using random days as comparator | 0.95 | 1.16 | 0.65 | 0.55 | 0.99 | 0.87 | 0.11 | 0.55 |
| PS stratification, using outpatient visits as comparator | 0.89 | 1.87 | 0.63 | 0.42 | 0.96 | 1.37 | 0.20 | 0.42 |
| PS stratification, using random days as comparator | 0.92 | 1.59 | 0.74 | 0.41 | 0.97 | 1.14 | 0.35 | 0.41 |
| PS weighting, using outpatient visits as comparator | 1.00 | 0.62 | 0.91 | 0.42 | 1.00 | 0.62 | 0.54 | 0.42 |
| PS weighting, using random days as comparator | 0.98 | 0.50 | 1.00 | 0.41 | 0.99 | 0.48 | 0.71 | 0.41 |
| Per−month PS matching, using outpatient visits as comparator | | | | | | | | |
| Per−month PS matching, using random days as comparator | | | | | | | | |
| **Historical comparator** | | | | | | | | |
| Unadjusted, using entire historic period | 0.69 | 2.77 | 0.86 | 0.38 | 0.97 | 0.50 | 0.60 | 0.38 |
| Age & sex adjusted, using entire historic period | 0.91 | 2.75 | 0.24 | 0.38 | 0.99 | 1.18 | 0.20 | 0.38 |
| Unadjusted, using TaR after historic visit | 0.58 | 2.73 | 1.38 | 0.38 | 0.98 | 0.39 | 0.82 | 0.38 |
| Age & sex adjusted, using TaR after historic visit | 0.89 | 2.54 | 0.27 | 0.38 | 0.99 | 1.08 | 0.27 | 0.38 |
| Unadjusted, using entire historic period, filtered | 0.69 | 2.53 | 0.86 | 0.40 | 0.97 | 0.47 | 0.59 | 0.40 |
| Age & sex adjusted, using entire historic period, filtered | 0.92 | 2.51 | 0.20 | 0.40 | 0.98 | 1.45 | 0.17 | 0.40 |
| Unadjusted, using TaR after historic visit, filtered | 0.59 | 2.50 | 1.38 | 0.40 | 0.98 | 0.36 | 0.81 | 0.40 |
| Age & sex adjusted, using TaR after historic visit, filtered | 0.90 | 2.33 | 0.25 | 0.40 | 0.99 | 1.19 | 0.25 | 0.40 |
| **SCCS / SCRI** | | | | | | | | |
| Unadjusted SCCS excluding pre−vaccination window | 0.90 | 2.44 | 0.30 | 0.39 | 0.97 | 1.54 | 0.25 | 0.39 |
| Age & season adjusted SCCS excluding pre−vaccination window | 0.90 | 2.43 | 0.30 | 0.39 | 0.98 | 1.55 | 0.26 | 0.39 |
| SCRI with prior control interval | 0.95 | 1.37 | 75.77 | 0.52 | 0.99 | 0.76 | 0.21 | 0.52 |
| SCRI with posterior control interval | 0.97 | 1.43 | 15.03 | 0.49 | 0.99 | 0.96 | 0.09 | 0.49 |
| Unadjusted SCCS excluding all pre−vaccination time | 0.91 | 2.11 | 0.35 | 0.41 | 0.98 | 1.24 | 0.26 | 0.41 |

Figure 113: The coverage, mean precision, mean squared error (MSE), and fraction non-estimable based on the negative control effect-size estimate at the end of the study period in the CCAE database for HPV vaccination (Gardasil 9).

### 14 Negative controls effect size estimates

Dots indicate the estimated effect size (x-axis) such as odds ratio or incidence rate ratio, and corresponding standard error (y-axis), which is linearly related to the width of the confidence interval. Estimates below the red dashed line have a one-sided p-value < 0.05, and filled dots indicate the LLR exceeds the CV.

#### 14.1 Negative controls effect size estimates in Optum EHR

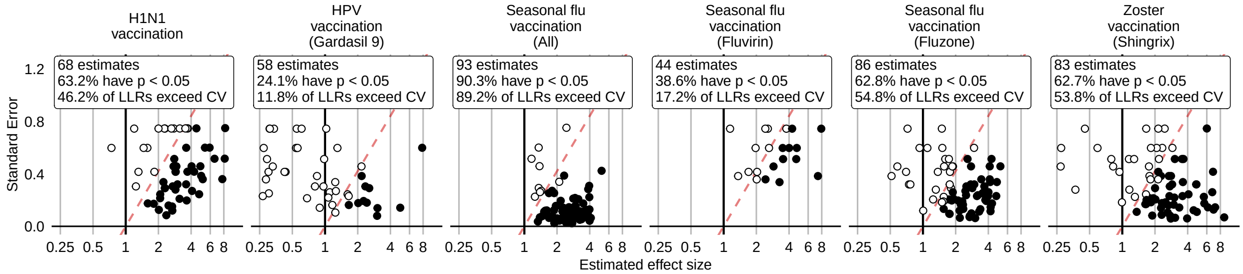

Figure 114: Negative control effect-size estimates and standard errors at the end of the study period using the HistoricalComparator method (Unadjusted, using entire historic period), in the Optum EHR database.

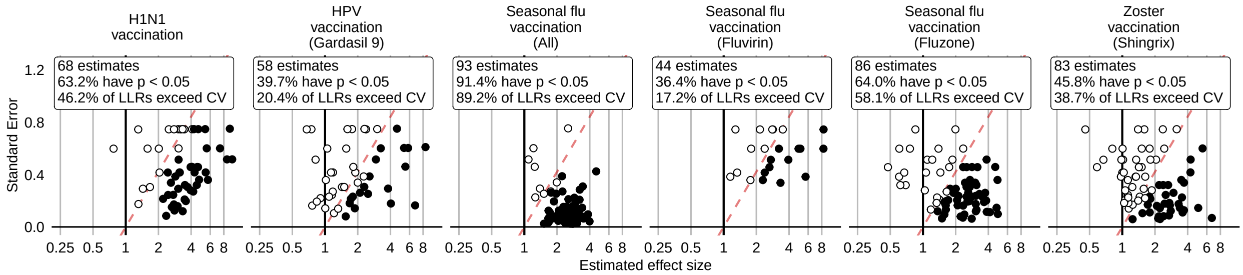

Figure 115: Negative control effect-size estimates and standard errors at the end of the study period using the HistoricalComparator method (Age & sex adjusted, using entire historic period), in the Optum EHR database.

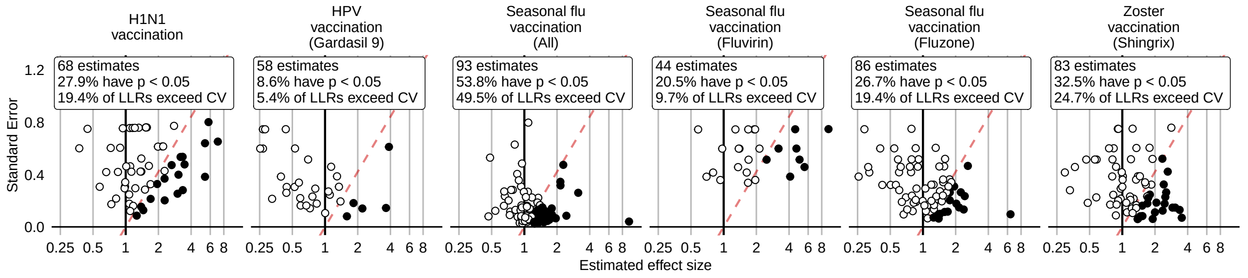

Figure 116: Negative control effect-size estimates and standard errors at the end of the study period using the HistoricalComparator method (Unadjusted, using TaR after historic visit), in the Optum EHR database.

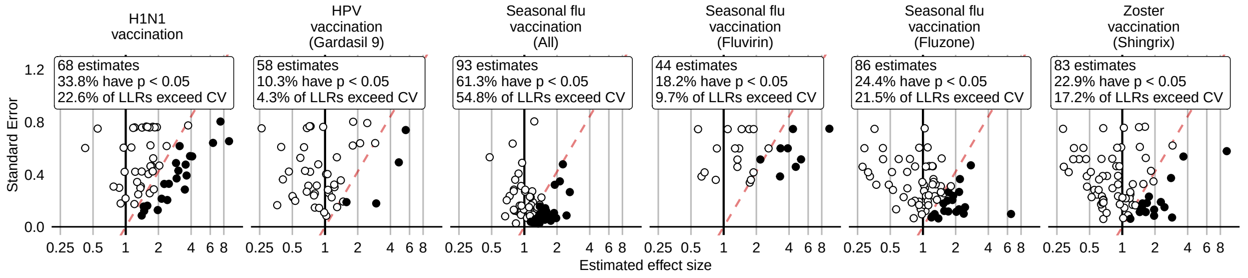

Figure 117: Negative control effect-size estimates and standard errors at the end of the study period using the HistoricalComparator method (Age & sex adjusted, using TaR after historic visit), in the Optum EHR database.

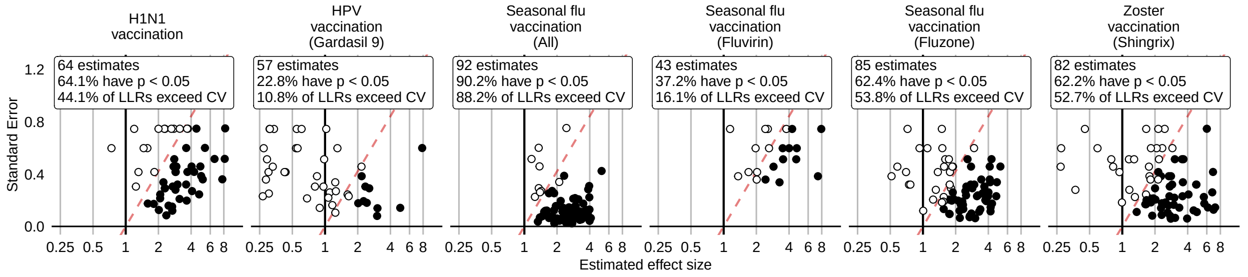

Figure 118: Negative control effect-size estimates and standard errors at the end of the study period using the HistoricalComparator method (Unadjusted, using entire historic period, filtered), in the Optum EHR database.

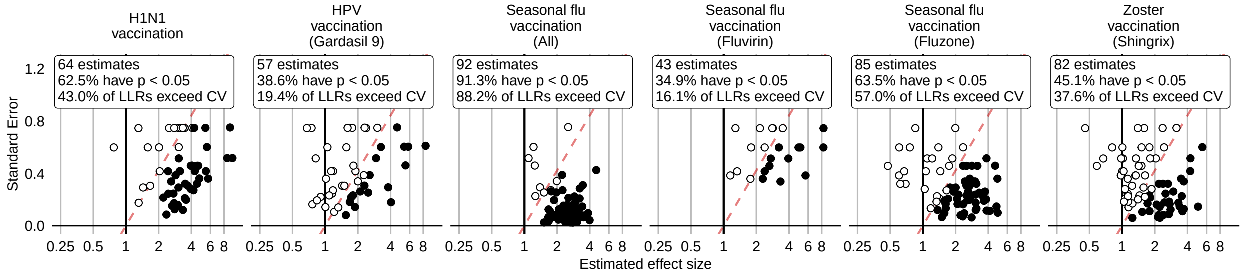

Figure 119: Negative control effect-size estimates and standard errors at the end of the study period using the HistoricalComparator method (Age & sex adjusted, using entire historic period, filtered), in the Optum EHR database.

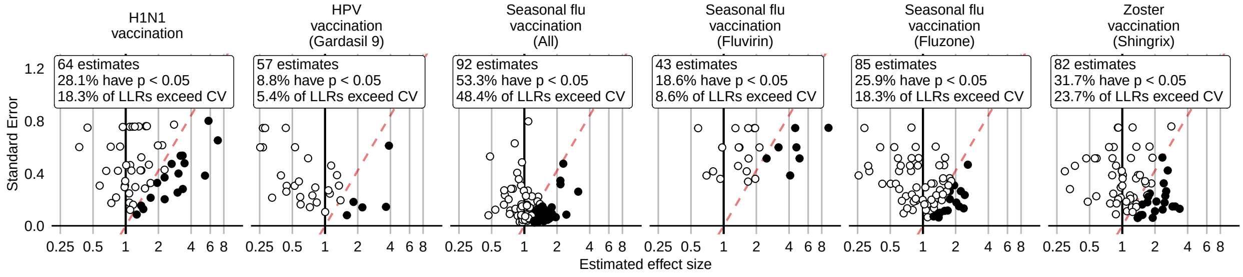

Figure 120: Negative control effect-size estimates and standard errors at the end of the study period using the HistoricalComparator method (Unadjusted, using TaR after historic visit, filtered), in the Optum EHR database.

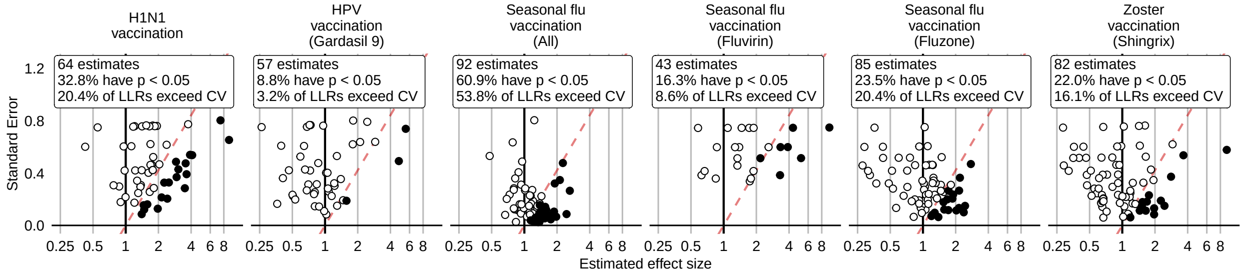

Figure 121: Negative control effect-size estimates and standard errors at the end of the study period using the HistoricalComparator method (Age & sex adjusted, using TaR after historic visit, filtered), in the Optum EHR database.

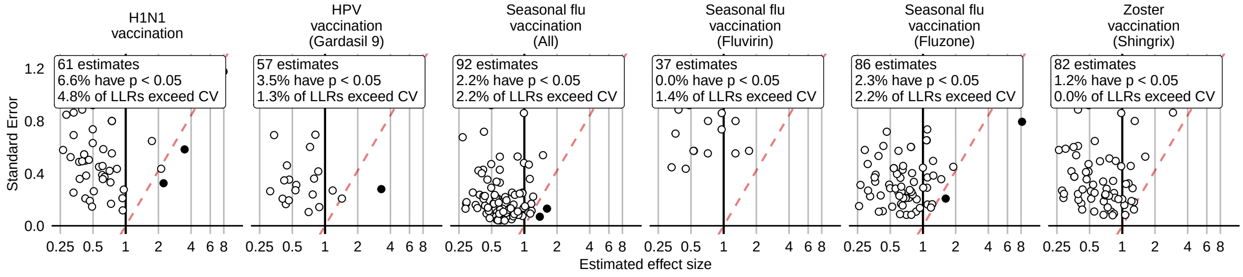

Figure 122: Negative control effect-size estimates and standard errors at the end of the study period using the CohortMethod method (Unadjusted, using outpatient visits as comparator), in the Optum EHR database.

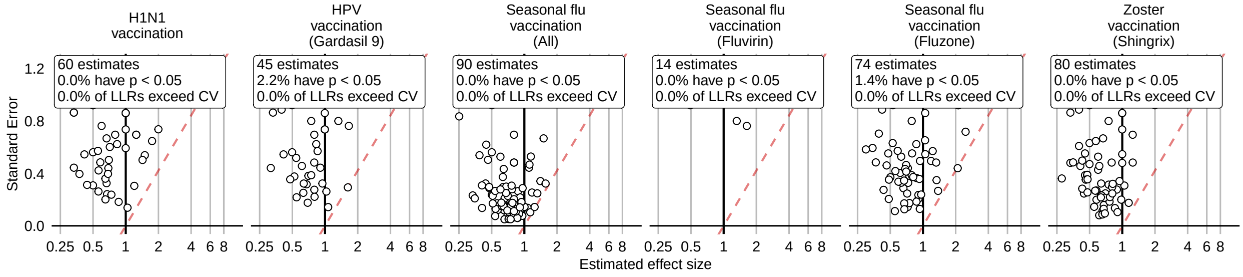

Figure 123: Negative control effect-size estimates and standard errors at the end of the study period using the CohortMethod method (PS matching, using outpatient visits as comparator), in the Optum EHR database.

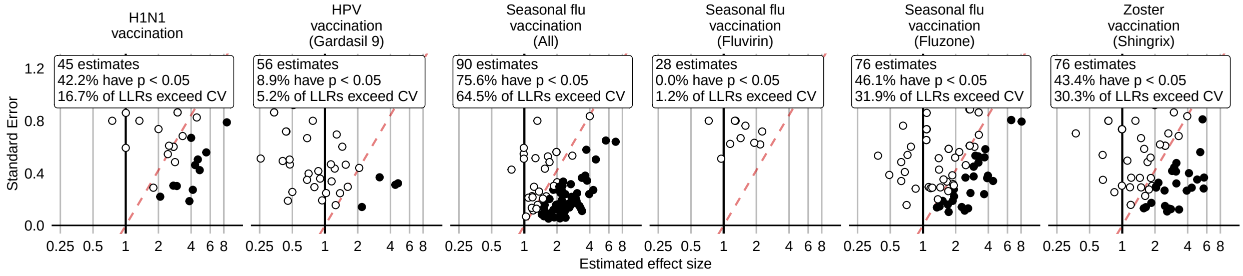

Figure 124: Negative control effect-size estimates and standard errors at the end of the study period using the CohortMethod method (Unadjusted, using random days as comparator), in the Optum EHR database.

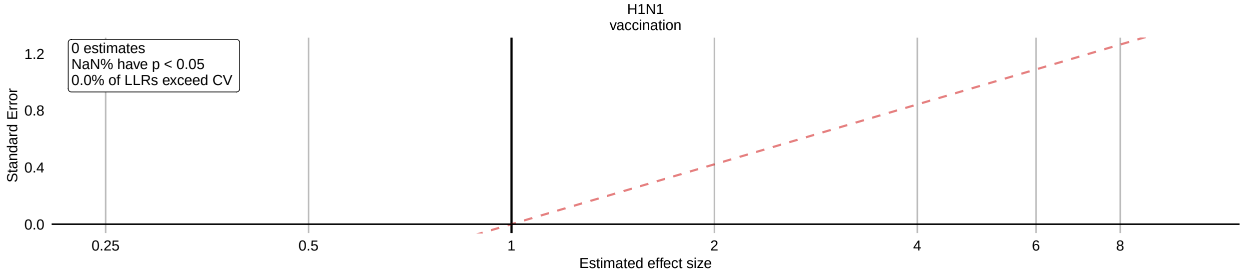

Figure 125: Negative control effect-size estimates and standard errors at the end of the study period using the CohortMethod method (PS matching, using random days as comparator), in the Optum EHR database.

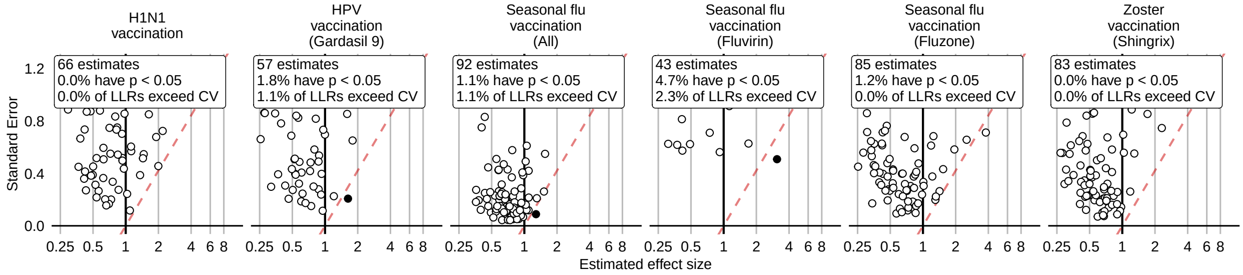

Figure 126: Negative control effect-size estimates and standard errors at the end of the study period using the CohortMethod method (PS stratification, using outpatient visits as comparator), in the Optum EHR database.

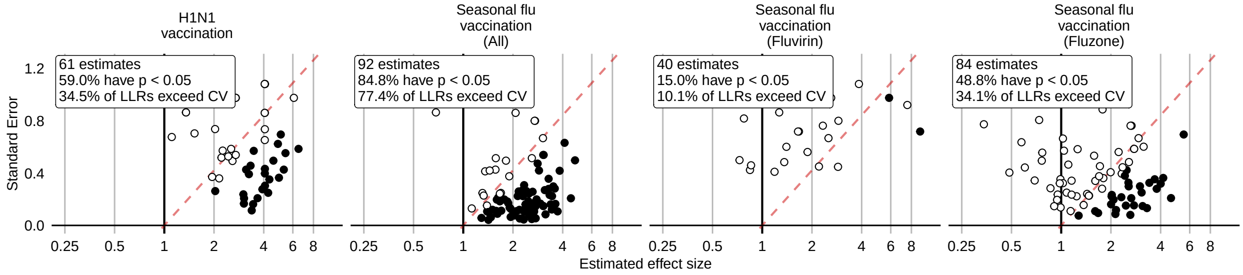

Figure 127: Negative control effect-size estimates and standard errors at the end of the study period using the CohortMethod method (PS stratification, using random days as comparator), in the Optum EHR database.

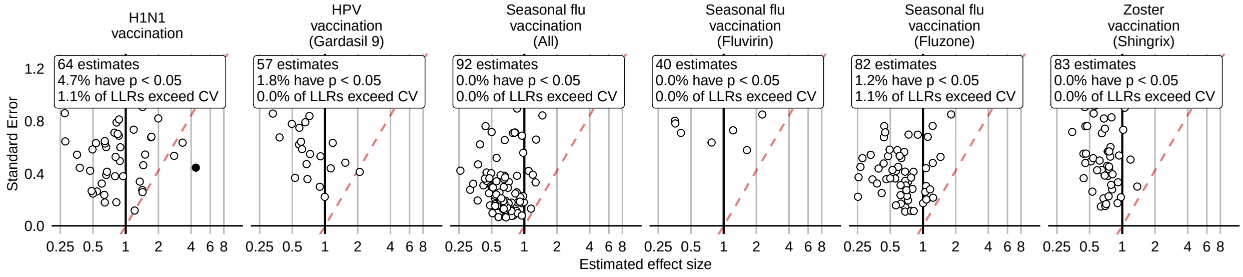

Figure 128: Negative control effect-size estimates and standard errors at the end of the study period using the CohortMethod method (PS weighting, using outpatient visits as comparator), in the Optum EHR database.

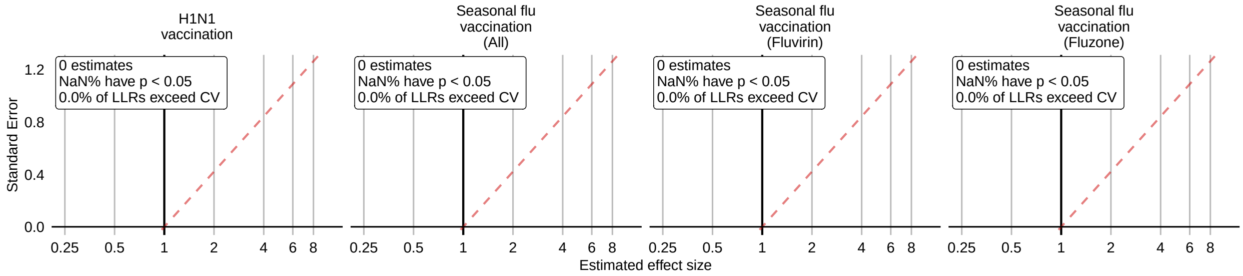

Figure 129: Negative control effect-size estimates and standard errors at the end of the study period using the CohortMethod method (PS weighting, using random days as comparator), in the Optum EHR database.

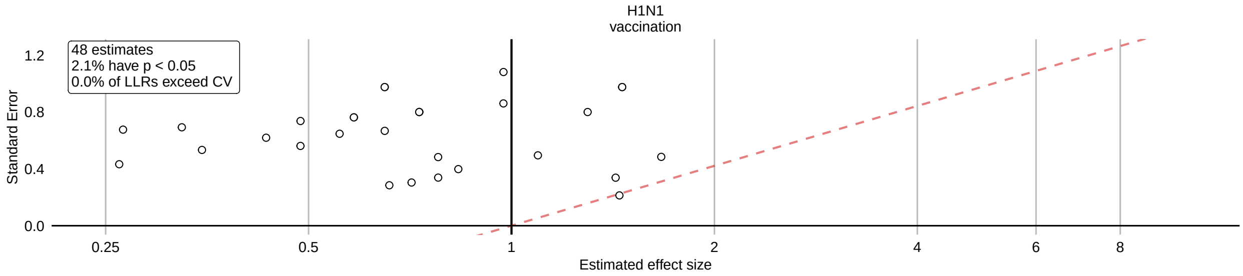

Figure 130: Negative control effect-size estimates and standard errors at the end of the study period using the CohortMethod method (Per-month PS matching, using outpatient visits as comparator), in the Optum EHR database.

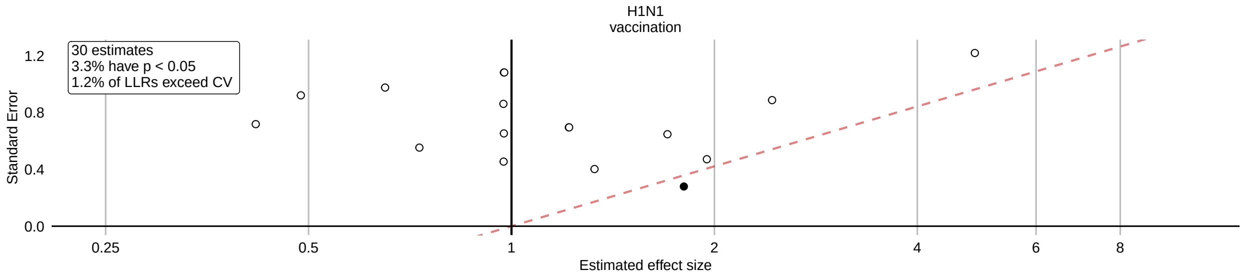

Figure 131: Negative control effect-size estimates and standard errors at the end of the study period using the CohortMethod method (Per-month PS matching, using random days as comparator), in the Optum EHR database.

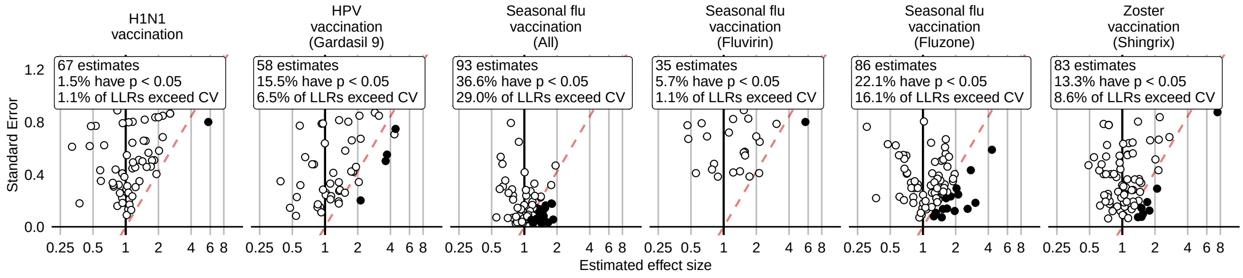

Figure 132: Negative control effect-size estimates and standard errors at the end of the study period using the SCCS method (Unadjusted SCCS excluding pre-vaccination window), in the Optum EHR database.

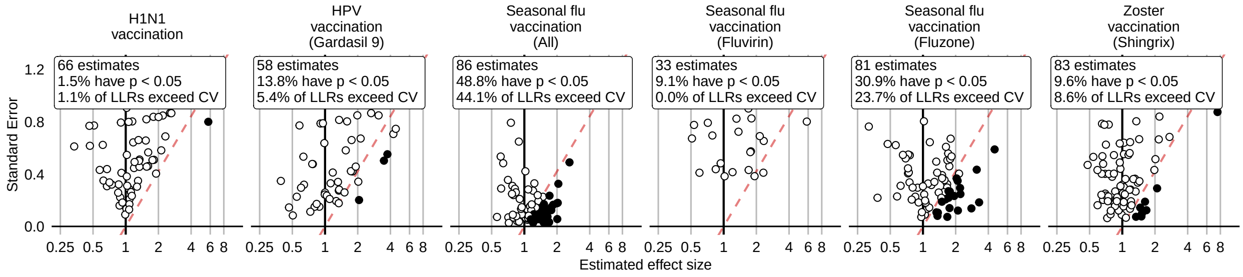

Figure 133: Negative control effect-size estimates and standard errors at the end of the study period using the SCCS method (Age & season adjusted SCCS excluding pre-vaccination window), in the Optum EHR database.

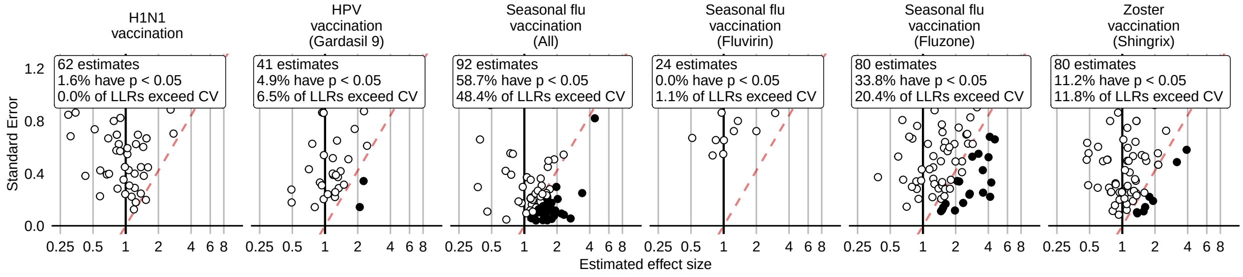

Figure 134: Negative control effect-size estimates and standard errors at the end of the study period using the SCCS method (SCRI with prior control interval), in the Optum EHR database.

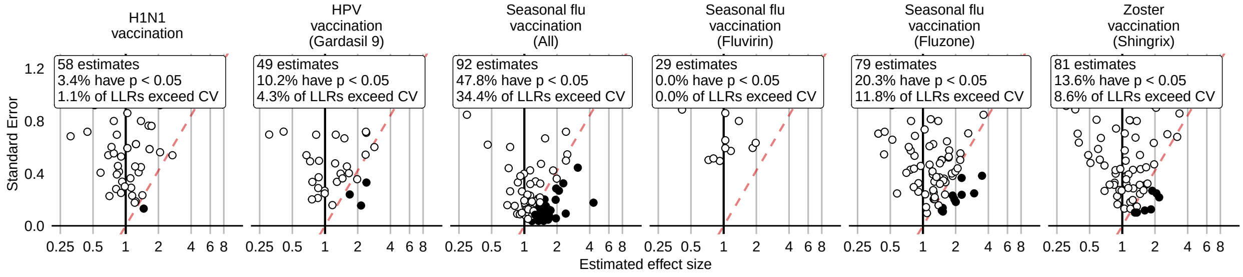

Figure 135: Negative control effect-size estimates and standard errors at the end of the study period using the SCCS method (SCRI with posterior control interval), in the Optum EHR database.

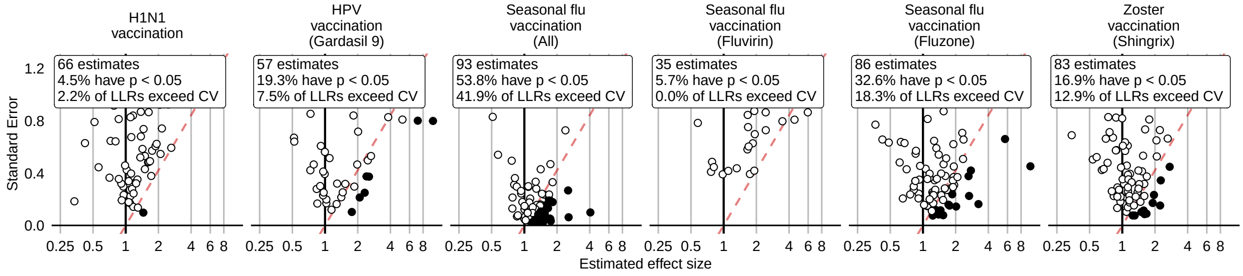

Figure 136: Negative control effect-size estimates and standard errors at the end of the study period using the SCCS method (Unadjusted SCCS excluding all pre-vaccination time), in the Optum EHR database.

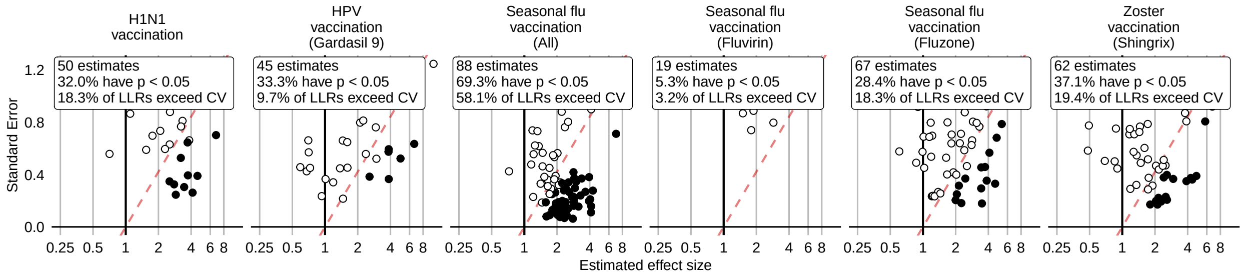

Figure 137: Negative control effect-size estimates and standard errors at the end of the study period using the CaseControl method (Age & sex adjusted, using random controls), in the Optum EHR database.

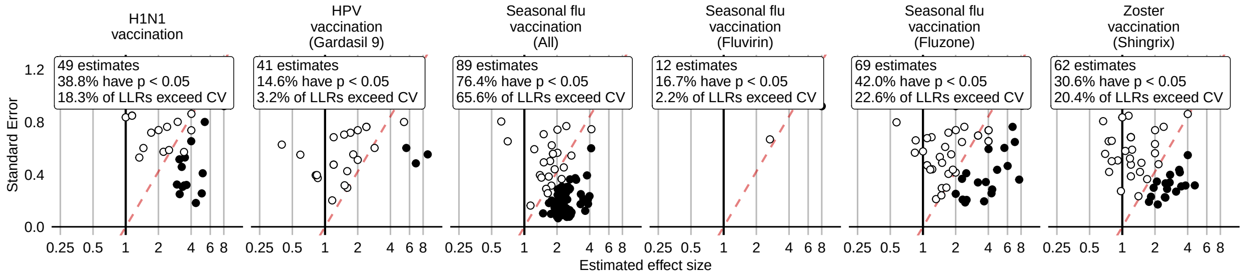

Figure 138: Negative control effect-size estimates and standard errors at the end of the study period using the CaseControl method (Age & sex matched controls), in the Optum EHR database.

#### 14.2 Negative controls effect size estimates in MDCD

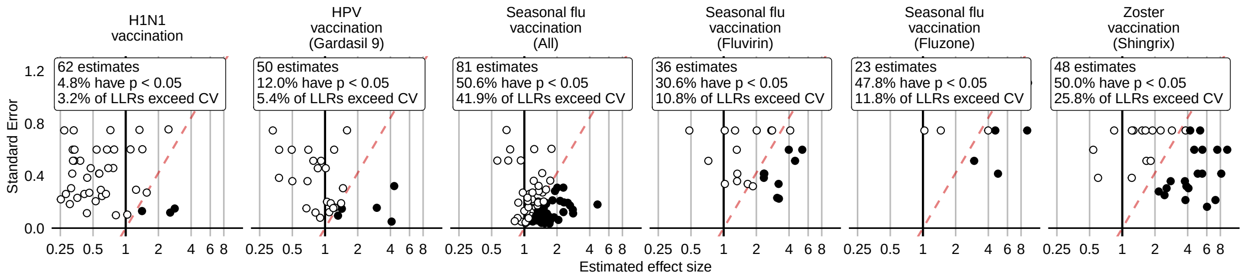

Figure 139: Negative control effect-size estimates and standard errors at the end of the study period using the HistoricalComparator method (Unadjusted, using entire historic period), in the MDCD database.

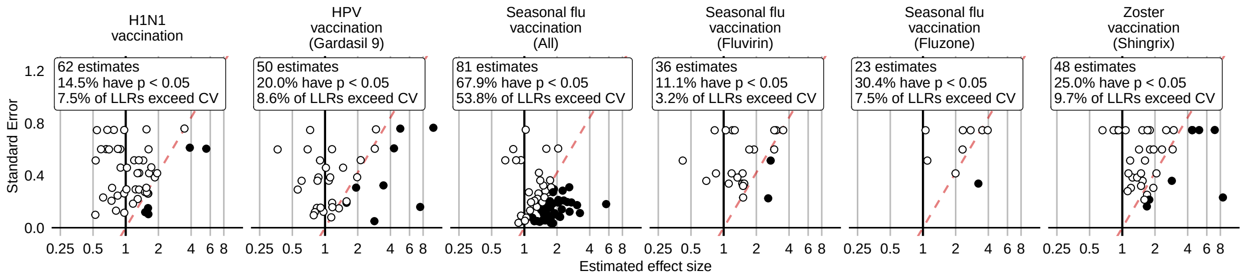

Figure 140: Negative control effect-size estimates and standard errors at the end of the study period using the HistoricalComparator method (Age & sex adjusted, using entire historic period), in the MDCD database.

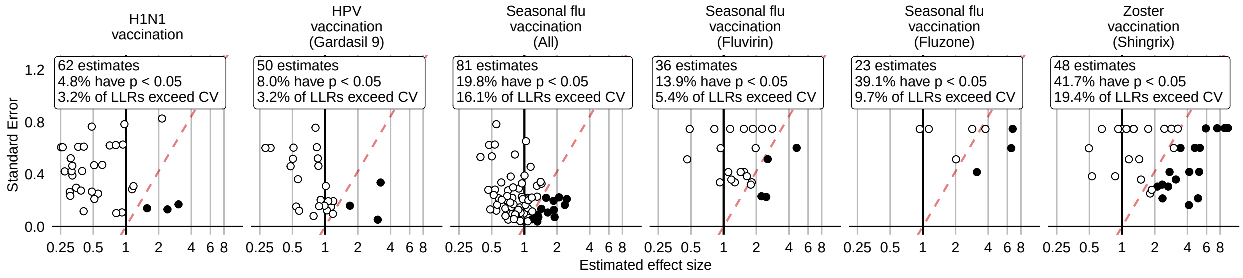

Figure 141: Negative control effect-size estimates and standard errors at the end of the study period using the HistoricalComparator method (Unadjusted, using TaR after historic visit), in the MDCD database.

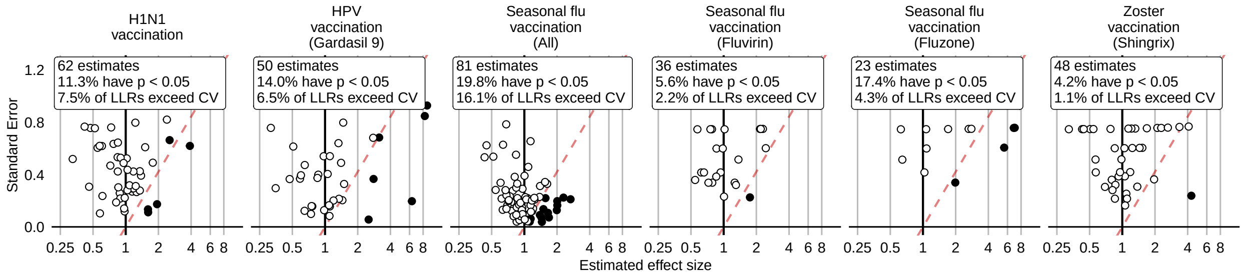

Figure 142: Negative control effect-size estimates and standard errors at the end of the study period using the HistoricalComparator method (Age & sex adjusted, using TaR after historic visit), in the MDCD database.

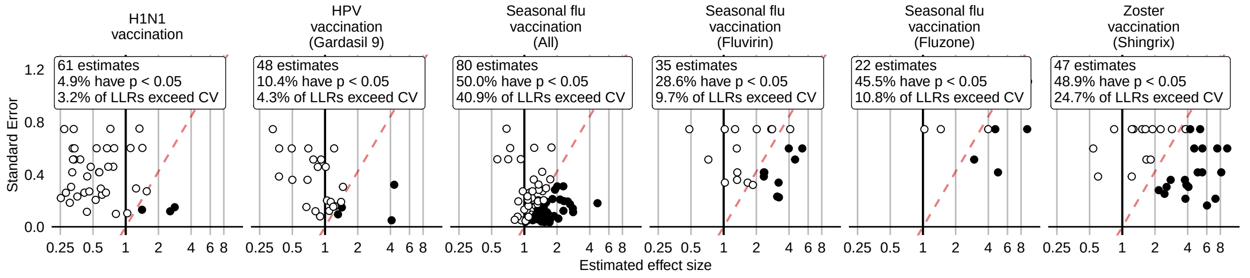

Figure 143: Negative control effect-size estimates and standard errors at the end of the study period using the HistoricalComparator method (Unadjusted, using entire historic period, filtered), in the MDCD database.

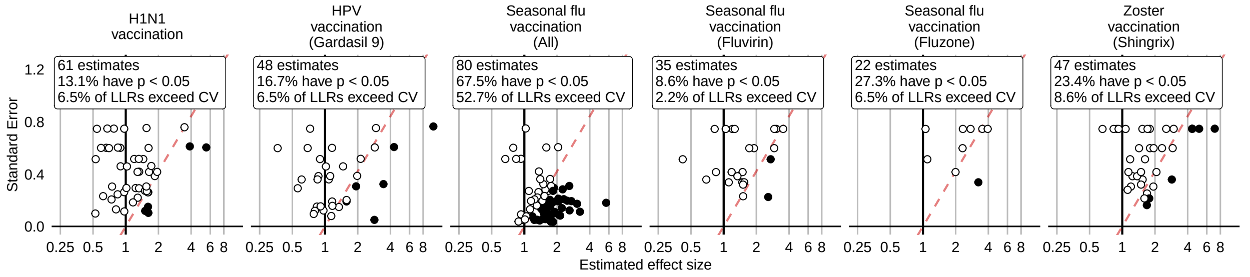

Figure 144: Negative control effect-size estimates and standard errors at the end of the study period using the HistoricalComparator method (Age & sex adjusted, using entire historic period, filtered), in the MDCD database.

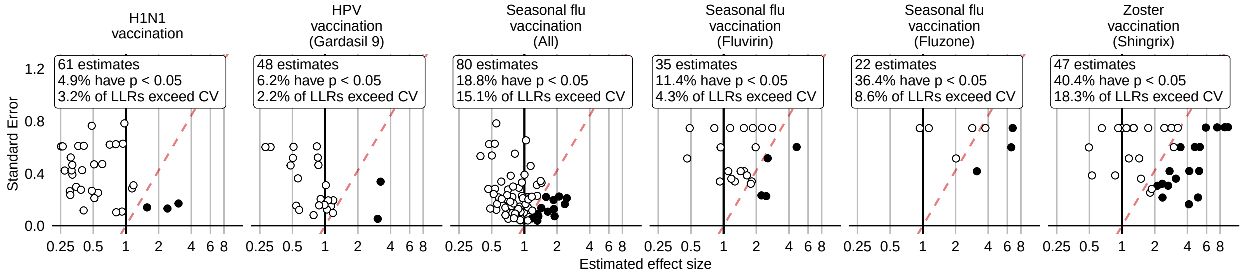

Figure 145: Negative control effect-size estimates and standard errors at the end of the study period using the HistoricalComparator method (Unadjusted, using TaR after historic visit, filtered), in the MDCD database.

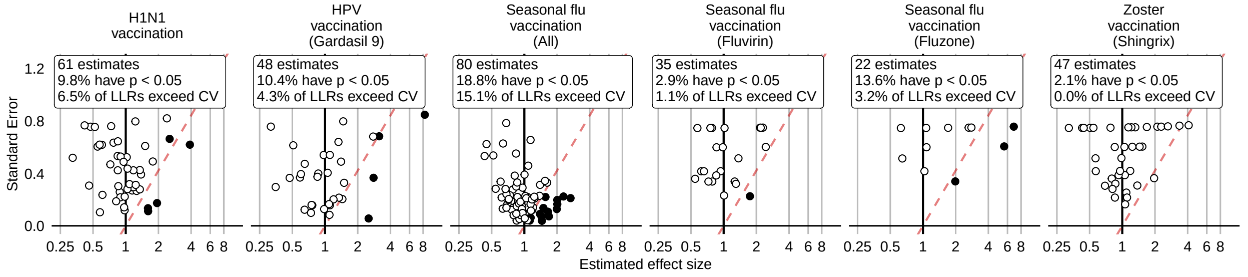

Figure 146: Negative control effect-size estimates and standard errors at the end of the study period using the HistoricalComparator method (Age & sex adjusted, using TaR after historic visit, filtered), in the MDCD database.

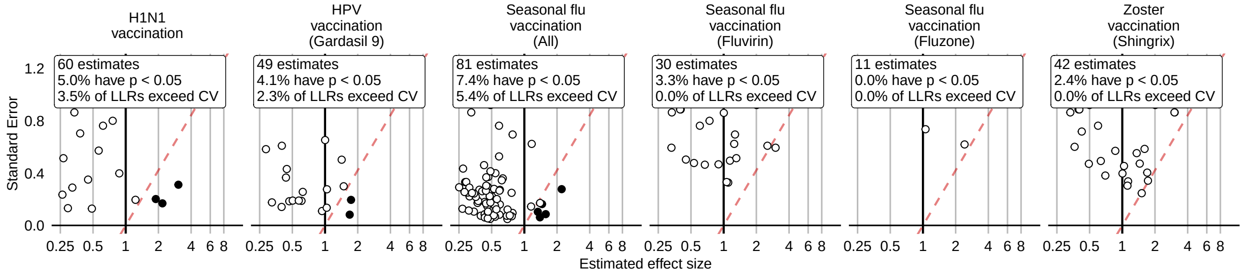

Figure 147: Negative control effect-size estimates and standard errors at the end of the study period using the CohortMethod method (Unadjusted, using outpatient visits as comparator), in the MDCD database.

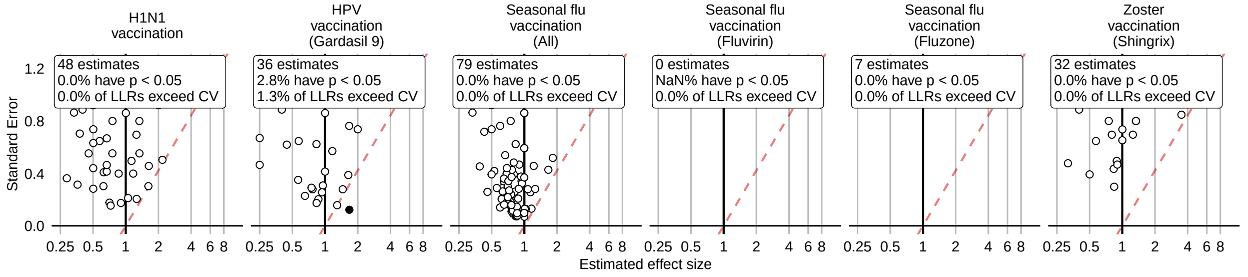

Figure 148: Negative control effect-size estimates and standard errors at the end of the study period using the CohortMethod method (PS matching, using outpatient visits as comparator), in the MDCD database.

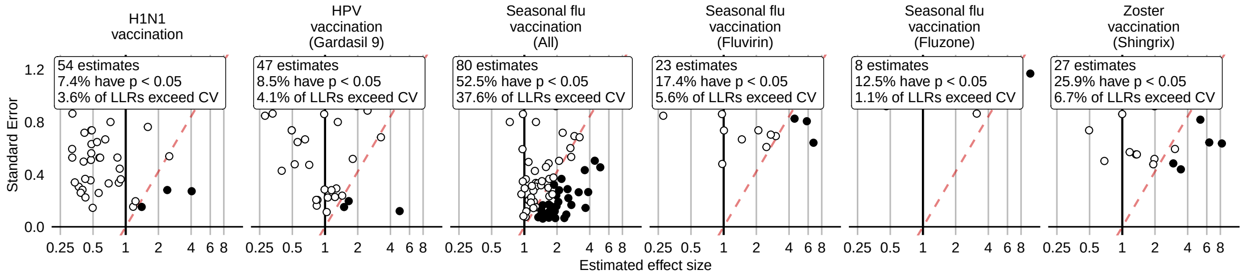

Figure 149: Negative control effect-size estimates and standard errors at the end of the study period using the CohortMethod method (Unadjusted, using random days as comparator), in the MDCD database.

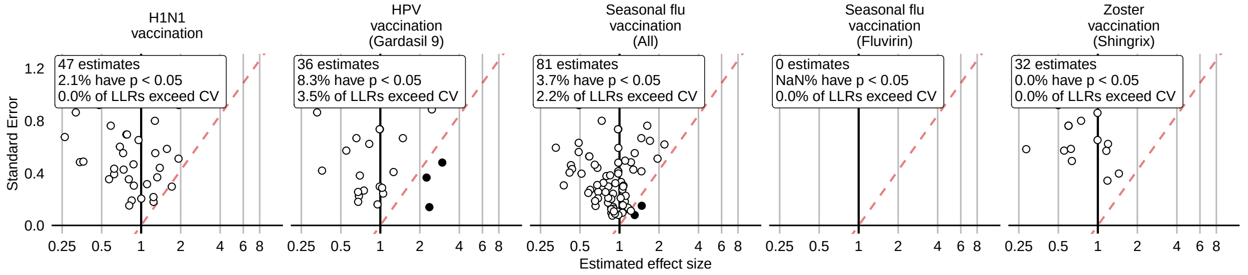

Figure 150: Negative control effect-size estimates and standard errors at the end of the study period using the CohortMethod method (PS matching, using random days as comparator), in the MDCD database.

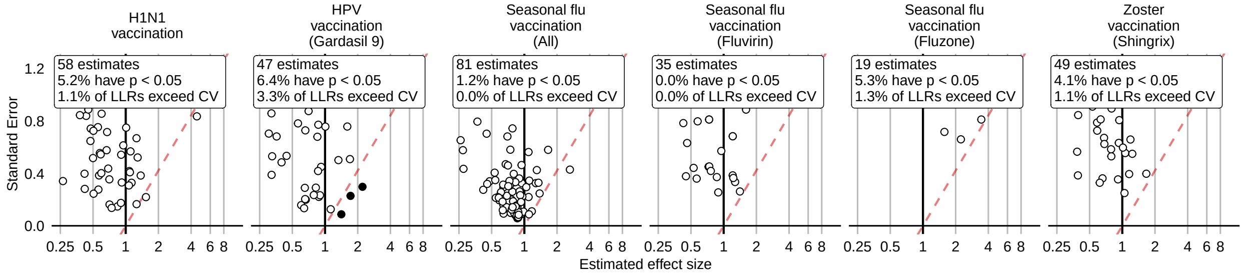

Figure 151: Negative control effect-size estimates and standard errors at the end of the study period using the CohortMethod method (PS stratification, using outpatient visits as comparator), in the MDCD database.

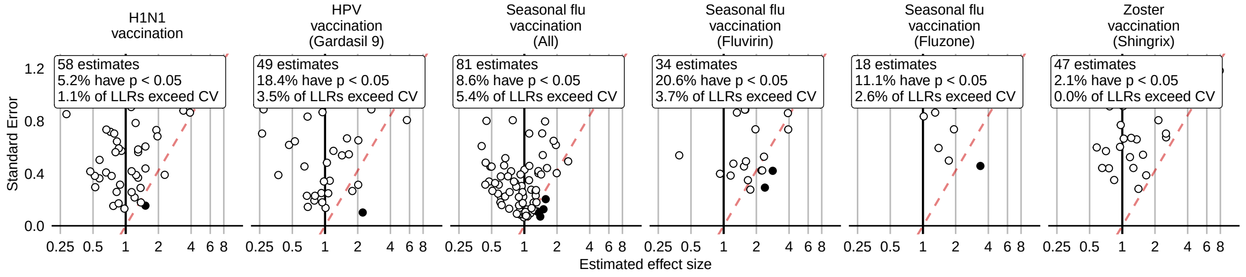

Figure 152: Negative control effect-size estimates and standard errors at the end of the study period using the CohortMethod method (PS stratification, using random days as comparator), in the MDCD database.

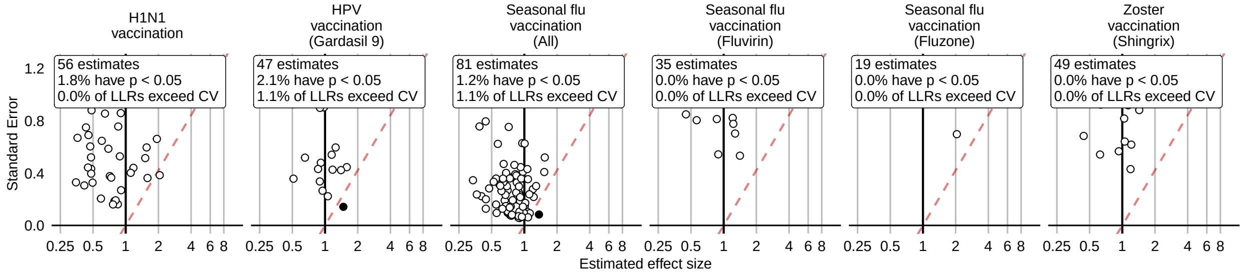

Figure 153: Negative control effect-size estimates and standard errors at the end of the study period using the CohortMethod method (PS weighting, using outpatient visits as comparator), in the MDCD database.

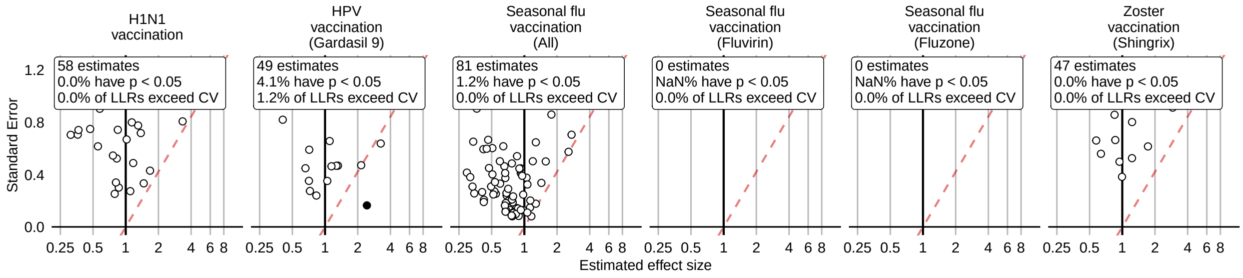

Figure 154: Negative control effect-size estimates and standard errors at the end of the study period using the CohortMethod method (PS weighting, using random days as comparator), in the MDCD database.

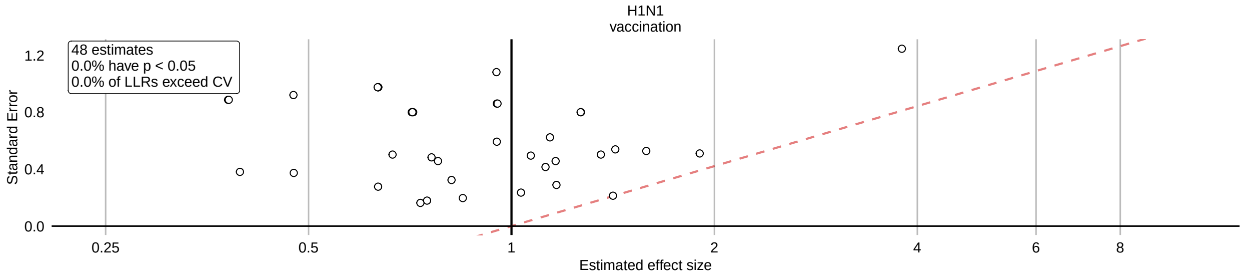

Figure 155: Negative control effect-size estimates and standard errors at the end of the study period using the CohortMethod method (Per-month PS matching, using outpatient visits as comparator), in the MDCD database.

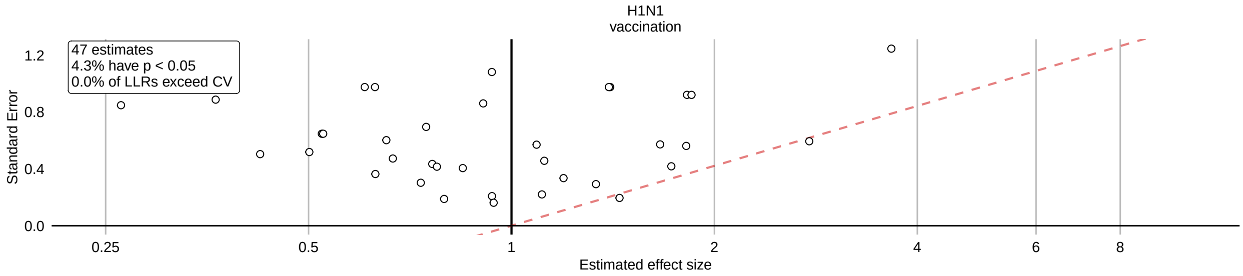

Figure 156: Negative control effect-size estimates and standard errors at the end of the study period using the CohortMethod method (Per-month PS matching, using random days as comparator), in the MDCD database.

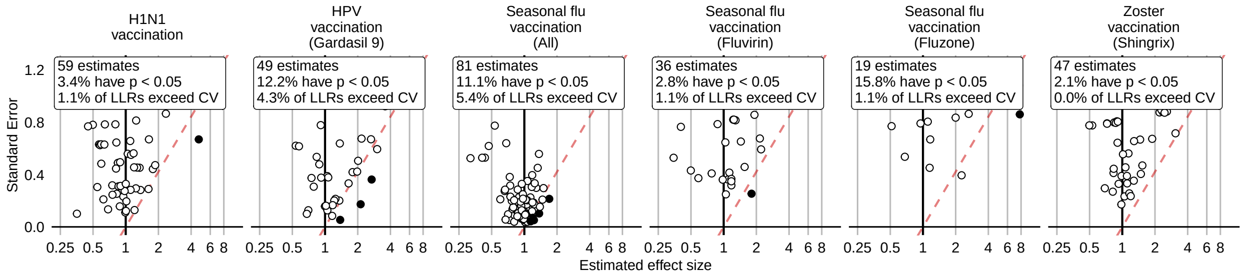

Figure 157: Negative control effect-size estimates and standard errors at the end of the study period using the SCCS method (Unadjusted SCCS excluding pre-vaccination window), in the MDCD database.

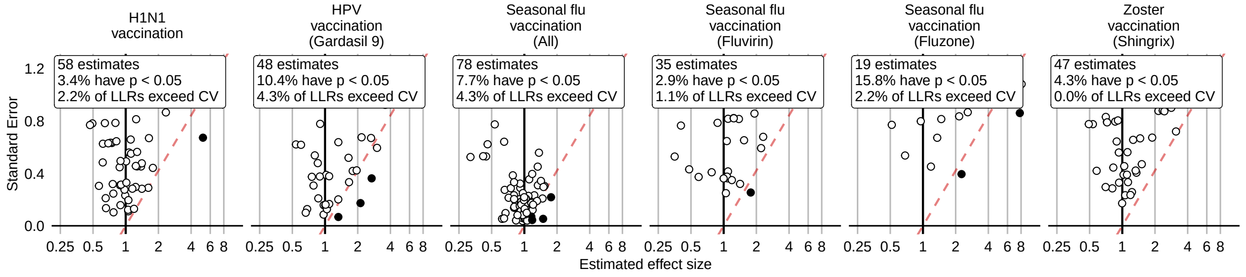

Figure 158: Negative control effect-size estimates and standard errors at the end of the study period using the SCCS method (Age & season adjusted SCCS excluding pre-vaccination window), in the MDCD database.

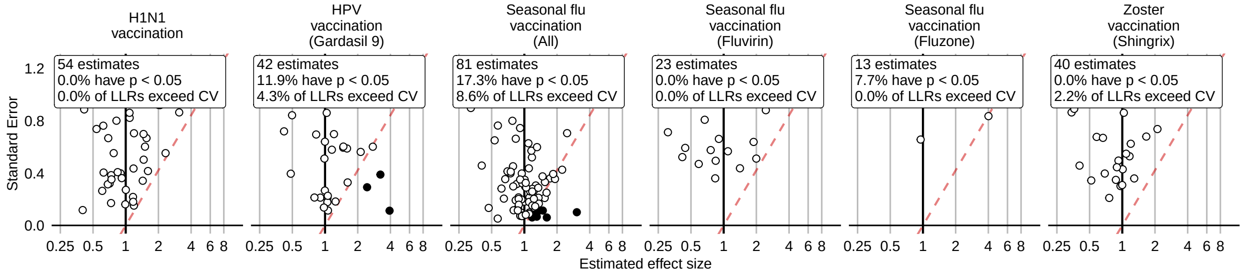

Figure 159: Negative control effect-size estimates and standard errors at the end of the study period using the SCCS method (SCRI with prior control interval), in the MDCD database.

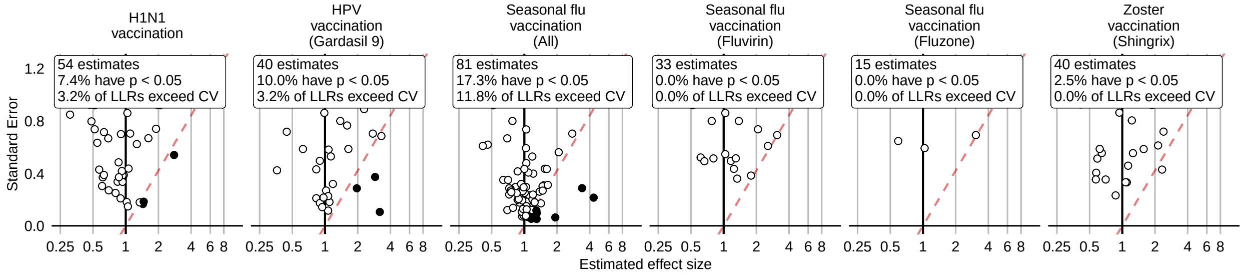

Figure 160: Negative control effect-size estimates and standard errors at the end of the study period using the SCCS method (SCRI with posterior control interval), in the MDCD database.

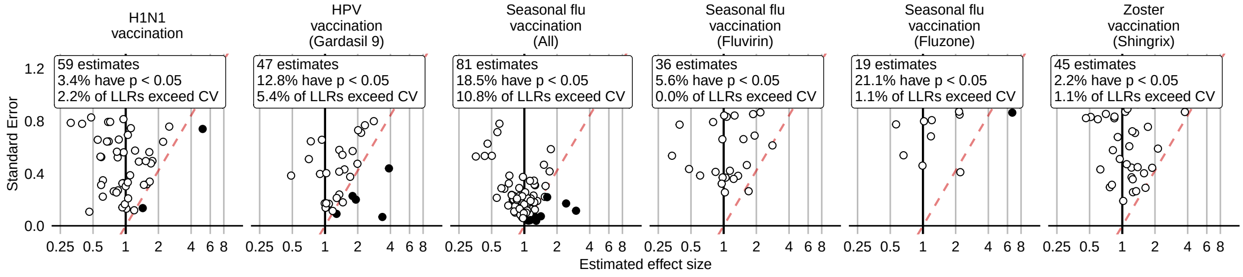

Figure 161: Negative control effect-size estimates and standard errors at the end of the study period using the SCCS method (Unadjusted SCCS excluding all pre-vaccination time), in the MDCD database.

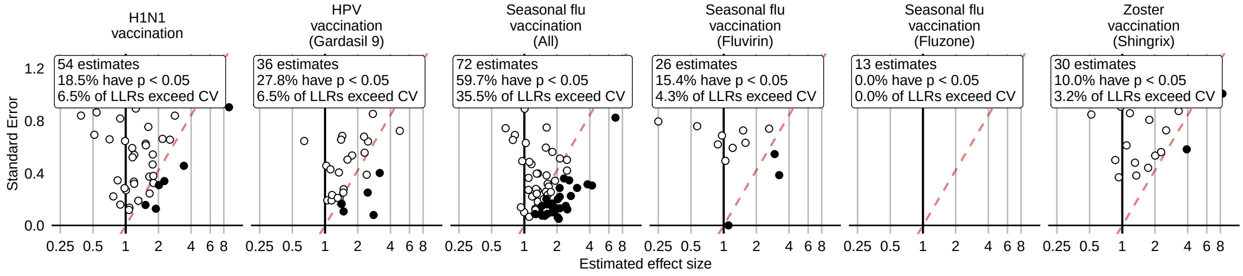

Figure 162: Negative control effect-size estimates and standard errors at the end of the study period using the CaseControl method (Age & sex adjusted, using random controls), in the MDCD database.

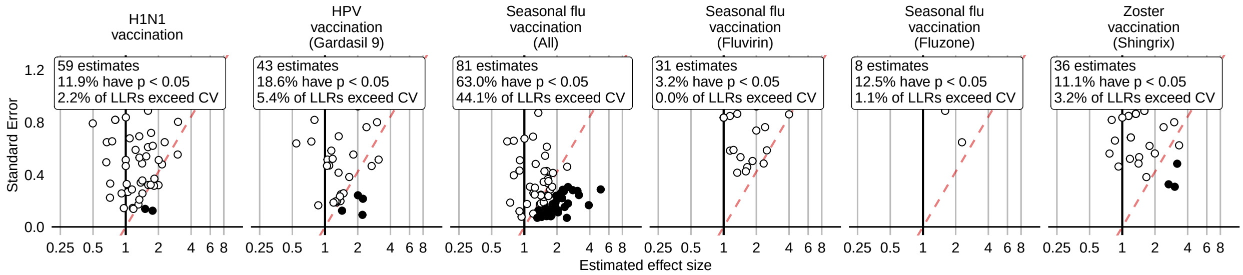

Figure 163: Negative control effect-size estimates and standard errors at the end of the study period using the CaseControl method (Age & sex matched controls), in the MDCD database.

#### 14.3 Negative controls effect size estimates in MDCR

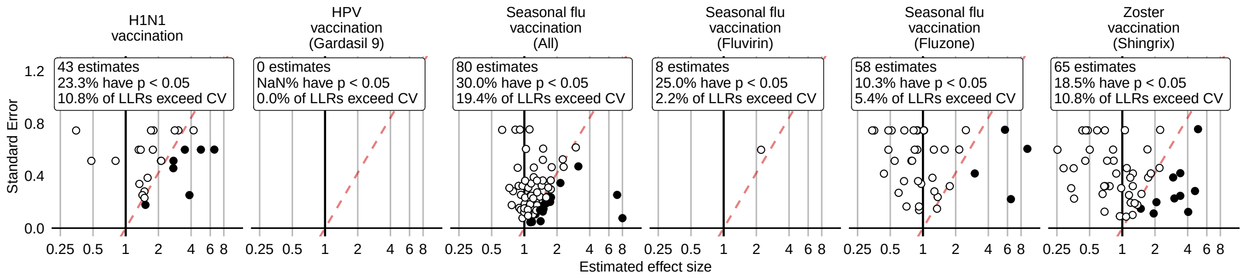

Figure 164: Negative control effect-size estimates and standard errors at the end of the study period using the HistoricalComparator method (Unadjusted, using entire historic period), in the MDCR database.

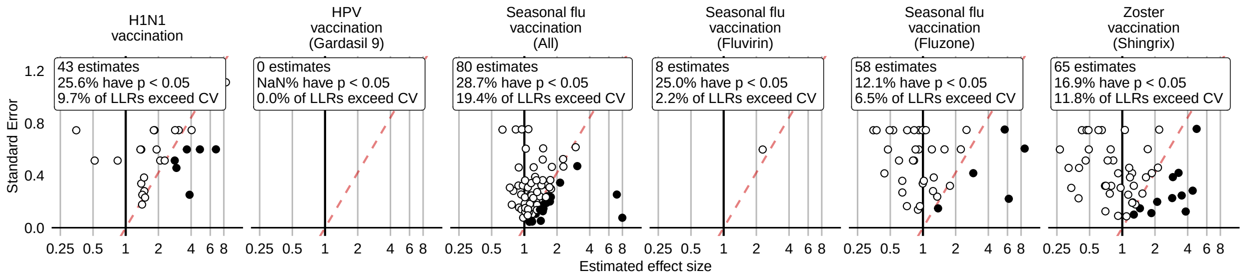

Figure 165: Negative control effect-size estimates and standard errors at the end of the study period using the HistoricalComparator method (Age & sex adjusted, using entire historic period), in the MDCR database.

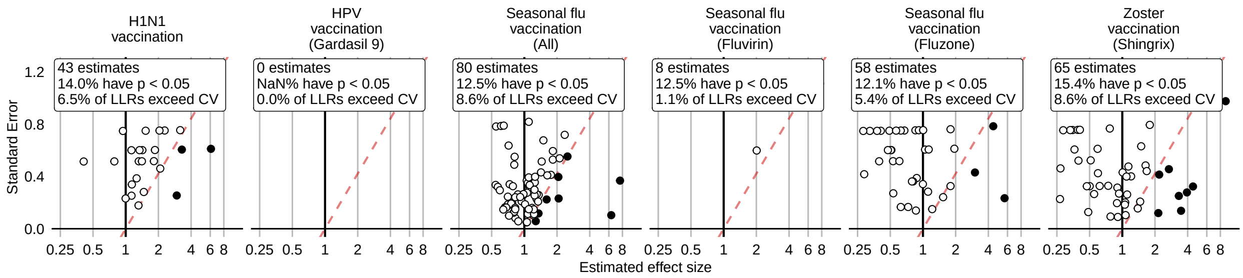

Figure 166: Negative control effect-size estimates and standard errors at the end of the study period using the HistoricalComparator method (Unadjusted, using TaR after historic visit), in the MDCR database.

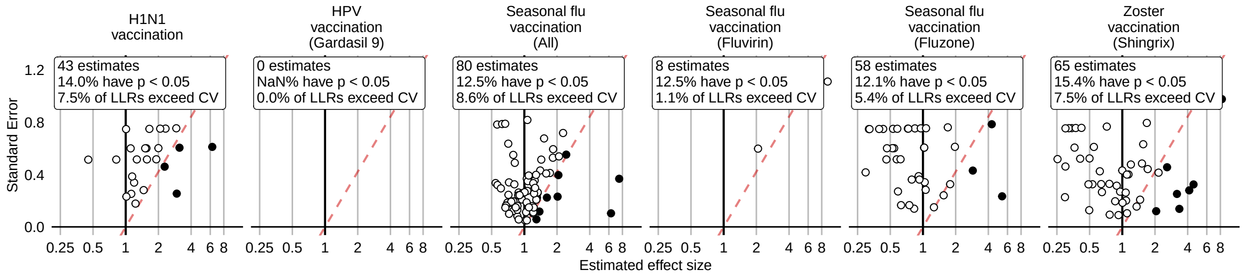

Figure 167: Negative control effect-size estimates and standard errors at the end of the study period using the HistoricalComparator method (Age & sex adjusted, using TaR after historic visit), in the MDCR database.

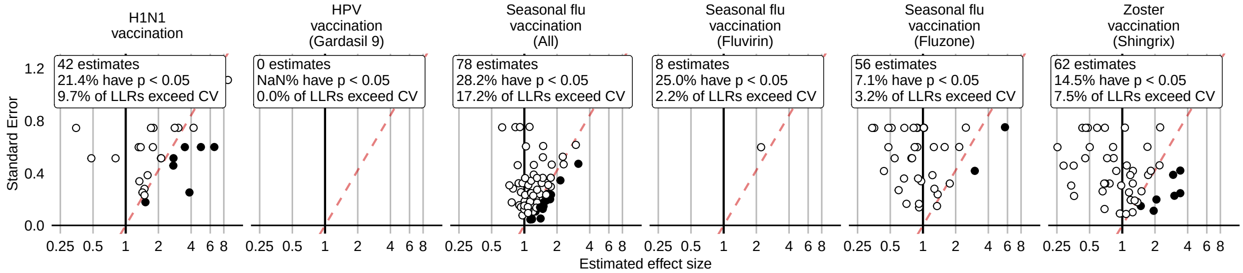

Figure 168: Negative control effect-size estimates and standard errors at the end of the study period using the HistoricalComparator method (Unadjusted, using entire historic period, filtered), in the MDCR database.

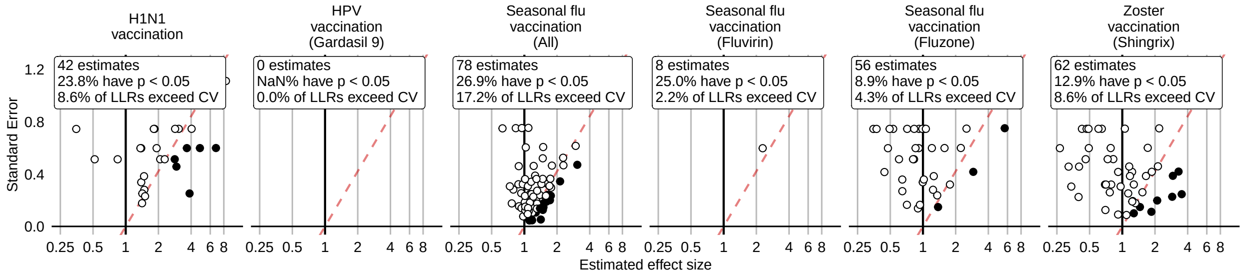

Figure 169: Negative control effect-size estimates and standard errors at the end of the study period using the HistoricalComparator method (Age & sex adjusted, using entire historic period, filtered), in the MDCR database.

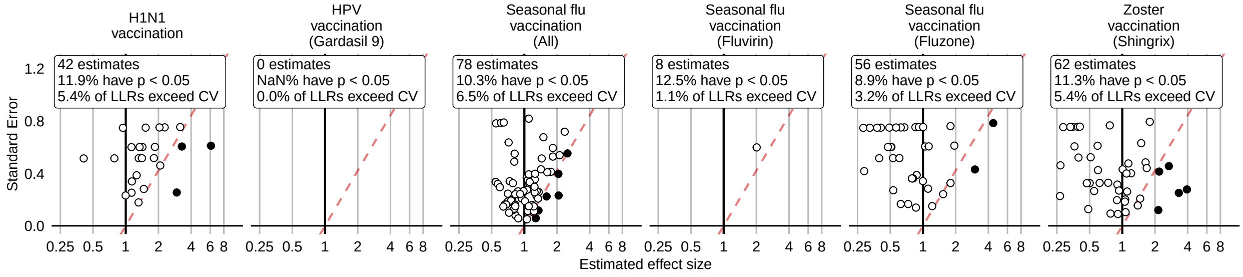

Figure 170: Negative control effect-size estimates and standard errors at the end of the study period using the HistoricalComparator method (Unadjusted, using TaR after historic visit, filtered), in the MDCR database.

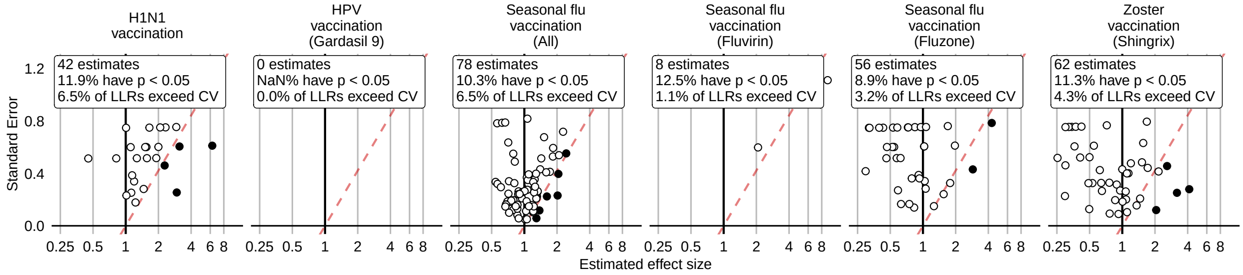

Figure 171: Negative control effect-size estimates and standard errors at the end of the study period using the HistoricalComparator method (Age & sex adjusted, using TaR after historic visit, filtered), in the MDCR database.

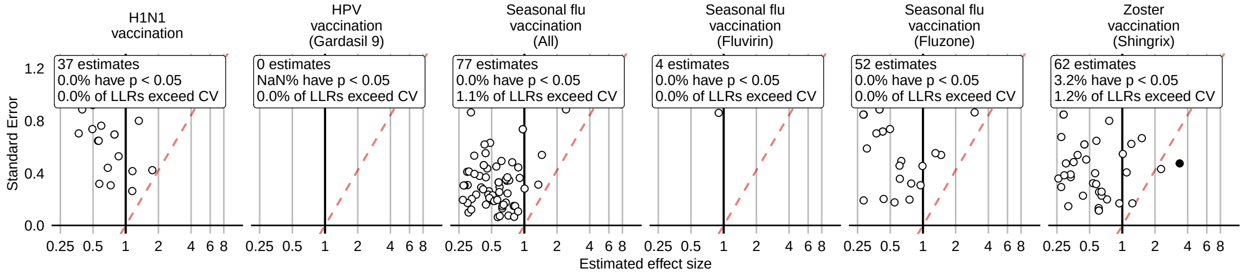

Figure 172: Negative control effect-size estimates and standard errors at the end of the study period using the CohortMethod method (Unadjusted, using outpatient visits as comparator), in the MDCR database.

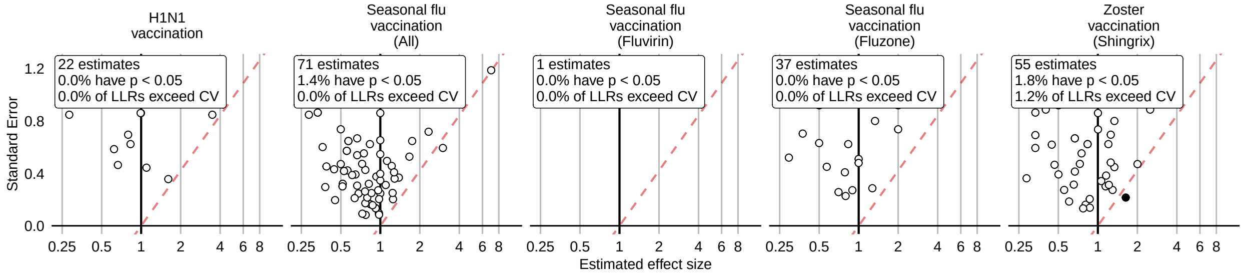

Figure 173: Negative control effect-size estimates and standard errors at the end of the study period using the CohortMethod method (PS matching, using outpatient visits as comparator), in the MDCR database.

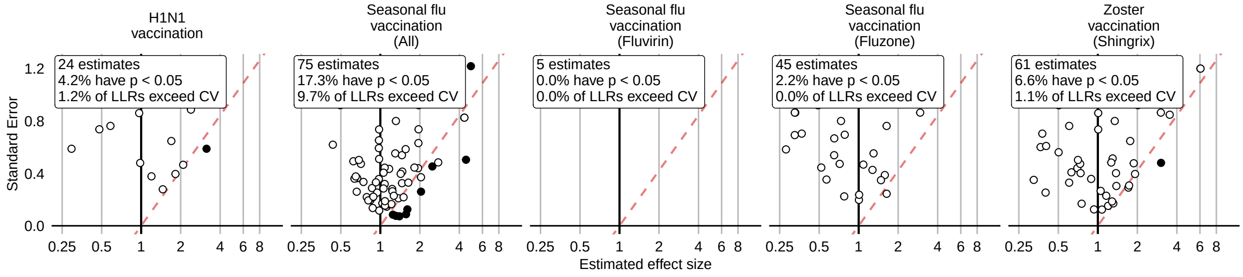

Figure 174: Negative control effect-size estimates and standard errors at the end of the study period using the CohortMethod method (Unadjusted, using random days as comparator), in the MDCR database.

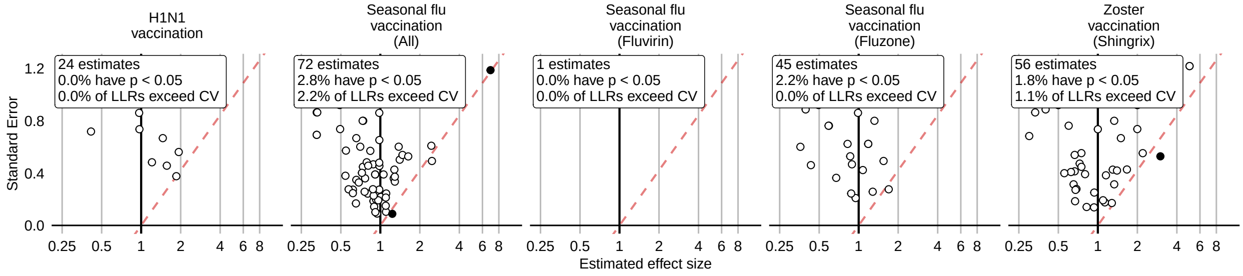

Figure 175: Negative control effect-size estimates and standard errors at the end of the study period using the CohortMethod method (PS matching, using random days as comparator), in the MDCR database.

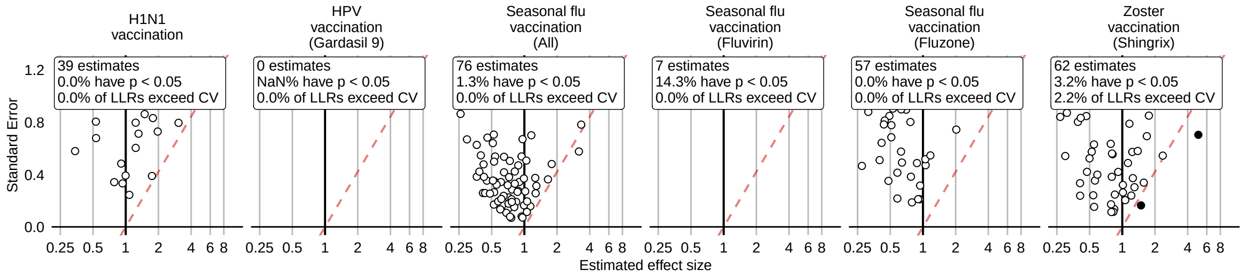

Figure 176: Negative control effect-size estimates and standard errors at the end of the study period using the CohortMethod method (PS stratification, using outpatient visits as comparator), in the MDCR database.

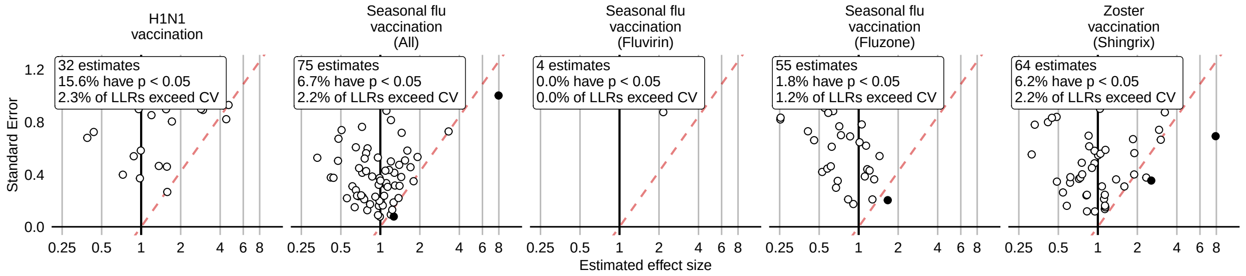

Figure 177: Negative control effect-size estimates and standard errors at the end of the study period using the CohortMethod method (PS stratification, using random days as comparator), in the MDCR database.

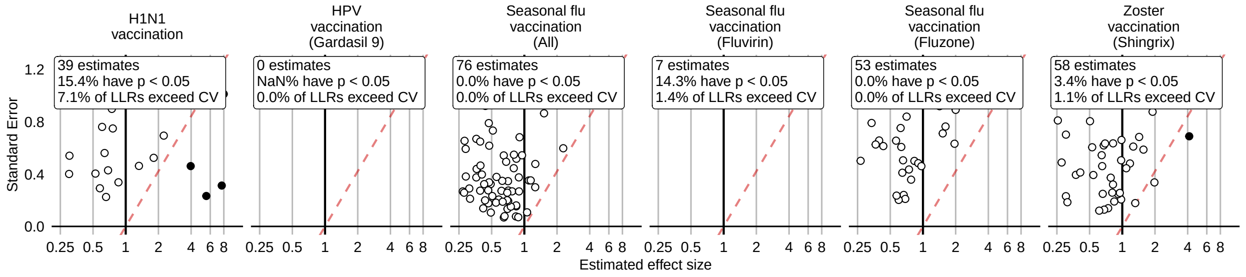

Figure 178: Negative control effect-size estimates and standard errors at the end of the study period using the CohortMethod method (PS weighting, using outpatient visits as comparator), in the MDCR database.

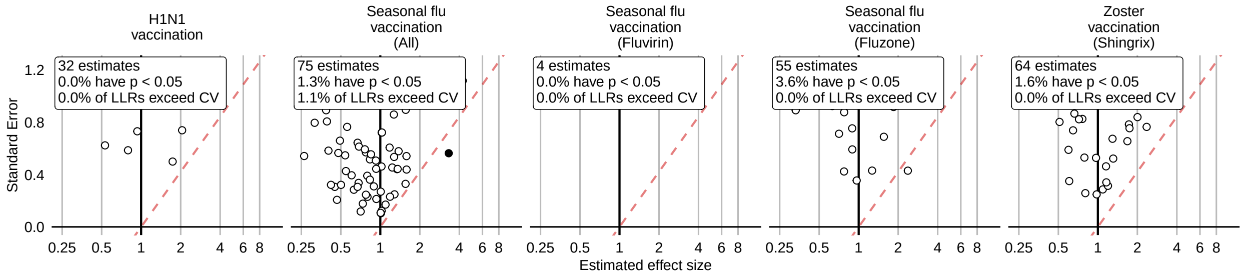

Figure 179: Negative control effect-size estimates and standard errors at the end of the study period using the CohortMethod method (PS weighting, using random days as comparator), in the MDCR database.

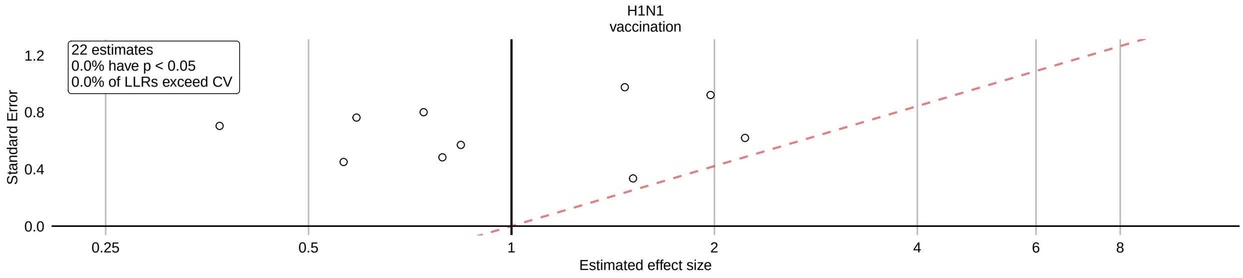

Figure 180: Negative control effect-size estimates and standard errors at the end of the study period using the CohortMethod method (Per-month PS matching, using outpatient visits as comparator), in the MDCR database.

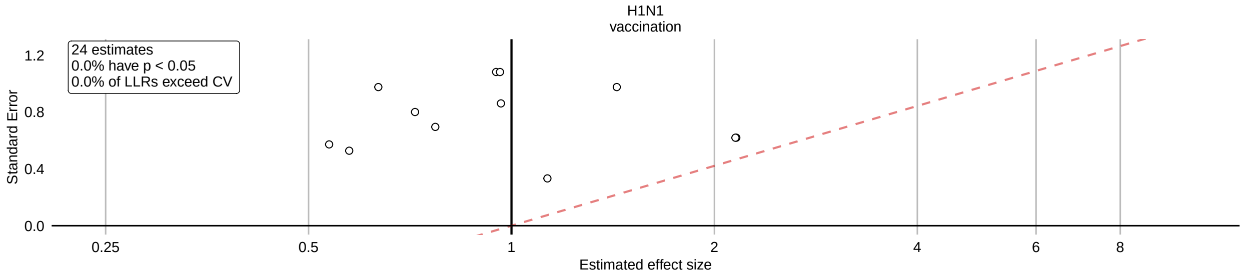

Figure 181: Negative control effect-size estimates and standard errors at the end of the study period using the CohortMethod method (Per-month PS matching, using random days as comparator), in the MDCR database.

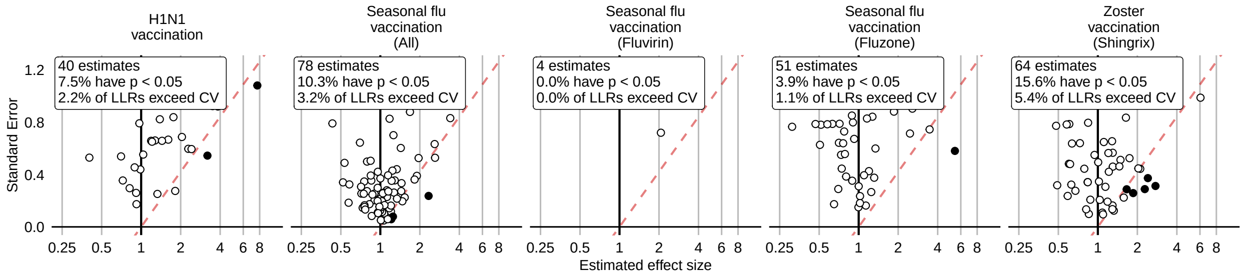

Figure 182: Negative control effect-size estimates and standard errors at the end of the study period using the SCCS method (Unadjusted SCCS excluding pre-vaccination window), in the MDCR database.

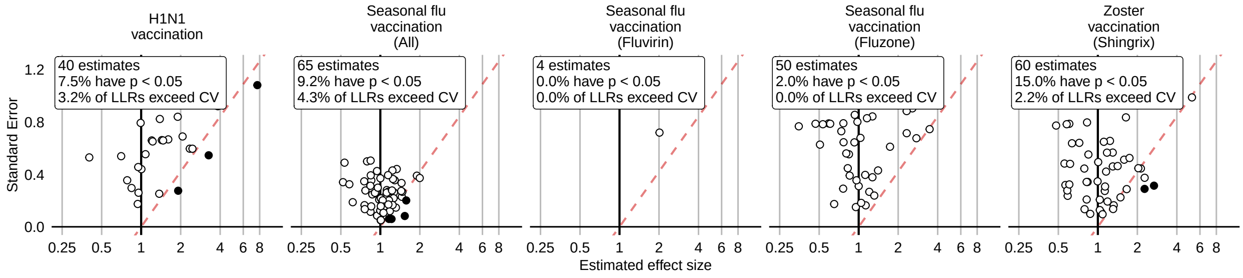

Figure 183: Negative control effect-size estimates and standard errors at the end of the study period using the SCCS method (Age & season adjusted SCCS excluding pre-vaccination window), in the MDCR database.

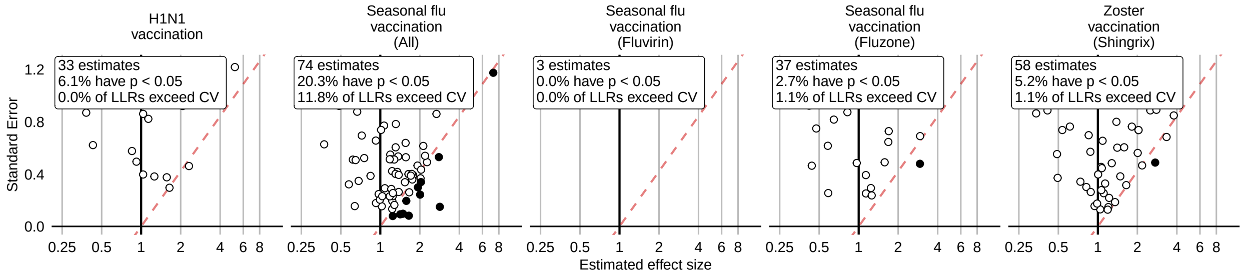

Figure 184: Negative control effect-size estimates and standard errors at the end of the study period using the SCCS method (SCRI with prior control interval), in the MDCR database.

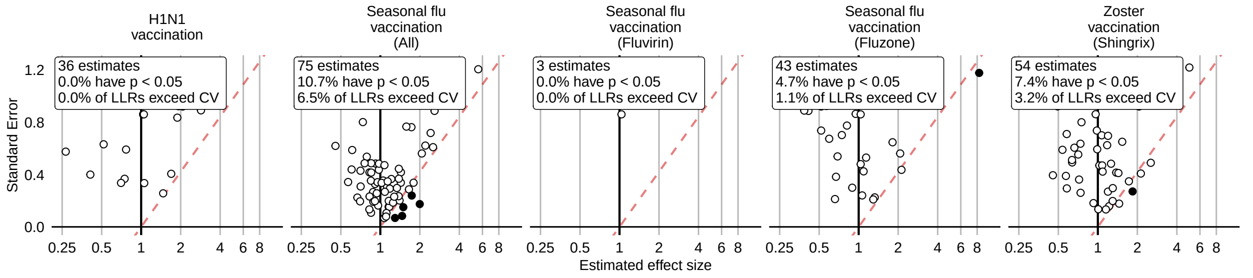

Figure 185: Negative control effect-size estimates and standard errors at the end of the study period using the SCCS method (SCRI with posterior control interval), in the MDCR database.

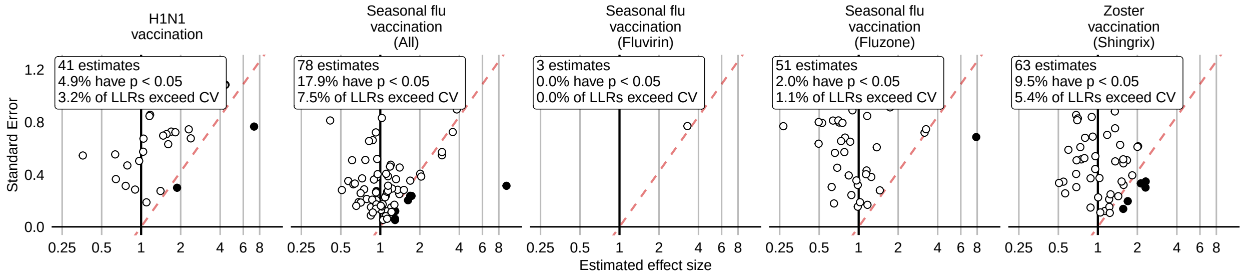

Figure 186: Negative control effect-size estimates and standard errors at the end of the study period using the SCCS method (Unadjusted SCCS excluding all pre-vaccination time), in the MDCR database.

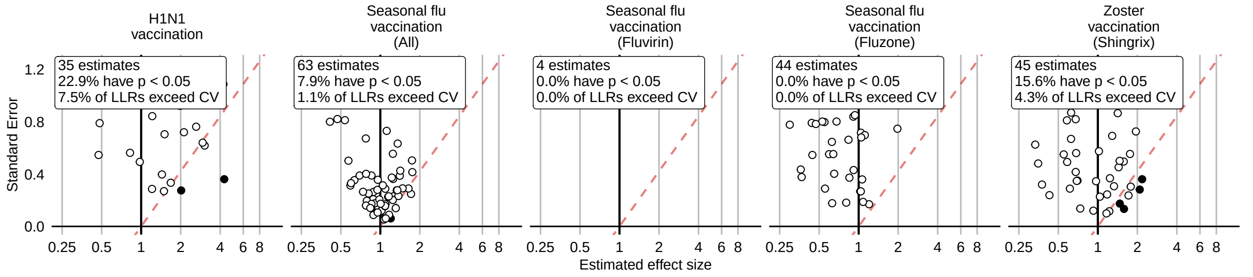

Figure 187: Negative control effect-size estimates and standard errors at the end of the study period using the CaseControl method (Age & sex adjusted, using random controls), in the MDCR database.

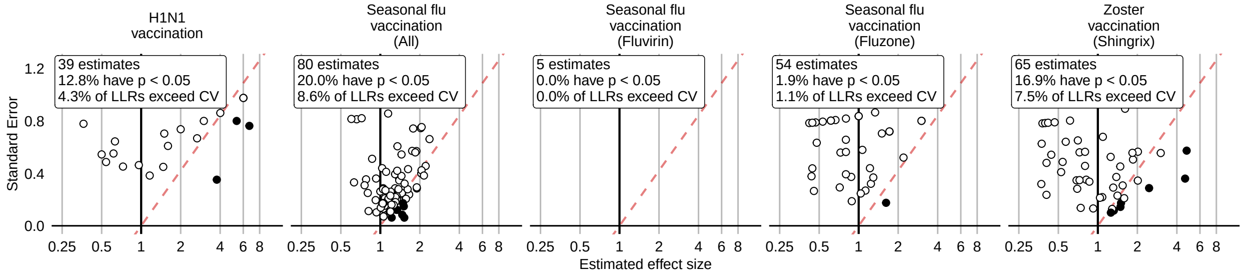

Figure 188: Negative control effect-size estimates and standard errors at the end of the study period using the CaseControl method (Age & sex matched controls), in the MDCR database.

#### 14.4 Negative controls effect size estimates in CCAE

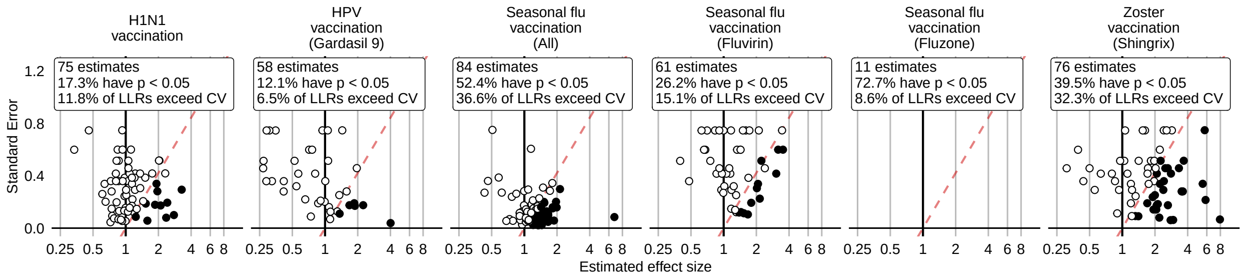

Figure 189: Negative control effect-size estimates and standard errors at the end of the study period using the HistoricalComparator method (Unadjusted, using entire historic period), in the CCAE database.

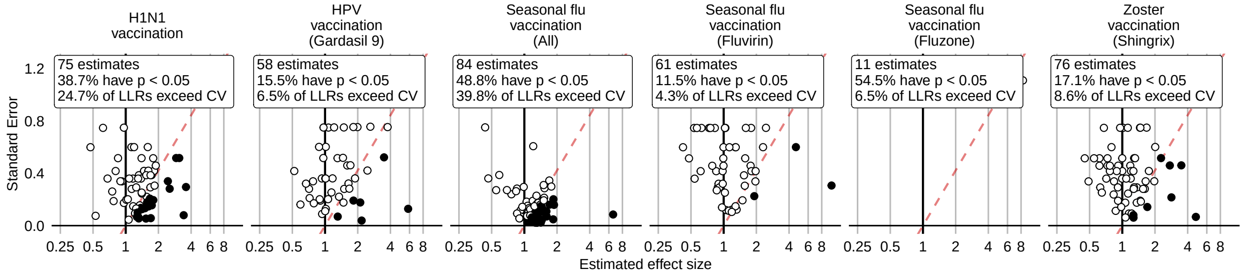

Figure 190: Negative control effect-size estimates and standard errors at the end of the study period using the HistoricalComparator method (Age & sex adjusted, using entire historic period), in the CCAE database.

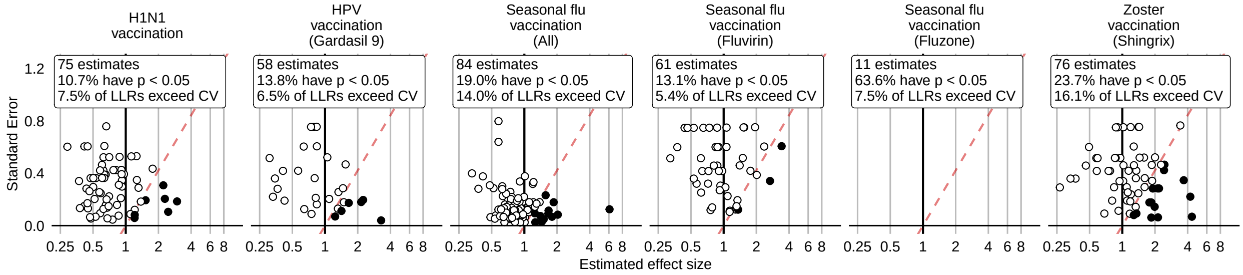

Figure 191: Negative control effect-size estimates and standard errors at the end of the study period using the HistoricalComparator method (Unadjusted, using TaR after historic visit), in the CCAE database.

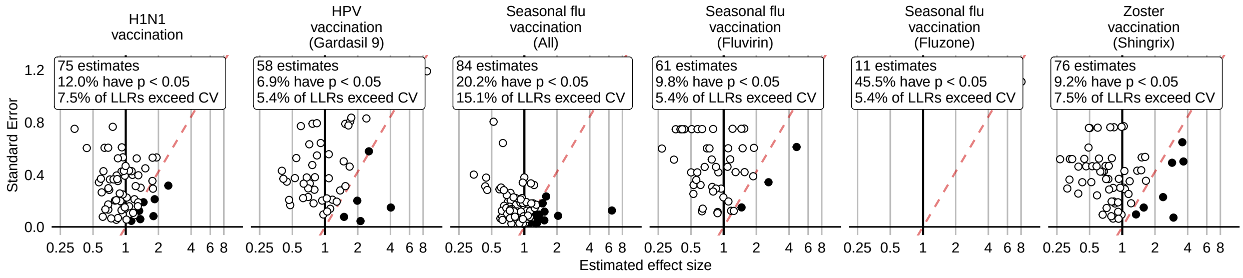

Figure 192: Negative control effect-size estimates and standard errors at the end of the study period using the HistoricalComparator method (Age & sex adjusted, using TaR after historic visit), in the CCAE database.

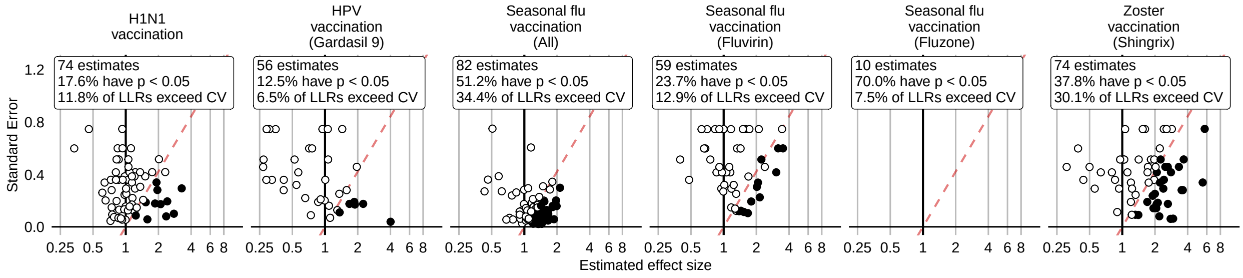

Figure 193: Negative control effect-size estimates and standard errors at the end of the study period using the HistoricalComparator method (Unadjusted, using entire historic period, filtered), in the CCAE database.

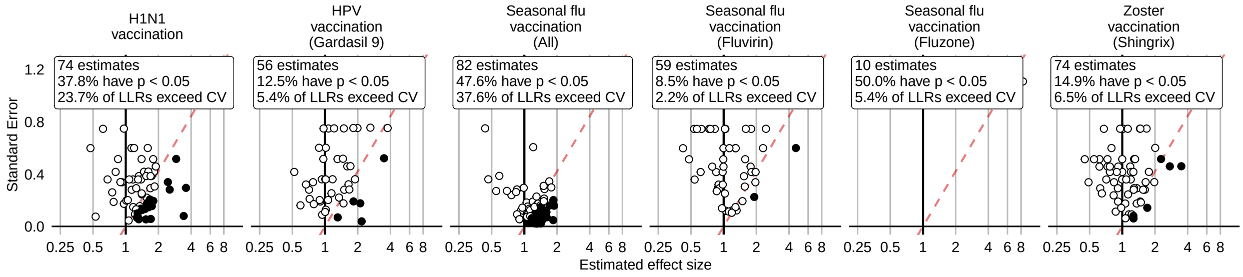

Figure 194: Negative control effect-size estimates and standard errors at the end of the study period using the HistoricalComparator method (Age & sex adjusted, using entire historic period, filtered), in the CCAE database.

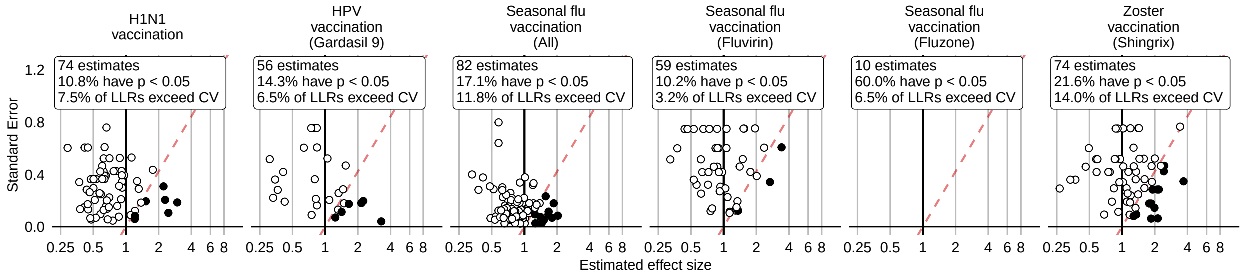

Figure 195: Negative control effect-size estimates and standard errors at the end of the study period using the HistoricalComparator method (Unadjusted, using TaR after historic visit, filtered), in the CCAE database.

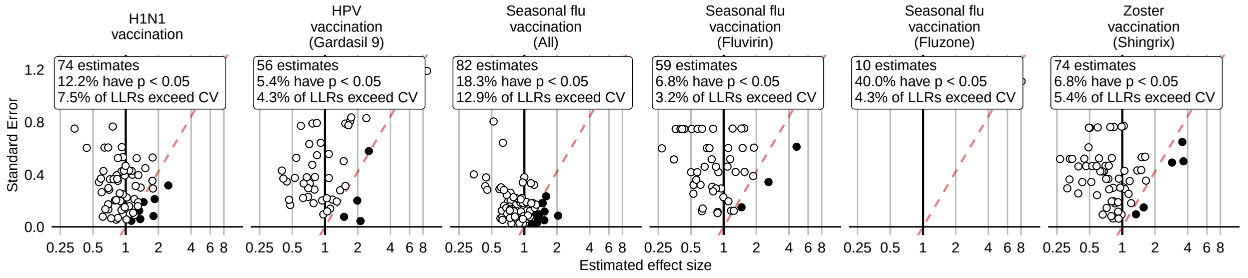

Figure 196: Negative control effect-size estimates and standard errors at the end of the study period using the HistoricalComparator method (Age & sex adjusted, using TaR after historic visit, filtered), in the CCAE database.

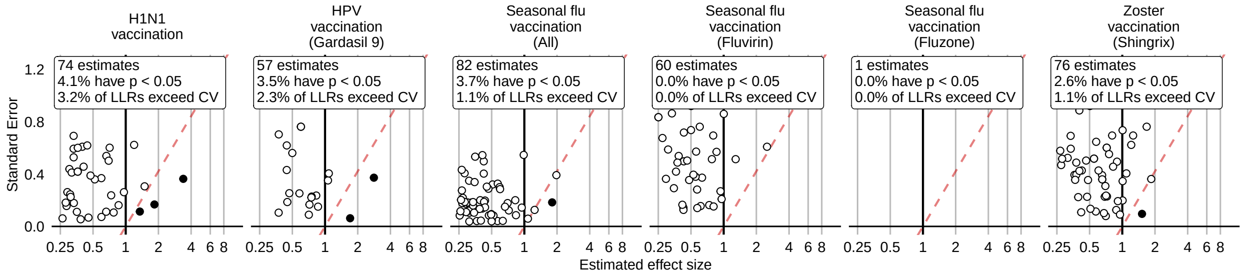

Figure 197: Negative control effect-size estimates and standard errors at the end of the study period using the CohortMethod method (Unadjusted, using outpatient visits as comparator), in the CCAE database.

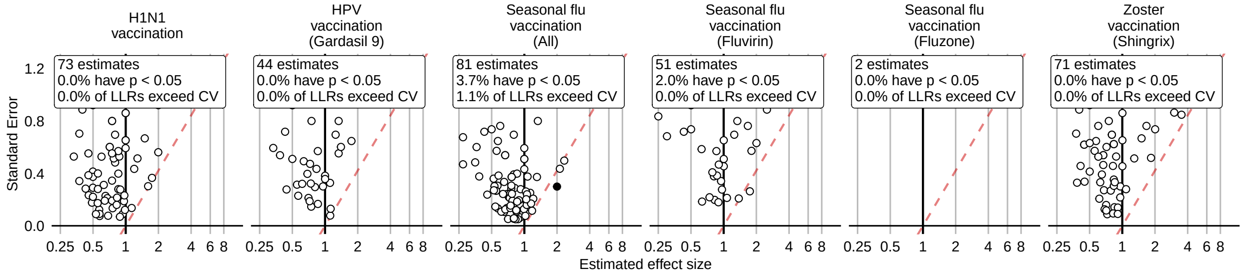

Figure 198: Negative control effect-size estimates and standard errors at the end of the study period using the CohortMethod method (PS matching, using outpatient visits as comparator), in the CCAE database.

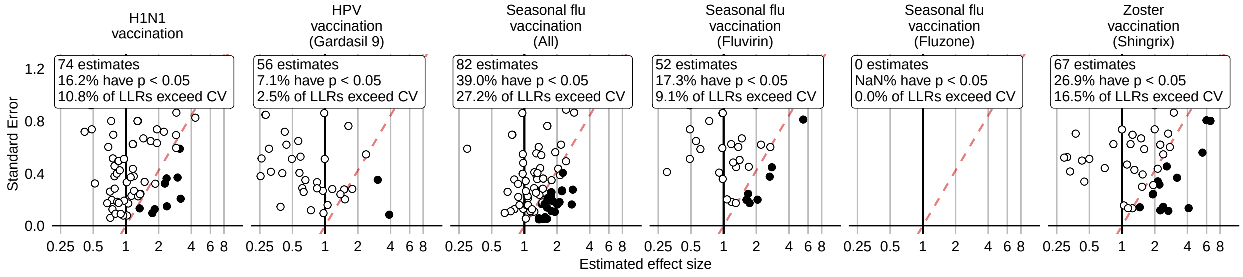

Figure 199: Negative control effect-size estimates and standard errors at the end of the study period using the CohortMethod method (Unadjusted, using random days as comparator), in the CCAE database.

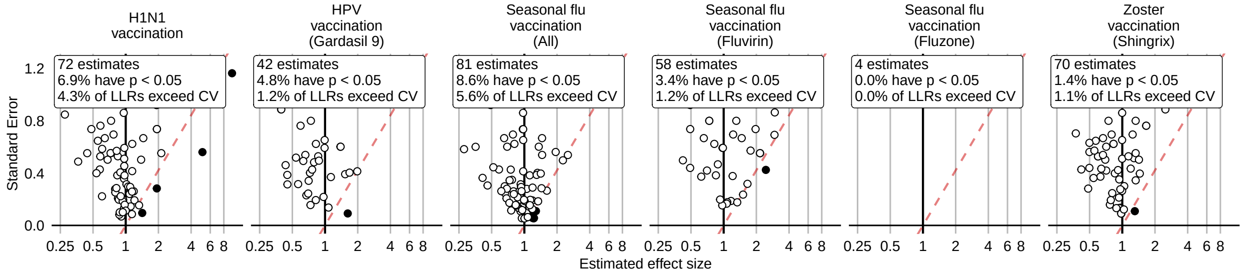

Figure 200: Negative control effect-size estimates and standard errors at the end of the study period using the CohortMethod method (PS matching, using random days as comparator), in the CCAE database.

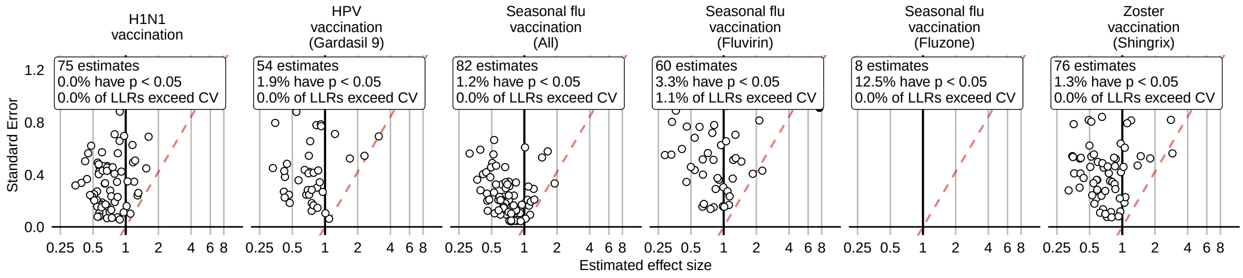

Figure 201: Negative control effect-size estimates and standard errors at the end of the study period using the CohortMethod method (PS stratification, using outpatient visits as comparator), in the CCAE database.

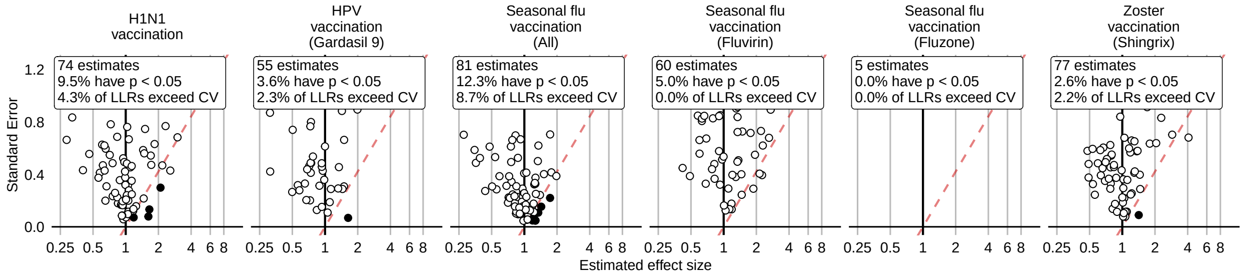

Figure 202: Negative control effect-size estimates and standard errors at the end of the study period using the CohortMethod method (PS stratification, using random days as comparator), in the CCAE database.

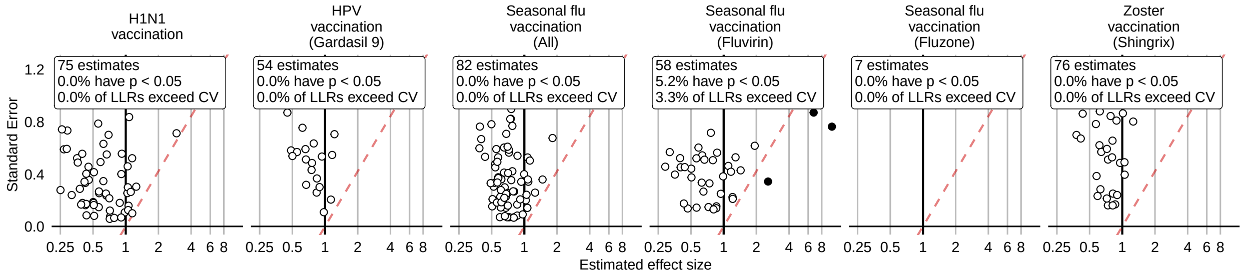

Figure 203: Negative control effect-size estimates and standard errors at the end of the study period using the CohortMethod method (PS weighting, using outpatient visits as comparator), in the CCAE database.

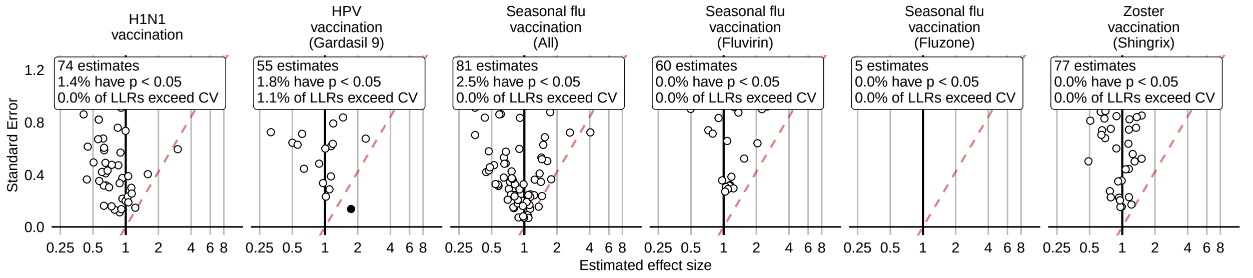

Figure 204: Negative control effect-size estimates and standard errors at the end of the study period using the CohortMethod method (PS weighting, using random days as comparator), in the CCAE database.

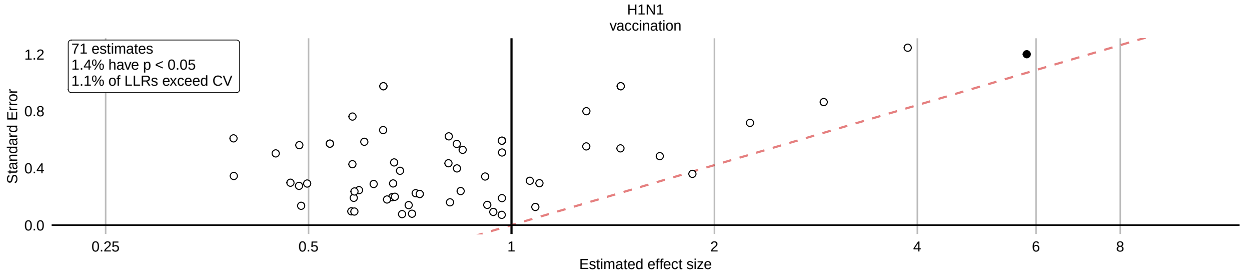

Figure 205: Negative control effect-size estimates and standard errors at the end of the study period using the CohortMethod method (Per-month PS matching, using outpatient visits as comparator), in the CCAE database.

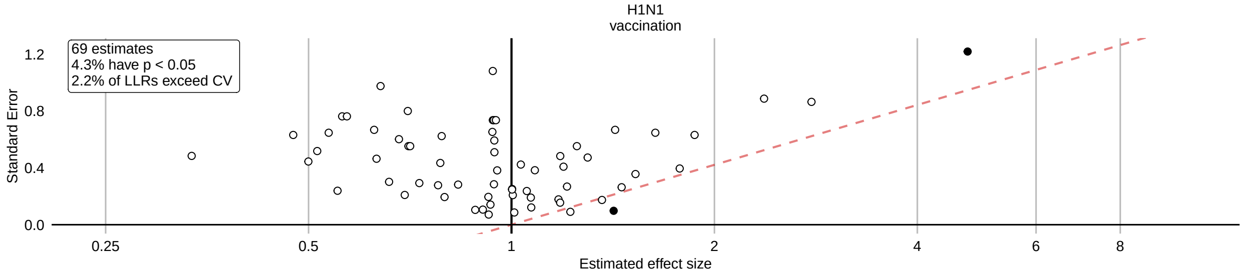

Figure 206: Negative control effect-size estimates and standard errors at the end of the study period using the CohortMethod method (Per-month PS matching, using random days as comparator), in the CCAE database.

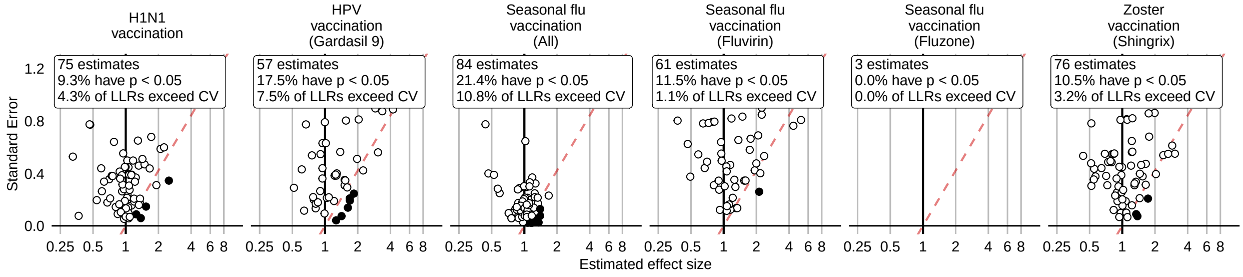

Figure 207: Negative control effect-size estimates and standard errors at the end of the study period using the SCCS method (Unadjusted SCCS excluding pre-vaccination window), in the CCAE database.

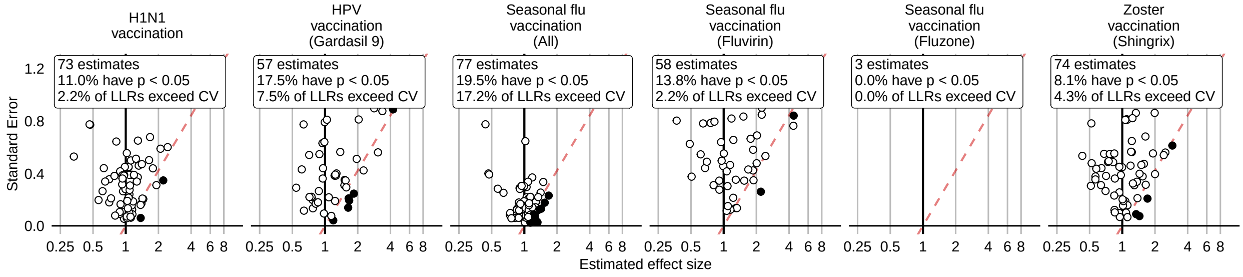

Figure 208: Negative control effect-size estimates and standard errors at the end of the study period using the SCCS method (Age & season adjusted SCCS excluding pre-vaccination window), in the CCAE database.

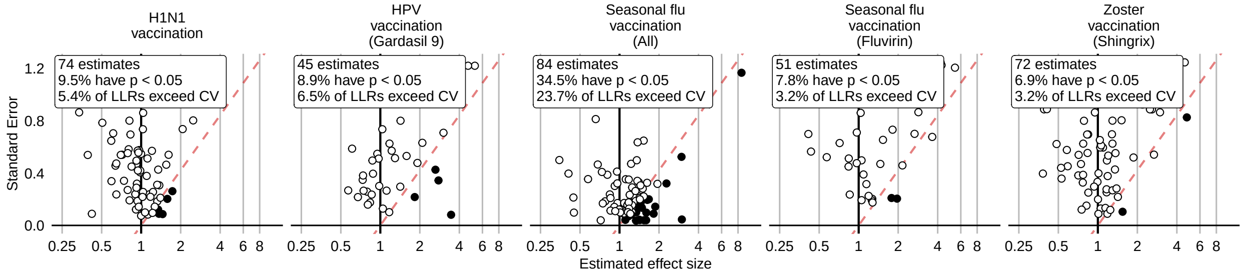

Figure 209: Negative control effect-size estimates and standard errors at the end of the study period using the SCCS method (SCRI with prior control interval), in the CCAE database.

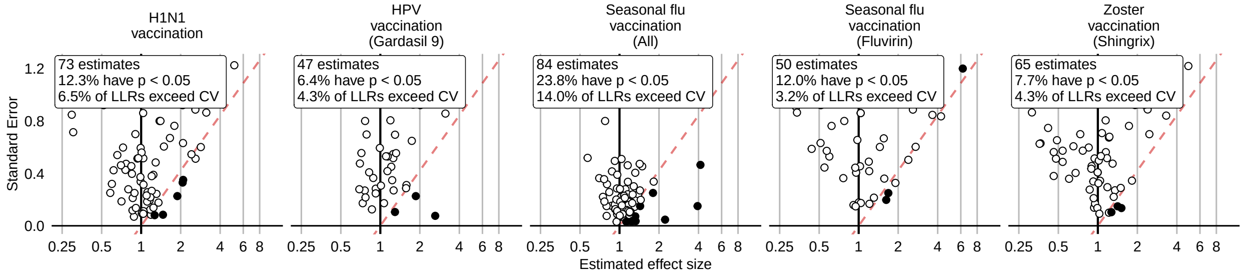

Figure 210: Negative control effect-size estimates and standard errors at the end of the study period using the SCCS method (SCRI with posterior control interval), in the CCAE database.

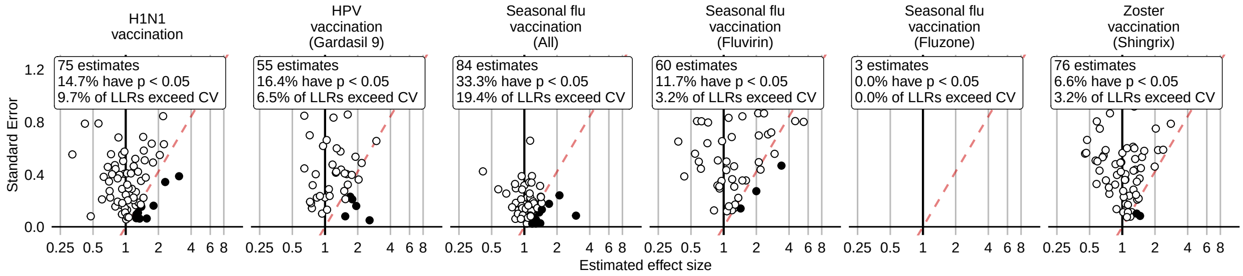

Figure 211: Negative control effect-size estimates and standard errors at the end of the study period using the SCCS method (Unadjusted SCCS excluding all pre-vaccination time), in the CCAE database.

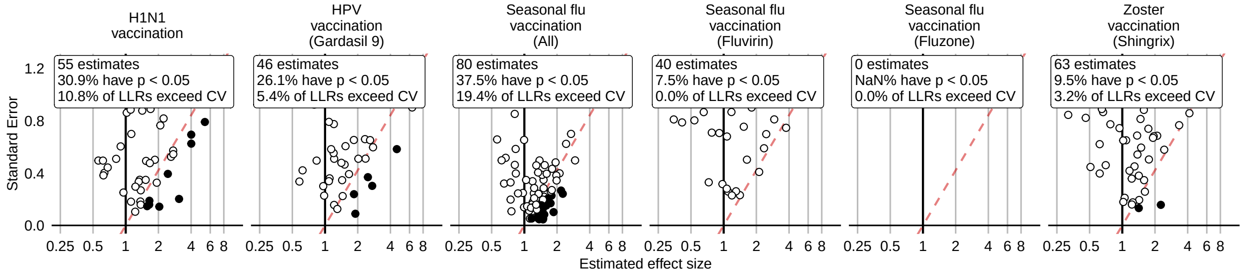

Figure 212: Negative control effect-size estimates and standard errors at the end of the study period using the CaseControl method (Age & sex adjusted, using random controls), in the CCAE database.

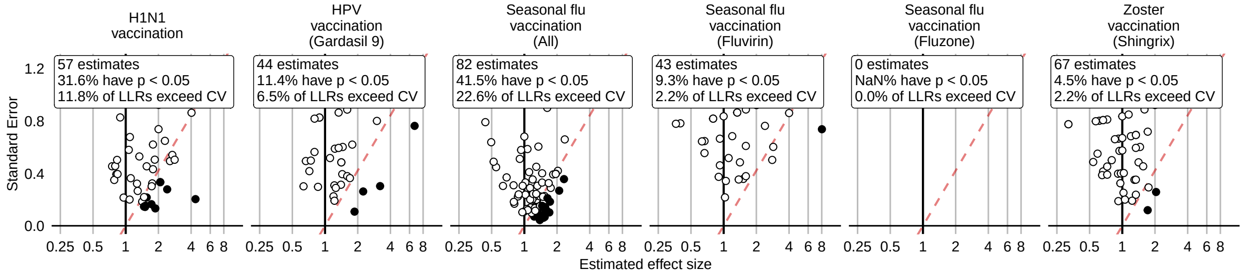

Figure 213: Negative control effect-size estimates and standard errors at the end of the study period using the CaseControl method (Age & sex matched controls), in the CCAE database.

### 15 Negative controls effect log likelihood ratios

Log likelihood ratios (LLR) for the negative controls at various points in time. Closed dots indicate the LLR in that period exceeded the critical value. The critical value depends on sample size within and across periods, and is therefore different for each control.

#### 15.1 Negative controls log likelihood ratios in Optum EHR

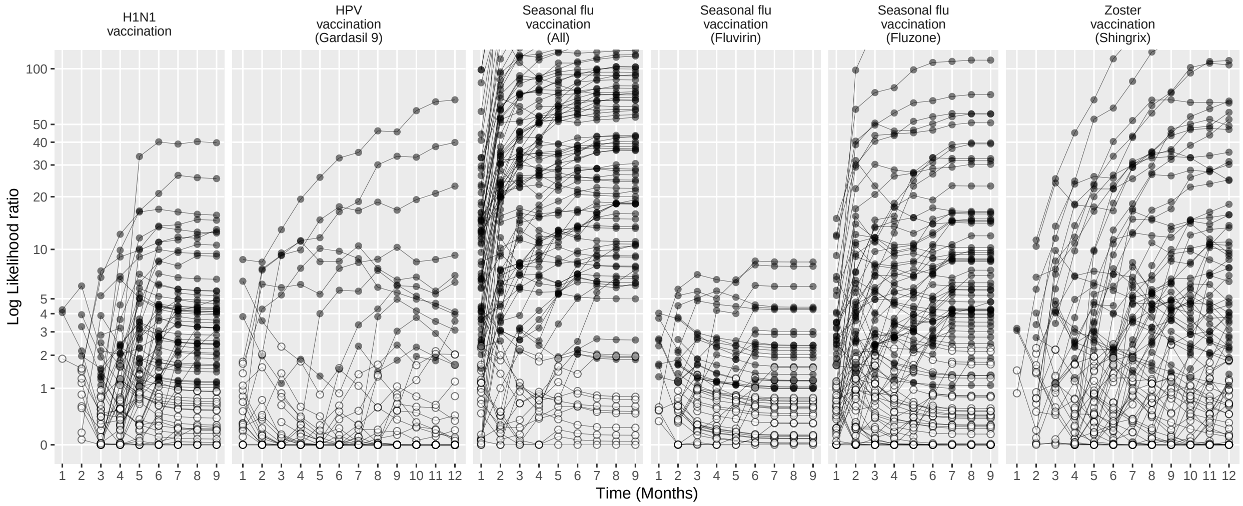

Figure 214: Negative control log likelihood ratios per month using the HistoricalComparator method (Unadjusted, using entire historic period), in the Optum EHR database.

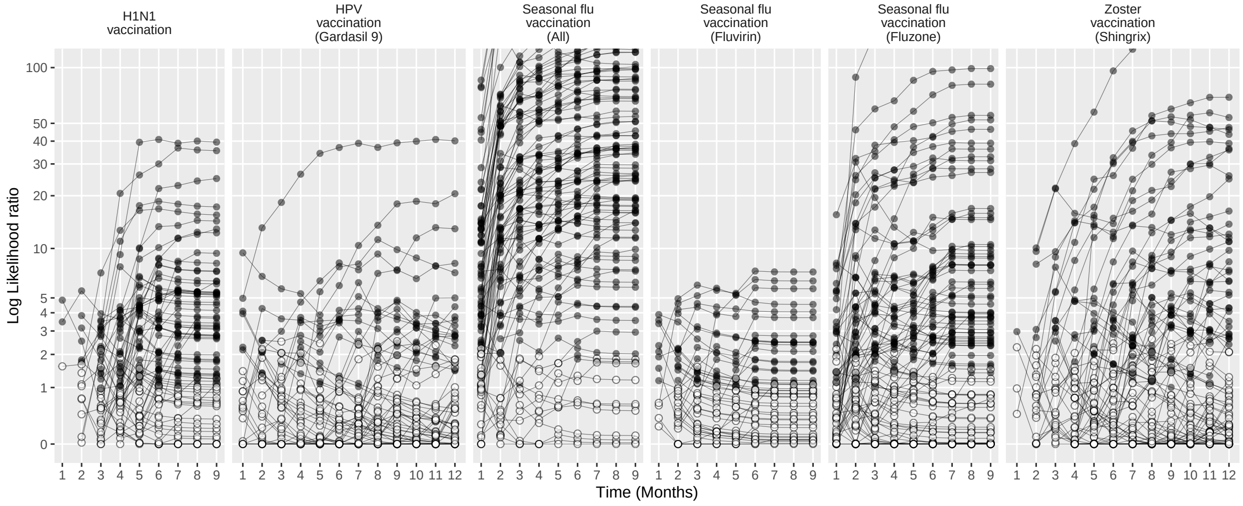

Figure 215: Negative control log likelihood ratios per month using the HistoricalComparator method (Age & sex adjusted, using entire historic period), in the Optum EHR database.

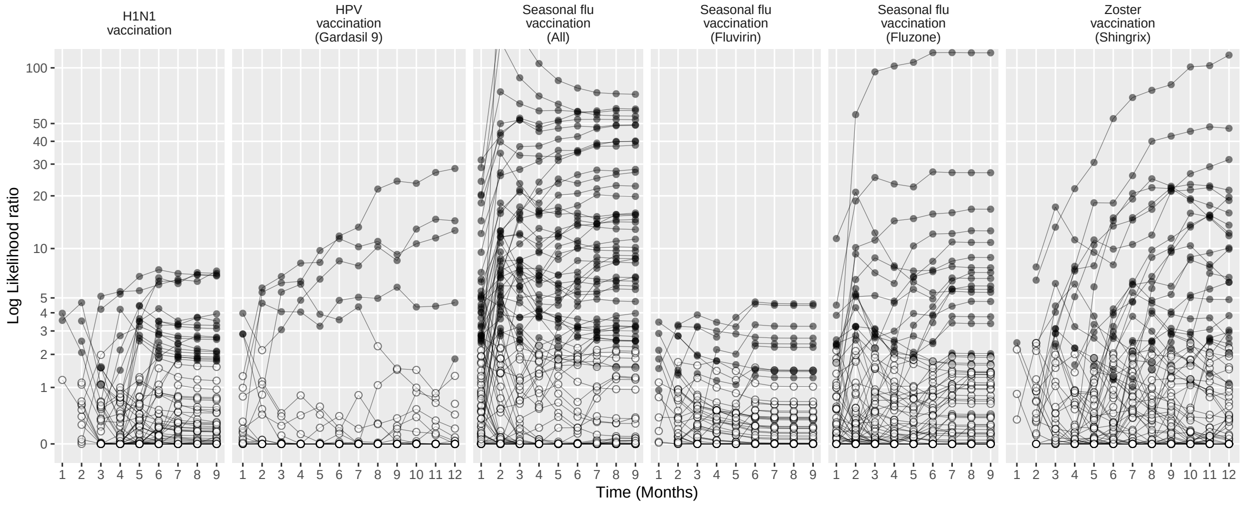

Figure 216: Negative control log likelihood ratios per month using the HistoricalComparator method (Unadjusted, using TaR after historic visit), in the Optum EHR database.

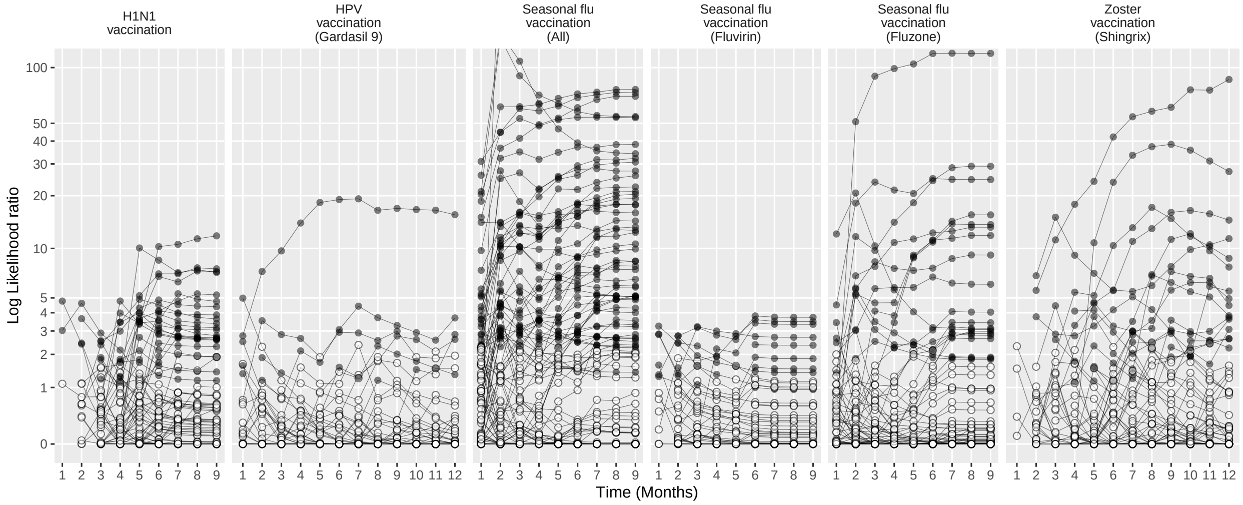

Figure 217: Negative control log likelihood ratios per month using the HistoricalComparator method (Age & sex adjusted, using TaR after historic visit), in the Optum EHR database.

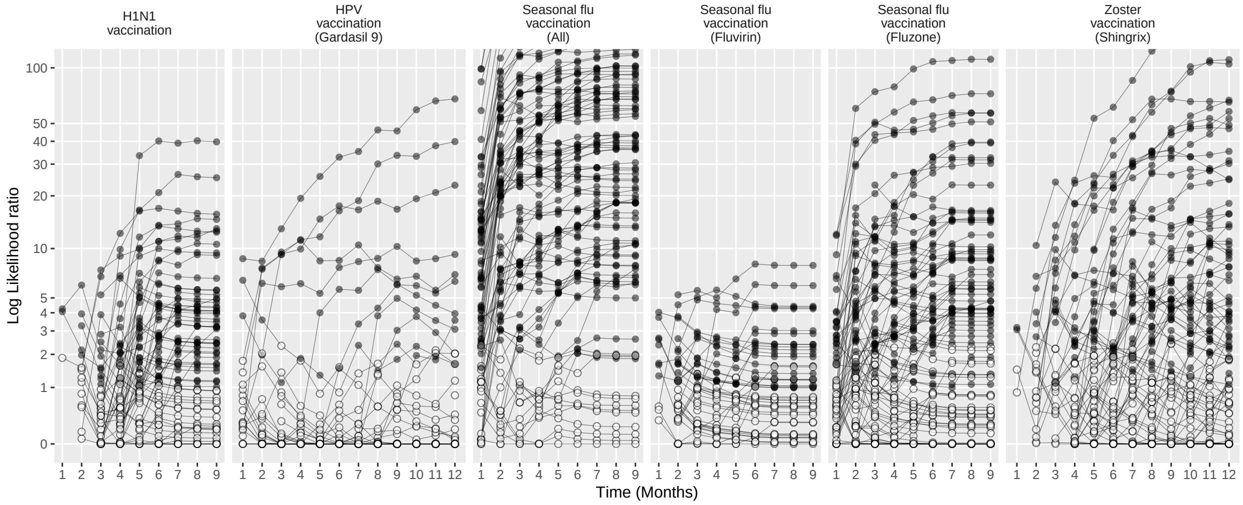

Figure 218: Negative control log likelihood ratios per month using the HistoricalComparator method (Unadjusted, using entire historic period, filtered), in the Optum EHR database.

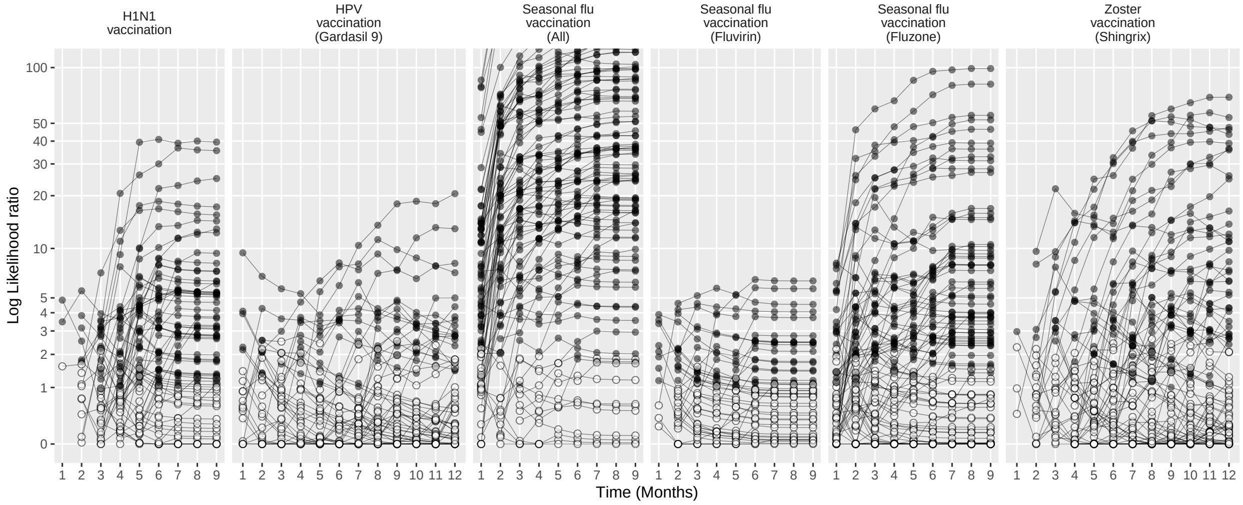

Figure 219: Negative control log likelihood ratios per month using the HistoricalComparator method (Age & sex adjusted, using entire historic period, filtered), in the Optum EHR database.

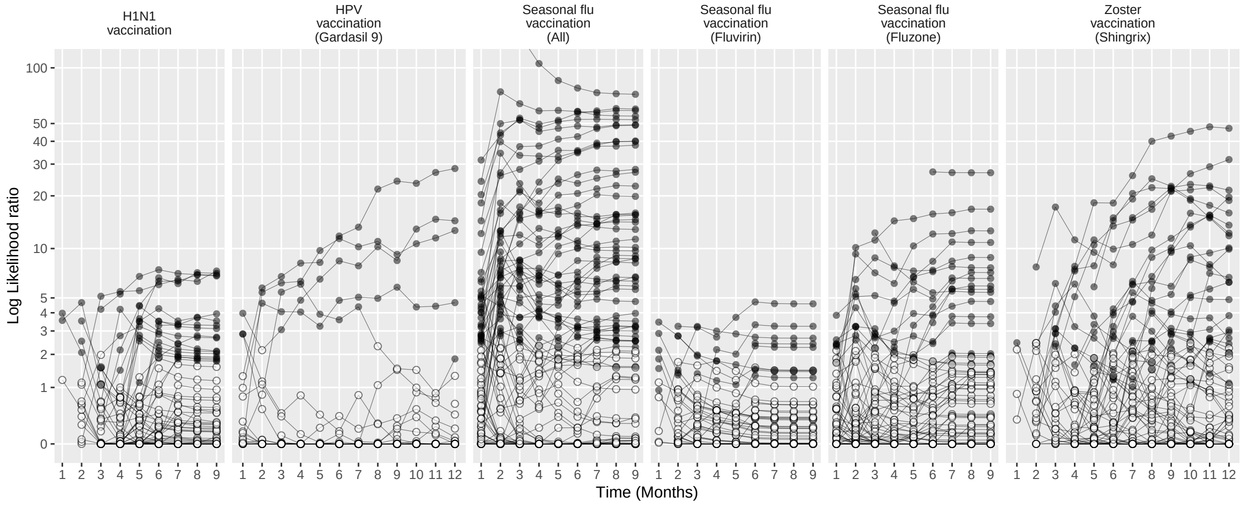

Figure 220: Negative control log likelihood ratios per month using the HistoricalComparator method (Unadjusted, using TaR after historic visit, filtered), in the Optum EHR database.

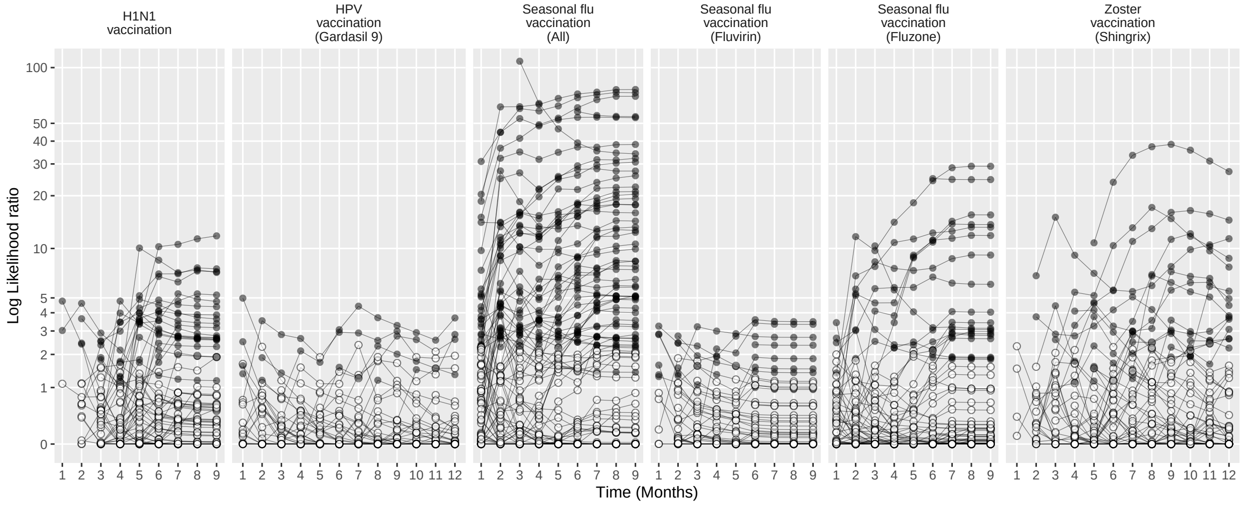

Figure 221: Negative control log likelihood ratios per month using the HistoricalComparator method (Age & sex adjusted, using TaR after historic visit, filtered), in the Optum EHR database.

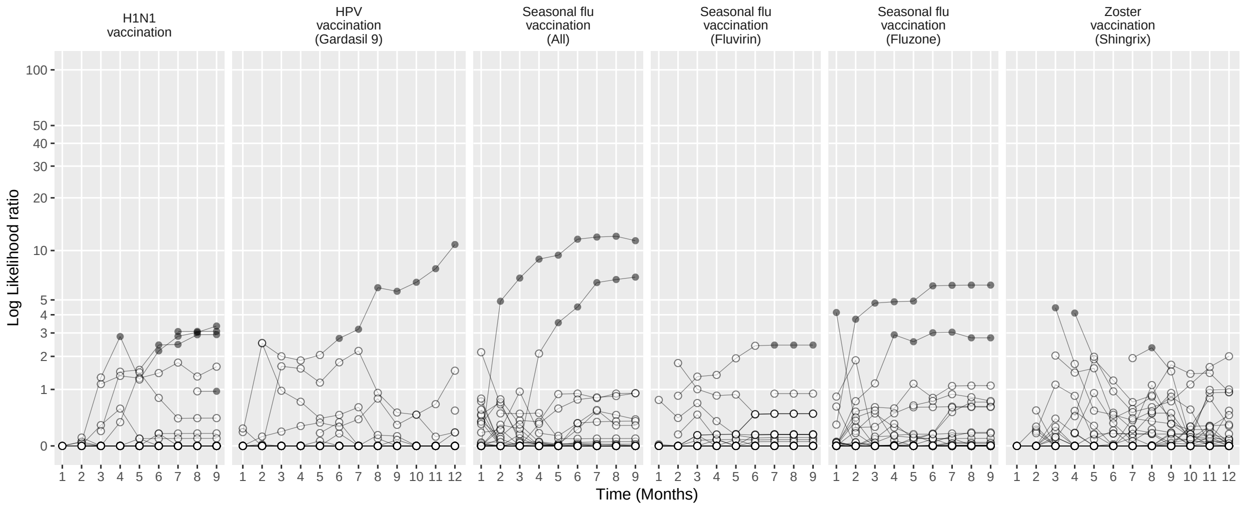

Figure 222: Negative control log likelihood ratios per month using the CohortMethod method (Unadjusted, using outpatient visits as comparator), in the Optum EHR database.

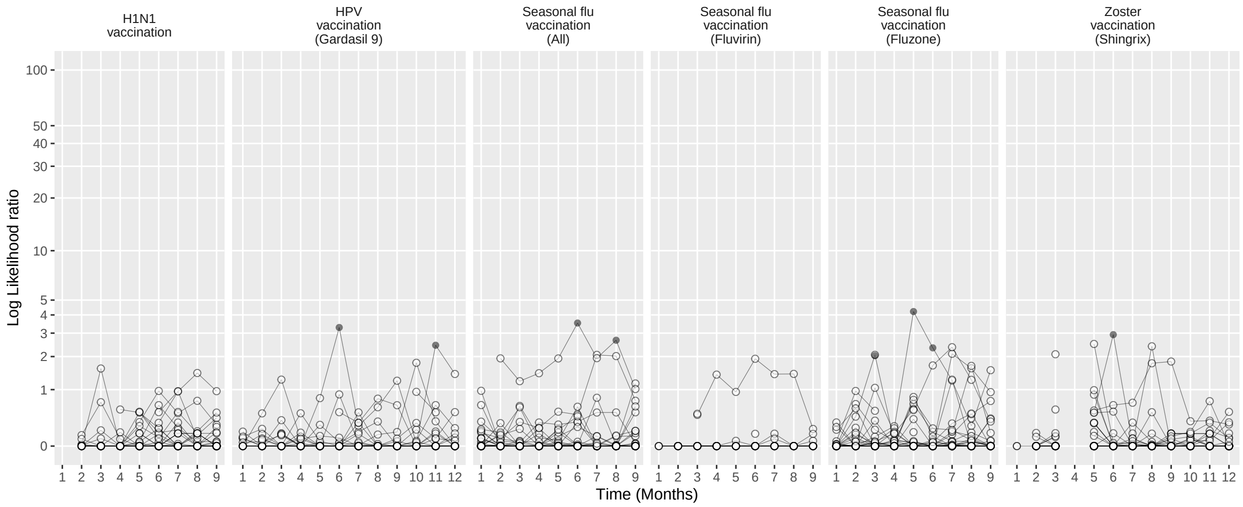

Figure 223: Negative control log likelihood ratios per month using the CohortMethod method (PS matching, using outpatient visits as comparator), in the Optum EHR database.

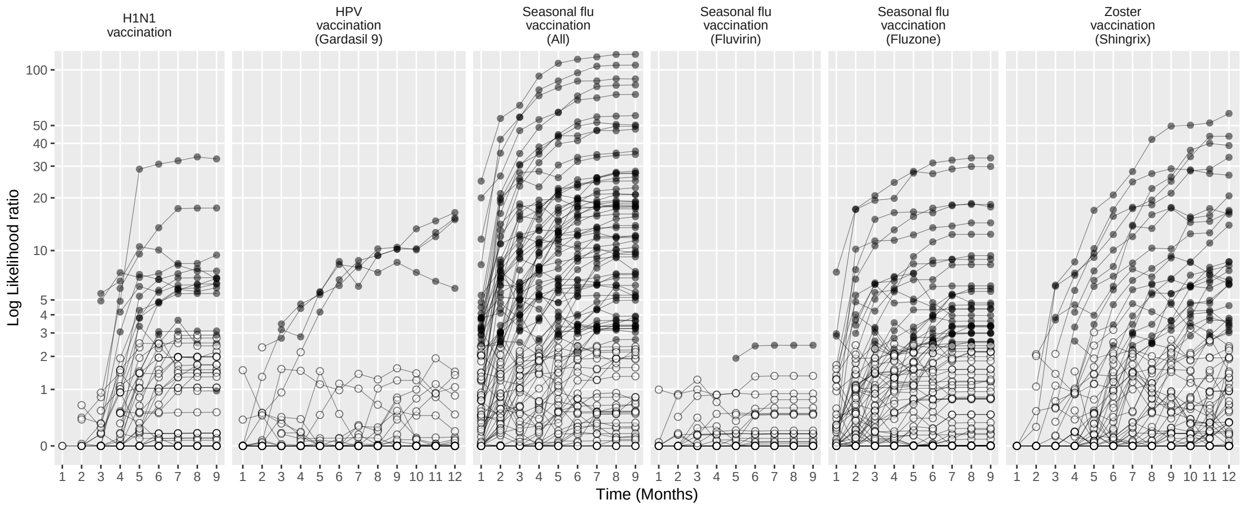

Figure 224: Negative control log likelihood ratios per month using the CohortMethod method (Unadjusted, using random days as comparator), in the Optum EHR database.

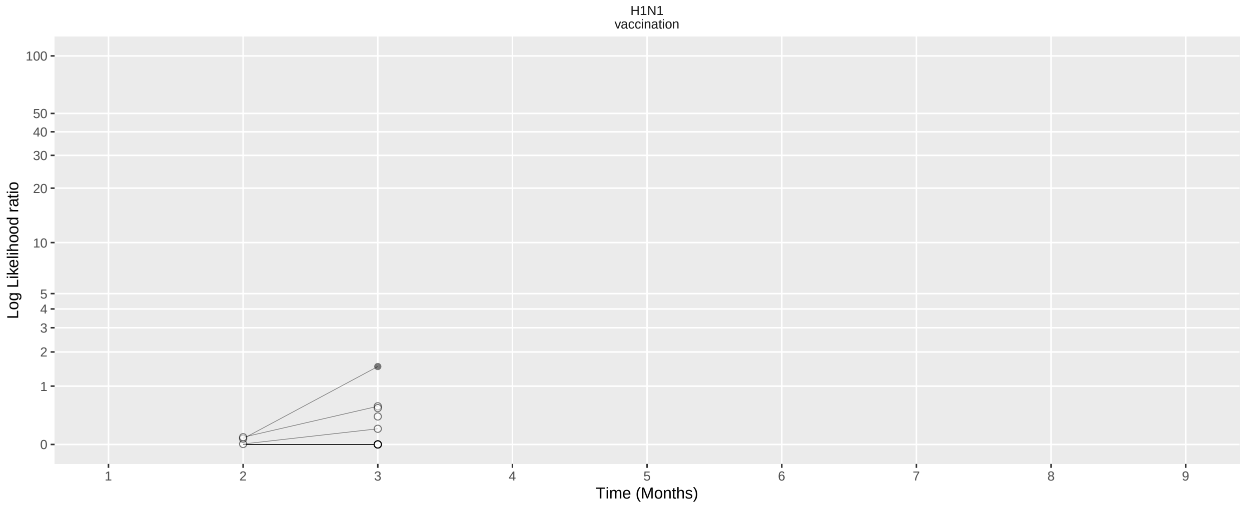

Figure 225: Negative control log likelihood ratios per month using the CohortMethod method (PS matching, using random days as comparator), in the Optum EHR database.

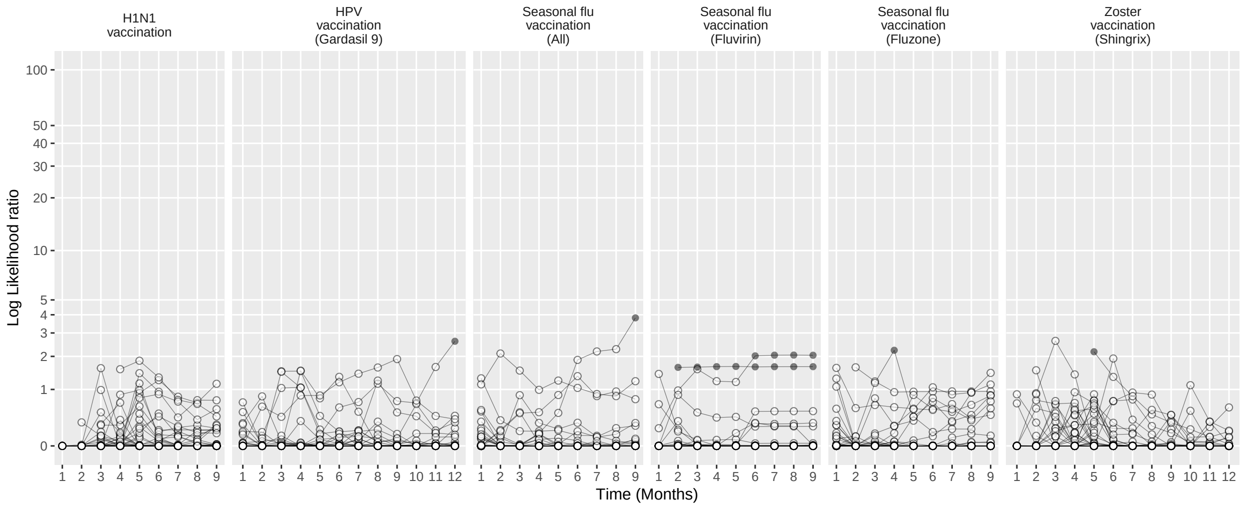

Figure 226: Negative control log likelihood ratios per month using the CohortMethod method (PS stratification, using outpatient visits as comparator), in the Optum EHR database.

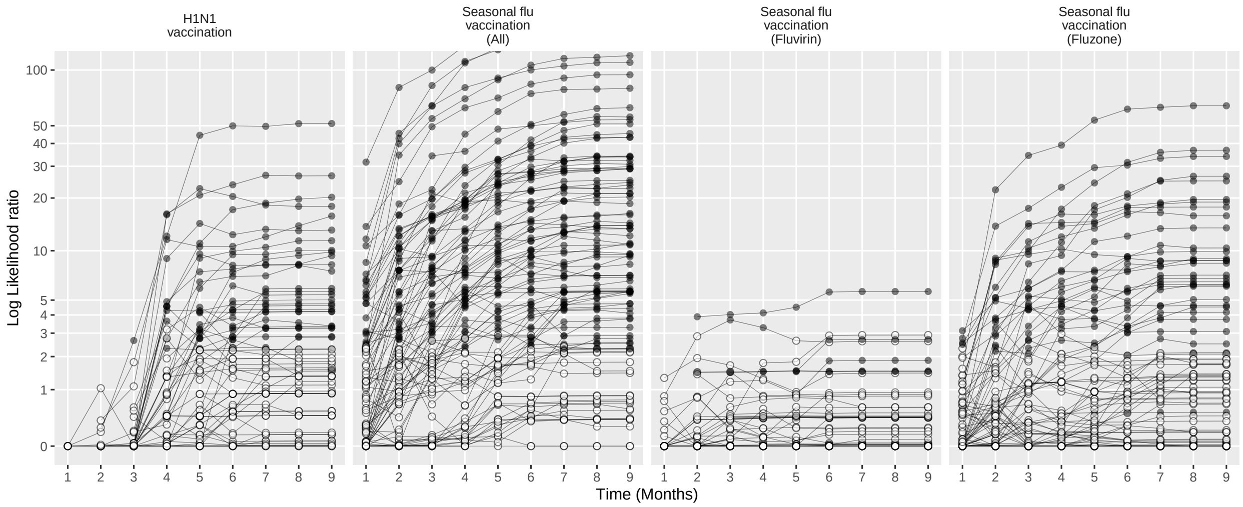

Figure 227: Negative control log likelihood ratios per month using the CohortMethod method (PS stratification, using random days as comparator), in the Optum EHR database.

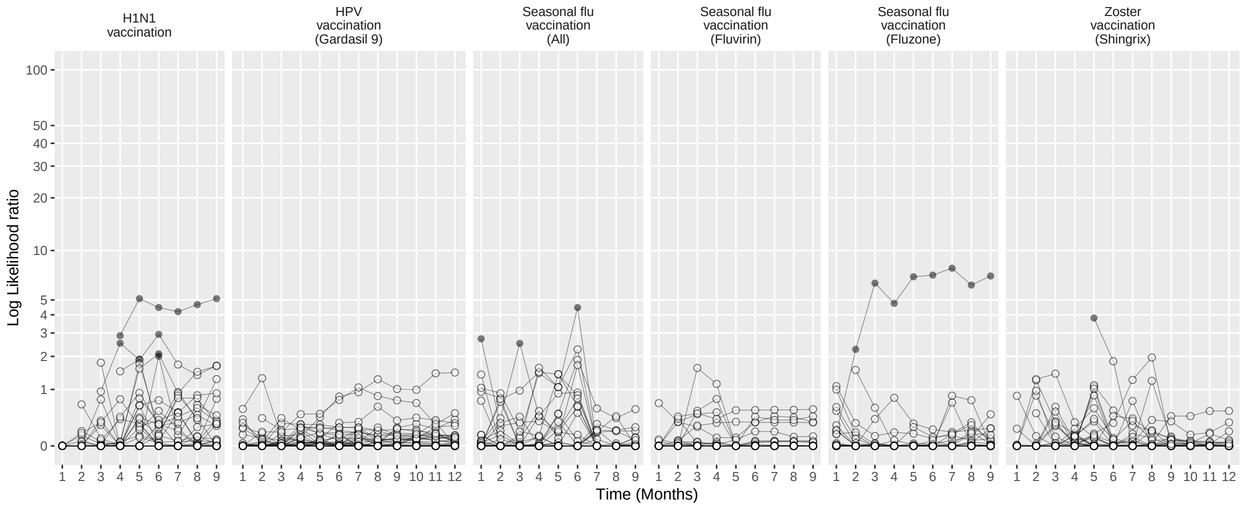

Figure 228: Negative control log likelihood ratios per month using the CohortMethod method (PS weighting, using outpatient visits as comparator), in the Optum EHR database.

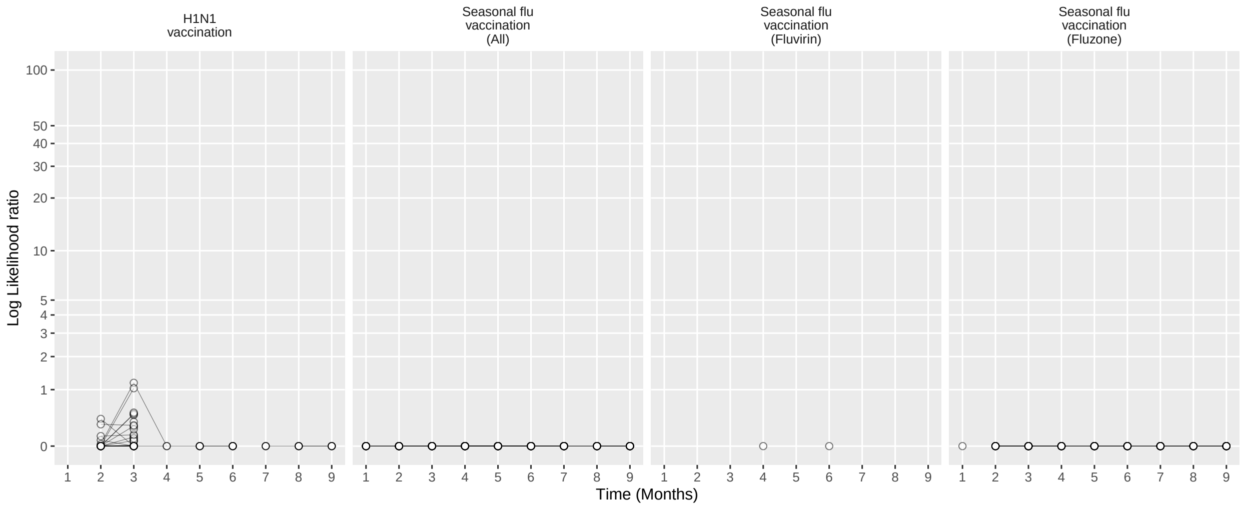

Figure 229: Negative control log likelihood ratios per month using the CohortMethod method (PS weighting, using random days as comparator), in the Optum EHR database.

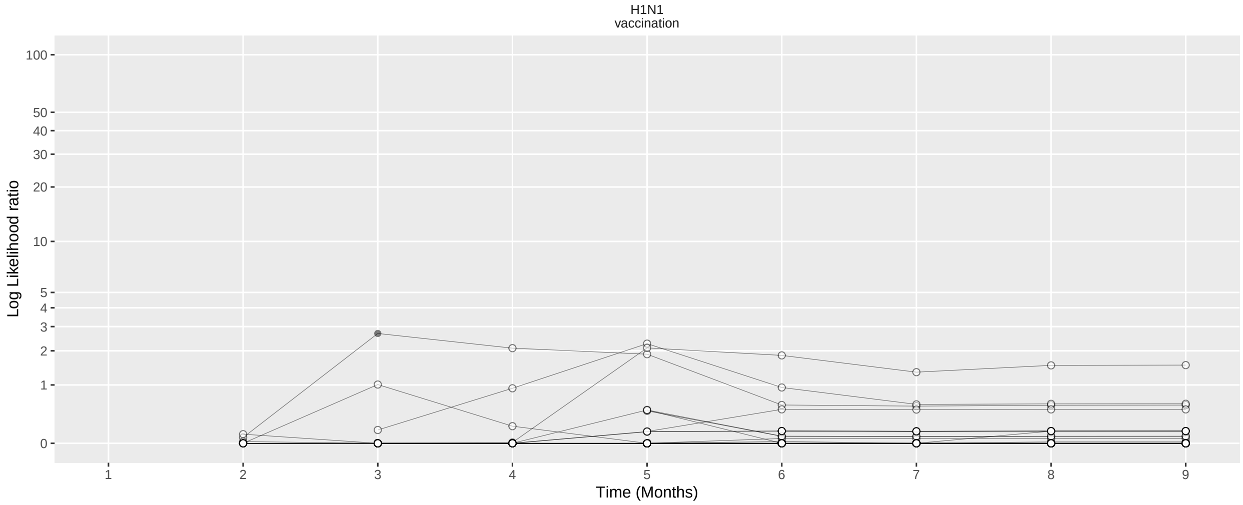

Figure 230: Negative control log likelihood ratios per month using the CohortMethod method (Per-month PS matching, using outpatient visits as comparator), in the Optum EHR database.

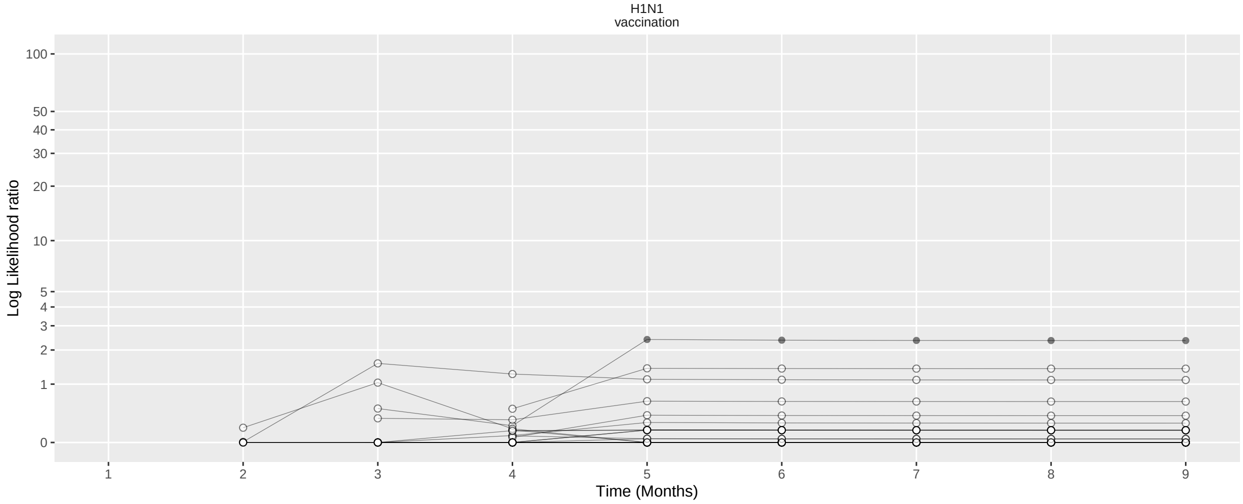

Figure 231: Negative control log likelihood ratios per month using the CohortMethod method (Per-month PS matching, using random days as comparator), in the Optum EHR database.

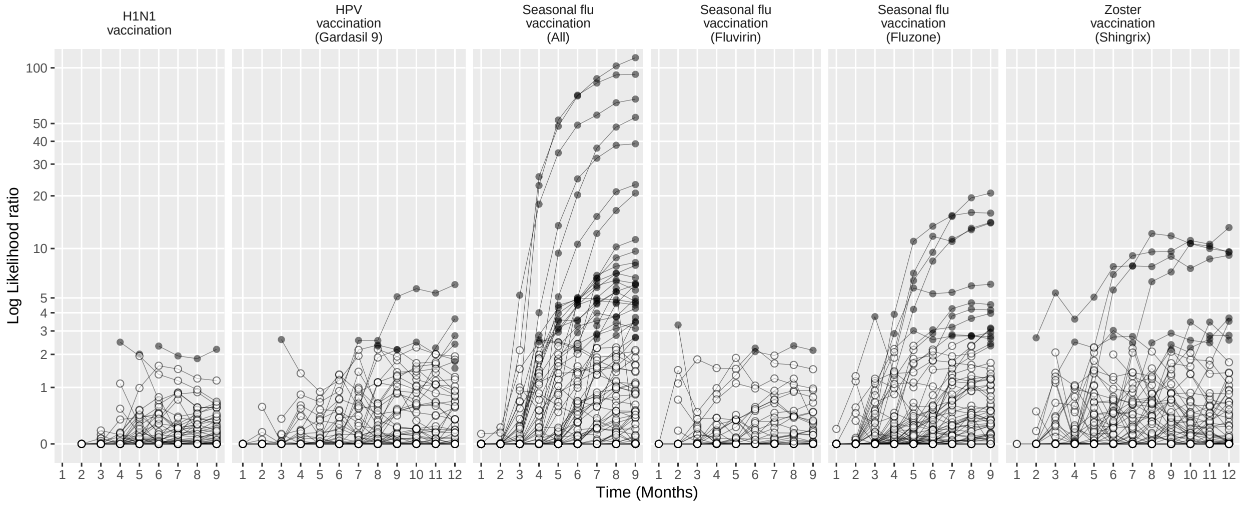

Figure 232: Negative control log likelihood ratios per month using the SCCS method (Unadjusted SCCS excluding pre-vaccination window), in the Optum EHR database.

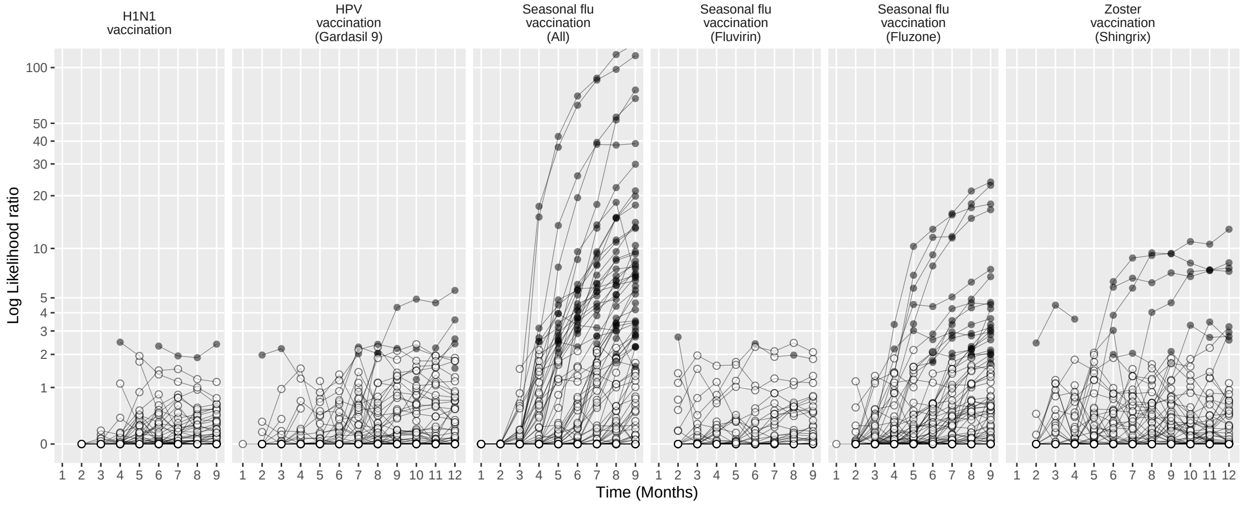

Figure 233: Negative control log likelihood ratios per month using the SCCS method (Age & season adjusted SCCS excluding pre-vaccination window), in the Optum EHR database.

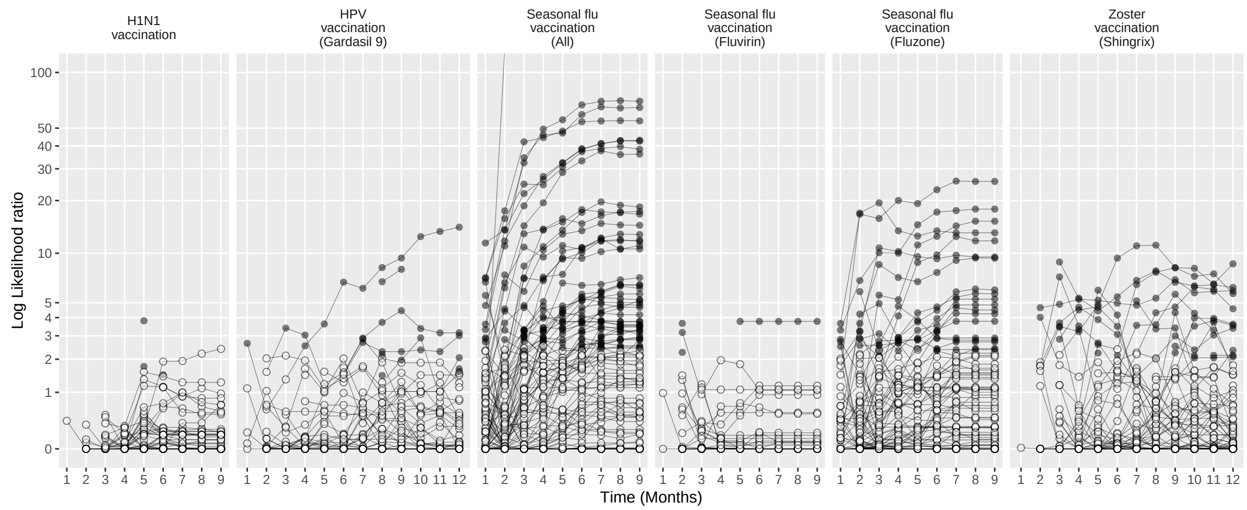

Figure 234: Negative control log likelihood ratios per month using the SCCS method (SCRI with prior control interval), in the Optum EHR database.

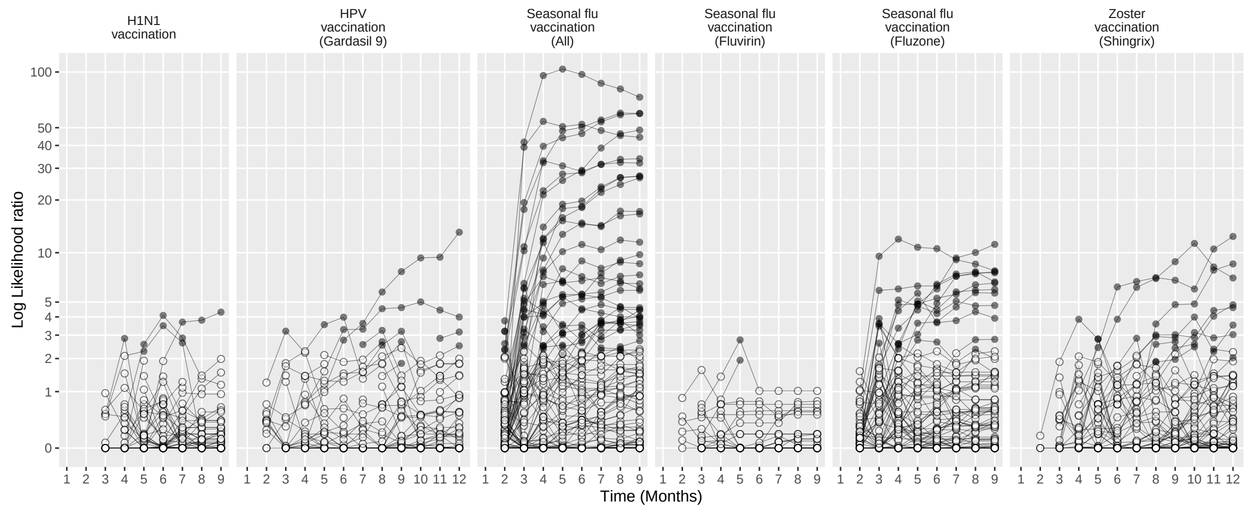

Figure 235: Negative control log likelihood ratios per month using the SCCS method (SCRI with posterior control interval), in the Optum EHR database.

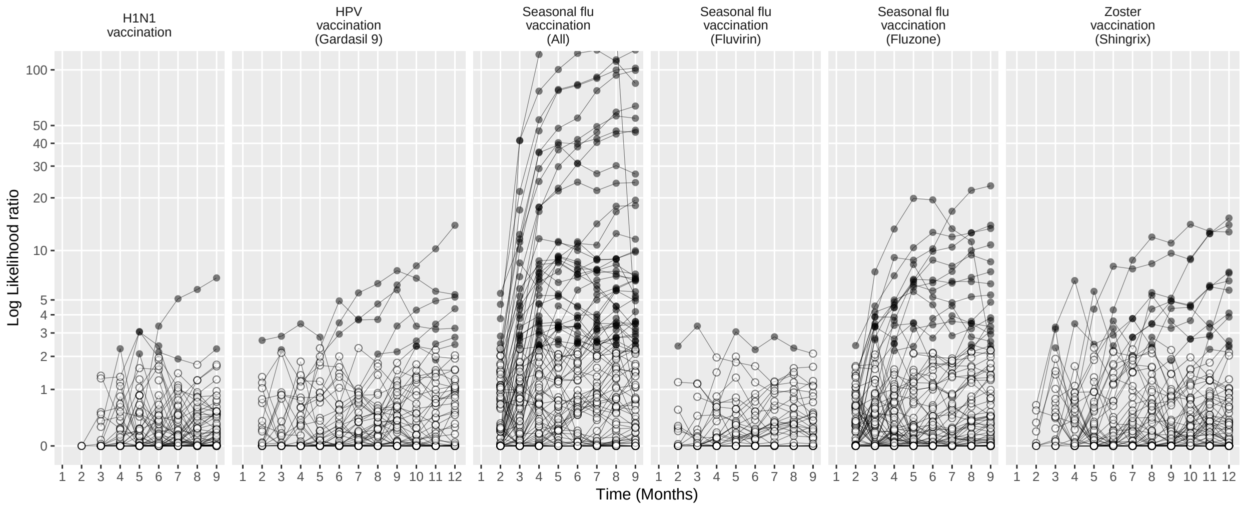

Figure 236: Negative control log likelihood ratios per month using the SCCS method (Unadjusted SCCS excluding all pre-vaccination time), in the Optum EHR database.

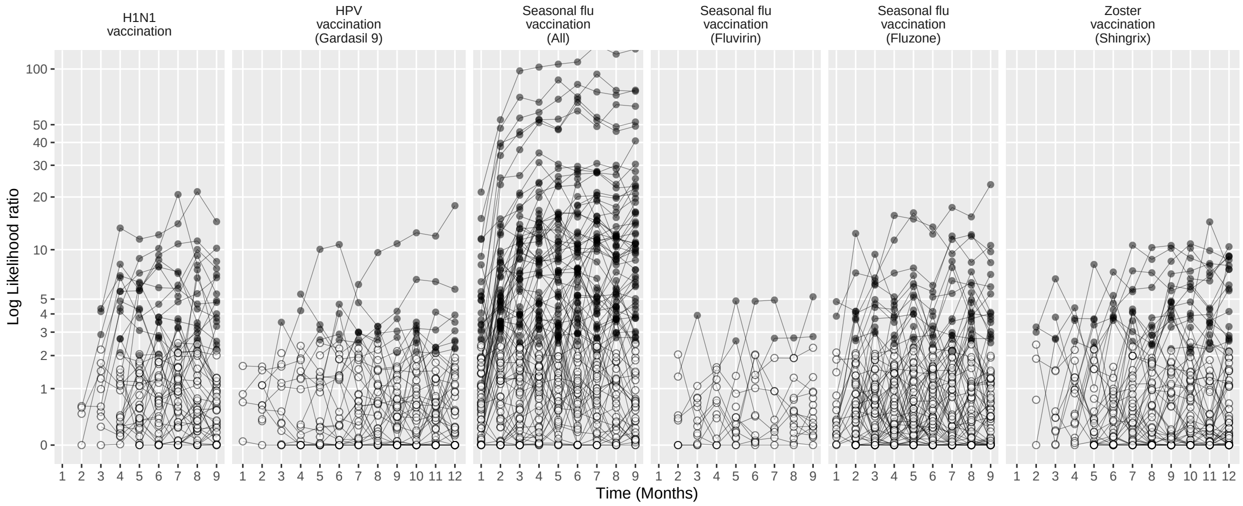

Figure 237: Negative control log likelihood ratios per month using the CaseControl method (Age & sex adjusted, using random controls), in the Optum EHR database.

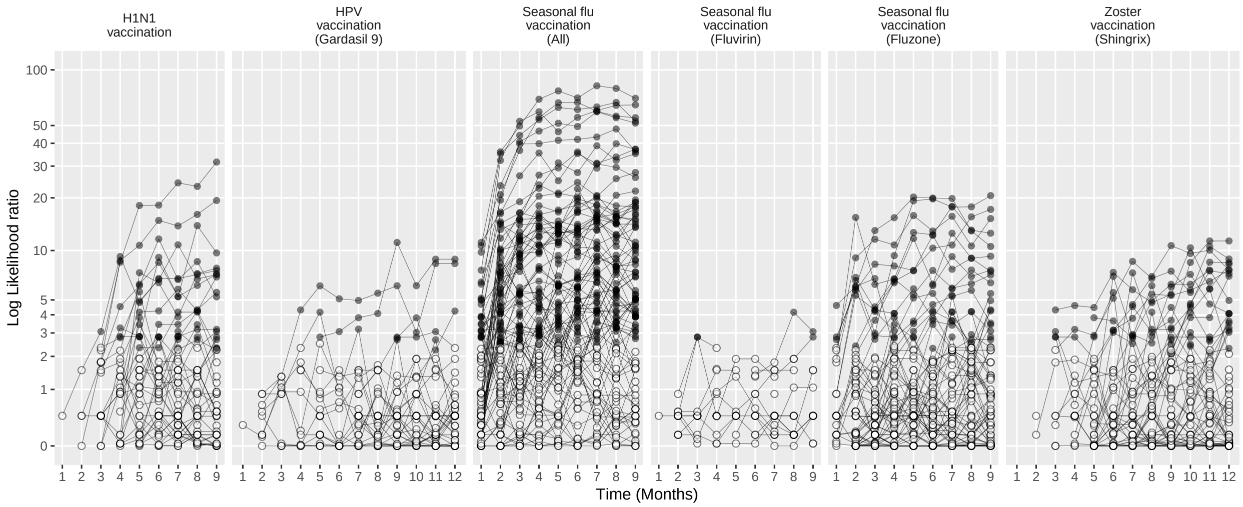

Figure 238: Negative control log likelihood ratios per month using the CaseControl method (Age & sex matched controls), in the Optum EHR database.

#### 15.2 Negative controls log likelihood ratios in MDCD

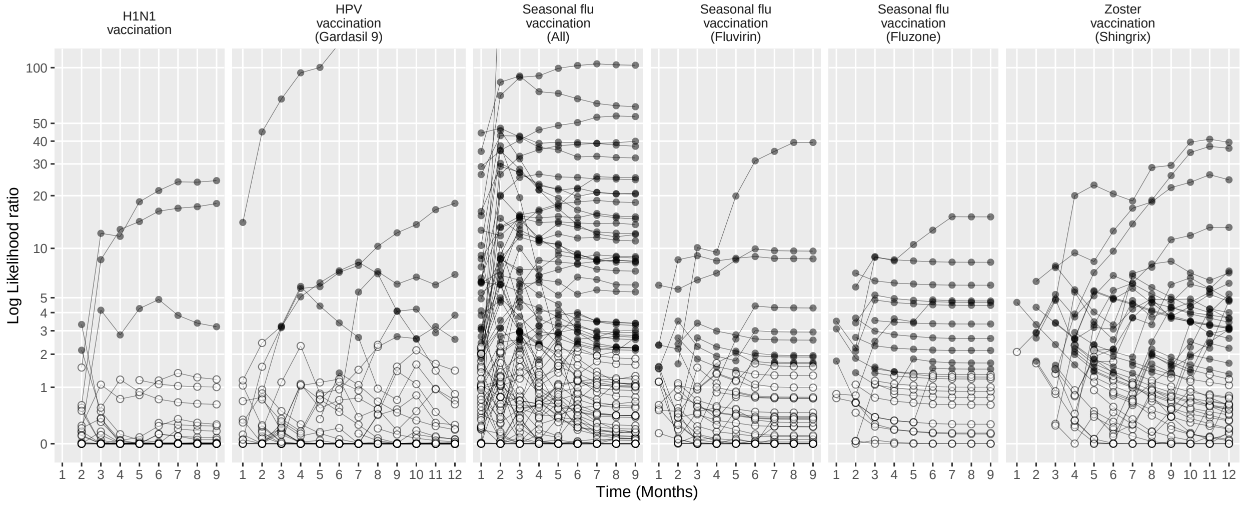

Figure 239: Negative control log likelihood ratios per month using the HistoricalComparator method (Unadjusted, using entire historic period), in the MDCD database.

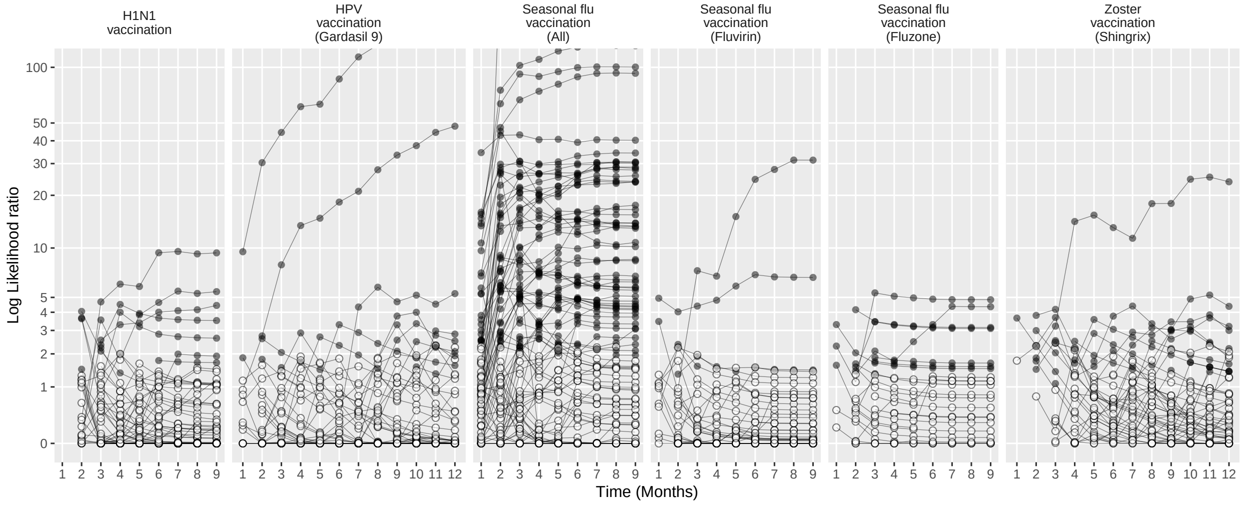

Figure 240: Negative control log likelihood ratios per month using the HistoricalComparator method (Age & sex adjusted, using entire historic period), in the MDCD database.

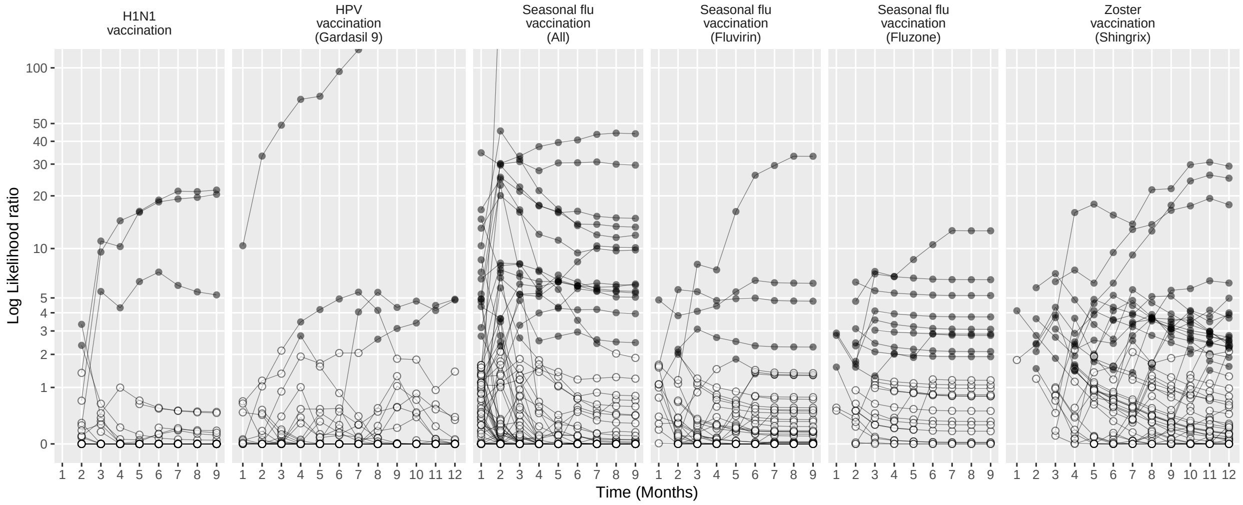

Figure 241: Negative control log likelihood ratios per month using the HistoricalComparator method (Unadjusted, using TaR after historic visit), in the MDCD database.

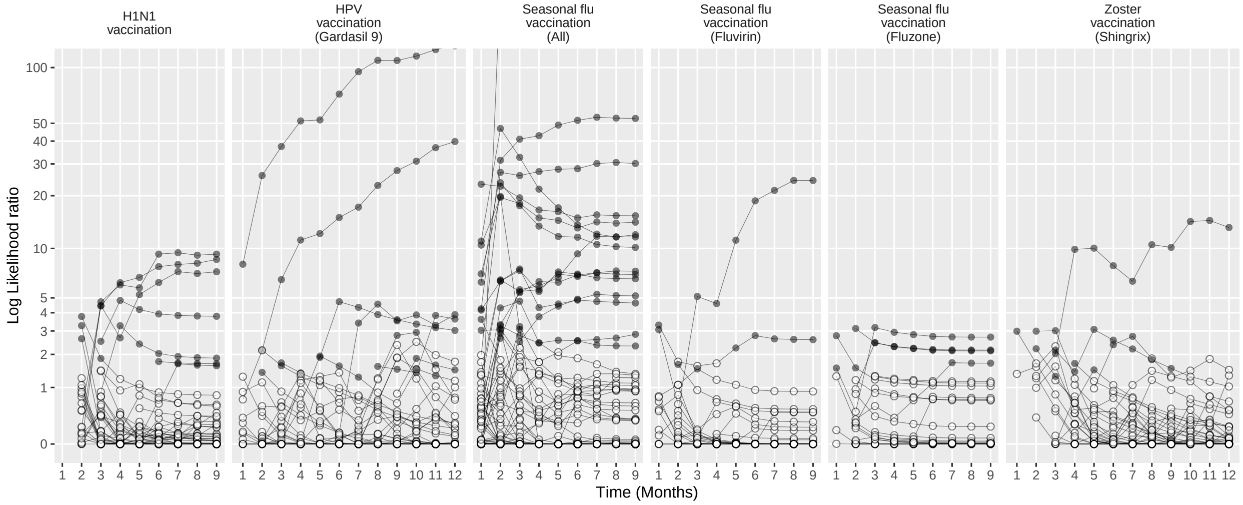

Figure 242: Negative control log likelihood ratios per month using the HistoricalComparator method (Age & sex adjusted, using TaR after historic visit), in the MDCD database.

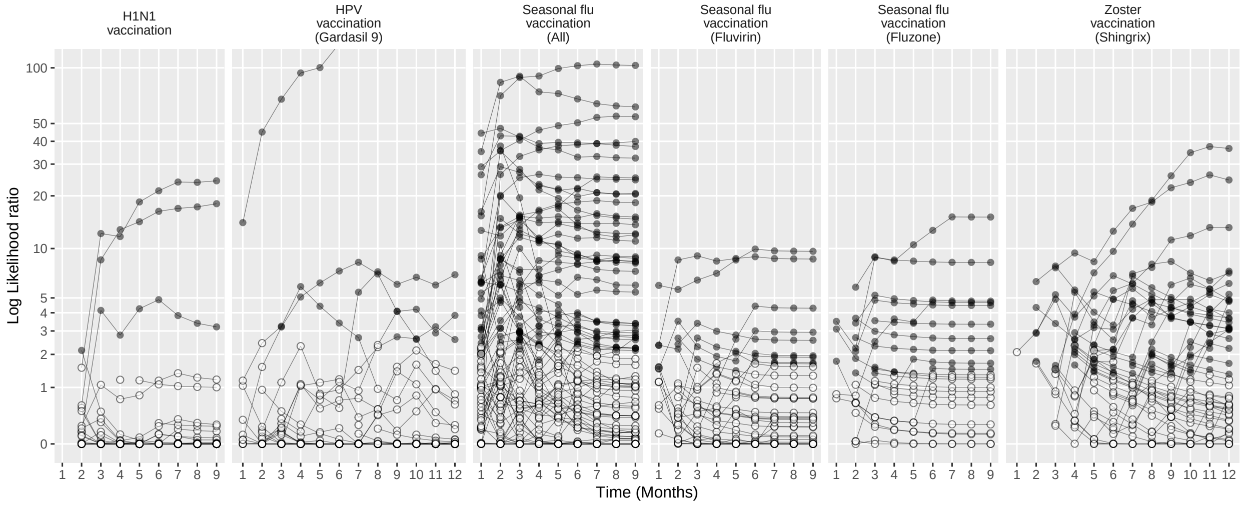

Figure 243: Negative control log likelihood ratios per month using the HistoricalComparator method (Unadjusted, using entire historic period, filtered), in the MDCD database.

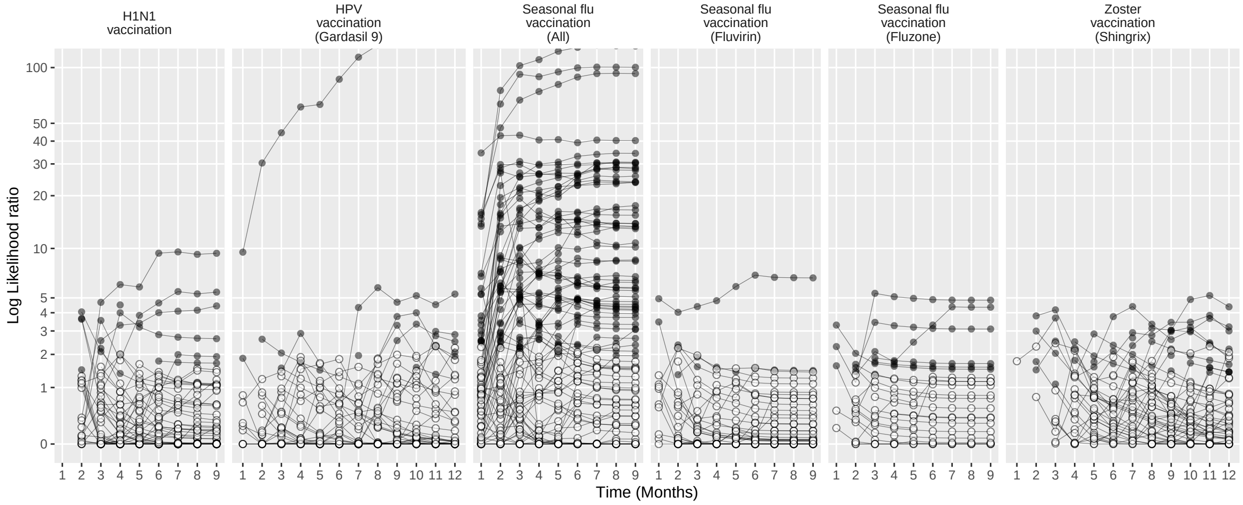

Figure 244: Negative control log likelihood ratios per month using the HistoricalComparator method (Age & sex adjusted, using entire historic period, filtered), in the MDCD database.

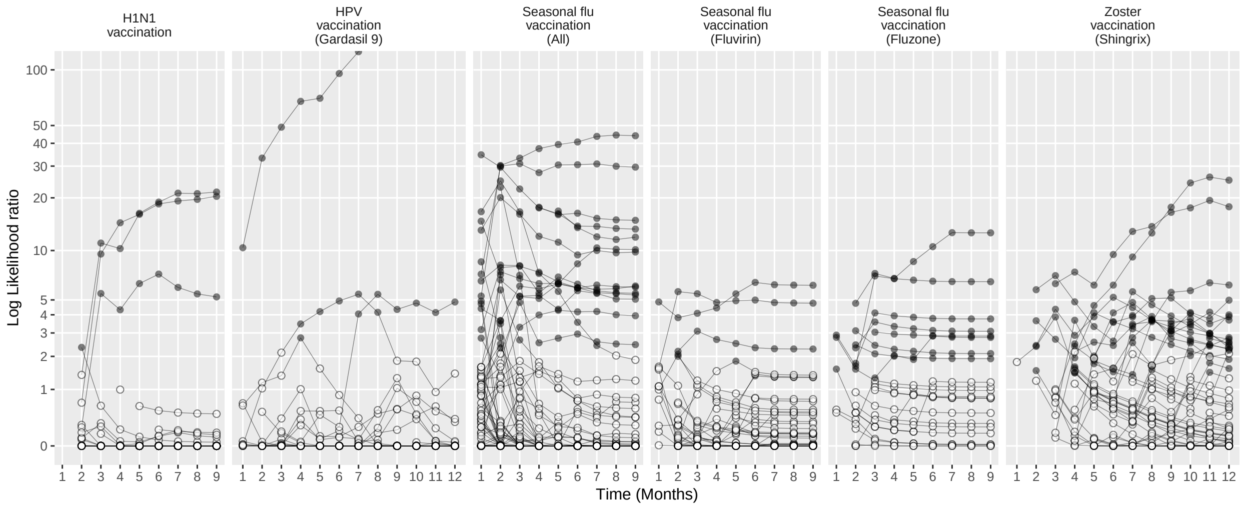

Figure 245: Negative control log likelihood ratios per month using the HistoricalComparator method (Unadjusted, using TaR after historic visit, filtered), in the MDCD database.

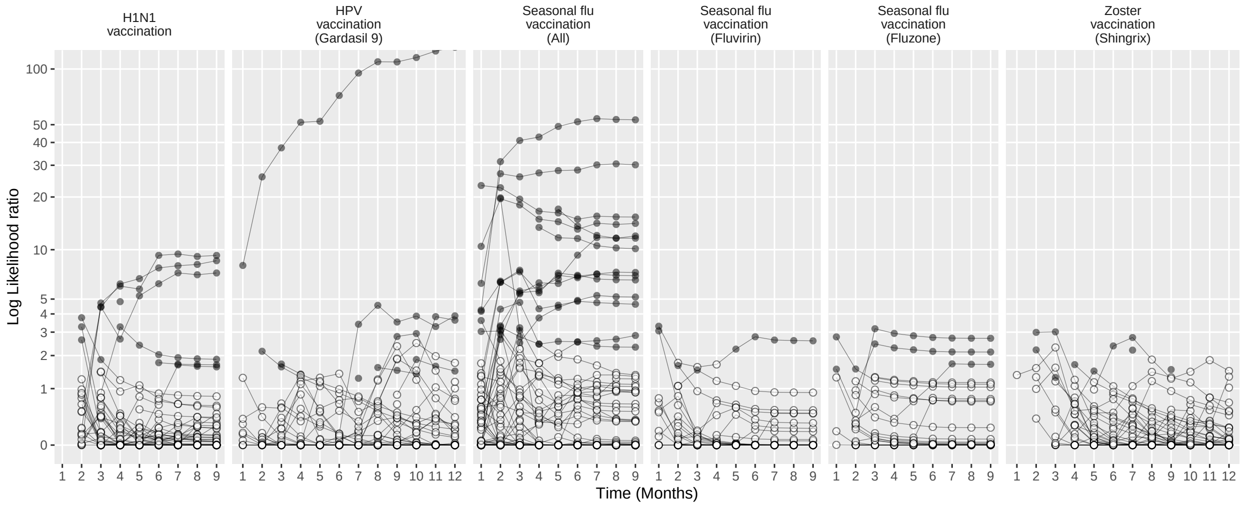

Figure 246: Negative control log likelihood ratios per month using the HistoricalComparator method (Age & sex adjusted, using TaR after historic visit, filtered), in the MDCD database.

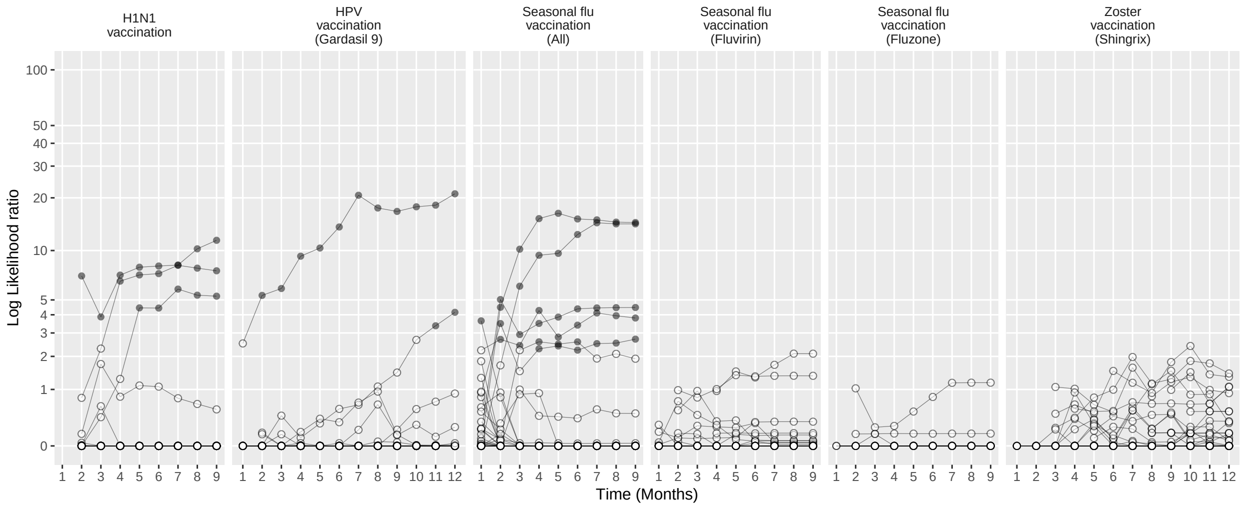

Figure 247: Negative control log likelihood ratios per month using the CohortMethod method (Unadjusted, using outpatient visits as comparator), in the MDCD database.

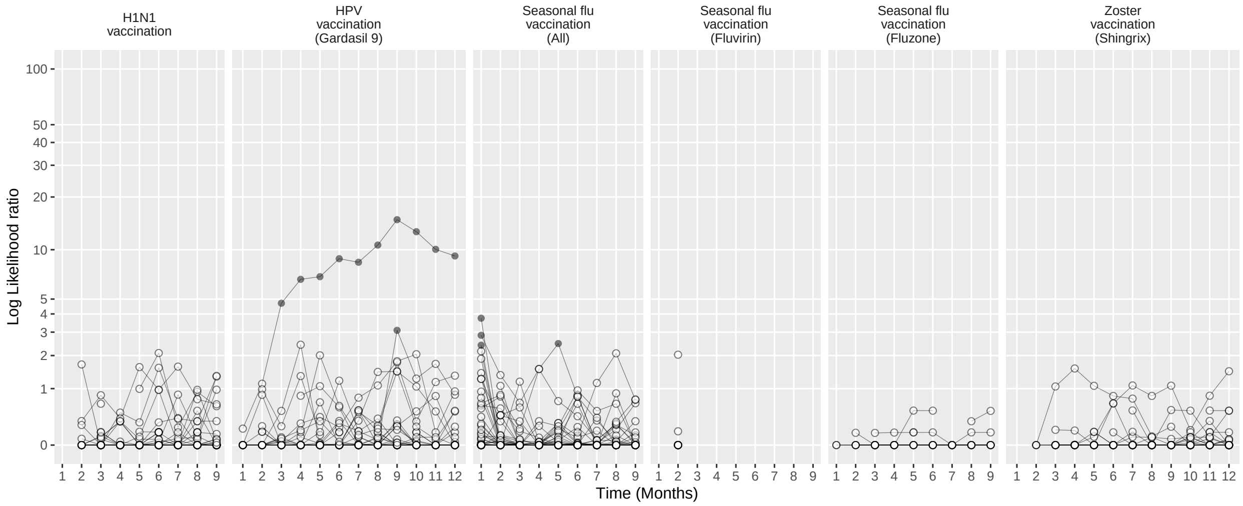

Figure 248: Negative control log likelihood ratios per month using the CohortMethod method (PS matching, using outpatient visits as comparator), in the MDCD database.

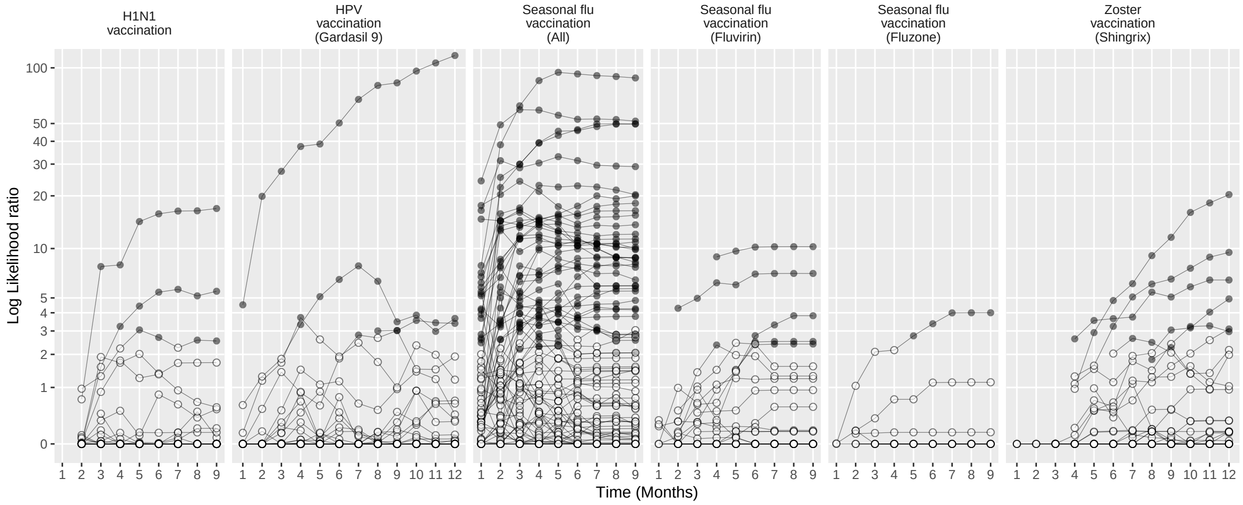

Figure 249: Negative control log likelihood ratios per month using the CohortMethod method (Unadjusted, using random days as comparator), in the MDCD database.

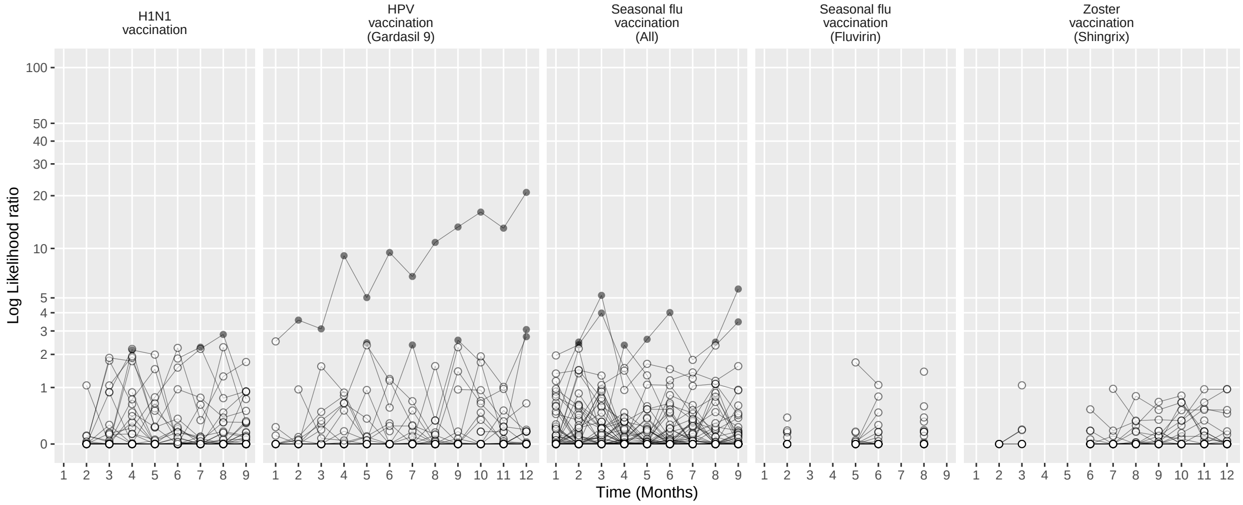

Figure 250: Negative control log likelihood ratios per month using the CohortMethod method (PS matching, using random days as comparator), in the MDCD database.

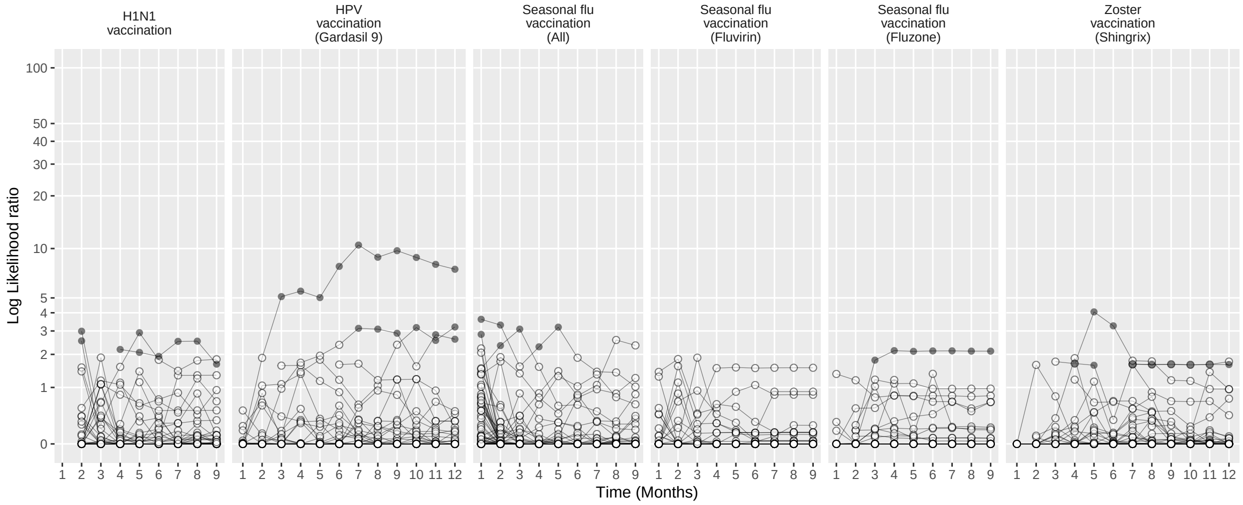

Figure 251: Negative control log likelihood ratios per month using the CohortMethod method (PS stratification, using outpatient visits as comparator), in the MDCD database.

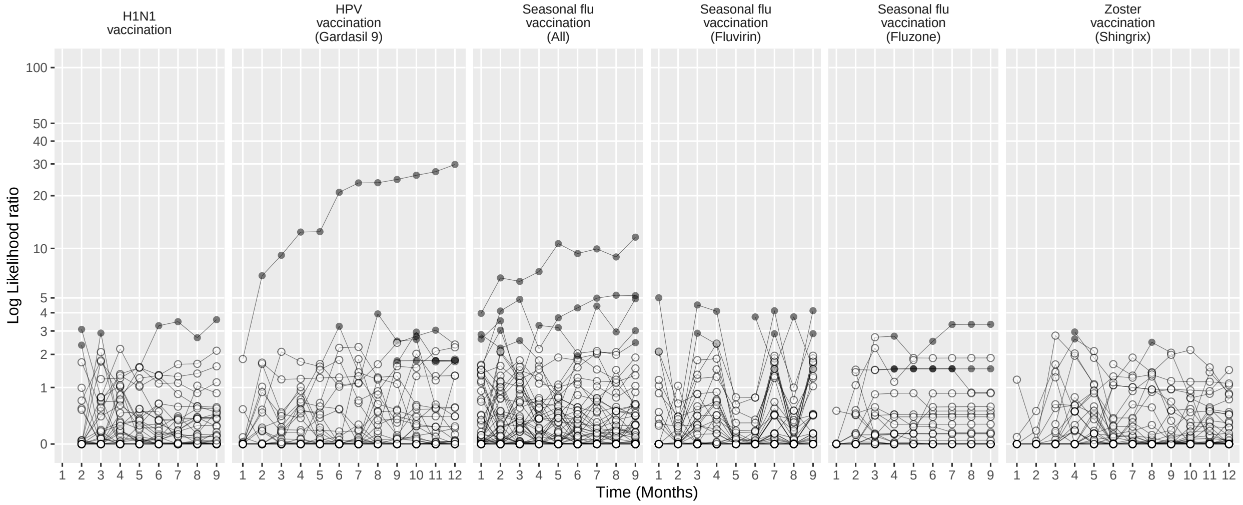

Figure 252: Negative control log likelihood ratios per month using the CohortMethod method (PS stratification, using random days as comparator), in the MDCD database.

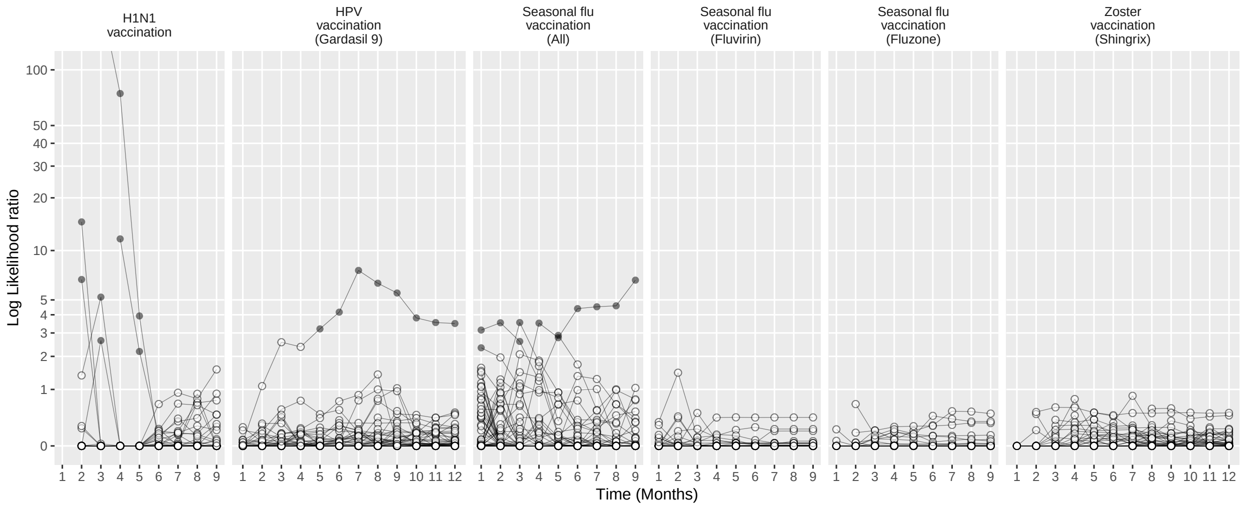

Figure 253: Negative control log likelihood ratios per month using the CohortMethod method (PS weighting, using outpatient visits as comparator), in the MDCD database.

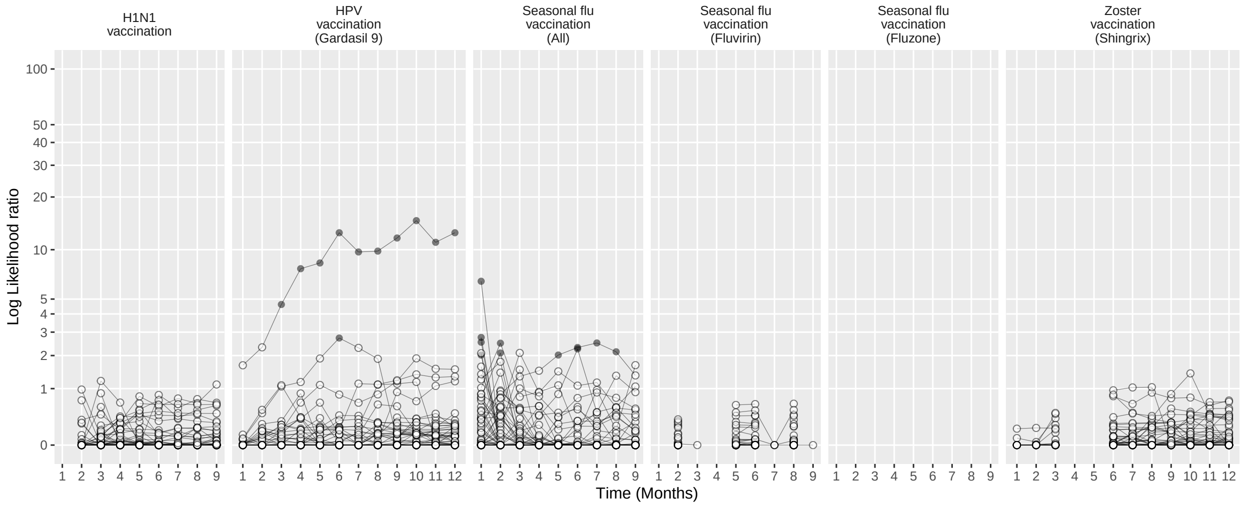

Figure 254: Negative control log likelihood ratios per month using the CohortMethod method (PS weighting, using random days as comparator), in the MDCD database.

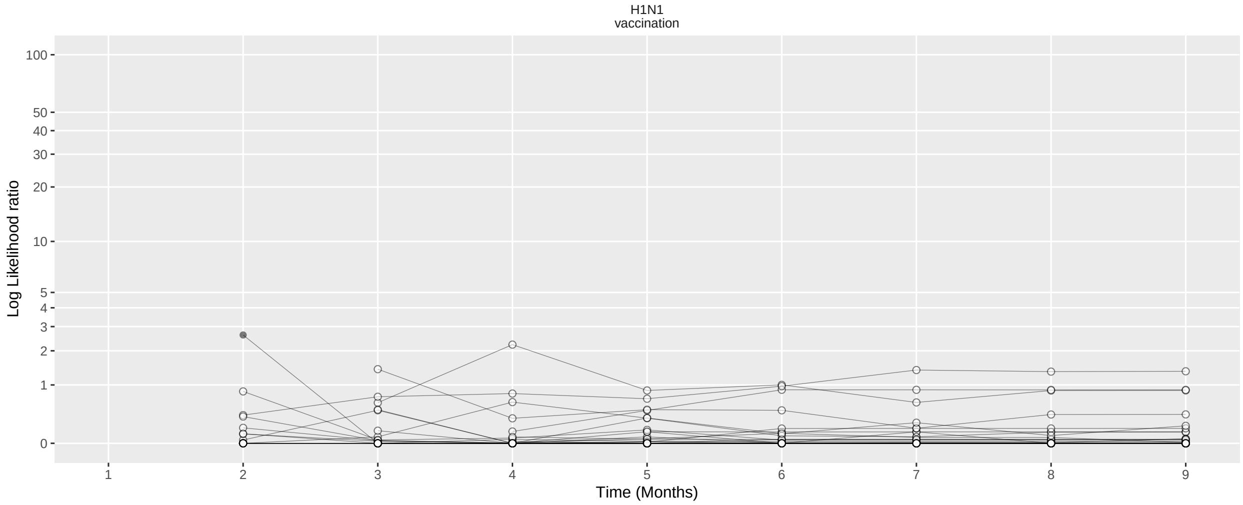

Figure 255: Negative control log likelihood ratios per month using the CohortMethod method (Per-month PS matching, using outpatient visits as comparator), in the MDCD database.

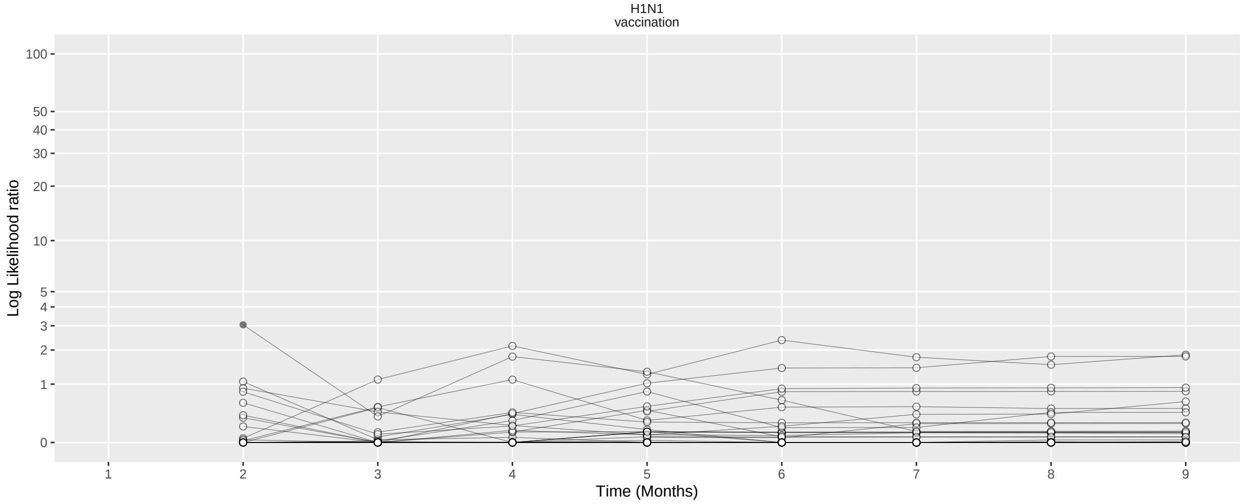

Figure 256: Negative control log likelihood ratios per month using the CohortMethod method (Per-month PS matching, using random days as comparator), in the MDCD database.

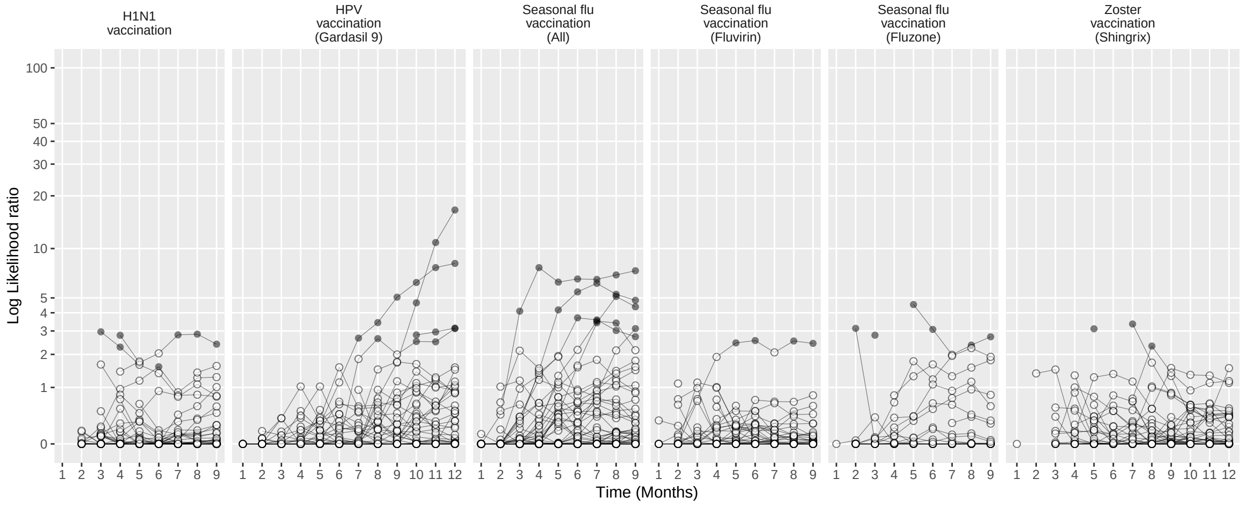

Figure 257: Negative control log likelihood ratios per month using the SCCS method (Unadjusted SCCS excluding pre-vaccination window), in the MDCD database.

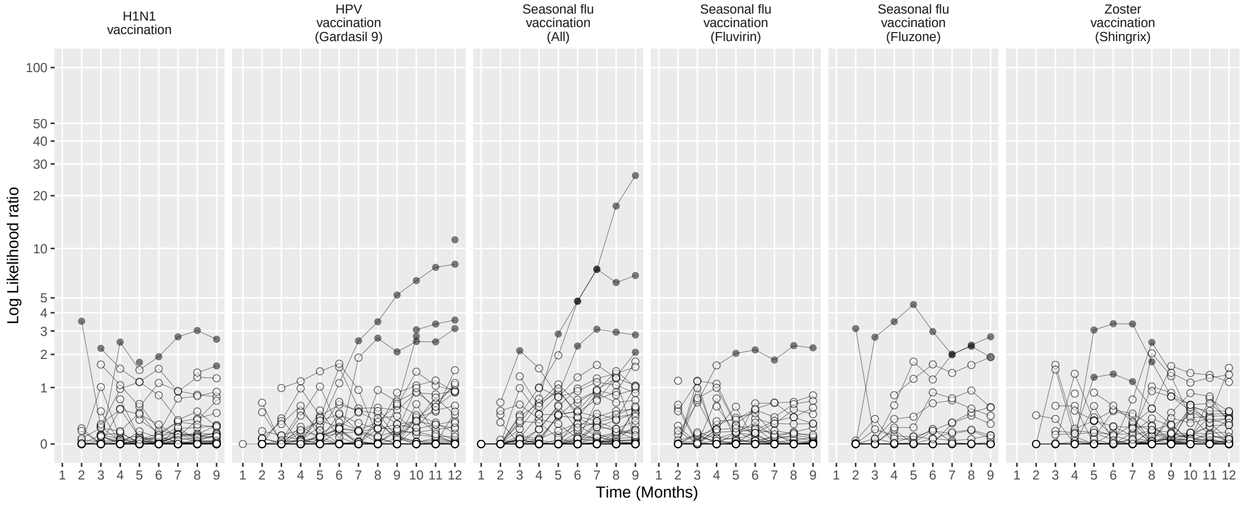

Figure 258: Negative control log likelihood ratios per month using the SCCS method (Age & season adjusted SCCS excluding pre-vaccination window), in the MDCD database.

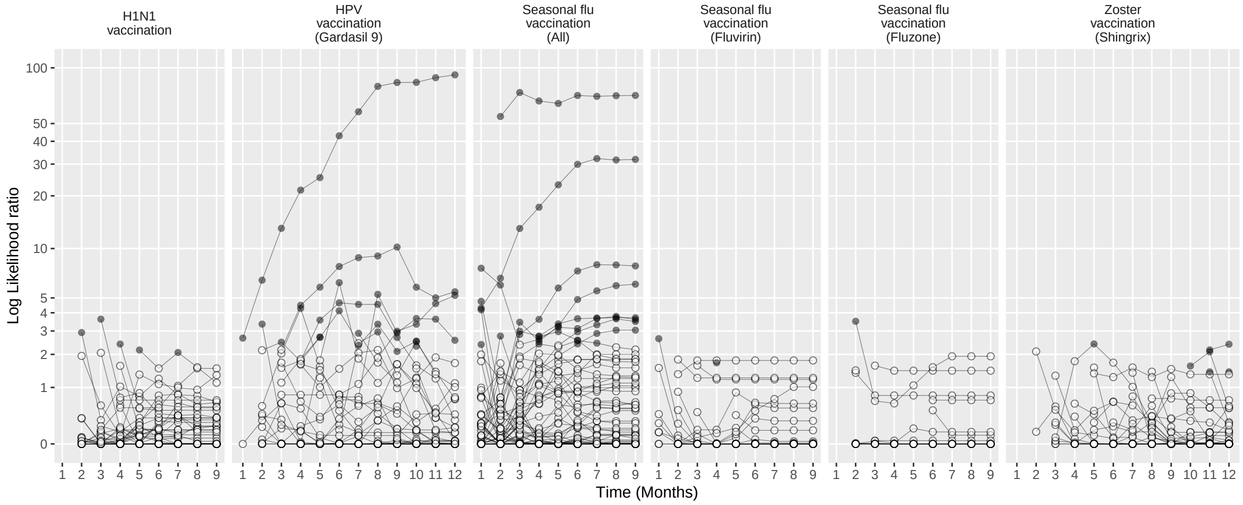

Figure 259: Negative control log likelihood ratios per month using the SCCS method (SCRI with prior control interval), in the MDCD database.

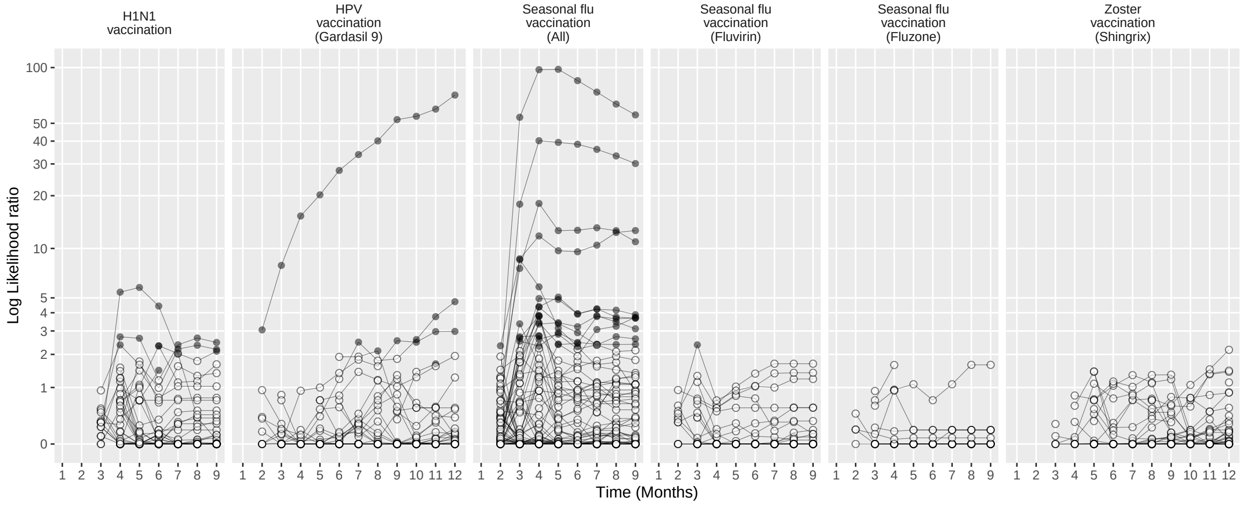

Figure 260: Negative control log likelihood ratios per month using the SCCS method (SCRI with posterior control interval), in the MDCD database.

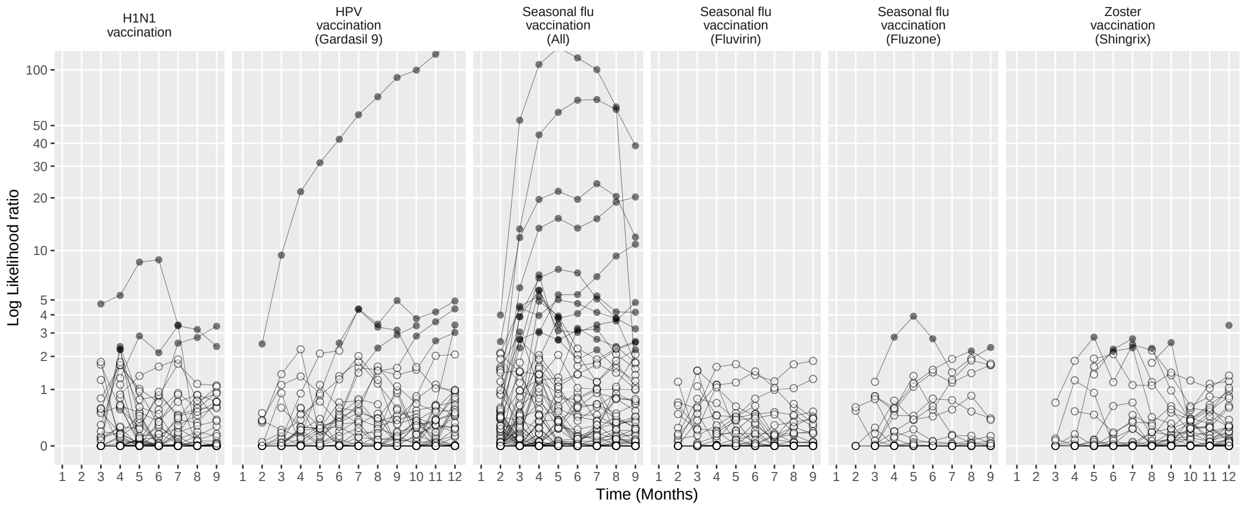

Figure 261: Negative control log likelihood ratios per month using the SCCS method (Unadjusted SCCS excluding all pre-vaccination time), in the MDCD database.

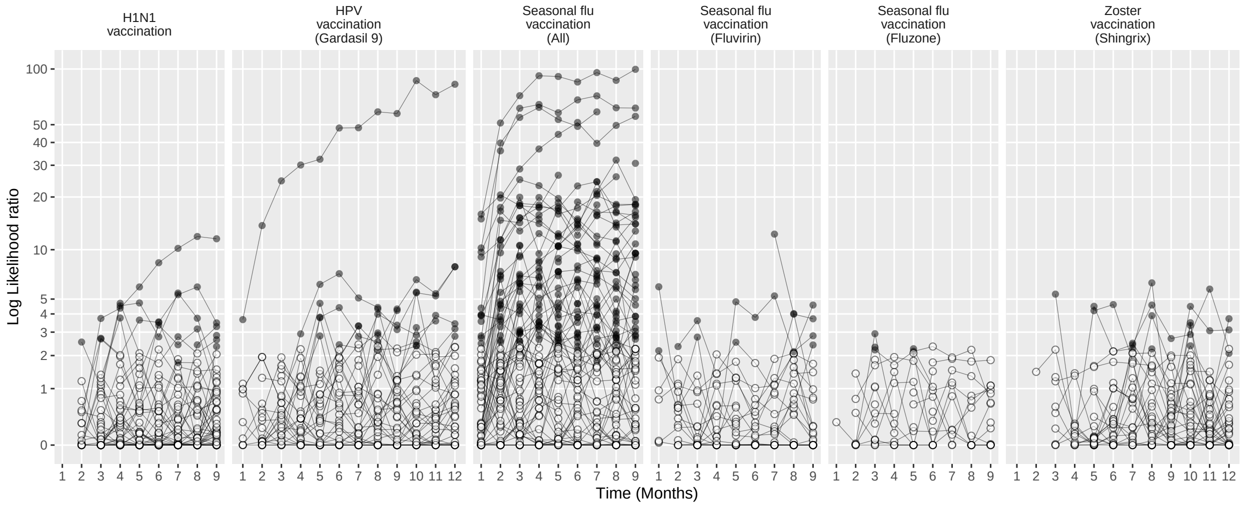

Figure 262: Negative control log likelihood ratios per month using the CaseControl method (Age & sex adjusted, using random controls), in the MDCD database.

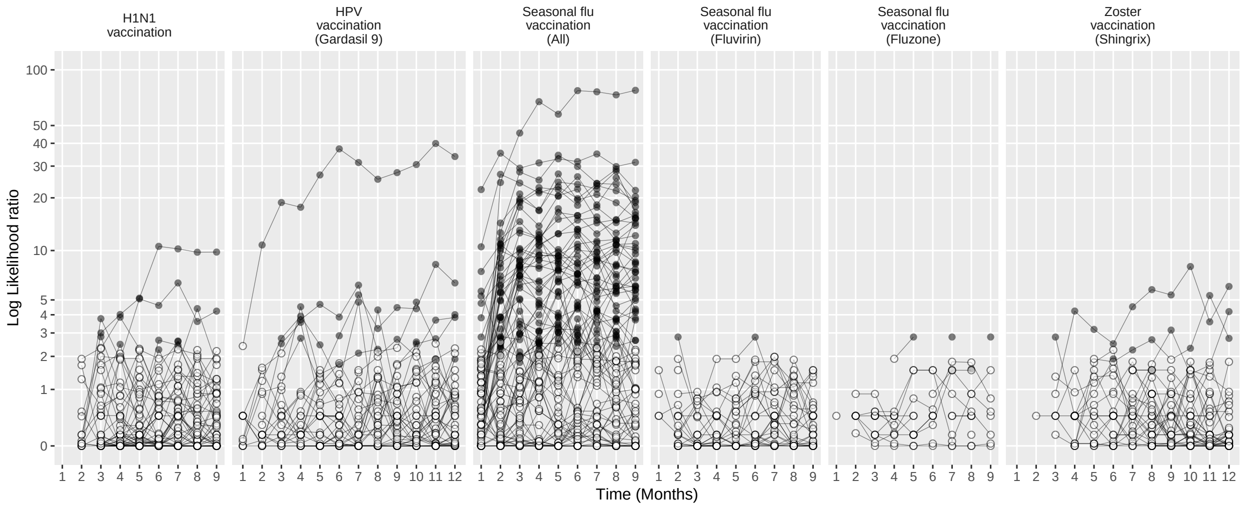

Figure 263: Negative control log likelihood ratios per month using the CaseControl method (Age & sex matched controls), in the MDCD database.

#### 15.3 Negative controls log likelihood ratios in MDCR

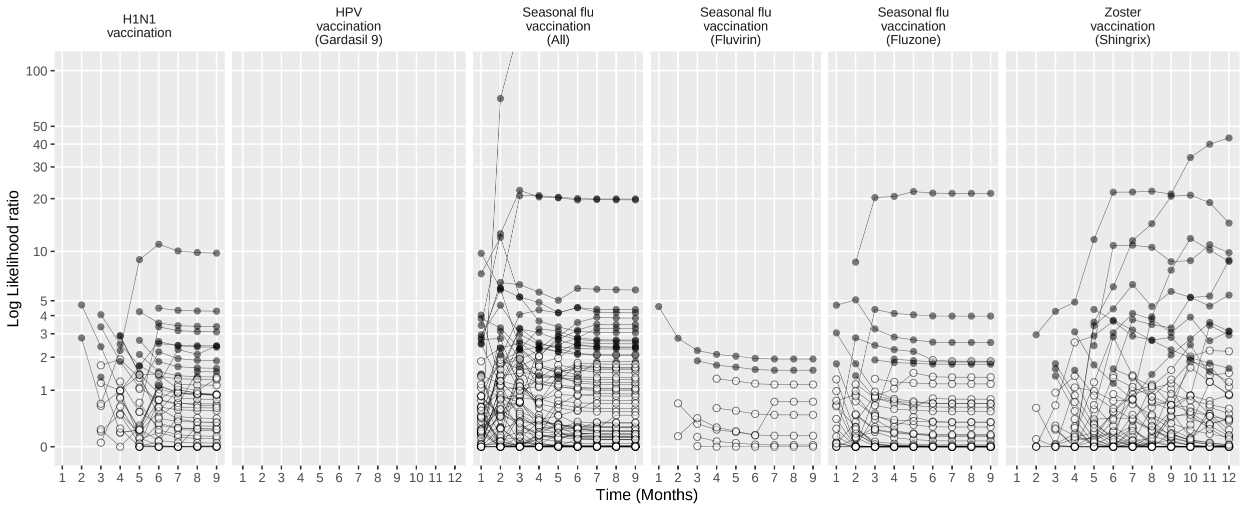

Figure 264: Negative control log likelihood ratios per month using the HistoricalComparator method (Unadjusted, using entire historic period), in the MDCR database.

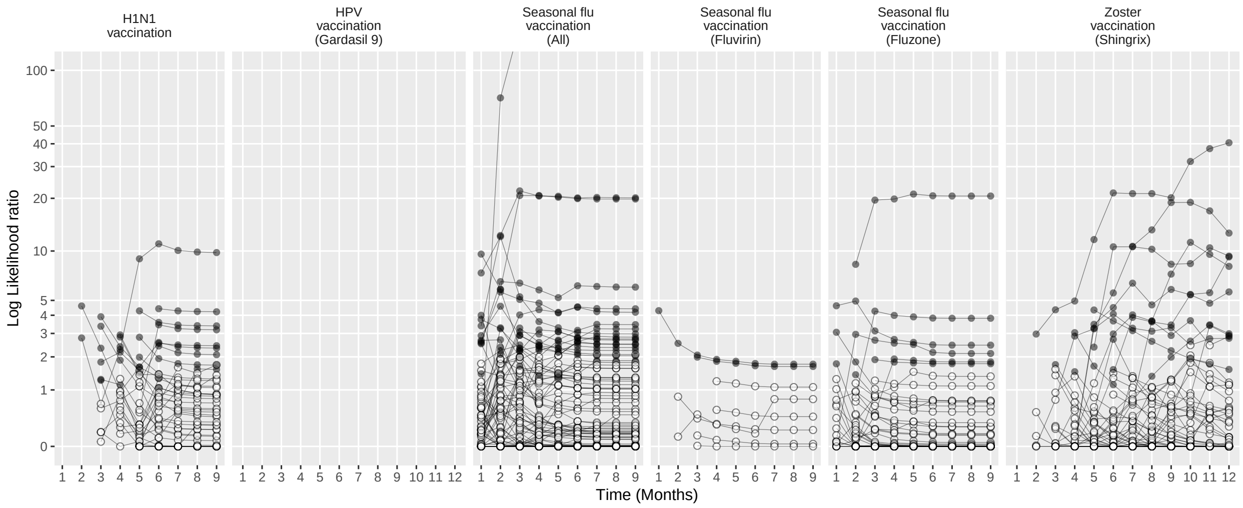

Figure 265: Negative control log likelihood ratios per month using the HistoricalComparator method (Age & sex adjusted, using entire historic period), in the MDCR database.

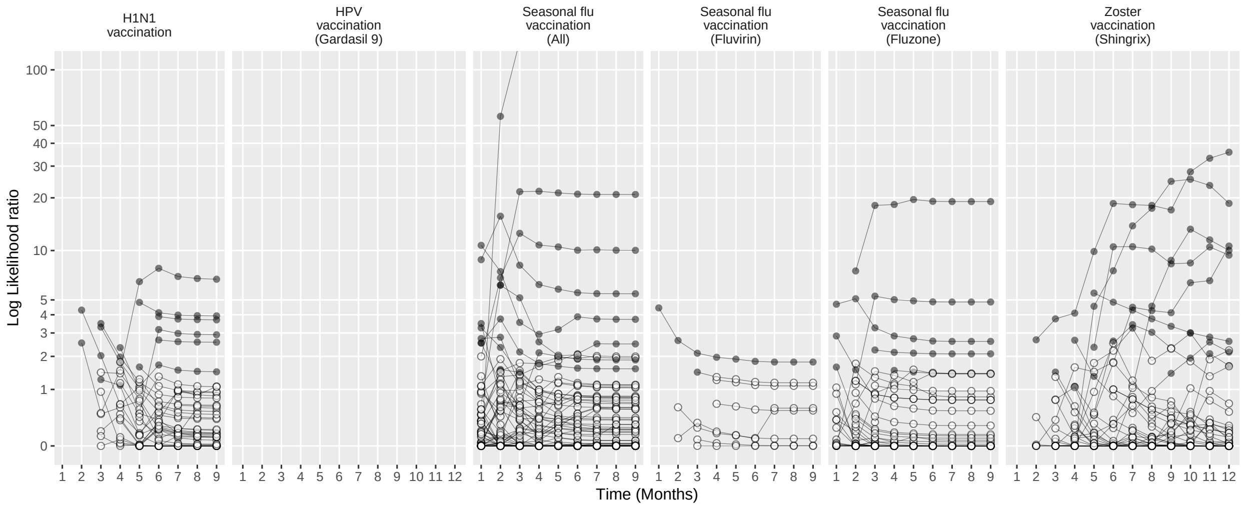

Figure 266: Negative control log likelihood ratios per month using the HistoricalComparator method (Unadjusted, using TaR after historic visit), in the MDCR database.

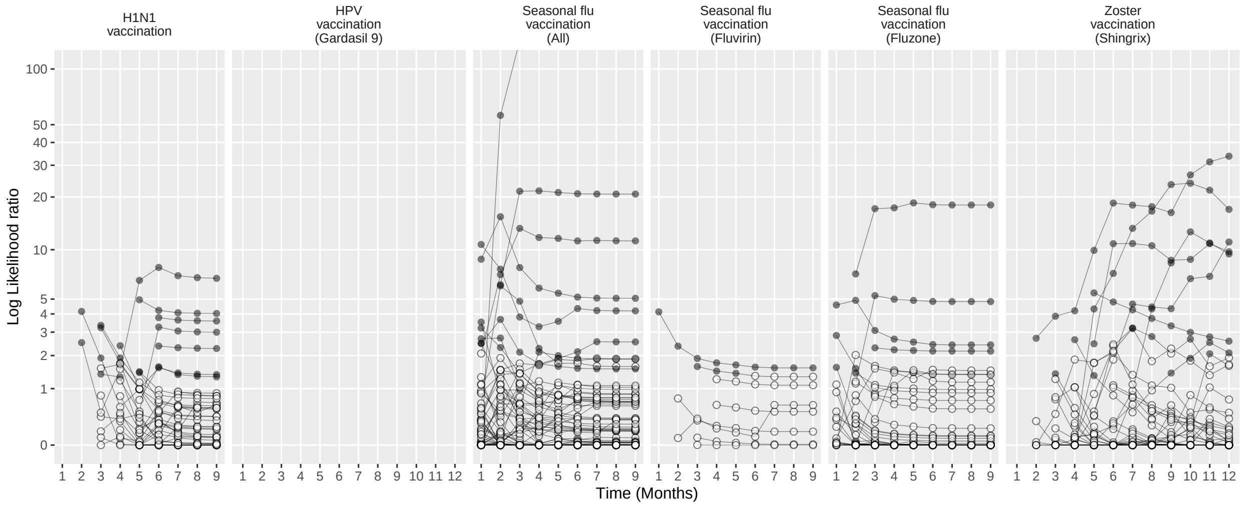

Figure 267: Negative control log likelihood ratios per month using the HistoricalComparator method (Age & sex adjusted, using TaR after historic visit), in the MDCR database.

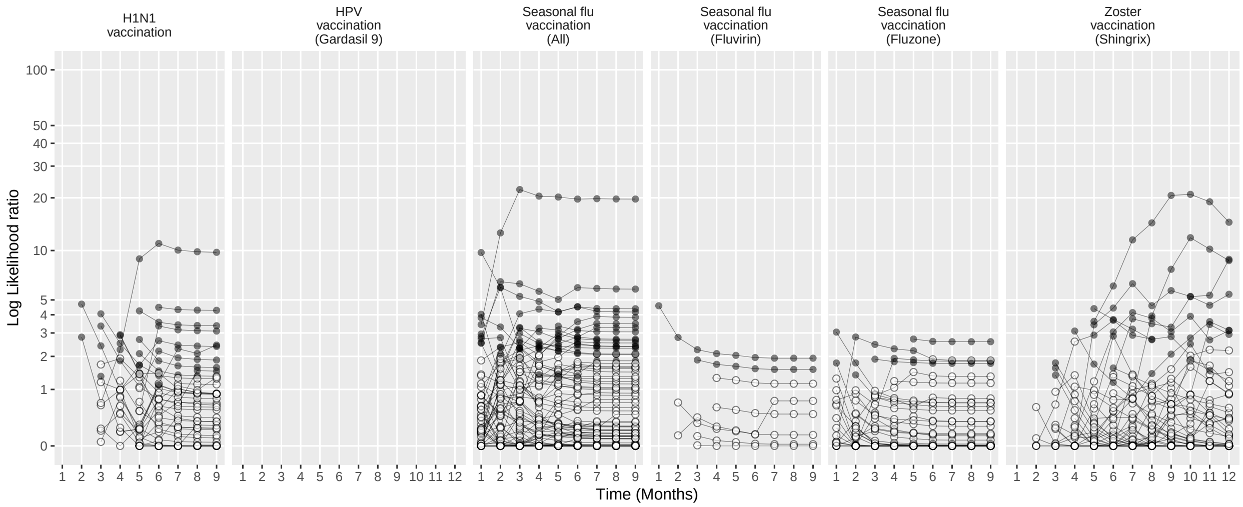

Figure 268: Negative control log likelihood ratios per month using the HistoricalComparator method (Unadjusted, using entire historic period, filtered), in the MDCR database.

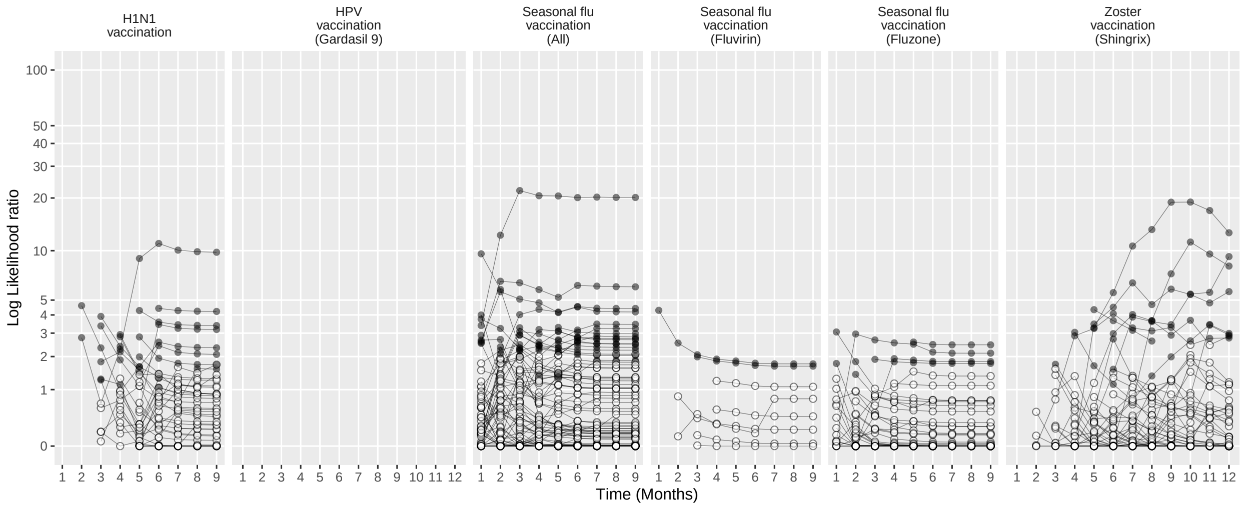

Figure 269: Negative control log likelihood ratios per month using the HistoricalComparator method (Age & sex adjusted, using entire historic period, filtered), in the MDCR database.

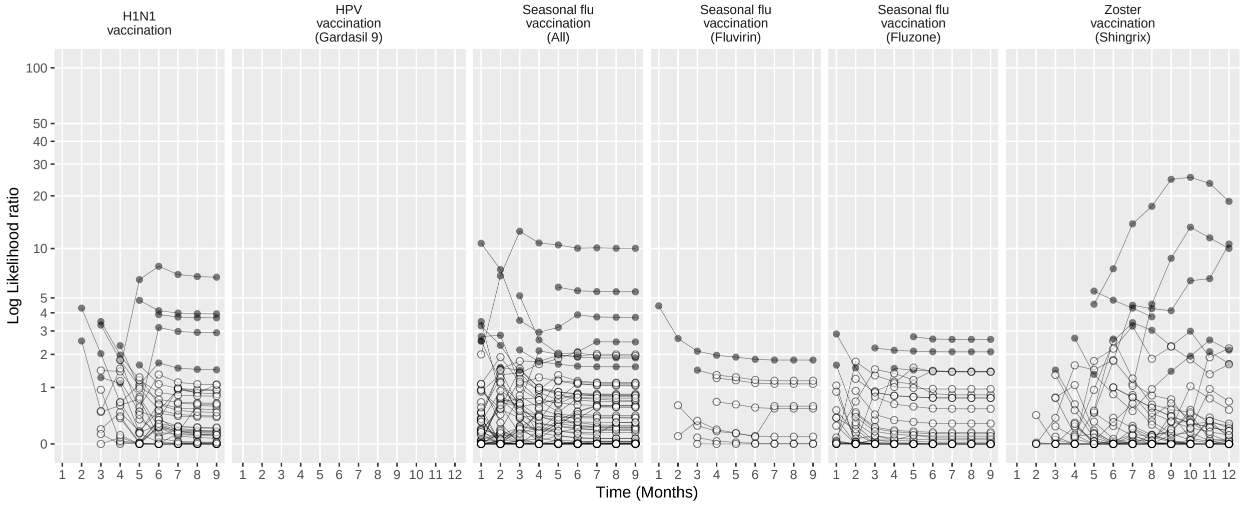

Figure 270: Negative control log likelihood ratios per month using the HistoricalComparator method (Unadjusted, using TaR after historic visit, filtered), in the MDCR database.

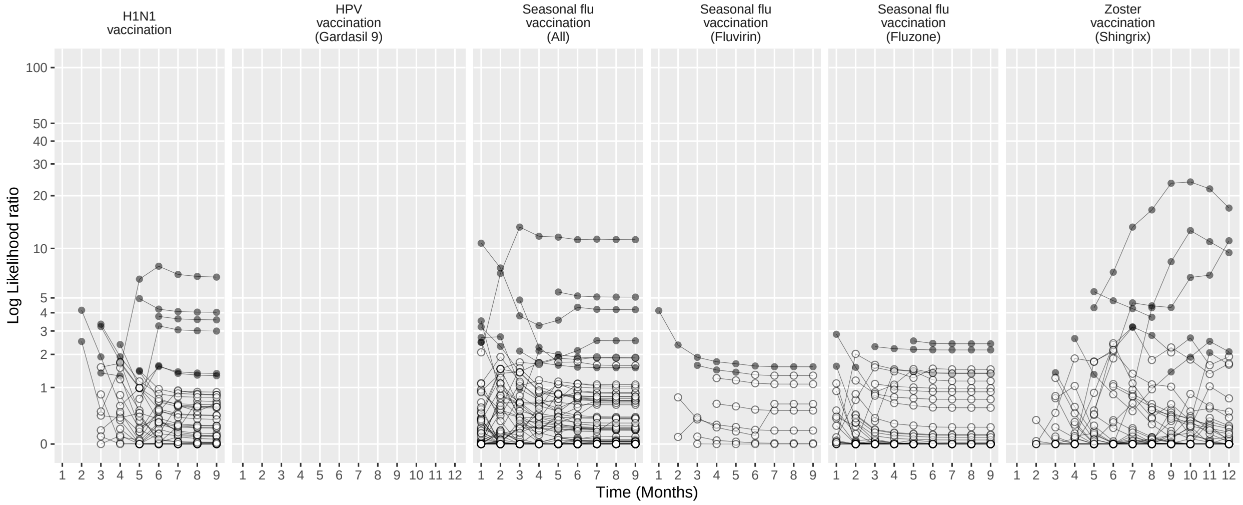

Figure 271: Negative control log likelihood ratios per month using the HistoricalComparator method (Age & sex adjusted, using TaR after historic visit, filtered), in the MDCR database.

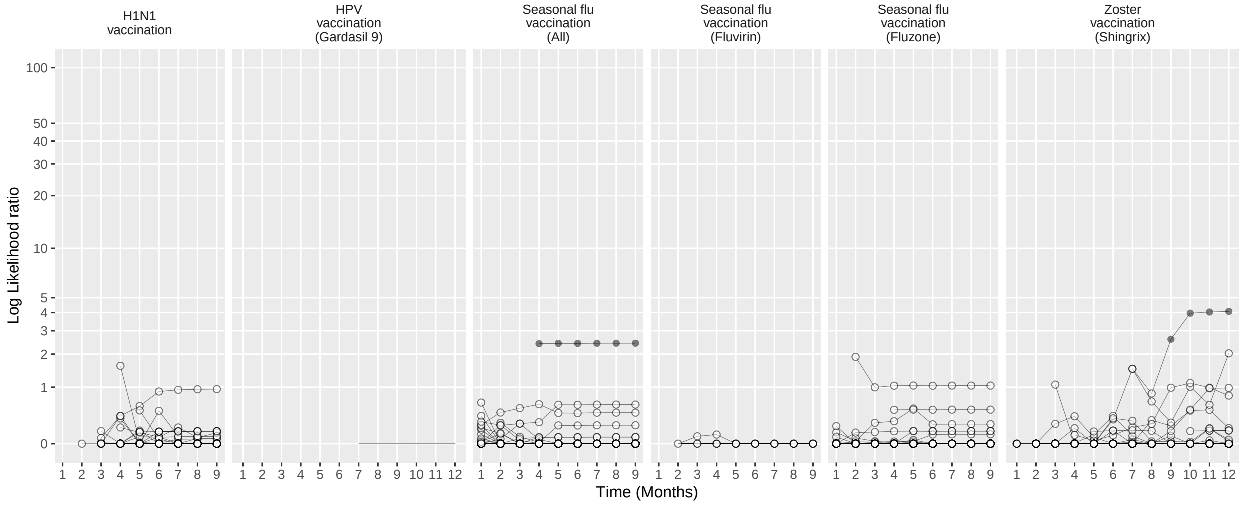

Figure 272: Negative control log likelihood ratios per month using the CohortMethod method (Unadjusted, using outpatient visits as comparator), in the MDCR database.

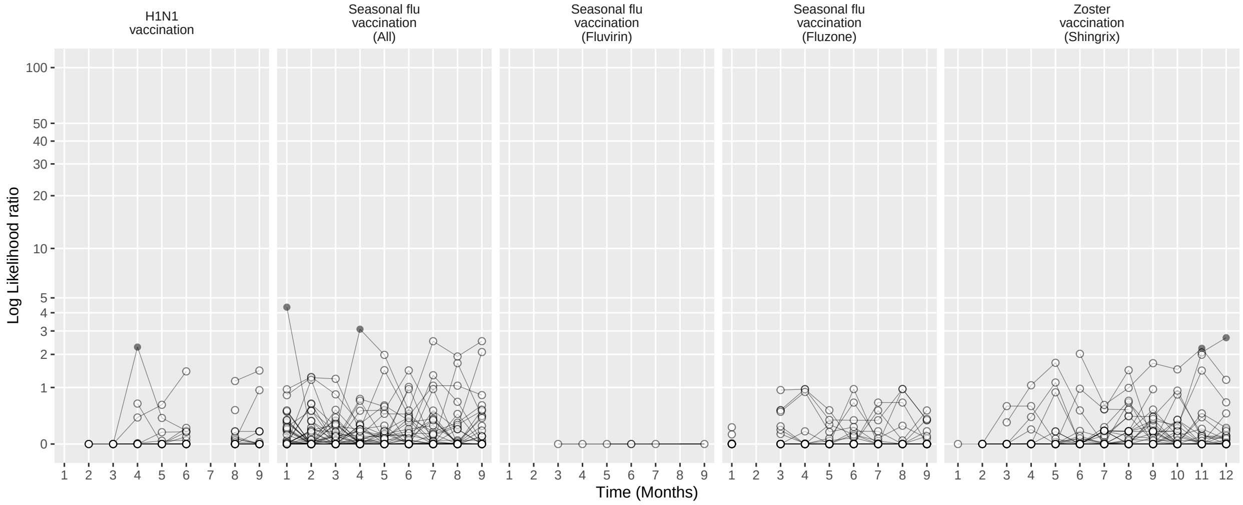

Figure 273: Negative control log likelihood ratios per month using the CohortMethod method (PS matching, using outpatient visits as comparator), in the MDCR database.

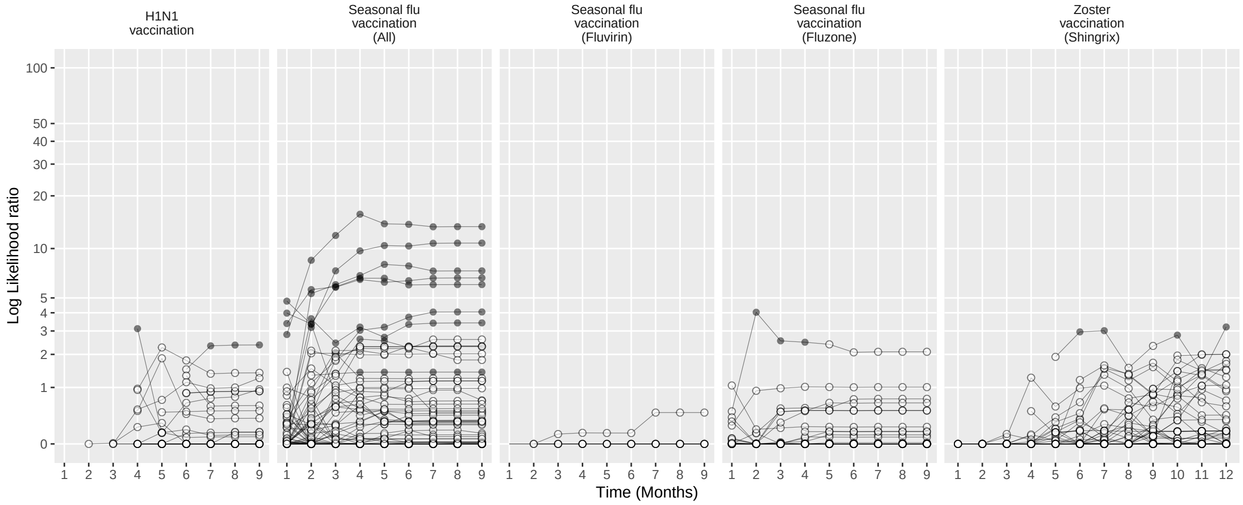

Figure 274: Negative control log likelihood ratios per month using the CohortMethod method (Unadjusted, using random days as comparator), in the MDCR database.

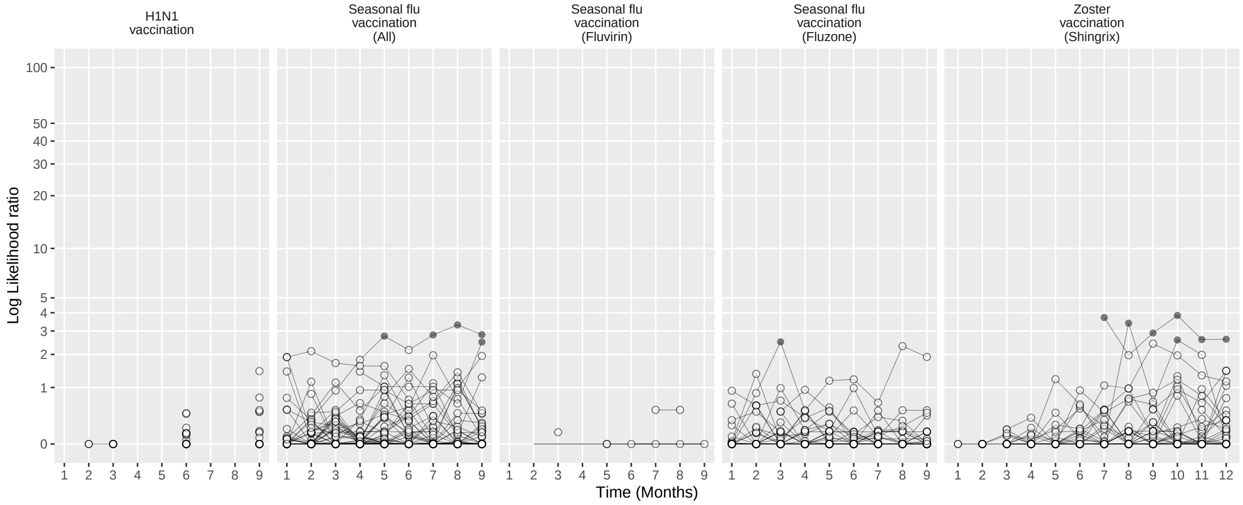

Figure 275: Negative control log likelihood ratios per month using the CohortMethod method (PS matching, using random days as comparator), in the MDCR database.

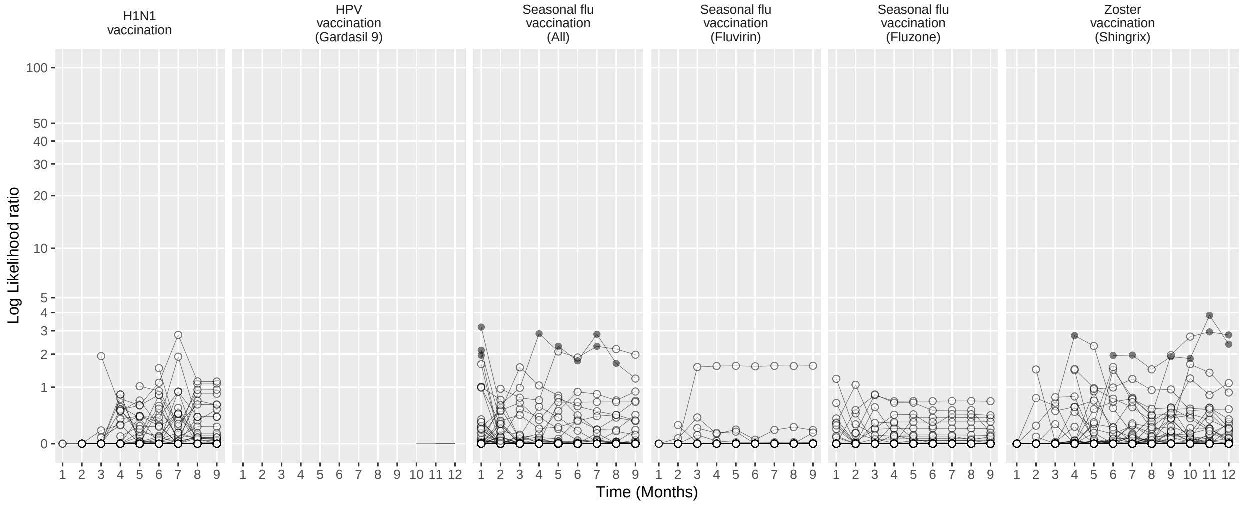

Figure 276: Negative control log likelihood ratios per month using the CohortMethod method (PS stratification, using outpatient visits as comparator), in the MDCR database.

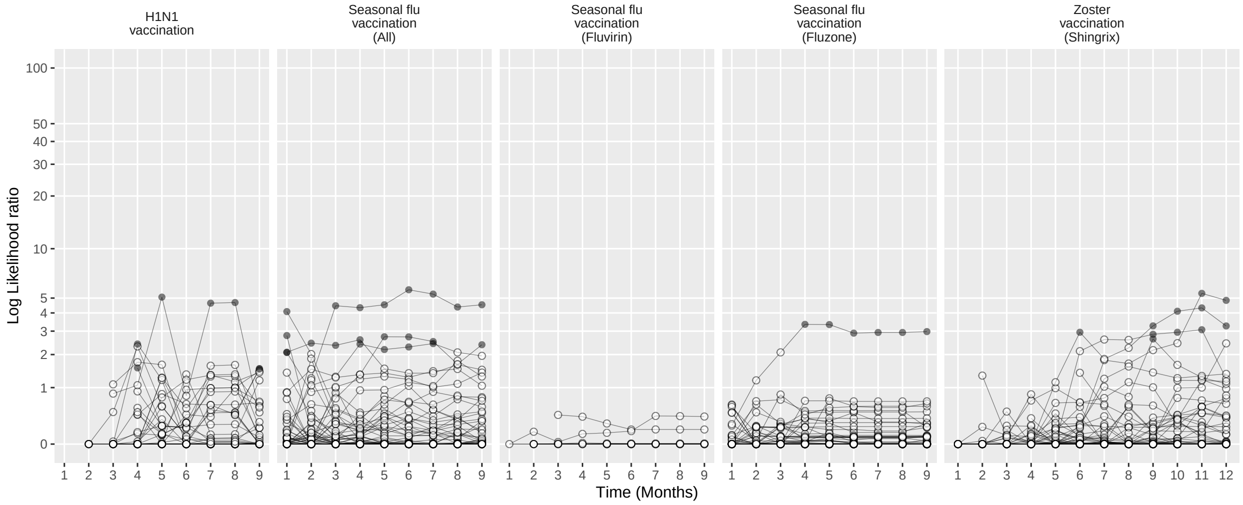

Figure 277: Negative control log likelihood ratios per month using the CohortMethod method (PS stratification, using random days as comparator), in the MDCR database.

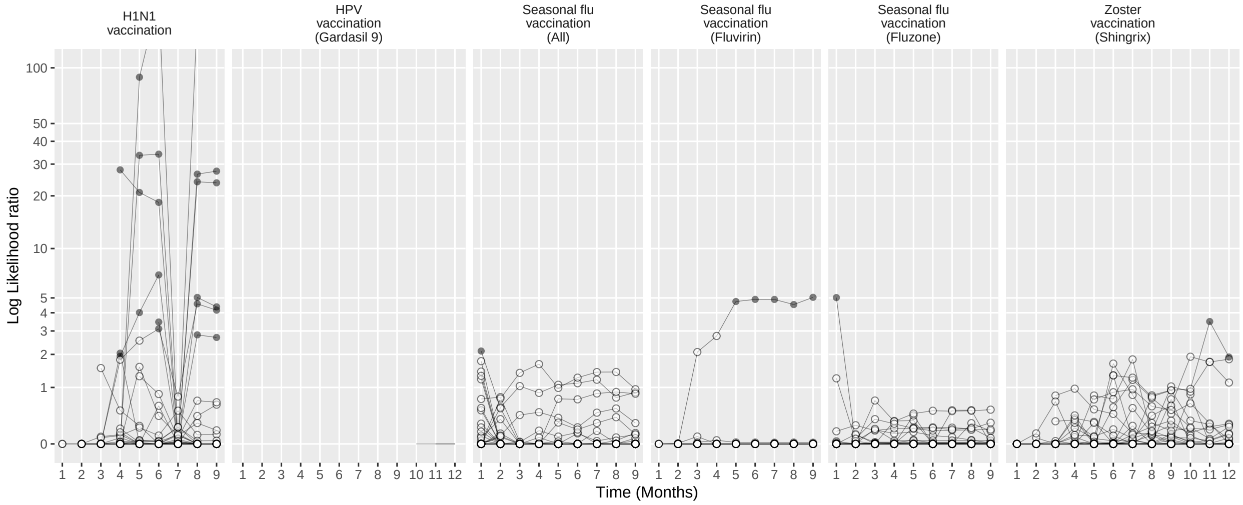

Figure 278: Negative control log likelihood ratios per month using the CohortMethod method (PS weighting, using outpatient visits as comparator), in the MDCR database.

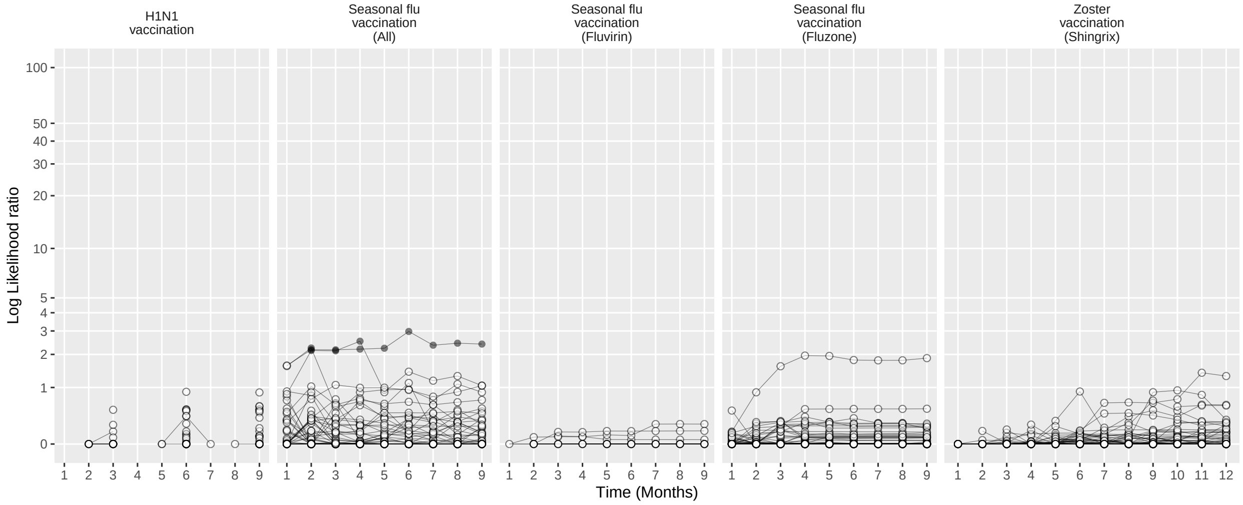

Figure 279: Negative control log likelihood ratios per month using the CohortMethod method (PS weighting, using random days as comparator), in the MDCR database.

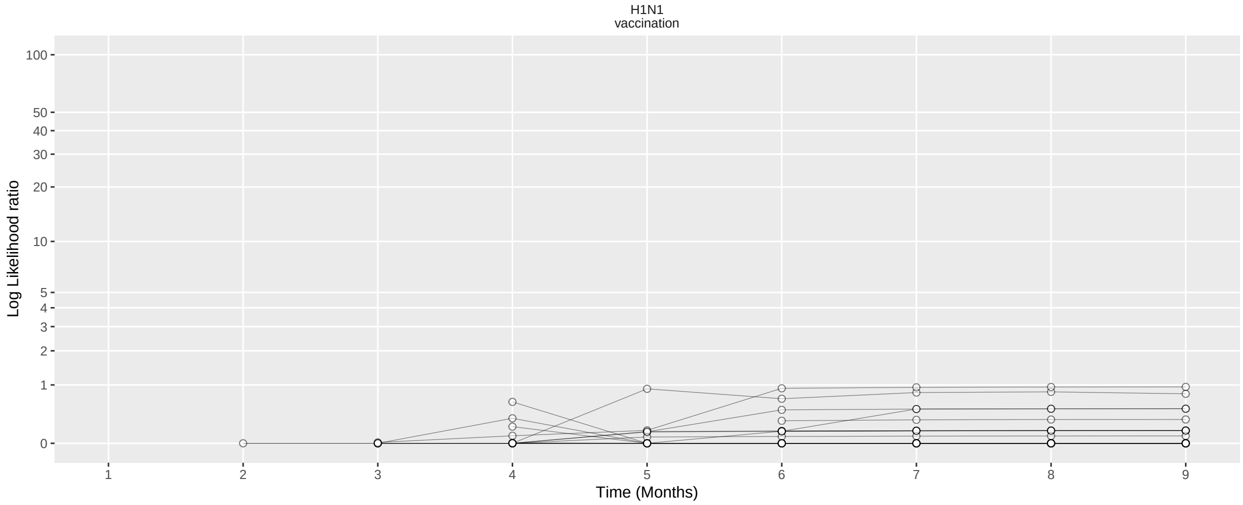

Figure 280: Negative control log likelihood ratios per month using the CohortMethod method (Per-month PS matching, using outpatient visits as comparator), in the MDCR database.

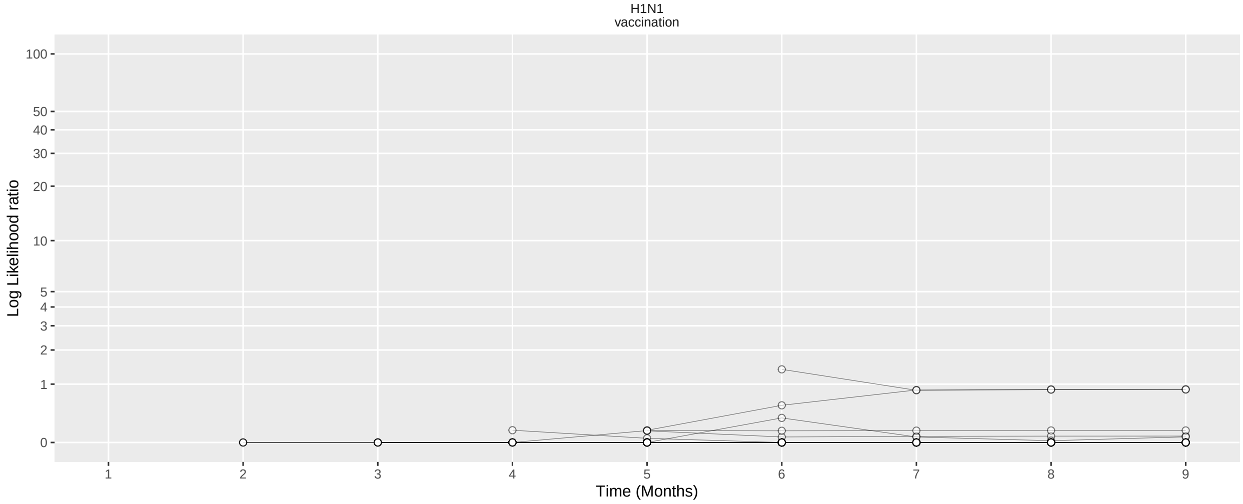

Figure 281: Negative control log likelihood ratios per month using the CohortMethod method (Per-month PS matching, using random days as comparator), in the MDCR database.

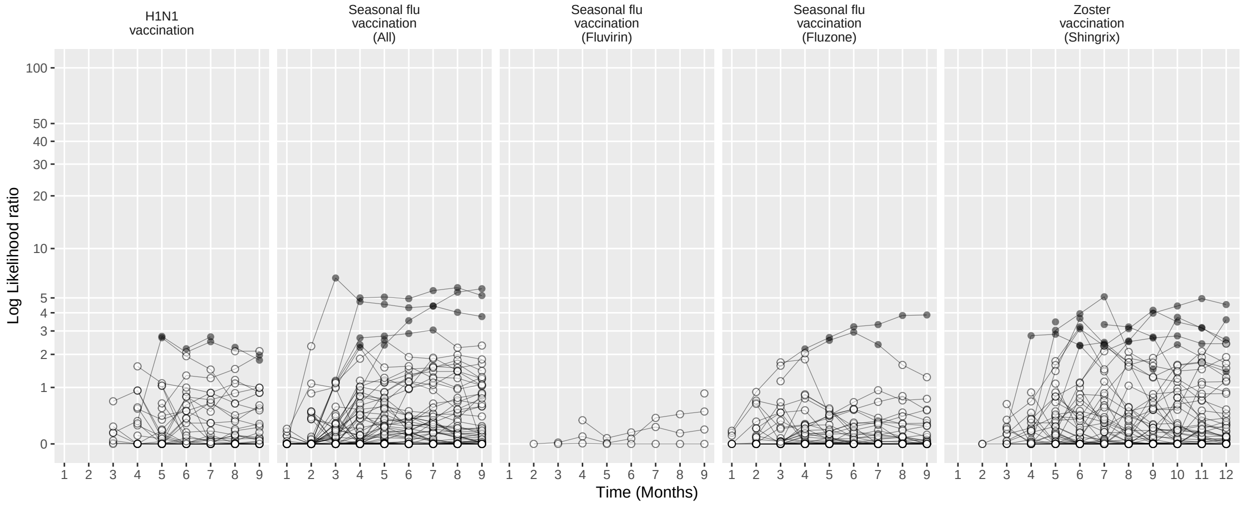

Figure 282: Negative control log likelihood ratios per month using the SCCS method (Unadjusted SCCS excluding pre-vaccination window), in the MDCR database.

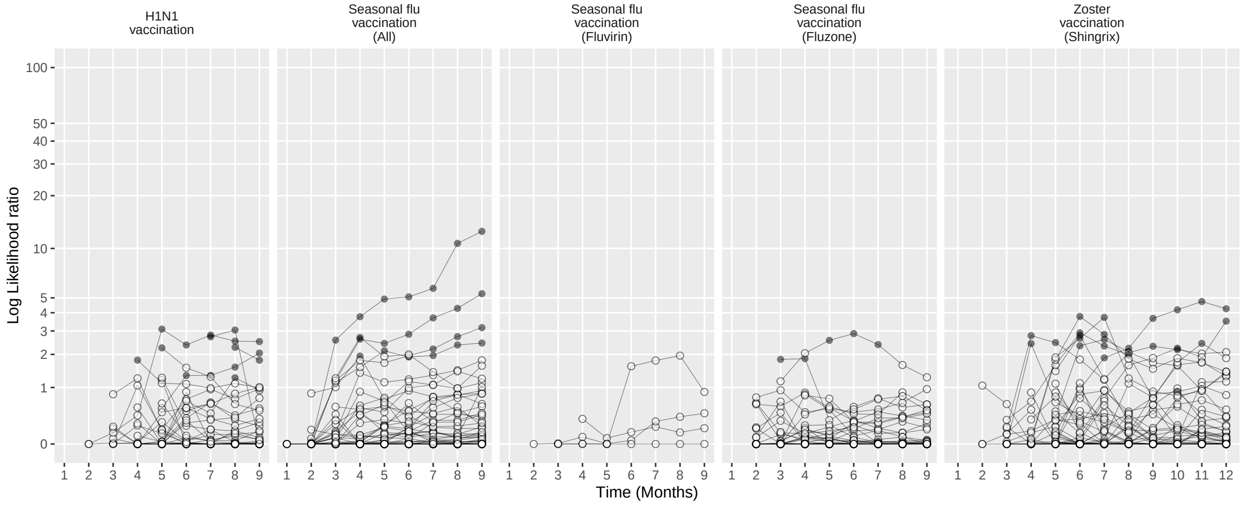

Figure 283: Negative control log likelihood ratios per month using the SCCS method (Age & season adjusted SCCS excluding pre-vaccination window), in the MDCR database.

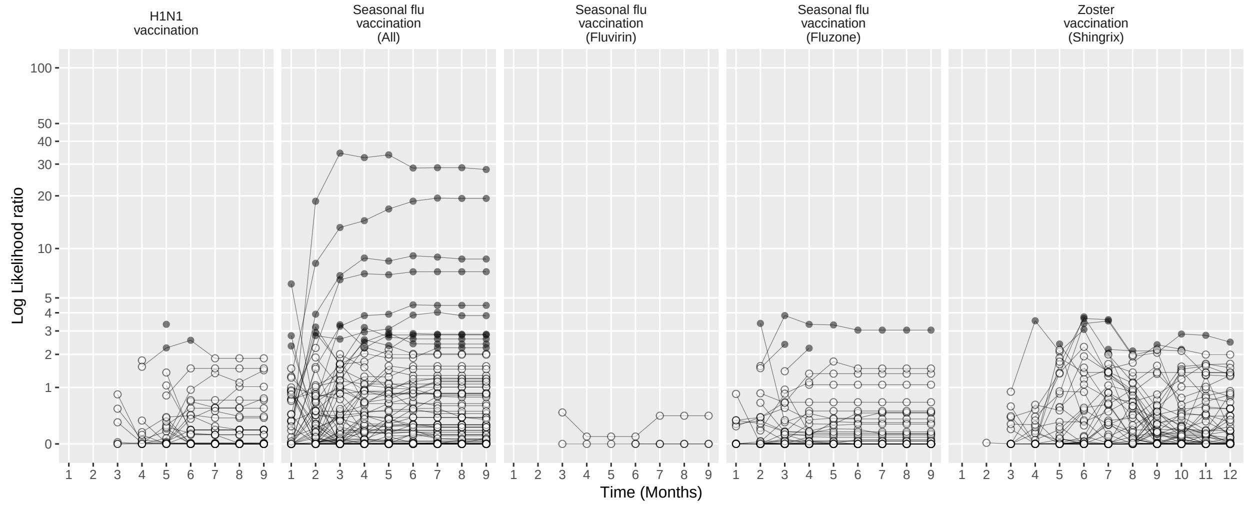

Figure 284: Negative control log likelihood ratios per month using the SCCS method (SCRI with prior control interval), in the MDCR database.

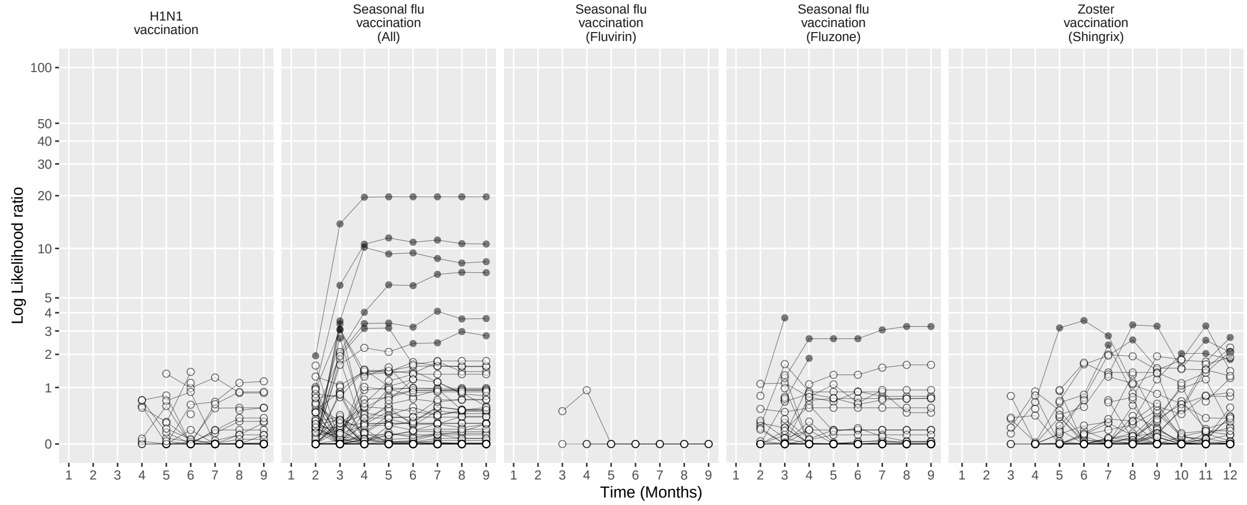

Figure 285: Negative control log likelihood ratios per month using the SCCS method (SCRI with posterior control interval), in the MDCR database.

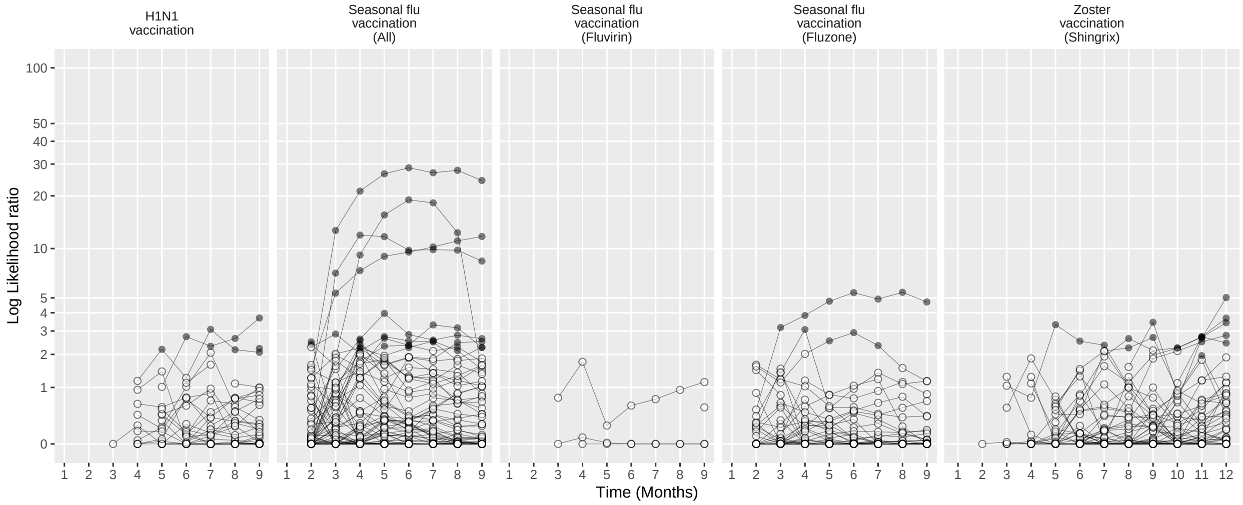

Figure 286: Negative control log likelihood ratios per month using the SCCS method (Unadjusted SCCS excluding all pre-vaccination time), in the MDCR database.

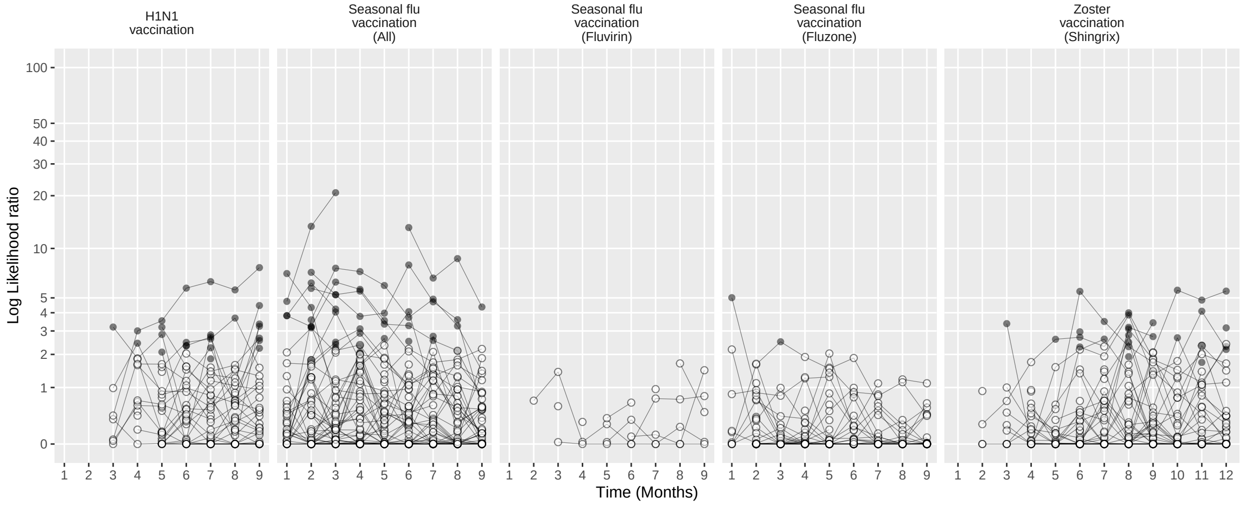

Figure 287: Negative control log likelihood ratios per month using the CaseControl method (Age & sex adjusted, using random controls), in the MDCR database.

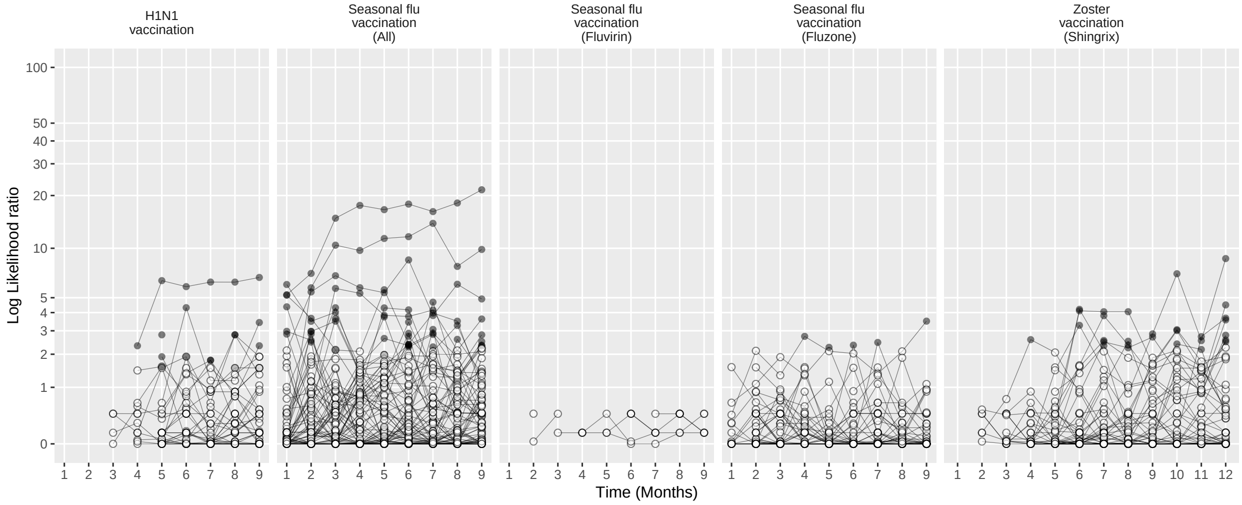

Figure 288: Negative control log likelihood ratios per month using the CaseControl method (Age & sex matched controls), in the MDCR database.

#### 15.4 Negative controls log likelihood ratios in CCAE

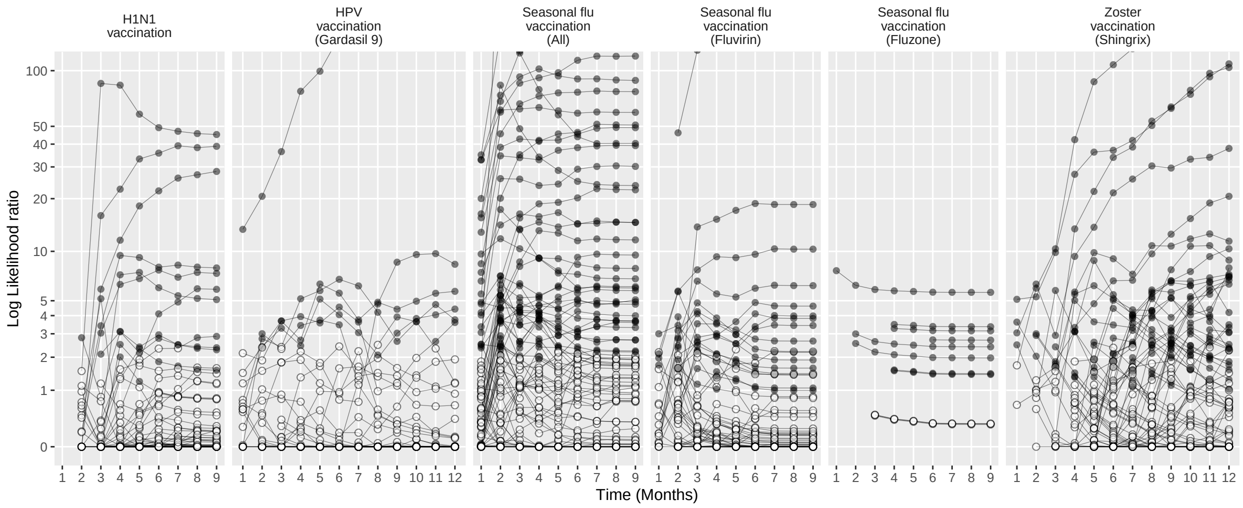

Figure 289: Negative control log likelihood ratios per month using the HistoricalComparator method (Unadjusted, using entire historic period), in the CCAE database.

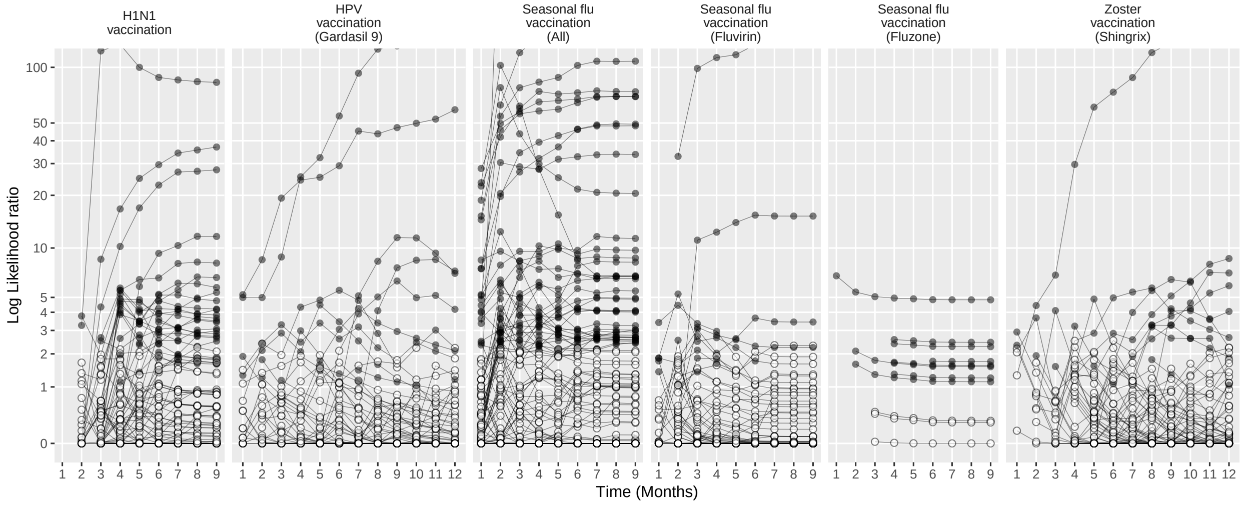

Figure 290: Negative control log likelihood ratios per month using the HistoricalComparator method (Age & sex adjusted, using entire historic period), in the CCAE database.

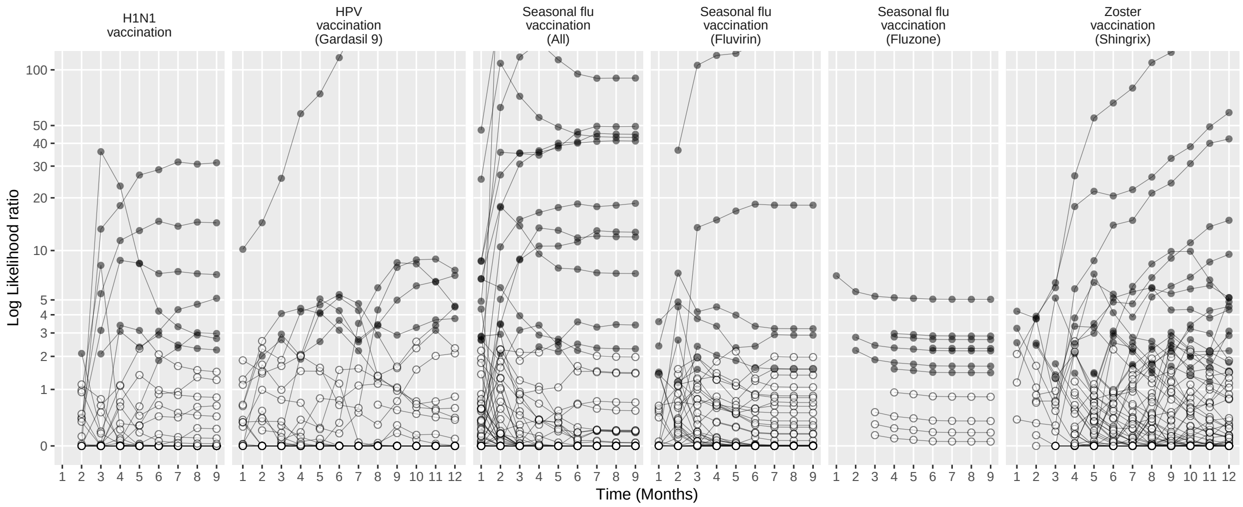

Figure 291: Negative control log likelihood ratios per month using the HistoricalComparator method (Unadjusted, using TaR after historic visit), in the CCAE database.

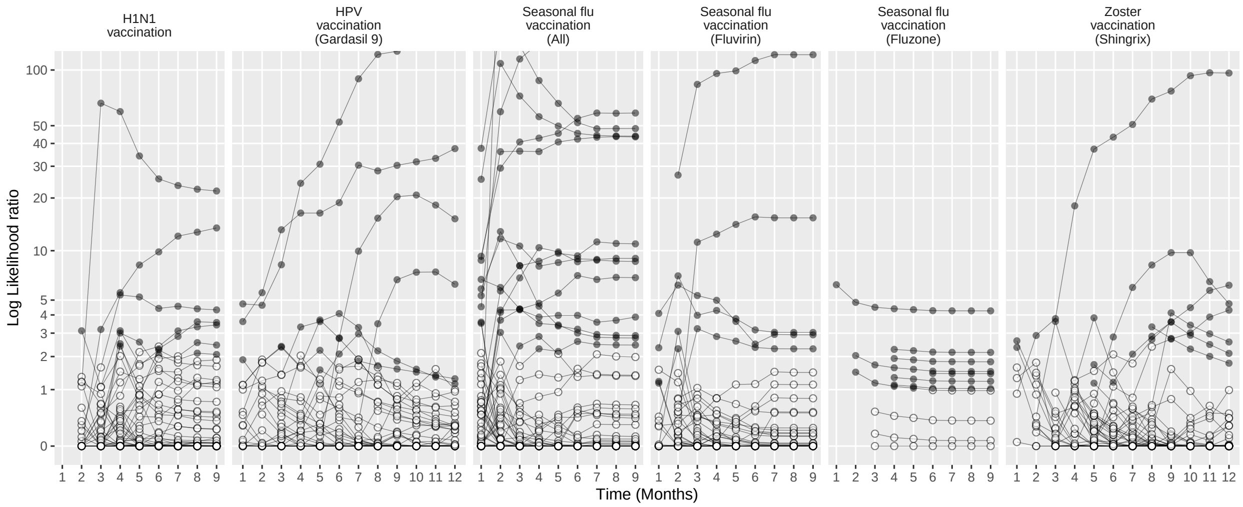

Figure 292: Negative control log likelihood ratios per month using the HistoricalComparator method (Age & sex adjusted, using TaR after historic visit), in the CCAE database.

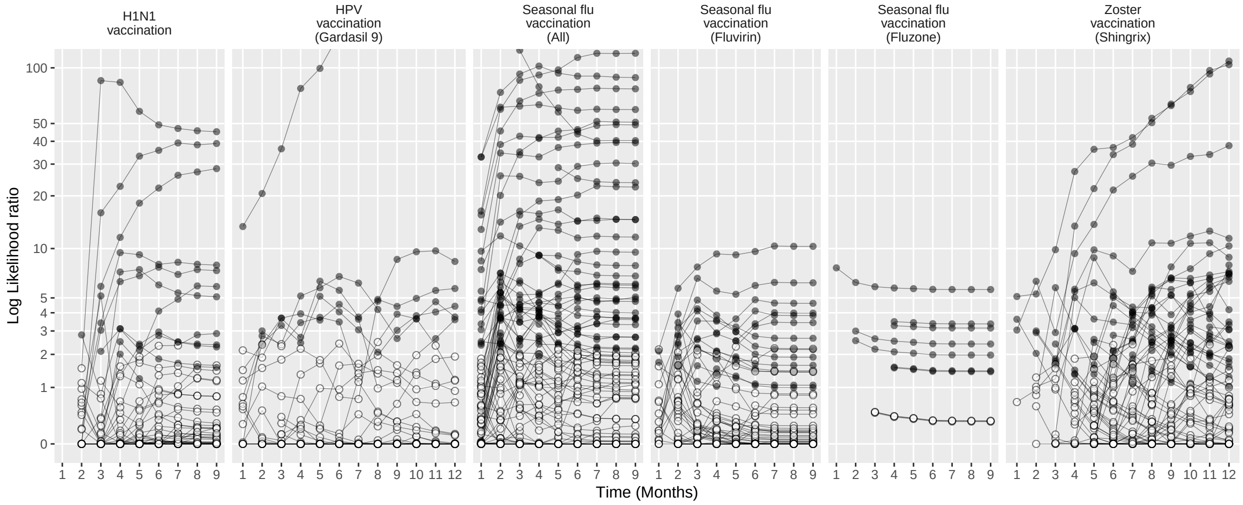

Figure 293: Negative control log likelihood ratios per month using the HistoricalComparator method (Unadjusted, using entire historic period, filtered), in the CCAE database.

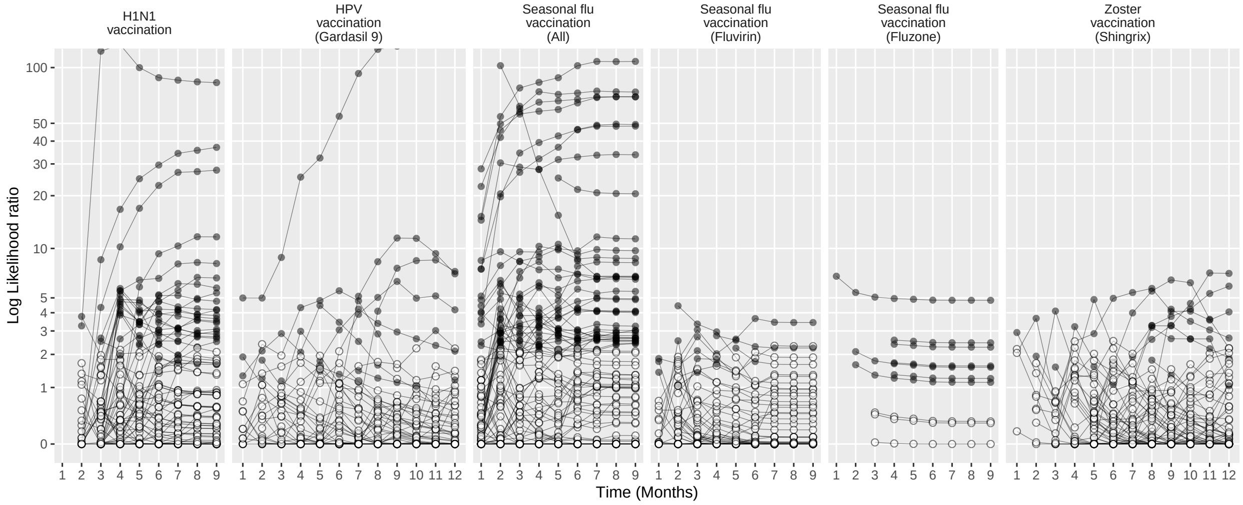

Figure 294: Negative control log likelihood ratios per month using the HistoricalComparator method (Age & sex adjusted, using entire historic period, filtered), in the CCAE database.

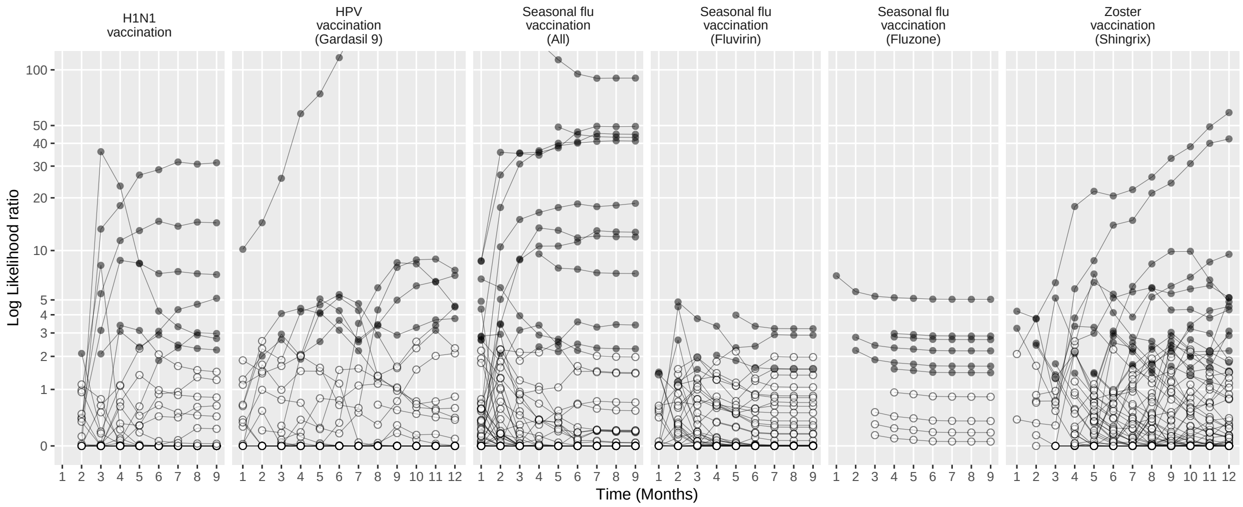

Figure 295: Negative control log likelihood ratios per month using the HistoricalComparator method (Unadjusted, using TaR after historic visit, filtered), in the CCAE database.

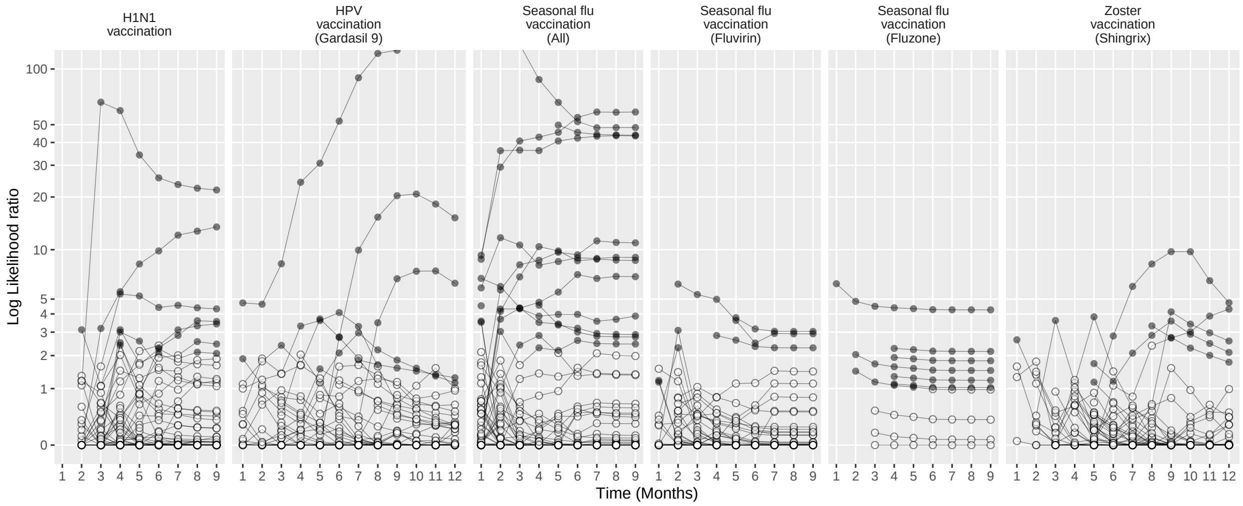

Figure 296: Negative control log likelihood ratios per month using the HistoricalComparator method (Age & sex adjusted, using TaR after historic visit, filtered), in the CCAE database.

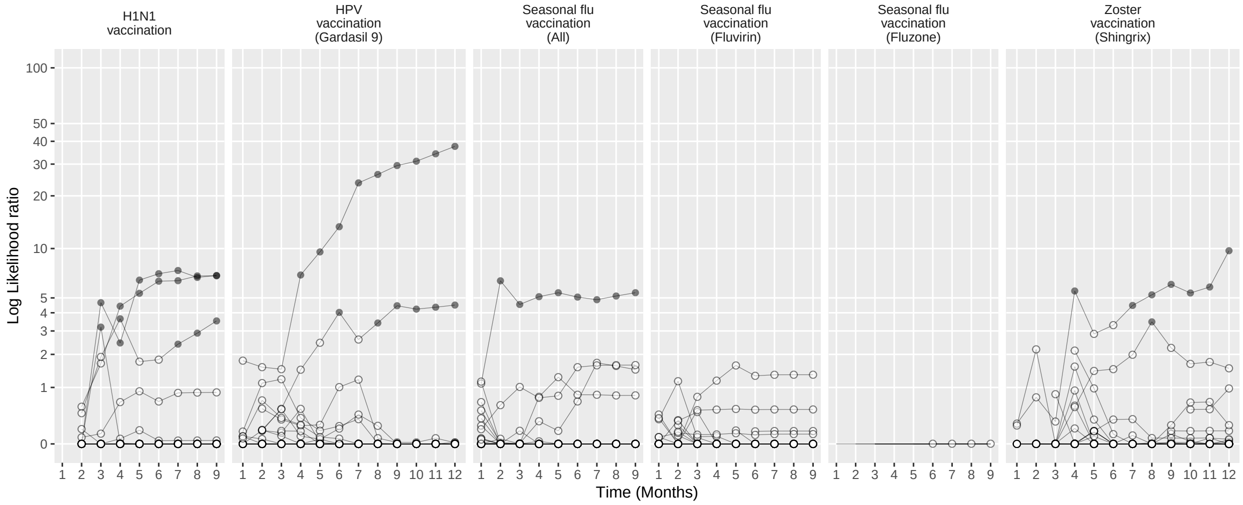

Figure 297: Negative control log likelihood ratios per month using the CohortMethod method (Unadjusted, using outpatient visits as comparator), in the CCAE database.

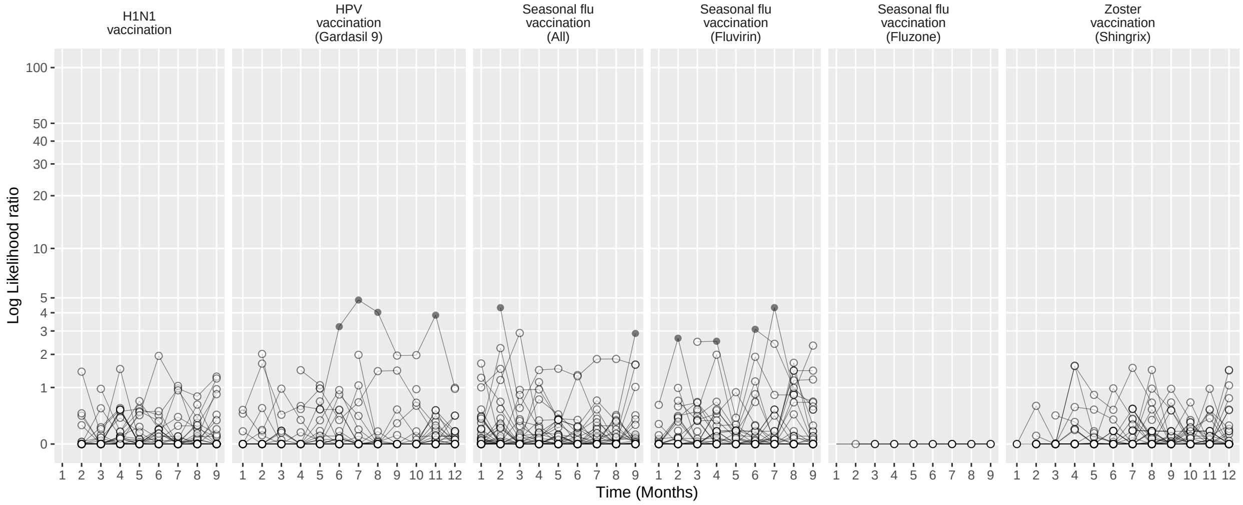

Figure 298: Negative control log likelihood ratios per month using the CohortMethod method (PS matching, using outpatient visits as comparator), in the CCAE database.

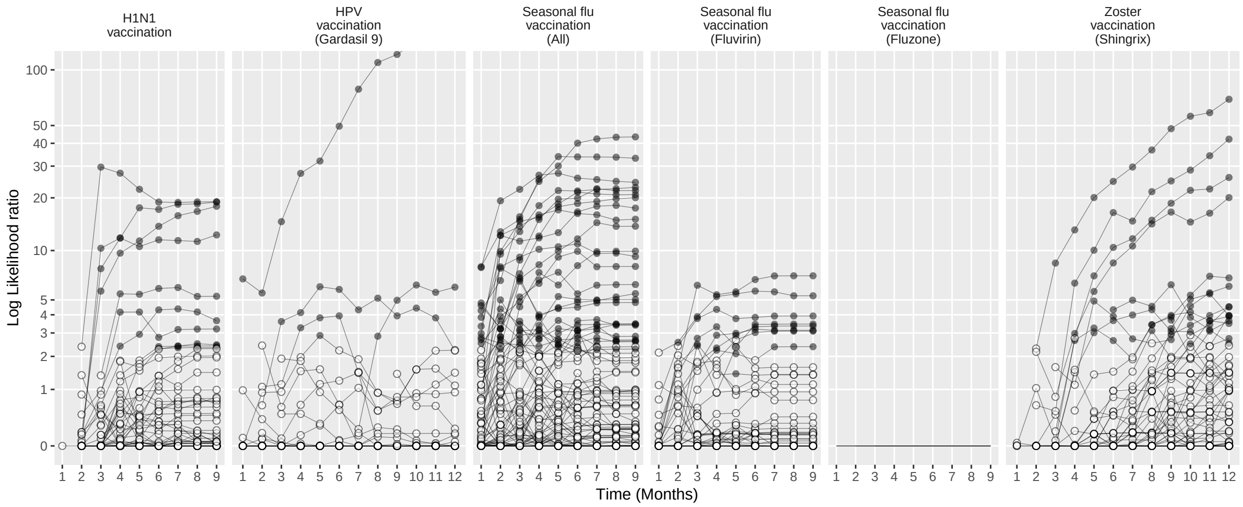

Figure 299: Negative control log likelihood ratios per month using the CohortMethod method (Unadjusted, using random days as comparator), in the CCAE database.

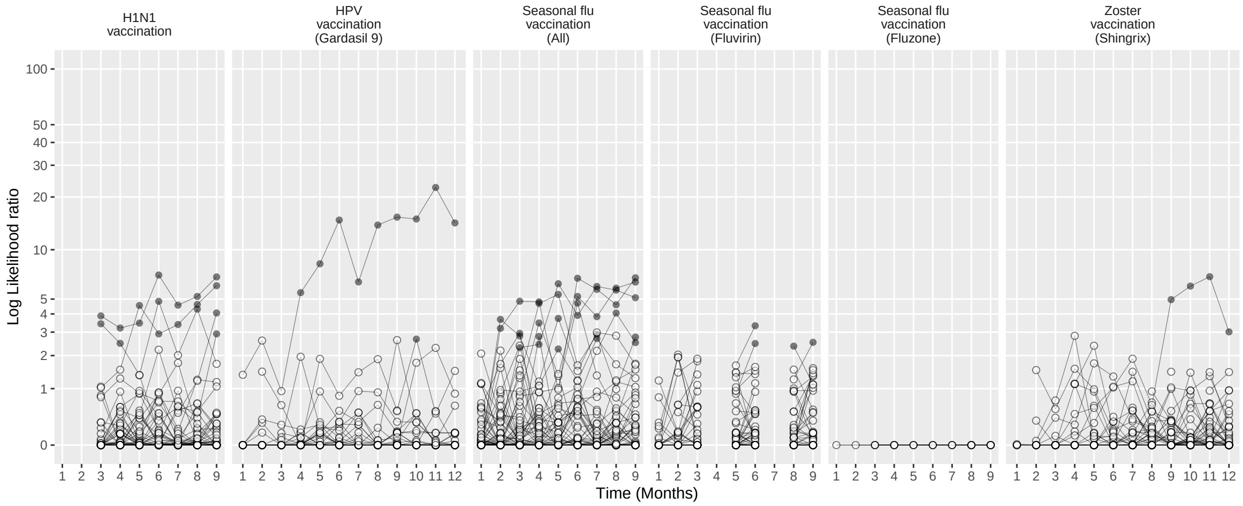

Figure 300: Negative control log likelihood ratios per month using the CohortMethod method (PS matching, using random days as comparator), in the CCAE database.

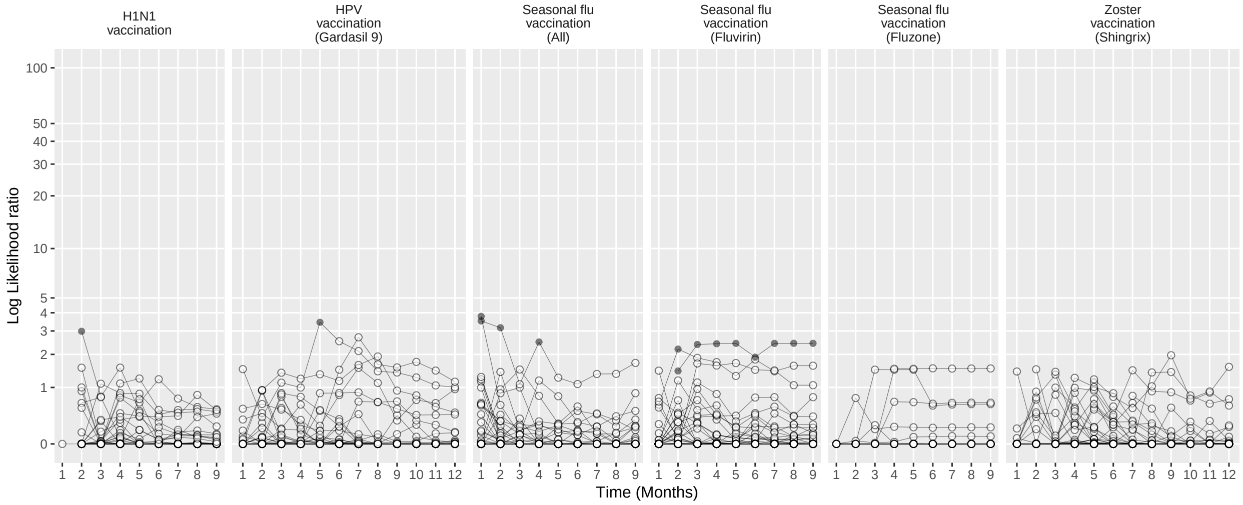

Figure 301: Negative control log likelihood ratios per month using the CohortMethod method (PS stratification, using outpatient visits as comparator), in the CCAE database.

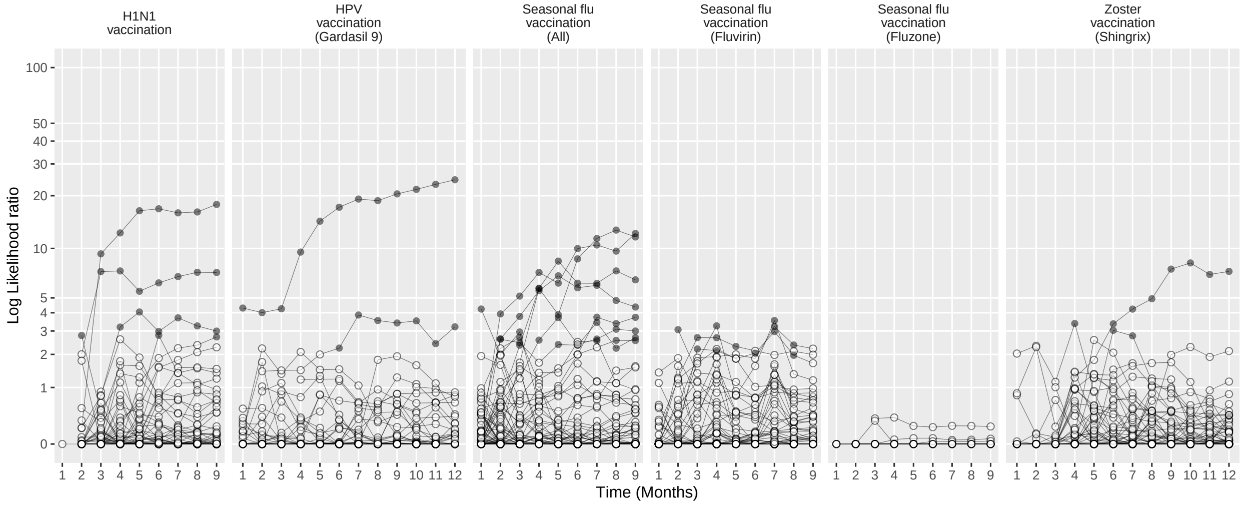

Figure 302: Negative control log likelihood ratios per month using the CohortMethod method (PS stratification, using random days as comparator), in the CCAE database.

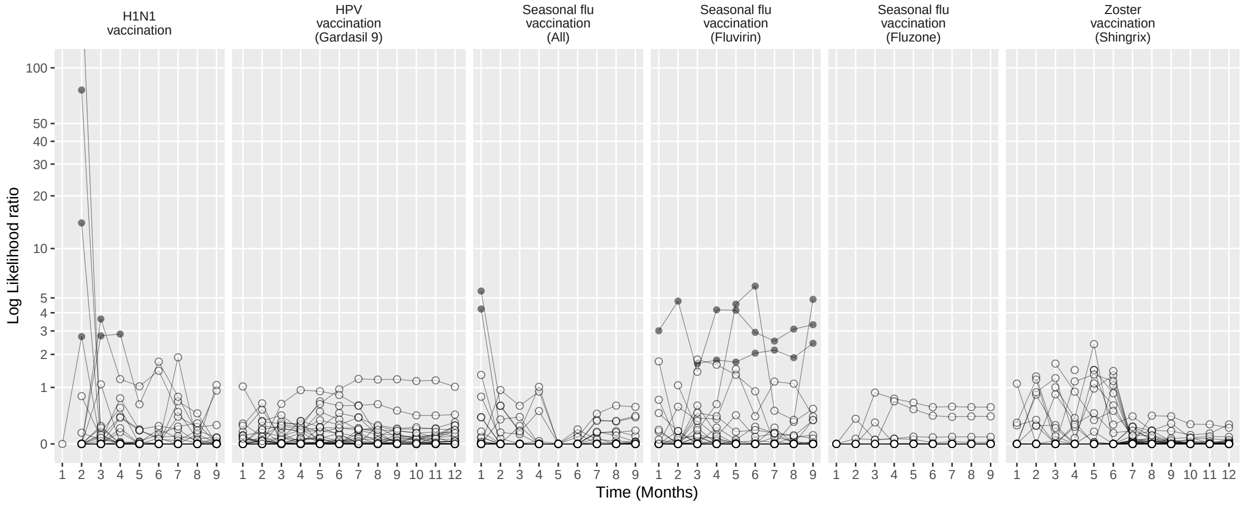

Figure 303: Negative control log likelihood ratios per month using the CohortMethod method (PS weighting, using outpatient visits as comparator), in the CCAE database.

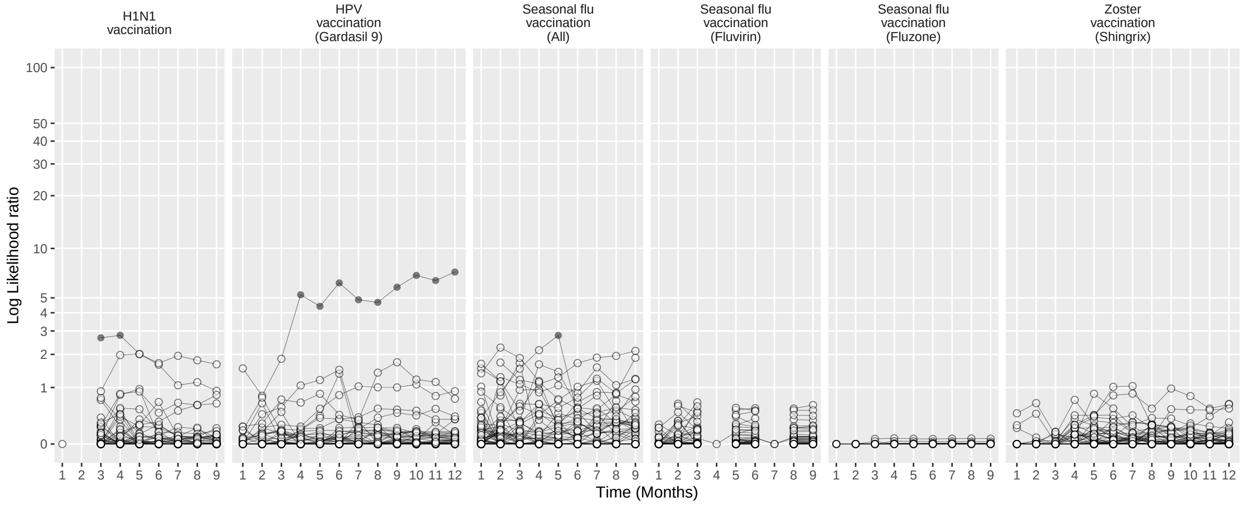

Figure 304: Negative control log likelihood ratios per month using the CohortMethod method (PS weighting, using random days as comparator), in the CCAE database.

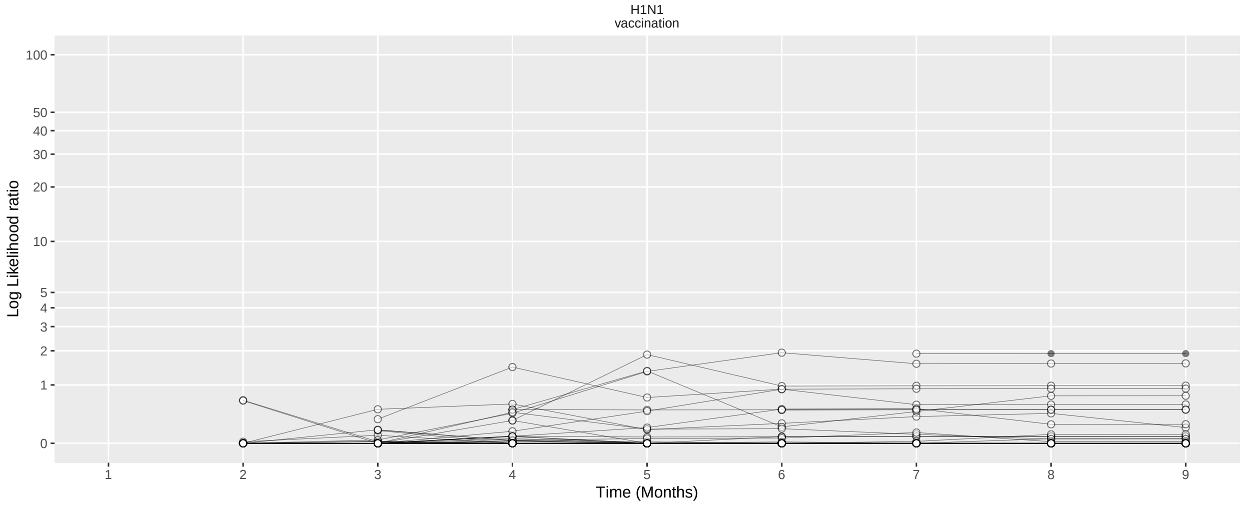

Figure 305: Negative control log likelihood ratios per month using the CohortMethod method (Per-month PS matching, using outpatient visits as comparator), in the CCAE database.

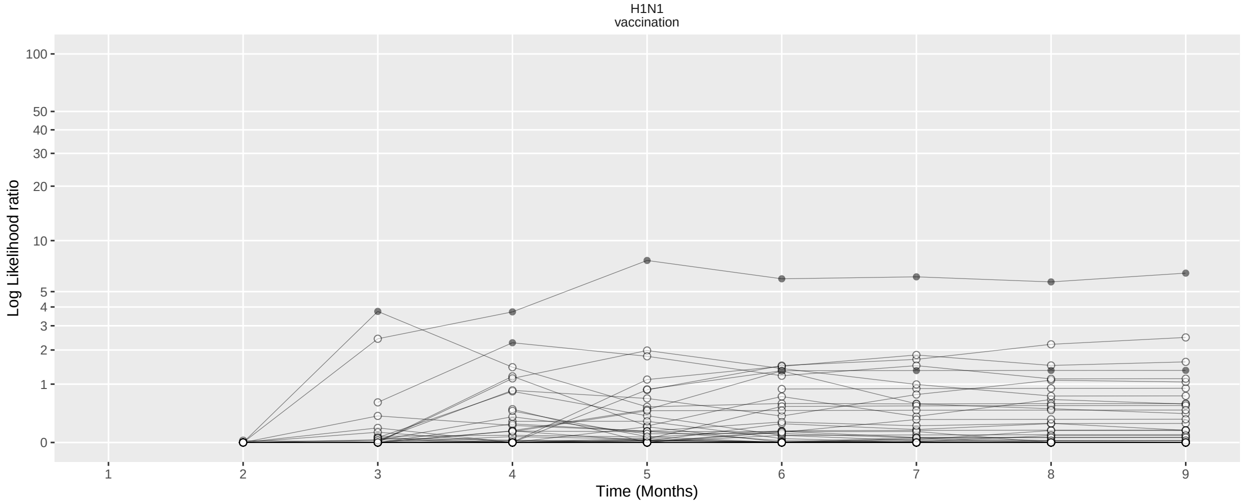

Figure 306: Negative control log likelihood ratios per month using the CohortMethod method (Per-month PS matching, using random days as comparator), in the CCAE database.

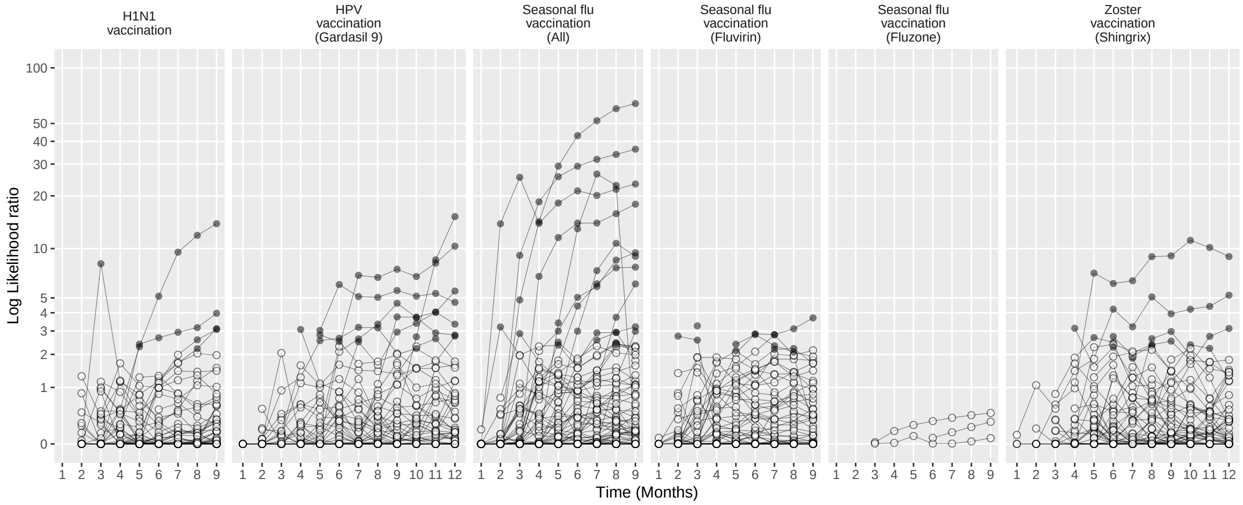

Figure 307: Negative control log likelihood ratios per month using the SCCS method (Unadjusted SCCS excluding pre-vaccination window), in the CCAE database.

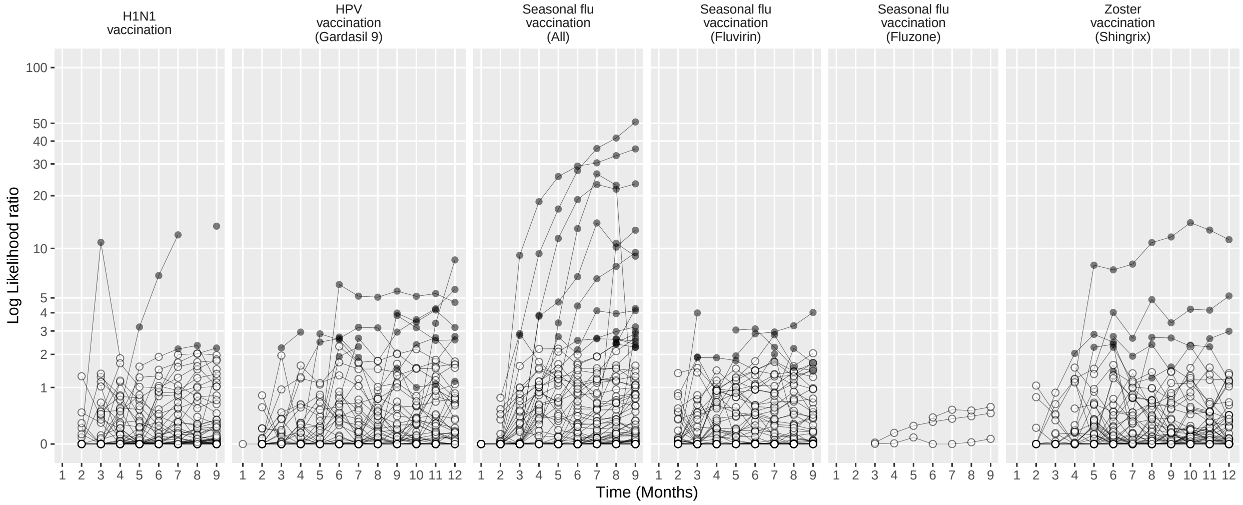

Figure 308: Negative control log likelihood ratios per month using the SCCS method (Age & season adjusted SCCS excluding pre-vaccination window), in the CCAE database.

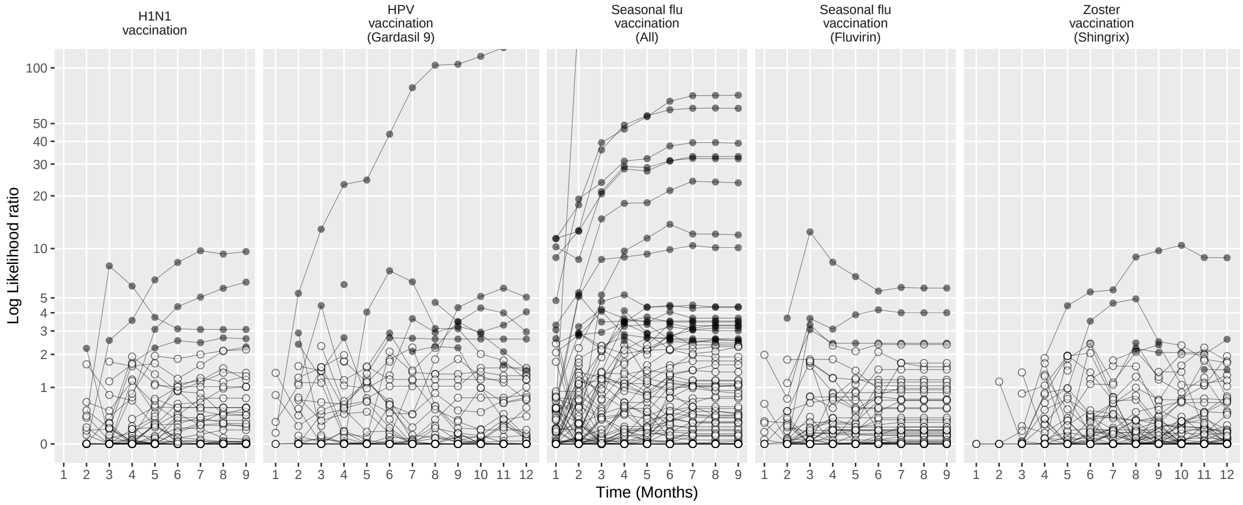

Figure 309: Negative control log likelihood ratios per month using the SCCS method (SCRI with prior control interval), in the CCAE database.

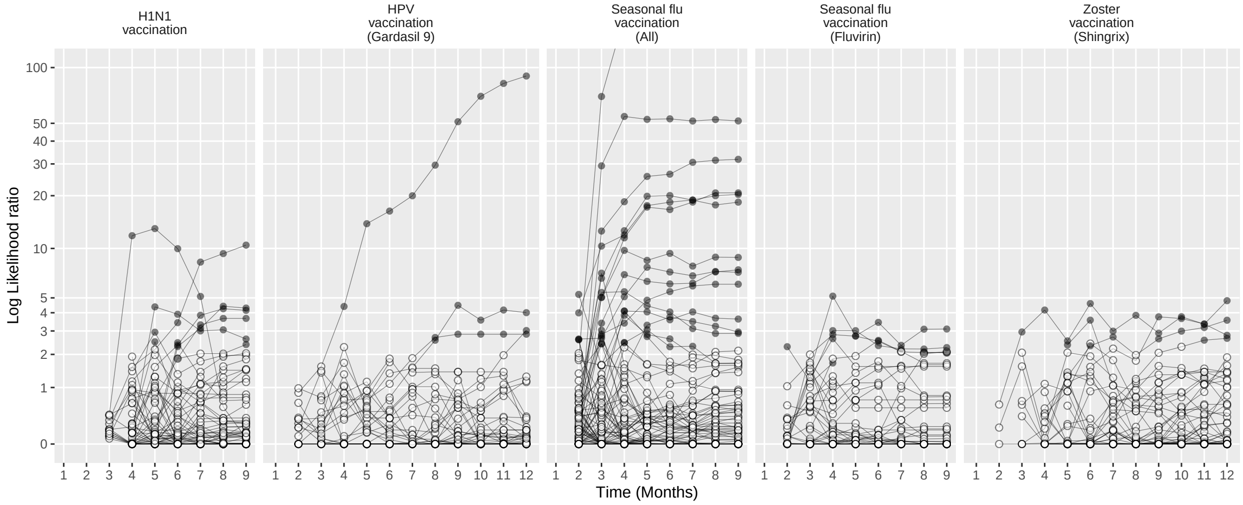

Figure 310: Negative control log likelihood ratios per month using the SCCS method (SCRI with posterior control interval), in the CCAE database.

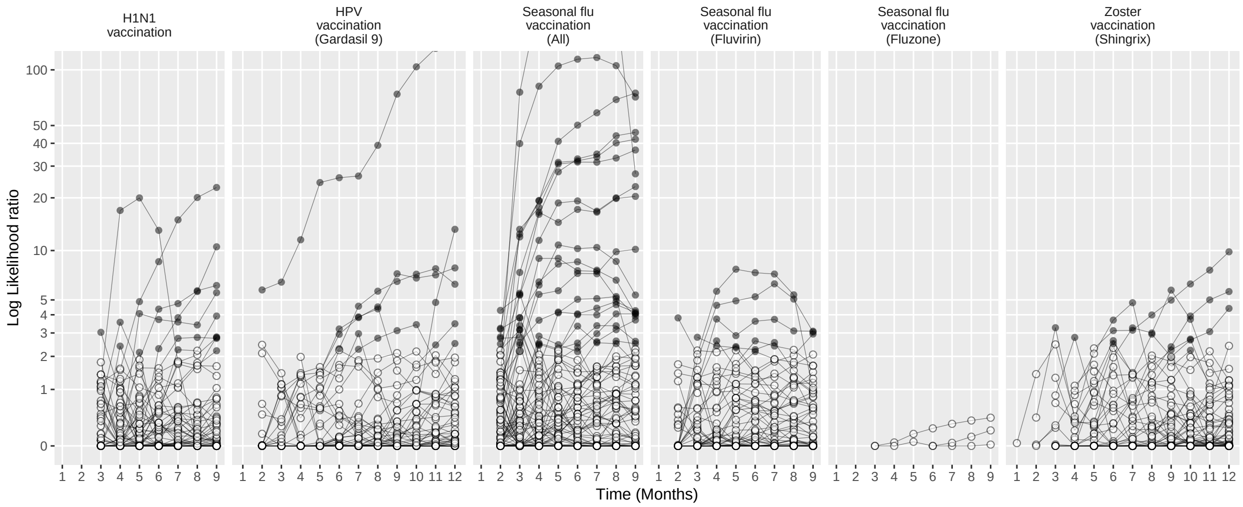

Figure 311: Negative control log likelihood ratios per month using the SCCS method (Unadjusted SCCS excluding all pre-vaccination time), in the CCAE database.

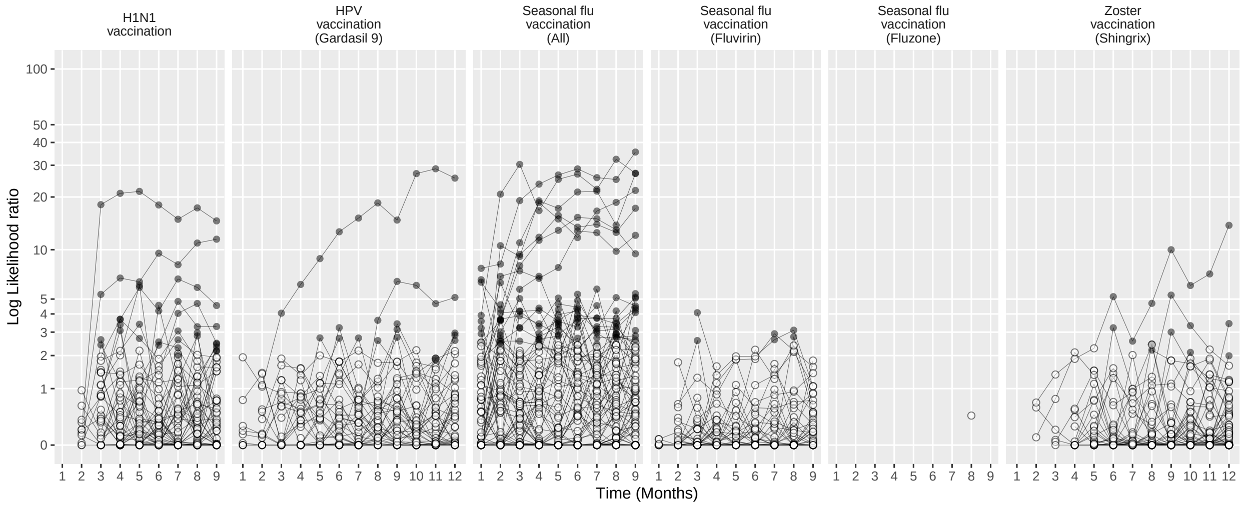

Figure 312: Negative control log likelihood ratios per month using the CaseControl method (Age & sex adjusted, using random controls), in the CCAE database.

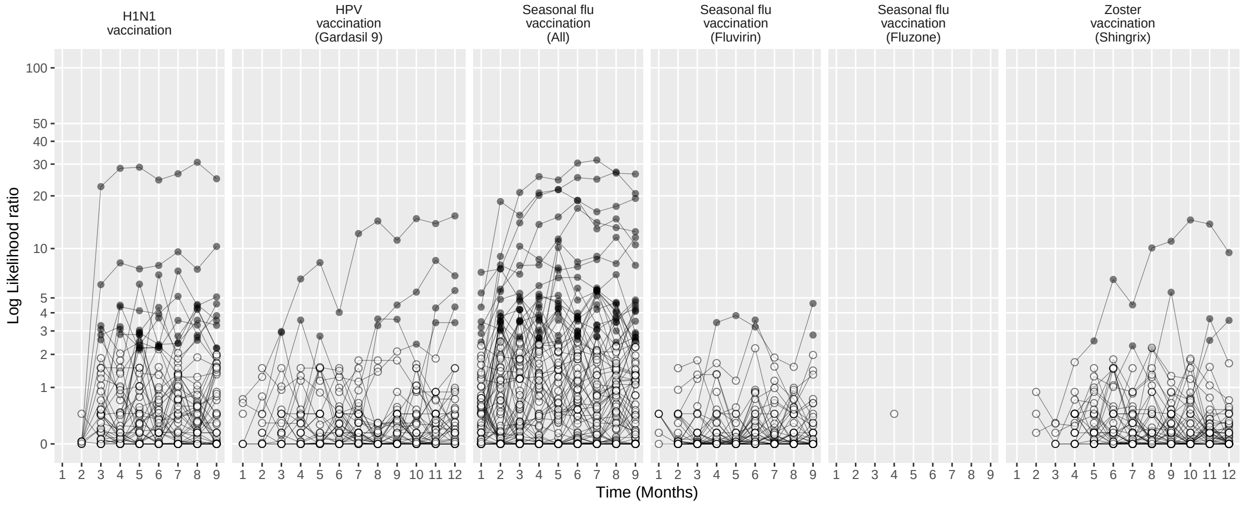

Figure 313: Negative control log likelihood ratios per month using the CaseControl method (Age & sex matched controls), in the CCAE database.
